# Evaluating Regional Diversity in Scientific Communication: A Comparative Analysis of COVID-19 Preprints and Peer-Reviewed Publications

**DOI:** 10.1101/2025.01.04.25319994

**Authors:** Dong Hyun Kim, Kyu Lee Jeon, Seng Chan You

**Affiliations:** Yonsei University College of Medicine, Seoul, Korea; Department of Biomedical Systems Informatics, Yonsei University College of Medicine, Seoul, Korea; Institute for Innovation in Digital Healthcare, Yonsei University, Seoul, Korea

## Abstract

**Background:** The unprecedented COVID-19 pandemic has triggered extensive global research, leading to an overwhelming surge in publications with surge of preprints. Despite the proliferation of preprints during the pandemic, the specific details of their implications for global diversity, along with their utility, remain underexplored. In this study, we assess the contribution of COVID-19 preprints in diverse aspects.

**Methods:** We collected COVID-19-related peer-reviewed papers and preprints from SCOPUS and MedRxiv, respectively, between December 2019 to November 2022. We analyzed four key aspects of scientific communication: 1) international co-authorship patterns using network analysis and eigenvector centrality, 2) publication patterns through relative ratio analysis comparing preprint to peer-reviewed paper counts, 3) social media dissemination through analysis of X (formerly Twitter) post quotations, and 4) citation impact by comparing citation counts between peer-reviewed papers with and without preprint history. All analyses were stratified by country income levels and geographical regions.

**Results:** Network analysis revealed higher co-authorship diversity in preprints, with Sub-Saharan Africa, Latin America, and the Caribbean showing 3.9 to 4.5 times higher eigenvector centrality compared to peer-reviewed papers. Countries with lower GDP showed significantly higher preprint publication ratios (correlation coefficient: −0.38, p-value < 0.001). Social media analysis demonstrated higher engagement with preprints, as evidenced by higher median numbers of social media quotations for preprints across all income groups. Peer-reviewed papers with preprint history received significantly higher citations (median: 10, IQR: 3-30) compared to those without (median: 5, IQR: 1-15, p-value < 0.001), particularly pronounced in low- and middle-income countries.

**Conclusion:** This study demonstrates the significant role of preprints in advancing regional diversity in scientific communication during the COVID-19 pandemic. Our findings show enhanced international collaboration through preprints, particularly benefiting researchers from lower-income regions, higher social media engagement across income groups, and increased citation impact for papers with preprint history. These results highlight preprints as an important tool for promoting more equitable global scientific discourse.

## Introduction

The COVID-19 pandemic has triggered extensive research efforts worldwide, with a substantial body of work being produced to understand and combat the virus.[1–5] However, much of this research has been concentrated in high-income countries, leading to disparities in the geographical representation of scientific output. Despite the global nature of the pandemic, researchers from low- and middle-income countries (LMICs) have been underrepresented, further highlighting the existing inequities in the global research landscape.

In this context, preprints have gained attention as a potential solution to overcome the limitations of traditional peer-reviewed journals. The conventional academic publishing process often faces issues such as lengthy and complex review processes, high publication costs, and sometimes bias based on authors’ affiliations or nationalities.[6] In contrast, preprints, which involve sharing preliminary manuscripts on public servers before formal peer review, offer advantages such as rapid feedback collection, idea prioritization, and improved access to research.[7, 8] The use of preprints surged during the COVID-19 pandemic, particularly in medical research.[2, 9–11] The preprint system enables swift dissemination of information in emergency situations, complementing the limitations of traditional publishing systems.

Although numerous preprints have been published via open server during this pandemic,[12] little attention has been given to their implications for the inclusive science communication.[13] Meanwhile, social media tools are increasingly part of the research workflow, offering new dissemination and communication possibilities to researchers.[14]

The aim of this study is to analyze the impact of COVID-19-related preprints and peer-reviewed papers on global scientific communication. Specifically, we assess the role of preprints in fostering more inclusive research practices by examining regional diversity in authorship, publication, and dissemination. Our analysis draws on data from both publication databases and social media (X, formerly Twitter) to compare the reach and engagement of preprints and peer-reviewed papers, with a focus on regional disparities.

## Methods

### Data Collection

This study collected data from Scopus and medRxiv to assess regional diversity in scientific communication during COVID-19. Scopus is a peer-reviewed literature database, while medRxiv is a health sciences preprint repository.[15, 16] COVID-19-related articles from December 1, 2019, to November 31, 2022, were collected using keywords such as “COVID-19”, “SARS-CoV-2”, and “coronavirus disease 2019” (**Supplementary Appendix 1**).[1] By April 20, 2023, metadata including titles, digital object identifiers (DOIs), and author affiliations were obtained, resulting in 277,366 papers from Scopus and 19,584 from medRxiv.

For regional analysis, the author’s affiliations were processed using the ‘hugofitipaldi/affiliation’ R package. X (formerly Twitter) data were integrated using Altmetric and the X/Twitter API, tracking posts linking to papers and categorizing users into demographic groups such as “public”, “researcher”, “practitioner”, and “science communicator”.[17] Citation data were retrieved via the CrossRef API to compare citation counts between peer-reviewed papers with and without preprint history.[18]

### Regional and Income-Level Categorization

For the analysis of regional and income-level diversity, country information was grouped based on the World Bank’s 2022 regional and income group classifications.[19] Income groups were defined as follows: low income (gross national income [GNI] per capita of $1,135 or less), lower-middle income ($1,136 to $4,465), upper-middle income ($4,466 to $13,845), and high income ($13,846 or more). Geographical regions were grouped according to the World Bank’s administrative classifications, including East Asia and Pacific, Europe and Central Asia, Latin America and the Caribbean, Middle East and North Africa, North America, South Asia, and Sub-Saharan Africa. To address previous findings that Europe and North America are often overrepresented in peer-reviewed journals, we distinguished Europe from Central Asia for clearer analysis.

### Global Co-authorship Networks

To identify patterns of international collaboration between regions, the Inter-regional Co-authorship Count was defined to quantify the number of collaborative papers between researchers from different regions. This was calculated as follow:

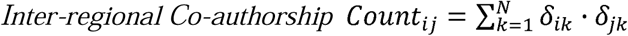

where *N* represents the total number of papers, and *δ_ik_* and *δ_jk_* indicate whether paper *k* has authors from regions *i*, and *j*, respectively.

To evaluate the influence of each region, we used eigenvector centrality (EVC) and calculated the Co-authorship Enhancement Index (CEI), defined as:

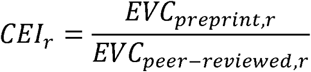

This ratio indicates whether a region *r* was more central in the preprint network compared to the peer-reviewed network. Eigenvector centrality assigns scores to nodes (regions) based on their connections to influential nodes, weighted by normalized inter-regional co-authorship counts. This measure highlights which regions hold the most central and influential positions in global research collaboration, allowing us to compare the roles of regions in preprint versus peer-reviewed networks.

### Publication of Research

To analyze differences in publication between preprints and peer-reviewed articles across different regions, we calculated the Publication Relative Ratio (PubRR) for each country *c*.

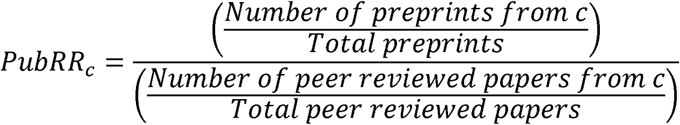

PubRR represents the relative ratio of a country’s share of first authors preprints to its share in peer-reviewed papers. This ratio allows us to determine whether researchers in a country are more inclined to publish preprints compared to peer-reviewed articles.

### Social Media Dissemination of Research

To evaluate how research is disseminated on social media, we analyzed X posts of preprints and peer-reviewed papers. Social Media Quotation Count quantifies the number of X posts referencing papers from a country. A higher quotation count indicates broader dissemination of that country’s research on X.

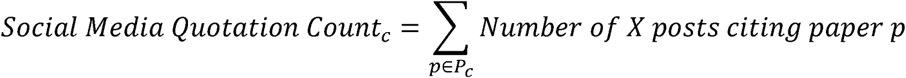

*where P_c_* represents the set of papers with first authors from country *c*.

Social Media Quotation Relative Ratio (QuoRR) compares the share of social media quotation count for preprints versus peer-reviewed articles for each country.

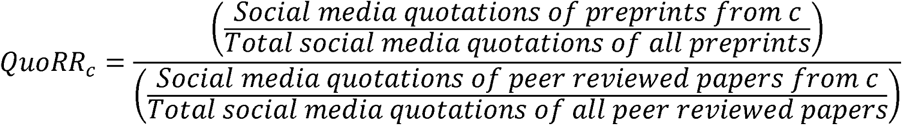

### Social Media Readership Patterns

To understand how social media users engage with research, we used the Social Media Readership Relative Ratio (ReadRR). ReadRR measures the relative ratio of social media users citing preprints versus peer-reviewed articles from a particular country

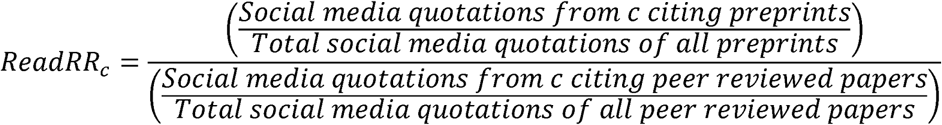

To explore how the general public engages with research output in each country through social media, we calculated the Public Engagement Relative Ratio (PERR). PERR was used to assess whether public users engage more with preprints than peer-reviewed papers from a given country.

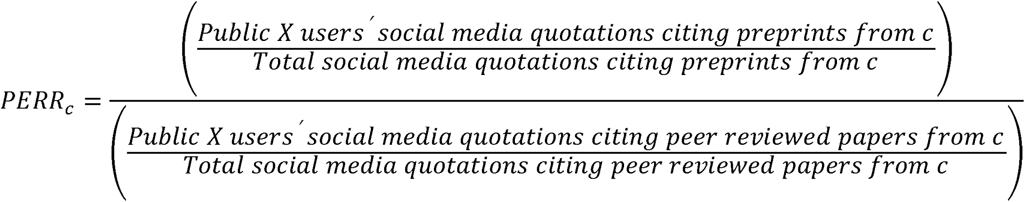

### Relative Ratio Interpretation

All relative ratios (PubRR, QuoRR, ReadRR, and PERR) are interpreted as follows:

- *Value > 1: greater engagement for preprints compared to peer-reviewed papers.*
- *Value = 1: equal engagement.*
- *Value < 1: lower engagement for preprints compared to peer-reviewed papers.*

These metrics provide a comprehensive understanding of collaboration, publication, social media dissemination and user engagement across different regions during the COVID-19 pandemic.

### Statistical Analyses

The coefficient of variation (CV) for inter-regional co-authorship counts was calculated to assess variability in collaboration among regions. Pearson’s correlation analyses were performed to examine the relationships between GDP and four relative ratio metrics. Scatter plots and R-squared values were used to visualize these relationships. The Wilcoxon signed-rank test was used to compare the median number of social media posts quoting preprints versus peer-reviewed papers within income groups. A two-sample proportion test was employed to compare the ratio of social media quotation counts to total paper counts across Scopus and medRxiv. Chi-squared tests were used to compare user demographics citing different publication types, and the Wilcoxon rank-sum test assessed citation differences between peer-reviewed papers with and without preprint versions.

## Results

### Data Collection

We collected 15,413,656 X posts referencing 277,366 papers from Scopus and 2,718,034 X posts referencing 19,584 papers from medRxiv, authored by users from 201 and 213 countries, respectively. For regional diversity analysis, we categorized countries of first authors and X users by income level and geographical region, excluding entries without identifiable country information (**Supplementary File 1**). The final categorization, which is shown in **Supplementary Figure 1**, included:

1. Country-classified Papers: We categorized papers based on the country of the first author for 250,442 Scopus papers (192 countries) and 13,952 medRxiv papers (147 countries) (**Supplementary Table 1**).
2. Country-classified X Posts: We categorized X posts based on the country of the paper’s first author (14,325,011 posts for Scopus and 1,853,738 for medRxiv) (**Supplementary Table 2 A**). Additionally, we categorized posts based on the users’ country (5,910,994 posts for Scopus and 1,010,582 for medRxiv) (**Supplementary Table 2 B**).

### Global Co-authorship Networks

Inter-regional co-authorship counts were calculated across 8 regions, resulting in 28 pairwise combinations for each data source. The average co-authorship count was 196,572.30 for Scopus and 6,833.64 for medRxiv, with a coefficient of variation (CV) of 2.70 for Scopus and 1.95 for medRxiv. This indicates a wider disparity in regional collaboration for peer-reviewed articles compared to preprints (**Table 1**). Network graphs visualizing these counts are shown in **Figure 1**, with detailed matrix and counts provided in **Supplementary Figure 2** and **Supplementary Table 3**. The co-authorship enhancement index (CEI)—calculated as the ratio of EVC for preprints to peer-reviewed publications—was highest in North America (7.66), followed by Sub-Saharan Africa (4.53), Central Asia (4.29), Latin America & Caribbean (3.90), Middle East & North Africa (2.19), East Asia & Pacific (1.76), and South Asia (1.07) (**Table 2**). **Supplementary File 2** contains inter-country co-authorship networks for peer-reviewed papers and preprints, while **Supplementary File 3** provides interactive visualizations for scatter plots, geographic heatmaps, and bar plots.

**Table 1.** Coefficient of variation in inter-regional co-authorship network.

|  | Mean | SD | CV |
| --- | --- | --- | --- |
| Preprint | 6833.6 | 13,300.3 | 1.95 |
| Peer-reviewed paper | 196,572.3 | 530,283.8 | 2.70 |

CV was calculated based on inter-regional co-authorships between all possible pairs of regions, involving 36 unique combinations (8 regions taken 2 at a time), to reflect the weight of edges across both preprints and peer-reviewed papers. Our findings revealed a skewed distribution of international co-authorship in peer-reviewed papers, heavily favoring European countries. In contrast, preprints demonstrated a more balanced co-authorship pattern, as indicated by a CV of 1.95 for preprints compared to 2.70 for peer-reviewed papers, highlighting the greater concentration of collaborations in the latter.
**Abbreviations:** CV, Coefficient of Variation; SD, Standard Deviation.

**Figure 1.**
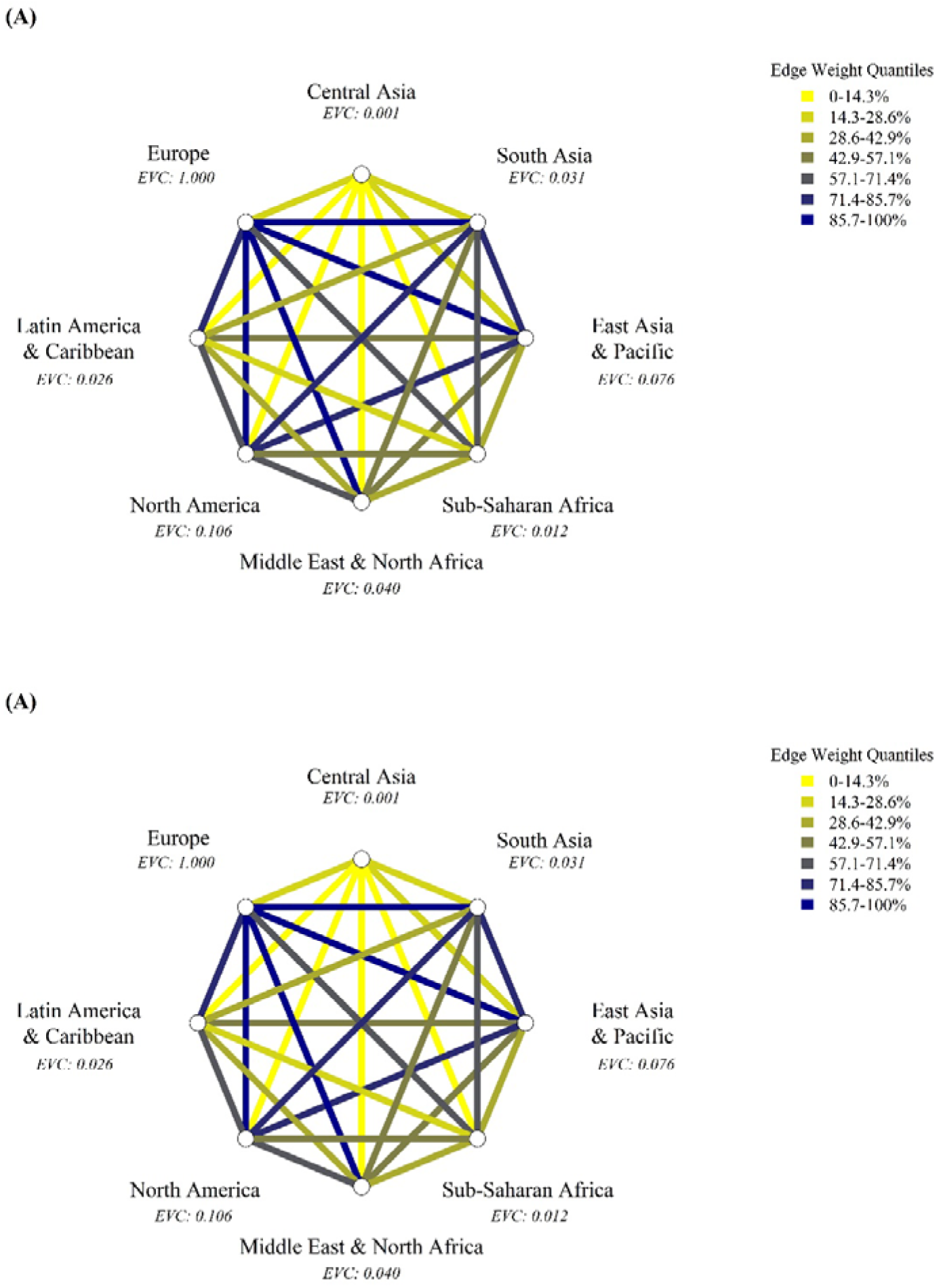
Network visualization on inter-regional co-authorship (A) Peer-reviewed Papers (B) Preprints. Figure 1, which is a network visualization on inter-regional co-authorship among regions for (a) Peer-reviewed papers and (b) Preprints, depicts regions as nodes and established connections as edges. It was made by first extracting regions for each distinct institution when papers had multiple affiliations then forming inter-regional co-authorship connections between them. The color of the edges between nodes indicates the strength of collaboration between two regions, categorized into six quantile levels. The first quantile (0-16.7) shows yellow for the lowest strength and last quantile (83.3-100) shows darkblue for the highest collaboration strength. The quantile cutoff values for each type are as follows: for peer-reviewed papers, the quantile cutoffs are 2,725.2, 33,622.9, 55,040.8, 108,458.0, and 221,283.1; for preprints, the cutoffs are 181.4, 960.8, 2,186.1, 4,900.5, and 6,888.6. The CV, EVC, and CEI was calculated from this network. **Abbreviations:** CV, Coefficient of Variation; EVC, Eigenvector Centrality; CEI, Co-authorship Enhancement Index.

**Table 2.** Eigenvector centrality in inter-regional co-authorship network.

|  | Peer-reviewed paper | Preprint | CEI |
| --- | --- | --- | --- |
| Europe | 1.000 | 1.000 | 1.00 |
| North America | 0.106 | 0.812 | 7.66 |
| East Asia & Pacific | 0.077 | 0.135 | 1.76 |
| Latin America & Caribbean | 0.026 | 0.103 | 3.90 |
| Middle East & North Africa | 0.040 | 0.088 | 2.19 |
| Sub-Saharan Africa | 0.012 | 0.055 | 4.53 |
| South Asia | 0.031 | 0.033 | 1.07 |
| Central Asia | 0.008 | 0.003 | 4.29 |

### Publication of Research

The Publication Relative Ratio (PubRR) was calculated to compare the number of papers authored by researchers from each country across Scopus and medRxiv. A significant negative correlation between PubRR and GDP was observed (Pearson correlation coefficient: −0.38, p-value < 0.001), indicating that researchers from countries with lower GDPs are more likely to publish in preprint repositories (**Figure 2 A**). Over half of the countries in North America, Latin America, Central Asia, and Sub-Saharan Africa displayed PubRR values greater than 1 (**Figure 2 B, Supplementary Figure 3**). High PubRR values were recorded in countries such as Turkmenistan (35.9), South Sudan (33.3), Mali (15.9), Cameroon (11.2) and Ukraine (9.9) (**Supplementary Table 4**). Preprints surged rapidly during COVID-19, reached half-publications faster, and had a 196 day lead over corresponding peer-reviewed publications (**Supplementary Figure 4**).

**Figure 2.**
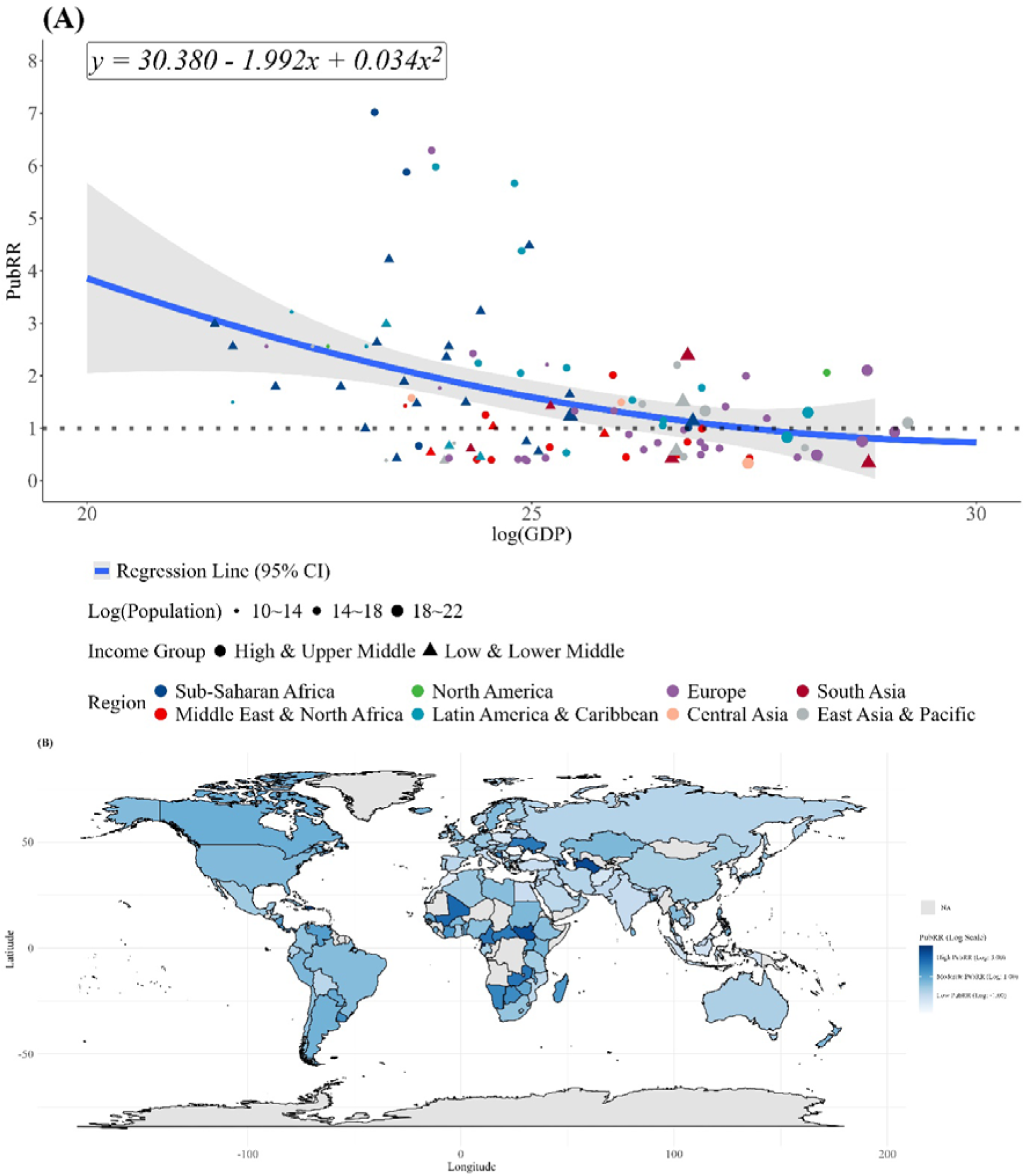
(A) Scatter plot of the country’s GDP and PubRR, (B) Geographic heatmap of PubRR. Figure 2 was made by first examining the count of papers published in each country based on the 1st author’s affiliation then calculating PubRR. Top 10% and bottom 10% outliers of PubRR values were excluded. In Figure (a), X-axis and Y-axis of the scatter plot shows log(GDP) and PubRR value of each country. It also shows Log(Population) value, Income Type (HMIC/LMIC) and Region of each country. The second-order polynomial trend line is shown to be *y* = 30.38 - 1.99*x* + 0.03*x*^2^(*R*^2^ = 0.159), and the Pearson correlation coefficient −0.28 with a *p*-value < 0.001. Figure (b) shows PubRR value on a geographical heatmap with countries with missing PubRR values colored as gray. **Abbreviations:** GDP, Gross Domestic Product; PubRR, Publication Relative Ratio; CI, Confidence Interval; Log, Common Logarithm; HMIC, High-Middle Income Countries; LMIC, Low-Middle Income Countries; NA, Not Available.

### Social Media Dissemination of Research

We analyzed the distribution of posts on platform X that referenced preprints and peer-reviewed papers by first authors from each country. Across all income groups, the median Social Media Quotation Count for medRxiv was significantly higher than for Scopus (Wilcoxon test, p-value < 0.001) (**Figure 3 A, Supplementary Table 5**). A significant negative correlation was found between Social Media Quotation Relative Ratio (QuoRR) and GDP (Pearson correlation coefficient: −0.17, p-value = 0.046), indicating that countries with lower GDPs had a higher relative share of social media quotations for preprints (**Figure 3 B**). Approximately half of the countries in North America, Sub-Saharan Africa, and North-Middle Africa had QuoRR values greater than 1, suggesting relatively higher social media attention for preprints in these regions (**Figure 3 C, Supplementary Figure 5**). High QuoRR values were observed in countries like Cameroon (80.8), South Sudan (62.9), Sudan (36.7), Turkmenistan (19.3), and Kazakhstan (13.1) (**Supplementary Table 6**).

**Figure 3.**
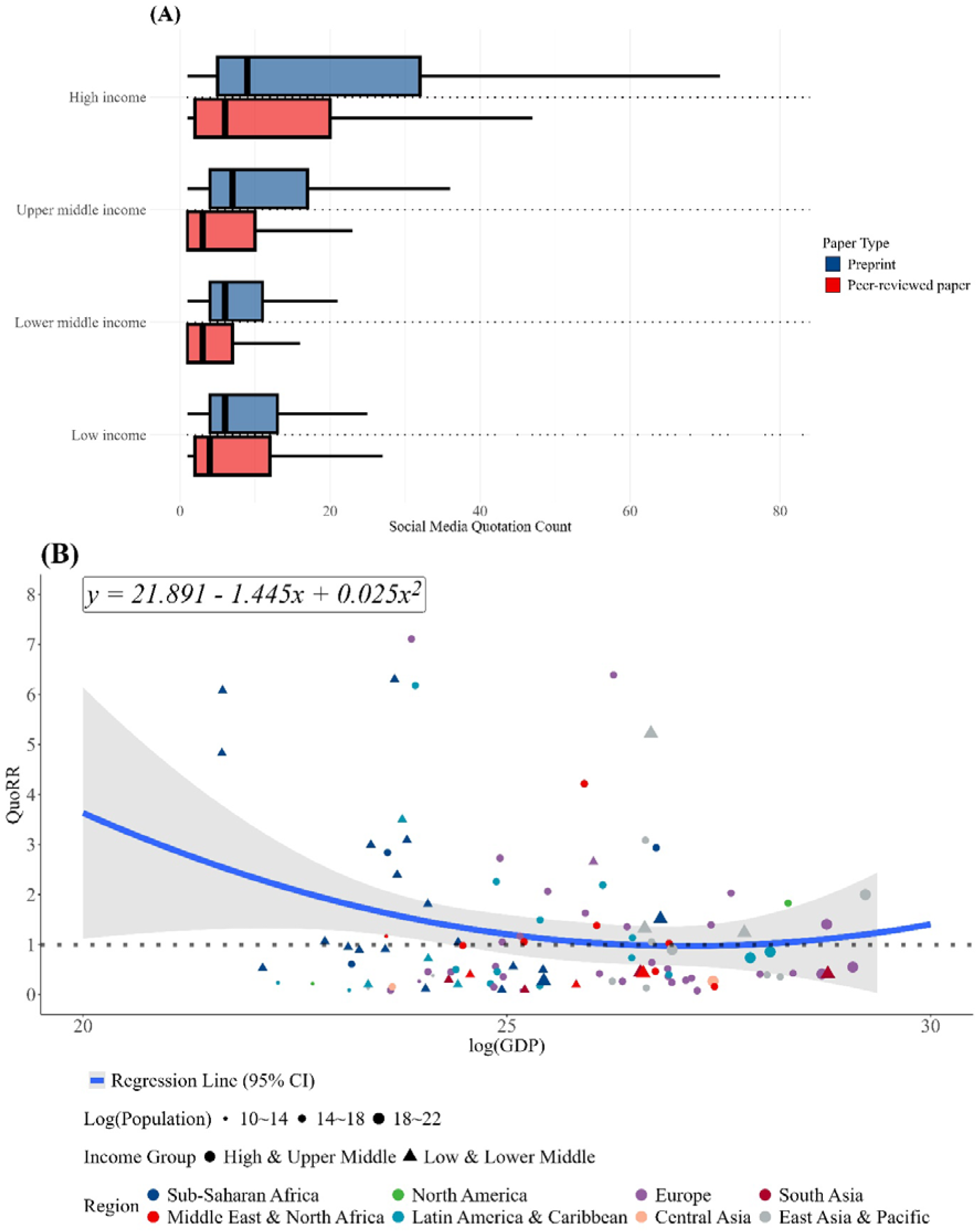
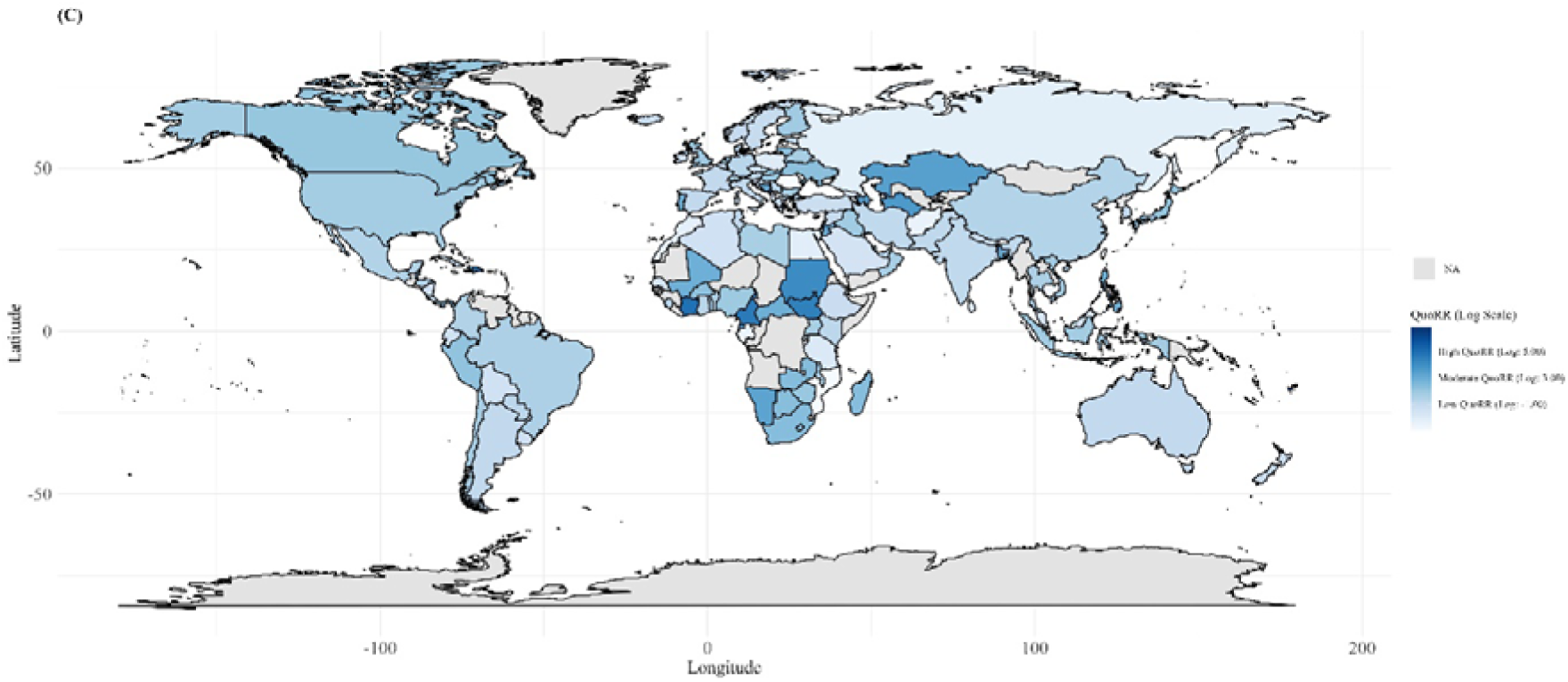
(A) Boxplot of Social Media Quotation Count by Income and Paper Type, (B) Scatter plot of the country’s GDP and QuoRR (C) Geographical Heatmap of QuoRR. Figure 3 (a) is a set of boxplots for each paper type (preprints and peer-reviewed papers) based on the social media quotation counts of each paper. They are sub-grouped by the income group of the 1st author’s country. Outliers were excluded when drawing these boxplots. Five-number summaries for each boxplot starting from the top are (1, 5, 9, 32, 72), (1, 2, 6, 20, 47), (1, 4, 7, 17, 36), (1, 1, 3, 10, 23), (1, 4, 6, 11, 21), (1, 1, 3, 7, 16), (1, 4, 6, 13, 25) and (1, 2, 4, 12, 27). Figure 3 (b), (c) was made by examining the social media quotation counts of papers published in each country then calculating QuoRR. They are drawn in the same way that Figure 2 was drawn. The second-order polynomial trend line is shown to be *y* = 21.89 - 1.45*x* + 0.03*x*^2^(*R*^2^ = 0.03), and the Pearson correlation coefficient −0.17 (p-value = 0.046). **Abbreviations:** GDP, Gross Domestic Product; QuoRR, Social Media Quotation Relative Ratio; CI, Confidence Interval; Log, Common Logarithm; HMIC, High-Middle Income Countries; LMIC, Low-Middle Income Countries; NA = Not Available.

### Social Media Readership Patterns

We compared social media engagement for preprints versus peer-reviewed papers across countries. No significant correlation was found between GDP and the Social Media Readership Relative Ratio (ReadRR) (Pearson correlation coefficient: −0.11, p-value = 0.136) (**Figure 4, Supplementary Figure 6**). Both types of papers show a similar decline in the social media quotation-to-paper ratio from high to low-income groups, with no significant difference between paper types (**Supplementary Figure 7**). The general public was the most frequent group citing both preprints and peer-reviewed papers, with a higher engagement rate for preprints (preprints: 88.26%, peer-reviewed papers: 85.75%, p-value < 0.001) (**Supplementary Figure 8**). Scientists, practitioners and science communicators showed greater engagement with peer-reviewed articles (p-value < 0.001). The Public Engagement Relative Ratio (PERR) exhibited a negative correlation with GDP (Pearson correlation coefficient: −0.20, p-value = 0.021). In North America and Central Asia, all countries had PERR values greater than 1. Similarly, in East Asia & Pacific, Sub-Saharan Africa, and Latin America & Caribbean, approximately 90% of countries had PERR greater than 1 (**Supplementary Figure 9**). High PERR values were observed in countries like Algeria (1.2), Palestine (4.4), Cameroon (1.3), Kazakhstan (1.2), Armenia (1.2), and Cambodia (1.2) **(Supplementary Table 7).**

**Figure 4.**
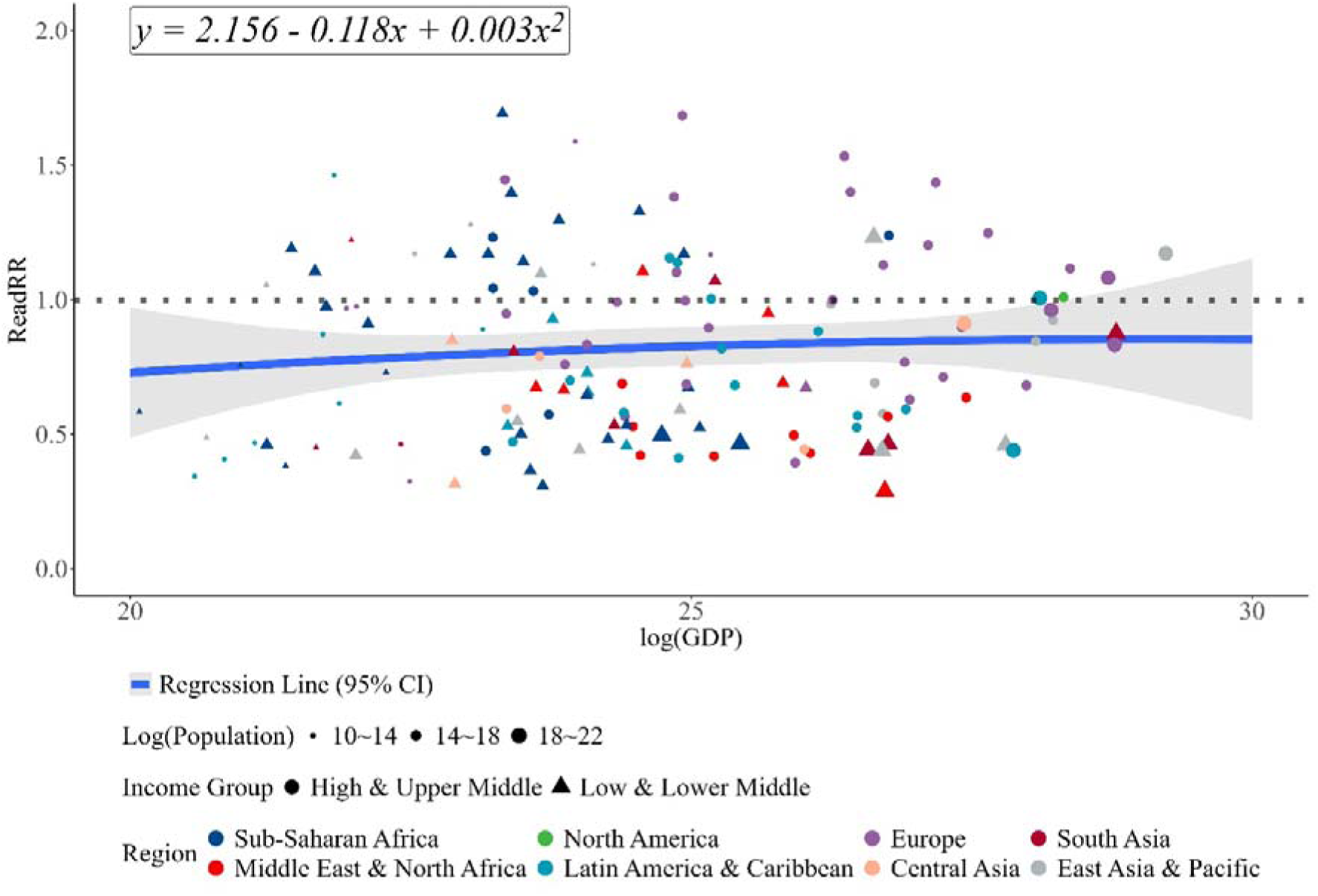
Scatter plot of the country’s GDP and ReadRR. Figure 4 was made by first examining social media quotation counts of peer-reviewed papers and preprints left by X users from each country then calculating ReadRR. It is drawn in the same way that Figure 2 was drawn. The second-order polynomial trend line is shown to be *y* = 2.16 - 0.12*x* + 0.003*x*^2^(*R*^2^ = 0.01), and the Pearson correlation coefficient −0.11 but with a non-significant p-value of 0.136. **Abbreviations**: GDP, Gross Domestic Product; ReadRR, Social Media Readership Relative Ratio; CI, Confidence Interval; Log, Common Logarithm; HMIC, High-Middle Income Countries; LMIC, Low-Middle Income Countries.

### Citation Analyses

We assessed the impact of prior preprint publication on subsequent citation counts in peer-reviewed journals. Papers with a preprint history had significantly higher citation counts (median: 10, Q1–Q3: 3–30) compared to those without (median: 5, Q1–Q3: 1–15) (p-value < 0.001) (**Figure 5**). This trend held true for papers from low- and middle-income countries (LMICs), with preprints receiving higher citation counts (median: 9, Q1-Q3: 2-25) than those without (median: 2, Q1-Q3: 0-8) (p-value < 0.001). The median citation counts by country for preprints and peer-reviewed papers are shown in **Supplementary Figure 10** and citation count statistics by region and income group for peer-reviewed papers and preprints in **Supplementary Table 8**. We also tracked changes in the income classification of first authors when preprints transitioned to peer-reviewed publications. Of these, 57% of preprints from low-income countries, 49% from lower-middle-income countries, and 22% from upper-middle-income countries were subsequently published with first authors from higher-income groups. Among LMIC preprints, 22 remained published by LMIC authors, while 26 transitioned to authorship from high-income countries (**Supplementary Table 9**).

**Figure 5.**
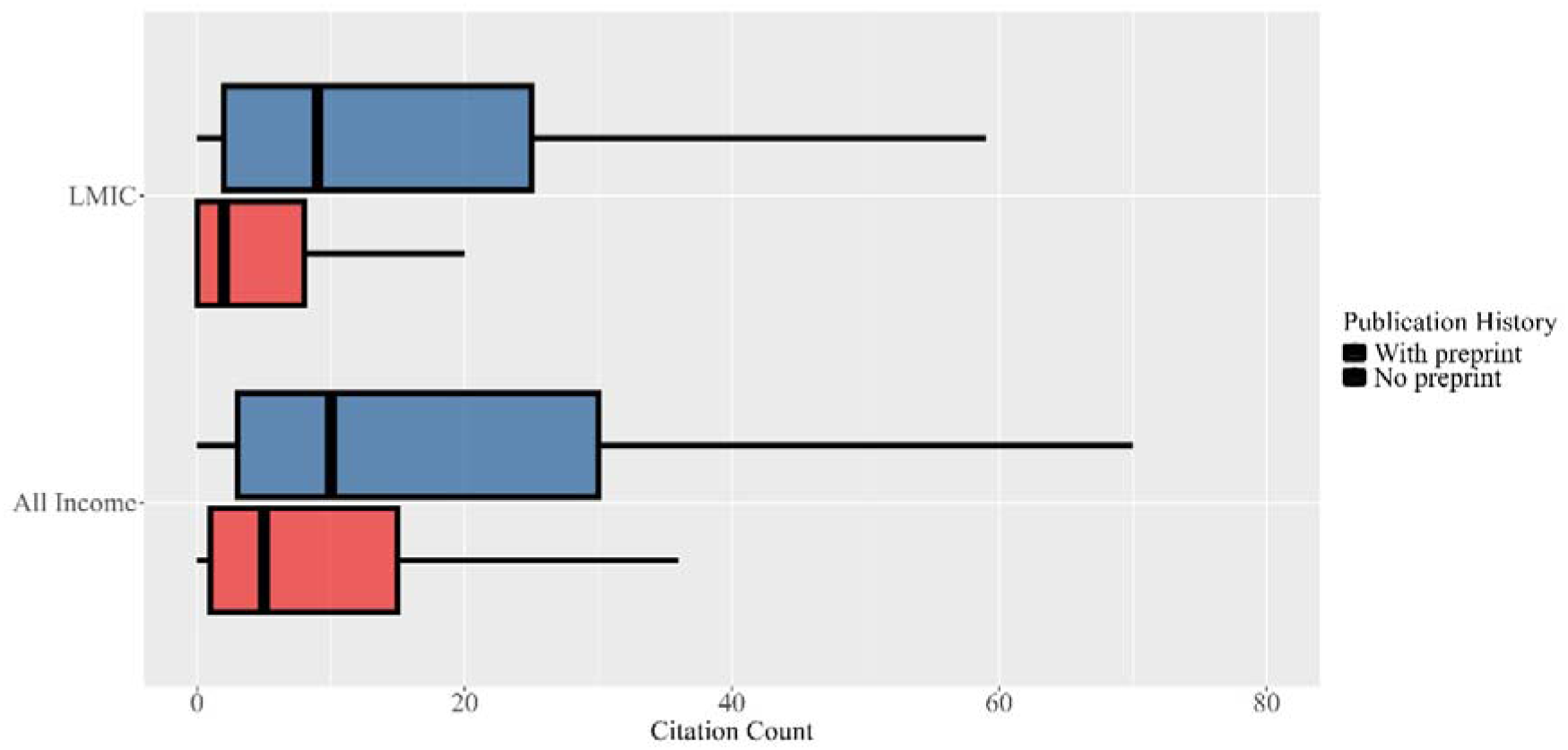
Boxplot of citation count by preprint publication history. The citation count distribution of peer-reviewed papers with and without preprint publication history is shown. Outliers were excluded. Peer-reviewed papers with preprint history exhibit higher median and quartile citation counts. The Wilcoxon rank-sum test indicates a significant increase in citations for papers with preprint history (median: 10, Q1-Q3: 3-30) versus those without (median: 5, Q1-Q3: 1-15), with a p-value < 0.001. This trend is also confirmed in LMIC countries, with median citations of 9 (Q1-Q3: 2-25) for papers with preprint history compared to 2 (Q1-Q3: 0-8) for those without, supported by a p-value < 0.001. **Abbreviations:** LMIC, Low-Middle Income Countries.

## Discussion

This study provides a comprehensive bibliometric analysis of how preprints influenced regional diversity in scientific communication during the COVID-19 pandemic. Our comparative analysis of preprints and peer-reviewed publications revealed distinct patterns across geographical regions and economic strata. Three key findings emerged: First, preprints significantly enhanced international collaboration by facilitating the inclusion of authors from regions traditionally underrepresented in peer-reviewed publications. Second, researchers from lower-GDP countries showed higher engagement with preprint platforms, suggesting these platforms serve as a more accessible publication route. Third, preprints demonstrated broader reach through social media engagement and, notably, papers that began as preprints achieved higher citation counts after peer review, particularly those from lower-GDP regions. These findings collectively indicate that preprints serve as an effective mechanism for democratizing scientific communication, enabling researchers from underrepresented regions to increase their visibility and impact in the global scientific discourse.

Our study found that peer-reviewed journals tend to publish papers from established collaborative networks, especially in affluent regions such as North America, Europe, and East Asia. This pattern likely represents several factors including long-standing research relationships,[20–22] greater access to funding,[23, 24] and the prestige of established institutions.[25–27] In contrast, our analysis revealed that preprints demonstrated a more balanced distribution of collaborations across regions, with particularly strong representation from lower-GDP countries. The higher utilization of preprint servers by researchers from lower-GDP countries compared to traditional peer-reviewed journals suggests that preprints provide a more accessible publication route for these researchers.

This accessibility proved especially crucial during the COVID-19 pandemic,[28] where rapid dissemination of research findings was essential for global public health responses.[29] By enabling researchers from lower-income regions to contribute more readily to the scientific discourse, preprints helped foster greater inclusivity in scientific communication during a critical period.

Our analysis further revealed that preprints were disseminated more extensively on social media, particularly by public users. This trend was especially pronounced among papers by researchers from lower-GDP countries, for whom social media served as a crucial tool to enhance visibility and facilitate engagement with a broader audience.[30, 31] The role of social media in disseminating research is critical, as it provides an alternative avenue for researchers who may lack the traditional means of publication and collaboration.[30, 32] Previous studies have shown that social media can significantly increase the citation impact of research, particularly for authors from institutions with fewer resources.[33, 34] This suggests that the use of social media not only facilitates the dissemination of research but also democratizes access to scientific knowledge, allowing researchers from lower GDP countries to engage more actively in global discourse.

Our analyses also indicate that peer-reviewed papers that originated as preprints tend to receive a significantly higher median citation count compared to those that did not. This trend is especially pronounced for papers where LMIC researchers are the first authors. The increased visibility and dissemination afforded by preprints may enhance subsequent citations, as preprints often serve as a preliminary platform for sharing research findings, which can lead to greater engagement and discourse within the academic community.[35–37] However, it has been observed that LMIC authorship in preprints often shifts to higher-income countries in peer-reviewed versions, likely due to disparities in funding, institutional support, and perceived credibility.[23, 38] This raises concerns about equity and representation in global research outputs.[39, 40]

Despite the advantages of preprints, concerns regarding their quality and credibility persist due to the absence of formal peer review prior to publication. Addressing these concerns requires fostering an open discussion environment with interactive feedback mechanisms.[41] Utilizing communication tools such as forums and collaborative platforms can enhance the quality assurance of preprints by allowing researchers to engage in constructive dialogue about the findings presented.[42] This approach is supported by the notion that preprints can be revised based on community input. Furthermore, the integration of communication tools such as social media can facilitate these discussions, allowing researchers to share insights and critiques in real-time. The use of interactive feedback mechanisms can help establish frameworks, allowing for a more robust evaluation of preprints. This aligns with the findings of Weissgerber et al., who noted that the overwhelming influx of preprints during the pandemic necessitated better monitoring and assess ment strategies.[43] By leveraging communication tools, researchers can create a collaborative environment that not only addresses quality concerns but also fosters a culture of transparency and accountability in scientific communication.

Although this study provides valuable insights, there are some limitations. One limitation relates to dataset attrition bias. A total of 9.7% of peer-reviewed papers and 28.8% of preprints were excluded due to incomplete affiliation details, despite the use of advanced methods to extract country information. This higher exclusion rate for preprints is largely due to the preprint system’s reliance on free-format entries for affiliations, lacking the standardized format used in peer-reviewed papers, which complicates accurate geographic extraction. Additionally, attempts to automate web-API searches were hindered by inconsistent author identification. The reliance on SCOPUS for peer-reviewed papers and MedRxiv for preprints may have excluded relevant publications from other platforms.[44] English-language queries may have missed non-English titles, particularly early COVID-19 studies from China.[45] Furthermore, the country data were based on first authors’ affiliations rather than their origin, and this study did not analyze time trends or journal impact factors, which could further refine the findings. Future research could address these aspects.

## Conclusion

The COVID-19 pandemic has clearly demonstrated how inequalities in scholarly communication can be exacerbated during a global health crisis. While it is unlikely that preprints will fully replace peer-reviewed journals in the medical field, they have proven to be an important tool in mitigating these inequalities. With specific and measurable objectives, collaborative efforts can contribute to the development of an inclusive and sustainable scientific communication environment.

## Data Availability

All data produced in the present study are available upon reasonable request to the authors

## Acknowledgement

We extend our sincere gratitude to Dr. Joshua Wallach from Emory University for his invaluable insights and constructive feedback that significantly enhanced this meta-research study.

