## Supplementary material for "Evaluating Regional Diversity in Scientific Communication: A Comparative Analysis of COVID-19 Preprints and Peer-Reviewed Publications": vis-network.min.js

```
/**
 * vis-network
 * https://visjs.github.io/vis-network/
 *
 * A dynamic, browser-based visualization library.
 *
 * @version 0.0.0-no-version
 * @date    2021-09-28T12:03:00.413Z
 *
 * @copyright (c) 2011-2017 Almende B.V, http://almende.com
 * @copyright (c) 2017-2019 visjs contributors, https://github.com/visjs
 *
 * @license
 * vis.js is dual licensed under both
 *
 *   1. The Apache 2.0 License
 *      http://www.apache.org/licenses/LICENSE-2.0
 *
 *   and
 *
 *   2. The MIT License
 *      http://opensource.org/licenses/MIT
 *
 * vis.js may be distributed under either license.
 */
!function(t,e){"object"==typeof exports&&"undefined"!=typeof module?e(exports):"function"==typeof define&&define.amd?define(["exports"],e):e((t="undefined"!=typeof globalThis?globalThis:t||self).vis=t.vis||{})}(this,(function(t){"use strict";var e="undefined"!=typeof globalThis?globalThis:"undefined"!=typeof window?window:"undefined"!=typeof global?global:"undefined"!=typeof self?self:{};function i(t){return t&&t.__esModule&&Object.prototype.hasOwnProperty.call(t,"default")?t.default:t}function n(t,e){return t(e={exports:{}},e.exports),e.exports}var o,r,s=function(t){return t&&t.Math==Math&&t},a=s("object"==typeof globalThis&&globalThis)||s("object"==typeof window&&window)||s("object"==typeof self&&self)||s("object"==typeof e&&e)||function(){return this}()||Function("return this")(),h=function(t){try{return!!t()}catch(t){return!0}},l=!h((function(){return 7!=Object.defineProperty({},1,{get:function(){return 7}})[1]})),d={}.propertyIsEnumerable,c=Object.getOwnPropertyDescriptor,u={f:c&&!d.call({1:2},1)?function(t){var e=c(this,t);return!!e&&e.enumerable}:d},f=function(t,e){return{enumerable:!(1&t),configurable:!(2&t),writable:!(4&t),value:e}},p={}.toString,v=function(t){return p.call(t).slice(8,-1)},g="".split,y=h((function(){return!Object("z").propertyIsEnumerable(0)}))?function(t){return"String"==v(t)?g.call(t,""):Object(t)}:Object,m=function(t){if(null==t)throw TypeError("Can't call method on "+t);return t},b=function(t){return y(m(t))},w=function(t){return"object"==typeof t?null!==t:"function"==typeof t},k={},_=function(t){return"function"==typeof t?t:void 0},x=function(t,e){return arguments.length<2?_(k[t])||_(a[t]):k[t]&&k[t][e]||a[t]&&a[t][e]},E=x("navigator","userAgent")||"",O=a.process,C=a.Deno,S=O&&O.versions||C&&C.version,T=S&&S.v8;T?r=(o=T.split("."))[0]<4?1:o[0]+o[1]:E&&(!(o=E.match(/Edge\/(\d+)/))||o[1]>=74)&&(o=E.match(/Chrome\/(\d+)/))&&(r=o[1]);var M=r&&+r,P=!!Object.getOwnPropertySymbols&&!h((function(){var t=Symbol();return!String(t)||!(Object(t)instanceof Symbol)||!Symbol.sham&&M&&M<41})),D=P&&!Symbol.sham&&"symbol"==typeof Symbol.iterator,I=D?function(t){return"symbol"==typeof t}:function(t){var e=x("Symbol");return"function"==typeof e&&Object(t)instanceof e},B="__core-js_shared__",z=a[B]||function(t,e){try{Object.defineProperty(a,t,{value:e,configurable:!0,writable:!0})}catch(i){a[t]=e}return e}(B,{}),N=n((function(t){(t.exports=function(t,e){return z[t]||(z[t]=void 0!==e?e:{})})("versions",[]).push({version:"3.16.1",mode:"pure",copyright:"© 2021 Denis Pushkarev (zloirock.ru)"})})),A=function(t){return Object(m(t))},F={}.hasOwnProperty,j=Object.hasOwn||function(t,e){return F.call(A(t),e)},R=0,L=Math.random(),H=function(t){return"Symbol("+String(void 0===t?"":t)+")_"+(++R+L).toString(36)},W=N("wks"),q=a.Symbol,V=D?q:q&&q.withoutSetter||H,U=function(t){return j(W,t)&&(P||"string"==typeof W[t])||(P&&j(q,t)?W[t]=q[t]:W[t]=V("Symbol."+t)),W[t]},Y=U("toPrimitive"),X=function(t,e){if(!w(t)||I(t))return t;var i,n=t[Y];if(void 0!==n){if(void 0===e&&(e="default"),i=n.call(t,e),!w(i)||I(i))return i;throw TypeError("Can't convert object to primitive value")}return void 0===e&&(e="number"),function(t,e){var i,n;if("string"===e&&"function"==typeof(i=t.toString)&&!w(n=i.call(t)))return n;if("function"==typeof(i=t.valueOf)&&!w(n=i.call(t)))return n;if("string"!==e&&"function"==typeof(i=t.toString)&&!w(n=i.call(t)))return n;throw TypeError("Can't convert object to primitive value")}(t,e)},G=function(t){var e=X(t,"string");return I(e)?e:String(e)},K=a.document,$=w(K)&&w(K.createElement),Z=function(t){return $?K.createElement(t):{}},Q=!l&&!h((function(){return 7!=Object.defineProperty(Z("div"),"a",{get:function(){return 7}}).a})),J=Object.getOwnPropertyDescriptor,tt={f:l?J:function(t,e){if(t=b(t),e=G(e),Q)try{return J(t,e)}catch(t){}if(j(t,e))return f(!u.f.call(t,e),t[e])}},et=/#|\.prototype\./,it=function(t,e){var i=ot[nt(t)];return i==st||i!=rt&&("function"==typeof e?h(e):!!e)},nt=it.normalize=function(t){return String(t).replace(et,".").toLowerCase()},ot=it.data={},rt=it.NATIVE="N",st=it.POLYFILL="P",at=it,ht=function(t){if("function"!=typeof t)throw TypeError(String(t)+" is not a function");return t},lt=function(t,e,i){if(ht(t),void 0===e)return t;switch(i){case 0:return function(){return t.call(e)};case 1:return function(i){return t.call(e,i)};case 2:return function(i,n){return t.call(e,i,n)};case 3:return function(i,n,o){return t.call(e,i,n,o)}}return function(){return t.apply(e,arguments)}},dt=function(t){if(!w(t))throw TypeError(String(t)+" is not an object");return t},ct=Object.defineProperty,ut={f:l?ct:function(t,e,i){if(dt(t),e=G(e),dt(i),Q)try{return ct(t,e,i)}catch(t){}if("get"in i||"set"in i)throw TypeError("Accessors not supported");return"value"in i&&(t[e]=i.value),t}},ft=l?function(t,e,i){return ut.f(t,e,f(1,i))}:function(t,e,i){return t[e]=i,t},pt=tt.f,vt=function(t){var e=function(e,i,n){if(this instanceof t){switch(arguments.length){case 0:return new t;case 1:return new t(e);case 2:return new t(e,i)}return new t(e,i,n)}return t.apply(this,arguments)};return e.prototype=t.prototype,e},gt=function(t,e){var i,n,o,r,s,h,l,d,c=t.target,u=t.global,f=t.stat,p=t.proto,v=u?a:f?a[c]:(a[c]||{}).prototype,g=u?k:k[c]||(k[c]={}),y=g.prototype;for(o in e)i=!at(u?o:c+(f?".":"#")+o,t.forced)&&v&&j(v,o),s=g[o],i&&(h=t.noTargetGet?(d=pt(v,o))&&d.value:v[o]),r=i&&h?h:e[o],i&&typeof s==typeof r||(l=t.bind&&i?lt(r,a):t.wrap&&i?vt(r):p&&"function"==typeof r?lt(Function.call,r):r,(t.sham||r&&r.sham||s&&s.sham)&&ft(l,"sham",!0),g[o]=l,p&&(j(k,n=c+"Prototype")||ft(k,n,{}),k[n][o]=r,t.real&&y&&!y[o]&&ft(y,o,r)))},yt=Math.ceil,mt=Math.floor,bt=function(t){return isNaN(t=+t)?0:(t>0?mt:yt)(t)},wt=Math.min,kt=function(t){return t>0?wt(bt(t),9007199254740991):0},_t=Math.max,xt=Math.min,Et=function(t,e){var i=bt(t);return i<0?_t(i+e,0):xt(i,e)},Ot=function(t){return function(e,i,n){var o,r=b(e),s=kt(r.length),a=Et(n,s);if(t&&i!=i){for(;s>a;)if((o=r[a++])!=o)return!0}else for(;s>a;a++)if((t||a in r)&&r[a]===i)return t||a||0;return!t&&-1}},Ct={includes:Ot(!0),indexOf:Ot(!1)},St={},Tt=Ct.indexOf,Mt=function(t,e){var i,n=b(t),o=0,r=[];for(i in n)!j(St,i)&&j(n,i)&&r.push(i);for(;e.length>o;)j(n,i=e[o++])&&(~Tt(r,i)||r.push(i));return r},Pt=["constructor","hasOwnProperty","isPrototypeOf","propertyIsEnumerable","toLocaleString","toString","valueOf"],Dt=Object.keys||function(t){return Mt(t,Pt)},It={f:Object.getOwnPropertySymbols},Bt=Object.assign,zt=Object.defineProperty,Nt=!Bt||h((function(){if(l&&1!==Bt({b:1},Bt(zt({},"a",{enumerable:!0,get:function(){zt(this,"b",{value:3,enumerable:!1})}}),{b:2})).b)return!0;var t={},e={},i=Symbol(),n="abcdefghijklmnopqrst";return t[i]=7,n.split("").forEach((function(t){e[t]=t})),7!=Bt({},t)[i]||Dt(Bt({},e)).join("")!=n}))?function(t,e){for(var i=A(t),n=arguments.length,o=1,r=It.f,s=u.f;n>o;)for(var a,h=y(arguments[o++]),d=r?Dt(h).concat(r(h)):Dt(h),c=d.length,f=0;c>f;)a=d[f++],l&&!s.call(h,a)||(i[a]=h[a]);return i}:Bt;gt({target:"Object",stat:!0,forced:Object.assign!==Nt},{assign:Nt});var At=k.Object.assign,Ft=[].slice,jt={},Rt=function(t,e,i){if(!(e in jt)){for(var n=[],o=0;o<e;o++)n[o]="a["+o+"]";jt[e]=Function("C,a","return new C("+n.join(",")+")")}return jt[e](t,i)},Lt=Function.bind||function(t){var e=ht(this),i=Ft.call(arguments,1),n=function(){var o=i.concat(Ft.call(arguments));return this instanceof n?Rt(e,o.length,o):e.apply(t,o)};return w(e.prototype)&&(n.prototype=e.prototype),n};gt({target:"Function",proto:!0},{bind:Lt});var Ht=function(t){return k[t+"Prototype"]},Wt=Ht("Function").bind,qt=Function.prototype,Vt=function(t){var e=t.bind;return t===qt||t instanceof Function&&e===qt.bind?Wt:e};function Ut(t,e,i,n){t.beginPath(),t.arc(e,i,n,0,2*Math.PI,!1),t.closePath()}function Yt(t,e,i,n,o,r){var s=Math.PI/180;n-2*r<0&&(r=n/2),o-2*r<0&&(r=o/2),t.beginPath(),t.moveTo(e+r,i),t.lineTo(e+n-r,i),t.arc(e+n-r,i+r,r,270*s,360*s,!1),t.lineTo(e+n,i+o-r),t.arc(e+n-r,i+o-r,r,0,90*s,!1),t.lineTo(e+r,i+o),t.arc(e+r,i+o-r,r,90*s,180*s,!1),t.lineTo(e,i+r),t.arc(e+r,i+r,r,180*s,270*s,!1),t.closePath()}function Xt(t,e,i,n,o){var r=.5522848,s=n/2*r,a=o/2*r,h=e+n,l=i+o,d=e+n/2,c=i+o/2;t.beginPath(),t.moveTo(e,c),t.bezierCurveTo(e,c-a,d-s,i,d,i),t.bezierCurveTo(d+s,i,h,c-a,h,c),t.bezierCurveTo(h,c+a,d+s,l,d,l),t.bezierCurveTo(d-s,l,e,c+a,e,c),t.closePath()}function Gt(t,e,i,n,o){var r=o*(1/3),s=.5522848,a=n/2*s,h=r/2*s,l=e+n,d=i+r,c=e+n/2,u=i+r/2,f=i+(o-r/2),p=i+o;t.beginPath(),t.moveTo(l,u),t.bezierCurveTo(l,u+h,c+a,d,c,d),t.bezierCurveTo(c-a,d,e,u+h,e,u),t.bezierCurveTo(e,u-h,c-a,i,c,i),t.bezierCurveTo(c+a,i,l,u-h,l,u),t.lineTo(l,f),t.bezierCurveTo(l,f+h,c+a,p,c,p),t.bezierCurveTo(c-a,p,e,f+h,e,f),t.lineTo(e,u)}function Kt(t,e,i,n,o,r){t.beginPath(),t.moveTo(e,i);for(var s=r.length,a=n-e,h=o-i,l=h/a,d=Math.sqrt(a*a+h*h),c=0,u=!0,f=0,p=+r[0];d>=.1;)(p=+r[c++%s])>d&&(p=d),f=Math.sqrt(p*p/(1+l*l)),e+=f=a<0?-f:f,i+=l*f,!0===u?t.lineTo(e,i):t.moveTo(e,i),d-=p,u=!u}var $t={circle:Ut,dashedLine:Kt,database:Gt,diamond:function(t,e,i,n){t.beginPath(),t.lineTo(e,i+n),t.lineTo(e+n,i),t.lineTo(e,i-n),t.lineTo(e-n,i),t.closePath()},ellipse:Xt,ellipse_vis:Xt,hexagon:function(t,e,i,n){t.beginPath();var o=2*Math.PI/6;t.moveTo(e+n,i);for(var r=1;r<6;r++)t.lineTo(e+n*Math.cos(o*r),i+n*Math.sin(o*r));t.closePath()},roundRect:Yt,square:function(t,e,i,n){t.beginPath(),t.rect(e-n,i-n,2*n,2*n),t.closePath()},star:function(t,e,i,n){t.beginPath(),i+=.1*(n*=.82);for(var o=0;o<10;o++){var r=o%2==0?1.3*n:.5*n;t.lineTo(e+r*Math.sin(2*o*Math.PI/10),i-r*Math.cos(2*o*Math.PI/10))}t.closePath()},triangle:function(t,e,i,n){t.beginPath(),i+=.275*(n*=1.15);var o=2*n,r=o/2,s=Math.sqrt(3)/6*o,a=Math.sqrt(o*o-r*r);t.moveTo(e,i-(a-s)),t.lineTo(e+r,i+s),t.lineTo(e-r,i+s),t.lineTo(e,i-(a-s)),t.closePath()},triangleDown:function(t,e,i,n){t.beginPath(),i-=.275*(n*=1.15);var o=2*n,r=o/2,s=Math.sqrt(3)/6*o,a=Math.sqrt(o*o-r*r);t.moveTo(e,i+(a-s)),t.lineTo(e+r,i-s),t.lineTo(e-r,i-s),t.lineTo(e,i+(a-s)),t.closePath()}};var Zt=n((function(t){function e(t){if(t)return function(t){for(var i in e.prototype)t[i]=e.prototype[i];return t}(t)}t.exports=e,e.prototype.on=e.prototype.addEventListener=function(t,e){return this._callbacks=this._callbacks||{},(this._callbacks["$"+t]=this._callbacks["$"+t]||[]).push(e),this},e.prototype.once=function(t,e){function i(){this.off(t,i),e.apply(this,arguments)}return i.fn=e,this.on(t,i),this},e.prototype.off=e.prototype.removeListener=e.prototype.removeAllListeners=e.prototype.removeEventListener=function(t,e){if(this._callbacks=this._callbacks||{},0==arguments.length)return this._callbacks={},this;var i,n=this._callbacks["$"+t];if(!n)return this;if(1==arguments.length)return delete this._callbacks["$"+t],this;for(var o=0;o<n.length;o++)if((i=n[o])===e||i.fn===e){n.splice(o,1);break}return 0===n.length&&delete this._callbacks["$"+t],this},e.prototype.emit=function(t){this._callbacks=this._callbacks||{};for(var e=new Array(arguments.length-1),i=this._callbacks["$"+t],n=1;n<arguments.length;n++)e[n-1]=arguments[n];if(i){n=0;for(var o=(i=i.slice(0)).length;n<o;++n)i[n].apply(this,e)}return this},e.prototype.listeners=function(t){return this._callbacks=this._callbacks||{},this._callbacks["$"+t]||[]},e.prototype.hasListeners=function(t){return!!this.listeners(t).length}})),Qt=function(t){if(I(t))throw TypeError("Cannot convert a Symbol value to a string");return String(t)},Jt=function(t){return function(e,i){var n,o,r=Qt(m(e)),s=bt(i),a=r.length;return s<0||s>=a?t?"":void 0:(n=r.charCodeAt(s))<55296||n>56319||s+1===a||(o=r.charCodeAt(s+1))<56320||o>57343?t?r.charAt(s):n:t?r.slice(s,s+2):o-56320+(n-55296<<10)+65536}},te={codeAt:Jt(!1),charAt:Jt(!0)},ee=Function.toString;"function"!=typeof z.inspectSource&&(z.inspectSource=function(t){return ee.call(t)});var ie,ne,oe,re=z.inspectSource,se=a.WeakMap,ae="function"==typeof se&&/native code/.test(re(se)),he=N("keys"),le=function(t){return he[t]||(he[t]=H(t))},de="Object already initialized",ce=a.WeakMap;if(ae||z.state){var ue=z.state||(z.state=new ce),fe=ue.get,pe=ue.has,ve=ue.set;ie=function(t,e){if(pe.call(ue,t))throw new TypeError(de);return e.facade=t,ve.call(ue,t,e),e},ne=function(t){return fe.call(ue,t)||{}},oe=function(t){return pe.call(ue,t)}}else{var ge=le("state");St[ge]=!0,ie=function(t,e){if(j(t,ge))throw new TypeError(de);return e.facade=t,ft(t,ge,e),e},ne=function(t){return j(t,ge)?t[ge]:{}},oe=function(t){return j(t,ge)}}var ye,me,be,we={set:ie,get:ne,has:oe,enforce:function(t){return oe(t)?ne(t):ie(t,{})},getterFor:function(t){return function(e){var i;if(!w(e)||(i=ne(e)).type!==t)throw TypeError("Incompatible receiver, "+t+" required");return i}}},ke=!h((function(){function t(){}return t.prototype.constructor=null,Object.getPrototypeOf(new t)!==t.prototype})),_e=le("IE_PROTO"),xe=Object.prototype,Ee=ke?Object.getPrototypeOf:function(t){return t=A(t),j(t,_e)?t[_e]:"function"==typeof t.constructor&&t instanceof t.constructor?t.constructor.prototype:t instanceof Object?xe:null},Oe=U("iterator"),Ce=!1;[].keys&&("next"in(be=[].keys())?(me=Ee(Ee(be)))!==Object.prototype&&(ye=me):Ce=!0);var Se=null==ye||h((function(){var t={};return ye[Oe].call(t)!==t}));Se&&(ye={}),Se&&!j(ye,Oe)&&ft(ye,Oe,(function(){return this}));var Te,Me={IteratorPrototype:ye,BUGGY_SAFARI_ITERATORS:Ce},Pe=l?Object.defineProperties:function(t,e){dt(t);for(var i,n=Dt(e),o=n.length,r=0;o>r;)ut.f(t,i=n[r++],e[i]);return t},De=x("document","documentElement"),Ie=le("IE_PROTO"),Be=function(){},ze=function(t){return"<script>"+t+"</"+"script>"},Ne=function(t){t.write(ze("")),t.close();var e=t.parentWindow.Object;return t=null,e},Ae=function(){try{Te=new ActiveXObject("htmlfile")}catch(t){}Ae=document.domain&&Te?Ne(Te):function(){var t,e=Z("iframe");if(e.style)return e.style.display="none",De.appendChild(e),e.src=String("javascript:"),(t=e.contentWindow.document).open(),t.write(ze("document.F=Object")),t.close(),t.F}()||Ne(Te);for(var t=Pt.length;t--;)delete Ae.prototype[Pt[t]];return Ae()};St[Ie]=!0;var Fe=Object.create||function(t,e){var i;return null!==t?(Be.prototype=dt(t),i=new Be,Be.prototype=null,i[Ie]=t):i=Ae(),void 0===e?i:Pe(i,e)},je={};je[U("toStringTag")]="z";var Re="[object z]"===String(je),Le=U("toStringTag"),He="Arguments"==v(function(){return arguments}()),We=Re?v:function(t){var e,i,n;return void 0===t?"Undefined":null===t?"Null":"string"==typeof(i=function(t,e){try{return t[e]}catch(t){}}(e=Object(t),Le))?i:He?v(e):"Object"==(n=v(e))&&"function"==typeof e.callee?"Arguments":n},qe=Re?{}.toString:function(){return"[object "+We(this)+"]"},Ve=ut.f,Ue=U("toStringTag"),Ye=function(t,e,i,n){if(t){var o=i?t:t.prototype;j(o,Ue)||Ve(o,Ue,{configurable:!0,value:e}),n&&!Re&&ft(o,"toString",qe)}},Xe={},Ge=Me.IteratorPrototype,Ke=function(){return this},$e=Object.setPrototypeOf||("__proto__"in{}?function(){var t,e=!1,i={};try{(t=Object.getOwnPropertyDescriptor(Object.prototype,"__proto__").set).call(i,[]),e=i instanceof Array}catch(t){}return function(i,n){return dt(i),function(t){if(!w(t)&&null!==t)throw TypeError("Can't set "+String(t)+" as a prototype")}(n),e?t.call(i,n):i.__proto__=n,i}}():void 0),Ze=function(t,e,i,n){n&&n.enumerable?t[e]=i:ft(t,e,i)},Qe=Me.IteratorPrototype,Je=Me.BUGGY_SAFARI_ITERATORS,ti=U("iterator"),ei="keys",ii="values",ni="entries",oi=function(){return this},ri=function(t,e,i,n,o,r,s){!function(t,e,i){var n=e+" Iterator";t.prototype=Fe(Ge,{next:f(1,i)}),Ye(t,n,!1,!0),Xe[n]=Ke}(i,e,n);var a,h,l,d=function(t){if(t===o&&g)return g;if(!Je&&t in p)return p[t];switch(t){case ei:case ii:case ni:return function(){return new i(this,t)}}return function(){return new i(this)}},c=e+" Iterator",u=!1,p=t.prototype,v=p[ti]||p["@@iterator"]||o&&p[o],g=!Je&&v||d(o),y="Array"==e&&p.entries||v;if(y&&(a=Ee(y.call(new t)),Qe!==Object.prototype&&a.next&&(Ye(a,c,!0,!0),Xe[c]=oi)),o==ii&&v&&v.name!==ii&&(u=!0,g=function(){return v.call(this)}),s&&p[ti]!==g&&ft(p,ti,g),Xe[e]=g,o)if(h={values:d(ii),keys:r?g:d(ei),entries:d(ni)},s)for(l in h)(Je||u||!(l in p))&&Ze(p,l,h[l]);else gt({target:e,proto:!0,forced:Je||u},h);return h},si=te.charAt,ai="String Iterator",hi=we.set,li=we.getterFor(ai);ri(String,"String",(function(t){hi(this,{type:ai,string:Qt(t),index:0})}),(function(){var t,e=li(this),i=e.string,n=e.index;return n>=i.length?{value:void 0,done:!0}:(t=si(i,n),e.index+=t.length,{value:t,done:!1})}));var di=function(t){var e=t.return;if(void 0!==e)return dt(e.call(t)).value},ci=function(t,e,i,n){try{return n?e(dt(i)[0],i[1]):e(i)}catch(e){throw di(t),e}},ui=U("iterator"),fi=Array.prototype,pi=function(t){return void 0!==t&&(Xe.Array===t||fi[ui]===t)},vi=function(t,e,i){var n=G(e);n in t?ut.f(t,n,f(0,i)):t[n]=i},gi=U("iterator"),yi=function(t){if(null!=t)return t[gi]||t["@@iterator"]||Xe[We(t)]},mi=U("iterator"),bi=!1;try{var wi=0,ki={next:function(){return{done:!!wi++}},return:function(){bi=!0}};ki[mi]=function(){return this},Array.from(ki,(function(){throw 2}))}catch(t){}var _i=!function(t,e){if(!e&&!bi)return!1;var i=!1;try{var n={};n[mi]=function(){return{next:function(){return{done:i=!0}}}},t(n)}catch(t){}return i}((function(t){Array.from(t)}));gt({target:"Array",stat:!0,forced:_i},{from:function(t){var e,i,n,o,r,s,a=A(t),h="function"==typeof this?this:Array,l=arguments.length,d=l>1?arguments[1]:void 0,c=void 0!==d,u=yi(a),f=0;if(c&&(d=lt(d,l>2?arguments[2]:void 0,2)),null==u||h==Array&&pi(u))for(i=new h(e=kt(a.length));e>f;f++)s=c?d(a[f],f):a[f],vi(i,f,s);else for(r=(o=u.call(a)).next,i=new h;!(n=r.call(o)).done;f++)s=c?ci(o,d,[n.value,f],!0):n.value,vi(i,f,s);return i.length=f,i}});var xi=k.Array.from,Ei=xi,Oi="Array Iterator",Ci=we.set,Si=we.getterFor(Oi);ri(Array,"Array",(function(t,e){Ci(this,{type:Oi,target:b(t),index:0,kind:e})}),(function(){var t=Si(this),e=t.target,i=t.kind,n=t.index++;return!e||n>=e.length?(t.target=void 0,{value:void 0,done:!0}):"keys"==i?{value:n,done:!1}:"values"==i?{value:e[n],done:!1}:{value:[n,e[n]],done:!1}}),"values"),Xe.Arguments=Xe.Array;var Ti=yi,Mi=U("toStringTag");for(var Pi in{CSSRuleList:0,CSSStyleDeclaration:0,CSSValueList:0,ClientRectList:0,DOMRectList:0,DOMStringList:0,DOMTokenList:1,DataTransferItemList:0,FileList:0,HTMLAllCollection:0,HTMLCollection:0,HTMLFormElement:0,HTMLSelectElement:0,MediaList:0,MimeTypeArray:0,NamedNodeMap:0,NodeList:1,PaintRequestList:0,Plugin:0,PluginArray:0,SVGLengthList:0,SVGNumberList:0,SVGPathSegList:0,SVGPointList:0,SVGStringList:0,SVGTransformList:0,SourceBufferList:0,StyleSheetList:0,TextTrackCueList:0,TextTrackList:0,TouchList:0}){var Di=a[Pi],Ii=Di&&Di.prototype;Ii&&We(Ii)!==Mi&&ft(Ii,Mi,Pi),Xe[Pi]=Xe.Array}var Bi=Ti,zi=Array.isArray||function(t){return"Array"==v(t)},Ni=Pt.concat("length","prototype"),Ai={f:Object.getOwnPropertyNames||function(t){return Mt(t,Ni)}},Fi=Ai.f,ji={}.toString,Ri="object"==typeof window&&window&&Object.getOwnPropertyNames?Object.getOwnPropertyNames(window):[],Li={f:function(t){return Ri&&"[object Window]"==ji.call(t)?function(t){try{return Fi(t)}catch(t){return Ri.slice()}}(t):Fi(b(t))}},Hi={f:U},Wi=ut.f,qi=function(t){var e=k.Symbol||(k.Symbol={});j(e,t)||Wi(e,t,{value:Hi.f(t)})},Vi=U("species"),Ui=function(t,e){return new(function(t){var e;return zi(t)&&("function"!=typeof(e=t.constructor)||e!==Array&&!zi(e.prototype)?w(e)&&null===(e=e[Vi])&&(e=void 0):e=void 0),void 0===e?Array:e}(t))(0===e?0:e)},Yi=[].push,Xi=function(t){var e=1==t,i=2==t,n=3==t,o=4==t,r=6==t,s=7==t,a=5==t||r;return function(h,l,d,c){for(var u,f,p=A(h),v=y(p),g=lt(l,d,3),m=kt(v.length),b=0,w=c||Ui,k=e?w(h,m):i||s?w(h,0):void 0;m>b;b++)if((a||b in v)&&(f=g(u=v[b],b,p),t))if(e)k[b]=f;else if(f)switch(t){case 3:return!0;case 5:return u;case 6:return b;case 2:Yi.call(k,u)}else switch(t){case 4:return!1;case 7:Yi.call(k,u)}return r?-1:n||o?o:k}},Gi={forEach:Xi(0),map:Xi(1),filter:Xi(2),some:Xi(3),every:Xi(4),find:Xi(5),findIndex:Xi(6),filterReject:Xi(7)},Ki=Gi.forEach,$i=le("hidden"),Zi="Symbol",Qi=U("toPrimitive"),Ji=we.set,tn=we.getterFor(Zi),en=Object.prototype,nn=a.Symbol,on=x("JSON","stringify"),rn=tt.f,sn=ut.f,an=Li.f,hn=u.f,ln=N("symbols"),dn=N("op-symbols"),cn=N("string-to-symbol-registry"),un=N("symbol-to-string-registry"),fn=N("wks"),pn=a.QObject,vn=!pn||!pn.prototype||!pn.prototype.findChild,gn=l&&h((function(){return 7!=Fe(sn({},"a",{get:function(){return sn(this,"a",{value:7}).a}})).a}))?function(t,e,i){var n=rn(en,e);n&&delete en[e],sn(t,e,i),n&&t!==en&&sn(en,e,n)}:sn,yn=function(t,e){var i=ln[t]=Fe(nn.prototype);return Ji(i,{type:Zi,tag:t,description:e}),l||(i.description=e),i},mn=function(t,e,i){t===en&&mn(dn,e,i),dt(t);var n=G(e);return dt(i),j(ln,n)?(i.enumerable?(j(t,$i)&&t[$i][n]&&(t[$i][n]=!1),i=Fe(i,{enumerable:f(0,!1)})):(j(t,$i)||sn(t,$i,f(1,{})),t[$i][n]=!0),gn(t,n,i)):sn(t,n,i)},bn=function(t,e){dt(t);var i=b(e),n=Dt(i).concat(xn(i));return Ki(n,(function(e){l&&!wn.call(i,e)||mn(t,e,i[e])})),t},wn=function(t){var e=G(t),i=hn.call(this,e);return!(this===en&&j(ln,e)&&!j(dn,e))&&(!(i||!j(this,e)||!j(ln,e)||j(this,$i)&&this[$i][e])||i)},kn=function(t,e){var i=b(t),n=G(e);if(i!==en||!j(ln,n)||j(dn,n)){var o=rn(i,n);return!o||!j(ln,n)||j(i,$i)&&i[$i][n]||(o.enumerable=!0),o}},_n=function(t){var e=an(b(t)),i=[];return Ki(e,(function(t){j(ln,t)||j(St,t)||i.push(t)})),i},xn=function(t){var e=t===en,i=an(e?dn:b(t)),n=[];return Ki(i,(function(t){!j(ln,t)||e&&!j(en,t)||n.push(ln[t])})),n};if(P||(Ze((nn=function(){if(this instanceof nn)throw TypeError("Symbol is not a constructor");var t=arguments.length&&void 0!==arguments[0]?Qt(arguments[0]):void 0,e=H(t),i=function(t){this===en&&i.call(dn,t),j(this,$i)&&j(this[$i],e)&&(this[$i][e]=!1),gn(this,e,f(1,t))};return l&&vn&&gn(en,e,{configurable:!0,set:i}),yn(e,t)}).prototype,"toString",(function(){return tn(this).tag})),Ze(nn,"withoutSetter",(function(t){return yn(H(t),t)})),u.f=wn,ut.f=mn,tt.f=kn,Ai.f=Li.f=_n,It.f=xn,Hi.f=function(t){return yn(U(t),t)},l&&sn(nn.prototype,"description",{configurable:!0,get:function(){return tn(this).description}})),gt({global:!0,wrap:!0,forced:!P,sham:!P},{Symbol:nn}),Ki(Dt(fn),(function(t){qi(t)})),gt({target:Zi,stat:!0,forced:!P},{for:function(t){var e=Qt(t);if(j(cn,e))return cn[e];var i=nn(e);return cn[e]=i,un[i]=e,i},keyFor:function(t){if(!I(t))throw TypeError(t+" is not a symbol");if(j(un,t))return un[t]},useSetter:function(){vn=!0},useSimple:function(){vn=!1}}),gt({target:"Object",stat:!0,forced:!P,sham:!l},{create:function(t,e){return void 0===e?Fe(t):bn(Fe(t),e)},defineProperty:mn,defineProperties:bn,getOwnPropertyDescriptor:kn}),gt({target:"Object",stat:!0,forced:!P},{getOwnPropertyNames:_n,getOwnPropertySymbols:xn}),gt({target:"Object",stat:!0,forced:h((function(){It.f(1)}))},{getOwnPropertySymbols:function(t){return It.f(A(t))}}),on){var En=!P||h((function(){var t=nn();return"[null]"!=on([t])||"{}"!=on({a:t})||"{}"!=on(Object(t))}));gt({target:"JSON",stat:!0,forced:En},{stringify:function(t,e,i){for(var n,o=[t],r=1;arguments.length>r;)o.push(arguments[r++]);if(n=e,(w(e)||void 0!==t)&&!I(t))return zi(e)||(e=function(t,e){if("function"==typeof n&&(e=n.call(this,t,e)),!I(e))return e}),o[1]=e,on.apply(null,o)}})}nn.prototype[Qi]||ft(nn.prototype,Qi,nn.prototype.valueOf),Ye(nn,Zi),St[$i]=!0;var On=k.Object.getOwnPropertySymbols,Cn=tt.f,Sn=h((function(){Cn(1)}));gt({target:"Object",stat:!0,forced:!l||Sn,sham:!l},{getOwnPropertyDescriptor:function(t,e){return Cn(b(t),e)}});var Tn=n((function(t){var e=k.Object,i=t.exports=function(t,i){return e.getOwnPropertyDescriptor(t,i)};e.getOwnPropertyDescriptor.sham&&(i.sham=!0)})),Mn=Tn,Pn=x("Reflect","ownKeys")||function(t){var e=Ai.f(dt(t)),i=It.f;return i?e.concat(i(t)):e};gt({target:"Object",stat:!0,sham:!l},{getOwnPropertyDescriptors:function(t){for(var e,i,n=b(t),o=tt.f,r=Pn(n),s={},a=0;r.length>a;)void 0!==(i=o(n,e=r[a++]))&&vi(s,e,i);return s}});var Dn=k.Object.getOwnPropertyDescriptors;gt({target:"Object",stat:!0,forced:!l,sham:!l},{defineProperties:Pe});var In=n((function(t){var e=k.Object,i=t.exports=function(t,i){return e.defineProperties(t,i)};e.defineProperties.sham&&(i.sham=!0)}));gt({target:"Object",stat:!0,forced:!l,sham:!l},{defineProperty:ut.f});var Bn=n((function(t){var e=k.Object,i=t.exports=function(t,i,n){return e.defineProperty(t,i,n)};e.defineProperty.sham&&(i.sham=!0)})),zn=Bn,Nn=i(n((function(t){t.exports=function(t,e){if(!(t instanceof e))throw new TypeError("Cannot call a class as a function")},t.exports.default=t.exports,t.exports.__esModule=!0}))),An=Bn,Fn=i(n((function(t){function e(t,e){for(var i=0;i<e.length;i++){var n=e[i];n.enumerable=n.enumerable||!1,n.configurable=!0,"value"in n&&(n.writable=!0),An(t,n.key,n)}}t.exports=function(t,i,n){return i&&e(t.prototype,i),n&&e(t,n),t},t.exports.default=t.exports,t.exports.__esModule=!0}))),jn=i(n((function(t){t.exports=function(t,e,i){return e in t?An(t,e,{value:i,enumerable:!0,configurable:!0,writable:!0}):t[e]=i,t},t.exports.default=t.exports,t.exports.__esModule=!0})));gt({target:"Array",stat:!0},{isArray:zi});var Rn=k.Array.isArray,Ln=Rn,Hn=n((function(t){t.exports=function(t){if(Ln(t))return t},t.exports.default=t.exports,t.exports.__esModule=!0}));i(Hn);var Wn=U("species"),qn=function(t){return M>=51||!h((function(){var e=[];return(e.constructor={})[Wn]=function(){return{foo:1}},1!==e[t](Boolean).foo}))},Vn=U("isConcatSpreadable"),Un=9007199254740991,Yn="Maximum allowed index exceeded",Xn=M>=51||!h((function(){var t=[];return t[Vn]=!1,t.concat()[0]!==t})),Gn=qn("concat"),Kn=function(t){if(!w(t))return!1;var e=t[Vn];return void 0!==e?!!e:zi(t)};gt({target:"Array",proto:!0,forced:!Xn||!Gn},{concat:function(t){var e,i,n,o,r,s=A(this),a=Ui(s,0),h=0;for(e=-1,n=arguments.length;e<n;e++)if(Kn(r=-1===e?s:arguments[e])){if(h+(o=kt(r.length))>Un)throw TypeError(Yn);for(i=0;i<o;i++,h++)i in r&&vi(a,h,r[i])}else{if(h>=Un)throw TypeError(Yn);vi(a,h++,r)}return a.length=h,a}}),qi("asyncIterator"),qi("hasInstance"),qi("isConcatSpreadable"),qi("iterator"),qi("match"),qi("matchAll"),qi("replace"),qi("search"),qi("species"),qi("split"),qi("toPrimitive"),qi("toStringTag"),qi("unscopables"),Ye(a.JSON,"JSON",!0);var $n=k.Symbol;qi("asyncDispose"),qi("dispose"),qi("matcher"),qi("metadata"),qi("observable"),qi("patternMatch"),qi("replaceAll");var Zn=$n,Qn=n((function(t){t.exports=function(t,e){var i=null==t?null:void 0!==Zn&&Bi(t)||t["@@iterator"];if(null!=i){var n,o,r=[],s=!0,a=!1;try{for(i=i.call(t);!(s=(n=i.next()).done)&&(r.push(n.value),!e||r.length!==e);s=!0);}catch(t){a=!0,o=t}finally{try{s||null==i.return||i.return()}finally{if(a)throw o}}return r}},t.exports.default=t.exports,t.exports.__esModule=!0}));i(Qn);var Jn=qn("slice"),to=U("species"),eo=[].slice,io=Math.max;gt({target:"Array",proto:!0,forced:!Jn},{slice:function(t,e){var i,n,o,r=b(this),s=kt(r.length),a=Et(t,s),h=Et(void 0===e?s:e,s);if(zi(r)&&("function"!=typeof(i=r.constructor)||i!==Array&&!zi(i.prototype)?w(i)&&null===(i=i[to])&&(i=void 0):i=void 0,i===Array||void 0===i))return eo.call(r,a,h);for(n=new(void 0===i?Array:i)(io(h-a,0)),o=0;a<h;a++,o++)a in r&&vi(n,o,r[a]);return n.length=o,n}});var no=Ht("Array").slice,oo=Array.prototype,ro=function(t){var e=t.slice;return t===oo||t instanceof Array&&e===oo.slice?no:e},so=ro,ao=xi,ho=n((function(t){t.exports=function(t,e){(null==e||e>t.length)&&(e=t.length);for(var i=0,n=new Array(e);i<e;i++)n[i]=t[i];return n},t.exports.default=t.exports,t.exports.__esModule=!0}));i(ho);var lo=n((function(t){t.exports=function(t,e){var i;if(t){if("string"==typeof t)return ho(t,e);var n=so(i=Object.prototype.toString.call(t)).call(i,8,-1);return"Object"===n&&t.constructor&&(n=t.constructor.name),"Map"===n||"Set"===n?ao(t):"Arguments"===n||/^(?:Ui|I)nt(?:8|16|32)(?:Clamped)?Array$/.test(n)?ho(t,e):void 0}},t.exports.default=t.exports,t.exports.__esModule=!0}));i(lo);var co=n((function(t){t.exports=function(){throw new TypeError("Invalid attempt to destructure non-iterable instance.\nIn order to be iterable, non-array objects must have a [Symbol.iterator]() method.")},t.exports.default=t.exports,t.exports.__esModule=!0}));i(co);var uo=i(n((function(t){t.exports=function(t,e){return Hn(t)||Qn(t,e)||lo(t,e)||co()},t.exports.default=t.exports,t.exports.__esModule=!0}))),fo=Hi.f("iterator"),po=fo,vo=n((function(t){function e(i){return"function"==typeof Zn&&"symbol"==typeof po?(t.exports=e=function(t){return typeof t},t.exports.default=t.exports,t.exports.__esModule=!0):(t.exports=e=function(t){return t&&"function"==typeof Zn&&t.constructor===Zn&&t!==Zn.prototype?"symbol":typeof t},t.exports.default=t.exports,t.exports.__esModule=!0),e(i)}t.exports=e,t.exports.default=t.exports,t.exports.__esModule=!0})),go=i(vo),yo=n((function(t){t.exports=function(t){if(Ln(t))return ho(t)},t.exports.default=t.exports,t.exports.__esModule=!0}));i(yo);var mo=n((function(t){t.exports=function(t){if(void 0!==Zn&&null!=Bi(t)||null!=t["@@iterator"])return ao(t)},t.exports.default=t.exports,t.exports.__esModule=!0}));i(mo);var bo=n((function(t){t.exports=function(){throw new TypeError("Invalid attempt to spread non-iterable instance.\nIn order to be iterable, non-array objects must have a [Symbol.iterator]() method.")},t.exports.default=t.exports,t.exports.__esModule=!0}));i(bo);var wo=i(n((function(t){t.exports=function(t){return yo(t)||mo(t)||lo(t)||bo()},t.exports.default=t.exports,t.exports.__esModule=!0}))),ko=$n,_o=Ht("Array").concat,xo=Array.prototype,Eo=function(t){var e=t.concat;return t===xo||t instanceof Array&&e===xo.concat?_o:e},Oo=ro;gt({target:"Reflect",stat:!0},{ownKeys:Pn});var Co=k.Reflect.ownKeys,So=Rn,To=Gi.map,Mo=qn("map");gt({target:"Array",proto:!0,forced:!Mo},{map:function(t){return To(this,t,arguments.length>1?arguments[1]:void 0)}});var Po=Ht("Array").map,Do=Array.prototype,Io=function(t){var e=t.map;return t===Do||t instanceof Array&&e===Do.map?Po:e},Bo=h((function(){Dt(1)}));gt({target:"Object",stat:!0,forced:Bo},{keys:function(t){return Dt(A(t))}});var zo=k.Object.keys;gt({target:"Date",stat:!0},{now:function(){return(new Date).getTime()}});var No=k.Date.now,Ao=function(t,e){var i=[][t];return!!i&&h((function(){i.call(null,e||function(){throw 1},1)}))},Fo=Gi.forEach,jo=Ao("forEach")?[].forEach:function(t){return Fo(this,t,arguments.length>1?arguments[1]:void 0)};gt({target:"Array",proto:!0,forced:[].forEach!=jo},{forEach:jo});var Ro=Ht("Array").forEach,Lo=Array.prototype,Ho={DOMTokenList:!0,NodeList:!0},Wo=function(t){var e=t.forEach;return t===Lo||t instanceof Array&&e===Lo.forEach||Ho.hasOwnProperty(We(t))?Ro:e},qo=[].reverse,Vo=[1,2];gt({target:"Array",proto:!0,forced:String(Vo)===String(Vo.reverse())},{reverse:function(){return zi(this)&&(this.length=this.length),qo.call(this)}});var Uo=Ht("Array").reverse,Yo=Array.prototype,Xo=function(t){var e=t.reverse;return t===Yo||t instanceof Array&&e===Yo.reverse?Uo:e},Go=qn("splice"),Ko=Math.max,$o=Math.min,Zo=9007199254740991,Qo="Maximum allowed length exceeded";gt({target:"Array",proto:!0,forced:!Go},{splice:function(t,e){var i,n,o,r,s,a,h=A(this),l=kt(h.length),d=Et(t,l),c=arguments.length;if(0===c?i=n=0:1===c?(i=0,n=l-d):(i=c-2,n=$o(Ko(bt(e),0),l-d)),l+i-n>Zo)throw TypeError(Qo);for(o=Ui(h,n),r=0;r<n;r++)(s=d+r)in h&&vi(o,r,h[s]);if(o.length=n,i<n){for(r=d;r<l-n;r++)a=r+i,(s=r+n)in h?h[a]=h[s]:delete h[a];for(r=l;r>l-n+i;r--)delete h[r-1]}else if(i>n)for(r=l-n;r>d;r--)a=r+i-1,(s=r+n-1)in h?h[a]=h[s]:delete h[a];for(r=0;r<i;r++)h[r+d]=arguments[r+2];return h.length=l-n+i,o}});var Jo=Ht("Array").splice,tr=Array.prototype,er=function(t){var e=t.splice;return t===tr||t instanceof Array&&e===tr.splice?Jo:e},ir=Ct.includes;gt({target:"Array",proto:!0},{includes:function(t){return ir(this,t,arguments.length>1?arguments[1]:void 0)}});var nr=Ht("Array").includes,or=U("match"),rr=function(t){if(function(t){var e;return w(t)&&(void 0!==(e=t[or])?!!e:"RegExp"==v(t))}(t))throw TypeError("The method doesn't accept regular expressions");return t},sr=U("match");gt({target:"String",proto:!0,forced:!function(t){var e=/./;try{"/./"[t](e)}catch(i){try{return e[sr]=!1,"/./"[t](e)}catch(t){}}return!1}("includes")},{includes:function(t){return!!~Qt(m(this)).indexOf(Qt(rr(t)),arguments.length>1?arguments[1]:void 0)}});var ar=Ht("String").includes,hr=Array.prototype,lr=String.prototype,dr=function(t){var e=t.includes;return t===hr||t instanceof Array&&e===hr.includes?nr:"string"==typeof t||t===lr||t instanceof String&&e===lr.includes?ar:e},cr=h((function(){Ee(1)}));gt({target:"Object",stat:!0,forced:cr,sham:!ke},{getPrototypeOf:function(t){return Ee(A(t))}});var ur=k.Object.getPrototypeOf,fr=ur,pr=Gi.filter,vr=qn("filter");gt({target:"Array",proto:!0,forced:!vr},{filter:function(t){return pr(this,t,arguments.length>1?arguments[1]:void 0)}});var gr=Ht("Array").filter,yr=Array.prototype,mr=function(t){var e=t.filter;return t===yr||t instanceof Array&&e===yr.filter?gr:e},br=u.f,wr=function(t){return function(e){for(var i,n=b(e),o=Dt(n),r=o.length,s=0,a=[];r>s;)i=o[s++],l&&!br.call(n,i)||a.push(t?[i,n[i]]:n[i]);return a}},kr={entries:wr(!0),values:wr(!1)}.values;gt({target:"Object",stat:!0},{values:function(t){return kr(t)}});var _r=k.Object.values,xr="\t\n\v\f\r                　\u2028\u2029\ufeff",Er="["+xr+"]",Or=RegExp("^"+Er+Er+"*"),Cr=RegExp(Er+Er+"*$"),Sr=function(t){return function(e){var i=Qt(m(e));return 1&t&&(i=i.replace(Or,"")),2&t&&(i=i.replace(Cr,"")),i}},Tr={start:Sr(1),end:Sr(2),trim:Sr(3)},Mr=Tr.trim,Pr=a.parseInt,Dr=/^[+-]?0[Xx]/,Ir=8!==Pr(xr+"08")||22!==Pr(xr+"0x16")?function(t,e){var i=Mr(Qt(t));return Pr(i,e>>>0||(Dr.test(i)?16:10))}:Pr;gt({global:!0,forced:parseInt!=Ir},{parseInt:Ir});var Br=k.parseInt,zr=Ct.indexOf,Nr=[].indexOf,Ar=!!Nr&&1/[1].indexOf(1,-0)<0,Fr=Ao("indexOf");gt({target:"Array",proto:!0,forced:Ar||!Fr},{indexOf:function(t){return Ar?Nr.apply(this,arguments)||0:zr(this,t,arguments.length>1?arguments[1]:void 0)}});var jr,Rr=Ht("Array").indexOf,Lr=Array.prototype,Hr=function(t){var e=t.indexOf;return t===Lr||t instanceof Array&&e===Lr.indexOf?Rr:e},Wr=Tr.trim;gt({target:"String",proto:!0,forced:(jr="trim",h((function(){return!!xr[jr]()||"​᠎"!="​᠎"[jr]()||xr[jr].name!==jr})))},{trim:function(){return Wr(this)}});var qr=Ht("String").trim,Vr=String.prototype,Ur=function(t){var e=t.trim;return"string"==typeof t||t===Vr||t instanceof String&&e===Vr.trim?qr:e};gt({target:"Object",stat:!0,sham:!l},{create:Fe});var Yr=k.Object,Xr=function(t,e){return Yr.create(t,e)},Gr=Xr,Kr=x("JSON","stringify"),$r=/[\uD800-\uDFFF]/g,Zr=/^[\uD800-\uDBFF]$/,Qr=/^[\uDC00-\uDFFF]$/,Jr=function(t,e,i){var n=i.charAt(e-1),o=i.charAt(e+1);return Zr.test(t)&&!Qr.test(o)||Qr.test(t)&&!Zr.test(n)?"\\u"+t.charCodeAt(0).toString(16):t},ts=h((function(){return'"\\udf06\\ud834"'!==Kr("\udf06\ud834")||'"\\udead"'!==Kr("\udead")}));Kr&&gt({target:"JSON",stat:!0,forced:ts},{stringify:function(t,e,i){var n=Kr.apply(null,arguments);return"string"==typeof n?n.replace($r,Jr):n}}),k.JSON||(k.JSON={stringify:JSON.stringify});var es=function(t,e,i){return k.JSON.stringify.apply(null,arguments)},is=[].slice,ns=/MSIE .\./.test(E),os=function(t){return function(e,i){var n=arguments.length>2,o=n?is.call(arguments,2):void 0;return t(n?function(){("function"==typeof e?e:Function(e)).apply(this,o)}:e,i)}};gt({global:!0,bind:!0,forced:ns},{setTimeout:os(a.setTimeout),setInterval:os(a.setInterval)});var rs=k.setTimeout;gt({target:"Array",proto:!0},{fill:function(t){for(var e=A(this),i=kt(e.length),n=arguments.length,o=Et(n>1?arguments[1]:void 0,i),r=n>2?arguments[2]:void 0,s=void 0===r?i:Et(r,i);s>o;)e[o++]=t;return e}});var ss=Ht("Array").fill,as=Array.prototype,hs=function(t){var e=t.fill;return t===as||t instanceof Array&&e===as.fill?ss:e};function ls(){return(ls=Object.assign||function(t){for(var e=1;e<arguments.length;e++){var i=arguments[e];for(var n in i)Object.prototype.hasOwnProperty.call(i,n)&&(t[n]=i[n])}return t}).apply(this,arguments)}function ds(t,e){t.prototype=Object.create(e.prototype),t.prototype.constructor=t,t.__proto__=e}function cs(t){if(void 0===t)throw new ReferenceError("this hasn't been initialised - super() hasn't been called");return t}var us,fs="function"!=typeof Object.assign?function(t){if(null==t)throw new TypeError("Cannot convert undefined or null to object");for(var e=Object(t),i=1;i<arguments.length;i++){var n=arguments[i];if(null!=n)for(var o in n)n.hasOwnProperty(o)&&(e[o]=n[o])}return e}:Object.assign,ps=["","webkit","Moz","MS","ms","o"],vs="undefined"==typeof document?{style:{}}:document.createElement("div"),gs=Math.round,ys=Math.abs,ms=Date.now;function bs(t,e){for(var i,n,o=e[0].toUpperCase()+e.slice(1),r=0;r<ps.length;){if((n=(i=ps[r])?i+o:e)in t)return n;r++}}us="undefined"==typeof window?{}:window;var ws=bs(vs.style,"touchAction"),ks=void 0!==ws;var _s="compute",xs="auto",Es="manipulation",Os="none",Cs="pan-x",Ss="pan-y",Ts=function(){if(!ks)return!1;var t={},e=us.CSS&&us.CSS.supports;return["auto","manipulation","pan-y","pan-x","pan-x pan-y","none"].forEach((function(i){return t[i]=!e||us.CSS.supports("touch-action",i)})),t}(),Ms="ontouchstart"in us,Ps=void 0!==bs(us,"PointerEvent"),Ds=Ms&&/mobile|tablet|ip(ad|hone|od)|android/i.test(navigator.userAgent),Is="touch",Bs="mouse",zs=16,Ns=24,As=["x","y"],Fs=["clientX","clientY"];function js(t,e,i){var n;if(t)if(t.forEach)t.forEach(e,i);else if(void 0!==t.length)for(n=0;n<t.length;)e.call(i,t[n],n,t),n++;else for(n in t)t.hasOwnProperty(n)&&e.call(i,t[n],n,t)}function Rs(t,e){return"function"==typeof t?t.apply(e&&e[0]||void 0,e):t}function Ls(t,e){return t.indexOf(e)>-1}var Hs=function(){function t(t,e){this.manager=t,this.set(e)}var e=t.prototype;return e.set=function(t){t===_s&&(t=this.compute()),ks&&this.manager.element.style&&Ts[t]&&(this.manager.element.style[ws]=t),this.actions=t.toLowerCase().trim()},e.update=function(){this.set(this.manager.options.touchAction)},e.compute=function(){var t=[];return js(this.manager.recognizers,(function(e){Rs(e.options.enable,[e])&&(t=t.concat(e.getTouchAction()))})),function(t){if(Ls(t,Os))return Os;var e=Ls(t,Cs),i=Ls(t,Ss);return e&&i?Os:e||i?e?Cs:Ss:Ls(t,Es)?Es:xs}(t.join(" "))},e.preventDefaults=function(t){var e=t.srcEvent,i=t.offsetDirection;if(this.manager.session.prevented)e.preventDefault();else{var n=this.actions,o=Ls(n,Os)&&!Ts.none,r=Ls(n,Ss)&&!Ts["pan-y"],s=Ls(n,Cs)&&!Ts["pan-x"];if(o){var a=1===t.pointers.length,h=t.distance<2,l=t.deltaTime<250;if(a&&h&&l)return}if(!s||!r)return o||r&&6&i||s&&i&Ns?this.preventSrc(e):void 0}},e.preventSrc=function(t){this.manager.session.prevented=!0,t.preventDefault()},t}();function Ws(t,e){for(;t;){if(t===e)return!0;t=t.parentNode}return!1}function qs(t){var e=t.length;if(1===e)return{x:gs(t[0].clientX),y:gs(t[0].clientY)};for(var i=0,n=0,o=0;o<e;)i+=t[o].clientX,n+=t[o].clientY,o++;return{x:gs(i/e),y:gs(n/e)}}function Vs(t){for(var e=[],i=0;i<t.pointers.length;)e[i]={clientX:gs(t.pointers[i].clientX),clientY:gs(t.pointers[i].clientY)},i++;return{timeStamp:ms(),pointers:e,center:qs(e),deltaX:t.deltaX,deltaY:t.deltaY}}function Us(t,e,i){i||(i=As);var n=e[i[0]]-t[i[0]],o=e[i[1]]-t[i[1]];return Math.sqrt(n*n+o*o)}function Ys(t,e,i){i||(i=As);var n=e[i[0]]-t[i[0]],o=e[i[1]]-t[i[1]];return 180*Math.atan2(o,n)/Math.PI}function Xs(t,e){return t===e?1:ys(t)>=ys(e)?t<0?2:4:e<0?8:zs}function Gs(t,e,i){return{x:e/t||0,y:i/t||0}}function Ks(t,e){var i=t.session,n=e.pointers,o=n.length;i.firstInput||(i.firstInput=Vs(e)),o>1&&!i.firstMultiple?i.firstMultiple=Vs(e):1===o&&(i.firstMultiple=!1);var r=i.firstInput,s=i.firstMultiple,a=s?s.center:r.center,h=e.center=qs(n);e.timeStamp=ms(),e.deltaTime=e.timeStamp-r.timeStamp,e.angle=Ys(a,h),e.distance=Us(a,h),function(t,e){var i=e.center,n=t.offsetDelta||{},o=t.prevDelta||{},r=t.prevInput||{};1!==e.eventType&&4!==r.eventType||(o=t.prevDelta={x:r.deltaX||0,y:r.deltaY||0},n=t.offsetDelta={x:i.x,y:i.y}),e.deltaX=o.x+(i.x-n.x),e.deltaY=o.y+(i.y-n.y)}(i,e),e.offsetDirection=Xs(e.deltaX,e.deltaY);var l,d,c=Gs(e.deltaTime,e.deltaX,e.deltaY);e.overallVelocityX=c.x,e.overallVelocityY=c.y,e.overallVelocity=ys(c.x)>ys(c.y)?c.x:c.y,e.scale=s?(l=s.pointers,Us((d=n)[0],d[1],Fs)/Us(l[0],l[1],Fs)):1,e.rotation=s?function(t,e){return Ys(e[1],e[0],Fs)+Ys(t[1],t[0],Fs)}(s.pointers,n):0,e.maxPointers=i.prevInput?e.pointers.length>i.prevInput.maxPointers?e.pointers.length:i.prevInput.maxPointers:e.pointers.length,function(t,e){var i,n,o,r,s=t.lastInterval||e,a=e.timeStamp-s.timeStamp;if(8!==e.eventType&&(a>25||void 0===s.velocity)){var h=e.deltaX-s.deltaX,l=e.deltaY-s.deltaY,d=Gs(a,h,l);n=d.x,o=d.y,i=ys(d.x)>ys(d.y)?d.x:d.y,r=Xs(h,l),t.lastInterval=e}else i=s.velocity,n=s.velocityX,o=s.velocityY,r=s.direction;e.velocity=i,e.velocityX=n,e.velocityY=o,e.direction=r}(i,e);var u,f=t.element,p=e.srcEvent;Ws(u=p.composedPath?p.composedPath()[0]:p.path?p.path[0]:p.target,f)&&(f=u),e.target=f}function $s(t,e,i){var n=i.pointers.length,o=i.changedPointers.length,r=1&e&&n-o==0,s=12&e&&n-o==0;i.isFirst=!!r,i.isFinal=!!s,r&&(t.session={}),i.eventType=e,Ks(t,i),t.emit("hammer.input",i),t.recognize(i),t.session.prevInput=i}function Zs(t){return t.trim().split(/\s+/g)}function Qs(t,e,i){js(Zs(e),(function(e){t.addEventListener(e,i,!1)}))}function Js(t,e,i){js(Zs(e),(function(e){t.removeEventListener(e,i,!1)}))}function ta(t){var e=t.ownerDocument||t;return e.defaultView||e.parentWindow||window}var ea=function(){function t(t,e){var i=this;this.manager=t,this.callback=e,this.element=t.element,this.target=t.options.inputTarget,this.domHandler=function(e){Rs(t.options.enable,[t])&&i.handler(e)},this.init()}var e=t.prototype;return e.handler=function(){},e.init=function(){this.evEl&&Qs(this.element,this.evEl,this.domHandler),this.evTarget&&Qs(this.target,this.evTarget,this.domHandler),this.evWin&&Qs(ta(this.element),this.evWin,this.domHandler)},e.destroy=function(){this.evEl&&Js(this.element,this.evEl,this.domHandler),this.evTarget&&Js(this.target,this.evTarget,this.domHandler),this.evWin&&Js(ta(this.element),this.evWin,this.domHandler)},t}();function ia(t,e,i){if(t.indexOf&&!i)return t.indexOf(e);for(var n=0;n<t.length;){if(i&&t[n][i]==e||!i&&t[n]===e)return n;n++}return-1}var na={pointerdown:1,pointermove:2,pointerup:4,pointercancel:8,pointerout:8},oa={2:Is,3:"pen",4:Bs,5:"kinect"},ra="pointerdown",sa="pointermove pointerup pointercancel";us.MSPointerEvent&&!us.PointerEvent&&(ra="MSPointerDown",sa="MSPointerMove MSPointerUp MSPointerCancel");var aa=function(t){function e(){var i,n=e.prototype;return n.evEl=ra,n.evWin=sa,(i=t.apply(this,arguments)||this).store=i.manager.session.pointerEvents=[],i}return ds(e,t),e.prototype.handler=function(t){var e=this.store,i=!1,n=t.type.toLowerCase().replace("ms",""),o=na[n],r=oa[t.pointerType]||t.pointerType,s=r===Is,a=ia(e,t.pointerId,"pointerId");1&o&&(0===t.button||s)?a<0&&(e.push(t),a=e.length-1):12&o&&(i=!0),a<0||(e[a]=t,this.callback(this.manager,o,{pointers:e,changedPointers:[t],pointerType:r,srcEvent:t}),i&&e.splice(a,1))},e}(ea);function ha(t){return Array.prototype.slice.call(t,0)}function la(t,e,i){for(var n=[],o=[],r=0;r<t.length;){var s=e?t[r][e]:t[r];ia(o,s)<0&&n.push(t[r]),o[r]=s,r++}return i&&(n=e?n.sort((function(t,i){return t[e]>i[e]})):n.sort()),n}var da={touchstart:1,touchmove:2,touchend:4,touchcancel:8},ca="touchstart touchmove touchend touchcancel",ua=function(t){function e(){var i;return e.prototype.evTarget=ca,(i=t.apply(this,arguments)||this).targetIds={},i}return ds(e,t),e.prototype.handler=function(t){var e=da[t.type],i=fa.call(this,t,e);i&&this.callback(this.manager,e,{pointers:i[0],changedPointers:i[1],pointerType:Is,srcEvent:t})},e}(ea);function fa(t,e){var i,n,o=ha(t.touches),r=this.targetIds;if(3&e&&1===o.length)return r[o[0].identifier]=!0,[o,o];var s=ha(t.changedTouches),a=[],h=this.target;if(n=o.filter((function(t){return Ws(t.target,h)})),1===e)for(i=0;i<n.length;)r[n[i].identifier]=!0,i++;for(i=0;i<s.length;)r[s[i].identifier]&&a.push(s[i]),12&e&&delete r[s[i].identifier],i++;return a.length?[la(n.concat(a),"identifier",!0),a]:void 0}var pa={mousedown:1,mousemove:2,mouseup:4},va="mousedown",ga="mousemove mouseup",ya=function(t){function e(){var i,n=e.prototype;return n.evEl=va,n.evWin=ga,(i=t.apply(this,arguments)||this).pressed=!1,i}return ds(e,t),e.prototype.handler=function(t){var e=pa[t.type];1&e&&0===t.button&&(this.pressed=!0),2&e&&1!==t.which&&(e=4),this.pressed&&(4&e&&(this.pressed=!1),this.callback(this.manager,e,{pointers:[t],changedPointers:[t],pointerType:Bs,srcEvent:t}))},e}(ea);function ma(t){var e=t.changedPointers[0];if(e.identifier===this.primaryTouch){var i={x:e.clientX,y:e.clientY},n=this.lastTouches;this.lastTouches.push(i);setTimeout((function(){var t=n.indexOf(i);t>-1&&n.splice(t,1)}),2500)}}function ba(t,e){1&t?(this.primaryTouch=e.changedPointers[0].identifier,ma.call(this,e)):12&t&&ma.call(this,e)}function wa(t){for(var e=t.srcEvent.clientX,i=t.srcEvent.clientY,n=0;n<this.lastTouches.length;n++){var o=this.lastTouches[n],r=Math.abs(e-o.x),s=Math.abs(i-o.y);if(r<=25&&s<=25)return!0}return!1}var ka=function(){return function(t){function e(e,i){var n;return(n=t.call(this,e,i)||this).handler=function(t,e,i){var o=i.pointerType===Is,r=i.pointerType===Bs;if(!(r&&i.sourceCapabilities&&i.sourceCapabilities.firesTouchEvents)){if(o)ba.call(cs(cs(n)),e,i);else if(r&&wa.call(cs(cs(n)),i))return;n.callback(t,e,i)}},n.touch=new ua(n.manager,n.handler),n.mouse=new ya(n.manager,n.handler),n.primaryTouch=null,n.lastTouches=[],n}return ds(e,t),e.prototype.destroy=function(){this.touch.destroy(),this.mouse.destroy()},e}(ea)}();function _a(t,e,i){return!!Array.isArray(t)&&(js(t,i[e],i),!0)}var xa=32,Ea=1;function Oa(t,e){var i=e.manager;return i?i.get(t):t}function Ca(t){return 16&t?"cancel":8&t?"end":4&t?"move":2&t?"start":""}var Sa=function(){function t(t){void 0===t&&(t={}),this.options=ls({enable:!0},t),this.id=Ea++,this.manager=null,this.state=1,this.simultaneous={},this.requireFail=[]}var e=t.prototype;return e.set=function(t){return fs(this.options,t),this.manager&&this.manager.touchAction.update(),this},e.recognizeWith=function(t){if(_a(t,"recognizeWith",this))return this;var e=this.simultaneous;return e[(t=Oa(t,this)).id]||(e[t.id]=t,t.recognizeWith(this)),this},e.dropRecognizeWith=function(t){return _a(t,"dropRecognizeWith",this)||(t=Oa(t,this),delete this.simultaneous[t.id]),this},e.requireFailure=function(t){if(_a(t,"requireFailure",this))return this;var e=this.requireFail;return-1===ia(e,t=Oa(t,this))&&(e.push(t),t.requireFailure(this)),this},e.dropRequireFailure=function(t){if(_a(t,"dropRequireFailure",this))return this;t=Oa(t,this);var e=ia(this.requireFail,t);return e>-1&&this.requireFail.splice(e,1),this},e.hasRequireFailures=function(){return this.requireFail.length>0},e.canRecognizeWith=function(t){return!!this.simultaneous[t.id]},e.emit=function(t){var e=this,i=this.state;function n(i){e.manager.emit(i,t)}i<8&&n(e.options.event+Ca(i)),n(e.options.event),t.additionalEvent&&n(t.additionalEvent),i>=8&&n(e.options.event+Ca(i))},e.tryEmit=function(t){if(this.canEmit())return this.emit(t);this.state=xa},e.canEmit=function(){for(var t=0;t<this.requireFail.length;){if(!(33&this.requireFail[t].state))return!1;t++}return!0},e.recognize=function(t){var e=fs({},t);if(!Rs(this.options.enable,[this,e]))return this.reset(),void(this.state=xa);56&this.state&&(this.state=1),this.state=this.process(e),30&this.state&&this.tryEmit(e)},e.process=function(t){},e.getTouchAction=function(){},e.reset=function(){},t}(),Ta=function(t){function e(e){var i;return void 0===e&&(e={}),(i=t.call(this,ls({event:"tap",pointers:1,taps:1,interval:300,time:250,threshold:9,posThreshold:10},e))||this).pTime=!1,i.pCenter=!1,i._timer=null,i._input=null,i.count=0,i}ds(e,t);var i=e.prototype;return i.getTouchAction=function(){return[Es]},i.process=function(t){var e=this,i=this.options,n=t.pointers.length===i.pointers,o=t.distance<i.threshold,r=t.deltaTime<i.time;if(this.reset(),1&t.eventType&&0===this.count)return this.failTimeout();if(o&&r&&n){if(4!==t.eventType)return this.failTimeout();var s=!this.pTime||t.timeStamp-this.pTime<i.interval,a=!this.pCenter||Us(this.pCenter,t.center)<i.posThreshold;if(this.pTime=t.timeStamp,this.pCenter=t.center,a&&s?this.count+=1:this.count=1,this._input=t,0===this.count%i.taps)return this.hasRequireFailures()?(this._timer=setTimeout((function(){e.state=8,e.tryEmit()}),i.interval),2):8}return xa},i.failTimeout=function(){var t=this;return this._timer=setTimeout((function(){t.state=xa}),this.options.interval),xa},i.reset=function(){clearTimeout(this._timer)},i.emit=function(){8===this.state&&(this._input.tapCount=this.count,this.manager.emit(this.options.event,this._input))},e}(Sa),Ma=function(t){function e(e){return void 0===e&&(e={}),t.call(this,ls({pointers:1},e))||this}ds(e,t);var i=e.prototype;return i.attrTest=function(t){var e=this.options.pointers;return 0===e||t.pointers.length===e},i.process=function(t){var e=this.state,i=t.eventType,n=6&e,o=this.attrTest(t);return n&&(8&i||!o)?16|e:n||o?4&i?8|e:2&e?4|e:2:xa},e}(Sa);function Pa(t){return t===zs?"down":8===t?"up":2===t?"left":4===t?"right":""}var Da=function(t){function e(e){var i;return void 0===e&&(e={}),(i=t.call(this,ls({event:"pan",threshold:10,pointers:1,direction:30},e))||this).pX=null,i.pY=null,i}ds(e,t);var i=e.prototype;return i.getTouchAction=function(){var t=this.options.direction,e=[];return 6&t&&e.push(Ss),t&Ns&&e.push(Cs),e},i.directionTest=function(t){var e=this.options,i=!0,n=t.distance,o=t.direction,r=t.deltaX,s=t.deltaY;return o&e.direction||(6&e.direction?(o=0===r?1:r<0?2:4,i=r!==this.pX,n=Math.abs(t.deltaX)):(o=0===s?1:s<0?8:zs,i=s!==this.pY,n=Math.abs(t.deltaY))),t.direction=o,i&&n>e.threshold&&o&e.direction},i.attrTest=function(t){return Ma.prototype.attrTest.call(this,t)&&(2&this.state||!(2&this.state)&&this.directionTest(t))},i.emit=function(e){this.pX=e.deltaX,this.pY=e.deltaY;var i=Pa(e.direction);i&&(e.additionalEvent=this.options.event+i),t.prototype.emit.call(this,e)},e}(Ma),Ia=function(t){function e(e){return void 0===e&&(e={}),t.call(this,ls({event:"swipe",threshold:10,velocity:.3,direction:30,pointers:1},e))||this}ds(e,t);var i=e.prototype;return i.getTouchAction=function(){return Da.prototype.getTouchAction.call(this)},i.attrTest=function(e){var i,n=this.options.direction;return 30&n?i=e.overallVelocity:6&n?i=e.overallVelocityX:n&Ns&&(i=e.overallVelocityY),t.prototype.attrTest.call(this,e)&&n&e.offsetDirection&&e.distance>this.options.threshold&&e.maxPointers===this.options.pointers&&ys(i)>this.options.velocity&&4&e.eventType},i.emit=function(t){var e=Pa(t.offsetDirection);e&&this.manager.emit(this.options.event+e,t),this.manager.emit(this.options.event,t)},e}(Ma),Ba=function(t){function e(e){return void 0===e&&(e={}),t.call(this,ls({event:"pinch",threshold:0,pointers:2},e))||this}ds(e,t);var i=e.prototype;return i.getTouchAction=function(){return[Os]},i.attrTest=function(e){return t.prototype.attrTest.call(this,e)&&(Math.abs(e.scale-1)>this.options.threshold||2&this.state)},i.emit=function(e){if(1!==e.scale){var i=e.scale<1?"in":"out";e.additionalEvent=this.options.event+i}t.prototype.emit.call(this,e)},e}(Ma),za=function(t){function e(e){return void 0===e&&(e={}),t.call(this,ls({event:"rotate",threshold:0,pointers:2},e))||this}ds(e,t);var i=e.prototype;return i.getTouchAction=function(){return[Os]},i.attrTest=function(e){return t.prototype.attrTest.call(this,e)&&(Math.abs(e.rotation)>this.options.threshold||2&this.state)},e}(Ma),Na=function(t){function e(e){var i;return void 0===e&&(e={}),(i=t.call(this,ls({event:"press",pointers:1,time:251,threshold:9},e))||this)._timer=null,i._input=null,i}ds(e,t);var i=e.prototype;return i.getTouchAction=function(){return[xs]},i.process=function(t){var e=this,i=this.options,n=t.pointers.length===i.pointers,o=t.distance<i.threshold,r=t.deltaTime>i.time;if(this._input=t,!o||!n||12&t.eventType&&!r)this.reset();else if(1&t.eventType)this.reset(),this._timer=setTimeout((function(){e.state=8,e.tryEmit()}),i.time);else if(4&t.eventType)return 8;return xa},i.reset=function(){clearTimeout(this._timer)},i.emit=function(t){8===this.state&&(t&&4&t.eventType?this.manager.emit(this.options.event+"up",t):(this._input.timeStamp=ms(),this.manager.emit(this.options.event,this._input)))},e}(Sa),Aa={domEvents:!1,touchAction:_s,enable:!0,inputTarget:null,inputClass:null,cssProps:{userSelect:"none",touchSelect:"none",touchCallout:"none",contentZooming:"none",userDrag:"none",tapHighlightColor:"rgba(0,0,0,0)"}},Fa=[[za,{enable:!1}],[Ba,{enable:!1},["rotate"]],[Ia,{direction:6}],[Da,{direction:6},["swipe"]],[Ta],[Ta,{event:"doubletap",taps:2},["tap"]],[Na]];function ja(t,e){var i,n=t.element;n.style&&(js(t.options.cssProps,(function(o,r){i=bs(n.style,r),e?(t.oldCssProps[i]=n.style[i],n.style[i]=o):n.style[i]=t.oldCssProps[i]||""})),e||(t.oldCssProps={}))}var Ra=function(){function t(t,e){var i,n=this;this.options=fs({},Aa,e||{}),this.options.inputTarget=this.options.inputTarget||t,this.handlers={},this.session={},this.recognizers=[],this.oldCssProps={},this.element=t,this.input=new((i=this).options.inputClass||(Ps?aa:Ds?ua:Ms?ka:ya))(i,$s),this.touchAction=new Hs(this,this.options.touchAction),ja(this,!0),js(this.options.recognizers,(function(t){var e=n.add(new t[0](t[1]));t[2]&&e.recognizeWith(t[2]),t[3]&&e.requireFailure(t[3])}),this)}var e=t.prototype;return e.set=function(t){return fs(this.options,t),t.touchAction&&this.touchAction.update(),t.inputTarget&&(this.input.destroy(),this.input.target=t.inputTarget,this.input.init()),this},e.stop=function(t){this.session.stopped=t?2:1},e.recognize=function(t){var e=this.session;if(!e.stopped){var i;this.touchAction.preventDefaults(t);var n=this.recognizers,o=e.curRecognizer;(!o||o&&8&o.state)&&(e.curRecognizer=null,o=null);for(var r=0;r<n.length;)i=n[r],2===e.stopped||o&&i!==o&&!i.canRecognizeWith(o)?i.reset():i.recognize(t),!o&&14&i.state&&(e.curRecognizer=i,o=i),r++}},e.get=function(t){if(t instanceof Sa)return t;for(var e=this.recognizers,i=0;i<e.length;i++)if(e[i].options.event===t)return e[i];return null},e.add=function(t){if(_a(t,"add",this))return this;var e=this.get(t.options.event);return e&&this.remove(e),this.recognizers.push(t),t.manager=this,this.touchAction.update(),t},e.remove=function(t){if(_a(t,"remove",this))return this;var e=this.get(t);if(t){var i=this.recognizers,n=ia(i,e);-1!==n&&(i.splice(n,1),this.touchAction.update())}return this},e.on=function(t,e){if(void 0===t||void 0===e)return this;var i=this.handlers;return js(Zs(t),(function(t){i[t]=i[t]||[],i[t].push(e)})),this},e.off=function(t,e){if(void 0===t)return this;var i=this.handlers;return js(Zs(t),(function(t){e?i[t]&&i[t].splice(ia(i[t],e),1):delete i[t]})),this},e.emit=function(t,e){this.options.domEvents&&function(t,e){var i=document.createEvent("Event");i.initEvent(t,!0,!0),i.gesture=e,e.target.dispatchEvent(i)}(t,e);var i=this.handlers[t]&&this.handlers[t].slice();if(i&&i.length){e.type=t,e.preventDefault=function(){e.srcEvent.preventDefault()};for(var n=0;n<i.length;)i[n](e),n++}},e.destroy=function(){this.element&&ja(this,!1),this.handlers={},this.session={},this.input.destroy(),this.element=null},t}(),La={touchstart:1,touchmove:2,touchend:4,touchcancel:8},Ha="touchstart",Wa="touchstart touchmove touchend touchcancel",qa=function(t){function e(){var i,n=e.prototype;return n.evTarget=Ha,n.evWin=Wa,(i=t.apply(this,arguments)||this).started=!1,i}return ds(e,t),e.prototype.handler=function(t){var e=La[t.type];if(1===e&&(this.started=!0),this.started){var i=Va.call(this,t,e);12&e&&i[0].length-i[1].length==0&&(this.started=!1),this.callback(this.manager,e,{pointers:i[0],changedPointers:i[1],pointerType:Is,srcEvent:t})}},e}(ea);function Va(t,e){var i=ha(t.touches),n=ha(t.changedTouches);return 12&e&&(i=la(i.concat(n),"identifier",!0)),[i,n]}function Ua(t,e,i){var n="DEPRECATED METHOD: "+e+"\n"+i+" AT \n";return function(){var e=new Error("get-stack-trace"),i=e&&e.stack?e.stack.replace(/^[^\(]+?[\n$]/gm,"").replace(/^\s+at\s+/gm,"").replace(/^Object.<anonymous>\s*\(/gm,"{anonymous}()@"):"Unknown Stack Trace",o=window.console&&(window.console.warn||window.console.log);return o&&o.call(window.console,n,i),t.apply(this,arguments)}}var Ya=Ua((function(t,e,i){for(var n=Object.keys(e),o=0;o<n.length;)(!i||i&&void 0===t[n[o]])&&(t[n[o]]=e[n[o]]),o++;return t}),"extend","Use `assign`."),Xa=Ua((function(t,e){return Ya(t,e,!0)}),"merge","Use `assign`.");function Ga(t,e,i){var n,o=e.prototype;(n=t.prototype=Object.create(o)).constructor=t,n._super=o,i&&fs(n,i)}function Ka(t,e){return function(){return t.apply(e,arguments)}}var $a=function(){var t=function(t,e){return void 0===e&&(e={}),new Ra(t,ls({recognizers:Fa.concat()},e))};return t.VERSION="2.0.17-rc",t.DIRECTION_ALL=30,t.DIRECTION_DOWN=zs,t.DIRECTION_LEFT=2,t.DIRECTION_RIGHT=4,t.DIRECTION_UP=8,t.DIRECTION_HORIZONTAL=6,t.DIRECTION_VERTICAL=Ns,t.DIRECTION_NONE=1,t.DIRECTION_DOWN=zs,t.INPUT_START=1,t.INPUT_MOVE=2,t.INPUT_END=4,t.INPUT_CANCEL=8,t.STATE_POSSIBLE=1,t.STATE_BEGAN=2,t.STATE_CHANGED=4,t.STATE_ENDED=8,t.STATE_RECOGNIZED=8,t.STATE_CANCELLED=16,t.STATE_FAILED=xa,t.Manager=Ra,t.Input=ea,t.TouchAction=Hs,t.TouchInput=ua,t.MouseInput=ya,t.PointerEventInput=aa,t.TouchMouseInput=ka,t.SingleTouchInput=qa,t.Recognizer=Sa,t.AttrRecognizer=Ma,t.Tap=Ta,t.Pan=Da,t.Swipe=Ia,t.Pinch=Ba,t.Rotate=za,t.Press=Na,t.on=Qs,t.off=Js,t.each=js,t.merge=Xa,t.extend=Ya,t.bindFn=Ka,t.assign=fs,t.inherit=Ga,t.bindFn=Ka,t.prefixed=bs,t.toArray=ha,t.inArray=ia,t.uniqueArray=la,t.splitStr=Zs,t.boolOrFn=Rs,t.hasParent=Ws,t.addEventListeners=Qs,t.removeEventListeners=Js,t.defaults=fs({},Aa,{preset:Fa}),t}();function Za(t,e){var i=zo(t);if(On){var n=On(t);e&&(n=mr(n).call(n,(function(e){return Mn(t,e).enumerable}))),i.push.apply(i,n)}return i}function Qa(t){for(var e=1;e<arguments.length;e++){var i,n=null!=arguments[e]?arguments[e]:{};if(e%2)Wo(i=Za(Object(n),!0)).call(i,(function(e){jn(t,e,n[e])}));else if(Dn)In(t,Dn(n));else{var o;Wo(o=Za(Object(n))).call(o,(function(e){zn(t,e,Mn(n,e))}))}}return t}function Ja(t,e){var i=void 0!==ko&&Bi(t)||t["@@iterator"];if(!i){if(So(t)||(i=function(t,e){var i;if(!t)return;if("string"==typeof t)return th(t,e);var n=Oo(i=Object.prototype.toString.call(t)).call(i,8,-1);"Object"===n&&t.constructor&&(n=t.constructor.name);if("Map"===n||"Set"===n)return Ei(t);if("Arguments"===n||/^(?:Ui|I)nt(?:8|16|32)(?:Clamped)?Array$/.test(n))return th(t,e)}(t))||e&&t&&"number"==typeof t.length){i&&(t=i);var n=0,o=function(){};return{s:o,n:function(){return n>=t.length?{done:!0}:{done:!1,value:t[n++]}},e:function(t){throw t},f:o}}throw new TypeError("Invalid attempt to iterate non-iterable instance.\nIn order to be iterable, non-array objects must have a [Symbol.iterator]() method.")}var r,s=!0,a=!1;return{s:function(){i=i.call(t)},n:function(){var t=i.next();return s=t.done,t},e:function(t){a=!0,r=t},f:function(){try{s||null==i.return||i.return()}finally{if(a)throw r}}}}function th(t,e){(null==e||e>t.length)&&(e=t.length);for(var i=0,n=new Array(e);i<e;i++)n[i]=t[i];return n}var eh=ko("DELETE");function ih(t){for(var e,i=arguments.length,n=new Array(i>1?i-1:0),o=1;o<i;o++)n[o-1]=arguments[o];return nh.apply(void 0,Eo(e=[{},t]).call(e,n))}function nh(){var t=oh.apply(void 0,arguments);return sh(t),t}function oh(){for(var t=arguments.length,e=new Array(t),i=0;i<t;i++)e[i]=arguments[i];if(e.length<2)return e[0];var n;if(e.length>2)return oh.apply(void 0,Eo(n=[nh(e[0],e[1])]).call(n,wo(Oo(e).call(e,2))));var o,r=e[0],s=e[1],a=Ja(Co(s));try{for(a.s();!(o=a.n()).done;){var h=o.value;Object.prototype.propertyIsEnumerable.call(s,h)&&(s[h]===eh?delete r[h]:null===r[h]||null===s[h]||"object"!==go(r[h])||"object"!==go(s[h])||So(r[h])||So(s[h])?r[h]=rh(s[h]):r[h]=oh(r[h],s[h]))}}catch(t){a.e(t)}finally{a.f()}return r}function rh(t){return So(t)?Io(t).call(t,(function(t){return rh(t)})):"object"===go(t)&&null!==t?oh({},t):t}function sh(t){for(var e=0,i=zo(t);e<i.length;e++){var n=i[e];t[n]===eh?delete t[n]:"object"===go(t[n])&&null!==t[n]&&sh(t[n])}}function ah(){for(var t=arguments.length,e=new Array(t),i=0;i<t;i++)e[i]=arguments[i];return hh(e.length?e:[No()])}function hh(t){var e=function(){for(var t=lh(),e=t(" "),i=t(" "),n=t(" "),o=0;o<arguments.length;o++)(e-=t(o<0||arguments.length<=o?void 0:arguments[o]))<0&&(e+=1),(i-=t(o<0||arguments.length<=o?void 0:arguments[o]))<0&&(i+=1),(n-=t(o<0||arguments.length<=o?void 0:arguments[o]))<0&&(n+=1);return[e,i,n]}(t),i=uo(e,3),n=i[0],o=i[1],r=i[2],s=1,a=function(){var t=2091639*n+2.3283064365386963e-10*s;return n=o,o=r,r=t-(s=0|t)};return a.uint32=function(){return 4294967296*a()},a.fract53=function(){return a()+11102230246251565e-32*(2097152*a()|0)},a.algorithm="Alea",a.seed=t,a.version="0.9",a}function lh(){var t=4022871197;return function(e){for(var i=e.toString(),n=0;n<i.length;n++){var o=.02519603282416938*(t+=i.charCodeAt(n));o-=t=o>>>0,t=(o*=t)>>>0,t+=4294967296*(o-=t)}return 2.3283064365386963e-10*(t>>>0)}}var dh="undefined"!=typeof window?window.Hammer||$a:function(){return{on:t=function(){},off:t,destroy:t,emit:t,get:function(){return{set:t}}};var t};function ch(t){var e,i=this;this._cleanupQueue=[],this.active=!1,this._dom={container:t,overlay:document.createElement("div")},this._dom.overlay.classList.add("vis-overlay"),this._dom.container.appendChild(this._dom.overlay),this._cleanupQueue.push((function(){i._dom.overlay.parentNode.removeChild(i._dom.overlay)}));var n=dh(this._dom.overlay);n.on("tap",Vt(e=this._onTapOverlay).call(e,this)),this._cleanupQueue.push((function(){n.destroy()}));var o=["tap","doubletap","press","pinch","pan","panstart","panmove","panend"];Wo(o).call(o,(function(t){n.on(t,(function(t){t.srcEvent.stopPropagation()}))})),document&&document.body&&(this._onClick=function(e){(function(t,e){for(;t;){if(t===e)return!0;t=t.parentNode}return!1})(e.target,t)||i.deactivate()},document.body.addEventListener("click",this._onClick),this._cleanupQueue.push((function(){document.body.removeEventListener("click",i._onClick)}))),this._escListener=function(t){("key"in t?"Escape"===t.key:27===t.keyCode)&&i.deactivate()}}Zt(ch.prototype),ch.current=null,ch.prototype.destroy=function(){var t,e;this.deactivate();var i,n=Ja(Xo(t=er(e=this._cleanupQueue).call(e,0)).call(t));try{for(n.s();!(i=n.n()).done;){(0,i.value)()}}catch(t){n.e(t)}finally{n.f()}},ch.prototype.activate=function(){ch.current&&ch.current.deactivate(),ch.current=this,this.active=!0,this._dom.overlay.style.display="none",this._dom.container.classList.add("vis-active"),this.emit("change"),this.emit("activate"),document.body.addEventListener("keydown",this._escListener)},ch.prototype.deactivate=function(){this.active=!1,this._dom.overlay.style.display="block",this._dom.container.classList.remove("vis-active"),document.body.removeEventListener("keydown",this._escListener),this.emit("change"),this.emit("deactivate")},ch.prototype._onTapOverlay=function(t){this.activate(),t.srcEvent.stopPropagation()};var uh=/^\/?Date\((-?\d+)/i,fh=/^#?([a-f\d]{2})([a-f\d]{2})([a-f\d]{2})$/i,ph=/^#?([a-f\d])([a-f\d])([a-f\d])$/i,vh=/^rgb\( *(1?\d{1,2}|2[0-4]\d|25[0-5]) *, *(1?\d{1,2}|2[0-4]\d|25[0-5]) *, *(1?\d{1,2}|2[0-4]\d|25[0-5]) *\)$/i,gh=/^rgba\( *(1?\d{1,2}|2[0-4]\d|25[0-5]) *, *(1?\d{1,2}|2[0-4]\d|25[0-5]) *, *(1?\d{1,2}|2[0-4]\d|25[0-5]) *, *([01]|0?\.\d+) *\)$/i;function yh(t){return t instanceof Number||"number"==typeof t}function mh(t){if(t)for(;!0===t.hasChildNodes();){var e=t.firstChild;e&&(mh(e),t.removeChild(e))}}function bh(t){return t instanceof String||"string"==typeof t}function wh(t){return"object"===go(t)&&null!==t}function kh(t,e,i,n){var o=!1;!0===n&&(o=null===e[i]&&void 0!==t[i]),o?delete t[i]:t[i]=e[i]}function _h(t,e){var i=arguments.length>2&&void 0!==arguments[2]&&arguments[2];for(var n in t)if(void 0!==e[n])if(null===e[n]||"object"!==go(e[n]))kh(t,e,n,i);else{var o=t[n],r=e[n];wh(o)&&wh(r)&&_h(o,r,i)}}var xh=At;function Eh(t,e,i){var n=arguments.length>3&&void 0!==arguments[3]&&arguments[3];if(So(i))throw new TypeError("Arrays are not supported by deepExtend");for(var o=0;o<t.length;o++){var r=t[o];if(Object.prototype.hasOwnProperty.call(i,r))if(i[r]&&i[r].constructor===Object)void 0===e[r]&&(e[r]={}),e[r].constructor===Object?Ch(e[r],i[r],!1,n):kh(e,i,r,n);else{if(So(i[r]))throw new TypeError("Arrays are not supported by deepExtend");kh(e,i,r,n)}}return e}function Oh(t,e,i){var n=arguments.length>3&&void 0!==arguments[3]&&arguments[3];if(So(i))throw new TypeError("Arrays are not supported by deepExtend");for(var o in i)if(Object.prototype.hasOwnProperty.call(i,o)&&!dr(t).call(t,o))if(i[o]&&i[o].constructor===Object)void 0===e[o]&&(e[o]={}),e[o].constructor===Object?Ch(e[o],i[o]):kh(e,i,o,n);else if(So(i[o])){e[o]=[];for(var r=0;r<i[o].length;r++)e[o].push(i[o][r])}else kh(e,i,o,n);return e}function Ch(t,e){var i=arguments.length>2&&void 0!==arguments[2]&&arguments[2],n=arguments.length>3&&void 0!==arguments[3]&&arguments[3];for(var o in e)if(Object.prototype.hasOwnProperty.call(e,o)||!0===i)if("object"===go(e[o])&&null!==e[o]&&fr(e[o])===Object.prototype)void 0===t[o]?t[o]=Ch({},e[o],i):"object"===go(t[o])&&null!==t[o]&&fr(t[o])===Object.prototype?Ch(t[o],e[o],i):kh(t,e,o,n);else if(So(e[o])){var r;t[o]=Oo(r=e[o]).call(r)}else kh(t,e,o,n);return t}function Sh(t,e){var i;return Eo(i=[]).call(i,wo(t),[e])}function Th(t){return Oo(t).call(t)}function Mh(t){return t.getBoundingClientRect().left}function Ph(t){return t.getBoundingClientRect().top}function Dh(t,e){if(So(t))for(var i=t.length,n=0;n<i;n++)e(t[n],n,t);else for(var o in t)Object.prototype.hasOwnProperty.call(t,o)&&e(t[o],o,t)}var Ih=_r;function Bh(t,e,i,n){var o;t.addEventListener?(void 0===n&&(n=!1),"mousewheel"===e&&dr(o=navigator.userAgent).call(o,"Firefox")&&(e="DOMMouseScroll"),t.addEventListener(e,i,n)):t.attachEvent("on"+e,i)}function zh(t,e,i,n){var o;t.removeEventListener?(void 0===n&&(n=!1),"mousewheel"===e&&dr(o=navigator.userAgent).call(o,"Firefox")&&(e="DOMMouseScroll"),t.removeEventListener(e,i,n)):t.detachEvent("on"+e,i)}var Nh={asBoolean:function(t,e){return"function"==typeof t&&(t=t()),null!=t?0!=t:e||null},asNumber:function(t,e){return"function"==typeof t&&(t=t()),null!=t?Number(t)||e||null:e||null},asString:function(t,e){return"function"==typeof t&&(t=t()),null!=t?String(t):e||null},asSize:function(t,e){return"function"==typeof t&&(t=t()),bh(t)?t:yh(t)?t+"px":e||null},asElement:function(t,e){return"function"==typeof t&&(t=t()),t||e||null}};function Ah(t){var e;switch(t.length){case 3:case 4:return(e=ph.exec(t))?{r:Br(e[1]+e[1],16),g:Br(e[2]+e[2],16),b:Br(e[3]+e[3],16)}:null;case 6:case 7:return(e=fh.exec(t))?{r:Br(e[1],16),g:Br(e[2],16),b:Br(e[3],16)}:null;default:return null}}function Fh(t,e){if(dr(t).call(t,"rgba"))return t;if(dr(t).call(t,"rgb")){var i=t.substr(Hr(t).call(t,"(")+1).replace(")","").split(",");return"rgba("+i[0]+","+i[1]+","+i[2]+","+e+")"}var n=Ah(t);return null==n?t:"rgba("+n.r+","+n.g+","+n.b+","+e+")"}function jh(t,e,i){var n;return"#"+Oo(n=((1<<24)+(t<<16)+(e<<8)+i).toString(16)).call(n,1)}function Rh(t,e){if(bh(t)){var i=t;if(Xh(i)){var n,o=Io(n=i.substr(4).substr(0,i.length-5).split(",")).call(n,(function(t){return Br(t)}));i=jh(o[0],o[1],o[2])}if(!0===Yh(i)){var r=Uh(i),s={h:r.h,s:.8*r.s,v:Math.min(1,1.02*r.v)},a={h:r.h,s:Math.min(1,1.25*r.s),v:.8*r.v},h=Vh(a.h,a.s,a.v),l=Vh(s.h,s.s,s.v);return{background:i,border:h,highlight:{background:l,border:h},hover:{background:l,border:h}}}return{background:i,border:i,highlight:{background:i,border:i},hover:{background:i,border:i}}}return e?{background:t.background||e.background,border:t.border||e.border,highlight:bh(t.highlight)?{border:t.highlight,background:t.highlight}:{background:t.highlight&&t.highlight.background||e.highlight.background,border:t.highlight&&t.highlight.border||e.highlight.border},hover:bh(t.hover)?{border:t.hover,background:t.hover}:{border:t.hover&&t.hover.border||e.hover.border,background:t.hover&&t.hover.background||e.hover.background}}:{background:t.background||void 0,border:t.border||void 0,highlight:bh(t.highlight)?{border:t.highlight,background:t.highlight}:{background:t.highlight&&t.highlight.background||void 0,border:t.highlight&&t.highlight.border||void 0},hover:bh(t.hover)?{border:t.hover,background:t.hover}:{border:t.hover&&t.hover.border||void 0,background:t.hover&&t.hover.background||void 0}}}function Lh(t,e,i){t/=255,e/=255,i/=255;var n=Math.min(t,Math.min(e,i)),o=Math.max(t,Math.max(e,i));return n===o?{h:0,s:0,v:n}:{h:60*((t===n?3:i===n?1:5)-(t===n?e-i:i===n?t-e:i-t)/(o-n))/360,s:(o-n)/o,v:o}}var Hh=function(t){var e,i={};return Wo(e=t.split(";")).call(e,(function(t){if(""!=Ur(t).call(t)){var e,n,o=t.split(":"),r=Ur(e=o[0]).call(e),s=Ur(n=o[1]).call(n);i[r]=s}})),i},Wh=function(t){var e;return Io(e=zo(t)).call(e,(function(e){return e+": "+t[e]})).join("; ")};function qh(t,e,i){var n,o,r,s=Math.floor(6*t),a=6*t-s,h=i*(1-e),l=i*(1-a*e),d=i*(1-(1-a)*e);switch(s%6){case 0:n=i,o=d,r=h;break;case 1:n=l,o=i,r=h;break;case 2:n=h,o=i,r=d;break;case 3:n=h,o=l,r=i;break;case 4:n=d,o=h,r=i;break;case 5:n=i,o=h,r=l}return{r:Math.floor(255*n),g:Math.floor(255*o),b:Math.floor(255*r)}}function Vh(t,e,i){var n=qh(t,e,i);return jh(n.r,n.g,n.b)}function Uh(t){var e=Ah(t);if(!e)throw new TypeError("'".concat(t,"' is not a valid color."));return Lh(e.r,e.g,e.b)}function Yh(t){return/(^#[0-9A-F]{6}$)|(^#[0-9A-F]{3}$)/i.test(t)}function Xh(t){return vh.test(t)}function Gh(t){return gh.test(t)}function Kh(t){if(null===t||"object"!==go(t))return null;if(t instanceof Element)return t;var e=Gr(t);for(var i in t)Object.prototype.hasOwnProperty.call(t,i)&&"object"==go(t[i])&&(e[i]=Kh(t[i]));return e}function $h(t,e,i){var n=arguments.length>3&&void 0!==arguments[3]?arguments[3]:{},o=function(t){return null!=t},r=function(t){return null!==t&&"object"===go(t)},s=function(t){for(var e in t)if(Object.prototype.hasOwnProperty.call(t,e))return!1;return!0};if(!r(t))throw new Error("Parameter mergeTarget must be an object");if(!r(e))throw new Error("Parameter options must be an object");if(!o(i))throw new Error("Parameter option must have a value");if(!r(n))throw new Error("Parameter globalOptions must be an object");var a=function(t,e,i){r(t[i])||(t[i]={});var n=e[i],o=t[i];for(var s in n)Object.prototype.hasOwnProperty.call(n,s)&&(o[s]=n[s])},h=e[i],l=r(n)&&!s(n),d=l?n[i]:void 0,c=d?d.enabled:void 0;if(void 0!==h){if("boolean"==typeof h)return r(t[i])||(t[i]={}),void(t[i].enabled=h);if(null===h&&!r(t[i])){if(!o(d))return;t[i]=Gr(d)}if(r(h)){var u=!0;void 0!==h.enabled?u=h.enabled:void 0!==c&&(u=d.enabled),a(t,e,i),t[i].enabled=u}}}var Zh={linear:function(t){return t},easeInQuad:function(t){return t*t},easeOutQuad:function(t){return t*(2-t)},easeInOutQuad:function(t){return t<.5?2*t*t:(4-2*t)*t-1},easeInCubic:function(t){return t*t*t},easeOutCubic:function(t){return--t*t*t+1},easeInOutCubic:function(t){return t<.5?4*t*t*t:(t-1)*(2*t-2)*(2*t-2)+1},easeInQuart:function(t){return t*t*t*t},easeOutQuart:function(t){return 1- --t*t*t*t},easeInOutQuart:function(t){return t<.5?8*t*t*t*t:1-8*--t*t*t*t},easeInQuint:function(t){return t*t*t*t*t},easeOutQuint:function(t){return 1+--t*t*t*t*t},easeInOutQuint:function(t){return t<.5?16*t*t*t*t*t:1+16*--t*t*t*t*t}};function Qh(t,e){var i;So(e)||(e=[e]);var n,o=Ja(t);try{for(o.s();!(n=o.n()).done;){var r=n.value;if(r){i=r[e[0]];for(var s=1;s<e.length;s++)i&&(i=i[e[s]]);if(void 0!==i)break}}}catch(t){o.e(t)}finally{o.f()}return i}var Jh={black:"#000000",navy:"#000080",darkblue:"#00008B",mediumblue:"#0000CD",blue:"#0000FF",darkgreen:"#006400",green:"#008000",teal:"#008080",darkcyan:"#008B8B",deepskyblue:"#00BFFF",darkturquoise:"#00CED1",mediumspringgreen:"#00FA9A",lime:"#00FF00",springgreen:"#00FF7F",aqua:"#00FFFF",cyan:"#00FFFF",midnightblue:"#191970",dodgerblue:"#1E90FF",lightseagreen:"#20B2AA",forestgreen:"#228B22",seagreen:"#2E8B57",darkslategray:"#2F4F4F",limegreen:"#32CD32",mediumseagreen:"#3CB371",turquoise:"#40E0D0",royalblue:"#4169E1",steelblue:"#4682B4",darkslateblue:"#483D8B",mediumturquoise:"#48D1CC",indigo:"#4B0082",darkolivegreen:"#556B2F",cadetblue:"#5F9EA0",cornflowerblue:"#6495ED",mediumaquamarine:"#66CDAA",dimgray:"#696969",slateblue:"#6A5ACD",olivedrab:"#6B8E23",slategray:"#708090",lightslategray:"#778899",mediumslateblue:"#7B68EE",lawngreen:"#7CFC00",chartreuse:"#7FFF00",aquamarine:"#7FFFD4",maroon:"#800000",purple:"#800080",olive:"#808000",gray:"#808080",skyblue:"#87CEEB",lightskyblue:"#87CEFA",blueviolet:"#8A2BE2",darkred:"#8B0000",darkmagenta:"#8B008B",saddlebrown:"#8B4513",darkseagreen:"#8FBC8F",lightgreen:"#90EE90",mediumpurple:"#9370D8",darkviolet:"#9400D3",palegreen:"#98FB98",darkorchid:"#9932CC",yellowgreen:"#9ACD32",sienna:"#A0522D",brown:"#A52A2A",darkgray:"#A9A9A9",lightblue:"#ADD8E6",greenyellow:"#ADFF2F",paleturquoise:"#AFEEEE",lightsteelblue:"#B0C4DE",powderblue:"#B0E0E6",firebrick:"#B22222",darkgoldenrod:"#B8860B",mediumorchid:"#BA55D3",rosybrown:"#BC8F8F",darkkhaki:"#BDB76B",silver:"#C0C0C0",mediumvioletred:"#C71585",indianred:"#CD5C5C",peru:"#CD853F",chocolate:"#D2691E",tan:"#D2B48C",lightgrey:"#D3D3D3",palevioletred:"#D87093",thistle:"#D8BFD8",orchid:"#DA70D6",goldenrod:"#DAA520",crimson:"#DC143C",gainsboro:"#DCDCDC",plum:"#DDA0DD",burlywood:"#DEB887",lightcyan:"#E0FFFF",lavender:"#E6E6FA",darksalmon:"#E9967A",violet:"#EE82EE",palegoldenrod:"#EEE8AA",lightcoral:"#F08080",khaki:"#F0E68C",aliceblue:"#F0F8FF",honeydew:"#F0FFF0",azure:"#F0FFFF",sandybrown:"#F4A460",wheat:"#F5DEB3",beige:"#F5F5DC",whitesmoke:"#F5F5F5",mintcream:"#F5FFFA",ghostwhite:"#F8F8FF",salmon:"#FA8072",antiquewhite:"#FAEBD7",linen:"#FAF0E6",lightgoldenrodyellow:"#FAFAD2",oldlace:"#FDF5E6",red:"#FF0000",fuchsia:"#FF00FF",magenta:"#FF00FF",deeppink:"#FF1493",orangered:"#FF4500",tomato:"#FF6347",hotpink:"#FF69B4",coral:"#FF7F50",darkorange:"#FF8C00",lightsalmon:"#FFA07A",orange:"#FFA500",lightpink:"#FFB6C1",pink:"#FFC0CB",gold:"#FFD700",peachpuff:"#FFDAB9",navajowhite:"#FFDEAD",moccasin:"#FFE4B5",bisque:"#FFE4C4",mistyrose:"#FFE4E1",blanchedalmond:"#FFEBCD",papayawhip:"#FFEFD5",lavenderblush:"#FFF0F5",seashell:"#FFF5EE",cornsilk:"#FFF8DC",lemonchiffon:"#FFFACD",floralwhite:"#FFFAF0",snow:"#FFFAFA",yellow:"#FFFF00",lightyellow:"#FFFFE0",ivory:"#FFFFF0",white:"#FFFFFF"},tl=function(){function t(){var e=arguments.length>0&&void 0!==arguments[0]?arguments[0]:1;Nn(this,t),this.pixelRatio=e,this.generated=!1,this.centerCoordinates={x:144.5,y:144.5},this.r=289*.49,this.color={r:255,g:255,b:255,a:1},this.hueCircle=void 0,this.initialColor={r:255,g:255,b:255,a:1},this.previousColor=void 0,this.applied=!1,this.updateCallback=function(){},this.closeCallback=function(){},this._create()}return Fn(t,[{key:"insertTo",value:function(t){void 0!==this.hammer&&(this.hammer.destroy(),this.hammer=void 0),this.container=t,this.container.appendChild(this.frame),this._bindHammer(),this._setSize()}},{key:"setUpdateCallback",value:function(t){if("function"!=typeof t)throw new Error("Function attempted to set as colorPicker update callback is not a function.");this.updateCallback=t}},{key:"setCloseCallback",value:function(t){if("function"!=typeof t)throw new Error("Function attempted to set as colorPicker closing callback is not a function.");this.closeCallback=t}},{key:"_isColorString",value:function(t){if("string"==typeof t)return Jh[t]}},{key:"setColor",value:function(t){var e=!(arguments.length>1&&void 0!==arguments[1])||arguments[1];if("none"!==t){var i,n=this._isColorString(t);if(void 0!==n&&(t=n),!0===bh(t)){if(!0===Xh(t)){var o=t.substr(4).substr(0,t.length-5).split(",");i={r:o[0],g:o[1],b:o[2],a:1}}else if(!0===Gh(t)){var r=t.substr(5).substr(0,t.length-6).split(",");i={r:r[0],g:r[1],b:r[2],a:r[3]}}else if(!0===Yh(t)){var s=Ah(t);i={r:s.r,g:s.g,b:s.b,a:1}}}else if(t instanceof Object&&void 0!==t.r&&void 0!==t.g&&void 0!==t.b){var a=void 0!==t.a?t.a:"1.0";i={r:t.r,g:t.g,b:t.b,a:a}}if(void 0===i)throw new Error("Unknown color passed to the colorPicker. Supported are strings: rgb, hex, rgba. Object: rgb ({r:r,g:g,b:b,[a:a]}). Supplied: "+es(t));this._setColor(i,e)}}},{key:"show",value:function(){void 0!==this.closeCallback&&(this.closeCallback(),this.closeCallback=void 0),this.applied=!1,this.frame.style.display="block",this._generateHueCircle()}},{key:"_hide",value:function(){var t=this,e=!(arguments.length>0&&void 0!==arguments[0])||arguments[0];!0===e&&(this.previousColor=At({},this.color)),!0===this.applied&&this.updateCallback(this.initialColor),this.frame.style.display="none",rs((function(){void 0!==t.closeCallback&&(t.closeCallback(),t.closeCallback=void 0)}),0)}},{key:"_save",value:function(){this.updateCallback(this.color),this.applied=!1,this._hide()}},{key:"_apply",value:function(){this.applied=!0,this.updateCallback(this.color),this._updatePicker(this.color)}},{key:"_loadLast",value:function(){void 0!==this.previousColor?this.setColor(this.previousColor,!1):alert("There is no last color to load...")}},{key:"_setColor",value:function(t){var e=!(arguments.length>1&&void 0!==arguments[1])||arguments[1];!0===e&&(this.initialColor=At({},t)),this.color=t;var i=Lh(t.r,t.g,t.b),n=2*Math.PI,o=this.r*i.s,r=this.centerCoordinates.x+o*Math.sin(n*i.h),s=this.centerCoordinates.y+o*Math.cos(n*i.h);this.colorPickerSelector.style.left=r-.5*this.colorPickerSelector.clientWidth+"px",this.colorPickerSelector.style.top=s-.5*this.colorPickerSelector.clientHeight+"px",this._updatePicker(t)}},{key:"_setOpacity",value:function(t){this.color.a=t/100,this._updatePicker(this.color)}},{key:"_setBrightness",value:function(t){var e=Lh(this.color.r,this.color.g,this.color.b);e.v=t/100;var i=qh(e.h,e.s,e.v);i.a=this.color.a,this.color=i,this._updatePicker()}},{key:"_updatePicker",value:function(){var t=arguments.length>0&&void 0!==arguments[0]?arguments[0]:this.color,e=Lh(t.r,t.g,t.b),i=this.colorPickerCanvas.getContext("2d");void 0===this.pixelRation&&(this.pixelRatio=(window.devicePixelRatio||1)/(i.webkitBackingStorePixelRatio||i.mozBackingStorePixelRatio||i.msBackingStorePixelRatio||i.oBackingStorePixelRatio||i.backingStorePixelRatio||1)),i.setTransform(this.pixelRatio,0,0,this.pixelRatio,0,0);var n=this.colorPickerCanvas.clientWidth,o=this.colorPickerCanvas.clientHeight;i.clearRect(0,0,n,o),i.putImageData(this.hueCircle,0,0),i.fillStyle="rgba(0,0,0,"+(1-e.v)+")",hs(i).call(i),this.brightnessRange.value=100*e.v,this.opacityRange.value=100*t.a,this.initialColorDiv.style.backgroundColor="rgba("+this.initialColor.r+","+this.initialColor.g+","+this.initialColor.b+","+this.initialColor.a+")",this.newColorDiv.style.backgroundColor="rgba("+this.color.r+","+this.color.g+","+this.color.b+","+this.color.a+")"}},{key:"_setSize",value:function(){this.colorPickerCanvas.style.width="100%",this.colorPickerCanvas.style.height="100%",this.colorPickerCanvas.width=289*this.pixelRatio,this.colorPickerCanvas.height=289*this.pixelRatio}},{key:"_create",value:function(){var t,e,i,n;if(this.frame=document.createElement("div"),this.frame.className="vis-color-picker",this.colorPickerDiv=document.createElement("div"),this.colorPickerSelector=document.createElement("div"),this.colorPickerSelector.className="vis-selector",this.colorPickerDiv.appendChild(this.colorPickerSelector),this.colorPickerCanvas=document.createElement("canvas"),this.colorPickerDiv.appendChild(this.colorPickerCanvas),this.colorPickerCanvas.getContext){var o=this.colorPickerCanvas.getContext("2d");this.pixelRatio=(window.devicePixelRatio||1)/(o.webkitBackingStorePixelRatio||o.mozBackingStorePixelRatio||o.msBackingStorePixelRatio||o.oBackingStorePixelRatio||o.backingStorePixelRatio||1),this.colorPickerCanvas.getContext("2d").setTransform(this.pixelRatio,0,0,this.pixelRatio,0,0)}else{var r=document.createElement("DIV");r.style.color="red",r.style.fontWeight="bold",r.style.padding="10px",r.innerText="Error: your browser does not support HTML canvas",this.colorPickerCanvas.appendChild(r)}this.colorPickerDiv.className="vis-color",this.opacityDiv=document.createElement("div"),this.opacityDiv.className="vis-opacity",this.brightnessDiv=document.createElement("div"),this.brightnessDiv.className="vis-brightness",this.arrowDiv=document.createElement("div"),this.arrowDiv.className="vis-arrow",this.opacityRange=document.createElement("input");try{this.opacityRange.type="range",this.opacityRange.min="0",this.opacityRange.max="100"}catch(t){}this.opacityRange.value="100",this.opacityRange.className="vis-range",this.brightnessRange=document.createElement("input");try{this.brightnessRange.type="range",this.brightnessRange.min="0",this.brightnessRange.max="100"}catch(t){}this.brightnessRange.value="100",this.brightnessRange.className="vis-range",this.opacityDiv.appendChild(this.opacityRange),this.brightnessDiv.appendChild(this.brightnessRange);var s=this;this.opacityRange.onchange=function(){s._setOpacity(this.value)},this.opacityRange.oninput=function(){s._setOpacity(this.value)},this.brightnessRange.onchange=function(){s._setBrightness(this.value)},this.brightnessRange.oninput=function(){s._setBrightness(this.value)},this.brightnessLabel=document.createElement("div"),this.brightnessLabel.className="vis-label vis-brightness",this.brightnessLabel.innerText="brightness:",this.opacityLabel=document.createElement("div"),this.opacityLabel.className="vis-label vis-opacity",this.opacityLabel.innerText="opacity:",this.newColorDiv=document.createElement("div"),this.newColorDiv.className="vis-new-color",this.newColorDiv.innerText="new",this.initialColorDiv=document.createElement("div"),this.initialColorDiv.className="vis-initial-color",this.initialColorDiv.innerText="initial",this.cancelButton=document.createElement("div"),this.cancelButton.className="vis-button vis-cancel",this.cancelButton.innerText="cancel",this.cancelButton.onclick=Vt(t=this._hide).call(t,this,!1),this.applyButton=document.createElement("div"),this.applyButton.className="vis-button vis-apply",this.applyButton.innerText="apply",this.applyButton.onclick=Vt(e=this._apply).call(e,this),this.saveButton=document.createElement("div"),this.saveButton.className="vis-button vis-save",this.saveButton.innerText="save",this.saveButton.onclick=Vt(i=this._save).call(i,this),this.loadButton=document.createElement("div"),this.loadButton.className="vis-button vis-load",this.loadButton.innerText="load last",this.loadButton.onclick=Vt(n=this._loadLast).call(n,this),this.frame.appendChild(this.colorPickerDiv),this.frame.appendChild(this.arrowDiv),this.frame.appendChild(this.brightnessLabel),this.frame.appendChild(this.brightnessDiv),this.frame.appendChild(this.opacityLabel),this.frame.appendChild(this.opacityDiv),this.frame.appendChild(this.newColorDiv),this.frame.appendChild(this.initialColorDiv),this.frame.appendChild(this.cancelButton),this.frame.appendChild(this.applyButton),this.frame.appendChild(this.saveButton),this.frame.appendChild(this.loadButton)}},{key:"_bindHammer",value:function(){var t=this;this.drag={},this.pinch={},this.hammer=new dh(this.colorPickerCanvas),this.hammer.get("pinch").set({enable:!0}),this.hammer.on("hammer.input",(function(e){e.isFirst&&t._moveSelector(e)})),this.hammer.on("tap",(function(e){t._moveSelector(e)})),this.hammer.on("panstart",(function(e){t._moveSelector(e)})),this.hammer.on("panmove",(function(e){t._moveSelector(e)})),this.hammer.on("panend",(function(e){t._moveSelector(e)}))}},{key:"_generateHueCircle",value:function(){if(!1===this.generated){var t=this.colorPickerCanvas.getContext("2d");void 0===this.pixelRation&&(this.pixelRatio=(window.devicePixelRatio||1)/(t.webkitBackingStorePixelRatio||t.mozBackingStorePixelRatio||t.msBackingStorePixelRatio||t.oBackingStorePixelRatio||t.backingStorePixelRatio||1)),t.setTransform(this.pixelRatio,0,0,this.pixelRatio,0,0);var e,i,n,o,r=this.colorPickerCanvas.clientWidth,s=this.colorPickerCanvas.clientHeight;t.clearRect(0,0,r,s),this.centerCoordinates={x:.5*r,y:.5*s},this.r=.49*r;var a,h=2*Math.PI/360,l=1/this.r;for(n=0;n<360;n++)for(o=0;o<this.r;o++)e=this.centerCoordinates.x+o*Math.sin(h*n),i=this.centerCoordinates.y+o*Math.cos(h*n),a=qh(.002777777777777778*n,o*l,1),t.fillStyle="rgb("+a.r+","+a.g+","+a.b+")",t.fillRect(e-.5,i-.5,2,2);t.strokeStyle="rgba(0,0,0,1)",t.stroke(),this.hueCircle=t.getImageData(0,0,r,s)}this.generated=!0}},{key:"_moveSelector",value:function(t){var e=this.colorPickerDiv.getBoundingClientRect(),i=t.center.x-e.left,n=t.center.y-e.top,o=.5*this.colorPickerDiv.clientHeight,r=.5*this.colorPickerDiv.clientWidth,s=i-r,a=n-o,h=Math.atan2(s,a),l=.98*Math.min(Math.sqrt(s*s+a*a),r),d=Math.cos(h)*l+o,c=Math.sin(h)*l+r;this.colorPickerSelector.style.top=d-.5*this.colorPickerSelector.clientHeight+"px",this.colorPickerSelector.style.left=c-.5*this.colorPickerSelector.clientWidth+"px";var u=h/(2*Math.PI);u=u<0?u+1:u;var f=l/this.r,p=Lh(this.color.r,this.color.g,this.color.b);p.h=u,p.s=f;var v=qh(p.h,p.s,p.v);v.a=this.color.a,this.color=v,this.initialColorDiv.style.backgroundColor="rgba("+this.initialColor.r+","+this.initialColor.g+","+this.initialColor.b+","+this.initialColor.a+")",this.newColorDiv.style.backgroundColor="rgba("+this.color.r+","+this.color.g+","+this.color.b+","+this.color.a+")"}}]),t}();function el(){for(var t=arguments.length,e=new Array(t),i=0;i<t;i++)e[i]=arguments[i];if(e.length<1)throw new TypeError("Invalid arguments.");if(1===e.length)return document.createTextNode(e[0]);var n=document.createElement(e[0]);return n.appendChild(el.apply(void 0,wo(Oo(e).call(e,1)))),n}var il,nl=function(){function t(e,i,n){var o=arguments.length>3&&void 0!==arguments[3]?arguments[3]:1,r=arguments.length>4&&void 0!==arguments[4]?arguments[4]:function(){return!1};Nn(this,t),this.parent=e,this.changedOptions=[],this.container=i,this.allowCreation=!1,this.hideOption=r,this.options={},this.initialized=!1,this.popupCounter=0,this.defaultOptions={enabled:!1,filter:!0,container:void 0,showButton:!0},At(this.options,this.defaultOptions),this.configureOptions=n,this.moduleOptions={},this.domElements=[],this.popupDiv={},this.popupLimit=5,this.popupHistory={},this.colorPicker=new tl(o),this.wrapper=void 0}return Fn(t,[{key:"setOptions",value:function(t){if(void 0!==t){this.popupHistory={},this._removePopup();var e=!0;if("string"==typeof t)this.options.filter=t;else if(So(t))this.options.filter=t.join();else if("object"===go(t)){if(null==t)throw new TypeError("options cannot be null");void 0!==t.container&&(this.options.container=t.container),void 0!==mr(t)&&(this.options.filter=mr(t)),void 0!==t.showButton&&(this.options.showButton=t.showButton),void 0!==t.enabled&&(e=t.enabled)}else"boolean"==typeof t?(this.options.filter=!0,e=t):"function"==typeof t&&(this.options.filter=t,e=!0);!1===mr(this.options)&&(e=!1),this.options.enabled=e}this._clean()}},{key:"setModuleOptions",value:function(t){this.moduleOptions=t,!0===this.options.enabled&&(this._clean(),void 0!==this.options.container&&(this.container=this.options.container),this._create())}},{key:"_create",value:function(){this._clean(),this.changedOptions=[];var t=mr(this.options),e=0,i=!1;for(var n in this.configureOptions)Object.prototype.hasOwnProperty.call(this.configureOptions,n)&&(this.allowCreation=!1,i=!1,"function"==typeof t?i=(i=t(n,[]))||this._handleObject(this.configureOptions[n],[n],!0):!0!==t&&-1===Hr(t).call(t,n)||(i=!0),!1!==i&&(this.allowCreation=!0,e>0&&this._makeItem([]),this._makeHeader(n),this._handleObject(this.configureOptions[n],[n])),e++);this._makeButton(),this._push()}},{key:"_push",value:function(){this.wrapper=document.createElement("div"),this.wrapper.className="vis-configuration-wrapper",this.container.appendChild(this.wrapper);for(var t=0;t<this.domElements.length;t++)this.wrapper.appendChild(this.domElements[t]);this._showPopupIfNeeded()}},{key:"_clean",value:function(){for(var t=0;t<this.domElements.length;t++)this.wrapper.removeChild(this.domElements[t]);void 0!==this.wrapper&&(this.container.removeChild(this.wrapper),this.wrapper=void 0),this.domElements=[],this._removePopup()}},{key:"_getValue",value:function(t){for(var e=this.moduleOptions,i=0;i<t.length;i++){if(void 0===e[t[i]]){e=void 0;break}e=e[t[i]]}return e}},{key:"_makeItem",value:function(t){if(!0===this.allowCreation){var e=document.createElement("div");e.className="vis-configuration vis-config-item vis-config-s"+t.length;for(var i=arguments.length,n=new Array(i>1?i-1:0),o=1;o<i;o++)n[o-1]=arguments[o];return Wo(n).call(n,(function(t){e.appendChild(t)})),this.domElements.push(e),this.domElements.length}return 0}},{key:"_makeHeader",value:function(t){var e=document.createElement("div");e.className="vis-configuration vis-config-header",e.innerText=t,this._makeItem([],e)}},{key:"_makeLabel",value:function(t,e){var i=arguments.length>2&&void 0!==arguments[2]&&arguments[2],n=document.createElement("div");if(n.className="vis-configuration vis-config-label vis-config-s"+e.length,!0===i){for(;n.firstChild;)n.removeChild(n.firstChild);n.appendChild(el("i","b",t))}else n.innerText=t+":";return n}},{key:"_makeDropdown",value:function(t,e,i){var n=document.createElement("select");n.className="vis-configuration vis-config-select";var o=0;void 0!==e&&-1!==Hr(t).call(t,e)&&(o=Hr(t).call(t,e));for(var r=0;r<t.length;r++){var s=document.createElement("option");s.value=t[r],r===o&&(s.selected="selected"),s.innerText=t[r],n.appendChild(s)}var a=this;n.onchange=function(){a._update(this.value,i)};var h=this._makeLabel(i[i.length-1],i);this._makeItem(i,h,n)}},{key:"_makeRange",value:function(t,e,i){var n=t[0],o=t[1],r=t[2],s=t[3],a=document.createElement("input");a.className="vis-configuration vis-config-range";try{a.type="range",a.min=o,a.max=r}catch(t){}a.step=s;var h="",l=0;if(void 0!==e){var d=1.2;e<0&&e*d<o?(a.min=Math.ceil(e*d),l=a.min,h="range increased"):e/d<o&&(a.min=Math.ceil(e/d),l=a.min,h="range increased"),e*d>r&&1!==r&&(a.max=Math.ceil(e*d),l=a.max,h="range increased"),a.value=e}else a.value=n;var c=document.createElement("input");c.className="vis-configuration vis-config-rangeinput",c.value=a.value;var u=this;a.onchange=function(){c.value=this.value,u._update(Number(this.value),i)},a.oninput=function(){c.value=this.value};var f=this._makeLabel(i[i.length-1],i),p=this._makeItem(i,f,a,c);""!==h&&this.popupHistory[p]!==l&&(this.popupHistory[p]=l,this._setupPopup(h,p))}},{key:"_makeButton",value:function(){var t=this;if(!0===this.options.showButton){var e=document.createElement("div");e.className="vis-configuration vis-config-button",e.innerText="generate options",e.onclick=function(){t._printOptions()},e.onmouseover=function(){e.className="vis-configuration vis-config-button hover"},e.onmouseout=function(){e.className="vis-configuration vis-config-button"},this.optionsContainer=document.createElement("div"),this.optionsContainer.className="vis-configuration vis-config-option-container",this.domElements.push(this.optionsContainer),this.domElements.push(e)}}},{key:"_setupPopup",value:function(t,e){var i=this;if(!0===this.initialized&&!0===this.allowCreation&&this.popupCounter<this.popupLimit){var n=document.createElement("div");n.id="vis-configuration-popup",n.className="vis-configuration-popup",n.innerText=t,n.onclick=function(){i._removePopup()},this.popupCounter+=1,this.popupDiv={html:n,index:e}}}},{key:"_removePopup",value:function(){void 0!==this.popupDiv.html&&(this.popupDiv.html.parentNode.removeChild(this.popupDiv.html),clearTimeout(this.popupDiv.hideTimeout),clearTimeout(this.popupDiv.deleteTimeout),this.popupDiv={})}},{key:"_showPopupIfNeeded",value:function(){var t=this;if(void 0!==this.popupDiv.html){var e=this.domElements[this.popupDiv.index].getBoundingClientRect();this.popupDiv.html.style.left=e.left+"px",this.popupDiv.html.style.top=e.top-30+"px",document.body.appendChild(this.popupDiv.html),this.popupDiv.hideTimeout=rs((function(){t.popupDiv.html.style.opacity=0}),1500),this.popupDiv.deleteTimeout=rs((function(){t._removePopup()}),1800)}}},{key:"_makeCheckbox",value:function(t,e,i){var n=document.createElement("input");n.type="checkbox",n.className="vis-configuration vis-config-checkbox",n.checked=t,void 0!==e&&(n.checked=e,e!==t&&("object"===go(t)?e!==t.enabled&&this.changedOptions.push({path:i,value:e}):this.changedOptions.push({path:i,value:e})));var o=this;n.onchange=function(){o._update(this.checked,i)};var r=this._makeLabel(i[i.length-1],i);this._makeItem(i,r,n)}},{key:"_makeTextInput",value:function(t,e,i){var n=document.createElement("input");n.type="text",n.className="vis-configuration vis-config-text",n.value=e,e!==t&&this.changedOptions.push({path:i,value:e});var o=this;n.onchange=function(){o._update(this.value,i)};var r=this._makeLabel(i[i.length-1],i);this._makeItem(i,r,n)}},{key:"_makeColorField",value:function(t,e,i){var n=this,o=t[1],r=document.createElement("div");"none"!==(e=void 0===e?o:e)?(r.className="vis-configuration vis-config-colorBlock",r.style.backgroundColor=e):r.className="vis-configuration vis-config-colorBlock none",e=void 0===e?o:e,r.onclick=function(){n._showColorPicker(e,r,i)};var s=this._makeLabel(i[i.length-1],i);this._makeItem(i,s,r)}},{key:"_showColorPicker",value:function(t,e,i){var n=this;e.onclick=function(){},this.colorPicker.insertTo(e),this.colorPicker.show(),this.colorPicker.setColor(t),this.colorPicker.setUpdateCallback((function(t){var o="rgba("+t.r+","+t.g+","+t.b+","+t.a+")";e.style.backgroundColor=o,n._update(o,i)})),this.colorPicker.setCloseCallback((function(){e.onclick=function(){n._showColorPicker(t,e,i)}}))}},{key:"_handleObject",value:function(t){var e=arguments.length>1&&void 0!==arguments[1]?arguments[1]:[],i=arguments.length>2&&void 0!==arguments[2]&&arguments[2],n=!1,o=mr(this.options),r=!1;for(var s in t)if(Object.prototype.hasOwnProperty.call(t,s)){n=!0;var a=t[s],h=Sh(e,s);if("function"==typeof o&&!1===(n=o(s,e))&&!So(a)&&"string"!=typeof a&&"boolean"!=typeof a&&a instanceof Object&&(this.allowCreation=!1,n=this._handleObject(a,h,!0),this.allowCreation=!1===i),!1!==n){r=!0;var l=this._getValue(h);if(So(a))this._handleArray(a,l,h);else if("string"==typeof a)this._makeTextInput(a,l,h);else if("boolean"==typeof a)this._makeCheckbox(a,l,h);else if(a instanceof Object){if(!this.hideOption(e,s,this.moduleOptions))if(void 0!==a.enabled){var d=Sh(h,"enabled"),c=this._getValue(d);if(!0===c){var u=this._makeLabel(s,h,!0);this._makeItem(h,u),r=this._handleObject(a,h)||r}else this._makeCheckbox(a,c,h)}else{var f=this._makeLabel(s,h,!0);this._makeItem(h,f),r=this._handleObject(a,h)||r}}else console.error("dont know how to handle",a,s,h)}}return r}},{key:"_handleArray",value:function(t,e,i){"string"==typeof t[0]&&"color"===t[0]?(this._makeColorField(t,e,i),t[1]!==e&&this.changedOptions.push({path:i,value:e})):"string"==typeof t[0]?(this._makeDropdown(t,e,i),t[0]!==e&&this.changedOptions.push({path:i,value:e})):"number"==typeof t[0]&&(this._makeRange(t,e,i),t[0]!==e&&this.changedOptions.push({path:i,value:Number(e)}))}},{key:"_update",value:function(t,e){var i=this._constructOptions(t,e);this.parent.body&&this.parent.body.emitter&&this.parent.body.emitter.emit&&this.parent.body.emitter.emit("configChange",i),this.initialized=!0,this.parent.setOptions(i)}},{key:"_constructOptions",value:function(t,e){var i=arguments.length>2&&void 0!==arguments[2]?arguments[2]:{},n=i;t="false"!==(t="true"===t||t)&&t;for(var o=0;o<e.length;o++)"global"!==e[o]&&(void 0===n[e[o]]&&(n[e[o]]={}),o!==e.length-1?n=n[e[o]]:n[e[o]]=t);return i}},{key:"_printOptions",value:function(){for(var t=this.getOptions();this.optionsContainer.firstChild;)this.optionsContainer.removeChild(this.optionsContainer.firstChild);this.optionsContainer.appendChild(el("pre","const options = "+es(t,null,2)))}},{key:"getOptions",value:function(){for(var t={},e=0;e<this.changedOptions.length;e++)this._constructOptions(this.changedOptions[e].value,this.changedOptions[e].path,t);return t}}]),t}(),ol=!1,rl="background: #FFeeee; color: #dd0000",sl=ch,al=tl,hl=nl,ll=dh,dl=function(){function t(e,i){Nn(this,t),this.container=e,this.overflowMethod=i||"cap",this.x=0,this.y=0,this.padding=5,this.hidden=!1,this.frame=document.createElement("div"),this.frame.className="vis-tooltip",this.container.appendChild(this.frame)}return Fn(t,[{key:"setPosition",value:function(t,e){this.x=Br(t),this.y=Br(e)}},{key:"setText",value:function(t){if(t instanceof Element){for(;this.frame.firstChild;)this.frame.removeChild(this.frame.firstChild);this.frame.appendChild(t)}else this.frame.innerText=t}},{key:"show",value:function(t){if(void 0===t&&(t=!0),!0===t){var e=this.frame.clientHeight,i=this.frame.clientWidth,n=this.frame.parentNode.clientHeight,o=this.frame.parentNode.clientWidth,r=0,s=0;if("flip"==this.overflowMethod){var a=!1,h=!0;this.y-e<this.padding&&(h=!1),this.x+i>o-this.padding&&(a=!0),r=a?this.x-i:this.x,s=h?this.y-e:this.y}else(s=this.y-e)+e+this.padding>n&&(s=n-e-this.padding),s<this.padding&&(s=this.padding),(r=this.x)+i+this.padding>o&&(r=o-i-this.padding),r<this.padding&&(r=this.padding);this.frame.style.left=r+"px",this.frame.style.top=s+"px",this.frame.style.visibility="visible",this.hidden=!1}else this.hide()}},{key:"hide",value:function(){this.hidden=!0,this.frame.style.left="0",this.frame.style.top="0",this.frame.style.visibility="hidden"}},{key:"destroy",value:function(){this.frame.parentNode.removeChild(this.frame)}}]),t}(),cl=rl,ul=function(){function t(){Nn(this,t)}return Fn(t,null,[{key:"validate",value:function(e,i,n){ol=!1,il=i;var o=i;return void 0!==n&&(o=i[n]),t.parse(e,o,[]),ol}},{key:"parse",value:function(e,i,n){for(var o in e)Object.prototype.hasOwnProperty.call(e,o)&&t.check(o,e,i,n)}},{key:"check",value:function(e,i,n,o){if(void 0!==n[e]||void 0!==n.__any__){var r=e,s=!0;void 0===n[e]&&void 0!==n.__any__&&(r="__any__",s="object"===t.getType(i[e]));var a=n[r];s&&void 0!==a.__type__&&(a=a.__type__),t.checkFields(e,i,n,r,a,o)}else t.getSuggestion(e,n,o)}},{key:"checkFields",value:function(e,i,n,o,r,s){var a=function(i){console.error("%c"+i+t.printLocation(s,e),rl)},h=t.getType(i[e]),l=r[h];void 0!==l?"array"===t.getType(l)&&-1===Hr(l).call(l,i[e])?(a('Invalid option detected in "'+e+'". Allowed values are:'+t.print(l)+' not "'+i[e]+'". '),ol=!0):"object"===h&&"__any__"!==o&&(s=Sh(s,e),t.parse(i[e],n[o],s)):void 0===r.any&&(a('Invalid type received for "'+e+'". Expected: '+t.print(zo(r))+". Received ["+h+'] "'+i[e]+'"'),ol=!0)}},{key:"getType",value:function(t){var e=go(t);return"object"===e?null===t?"null":t instanceof Boolean?"boolean":t instanceof Number?"number":t instanceof String?"string":So(t)?"array":t instanceof Date?"date":void 0!==t.nodeType?"dom":!0===t._isAMomentObject?"moment":"object":"number"===e?"number":"boolean"===e?"boolean":"string"===e?"string":void 0===e?"undefined":e}},{key:"getSuggestion",value:function(e,i,n){var o,r=t.findInOptions(e,i,n,!1),s=t.findInOptions(e,il,[],!0);o=void 0!==r.indexMatch?" in "+t.printLocation(r.path,e,"")+'Perhaps it was incomplete? Did you mean: "'+r.indexMatch+'"?\n\n':s.distance<=4&&r.distance>s.distance?" in "+t.printLocation(r.path,e,"")+"Perhaps it was misplaced? Matching option found at: "+t.printLocation(s.path,s.closestMatch,""):r.distance<=8?'. Did you mean "'+r.closestMatch+'"?'+t.printLocation(r.path,e):". Did you mean one of these: "+t.print(zo(i))+t.printLocation(n,e),console.error('%cUnknown option detected: "'+e+'"'+o,rl),ol=!0}},{key:"findInOptions",value:function(e,i,n){var o=arguments.length>3&&void 0!==arguments[3]&&arguments[3],r=1e9,s="",a=[],h=e.toLowerCase(),l=void 0;for(var d in i){var c=void 0;if(void 0!==i[d].__type__&&!0===o){var u=t.findInOptions(e,i[d],Sh(n,d));r>u.distance&&(s=u.closestMatch,a=u.path,r=u.distance,l=u.indexMatch)}else{var f;-1!==Hr(f=d.toLowerCase()).call(f,h)&&(l=d),r>(c=t.levenshteinDistance(e,d))&&(s=d,a=Th(n),r=c)}}return{closestMatch:s,path:a,distance:r,indexMatch:l}}},{key:"printLocation",value:function(t,e){for(var i=arguments.length>2&&void 0!==arguments[2]?arguments[2]:"Problem value found at: \n",n="\n\n"+i+"options = {\n",o=0;o<t.length;o++){for(var r=0;r<o+1;r++)n+="  ";n+=t[o]+": {\n"}for(var s=0;s<t.length+1;s++)n+="  ";n+=e+"\n";for(var a=0;a<t.length+1;a++){for(var h=0;h<t.length-a;h++)n+="  ";n+="}\n"}return n+"\n\n"}},{key:"print",value:function(t){return es(t).replace(/(")|(\[)|(\])|(,"__type__")/g,"").replace(/(,)/g,", ")}},{key:"levenshteinDistance",value:function(t,e){if(0===t.length)return e.length;if(0===e.length)return t.length;var i,n,o=[];for(i=0;i<=e.length;i++)o[i]=[i];for(n=0;n<=t.length;n++)o[0][n]=n;for(i=1;i<=e.length;i++)for(n=1;n<=t.length;n++)e.charAt(i-1)==t.charAt(n-1)?o[i][n]=o[i-1][n-1]:o[i][n]=Math.min(o[i-1][n-1]+1,Math.min(o[i][n-1]+1,o[i-1][n]+1));return o[e.length][t.length]}}]),t}(),fl=Object.freeze({__proto__:null,Activator:sl,Alea:ah,ColorPicker:al,Configurator:hl,DELETE:eh,HSVToHex:Vh,HSVToRGB:qh,Hammer:ll,Popup:dl,RGBToHSV:Lh,RGBToHex:jh,VALIDATOR_PRINT_STYLE:cl,Validator:ul,addClassName:function(t,e){var i=t.className.split(" "),n=e.split(" ");i=Eo(i).call(i,mr(n).call(n,(function(t){return!dr(i).call(i,t)}))),t.className=i.join(" ")},addCssText:function(t,e){var i=Hh(t.style.cssText),n=Hh(e),o=Qa(Qa({},i),n);t.style.cssText=Wh(o)},addEventListener:Bh,binarySearchCustom:function(t,e,i,n){for(var o=0,r=0,s=t.length-1;r<=s&&o<1e4;){var a=Math.floor((r+s)/2),h=t[a],l=e(void 0===n?h[i]:h[i][n]);if(0==l)return a;-1==l?r=a+1:s=a-1,o++}return-1},binarySearchValue:function(t,e,i,n,o){var r,s,a,h,l=0,d=0,c=t.length-1;for(o=null!=o?o:function(t,e){return t==e?0:t<e?-1:1};d<=c&&l<1e4;){if(h=Math.floor(.5*(c+d)),r=t[Math.max(0,h-1)][i],s=t[h][i],a=t[Math.min(t.length-1,h+1)][i],0==o(s,e))return h;if(o(r,e)<0&&o(s,e)>0)return"before"==n?Math.max(0,h-1):h;if(o(s,e)<0&&o(a,e)>0)return"before"==n?h:Math.min(t.length-1,h+1);o(s,e)<0?d=h+1:c=h-1,l++}return-1},bridgeObject:Kh,copyAndExtendArray:Sh,copyArray:Th,deepExtend:Ch,deepObjectAssign:nh,easingFunctions:Zh,equalArray:function(t,e){if(t.length!==e.length)return!1;for(var i=0,n=t.length;i<n;i++)if(t[i]!=e[i])return!1;return!0},extend:xh,fillIfDefined:_h,forEach:Dh,getAbsoluteLeft:Mh,getAbsoluteRight:function(t){return t.getBoundingClientRect().right},getAbsoluteTop:Ph,getScrollBarWidth:function(){var t=document.createElement("p");t.style.width="100%",t.style.height="200px";var e=document.createElement("div");e.style.position="absolute",e.style.top="0px",e.style.left="0px",e.style.visibility="hidden",e.style.width="200px",e.style.height="150px",e.style.overflow="hidden",e.appendChild(t),document.body.appendChild(e);var i=t.offsetWidth;e.style.overflow="scroll";var n=t.offsetWidth;return i==n&&(n=e.clientWidth),document.body.removeChild(e),i-n},getTarget:function(){var t=arguments.length>0&&void 0!==arguments[0]?arguments[0]:window.event,e=null;return t&&(t.target?e=t.target:t.srcElement&&(e=t.srcElement)),e instanceof Element&&(null==e.nodeType||3!=e.nodeType||(e=e.parentNode)instanceof Element)?e:null},getType:function(t){var e=go(t);return"object"===e?null===t?"null":t instanceof Boolean?"Boolean":t instanceof Number?"Number":t instanceof String?"String":So(t)?"Array":t instanceof Date?"Date":"Object":"number"===e?"Number":"boolean"===e?"Boolean":"string"===e?"String":void 0===e?"undefined":e},hasParent:function(t,e){for(var i=t;i;){if(i===e)return!0;if(!i.parentNode)return!1;i=i.parentNode}return!1},hexToHSV:Uh,hexToRGB:Ah,insertSort:function(t,e){for(var i=0;i<t.length;i++){var n=t[i],o=void 0;for(o=i;o>0&&e(n,t[o-1])<0;o--)t[o]=t[o-1];t[o]=n}return t},isDate:function(t){if(t instanceof Date)return!0;if(bh(t)){if(uh.exec(t))return!0;if(!isNaN(Date.parse(t)))return!0}return!1},isNumber:yh,isObject:wh,isString:bh,isValidHex:Yh,isValidRGB:Xh,isValidRGBA:Gh,mergeOptions:$h,option:Nh,overrideOpacity:Fh,parseColor:Rh,preventDefault:function(t){t||(t=window.event),t&&(t.preventDefault?t.preventDefault():t.returnValue=!1)},pureDeepObjectAssign:ih,recursiveDOMDelete:mh,removeClassName:function(t,e){var i=t.className.split(" "),n=e.split(" ");i=mr(i).call(i,(function(t){return!dr(n).call(n,t)})),t.className=i.join(" ")},removeCssText:function(t,e){var i=Hh(t.style.cssText),n=Hh(e);for(var o in n)Object.prototype.hasOwnProperty.call(n,o)&&delete i[o];t.style.cssText=Wh(i)},removeEventListener:zh,selectiveBridgeObject:function(t,e){if(null!==e&&"object"===go(e)){for(var i=Gr(e),n=0;n<t.length;n++)Object.prototype.hasOwnProperty.call(e,t[n])&&"object"==go(e[t[n]])&&(i[t[n]]=Kh(e[t[n]]));return i}return null},selectiveDeepExtend:Eh,selectiveExtend:function(t,e){if(!So(t))throw new Error("Array with property names expected as first argument");for(var i=arguments.length,n=new Array(i>2?i-2:0),o=2;o<i;o++)n[o-2]=arguments[o];for(var r=0,s=n;r<s.length;r++)for(var a=s[r],h=0;h<t.length;h++){var l=t[h];a&&Object.prototype.hasOwnProperty.call(a,l)&&(e[l]=a[l])}return e},selectiveNotDeepExtend:Oh,throttle:function(t){var e=!1;return function(){e||(e=!0,requestAnimationFrame((function(){e=!1,t()})))}},toArray:Ih,topMost:Qh,updateProperty:function(t,e,i){return t[e]!==i&&(t[e]=i,!0)}});function pl(t){return _l=t,function(){var t={};xl=0,void(El=_l.charAt(0)),Nl(),"strict"===Ol&&(t.strict=!0,Nl());"graph"!==Ol&&"digraph"!==Ol||(t.type=Ol,Nl());Cl===bl&&(t.id=Ol,Nl());if("{"!=Ol)throw Hl("Angle bracket { expected");if(Nl(),Al(t),"}"!=Ol)throw Hl("Angle bracket } expected");if(Nl(),""!==Ol)throw Hl("End of file expected");return Nl(),delete t.node,delete t.edge,delete t.graph,t}()}var vl={fontsize:"font.size",fontcolor:"font.color",labelfontcolor:"font.color",fontname:"font.face",color:["color.border","color.background"],fillcolor:"color.background",tooltip:"title",labeltooltip:"title"},gl=Gr(vl);gl.color="color.color",gl.style="dashes";var yl=0,ml=1,bl=2,wl=3,kl={"{":!0,"}":!0,"[":!0,"]":!0,";":!0,"=":!0,",":!0,"->":!0,"--":!0},_l="",xl=0,El="",Ol="",Cl=yl;function Sl(){xl++,El=_l.charAt(xl)}function Tl(){return _l.charAt(xl+1)}function Ml(t){var e=t.charCodeAt(0);return e<47?35===e||46===e:e<59?e>47:e<91?e>64:e<96?95===e:e<123&&e>96}function Pl(t,e){if(t||(t={}),e)for(var i in e)e.hasOwnProperty(i)&&(t[i]=e[i]);return t}function Dl(t,e,i){for(var n=e.split("."),o=t;n.length;){var r=n.shift();n.length?(o[r]||(o[r]={}),o=o[r]):o[r]=i}}function Il(t,e){for(var i,n,o=null,r=[t],s=t;s.parent;)r.push(s.parent),s=s.parent;if(s.nodes)for(i=0,n=s.nodes.length;i<n;i++)if(e.id===s.nodes[i].id){o=s.nodes[i];break}for(o||(o={id:e.id},t.node&&(o.attr=Pl(o.attr,t.node))),i=r.length-1;i>=0;i--){var a,h=r[i];h.nodes||(h.nodes=[]),-1===Hr(a=h.nodes).call(a,o)&&h.nodes.push(o)}e.attr&&(o.attr=Pl(o.attr,e.attr))}function Bl(t,e){if(t.edges||(t.edges=[]),t.edges.push(e),t.edge){var i=Pl({},t.edge);e.attr=Pl(i,e.attr)}}function zl(t,e,i,n,o){var r={from:e,to:i,type:n};return t.edge&&(r.attr=Pl({},t.edge)),r.attr=Pl(r.attr||{},o),null!=o&&o.hasOwnProperty("arrows")&&null!=o.arrows&&(r.arrows={to:{enabled:!0,type:o.arrows.type}},o.arrows=null),r}function Nl(){for(Cl=yl,Ol="";" "===El||"\t"===El||"\n"===El||"\r"===El;)Sl();do{var t=!1;if("#"===El){for(var e=xl-1;" "===_l.charAt(e)||"\t"===_l.charAt(e);)e--;if("\n"===_l.charAt(e)||""===_l.charAt(e)){for(;""!=El&&"\n"!=El;)Sl();t=!0}}if("/"===El&&"/"===Tl()){for(;""!=El&&"\n"!=El;)Sl();t=!0}if("/"===El&&"*"===Tl()){for(;""!=El;){if("*"===El&&"/"===Tl()){Sl(),Sl();break}Sl()}t=!0}for(;" "===El||"\t"===El||"\n"===El||"\r"===El;)Sl()}while(t);if(""!==El){var i=El+Tl();if(kl[i])return Cl=ml,Ol=i,Sl(),void Sl();if(kl[El])return Cl=ml,Ol=El,void Sl();if(Ml(El)||"-"===El){for(Ol+=El,Sl();Ml(El);)Ol+=El,Sl();return"false"===Ol?Ol=!1:"true"===Ol?Ol=!0:isNaN(Number(Ol))||(Ol=Number(Ol)),void(Cl=bl)}if('"'===El){for(Sl();""!=El&&('"'!=El||'"'===El&&'"'===Tl());)'"'===El?(Ol+=El,Sl()):"\\"===El&&"n"===Tl()?(Ol+="\n",Sl()):Ol+=El,Sl();if('"'!=El)throw Hl('End of string " expected');return Sl(),void(Cl=bl)}for(Cl=wl;""!=El;)Ol+=El,Sl();throw new SyntaxError('Syntax error in part "'+Wl(Ol,30)+'"')}Cl=ml}function Al(t){for(;""!==Ol&&"}"!=Ol;)Fl(t),";"===Ol&&Nl()}function Fl(t){var e=jl(t);if(e)Rl(t,e);else if(!function(t){if("node"===Ol)return Nl(),t.node=Ll(),"node";if("edge"===Ol)return Nl(),t.edge=Ll(),"edge";if("graph"===Ol)return Nl(),t.graph=Ll(),"graph";return null}(t)){if(Cl!=bl)throw Hl("Identifier expected");var i=Ol;if(Nl(),"="===Ol){if(Nl(),Cl!=bl)throw Hl("Identifier expected");t[i]=Ol,Nl()}else!function(t,e){var i={id:e},n=Ll();n&&(i.attr=n);Il(t,i),Rl(t,e)}(t,i)}}function jl(t){var e=null;if("subgraph"===Ol&&((e={}).type="subgraph",Nl(),Cl===bl&&(e.id=Ol,Nl())),"{"===Ol){if(Nl(),e||(e={}),e.parent=t,e.node=t.node,e.edge=t.edge,e.graph=t.graph,Al(e),"}"!=Ol)throw Hl("Angle bracket } expected");Nl(),delete e.node,delete e.edge,delete e.graph,delete e.parent,t.subgraphs||(t.subgraphs=[]),t.subgraphs.push(e)}return e}function Rl(t,e){for(;"->"===Ol||"--"===Ol;){var i,n=Ol;Nl();var o=jl(t);if(o)i=o;else{if(Cl!=bl)throw Hl("Identifier or subgraph expected");Il(t,{id:i=Ol}),Nl()}Bl(t,zl(t,e,i,n,Ll())),e=i}}function Ll(){for(var t,e,i=null,n={dashed:!0,solid:!1,dotted:[1,5]},o={dot:"circle",box:"box",crow:"crow",curve:"curve",icurve:"inv_curve",normal:"triangle",inv:"inv_triangle",diamond:"diamond",tee:"bar",vee:"vee"},r=new Array,s=new Array;"["===Ol;){for(Nl(),i={};""!==Ol&&"]"!=Ol;){if(Cl!=bl)throw Hl("Attribute name expected");var a=Ol;if(Nl(),"="!=Ol)throw Hl("Equal sign = expected");if(Nl(),Cl!=bl)throw Hl("Attribute value expected");var h=Ol;"style"===a&&(h=n[h]),"arrowhead"===a&&(a="arrows",h={to:{enabled:!0,type:o[h]}}),"arrowtail"===a&&(a="arrows",h={from:{enabled:!0,type:o[h]}}),r.push({attr:i,name:a,value:h}),s.push(a),Nl(),","==Ol&&Nl()}if("]"!=Ol)throw Hl("Bracket ] expected");Nl()}if(dr(s).call(s,"dir")){var l={arrows:{}};for(t=0;t<r.length;t++)if("arrows"===r[t].name)if(null!=r[t].value.to)l.arrows.to=t;else{if(null==r[t].value.from)throw Hl("Invalid value of arrows");l.arrows.from=t}else"dir"===r[t].name&&(l.dir=t);var d,c,u=r[l.dir].value;if(!dr(s).call(s,"arrows"))if("both"===u)r.push({attr:r[l.dir].attr,name:"arrows",value:{to:{enabled:!0}}}),l.arrows.to=r.length-1,r.push({attr:r[l.dir].attr,name:"arrows",value:{from:{enabled:!0}}}),l.arrows.from=r.length-1;else if("forward"===u)r.push({attr:r[l.dir].attr,name:"arrows",value:{to:{enabled:!0}}}),l.arrows.to=r.length-1;else if("back"===u)r.push({attr:r[l.dir].attr,name:"arrows",value:{from:{enabled:!0}}}),l.arrows.from=r.length-1;else{if("none"!==u)throw Hl('Invalid dir type "'+u+'"');r.push({attr:r[l.dir].attr,name:"arrows",value:""}),l.arrows.to=r.length-1}if("both"===u)l.arrows.to&&l.arrows.from?(c=r[l.arrows.to].value.to.type,d=r[l.arrows.from].value.from.type,r[l.arrows.to]={attr:r[l.arrows.to].attr,name:r[l.arrows.to].name,value:{to:{enabled:!0,type:c},from:{enabled:!0,type:d}}},er(r).call(r,l.arrows.from,1)):l.arrows.to?(c=r[l.arrows.to].value.to.type,d="arrow",r[l.arrows.to]={attr:r[l.arrows.to].attr,name:r[l.arrows.to].name,value:{to:{enabled:!0,type:c},from:{enabled:!0,type:d}}}):l.arrows.from&&(c="arrow",d=r[l.arrows.from].value.from.type,r[l.arrows.from]={attr:r[l.arrows.from].attr,name:r[l.arrows.from].name,value:{to:{enabled:!0,type:c},from:{enabled:!0,type:d}}});else if("back"===u)l.arrows.to&&l.arrows.from?(c="",d=r[l.arrows.from].value.from.type,r[l.arrows.from]={attr:r[l.arrows.from].attr,name:r[l.arrows.from].name,value:{to:{enabled:!0,type:c},from:{enabled:!0,type:d}}}):l.arrows.to?(c="",d="arrow",l.arrows.from=l.arrows.to,r[l.arrows.from]={attr:r[l.arrows.from].attr,name:r[l.arrows.from].name,value:{to:{enabled:!0,type:c},from:{enabled:!0,type:d}}}):l.arrows.from&&(c="",d=r[l.arrows.from].value.from.type,r[l.arrows.to]={attr:r[l.arrows.from].attr,name:r[l.arrows.from].name,value:{to:{enabled:!0,type:c},from:{enabled:!0,type:d}}}),r[l.arrows.from]={attr:r[l.arrows.from].attr,name:r[l.arrows.from].name,value:{from:{enabled:!0,type:r[l.arrows.from].value.from.type}}};else if("none"===u){var f;r[f=l.arrows.to?l.arrows.to:l.arrows.from]={attr:r[f].attr,name:r[f].name,value:""}}else{if("forward"!==u)throw Hl('Invalid dir type "'+u+'"');l.arrows.to&&l.arrows.from||l.arrows.to?(c=r[l.arrows.to].value.to.type,d="",r[l.arrows.to]={attr:r[l.arrows.to].attr,name:r[l.arrows.to].name,value:{to:{enabled:!0,type:c},from:{enabled:!0,type:d}}}):l.arrows.from&&(c="arrow",d="",l.arrows.to=l.arrows.from,r[l.arrows.to]={attr:r[l.arrows.to].attr,name:r[l.arrows.to].name,value:{to:{enabled:!0,type:c},from:{enabled:!0,type:d}}}),r[l.arrows.to]={attr:r[l.arrows.to].attr,name:r[l.arrows.to].name,value:{to:{enabled:!0,type:r[l.arrows.to].value.to.type}}}}er(r).call(r,l.dir,1)}if(dr(s).call(s,"penwidth")){var p=[];for(e=r.length,t=0;t<e;t++)"width"!==r[t].name&&("penwidth"===r[t].name&&(r[t].name="width"),p.push(r[t]));r=p}for(e=r.length,t=0;t<e;t++)Dl(r[t].attr,r[t].name,r[t].value);return i}function Hl(t){return new SyntaxError(t+', got "'+Wl(Ol,30)+'" (char '+xl+")")}function Wl(t,e){return t.length<=e?t:t.substr(0,27)+"..."}function ql(t,e,i){for(var n=e.split("."),o=n.pop(),r=t,s=0;s<n.length;s++){var a=n[s];a in r||(r[a]={}),r=r[a]}return r[o]=i,t}function Vl(t,e){var i={};for(var n in t)if(t.hasOwnProperty(n)){var o=e[n];So(o)?Wo(o).call(o,(function(e){ql(i,e,t[n])})):ql(i,"string"==typeof o?o:n,t[n])}return i}function Ul(t){var e,i=pl(t),n={nodes:[],edges:[],options:{}};i.nodes&&Wo(e=i.nodes).call(e,(function(t){var e={id:t.id,label:String(t.label||t.id)};Pl(e,Vl(t.attr,vl)),e.image&&(e.shape="image"),n.nodes.push(e)}));if(i.edges){var o,r=function(t){var e={from:t.from,to:t.to};return Pl(e,Vl(t.attr,gl)),null==e.arrows&&"->"===t.type&&(e.arrows="to"),e};Wo(o=i.edges).call(o,(function(t){var e,i,o,s,a,h,l;(e=t.from instanceof Object?t.from.nodes:{id:t.from},i=t.to instanceof Object?t.to.nodes:{id:t.to},t.from instanceof Object&&t.from.edges)&&Wo(o=t.from.edges).call(o,(function(t){var e=r(t);n.edges.push(e)}));(a=i,h=function(e,i){var o=zl(n,e.id,i.id,t.type,t.attr),s=r(o);n.edges.push(s)},So(s=e)?Wo(s).call(s,(function(t){So(a)?Wo(a).call(a,(function(e){h(t,e)})):h(t,a)})):So(a)?Wo(a).call(a,(function(t){h(s,t)})):h(s,a),t.to instanceof Object&&t.to.edges)&&Wo(l=t.to.edges).call(l,(function(t){var e=r(t);n.edges.push(e)}))}))}return i.attr&&(n.options=i.attr),n}var Yl=Object.freeze({__proto__:null,parseDOT:pl,DOTToGraph:Ul});function Xl(t,e){var i,n={edges:{inheritColor:!1},nodes:{fixed:!1,parseColor:!1}};null!=e&&(null!=e.fixed&&(n.nodes.fixed=e.fixed),null!=e.parseColor&&(n.nodes.parseColor=e.parseColor),null!=e.inheritColor&&(n.edges.inheritColor=e.inheritColor));var o=t.edges,r=Io(o).call(o,(function(t){var e={from:t.source,id:t.id,to:t.target};return null!=t.attributes&&(e.attributes=t.attributes),null!=t.label&&(e.label=t.label),null!=t.attributes&&null!=t.attributes.title&&(e.title=t.attributes.title),"Directed"===t.type&&(e.arrows="to"),t.color&&!1===n.edges.inheritColor&&(e.color=t.color),e}));return{nodes:Io(i=t.nodes).call(i,(function(t){var e={id:t.id,fixed:n.nodes.fixed&&null!=t.x&&null!=t.y};return null!=t.attributes&&(e.attributes=t.attributes),null!=t.label&&(e.label=t.label),null!=t.size&&(e.size=t.size),null!=t.attributes&&null!=t.attributes.title&&(e.title=t.attributes.title),null!=t.title&&(e.title=t.title),null!=t.x&&(e.x=t.x),null!=t.y&&(e.y=t.y),null!=t.color&&(!0===n.nodes.parseColor?e.color=t.color:e.color={background:t.color,border:t.color,highlight:{background:t.color,border:t.color},hover:{background:t.color,border:t.color}}),e})),edges:r}}var Gl=Object.freeze({__proto__:null,parseGephi:Xl}),Kl=Object.freeze({__proto__:null,en:{addDescription:"Click in an empty space to place a new node.",addEdge:"Add Edge",addNode:"Add Node",back:"Back",close:"Close",createEdgeError:"Cannot link edges to a cluster.",del:"Delete selected",deleteClusterError:"Clusters cannot be deleted.",edgeDescription:"Click on a node and drag the edge to another node to connect them.",edit:"Edit",editClusterError:"Clusters cannot be edited.",editEdge:"Edit Edge",editEdgeDescription:"Click on the control points and drag them to a node to connect to it.",editNode:"Edit Node"},de:{addDescription:"Klicke auf eine freie Stelle, um einen neuen Knoten zu plazieren.",addEdge:"Kante hinzufügen",addNode:"Knoten hinzufügen",back:"Zurück",close:"Schließen",createEdgeError:"Es ist nicht möglich, Kanten mit Clustern zu verbinden.",del:"Lösche Auswahl",deleteClusterError:"Cluster können nicht gelöscht werden.",edgeDescription:"Klicke auf einen Knoten und ziehe die Kante zu einem anderen Knoten, um diese zu verbinden.",edit:"Editieren",editClusterError:"Cluster können nicht editiert werden.",editEdge:"Kante editieren",editEdgeDescription:"Klicke auf die Verbindungspunkte und ziehe diese auf einen Knoten, um sie zu verbinden.",editNode:"Knoten editieren"},es:{addDescription:"Haga clic en un lugar vacío para colocar un nuevo nodo.",addEdge:"Añadir arista",addNode:"Añadir nodo",back:"Atrás",close:"Cerrar",createEdgeError:"No se puede conectar una arista a un grupo.",del:"Eliminar selección",deleteClusterError:"No es posible eliminar grupos.",edgeDescription:"Haga clic en un nodo y arrastre la arista hacia otro nodo para conectarlos.",edit:"Editar",editClusterError:"No es posible editar grupos.",editEdge:"Editar arista",editEdgeDescription:"Haga clic en un punto de control y arrastrelo a un nodo para conectarlo.",editNode:"Editar nodo"},it:{addDescription:"Clicca per aggiungere un nuovo nodo",addEdge:"Aggiungi un vertice",addNode:"Aggiungi un nodo",back:"Indietro",close:"Chiudere",createEdgeError:"Non si possono collegare vertici ad un cluster",del:"Cancella la selezione",deleteClusterError:"I cluster non possono essere cancellati",edgeDescription:"Clicca su un nodo e trascinalo ad un altro nodo per connetterli.",edit:"Modifica",editClusterError:"I clusters non possono essere modificati.",editEdge:"Modifica il vertice",editEdgeDescription:"Clicca sui Punti di controllo e trascinali ad un nodo per connetterli.",editNode:"Modifica il nodo"},nl:{addDescription:"Klik op een leeg gebied om een nieuwe node te maken.",addEdge:"Link toevoegen",addNode:"Node toevoegen",back:"Terug",close:"Sluiten",createEdgeError:"Kan geen link maken naar een cluster.",del:"Selectie verwijderen",deleteClusterError:"Clusters kunnen niet worden verwijderd.",edgeDescription:"Klik op een node en sleep de link naar een andere node om ze te verbinden.",edit:"Wijzigen",editClusterError:"Clusters kunnen niet worden aangepast.",editEdge:"Link wijzigen",editEdgeDescription:"Klik op de verbindingspunten en sleep ze naar een node om daarmee te verbinden.",editNode:"Node wijzigen"},pt:{addDescription:"Clique em um espaço em branco para adicionar um novo nó",addEdge:"Adicionar aresta",addNode:"Adicionar nó",back:"Voltar",close:"Fechar",createEdgeError:"Não foi possível linkar arestas a um cluster.",del:"Remover selecionado",deleteClusterError:"Clusters não puderam ser removidos.",edgeDescription:"Clique em um nó e arraste a aresta até outro nó para conectá-los",edit:"Editar",editClusterError:"Clusters não puderam ser editados.",editEdge:"Editar aresta",editEdgeDescription:"Clique nos pontos de controle e os arraste para um nó para conectá-los",editNode:"Editar nó"},ru:{addDescription:"Кликните в свободное место, чтобы добавить новый узел.",addEdge:"Добавить ребро",addNode:"Добавить узел",back:"Назад",close:"Закрывать",createEdgeError:"Невозможно соединить ребра в кластер.",del:"Удалить выбранное",deleteClusterError:"Кластеры не могут быть удалены",edgeDescription:"Кликните на узел и протяните ребро к другому узлу, чтобы соединить их.",edit:"Редактировать",editClusterError:"Кластеры недоступны для редактирования.",editEdge:"Редактировать ребро",editEdgeDescription:"Кликните на контрольные точки и перетащите их в узел, чтобы подключиться к нему.",editNode:"Редактировать узел"},cn:{addDescription:"单击空白处放置新节点。",addEdge:"添加连接线",addNode:"添加节点",back:"返回",close:"關閉",createEdgeError:"无法将连接线连接到群集。",del:"删除选定",deleteClusterError:"无法删除群集。",edgeDescription:"单击某个节点并将该连接线拖动到另一个节点以连接它们。",edit:"编辑",editClusterError:"无法编辑群集。",editEdge:"编辑连接线",editEdgeDescription:"单击控制节点并将它们拖到节点上连接。",editNode:"编辑节点"},uk:{addDescription:"Kлікніть на вільне місце, щоб додати новий вузол.",addEdge:"Додати край",addNode:"Додати вузол",back:"Назад",close:"Закрити",createEdgeError:"Не можливо об'єднати краї в групу.",del:"Видалити обране",deleteClusterError:"Групи не можуть бути видалені.",edgeDescription:"Клікніть на вузол і перетягніть край до іншого вузла, щоб їх з'єднати.",edit:"Редагувати",editClusterError:"Групи недоступні для редагування.",editEdge:"Редагувати край",editEdgeDescription:"Клікніть на контрольні точки і перетягніть їх у вузол, щоб підключитися до нього.",editNode:"Редагувати вузол"},fr:{addDescription:"Cliquez dans un endroit vide pour placer un nœud.",addEdge:"Ajouter un lien",addNode:"Ajouter un nœud",back:"Retour",close:"Fermer",createEdgeError:"Impossible de créer un lien vers un cluster.",del:"Effacer la sélection",deleteClusterError:"Les clusters ne peuvent pas être effacés.",edgeDescription:"Cliquez sur un nœud et glissez le lien vers un autre nœud pour les connecter.",edit:"Éditer",editClusterError:"Les clusters ne peuvent pas être édités.",editEdge:"Éditer le lien",editEdgeDescription:"Cliquez sur les points de contrôle et glissez-les pour connecter un nœud.",editNode:"Éditer le nœud"},cs:{addDescription:"Kluknutím do prázdného prostoru můžete přidat nový vrchol.",addEdge:"Přidat hranu",addNode:"Přidat vrchol",back:"Zpět",close:"Zavřít",createEdgeError:"Nelze připojit hranu ke shluku.",del:"Smazat výběr",deleteClusterError:"Nelze mazat shluky.",edgeDescription:"Přetažením z jednoho vrcholu do druhého můžete spojit tyto vrcholy novou hranou.",edit:"Upravit",editClusterError:"Nelze upravovat shluky.",editEdge:"Upravit hranu",editEdgeDescription:"Přetažením kontrolního vrcholu hrany ji můžete připojit k jinému vrcholu.",editNode:"Upravit vrchol"}});var $l=function(){function t(){Nn(this,t),this.NUM_ITERATIONS=4,this.image=new Image,this.canvas=document.createElement("canvas")}return Fn(t,[{key:"init",value:function(){if(!this.initialized()){this.src=this.image.src;var t=this.image.width,e=this.image.height;this.width=t,this.height=e;var i=Math.floor(e/2),n=Math.floor(e/4),o=Math.floor(e/8),r=Math.floor(e/16),s=Math.floor(t/2),a=Math.floor(t/4),h=Math.floor(t/8),l=Math.floor(t/16);this.canvas.width=3*a,this.canvas.height=i,this.coordinates=[[0,0,s,i],[s,0,a,n],[s,n,h,o],[5*h,n,l,r]],this._fillMipMap()}}},{key:"initialized",value:function(){return void 0!==this.coordinates}},{key:"_fillMipMap",value:function(){var t=this.canvas.getContext("2d"),e=this.coordinates[0];t.drawImage(this.image,e[0],e[1],e[2],e[3]);for(var i=1;i<this.NUM_ITERATIONS;i++){var n=this.coordinates[i-1],o=this.coordinates[i];t.drawImage(this.canvas,n[0],n[1],n[2],n[3],o[0],o[1],o[2],o[3])}}},{key:"drawImageAtPosition",value:function(t,e,i,n,o,r){if(this.initialized())if(e>2){e*=.5;for(var s=0;e>2&&s<this.NUM_ITERATIONS;)e*=.5,s+=1;s>=this.NUM_ITERATIONS&&(s=this.NUM_ITERATIONS-1);var a=this.coordinates[s];t.drawImage(this.canvas,a[0],a[1],a[2],a[3],i,n,o,r)}else t.drawImage(this.image,i,n,o,r)}}]),t}(),Zl=function(){function t(e){Nn(this,t),this.images={},this.imageBroken={},this.callback=e}return Fn(t,[{key:"_tryloadBrokenUrl",value:function(t,e,i){void 0!==t&&void 0!==i&&(void 0!==e?(i.image.onerror=function(){console.error("Could not load brokenImage:",e)},i.image.src=e):console.warn("No broken url image defined"))}},{key:"_redrawWithImage",value:function(t){this.callback&&this.callback(t)}},{key:"load",value:function(t,e){var i=this,n=this.images[t];if(n)return n;var o=new $l;return this.images[t]=o,o.image.onload=function(){i._fixImageCoordinates(o.image),o.init(),i._redrawWithImage(o)},o.image.onerror=function(){console.error("Could not load image:",t),i._tryloadBrokenUrl(t,e,o)},o.image.src=t,o}},{key:"_fixImageCoordinates",value:function(t){0===t.width&&(document.body.appendChild(t),t.width=t.offsetWidth,t.height=t.offsetHeight,document.body.removeChild(t))}}]),t}(),Ql=!h((function(){return Object.isExtensible(Object.preventExtensions({}))})),Jl=n((function(t){var e=ut.f,i=!1,n=H("meta"),o=0,r=Object.isExtensible||function(){return!0},s=function(t){e(t,n,{value:{objectID:"O"+o++,weakData:{}}})},a=t.exports={enable:function(){a.enable=function(){},i=!0;var t=Ai.f,e=[].splice,o={};o[n]=1,t(o).length&&(Ai.f=function(i){for(var o=t(i),r=0,s=o.length;r<s;r++)if(o[r]===n){e.call(o,r,1);break}return o},gt({target:"Object",stat:!0,forced:!0},{getOwnPropertyNames:Li.f}))},fastKey:function(t,e){if(!w(t))return"symbol"==typeof t?t:("string"==typeof t?"S":"P")+t;if(!j(t,n)){if(!r(t))return"F";if(!e)return"E";s(t)}return t[n].objectID},getWeakData:function(t,e){if(!j(t,n)){if(!r(t))return!0;if(!e)return!1;s(t)}return t[n].weakData},onFreeze:function(t){return Ql&&i&&r(t)&&!j(t,n)&&s(t),t}};St[n]=!0}));Jl.enable,Jl.fastKey,Jl.getWeakData,Jl.onFreeze;var td=function(t,e){this.stopped=t,this.result=e},ed=function(t,e,i){var n,o,r,s,a,h,l,d=i&&i.that,c=!(!i||!i.AS_ENTRIES),u=!(!i||!i.IS_ITERATOR),f=!(!i||!i.INTERRUPTED),p=lt(e,d,1+c+f),v=function(t){return n&&di(n),new td(!0,t)},g=function(t){return c?(dt(t),f?p(t[0],t[1],v):p(t[0],t[1])):f?p(t,v):p(t)};if(u)n=t;else{if("function"!=typeof(o=yi(t)))throw TypeError("Target is not iterable");if(pi(o)){for(r=0,s=kt(t.length);s>r;r++)if((a=g(t[r]))&&a instanceof td)return a;return new td(!1)}n=o.call(t)}for(h=n.next;!(l=h.call(n)).done;){try{a=g(l.value)}catch(t){throw di(n),t}if("object"==typeof a&&a&&a instanceof td)return a}return new td(!1)},id=function(t,e,i){if(!(t instanceof e))throw TypeError("Incorrect "+(i?i+" ":"")+"invocation");return t},nd=ut.f,od=Gi.forEach,rd=we.set,sd=we.getterFor,ad=function(t,e,i){var n,o=-1!==t.indexOf("Map"),r=-1!==t.indexOf("Weak"),s=o?"set":"add",d=a[t],c=d&&d.prototype,u={};if(l&&"function"==typeof d&&(r||c.forEach&&!h((function(){(new d).entries().next()})))){n=e((function(e,i){rd(id(e,n,t),{type:t,collection:new d}),null!=i&&ed(i,e[s],{that:e,AS_ENTRIES:o})}));var f=sd(t);od(["add","clear","delete","forEach","get","has","set","keys","values","entries"],(function(t){var e="add"==t||"set"==t;!(t in c)||r&&"clear"==t||ft(n.prototype,t,(function(i,n){var o=f(this).collection;if(!e&&r&&!w(i))return"get"==t&&void 0;var s=o[t](0===i?0:i,n);return e?this:s}))})),r||nd(n.prototype,"size",{configurable:!0,get:function(){return f(this).collection.size}})}else n=i.getConstructor(e,t,o,s),Jl.enable();return Ye(n,t,!1,!0),u[t]=n,gt({global:!0,forced:!0},u),r||i.setStrong(n,t,o),n},hd=function(t,e,i){for(var n in e)i&&i.unsafe&&t[n]?t[n]=e[n]:Ze(t,n,e[n],i);return t},ld=U("species"),dd=ut.f,cd=Jl.fastKey,ud=we.set,fd=we.getterFor,pd={getConstructor:function(t,e,i,n){var o=t((function(t,r){id(t,o,e),ud(t,{type:e,index:Fe(null),first:void 0,last:void 0,size:0}),l||(t.size=0),null!=r&&ed(r,t[n],{that:t,AS_ENTRIES:i})})),r=fd(e),s=function(t,e,i){var n,o,s=r(t),h=a(t,e);return h?h.value=i:(s.last=h={index:o=cd(e,!0),key:e,value:i,previous:n=s.last,next:void 0,removed:!1},s.first||(s.first=h),n&&(n.next=h),l?s.size++:t.size++,"F"!==o&&(s.index[o]=h)),t},a=function(t,e){var i,n=r(t),o=cd(e);if("F"!==o)return n.index[o];for(i=n.first;i;i=i.next)if(i.key==e)return i};return hd(o.prototype,{clear:function(){for(var t=r(this),e=t.index,i=t.first;i;)i.removed=!0,i.previous&&(i.previous=i.previous.next=void 0),delete e[i.index],i=i.next;t.first=t.last=void 0,l?t.size=0:this.size=0},delete:function(t){var e=this,i=r(e),n=a(e,t);if(n){var o=n.next,s=n.previous;delete i.index[n.index],n.removed=!0,s&&(s.next=o),o&&(o.previous=s),i.first==n&&(i.first=o),i.last==n&&(i.last=s),l?i.size--:e.size--}return!!n},forEach:function(t){for(var e,i=r(this),n=lt(t,arguments.length>1?arguments[1]:void 0,3);e=e?e.next:i.first;)for(n(e.value,e.key,this);e&&e.removed;)e=e.previous},has:function(t){return!!a(this,t)}}),hd(o.prototype,i?{get:function(t){var e=a(this,t);return e&&e.value},set:function(t,e){return s(this,0===t?0:t,e)}}:{add:function(t){return s(this,t=0===t?0:t,t)}}),l&&dd(o.prototype,"size",{get:function(){return r(this).size}}),o},setStrong:function(t,e,i){var n=e+" Iterator",o=fd(e),r=fd(n);ri(t,e,(function(t,e){ud(this,{type:n,target:t,state:o(t),kind:e,last:void 0})}),(function(){for(var t=r(this),e=t.kind,i=t.last;i&&i.removed;)i=i.previous;return t.target&&(t.last=i=i?i.next:t.state.first)?"keys"==e?{value:i.key,done:!1}:"values"==e?{value:i.value,done:!1}:{value:[i.key,i.value],done:!1}:(t.target=void 0,{value:void 0,done:!0})}),i?"entries":"values",!i,!0),function(t){var e=x(t),i=ut.f;l&&e&&!e[ld]&&i(e,ld,{configurable:!0,get:function(){return this}})}(e)}};ad("Map",(function(t){return function(){return t(this,arguments.length?arguments[0]:void 0)}}),pd);var vd=k.Map,gd=function(){function t(){Nn(this,t),this.clear(),this._defaultIndex=0,this._groupIndex=0,this._defaultGroups=[{border:"#2B7CE9",background:"#97C2FC",highlight:{border:"#2B7CE9",background:"#D2E5FF"},hover:{border:"#2B7CE9",background:"#D2E5FF"}},{border:"#FFA500",background:"#FFFF00",highlight:{border:"#FFA500",background:"#FFFFA3"},hover:{border:"#FFA500",background:"#FFFFA3"}},{border:"#FA0A10",background:"#FB7E81",highlight:{border:"#FA0A10",background:"#FFAFB1"},hover:{border:"#FA0A10",background:"#FFAFB1"}},{border:"#41A906",background:"#7BE141",highlight:{border:"#41A906",background:"#A1EC76"},hover:{border:"#41A906",background:"#A1EC76"}},{border:"#E129F0",background:"#EB7DF4",highlight:{border:"#E129F0",background:"#F0B3F5"},hover:{border:"#E129F0",background:"#F0B3F5"}},{border:"#7C29F0",background:"#AD85E4",highlight:{border:"#7C29F0",background:"#D3BDF0"},hover:{border:"#7C29F0",background:"#D3BDF0"}},{border:"#C37F00",background:"#FFA807",highlight:{border:"#C37F00",background:"#FFCA66"},hover:{border:"#C37F00",background:"#FFCA66"}},{border:"#4220FB",background:"#6E6EFD",highlight:{border:"#4220FB",background:"#9B9BFD"},hover:{border:"#4220FB",background:"#9B9BFD"}},{border:"#FD5A77",background:"#FFC0CB",highlight:{border:"#FD5A77",background:"#FFD1D9"},hover:{border:"#FD5A77",background:"#FFD1D9"}},{border:"#4AD63A",background:"#C2FABC",highlight:{border:"#4AD63A",background:"#E6FFE3"},hover:{border:"#4AD63A",background:"#E6FFE3"}},{border:"#990000",background:"#EE0000",highlight:{border:"#BB0000",background:"#FF3333"},hover:{border:"#BB0000",background:"#FF3333"}},{border:"#FF6000",background:"#FF6000",highlight:{border:"#FF6000",background:"#FF6000"},hover:{border:"#FF6000",background:"#FF6000"}},{border:"#97C2FC",background:"#2B7CE9",highlight:{border:"#D2E5FF",background:"#2B7CE9"},hover:{border:"#D2E5FF",background:"#2B7CE9"}},{border:"#399605",background:"#255C03",highlight:{border:"#399605",background:"#255C03"},hover:{border:"#399605",background:"#255C03"}},{border:"#B70054",background:"#FF007E",highlight:{border:"#B70054",background:"#FF007E"},hover:{border:"#B70054",background:"#FF007E"}},{border:"#AD85E4",background:"#7C29F0",highlight:{border:"#D3BDF0",background:"#7C29F0"},hover:{border:"#D3BDF0",background:"#7C29F0"}},{border:"#4557FA",background:"#000EA1",highlight:{border:"#6E6EFD",background:"#000EA1"},hover:{border:"#6E6EFD",background:"#000EA1"}},{border:"#FFC0CB",background:"#FD5A77",highlight:{border:"#FFD1D9",background:"#FD5A77"},hover:{border:"#FFD1D9",background:"#FD5A77"}},{border:"#C2FABC",background:"#74D66A",highlight:{border:"#E6FFE3",background:"#74D66A"},hover:{border:"#E6FFE3",background:"#74D66A"}},{border:"#EE0000",background:"#990000",highlight:{border:"#FF3333",background:"#BB0000"},hover:{border:"#FF3333",background:"#BB0000"}}],this.options={},this.defaultOptions={useDefaultGroups:!0},At(this.options,this.defaultOptions)}return Fn(t,[{key:"setOptions",value:function(t){var e=["useDefaultGroups"];if(void 0!==t)for(var i in t)if(Object.prototype.hasOwnProperty.call(t,i)&&-1===Hr(e).call(e,i)){var n=t[i];this.add(i,n)}}},{key:"clear",value:function(){this._groups=new vd,this._groupNames=[]}},{key:"get",value:function(t){var e=!(arguments.length>1&&void 0!==arguments[1])||arguments[1],i=this._groups.get(t);if(void 0===i&&e)if(!1===this.options.useDefaultGroups&&this._groupNames.length>0){var n=this._groupIndex%this._groupNames.length;++this._groupIndex,(i={}).color=this._groups.get(this._groupNames[n]),this._groups.set(t,i)}else{var o=this._defaultIndex%this._defaultGroups.length;this._defaultIndex++,(i={}).color=this._defaultGroups[o],this._groups.set(t,i)}return i}},{key:"add",value:function(t,e){return this._groups.has(t)||this._groupNames.push(t),this._groups.set(t,e),e}}]),t}();gt({target:"Number",stat:!0},{isNaN:function(t){return t!=t}});var yd=k.Number.isNaN,md=a.isFinite,bd=Number.isFinite||function(t){return"number"==typeof t&&md(t)};gt({target:"Number",stat:!0},{isFinite:bd});var wd=k.Number.isFinite,kd=Gi.some,_d=Ao("some");gt({target:"Array",proto:!0,forced:!_d},{some:function(t){return kd(this,t,arguments.length>1?arguments[1]:void 0)}});var xd=Ht("Array").some,Ed=Array.prototype,Od=function(t){var e=t.some;return t===Ed||t instanceof Array&&e===Ed.some?xd:e},Cd=x("Reflect","construct"),Sd=h((function(){function t(){}return!(Cd((function(){}),[],t)instanceof t)})),Td=!h((function(){Cd((function(){}))})),Md=Sd||Td;gt({target:"Reflect",stat:!0,forced:Md,sham:Md},{construct:function(t,e){ht(t),dt(e);var i=arguments.length<3?t:ht(arguments[2]);if(Td&&!Sd)return Cd(t,e,i);if(t==i){switch(e.length){case 0:return new t;case 1:return new t(e[0]);case 2:return new t(e[0],e[1]);case 3:return new t(e[0],e[1],e[2]);case 4:return new t(e[0],e[1],e[2],e[3])}var n=[null];return n.push.apply(n,e),new(Lt.apply(t,n))}var o=i.prototype,r=Fe(w(o)?o:Object.prototype),s=Function.apply.call(t,r,e);return w(s)?s:r}});var Pd=k.Reflect.construct,Dd=n((function(t){t.exports=function(t){if(void 0===t)throw new ReferenceError("this hasn't been initialised - super() hasn't been called");return t},t.exports.default=t.exports,t.exports.__esModule=!0})),Id=i(Dd),Bd=Xr;gt({target:"Object",stat:!0},{setPrototypeOf:$e});var zd=k.Object.setPrototypeOf,Nd=n((function(t){function e(i,n){return t.exports=e=zd||function(t,e){return t.__proto__=e,t},t.exports.default=t.exports,t.exports.__esModule=!0,e(i,n)}t.exports=e,t.exports.default=t.exports,t.exports.__esModule=!0}));i(Nd);var Ad=i(n((function(t){t.exports=function(t,e){if("function"!=typeof e&&null!==e)throw new TypeError("Super expression must either be null or a function");t.prototype=Bd(e&&e.prototype,{constructor:{value:t,writable:!0,configurable:!0}}),e&&Nd(t,e)},t.exports.default=t.exports,t.exports.__esModule=!0}))),Fd=i(n((function(t){var e=vo.default;t.exports=function(t,i){if(i&&("object"===e(i)||"function"==typeof i))return i;if(void 0!==i)throw new TypeError("Derived constructors may only return object or undefined");return Dd(t)},t.exports.default=t.exports,t.exports.__esModule=!0}))),jd=ur,Rd=n((function(t){function e(i){return t.exports=e=zd?jd:function(t){return t.__proto__||jd(t)},t.exports.default=t.exports,t.exports.__esModule=!0,e(i)}t.exports=e,t.exports.default=t.exports,t.exports.__esModule=!0})),Ld=i(Rd),Hd=n((function(t){var e=function(t){var e,i=Object.prototype,n=i.hasOwnProperty,o="function"==typeof Symbol?Symbol:{},r=o.iterator||"@@iterator",s=o.asyncIterator||"@@asyncIterator",a=o.toStringTag||"@@toStringTag";function h(t,e,i){return Object.defineProperty(t,e,{value:i,enumerable:!0,configurable:!0,writable:!0}),t[e]}try{h({},"")}catch(t){h=function(t,e,i){return t[e]=i}}function l(t,e,i,n){var o=e&&e.prototype instanceof g?e:g,r=Object.create(o.prototype),s=new T(n||[]);return r._invoke=function(t,e,i){var n=c;return function(o,r){if(n===f)throw new Error("Generator is already running");if(n===p){if("throw"===o)throw r;return P()}for(i.method=o,i.arg=r;;){var s=i.delegate;if(s){var a=O(s,i);if(a){if(a===v)continue;return a}}if("next"===i.method)i.sent=i._sent=i.arg;else if("throw"===i.method){if(n===c)throw n=p,i.arg;i.dispatchException(i.arg)}else"return"===i.method&&i.abrupt("return",i.arg);n=f;var h=d(t,e,i);if("normal"===h.type){if(n=i.done?p:u,h.arg===v)continue;return{value:h.arg,done:i.done}}"throw"===h.type&&(n=p,i.method="throw",i.arg=h.arg)}}}(t,i,s),r}function d(t,e,i){try{return{type:"normal",arg:t.call(e,i)}}catch(t){return{type:"throw",arg:t}}}t.wrap=l;var c="suspendedStart",u="suspendedYield",f="executing",p="completed",v={};function g(){}function y(){}function m(){}var b={};b[r]=function(){return this};var w=Object.getPrototypeOf,k=w&&w(w(M([])));k&&k!==i&&n.call(k,r)&&(b=k);var _=m.prototype=g.prototype=Object.create(b);function x(t){["next","throw","return"].forEach((function(e){h(t,e,(function(t){return this._invoke(e,t)}))}))}function E(t,e){function i(o,r,s,a){var h=d(t[o],t,r);if("throw"!==h.type){var l=h.arg,c=l.value;return c&&"object"==typeof c&&n.call(c,"__await")?e.resolve(c.__await).then((function(t){i("next",t,s,a)}),(function(t){i("throw",t,s,a)})):e.resolve(c).then((function(t){l.value=t,s(l)}),(function(t){return i("throw",t,s,a)}))}a(h.arg)}var o;this._invoke=function(t,n){function r(){return new e((function(e,o){i(t,n,e,o)}))}return o=o?o.then(r,r):r()}}function O(t,i){var n=t.iterator[i.method];if(n===e){if(i.delegate=null,"throw"===i.method){if(t.iterator.return&&(i.method="return",i.arg=e,O(t,i),"throw"===i.method))return v;i.method="throw",i.arg=new TypeError("The iterator does not provide a 'throw' method")}return v}var o=d(n,t.iterator,i.arg);if("throw"===o.type)return i.method="throw",i.arg=o.arg,i.delegate=null,v;var r=o.arg;return r?r.done?(i[t.resultName]=r.value,i.next=t.nextLoc,"return"!==i.method&&(i.method="next",i.arg=e),i.delegate=null,v):r:(i.method="throw",i.arg=new TypeError("iterator result is not an object"),i.delegate=null,v)}function C(t){var e={tryLoc:t[0]};1 in t&&(e.catchLoc=t[1]),2 in t&&(e.finallyLoc=t[2],e.afterLoc=t[3]),this.tryEntries.push(e)}function S(t){var e=t.completion||{};e.type="normal",delete e.arg,t.completion=e}function T(t){this.tryEntries=[{tryLoc:"root"}],t.forEach(C,this),this.reset(!0)}function M(t){if(t){var i=t[r];if(i)return i.call(t);if("function"==typeof t.next)return t;if(!isNaN(t.length)){var o=-1,s=function i(){for(;++o<t.length;)if(n.call(t,o))return i.value=t[o],i.done=!1,i;return i.value=e,i.done=!0,i};return s.next=s}}return{next:P}}function P(){return{value:e,done:!0}}return y.prototype=_.constructor=m,m.constructor=y,y.displayName=h(m,a,"GeneratorFunction"),t.isGeneratorFunction=function(t){var e="function"==typeof t&&t.constructor;return!!e&&(e===y||"GeneratorFunction"===(e.displayName||e.name))},t.mark=function(t){return Object.setPrototypeOf?Object.setPrototypeOf(t,m):(t.__proto__=m,h(t,a,"GeneratorFunction")),t.prototype=Object.create(_),t},t.awrap=function(t){return{__await:t}},x(E.prototype),E.prototype[s]=function(){return this},t.AsyncIterator=E,t.async=function(e,i,n,o,r){void 0===r&&(r=Promise);var s=new E(l(e,i,n,o),r);return t.isGeneratorFunction(i)?s:s.next().then((function(t){return t.done?t.value:s.next()}))},x(_),h(_,a,"Generator"),_[r]=function(){return this},_.toString=function(){return"[object Generator]"},t.keys=function(t){var e=[];for(var i in t)e.push(i);return e.reverse(),function i(){for(;e.length;){var n=e.pop();if(n in t)return i.value=n,i.done=!1,i}return i.done=!0,i}},t.values=M,T.prototype={constructor:T,reset:function(t){if(this.prev=0,this.next=0,this.sent=this._sent=e,this.done=!1,this.delegate=null,this.method="next",this.arg=e,this.tryEntries.forEach(S),!t)for(var i in this)"t"===i.charAt(0)&&n.call(this,i)&&!isNaN(+i.slice(1))&&(this[i]=e)},stop:function(){this.done=!0;var t=this.tryEntries[0].completion;if("throw"===t.type)throw t.arg;return this.rval},dispatchException:function(t){if(this.done)throw t;var i=this;function o(n,o){return a.type="throw",a.arg=t,i.next=n,o&&(i.method="next",i.arg=e),!!o}for(var r=this.tryEntries.length-1;r>=0;--r){var s=this.tryEntries[r],a=s.completion;if("root"===s.tryLoc)return o("end");if(s.tryLoc<=this.prev){var h=n.call(s,"catchLoc"),l=n.call(s,"finallyLoc");if(h&&l){if(this.prev<s.catchLoc)return o(s.catchLoc,!0);if(this.prev<s.finallyLoc)return o(s.finallyLoc)}else if(h){if(this.prev<s.catchLoc)return o(s.catchLoc,!0)}else{if(!l)throw new Error("try statement without catch or finally");if(this.prev<s.finallyLoc)return o(s.finallyLoc)}}}},abrupt:function(t,e){for(var i=this.tryEntries.length-1;i>=0;--i){var o=this.tryEntries[i];if(o.tryLoc<=this.prev&&n.call(o,"finallyLoc")&&this.prev<o.finallyLoc){var r=o;break}}r&&("break"===t||"continue"===t)&&r.tryLoc<=e&&e<=r.finallyLoc&&(r=null);var s=r?r.completion:{};return s.type=t,s.arg=e,r?(this.method="next",this.next=r.finallyLoc,v):this.complete(s)},complete:function(t,e){if("throw"===t.type)throw t.arg;return"break"===t.type||"continue"===t.type?this.next=t.arg:"return"===t.type?(this.rval=this.arg=t.arg,this.method="return",this.next="end"):"normal"===t.type&&e&&(this.next=e),v},finish:function(t){for(var e=this.tryEntries.length-1;e>=0;--e){var i=this.tryEntries[e];if(i.finallyLoc===t)return this.complete(i.completion,i.afterLoc),S(i),v}},catch:function(t){for(var e=this.tryEntries.length-1;e>=0;--e){var i=this.tryEntries[e];if(i.tryLoc===t){var n=i.completion;if("throw"===n.type){var o=n.arg;S(i)}return o}}throw new Error("illegal catch attempt")},delegateYield:function(t,i,n){return this.delegate={iterator:M(t),resultName:i,nextLoc:n},"next"===this.method&&(this.arg=e),v}},t}(t.exports);try{regeneratorRuntime=e}catch(t){Function("r","regeneratorRuntime = r")(e)}})),Wd=function(t){return function(e,i,n,o){ht(i);var r=A(e),s=y(r),a=kt(r.length),h=t?a-1:0,l=t?-1:1;if(n<2)for(;;){if(h in s){o=s[h],h+=l;break}if(h+=l,t?h<0:a<=h)throw TypeError("Reduce of empty array with no initial value")}for(;t?h>=0:a>h;h+=l)h in s&&(o=i(o,s[h],h,r));return o}},qd={left:Wd(!1),right:Wd(!0)},Vd="process"==v(a.process),Ud=qd.left,Yd=Ao("reduce");gt({target:"Array",proto:!0,forced:!Yd||!Vd&&M>79&&M<83},{reduce:function(t){return Ud(this,t,arguments.length,arguments.length>1?arguments[1]:void 0)}});var Xd=Ht("Array").reduce,Gd=Array.prototype,Kd=function(t){var e=t.reduce;return t===Gd||t instanceof Array&&e===Gd.reduce?Xd:e},$d=function(t,e,i,n,o,r,s,a){for(var h,l=o,d=0,c=!!s&&lt(s,a,3);d<n;){if(d in i){if(h=c?c(i[d],d,e):i[d],r>0&&zi(h))l=$d(t,e,h,kt(h.length),l,r-1)-1;else{if(l>=9007199254740991)throw TypeError("Exceed the acceptable array length");t[l]=h}l++}d++}return l},Zd=$d;gt({target:"Array",proto:!0},{flatMap:function(t){var e,i=A(this),n=kt(i.length);return ht(t),(e=Ui(i,0)).length=Zd(e,i,i,n,0,1,t,arguments.length>1?arguments[1]:void 0),e}});var Qd=Ht("Array").flatMap,Jd=Array.prototype,tc=function(t){var e=t.flatMap;return t===Jd||t instanceof Array&&e===Jd.flatMap?Qd:e};ad("Set",(function(t){return function(){return t(this,arguments.length?arguments[0]:void 0)}}),pd);var ec=k.Set,ic=fo,nc=function(t){var e=yi(t);if("function"!=typeof e)throw TypeError(String(t)+" is not iterable");return dt(e.call(t))},oc=Math.floor,rc=function(t,e){var i=t.length,n=oc(i/2);return i<8?sc(t,e):ac(rc(t.slice(0,n),e),rc(t.slice(n),e),e)},sc=function(t,e){for(var i,n,o=t.length,r=1;r<o;){for(n=r,i=t[r];n&&e(t[n-1],i)>0;)t[n]=t[--n];n!==r++&&(t[n]=i)}return t},ac=function(t,e,i){for(var n=t.length,o=e.length,r=0,s=0,a=[];r<n||s<o;)r<n&&s<o?a.push(i(t[r],e[s])<=0?t[r++]:e[s++]):a.push(r<n?t[r++]:e[s++]);return a},hc=rc,lc=E.match(/firefox\/(\d+)/i),dc=!!lc&&+lc[1],cc=/MSIE|Trident/.test(E),uc=E.match(/AppleWebKit\/(\d+)\./),fc=!!uc&&+uc[1],pc=[],vc=pc.sort,gc=h((function(){pc.sort(void 0)})),yc=h((function(){pc.sort(null)})),mc=Ao("sort"),bc=!h((function(){if(M)return M<70;if(!(dc&&dc>3)){if(cc)return!0;if(fc)return fc<603;var t,e,i,n,o="";for(t=65;t<76;t++){switch(e=String.fromCharCode(t),t){case 66:case 69:case 70:case 72:i=3;break;case 68:case 71:i=4;break;default:i=2}for(n=0;n<47;n++)pc.push({k:e+n,v:i})}for(pc.sort((function(t,e){return e.v-t.v})),n=0;n<pc.length;n++)e=pc[n].k.charAt(0),o.charAt(o.length-1)!==e&&(o+=e);return"DGBEFHACIJK"!==o}}));gt({target:"Array",proto:!0,forced:gc||!yc||!mc||!bc},{sort:function(t){void 0!==t&&ht(t);var e=A(this);if(bc)return void 0===t?vc.call(e):vc.call(e,t);var i,n,o=[],r=kt(e.length);for(n=0;n<r;n++)n in e&&o.push(e[n]);for(i=(o=hc(o,function(t){return function(e,i){return void 0===i?-1:void 0===e?1:void 0!==t?+t(e,i)||0:Qt(e)>Qt(i)?1:-1}}(t))).length,n=0;n<i;)e[n]=o[n++];for(;n<r;)delete e[n++];return e}});var wc,kc=Ht("Array").sort,_c=Array.prototype,xc=function(t){var e=t.sort;return t===_c||t instanceof Array&&e===_c.sort?kc:e},Ec=Ht("Array").keys,Oc=Array.prototype,Cc={DOMTokenList:!0,NodeList:!0},Sc=function(t){var e=t.keys;return t===Oc||t instanceof Array&&e===Oc.keys||Cc.hasOwnProperty(We(t))?Ec:e},Tc=Ht("Array").values,Mc=Array.prototype,Pc={DOMTokenList:!0,NodeList:!0},Dc=function(t){var e=t.values;return t===Mc||t instanceof Array&&e===Mc.values||Pc.hasOwnProperty(We(t))?Tc:e},Ic=Ht("Array").entries,Bc=Array.prototype,zc={DOMTokenList:!0,NodeList:!0},Nc=function(t){var e=t.entries;return t===Bc||t instanceof Array&&e===Bc.entries||zc.hasOwnProperty(We(t))?Ic:e},Ac=new Uint8Array(16);function Fc(){if(!wc&&!(wc="undefined"!=typeof crypto&&crypto.getRandomValues&&crypto.getRandomValues.bind(crypto)||"undefined"!=typeof msCrypto&&"function"==typeof msCrypto.getRandomValues&&msCrypto.getRandomValues.bind(msCrypto)))throw new Error("crypto.getRandomValues() not supported. See https://github.com/uuidjs/uuid#getrandomvalues-not-supported");return wc(Ac)}var jc=/^(?:[0-9a-f]{8}-[0-9a-f]{4}-[1-5][0-9a-f]{3}-[89ab][0-9a-f]{3}-[0-9a-f]{12}|00000000-0000-0000-0000-000000000000)$/i;function Rc(t){return"string"==typeof t&&jc.test(t)}for(var Lc=[],Hc=0;Hc<256;++Hc)Lc.push((Hc+256).toString(16).substr(1));function Wc(t,e,i){var n=(t=t||{}).random||(t.rng||Fc)();if(n[6]=15&n[6]|64,n[8]=63&n[8]|128,e){i=i||0;for(var o=0;o<16;++o)e[i+o]=n[o];return e}return function(t){var e=arguments.length>1&&void 0!==arguments[1]?arguments[1]:0,i=(Lc[t[e+0]]+Lc[t[e+1]]+Lc[t[e+2]]+Lc[t[e+3]]+"-"+Lc[t[e+4]]+Lc[t[e+5]]+"-"+Lc[t[e+6]]+Lc[t[e+7]]+"-"+Lc[t[e+8]]+Lc[t[e+9]]+"-"+Lc[t[e+10]]+Lc[t[e+11]]+Lc[t[e+12]]+Lc[t[e+13]]+Lc[t[e+14]]+Lc[t[e+15]]).toLowerCase();if(!Rc(i))throw TypeError("Stringified UUID is invalid");return i}(n)}function qc(t,e){var i=zo(t);if(On){var n=On(t);e&&(n=mr(n).call(n,(function(e){return Mn(t,e).enumerable}))),i.push.apply(i,n)}return i}function Vc(t){for(var e=1;e<arguments.length;e++){var i,n=null!=arguments[e]?arguments[e]:{};if(e%2)Wo(i=qc(Object(n),!0)).call(i,(function(e){jn(t,e,n[e])}));else if(Dn)In(t,Dn(n));else{var o;Wo(o=qc(Object(n))).call(o,(function(e){zn(t,e,Mn(n,e))}))}}return t}function Uc(t){var e=function(){if("undefined"==typeof Reflect||!Pd)return!1;if(Pd.sham)return!1;if("function"==typeof Proxy)return!0;try{return Boolean.prototype.valueOf.call(Pd(Boolean,[],(function(){}))),!0}catch(t){return!1}}();return function(){var i,n=Ld(t);if(e){var o=Ld(this).constructor;i=Pd(n,arguments,o)}else i=n.apply(this,arguments);return Fd(this,i)}}function Yc(t,e){var i=void 0!==ko&&Bi(t)||t["@@iterator"];if(!i){if(So(t)||(i=function(t,e){var i;if(!t)return;if("string"==typeof t)return Xc(t,e);var n=Oo(i=Object.prototype.toString.call(t)).call(i,8,-1);"Object"===n&&t.constructor&&(n=t.constructor.name);if("Map"===n||"Set"===n)return Ei(t);if("Arguments"===n||/^(?:Ui|I)nt(?:8|16|32)(?:Clamped)?Array$/.test(n))return Xc(t,e)}(t))||e&&t&&"number"==typeof t.length){i&&(t=i);var n=0,o=function(){};return{s:o,n:function(){return n>=t.length?{done:!0}:{done:!1,value:t[n++]}},e:function(t){throw t},f:o}}throw new TypeError("Invalid attempt to iterate non-iterable instance.\nIn order to be iterable, non-array objects must have a [Symbol.iterator]() method.")}var r,s=!0,a=!1;return{s:function(){i=i.call(t)},n:function(){var t=i.next();return s=t.done,t},e:function(t){a=!0,r=t},f:function(){try{s||null==i.return||i.return()}finally{if(a)throw r}}}}function Xc(t,e){(null==e||e>t.length)&&(e=t.length);for(var i=0,n=new Array(e);i<e;i++)n[i]=t[i];return n}var Gc=function(){function t(e,i,n){var o,r,s;Nn(this,t),this._source=e,this._transformers=i,this._target=n,this._listeners={add:Vt(o=this._add).call(o,this),remove:Vt(r=this._remove).call(r,this),update:Vt(s=this._update).call(s,this)}}return Fn(t,[{key:"all",value:function(){return this._target.update(this._transformItems(this._source.get())),this}},{key:"start",value:function(){return this._source.on("add",this._listeners.add),this._source.on("remove",this._listeners.remove),this._source.on("update",this._listeners.update),this}},{key:"stop",value:function(){return this._source.off("add",this._listeners.add),this._source.off("remove",this._listeners.remove),this._source.off("update",this._listeners.update),this}},{key:"_transformItems",value:function(t){var e;return Kd(e=this._transformers).call(e,(function(t,e){return e(t)}),t)}},{key:"_add",value:function(t,e){null!=e&&this._target.add(this._transformItems(this._source.get(e.items)))}},{key:"_update",value:function(t,e){null!=e&&this._target.update(this._transformItems(this._source.get(e.items)))}},{key:"_remove",value:function(t,e){null!=e&&this._target.remove(this._transformItems(e.oldData))}}]),t}(),Kc=function(){function t(e){Nn(this,t),this._source=e,this._transformers=[]}return Fn(t,[{key:"filter",value:function(t){return this._transformers.push((function(e){return mr(e).call(e,t)})),this}},{key:"map",value:function(t){return this._transformers.push((function(e){return Io(e).call(e,t)})),this}},{key:"flatMap",value:function(t){return this._transformers.push((function(e){return tc(e).call(e,t)})),this}},{key:"to",value:function(t){return new Gc(this._source,this._transformers,t)}}]),t}();function $c(t){return"string"==typeof t||"number"==typeof t}var Zc=function(){function t(e){Nn(this,t),this._queue=[],this._timeout=null,this._extended=null,this.delay=null,this.max=1/0,this.setOptions(e)}return Fn(t,[{key:"setOptions",value:function(t){t&&void 0!==t.delay&&(this.delay=t.delay),t&&void 0!==t.max&&(this.max=t.max),this._flushIfNeeded()}},{key:"destroy",value:function(){if(this.flush(),this._extended){for(var t=this._extended.object,e=this._extended.methods,i=0;i<e.length;i++){var n=e[i];n.original?t[n.name]=n.original:delete t[n.name]}this._extended=null}}},{key:"replace",value:function(t,e){var i=this,n=t[e];if(!n)throw new Error("Method "+e+" undefined");t[e]=function(){for(var t=arguments.length,e=new Array(t),o=0;o<t;o++)e[o]=arguments[o];i.queue({args:e,fn:n,context:this})}}},{key:"queue",value:function(t){"function"==typeof t?this._queue.push({fn:t}):this._queue.push(t),this._flushIfNeeded()}},{key:"_flushIfNeeded",value:function(){var t=this;this._queue.length>this.max&&this.flush(),null!=this._timeout&&(clearTimeout(this._timeout),this._timeout=null),this.queue.length>0&&"number"==typeof this.delay&&(this._timeout=rs((function(){t.flush()}),this.delay))}},{key:"flush",value:function(){var t,e;Wo(t=er(e=this._queue).call(e,0)).call(t,(function(t){t.fn.apply(t.context||t.fn,t.args||[])}))}}],[{key:"extend",value:function(e,i){var n=new t(i);if(void 0!==e.flush)throw new Error("Target object already has a property flush");e.flush=function(){n.flush()};var o=[{name:"flush",original:void 0}];if(i&&i.replace)for(var r=0;r<i.replace.length;r++){var s=i.replace[r];o.push({name:s,original:e[s]}),n.replace(e,s)}return n._extended={object:e,methods:o},n}}]),t}(),Qc=function(){function t(){Nn(this,t),this._subscribers={"*":[],add:[],remove:[],update:[]},this.subscribe=t.prototype.on,this.unsubscribe=t.prototype.off}return Fn(t,[{key:"_trigger",value:function(t,e,i){var n,o;if("*"===t)throw new Error("Cannot trigger event *");Wo(n=Eo(o=[]).call(o,wo(this._subscribers[t]),wo(this._subscribers["*"]))).call(n,(function(n){n(t,e,null!=i?i:null)}))}},{key:"on",value:function(t,e){"function"==typeof e&&this._subscribers[t].push(e)}},{key:"off",value:function(t,e){var i;this._subscribers[t]=mr(i=this._subscribers[t]).call(i,(function(t){return t!==e}))}}]),t}(),Jc=function(t){function e(t){Nn(this,e),this._pairs=t}return Fn(e,[{key:t,value:Hd.mark((function t(){var e,i,n,o,r;return Hd.wrap((function(t){for(;;)switch(t.prev=t.next){case 0:e=Yc(this._pairs),t.prev=1,e.s();case 3:if((i=e.n()).done){t.next=9;break}return n=uo(i.value,2),o=n[0],r=n[1],t.next=7,[o,r];case 7:t.next=3;break;case 9:t.next=14;break;case 11:t.prev=11,t.t0=t.catch(1),e.e(t.t0);case 14:return t.prev=14,e.f(),t.finish(14);case 17:case"end":return t.stop()}}),t,this,[[1,11,14,17]])}))},{key:"entries",value:Hd.mark((function t(){var e,i,n,o,r;return Hd.wrap((function(t){for(;;)switch(t.prev=t.next){case 0:e=Yc(this._pairs),t.prev=1,e.s();case 3:if((i=e.n()).done){t.next=9;break}return n=uo(i.value,2),o=n[0],r=n[1],t.next=7,[o,r];case 7:t.next=3;break;case 9:t.next=14;break;case 11:t.prev=11,t.t0=t.catch(1),e.e(t.t0);case 14:return t.prev=14,e.f(),t.finish(14);case 17:case"end":return t.stop()}}),t,this,[[1,11,14,17]])}))},{key:"keys",value:Hd.mark((function t(){var e,i,n,o;return Hd.wrap((function(t){for(;;)switch(t.prev=t.next){case 0:e=Yc(this._pairs),t.prev=1,e.s();case 3:if((i=e.n()).done){t.next=9;break}return n=uo(i.value,1),o=n[0],t.next=7,o;case 7:t.next=3;break;case 9:t.next=14;break;case 11:t.prev=11,t.t0=t.catch(1),e.e(t.t0);case 14:return t.prev=14,e.f(),t.finish(14);case 17:case"end":return t.stop()}}),t,this,[[1,11,14,17]])}))},{key:"values",value:Hd.mark((function t(){var e,i,n,o;return Hd.wrap((function(t){for(;;)switch(t.prev=t.next){case 0:e=Yc(this._pairs),t.prev=1,e.s();case 3:if((i=e.n()).done){t.next=9;break}return n=uo(i.value,2),o=n[1],t.next=7,o;case 7:t.next=3;break;case 9:t.next=14;break;case 11:t.prev=11,t.t0=t.catch(1),e.e(t.t0);case 14:return t.prev=14,e.f(),t.finish(14);case 17:case"end":return t.stop()}}),t,this,[[1,11,14,17]])}))},{key:"toIdArray",value:function(){var t;return Io(t=wo(this._pairs)).call(t,(function(t){return t[0]}))}},{key:"toItemArray",value:function(){var t;return Io(t=wo(this._pairs)).call(t,(function(t){return t[1]}))}},{key:"toEntryArray",value:function(){return wo(this._pairs)}},{key:"toObjectMap",value:function(){var t,e=Gr(null),i=Yc(this._pairs);try{for(i.s();!(t=i.n()).done;){var n=uo(t.value,2),o=n[0],r=n[1];e[o]=r}}catch(t){i.e(t)}finally{i.f()}return e}},{key:"toMap",value:function(){return new vd(this._pairs)}},{key:"toIdSet",value:function(){return new ec(this.toIdArray())}},{key:"toItemSet",value:function(){return new ec(this.toItemArray())}},{key:"cache",value:function(){return new e(wo(this._pairs))}},{key:"distinct",value:function(t){var e,i=new ec,n=Yc(this._pairs);try{for(n.s();!(e=n.n()).done;){var o=uo(e.value,2),r=o[0],s=o[1];i.add(t(s,r))}}catch(t){n.e(t)}finally{n.f()}return i}},{key:"filter",value:function(t){var i=this._pairs;return new e(jn({},ic,Hd.mark((function e(){var n,o,r,s,a;return Hd.wrap((function(e){for(;;)switch(e.prev=e.next){case 0:n=Yc(i),e.prev=1,n.s();case 3:if((o=n.n()).done){e.next=10;break}if(r=uo(o.value,2),s=r[0],a=r[1],!t(a,s)){e.next=8;break}return e.next=8,[s,a];case 8:e.next=3;break;case 10:e.next=15;break;case 12:e.prev=12,e.t0=e.catch(1),n.e(e.t0);case 15:return e.prev=15,n.f(),e.finish(15);case 18:case"end":return e.stop()}}),e,null,[[1,12,15,18]])}))))}},{key:"forEach",value:function(t){var e,i=Yc(this._pairs);try{for(i.s();!(e=i.n()).done;){var n=uo(e.value,2),o=n[0];t(n[1],o)}}catch(t){i.e(t)}finally{i.f()}}},{key:"map",value:function(t){var i=this._pairs;return new e(jn({},ic,Hd.mark((function e(){var n,o,r,s,a;return Hd.wrap((function(e){for(;;)switch(e.prev=e.next){case 0:n=Yc(i),e.prev=1,n.s();case 3:if((o=n.n()).done){e.next=9;break}return r=uo(o.value,2),s=r[0],a=r[1],e.next=7,[s,t(a,s)];case 7:e.next=3;break;case 9:e.next=14;break;case 11:e.prev=11,e.t0=e.catch(1),n.e(e.t0);case 14:return e.prev=14,n.f(),e.finish(14);case 17:case"end":return e.stop()}}),e,null,[[1,11,14,17]])}))))}},{key:"max",value:function(t){var e=nc(this._pairs),i=e.next();if(i.done)return null;for(var n=i.value[1],o=t(i.value[1],i.value[0]);!(i=e.next()).done;){var r=uo(i.value,2),s=r[0],a=r[1],h=t(a,s);h>o&&(o=h,n=a)}return n}},{key:"min",value:function(t){var e=nc(this._pairs),i=e.next();if(i.done)return null;for(var n=i.value[1],o=t(i.value[1],i.value[0]);!(i=e.next()).done;){var r=uo(i.value,2),s=r[0],a=r[1],h=t(a,s);h<o&&(o=h,n=a)}return n}},{key:"reduce",value:function(t,e){var i,n=Yc(this._pairs);try{for(n.s();!(i=n.n()).done;){var o=uo(i.value,2),r=o[0];e=t(e,o[1],r)}}catch(t){n.e(t)}finally{n.f()}return e}},{key:"sort",value:function(t){var i=this;return new e(jn({},ic,(function(){var e;return nc(xc(e=wo(i._pairs)).call(e,(function(e,i){var n=uo(e,2),o=n[0],r=n[1],s=uo(i,2),a=s[0],h=s[1];return t(r,h,o,a)})))})))}}]),e}(ic);var tu=function(t){Ad(i,t);var e=Uc(i);function i(t,n){var o;return Nn(this,i),(o=e.call(this))._queue=null,t&&!So(t)&&(n=t,t=[]),o._options=n||{},o._data=new vd,o.length=0,o._idProp=o._options.fieldId||"id",t&&t.length&&o.add(t),o.setOptions(n),o}return Fn(i,[{key:"idProp",get:function(){return this._idProp}},{key:"setOptions",value:function(t){t&&void 0!==t.queue&&(!1===t.queue?this._queue&&(this._queue.destroy(),this._queue=null):(this._queue||(this._queue=Zc.extend(this,{replace:["add","update","remove"]})),t.queue&&"object"===go(t.queue)&&this._queue.setOptions(t.queue)))}},{key:"add",value:function(t,e){var i,n=this,o=[];if(So(t)){var r=Io(t).call(t,(function(t){return t[n._idProp]}));if(Od(r).call(r,(function(t){return n._data.has(t)})))throw new Error("A duplicate id was found in the parameter array.");for(var s=0,a=t.length;s<a;s++)i=this._addItem(t[s]),o.push(i)}else{if(!t||"object"!==go(t))throw new Error("Unknown dataType");i=this._addItem(t),o.push(i)}return o.length&&this._trigger("add",{items:o},e),o}},{key:"update",value:function(t,e){var i=this,n=[],o=[],r=[],s=[],a=this._idProp,h=function(t){var e=t[a];if(null!=e&&i._data.has(e)){var h=t,l=At({},i._data.get(e)),d=i._updateItem(h);o.push(d),s.push(h),r.push(l)}else{var c=i._addItem(t);n.push(c)}};if(So(t))for(var l=0,d=t.length;l<d;l++)t[l]&&"object"===go(t[l])?h(t[l]):console.warn("Ignoring input item, which is not an object at index "+l);else{if(!t||"object"!==go(t))throw new Error("Unknown dataType");h(t)}if(n.length&&this._trigger("add",{items:n},e),o.length){var c={items:o,oldData:r,data:s};this._trigger("update",c,e)}return Eo(n).call(n,o)}},{key:"updateOnly",value:function(t,e){var i,n=this;So(t)||(t=[t]);var o=Io(i=Io(t).call(t,(function(t){var e=n._data.get(t[n._idProp]);if(null==e)throw new Error("Updating non-existent items is not allowed.");return{oldData:e,update:t}}))).call(i,(function(t){var e=t.oldData,i=t.update,o=e[n._idProp],r=ih(e,i);return n._data.set(o,r),{id:o,oldData:e,updatedData:r}}));if(o.length){var r={items:Io(o).call(o,(function(t){return t.id})),oldData:Io(o).call(o,(function(t){return t.oldData})),data:Io(o).call(o,(function(t){return t.updatedData}))};return this._trigger("update",r,e),r.items}return[]}},{key:"get",value:function(t,e){var i=void 0,n=void 0,o=void 0;$c(t)?(i=t,o=e):So(t)?(n=t,o=e):o=t;var r,s=o&&"Object"===o.returnType?"Object":"Array",a=o&&mr(o),h=[],l=void 0,d=void 0,c=void 0;if(null!=i)(l=this._data.get(i))&&a&&!a(l)&&(l=void 0);else if(null!=n)for(var u=0,f=n.length;u<f;u++)null==(l=this._data.get(n[u]))||a&&!a(l)||h.push(l);else for(var p,v=0,g=(d=wo(Sc(p=this._data).call(p))).length;v<g;v++)c=d[v],null==(l=this._data.get(c))||a&&!a(l)||h.push(l);if(o&&o.order&&null==i&&this._sort(h,o.order),o&&o.fields){var y=o.fields;if(null!=i&&null!=l)l=this._filterFields(l,y);else for(var m=0,b=h.length;m<b;m++)h[m]=this._filterFields(h[m],y)}if("Object"==s){for(var w={},k=0,_=h.length;k<_;k++){var x=h[k];w[x[this._idProp]]=x}return w}return null!=i?null!==(r=l)&&void 0!==r?r:null:h}},{key:"getIds",value:function(t){var e=this._data,i=t&&mr(t),n=t&&t.order,o=wo(Sc(e).call(e)),r=[];if(i)if(n){for(var s=[],a=0,h=o.length;a<h;a++){var l=o[a],d=this._data.get(l);null!=d&&i(d)&&s.push(d)}this._sort(s,n);for(var c=0,u=s.length;c<u;c++)r.push(s[c][this._idProp])}else for(var f=0,p=o.length;f<p;f++){var v=o[f],g=this._data.get(v);null!=g&&i(g)&&r.push(g[this._idProp])}else if(n){for(var y=[],m=0,b=o.length;m<b;m++){var w=o[m];y.push(e.get(w))}this._sort(y,n);for(var k=0,_=y.length;k<_;k++)r.push(y[k][this._idProp])}else for(var x=0,E=o.length;x<E;x++){var O=o[x],C=e.get(O);null!=C&&r.push(C[this._idProp])}return r}},{key:"getDataSet",value:function(){return this}},{key:"forEach",value:function(t,e){var i=e&&mr(e),n=this._data,o=wo(Sc(n).call(n));if(e&&e.order)for(var r=this.get(e),s=0,a=r.length;s<a;s++){var h=r[s];t(h,h[this._idProp])}else for(var l=0,d=o.length;l<d;l++){var c=o[l],u=this._data.get(c);null==u||i&&!i(u)||t(u,c)}}},{key:"map",value:function(t,e){for(var i=e&&mr(e),n=[],o=this._data,r=wo(Sc(o).call(o)),s=0,a=r.length;s<a;s++){var h=r[s],l=this._data.get(h);null==l||i&&!i(l)||n.push(t(l,h))}return e&&e.order&&this._sort(n,e.order),n}},{key:"_filterFields",value:function(t,e){var i;return t?Kd(i=So(e)?e:zo(e)).call(i,(function(e,i){return e[i]=t[i],e}),{}):t}},{key:"_sort",value:function(t,e){if("string"==typeof e){var i=e;xc(t).call(t,(function(t,e){var n=t[i],o=e[i];return n>o?1:n<o?-1:0}))}else{if("function"!=typeof e)throw new TypeError("Order must be a function or a string");xc(t).call(t,e)}}},{key:"remove",value:function(t,e){for(var i=[],n=[],o=So(t)?t:[t],r=0,s=o.length;r<s;r++){var a=this._remove(o[r]);if(a){var h=a[this._idProp];null!=h&&(i.push(h),n.push(a))}}return i.length&&this._trigger("remove",{items:i,oldData:n},e),i}},{key:"_remove",value:function(t){var e;if($c(t)?e=t:t&&"object"===go(t)&&(e=t[this._idProp]),null!=e&&this._data.has(e)){var i=this._data.get(e)||null;return this._data.delete(e),--this.length,i}return null}},{key:"clear",value:function(t){for(var e,i=wo(Sc(e=this._data).call(e)),n=[],o=0,r=i.length;o<r;o++)n.push(this._data.get(i[o]));return this._data.clear(),this.length=0,this._trigger("remove",{items:i,oldData:n},t),i}},{key:"max",value:function(t){var e,i,n=null,o=null,r=Yc(Dc(e=this._data).call(e));try{for(r.s();!(i=r.n()).done;){var s=i.value,a=s[t];"number"==typeof a&&(null==o||a>o)&&(n=s,o=a)}}catch(t){r.e(t)}finally{r.f()}return n||null}},{key:"min",value:function(t){var e,i,n=null,o=null,r=Yc(Dc(e=this._data).call(e));try{for(r.s();!(i=r.n()).done;){var s=i.value,a=s[t];"number"==typeof a&&(null==o||a<o)&&(n=s,o=a)}}catch(t){r.e(t)}finally{r.f()}return n||null}},{key:"distinct",value:function(t){for(var e=this._data,i=wo(Sc(e).call(e)),n=[],o=0,r=0,s=i.length;r<s;r++){for(var a=i[r],h=e.get(a)[t],l=!1,d=0;d<o;d++)if(n[d]==h){l=!0;break}l||void 0===h||(n[o]=h,o++)}return n}},{key:"_addItem",value:function(t){var e=function(t,e){return null==t[e]&&(t[e]=Wc()),t}(t,this._idProp),i=e[this._idProp];if(this._data.has(i))throw new Error("Cannot add item: item with id "+i+" already exists");return this._data.set(i,e),++this.length,i}},{key:"_updateItem",value:function(t){var e=t[this._idProp];if(null==e)throw new Error("Cannot update item: item has no id (item: "+es(t)+")");var i=this._data.get(e);if(!i)throw new Error("Cannot update item: no item with id "+e+" found");return this._data.set(e,Vc(Vc({},i),t)),e}},{key:"stream",value:function(t){if(t){var e=this._data;return new Jc(jn({},ic,Hd.mark((function i(){var n,o,r,s;return Hd.wrap((function(i){for(;;)switch(i.prev=i.next){case 0:n=Yc(t),i.prev=1,n.s();case 3:if((o=n.n()).done){i.next=11;break}if(r=o.value,null==(s=e.get(r))){i.next=9;break}return i.next=9,[r,s];case 9:i.next=3;break;case 11:i.next=16;break;case 13:i.prev=13,i.t0=i.catch(1),n.e(i.t0);case 16:return i.prev=16,n.f(),i.finish(16);case 19:case"end":return i.stop()}}),i,null,[[1,13,16,19]])}))))}var i;return new Jc(jn({},ic,Vt(i=Nc(this._data)).call(i,this._data)))}}]),i}(Qc),eu=function(t){Ad(i,t);var e=Uc(i);function i(t,n){var o,r;return Nn(this,i),(r=e.call(this)).length=0,r._ids=new ec,r._options=n||{},r._listener=Vt(o=r._onEvent).call(o,Id(r)),r.setData(t),r}return Fn(i,[{key:"idProp",get:function(){return this.getDataSet().idProp}},{key:"setData",value:function(t){if(this._data){this._data.off&&this._data.off("*",this._listener);var e=this._data.getIds({filter:mr(this._options)}),i=this._data.get(e);this._ids.clear(),this.length=0,this._trigger("remove",{items:e,oldData:i})}if(null!=t){this._data=t;for(var n=this._data.getIds({filter:mr(this._options)}),o=0,r=n.length;o<r;o++){var s=n[o];this._ids.add(s)}this.length=n.length,this._trigger("add",{items:n})}else this._data=new tu;this._data.on&&this._data.on("*",this._listener)}},{key:"refresh",value:function(){for(var t=this._data.getIds({filter:mr(this._options)}),e=wo(this._ids),i={},n=[],o=[],r=[],s=0,a=t.length;s<a;s++){var h=t[s];i[h]=!0,this._ids.has(h)||(n.push(h),this._ids.add(h))}for(var l=0,d=e.length;l<d;l++){var c=e[l],u=this._data.get(c);null==u?console.error("If you see this, report it please."):i[c]||(o.push(c),r.push(u),this._ids.delete(c))}this.length+=n.length-o.length,n.length&&this._trigger("add",{items:n}),o.length&&this._trigger("remove",{items:o,oldData:r})}},{key:"get",value:function(t,e){if(null==this._data)return null;var i,n=null;$c(t)||So(t)?(n=t,i=e):i=t;var o=At({},this._options,i),r=mr(this._options),s=i&&mr(i);return r&&s&&(o.filter=function(t){return r(t)&&s(t)}),null==n?this._data.get(o):this._data.get(n,o)}},{key:"getIds",value:function(t){if(this._data.length){var e,i=mr(this._options),n=null!=t?mr(t):null;return e=n?i?function(t){return i(t)&&n(t)}:n:i,this._data.getIds({filter:e,order:t&&t.order})}return[]}},{key:"forEach",value:function(t,e){if(this._data){var i,n,o=mr(this._options),r=e&&mr(e);n=r?o?function(t){return o(t)&&r(t)}:r:o,Wo(i=this._data).call(i,t,{filter:n,order:e&&e.order})}}},{key:"map",value:function(t,e){if(this._data){var i,n,o=mr(this._options),r=e&&mr(e);return n=r?o?function(t){return o(t)&&r(t)}:r:o,Io(i=this._data).call(i,t,{filter:n,order:e&&e.order})}return[]}},{key:"getDataSet",value:function(){return this._data.getDataSet()}},{key:"stream",value:function(t){var e;return this._data.stream(t||jn({},ic,Vt(e=Sc(this._ids)).call(e,this._ids)))}},{key:"dispose",value:function(){var t;null!==(t=this._data)&&void 0!==t&&t.off&&this._data.off("*",this._listener);var e,n="This data view has already been disposed of.",o={get:function(){throw new Error(n)},set:function(){throw new Error(n)},configurable:!1},r=Yc(Co(i.prototype));try{for(r.s();!(e=r.n()).done;){var s=e.value;zn(this,s,o)}}catch(t){r.e(t)}finally{r.f()}}},{key:"_onEvent",value:function(t,e,i){if(e&&e.items&&this._data){var n=e.items,o=[],r=[],s=[],a=[],h=[],l=[];switch(t){case"add":for(var d=0,c=n.length;d<c;d++){var u=n[d];this.get(u)&&(this._ids.add(u),o.push(u))}break;case"update":for(var f=0,p=n.length;f<p;f++){var v=n[f];this.get(v)?this._ids.has(v)?(r.push(v),h.push(e.data[f]),a.push(e.oldData[f])):(this._ids.add(v),o.push(v)):this._ids.has(v)&&(this._ids.delete(v),s.push(v),l.push(e.oldData[f]))}break;case"remove":for(var g=0,y=n.length;g<y;g++){var m=n[g];this._ids.has(m)&&(this._ids.delete(m),s.push(m),l.push(e.oldData[g]))}}this.length+=o.length-s.length,o.length&&this._trigger("add",{items:o},i),r.length&&this._trigger("update",{items:r,oldData:a,data:h},i),s.length&&this._trigger("remove",{items:s,oldData:l},i)}}}]),i}(Qc);function iu(t,e){return"object"===go(e)&&null!==e&&t===e.idProp&&"function"==typeof e.add&&"function"==typeof e.clear&&"function"==typeof e.distinct&&"function"==typeof Wo(e)&&"function"==typeof e.get&&"function"==typeof e.getDataSet&&"function"==typeof e.getIds&&"number"==typeof e.length&&"function"==typeof Io(e)&&"function"==typeof e.max&&"function"==typeof e.min&&"function"==typeof e.off&&"function"==typeof e.on&&"function"==typeof e.remove&&"function"==typeof e.setOptions&&"function"==typeof e.stream&&"function"==typeof e.update&&"function"==typeof e.updateOnly}function nu(t,e){return"object"===go(e)&&null!==e&&t===e.idProp&&"function"==typeof Wo(e)&&"function"==typeof e.get&&"function"==typeof e.getDataSet&&"function"==typeof e.getIds&&"number"==typeof e.length&&"function"==typeof Io(e)&&"function"==typeof e.off&&"function"==typeof e.on&&"function"==typeof e.stream&&iu(t,e.getDataSet())}var ou=Object.freeze({__proto__:null,DELETE:eh,DataSet:tu,DataStream:Jc,DataView:eu,Queue:Zc,createNewDataPipeFrom:function(t){return new Kc(t)},isDataSetLike:iu,isDataViewLike:nu}),ru=Tr.trim,su=a.parseFloat,au=1/su(xr+"-0")!=-1/0?function(t){var e=ru(Qt(t)),i=su(e);return 0===i&&"-"==e.charAt(0)?-0:i}:su;gt({global:!0,forced:parseFloat!=au},{parseFloat:au});var hu=k.parseFloat,lu=Li.f,du=h((function(){return!Object.getOwnPropertyNames(1)}));gt({target:"Object",stat:!0,forced:du},{getOwnPropertyNames:lu});var cu=k.Object,uu=function(t){return cu.getOwnPropertyNames(t)};function fu(t,e){var i=["node","edge","label"],n=!0,o=Qh(e,"chosen");if("boolean"==typeof o)n=o;else if("object"===go(o)){if(-1===Hr(i).call(i,t))throw new Error("choosify: subOption '"+t+"' should be one of '"+i.join("', '")+"'");var r=Qh(e,["chosen",t]);"boolean"!=typeof r&&"function"!=typeof r||(n=r)}return n}function pu(t,e,i){if(t.width<=0||t.height<=0)return!1;if(void 0!==i){var n={x:e.x-i.x,y:e.y-i.y};if(0!==i.angle){var o=-i.angle;e={x:Math.cos(o)*n.x-Math.sin(o)*n.y,y:Math.sin(o)*n.x+Math.cos(o)*n.y}}else e=n}var r=t.x+t.width,s=t.y+t.width;return t.left<e.x&&r>e.x&&t.top<e.y&&s>e.y}function vu(t){return"string"==typeof t&&""!==t}function gu(t,e,i,n){var o=n.x,r=n.y;if("function"==typeof n.distanceToBorder){var s=n.distanceToBorder(t,e),a=Math.sin(e)*s,h=Math.cos(e)*s;h===s?(o+=s,r=n.y):a===s?(o=n.x,r-=s):(o+=h,r-=a)}else n.shape.width>n.shape.height?(o=n.x+.5*n.shape.width,r=n.y-i):(o=n.x+i,r=n.y-.5*n.shape.height);return{x:o,y:r}}var yu=function(){function t(e){Nn(this,t),this.measureText=e,this.current=0,this.width=0,this.height=0,this.lines=[]}return Fn(t,[{key:"_add",value:function(t,e){var i=arguments.length>2&&void 0!==arguments[2]?arguments[2]:"normal";void 0===this.lines[t]&&(this.lines[t]={width:0,height:0,blocks:[]});var n=e;void 0!==e&&""!==e||(n=" ");var o=this.measureText(n,i),r=At({},Dc(o));r.text=e,r.width=o.width,r.mod=i,void 0!==e&&""!==e||(r.width=0),this.lines[t].blocks.push(r),this.lines[t].width+=r.width}},{key:"curWidth",value:function(){var t=this.lines[this.current];return void 0===t?0:t.width}},{key:"append",value:function(t){var e=arguments.length>1&&void 0!==arguments[1]?arguments[1]:"normal";this._add(this.current,t,e)}},{key:"newLine",value:function(t){var e=arguments.length>1&&void 0!==arguments[1]?arguments[1]:"normal";this._add(this.current,t,e),this.current++}},{key:"determineLineHeights",value:function(){for(var t=0;t<this.lines.length;t++){var e=this.lines[t],i=0;if(void 0!==e.blocks)for(var n=0;n<e.blocks.length;n++){var o=e.blocks[n];i<o.height&&(i=o.height)}e.height=i}}},{key:"determineLabelSize",value:function(){for(var t=0,e=0,i=0;i<this.lines.length;i++){var n=this.lines[i];n.width>t&&(t=n.width),e+=n.height}this.width=t,this.height=e}},{key:"removeEmptyBlocks",value:function(){for(var t=[],e=0;e<this.lines.length;e++){var i=this.lines[e];if(0!==i.blocks.length&&(e!==this.lines.length-1||0!==i.width)){var n={};At(n,i),n.blocks=[];for(var o=void 0,r=[],s=0;s<i.blocks.length;s++){var a=i.blocks[s];0!==a.width?r.push(a):void 0===o&&(o=a)}0===r.length&&void 0!==o&&r.push(o),n.blocks=r,t.push(n)}}return t}},{key:"finalize",value:function(){this.determineLineHeights(),this.determineLabelSize();var t=this.removeEmptyBlocks();return{width:this.width,height:this.height,lines:t}}}]),t}(),mu={"<b>":/<b>/,"<i>":/<i>/,"<code>":/<code>/,"</b>":/<\/b>/,"</i>":/<\/i>/,"</code>":/<\/code>/,"*":/\*/,_:/_/,"`":/`/,afterBold:/[^*]/,afterItal:/[^_]/,afterMono:/[^`]/},bu=function(){function t(e){Nn(this,t),this.text=e,this.bold=!1,this.ital=!1,this.mono=!1,this.spacing=!1,this.position=0,this.buffer="",this.modStack=[],this.blocks=[]}return Fn(t,[{key:"mod",value:function(){return 0===this.modStack.length?"normal":this.modStack[0]}},{key:"modName",value:function(){return 0===this.modStack.length?"normal":"mono"===this.modStack[0]?"mono":this.bold&&this.ital?"boldital":this.bold?"bold":this.ital?"ital":void 0}},{key:"emitBlock",value:function(){this.spacing&&(this.add(" "),this.spacing=!1),this.buffer.length>0&&(this.blocks.push({text:this.buffer,mod:this.modName()}),this.buffer="")}},{key:"add",value:function(t){" "===t&&(this.spacing=!0),this.spacing&&(this.buffer+=" ",this.spacing=!1)," "!=t&&(this.buffer+=t)}},{key:"parseWS",value:function(t){return!!/[ \t]/.test(t)&&(this.mono?this.add(t):this.spacing=!0,!0)}},{key:"setTag",value:function(t){this.emitBlock(),this[t]=!0,this.modStack.unshift(t)}},{key:"unsetTag",value:function(t){this.emitBlock(),this[t]=!1,this.modStack.shift()}},{key:"parseStartTag",value:function(t,e){return!(this.mono||this[t]||!this.match(e))&&(this.setTag(t),!0)}},{key:"match",value:function(t){var e=!(arguments.length>1&&void 0!==arguments[1])||arguments[1],i=this.prepareRegExp(t),n=uo(i,2),o=n[0],r=n[1],s=o.test(this.text.substr(this.position,r));return s&&e&&(this.position+=r-1),s}},{key:"parseEndTag",value:function(t,e,i){var n=this.mod()===t;return!(!(n="mono"===t?n&&this.mono:n&&!this.mono)||!this.match(e))&&(void 0!==i?(this.position===this.text.length-1||this.match(i,!1))&&this.unsetTag(t):this.unsetTag(t),!0)}},{key:"replace",value:function(t,e){return!!this.match(t)&&(this.add(e),this.position+=length-1,!0)}},{key:"prepareRegExp",value:function(t){var e,i;if(t instanceof RegExp)i=t,e=1;else{var n=mu[t];i=void 0!==n?n:new RegExp(t),e=t.length}return[i,e]}}]),t}(),wu=function(){function t(e,i,n,o){var r=this;Nn(this,t),this.ctx=e,this.parent=i,this.selected=n,this.hover=o;this.lines=new yu((function(t,i){if(void 0===t)return 0;var s=r.parent.getFormattingValues(e,n,o,i),a=0;""!==t&&(a=r.ctx.measureText(t).width);return{width:a,values:s}}))}return Fn(t,[{key:"process",value:function(t){if(!vu(t))return this.lines.finalize();var e=this.parent.fontOptions;t=(t=t.replace(/\r\n/g,"\n")).replace(/\r/g,"\n");var i=String(t).split("\n"),n=i.length;if(e.multi)for(var o=0;o<n;o++){var r=this.splitBlocks(i[o],e.multi);if(void 0!==r)if(0!==r.length){if(e.maxWdt>0)for(var s=0;s<r.length;s++){var a=r[s].mod,h=r[s].text;this.splitStringIntoLines(h,a,!0)}else for(var l=0;l<r.length;l++){var d=r[l].mod,c=r[l].text;this.lines.append(c,d)}this.lines.newLine()}else this.lines.newLine("")}else if(e.maxWdt>0)for(var u=0;u<n;u++)this.splitStringIntoLines(i[u]);else for(var f=0;f<n;f++)this.lines.newLine(i[f]);return this.lines.finalize()}},{key:"decodeMarkupSystem",value:function(t){var e="none";return"markdown"===t||"md"===t?e="markdown":!0!==t&&"html"!==t||(e="html"),e}},{key:"splitHtmlBlocks",value:function(t){for(var e=new bu(t),i=function(t){return!!/&/.test(t)&&(e.replace(e.text,"&lt;","<")||e.replace(e.text,"&amp;","&")||e.add("&"),!0)};e.position<e.text.length;){var n=e.text.charAt(e.position);e.parseWS(n)||/</.test(n)&&(e.parseStartTag("bold","<b>")||e.parseStartTag("ital","<i>")||e.parseStartTag("mono","<code>")||e.parseEndTag("bold","</b>")||e.parseEndTag("ital","</i>")||e.parseEndTag("mono","</code>"))||i(n)||e.add(n),e.position++}return e.emitBlock(),e.blocks}},{key:"splitMarkdownBlocks",value:function(t){for(var e=this,i=new bu(t),n=!0,o=function(t){return!!/\\/.test(t)&&(i.position<e.text.length+1&&(i.position++,t=e.text.charAt(i.position),/ \t/.test(t)?i.spacing=!0:(i.add(t),n=!1)),!0)};i.position<i.text.length;){var r=i.text.charAt(i.position);i.parseWS(r)||o(r)||(n||i.spacing)&&(i.parseStartTag("bold","*")||i.parseStartTag("ital","_")||i.parseStartTag("mono","`"))||i.parseEndTag("bold","*","afterBold")||i.parseEndTag("ital","_","afterItal")||i.parseEndTag("mono","`","afterMono")||(i.add(r),n=!1),i.position++}return i.emitBlock(),i.blocks}},{key:"splitBlocks",value:function(t,e){var i=this.decodeMarkupSystem(e);return"none"===i?[{text:t,mod:"normal"}]:"markdown"===i?this.splitMarkdownBlocks(t):"html"===i?this.splitHtmlBlocks(t):void 0}},{key:"overMaxWidth",value:function(t){var e=this.ctx.measureText(t).width;return this.lines.curWidth()+e>this.parent.fontOptions.maxWdt}},{key:"getLongestFit",value:function(t){for(var e="",i=0;i<t.length;){var n=e+(""===e?"":" ")+t[i];if(this.overMaxWidth(n))break;e=n,i++}return i}},{key:"getLongestFitWord",value:function(t){for(var e=0;e<t.length&&!this.overMaxWidth(Oo(t).call(t,0,e));)e++;return e}},{key:"splitStringIntoLines",value:function(t){var e=arguments.length>1&&void 0!==arguments[1]?arguments[1]:"normal",i=arguments.length>2&&void 0!==arguments[2]&&arguments[2];this.parent.getFormattingValues(this.ctx,this.selected,this.hover,e);for(var n=(t=(t=t.replace(/^( +)/g,"$1\r")).replace(/([^\r][^ ]*)( +)/g,"$1\r$2\r")).split("\r");n.length>0;){var o=this.getLongestFit(n);if(0===o){var r=n[0],s=this.getLongestFitWord(r);this.lines.newLine(Oo(r).call(r,0,s),e),n[0]=Oo(r).call(r,s)}else{var a=o;" "===n[o-1]?o--:" "===n[a]&&a++;var h=Oo(n).call(n,0,o).join("");o==n.length&&i?this.lines.append(h,e):this.lines.newLine(h,e),n=Oo(n).call(n,a)}}}}]),t}(),ku=["bold","ital","boldital","mono"],_u=function(){function t(e,i){var n=arguments.length>2&&void 0!==arguments[2]&&arguments[2];Nn(this,t),this.body=e,this.pointToSelf=!1,this.baseSize=void 0,this.fontOptions={},this.setOptions(i),this.size={top:0,left:0,width:0,height:0,yLine:0},this.isEdgeLabel=n}return Fn(t,[{key:"setOptions",value:function(t){if(this.elementOptions=t,this.initFontOptions(t.font),vu(t.label)?this.labelDirty=!0:t.label=void 0,void 0!==t.font&&null!==t.font)if("string"==typeof t.font)this.baseSize=this.fontOptions.size;else if("object"===go(t.font)){var e=t.font.size;void 0!==e&&(this.baseSize=e)}}},{key:"initFontOptions",value:function(e){var i=this;Dh(ku,(function(t){i.fontOptions[t]={}})),t.parseFontString(this.fontOptions,e)?this.fontOptions.vadjust=0:Dh(e,(function(t,e){null!=t&&"object"!==go(t)&&(i.fontOptions[e]=t)}))}},{key:"constrain",value:function(t){var e={constrainWidth:!1,maxWdt:-1,minWdt:-1,constrainHeight:!1,minHgt:-1,valign:"middle"},i=Qh(t,"widthConstraint");if("number"==typeof i)e.maxWdt=Number(i),e.minWdt=Number(i);else if("object"===go(i)){var n=Qh(t,["widthConstraint","maximum"]);"number"==typeof n&&(e.maxWdt=Number(n));var o=Qh(t,["widthConstraint","minimum"]);"number"==typeof o&&(e.minWdt=Number(o))}var r=Qh(t,"heightConstraint");if("number"==typeof r)e.minHgt=Number(r);else if("object"===go(r)){var s=Qh(t,["heightConstraint","minimum"]);"number"==typeof s&&(e.minHgt=Number(s));var a=Qh(t,["heightConstraint","valign"]);"string"==typeof a&&("top"!==a&&"bottom"!==a||(e.valign=a))}return e}},{key:"update",value:function(t,e){this.setOptions(t,!0),this.propagateFonts(e),Ch(this.fontOptions,this.constrain(e)),this.fontOptions.chooser=fu("label",e)}},{key:"adjustSizes",value:function(t){var e=t?t.right+t.left:0;this.fontOptions.constrainWidth&&(this.fontOptions.maxWdt-=e,this.fontOptions.minWdt-=e);var i=t?t.top+t.bottom:0;this.fontOptions.constrainHeight&&(this.fontOptions.minHgt-=i)}},{key:"addFontOptionsToPile",value:function(t,e){for(var i=0;i<e.length;++i)this.addFontToPile(t,e[i])}},{key:"addFontToPile",value:function(t,e){if(void 0!==e&&void 0!==e.font&&null!==e.font){var i=e.font;t.push(i)}}},{key:"getBasicOptions",value:function(e){for(var i={},n=0;n<e.length;++n){var o=e[n],r={};t.parseFontString(r,o)&&(o=r),Dh(o,(function(t,e){void 0!==t&&(Object.prototype.hasOwnProperty.call(i,e)||(-1!==Hr(ku).call(ku,e)?i[e]={}:i[e]=t))}))}return i}},{key:"getFontOption",value:function(e,i,n){for(var o,r=0;r<e.length;++r){var s=e[r];if(Object.prototype.hasOwnProperty.call(s,i)){if(null==(o=s[i]))continue;var a={};if(t.parseFontString(a,o)&&(o=a),Object.prototype.hasOwnProperty.call(o,n))return o[n]}}if(Object.prototype.hasOwnProperty.call(this.fontOptions,n))return this.fontOptions[n];throw new Error("Did not find value for multi-font for property: '"+n+"'")}},{key:"getFontOptions",value:function(t,e){for(var i={},n=["color","size","face","mod","vadjust"],o=0;o<n.length;++o){var r=n[o];i[r]=this.getFontOption(t,e,r)}return i}},{key:"propagateFonts",value:function(t){var e=this,i=[];this.addFontOptionsToPile(i,t),this.fontOptions=this.getBasicOptions(i);for(var n=function(t){var n=ku[t],o=e.fontOptions[n];Dh(e.getFontOptions(i,n),(function(t,e){o[e]=t})),o.size=Number(o.size),o.vadjust=Number(o.vadjust)},o=0;o<ku.length;++o)n(o)}},{key:"draw",value:function(t,e,i,n,o){var r=arguments.length>5&&void 0!==arguments[5]?arguments[5]:"middle";if(void 0!==this.elementOptions.label){var s=this.fontOptions.size*this.body.view.scale;this.elementOptions.label&&s<this.elementOptions.scaling.label.drawThreshold-1||(s>=this.elementOptions.scaling.label.maxVisible&&(s=Number(this.elementOptions.scaling.label.maxVisible)/this.body.view.scale),this.calculateLabelSize(t,n,o,e,i,r),this._drawBackground(t),this._drawText(t,e,this.size.yLine,r,s))}}},{key:"_drawBackground",value:function(t){if(void 0!==this.fontOptions.background&&"none"!==this.fontOptions.background){t.fillStyle=this.fontOptions.background;var e=this.getSize();t.fillRect(e.left,e.top,e.width,e.height)}}},{key:"_drawText",value:function(t,e,i){var n=arguments.length>3&&void 0!==arguments[3]?arguments[3]:"middle",o=arguments.length>4?arguments[4]:void 0,r=this._setAlignment(t,e,i,n),s=uo(r,2);e=s[0],i=s[1],t.textAlign="left",e-=this.size.width/2,this.fontOptions.valign&&this.size.height>this.size.labelHeight&&("top"===this.fontOptions.valign&&(i-=(this.size.height-this.size.labelHeight)/2),"bottom"===this.fontOptions.valign&&(i+=(this.size.height-this.size.labelHeight)/2));for(var a=0;a<this.lineCount;a++){var h=this.lines[a];if(h&&h.blocks){var l=0;this.isEdgeLabel||"center"===this.fontOptions.align?l+=(this.size.width-h.width)/2:"right"===this.fontOptions.align&&(l+=this.size.width-h.width);for(var d=0;d<h.blocks.length;d++){var c=h.blocks[d];t.font=c.font;var u=this._getColor(c.color,o,c.strokeColor),f=uo(u,2),p=f[0],v=f[1];c.strokeWidth>0&&(t.lineWidth=c.strokeWidth,t.strokeStyle=v,t.lineJoin="round"),t.fillStyle=p,c.strokeWidth>0&&t.strokeText(c.text,e+l,i+c.vadjust),t.fillText(c.text,e+l,i+c.vadjust),l+=c.width}i+=h.height}}}},{key:"_setAlignment",value:function(t,e,i,n){if(this.isEdgeLabel&&"horizontal"!==this.fontOptions.align&&!1===this.pointToSelf){e=0,i=0;"top"===this.fontOptions.align?(t.textBaseline="alphabetic",i-=4):"bottom"===this.fontOptions.align?(t.textBaseline="hanging",i+=4):t.textBaseline="middle"}else t.textBaseline=n;return[e,i]}},{key:"_getColor",value:function(t,e,i){var n=t||"#000000",o=i||"#ffffff";if(e<=this.elementOptions.scaling.label.drawThreshold){var r=Math.max(0,Math.min(1,1-(this.elementOptions.scaling.label.drawThreshold-e)));n=Fh(n,r),o=Fh(o,r)}return[n,o]}},{key:"getTextSize",value:function(t){var e=arguments.length>1&&void 0!==arguments[1]&&arguments[1],i=arguments.length>2&&void 0!==arguments[2]&&arguments[2];return this._processLabel(t,e,i),{width:this.size.width,height:this.size.height,lineCount:this.lineCount}}},{key:"getSize",value:function(){var t=this.size.left,e=this.size.top-1;if(this.isEdgeLabel){var i=.5*-this.size.width;switch(this.fontOptions.align){case"middle":t=i,e=.5*-this.size.height;break;case"top":t=i,e=-(this.size.height+2);break;case"bottom":t=i,e=2}}return{left:t,top:e,width:this.size.width,height:this.size.height}}},{key:"calculateLabelSize",value:function(t,e,i){var n=arguments.length>3&&void 0!==arguments[3]?arguments[3]:0,o=arguments.length>4&&void 0!==arguments[4]?arguments[4]:0,r=arguments.length>5&&void 0!==arguments[5]?arguments[5]:"middle";this._processLabel(t,e,i),this.size.left=n-.5*this.size.width,this.size.top=o-.5*this.size.height,this.size.yLine=o+.5*(1-this.lineCount)*this.fontOptions.size,"hanging"===r&&(this.size.top+=.5*this.fontOptions.size,this.size.top+=4,this.size.yLine+=4)}},{key:"getFormattingValues",value:function(t,e,i,n){var o=function(t,e,i){return"normal"===e?"mod"===i?"":t[i]:void 0!==t[e][i]?t[e][i]:t[i]},r={color:o(this.fontOptions,n,"color"),size:o(this.fontOptions,n,"size"),face:o(this.fontOptions,n,"face"),mod:o(this.fontOptions,n,"mod"),vadjust:o(this.fontOptions,n,"vadjust"),strokeWidth:this.fontOptions.strokeWidth,strokeColor:this.fontOptions.strokeColor};(e||i)&&("normal"===n&&!0===this.fontOptions.chooser&&this.elementOptions.labelHighlightBold?r.mod="bold":"function"==typeof this.fontOptions.chooser&&this.fontOptions.chooser(r,this.elementOptions.id,e,i));var s="";return void 0!==r.mod&&""!==r.mod&&(s+=r.mod+" "),s+=r.size+"px "+r.face,t.font=s.replace(/"/g,""),r.font=t.font,r.height=r.size,r}},{key:"differentState",value:function(t,e){return t!==this.selectedState||e!==this.hoverState}},{key:"_processLabelText",value:function(t,e,i,n){return new wu(t,this,e,i).process(n)}},{key:"_processLabel",value:function(t,e,i){if(!1!==this.labelDirty||this.differentState(e,i)){var n=this._processLabelText(t,e,i,this.elementOptions.label);this.fontOptions.minWdt>0&&n.width<this.fontOptions.minWdt&&(n.width=this.fontOptions.minWdt),this.size.labelHeight=n.height,this.fontOptions.minHgt>0&&n.height<this.fontOptions.minHgt&&(n.height=this.fontOptions.minHgt),this.lines=n.lines,this.lineCount=n.lines.length,this.size.width=n.width,this.size.height=n.height,this.selectedState=e,this.hoverState=i,this.labelDirty=!1}}},{key:"visible",value:function(){return 0!==this.size.width&&0!==this.size.height&&void 0!==this.elementOptions.label&&!(this.fontOptions.size*this.body.view.scale<this.elementOptions.scaling.label.drawThreshold-1)}}],[{key:"parseFontString",value:function(t,e){if(!e||"string"!=typeof e)return!1;var i=e.split(" ");return t.size=+i[0].replace("px",""),t.face=i[1],t.color=i[2],!0}}]),t}(),xu=function(){function t(e,i,n){Nn(this,t),this.body=i,this.labelModule=n,this.setOptions(e),this.top=void 0,this.left=void 0,this.height=void 0,this.width=void 0,this.radius=void 0,this.margin=void 0,this.refreshNeeded=!0,this.boundingBox={top:0,left:0,right:0,bottom:0}}return Fn(t,[{key:"setOptions",value:function(t){this.options=t}},{key:"_setMargins",value:function(t){this.margin={},this.options.margin&&("object"==go(this.options.margin)?(this.margin.top=this.options.margin.top,this.margin.right=this.options.margin.right,this.margin.bottom=this.options.margin.bottom,this.margin.left=this.options.margin.left):(this.margin.top=this.options.margin,this.margin.right=this.options.margin,this.margin.bottom=this.options.margin,this.margin.left=this.options.margin)),t.adjustSizes(this.margin)}},{key:"_distanceToBorder",value:function(t,e){var i=this.options.borderWidth;return t&&this.resize(t),Math.min(Math.abs(this.width/2/Math.cos(e)),Math.abs(this.height/2/Math.sin(e)))+i}},{key:"enableShadow",value:function(t,e){e.shadow&&(t.shadowColor=e.shadowColor,t.shadowBlur=e.shadowSize,t.shadowOffsetX=e.shadowX,t.shadowOffsetY=e.shadowY)}},{key:"disableShadow",value:function(t,e){e.shadow&&(t.shadowColor="rgba(0,0,0,0)",t.shadowBlur=0,t.shadowOffsetX=0,t.shadowOffsetY=0)}},{key:"enableBorderDashes",value:function(t,e){if(!1!==e.borderDashes)if(void 0!==t.setLineDash){var i=e.borderDashes;!0===i&&(i=[5,15]),t.setLineDash(i)}else console.warn("setLineDash is not supported in this browser. The dashed borders cannot be used."),this.options.shapeProperties.borderDashes=!1,e.borderDashes=!1}},{key:"disableBorderDashes",value:function(t,e){!1!==e.borderDashes&&(void 0!==t.setLineDash?t.setLineDash([0]):(console.warn("setLineDash is not supported in this browser. The dashed borders cannot be used."),this.options.shapeProperties.borderDashes=!1,e.borderDashes=!1))}},{key:"needsRefresh",value:function(t,e){return!0===this.refreshNeeded?(this.refreshNeeded=!1,!0):void 0===this.width||this.labelModule.differentState(t,e)}},{key:"initContextForDraw",value:function(t,e){var i=e.borderWidth/this.body.view.scale;t.lineWidth=Math.min(this.width,i),t.strokeStyle=e.borderColor,t.fillStyle=e.color}},{key:"performStroke",value:function(t,e){var i=e.borderWidth/this.body.view.scale;t.save(),i>0&&(this.enableBorderDashes(t,e),t.stroke(),this.disableBorderDashes(t,e)),t.restore()}},{key:"performFill",value:function(t,e){t.save(),t.fillStyle=e.color,this.enableShadow(t,e),hs(t).call(t),this.disableShadow(t,e),t.restore(),this.performStroke(t,e)}},{key:"_addBoundingBoxMargin",value:function(t){this.boundingBox.left-=t,this.boundingBox.top-=t,this.boundingBox.bottom+=t,this.boundingBox.right+=t}},{key:"_updateBoundingBox",value:function(t,e,i,n,o){void 0!==i&&this.resize(i,n,o),this.left=t-this.width/2,this.top=e-this.height/2,this.boundingBox.left=this.left,this.boundingBox.top=this.top,this.boundingBox.bottom=this.top+this.height,this.boundingBox.right=this.left+this.width}},{key:"updateBoundingBox",value:function(t,e,i,n,o){this._updateBoundingBox(t,e,i,n,o)}},{key:"getDimensionsFromLabel",value:function(t,e,i){this.textSize=this.labelModule.getTextSize(t,e,i);var n=this.textSize.width,o=this.textSize.height;return 0===n&&(n=14,o=14),{width:n,height:o}}}]),t}();function Eu(t){var e=function(){if("undefined"==typeof Reflect||!Pd)return!1;if(Pd.sham)return!1;if("function"==typeof Proxy)return!0;try{return Boolean.prototype.valueOf.call(Pd(Boolean,[],(function(){}))),!0}catch(t){return!1}}();return function(){var i,n=Ld(t);if(e){var o=Ld(this).constructor;i=Pd(n,arguments,o)}else i=n.apply(this,arguments);return Fd(this,i)}}var Ou=function(t){Ad(i,t);var e=Eu(i);function i(t,n,o){var r;return Nn(this,i),(r=e.call(this,t,n,o))._setMargins(o),r}return Fn(i,[{key:"resize",value:function(t){var e=arguments.length>1&&void 0!==arguments[1]?arguments[1]:this.selected,i=arguments.length>2&&void 0!==arguments[2]?arguments[2]:this.hover;if(this.needsRefresh(e,i)){var n=this.getDimensionsFromLabel(t,e,i);this.width=n.width+this.margin.right+this.margin.left,this.height=n.height+this.margin.top+this.margin.bottom,this.radius=this.width/2}}},{key:"draw",value:function(t,e,i,n,o,r){this.resize(t,n,o),this.left=e-this.width/2,this.top=i-this.height/2,this.initContextForDraw(t,r),Yt(t,this.left,this.top,this.width,this.height,r.borderRadius),this.performFill(t,r),this.updateBoundingBox(e,i,t,n,o),this.labelModule.draw(t,this.left+this.textSize.width/2+this.margin.left,this.top+this.textSize.height/2+this.margin.top,n,o)}},{key:"updateBoundingBox",value:function(t,e,i,n,o){this._updateBoundingBox(t,e,i,n,o);var r=this.options.shapeProperties.borderRadius;this._addBoundingBoxMargin(r)}},{key:"distanceToBorder",value:function(t,e){t&&this.resize(t);var i=this.options.borderWidth;return Math.min(Math.abs(this.width/2/Math.cos(e)),Math.abs(this.height/2/Math.sin(e)))+i}}]),i}(xu);function Cu(t){var e=function(){if("undefined"==typeof Reflect||!Pd)return!1;if(Pd.sham)return!1;if("function"==typeof Proxy)return!0;try{return Boolean.prototype.valueOf.call(Pd(Boolean,[],(function(){}))),!0}catch(t){return!1}}();return function(){var i,n=Ld(t);if(e){var o=Ld(this).constructor;i=Pd(n,arguments,o)}else i=n.apply(this,arguments);return Fd(this,i)}}var Su=function(t){Ad(i,t);var e=Cu(i);function i(t,n,o){var r;return Nn(this,i),(r=e.call(this,t,n,o)).labelOffset=0,r.selected=!1,r}return Fn(i,[{key:"setOptions",value:function(t,e,i){this.options=t,void 0===e&&void 0===i||this.setImages(e,i)}},{key:"setImages",value:function(t,e){e&&this.selected?(this.imageObj=e,this.imageObjAlt=t):(this.imageObj=t,this.imageObjAlt=e)}},{key:"switchImages",value:function(t){var e=t&&!this.selected||!t&&this.selected;if(this.selected=t,void 0!==this.imageObjAlt&&e){var i=this.imageObj;this.imageObj=this.imageObjAlt,this.imageObjAlt=i}}},{key:"_getImagePadding",value:function(){var t={top:0,right:0,bottom:0,left:0};if(this.options.imagePadding){var e=this.options.imagePadding;"object"==go(e)?(t.top=e.top,t.right=e.right,t.bottom=e.bottom,t.left=e.left):(t.top=e,t.right=e,t.bottom=e,t.left=e)}return t}},{key:"_resizeImage",value:function(){var t,e;if(!1===this.options.shapeProperties.useImageSize){var i=1,n=1;this.imageObj.width&&this.imageObj.height&&(this.imageObj.width>this.imageObj.height?i=this.imageObj.width/this.imageObj.height:n=this.imageObj.height/this.imageObj.width),t=2*this.options.size*i,e=2*this.options.size*n}else{var o=this._getImagePadding();t=this.imageObj.width+o.left+o.right,e=this.imageObj.height+o.top+o.bottom}this.width=t,this.height=e,this.radius=.5*this.width}},{key:"_drawRawCircle",value:function(t,e,i,n){this.initContextForDraw(t,n),Ut(t,e,i,n.size),this.performFill(t,n)}},{key:"_drawImageAtPosition",value:function(t,e){if(0!=this.imageObj.width){t.globalAlpha=void 0!==e.opacity?e.opacity:1,this.enableShadow(t,e);var i=1;!0===this.options.shapeProperties.interpolation&&(i=this.imageObj.width/this.width/this.body.view.scale);var n=this._getImagePadding(),o=this.left+n.left,r=this.top+n.top,s=this.width-n.left-n.right,a=this.height-n.top-n.bottom;this.imageObj.drawImageAtPosition(t,i,o,r,s,a),this.disableShadow(t,e)}}},{key:"_drawImageLabel",value:function(t,e,i,n,o){var r=0;if(void 0!==this.height){r=.5*this.height;var s=this.labelModule.getTextSize(t,n,o);s.lineCount>=1&&(r+=s.height/2)}var a=i+r;this.options.label&&(this.labelOffset=r),this.labelModule.draw(t,e,a,n,o,"hanging")}}]),i}(xu);function Tu(t){var e=function(){if("undefined"==typeof Reflect||!Pd)return!1;if(Pd.sham)return!1;if("function"==typeof Proxy)return!0;try{return Boolean.prototype.valueOf.call(Pd(Boolean,[],(function(){}))),!0}catch(t){return!1}}();return function(){var i,n=Ld(t);if(e){var o=Ld(this).constructor;i=Pd(n,arguments,o)}else i=n.apply(this,arguments);return Fd(this,i)}}var Mu=function(t){Ad(i,t);var e=Tu(i);function i(t,n,o){var r;return Nn(this,i),(r=e.call(this,t,n,o))._setMargins(o),r}return Fn(i,[{key:"resize",value:function(t){var e=arguments.length>1&&void 0!==arguments[1]?arguments[1]:this.selected,i=arguments.length>2&&void 0!==arguments[2]?arguments[2]:this.hover;if(this.needsRefresh(e,i)){var n=this.getDimensionsFromLabel(t,e,i),o=Math.max(n.width+this.margin.right+this.margin.left,n.height+this.margin.top+this.margin.bottom);this.options.size=o/2,this.width=o,this.height=o,this.radius=this.width/2}}},{key:"draw",value:function(t,e,i,n,o,r){this.resize(t,n,o),this.left=e-this.width/2,this.top=i-this.height/2,this._drawRawCircle(t,e,i,r),this.updateBoundingBox(e,i),this.labelModule.draw(t,this.left+this.textSize.width/2+this.margin.left,i,n,o)}},{key:"updateBoundingBox",value:function(t,e){this.boundingBox.top=e-this.options.size,this.boundingBox.left=t-this.options.size,this.boundingBox.right=t+this.options.size,this.boundingBox.bottom=e+this.options.size}},{key:"distanceToBorder",value:function(t){return t&&this.resize(t),.5*this.width}}]),i}(Su);function Pu(t){var e=function(){if("undefined"==typeof Reflect||!Pd)return!1;if(Pd.sham)return!1;if("function"==typeof Proxy)return!0;try{return Boolean.prototype.valueOf.call(Pd(Boolean,[],(function(){}))),!0}catch(t){return!1}}();return function(){var i,n=Ld(t);if(e){var o=Ld(this).constructor;i=Pd(n,arguments,o)}else i=n.apply(this,arguments);return Fd(this,i)}}var Du=function(t){Ad(i,t);var e=Pu(i);function i(t,n,o,r,s){var a;return Nn(this,i),(a=e.call(this,t,n,o)).setImages(r,s),a}return Fn(i,[{key:"resize",value:function(t){var e=arguments.length>1&&void 0!==arguments[1]?arguments[1]:this.selected,i=arguments.length>2&&void 0!==arguments[2]?arguments[2]:this.hover,n=void 0===this.imageObj.src||void 0===this.imageObj.width||void 0===this.imageObj.height;if(n){var o=2*this.options.size;return this.width=o,this.height=o,void(this.radius=.5*this.width)}this.needsRefresh(e,i)&&this._resizeImage()}},{key:"draw",value:function(t,e,i,n,o,r){this.switchImages(n),this.resize();var s=e,a=i;"top-left"===this.options.shapeProperties.coordinateOrigin?(this.left=e,this.top=i,s+=this.width/2,a+=this.height/2):(this.left=e-this.width/2,this.top=i-this.height/2),this._drawRawCircle(t,s,a,r),t.save(),t.clip(),this._drawImageAtPosition(t,r),t.restore(),this._drawImageLabel(t,s,a,n,o),this.updateBoundingBox(e,i)}},{key:"updateBoundingBox",value:function(t,e){"top-left"===this.options.shapeProperties.coordinateOrigin?(this.boundingBox.top=e,this.boundingBox.left=t,this.boundingBox.right=t+2*this.options.size,this.boundingBox.bottom=e+2*this.options.size):(this.boundingBox.top=e-this.options.size,this.boundingBox.left=t-this.options.size,this.boundingBox.right=t+this.options.size,this.boundingBox.bottom=e+this.options.size),this.boundingBox.left=Math.min(this.boundingBox.left,this.labelModule.size.left),this.boundingBox.right=Math.max(this.boundingBox.right,this.labelModule.size.left+this.labelModule.size.width),this.boundingBox.bottom=Math.max(this.boundingBox.bottom,this.boundingBox.bottom+this.labelOffset)}},{key:"distanceToBorder",value:function(t){return t&&this.resize(t),.5*this.width}}]),i}(Su);function Iu(t){var e=function(){if("undefined"==typeof Reflect||!Pd)return!1;if(Pd.sham)return!1;if("function"==typeof Proxy)return!0;try{return Boolean.prototype.valueOf.call(Pd(Boolean,[],(function(){}))),!0}catch(t){return!1}}();return function(){var i,n=Ld(t);if(e){var o=Ld(this).constructor;i=Pd(n,arguments,o)}else i=n.apply(this,arguments);return Fd(this,i)}}var Bu=function(t){Ad(i,t);var e=Iu(i);function i(t,n,o){return Nn(this,i),e.call(this,t,n,o)}return Fn(i,[{key:"resize",value:function(t){var e=arguments.length>1&&void 0!==arguments[1]?arguments[1]:this.selected,i=arguments.length>2&&void 0!==arguments[2]?arguments[2]:this.hover,n=arguments.length>3&&void 0!==arguments[3]?arguments[3]:{size:this.options.size};if(this.needsRefresh(e,i)){var o,r;this.labelModule.getTextSize(t,e,i);var s=2*n.size;this.width=null!==(o=this.customSizeWidth)&&void 0!==o?o:s,this.height=null!==(r=this.customSizeHeight)&&void 0!==r?r:s,this.radius=.5*this.width}}},{key:"_drawShape",value:function(t,e,i,n,o,r,s,a){var h,l=this;return this.resize(t,r,s,a),this.left=n-this.width/2,this.top=o-this.height/2,this.initContextForDraw(t,a),(h=e,Object.prototype.hasOwnProperty.call($t,h)?$t[h]:function(t){for(var e=arguments.length,i=new Array(e>1?e-1:0),n=1;n<e;n++)i[n-1]=arguments[n];CanvasRenderingContext2D.prototype[h].call(t,i)})(t,n,o,a.size),this.performFill(t,a),void 0!==this.options.icon&&void 0!==this.options.icon.code&&(t.font=(r?"bold ":"")+this.height/2+"px "+(this.options.icon.face||"FontAwesome"),t.fillStyle=this.options.icon.color||"black",t.textAlign="center",t.textBaseline="middle",t.fillText(this.options.icon.code,n,o)),{drawExternalLabel:function(){if(void 0!==l.options.label){l.labelModule.calculateLabelSize(t,r,s,n,o,"hanging");var e=o+.5*l.height+.5*l.labelModule.size.height;l.labelModule.draw(t,n,e,r,s,"hanging")}l.updateBoundingBox(n,o)}}}},{key:"updateBoundingBox",value:function(t,e){this.boundingBox.top=e-this.options.size,this.boundingBox.left=t-this.options.size,this.boundingBox.right=t+this.options.size,this.boundingBox.bottom=e+this.options.size,void 0!==this.options.label&&this.labelModule.size.width>0&&(this.boundingBox.left=Math.min(this.boundingBox.left,this.labelModule.size.left),this.boundingBox.right=Math.max(this.boundingBox.right,this.labelModule.size.left+this.labelModule.size.width),this.boundingBox.bottom=Math.max(this.boundingBox.bottom,this.boundingBox.bottom+this.labelModule.size.height))}}]),i}(xu);function zu(t,e){var i=zo(t);if(On){var n=On(t);e&&(n=mr(n).call(n,(function(e){return Mn(t,e).enumerable}))),i.push.apply(i,n)}return i}function Nu(t){for(var e=1;e<arguments.length;e++){var i,n=null!=arguments[e]?arguments[e]:{};if(e%2)Wo(i=zu(Object(n),!0)).call(i,(function(e){jn(t,e,n[e])}));else if(Dn)In(t,Dn(n));else{var o;Wo(o=zu(Object(n))).call(o,(function(e){zn(t,e,Mn(n,e))}))}}return t}function Au(t){var e=function(){if("undefined"==typeof Reflect||!Pd)return!1;if(Pd.sham)return!1;if("function"==typeof Proxy)return!0;try{return Boolean.prototype.valueOf.call(Pd(Boolean,[],(function(){}))),!0}catch(t){return!1}}();return function(){var i,n=Ld(t);if(e){var o=Ld(this).constructor;i=Pd(n,arguments,o)}else i=n.apply(this,arguments);return Fd(this,i)}}var Fu=function(t){Ad(i,t);var e=Au(i);function i(t,n,o,r){var s;return Nn(this,i),(s=e.call(this,t,n,o,r)).ctxRenderer=r,s}return Fn(i,[{key:"draw",value:function(t,e,i,n,o,r){this.resize(t,n,o,r),this.left=e-this.width/2,this.top=i-this.height/2,t.save();var s=this.ctxRenderer({ctx:t,id:this.options.id,x:e,y:i,state:{selected:n,hover:o},style:Nu({},r),label:this.options.label});if(null!=s.drawNode&&s.drawNode(),t.restore(),s.drawExternalLabel){var a=s.drawExternalLabel;s.drawExternalLabel=function(){t.save(),a(),t.restore()}}return s.nodeDimensions&&(this.customSizeWidth=s.nodeDimensions.width,this.customSizeHeight=s.nodeDimensions.height),s}},{key:"distanceToBorder",value:function(t,e){return this._distanceToBorder(t,e)}}]),i}(Bu);function ju(t){var e=function(){if("undefined"==typeof Reflect||!Pd)return!1;if(Pd.sham)return!1;if("function"==typeof Proxy)return!0;try{return Boolean.prototype.valueOf.call(Pd(Boolean,[],(function(){}))),!0}catch(t){return!1}}();return function(){var i,n=Ld(t);if(e){var o=Ld(this).constructor;i=Pd(n,arguments,o)}else i=n.apply(this,arguments);return Fd(this,i)}}var Ru=function(t){Ad(i,t);var e=ju(i);function i(t,n,o){var r;return Nn(this,i),(r=e.call(this,t,n,o))._setMargins(o),r}return Fn(i,[{key:"resize",value:function(t,e,i){if(this.needsRefresh(e,i)){var n=this.getDimensionsFromLabel(t,e,i).width+this.margin.right+this.margin.left;this.width=n,this.height=n,this.radius=this.width/2}}},{key:"draw",value:function(t,e,i,n,o,r){this.resize(t,n,o),this.left=e-this.width/2,this.top=i-this.height/2,this.initContextForDraw(t,r),Gt(t,e-this.width/2,i-this.height/2,this.width,this.height),this.performFill(t,r),this.updateBoundingBox(e,i,t,n,o),this.labelModule.draw(t,this.left+this.textSize.width/2+this.margin.left,this.top+this.textSize.height/2+this.margin.top,n,o)}},{key:"distanceToBorder",value:function(t,e){return this._distanceToBorder(t,e)}}]),i}(xu);function Lu(t){var e=function(){if("undefined"==typeof Reflect||!Pd)return!1;if(Pd.sham)return!1;if("function"==typeof Proxy)return!0;try{return Boolean.prototype.valueOf.call(Pd(Boolean,[],(function(){}))),!0}catch(t){return!1}}();return function(){var i,n=Ld(t);if(e){var o=Ld(this).constructor;i=Pd(n,arguments,o)}else i=n.apply(this,arguments);return Fd(this,i)}}var Hu=function(t){Ad(i,t);var e=Lu(i);function i(t,n,o){return Nn(this,i),e.call(this,t,n,o)}return Fn(i,[{key:"draw",value:function(t,e,i,n,o,r){return this._drawShape(t,"diamond",4,e,i,n,o,r)}},{key:"distanceToBorder",value:function(t,e){return this._distanceToBorder(t,e)}}]),i}(Bu);function Wu(t){var e=function(){if("undefined"==typeof Reflect||!Pd)return!1;if(Pd.sham)return!1;if("function"==typeof Proxy)return!0;try{return Boolean.prototype.valueOf.call(Pd(Boolean,[],(function(){}))),!0}catch(t){return!1}}();return function(){var i,n=Ld(t);if(e){var o=Ld(this).constructor;i=Pd(n,arguments,o)}else i=n.apply(this,arguments);return Fd(this,i)}}var qu=function(t){Ad(i,t);var e=Wu(i);function i(t,n,o){return Nn(this,i),e.call(this,t,n,o)}return Fn(i,[{key:"draw",value:function(t,e,i,n,o,r){return this._drawShape(t,"circle",2,e,i,n,o,r)}},{key:"distanceToBorder",value:function(t){return t&&this.resize(t),this.options.size}}]),i}(Bu);function Vu(t){var e=function(){if("undefined"==typeof Reflect||!Pd)return!1;if(Pd.sham)return!1;if("function"==typeof Proxy)return!0;try{return Boolean.prototype.valueOf.call(Pd(Boolean,[],(function(){}))),!0}catch(t){return!1}}();return function(){var i,n=Ld(t);if(e){var o=Ld(this).constructor;i=Pd(n,arguments,o)}else i=n.apply(this,arguments);return Fd(this,i)}}var Uu=function(t){Ad(i,t);var e=Vu(i);function i(t,n,o){return Nn(this,i),e.call(this,t,n,o)}return Fn(i,[{key:"resize",value:function(t){var e=arguments.length>1&&void 0!==arguments[1]?arguments[1]:this.selected,i=arguments.length>2&&void 0!==arguments[2]?arguments[2]:this.hover;if(this.needsRefresh(e,i)){var n=this.getDimensionsFromLabel(t,e,i);this.height=2*n.height,this.width=n.width+n.height,this.radius=.5*this.width}}},{key:"draw",value:function(t,e,i,n,o,r){this.resize(t,n,o),this.left=e-.5*this.width,this.top=i-.5*this.height,this.initContextForDraw(t,r),Xt(t,this.left,this.top,this.width,this.height),this.performFill(t,r),this.updateBoundingBox(e,i,t,n,o),this.labelModule.draw(t,e,i,n,o)}},{key:"distanceToBorder",value:function(t,e){t&&this.resize(t);var i=.5*this.width,n=.5*this.height,o=Math.sin(e)*i,r=Math.cos(e)*n;return i*n/Math.sqrt(o*o+r*r)}}]),i}(xu);function Yu(t){var e=function(){if("undefined"==typeof Reflect||!Pd)return!1;if(Pd.sham)return!1;if("function"==typeof Proxy)return!0;try{return Boolean.prototype.valueOf.call(Pd(Boolean,[],(function(){}))),!0}catch(t){return!1}}();return function(){var i,n=Ld(t);if(e){var o=Ld(this).constructor;i=Pd(n,arguments,o)}else i=n.apply(this,arguments);return Fd(this,i)}}var Xu=function(t){Ad(i,t);var e=Yu(i);function i(t,n,o){var r;return Nn(this,i),(r=e.call(this,t,n,o))._setMargins(o),r}return Fn(i,[{key:"resize",value:function(t,e,i){this.needsRefresh(e,i)&&(this.iconSize={width:Number(this.options.icon.size),height:Number(this.options.icon.size)},this.width=this.iconSize.width+this.margin.right+this.margin.left,this.height=this.iconSize.height+this.margin.top+this.margin.bottom,this.radius=.5*this.width)}},{key:"draw",value:function(t,e,i,n,o,r){var s=this;return this.resize(t,n,o),this.options.icon.size=this.options.icon.size||50,this.left=e-this.width/2,this.top=i-this.height/2,this._icon(t,e,i,n,o,r),{drawExternalLabel:function(){if(void 0!==s.options.label){s.labelModule.draw(t,s.left+s.iconSize.width/2+s.margin.left,i+s.height/2+5,n)}s.updateBoundingBox(e,i)}}}},{key:"updateBoundingBox",value:function(t,e){if(this.boundingBox.top=e-.5*this.options.icon.size,this.boundingBox.left=t-.5*this.options.icon.size,this.boundingBox.right=t+.5*this.options.icon.size,this.boundingBox.bottom=e+.5*this.options.icon.size,void 0!==this.options.label&&this.labelModule.size.width>0){this.boundingBox.left=Math.min(this.boundingBox.left,this.labelModule.size.left),this.boundingBox.right=Math.max(this.boundingBox.right,this.labelModule.size.left+this.labelModule.size.width),this.boundingBox.bottom=Math.max(this.boundingBox.bottom,this.boundingBox.bottom+this.labelModule.size.height+5)}}},{key:"_icon",value:function(t,e,i,n,o,r){var s=Number(this.options.icon.size);void 0!==this.options.icon.code?(t.font=[null!=this.options.icon.weight?this.options.icon.weight:n?"bold":"",(null!=this.options.icon.weight&&n?5:0)+s+"px",this.options.icon.face].join(" "),t.fillStyle=this.options.icon.color||"black",t.textAlign="center",t.textBaseline="middle",this.enableShadow(t,r),t.fillText(this.options.icon.code,e,i),this.disableShadow(t,r)):console.error("When using the icon shape, you need to define the code in the icon options object. This can be done per node or globally.")}},{key:"distanceToBorder",value:function(t,e){return this._distanceToBorder(t,e)}}]),i}(xu);function Gu(t){var e=function(){if("undefined"==typeof Reflect||!Pd)return!1;if(Pd.sham)return!1;if("function"==typeof Proxy)return!0;try{return Boolean.prototype.valueOf.call(Pd(Boolean,[],(function(){}))),!0}catch(t){return!1}}();return function(){var i,n=Ld(t);if(e){var o=Ld(this).constructor;i=Pd(n,arguments,o)}else i=n.apply(this,arguments);return Fd(this,i)}}var Ku=function(t){Ad(i,t);var e=Gu(i);function i(t,n,o,r,s){var a;return Nn(this,i),(a=e.call(this,t,n,o)).setImages(r,s),a}return Fn(i,[{key:"resize",value:function(t){var e=arguments.length>1&&void 0!==arguments[1]?arguments[1]:this.selected,i=arguments.length>2&&void 0!==arguments[2]?arguments[2]:this.hover,n=void 0===this.imageObj.src||void 0===this.imageObj.width||void 0===this.imageObj.height;if(n){var o=2*this.options.size;return this.width=o,void(this.height=o)}this.needsRefresh(e,i)&&this._resizeImage()}},{key:"draw",value:function(t,e,i,n,o,r){t.save(),this.switchImages(n),this.resize();var s=e,a=i;if("top-left"===this.options.shapeProperties.coordinateOrigin?(this.left=e,this.top=i,s+=this.width/2,a+=this.height/2):(this.left=e-this.width/2,this.top=i-this.height/2),!0===this.options.shapeProperties.useBorderWithImage){var h=this.options.borderWidth,l=this.options.borderWidthSelected||2*this.options.borderWidth,d=(n?l:h)/this.body.view.scale;t.lineWidth=Math.min(this.width,d),t.beginPath();var c=n?this.options.color.highlight.border:o?this.options.color.hover.border:this.options.color.border,u=n?this.options.color.highlight.background:o?this.options.color.hover.background:this.options.color.background;void 0!==r.opacity&&(c=Fh(c,r.opacity),u=Fh(u,r.opacity)),t.strokeStyle=c,t.fillStyle=u,t.rect(this.left-.5*t.lineWidth,this.top-.5*t.lineWidth,this.width+t.lineWidth,this.height+t.lineWidth),hs(t).call(t),this.performStroke(t,r),t.closePath()}this._drawImageAtPosition(t,r),this._drawImageLabel(t,s,a,n,o),this.updateBoundingBox(e,i),t.restore()}},{key:"updateBoundingBox",value:function(t,e){this.resize(),"top-left"===this.options.shapeProperties.coordinateOrigin?(this.left=t,this.top=e):(this.left=t-this.width/2,this.top=e-this.height/2),this.boundingBox.left=this.left,this.boundingBox.top=this.top,this.boundingBox.bottom=this.top+this.height,this.boundingBox.right=this.left+this.width,void 0!==this.options.label&&this.labelModule.size.width>0&&(this.boundingBox.left=Math.min(this.boundingBox.left,this.labelModule.size.left),this.boundingBox.right=Math.max(this.boundingBox.right,this.labelModule.size.left+this.labelModule.size.width),this.boundingBox.bottom=Math.max(this.boundingBox.bottom,this.boundingBox.bottom+this.labelOffset))}},{key:"distanceToBorder",value:function(t,e){return this._distanceToBorder(t,e)}}]),i}(Su);function $u(t){var e=function(){if("undefined"==typeof Reflect||!Pd)return!1;if(Pd.sham)return!1;if("function"==typeof Proxy)return!0;try{return Boolean.prototype.valueOf.call(Pd(Boolean,[],(function(){}))),!0}catch(t){return!1}}();return function(){var i,n=Ld(t);if(e){var o=Ld(this).constructor;i=Pd(n,arguments,o)}else i=n.apply(this,arguments);return Fd(this,i)}}var Zu=function(t){Ad(i,t);var e=$u(i);function i(t,n,o){return Nn(this,i),e.call(this,t,n,o)}return Fn(i,[{key:"draw",value:function(t,e,i,n,o,r){return this._drawShape(t,"square",2,e,i,n,o,r)}},{key:"distanceToBorder",value:function(t,e){return this._distanceToBorder(t,e)}}]),i}(Bu);function Qu(t){var e=function(){if("undefined"==typeof Reflect||!Pd)return!1;if(Pd.sham)return!1;if("function"==typeof Proxy)return!0;try{return Boolean.prototype.valueOf.call(Pd(Boolean,[],(function(){}))),!0}catch(t){return!1}}();return function(){var i,n=Ld(t);if(e){var o=Ld(this).constructor;i=Pd(n,arguments,o)}else i=n.apply(this,arguments);return Fd(this,i)}}var Ju=function(t){Ad(i,t);var e=Qu(i);function i(t,n,o){return Nn(this,i),e.call(this,t,n,o)}return Fn(i,[{key:"draw",value:function(t,e,i,n,o,r){return this._drawShape(t,"hexagon",4,e,i,n,o,r)}},{key:"distanceToBorder",value:function(t,e){return this._distanceToBorder(t,e)}}]),i}(Bu);function tf(t){var e=function(){if("undefined"==typeof Reflect||!Pd)return!1;if(Pd.sham)return!1;if("function"==typeof Proxy)return!0;try{return Boolean.prototype.valueOf.call(Pd(Boolean,[],(function(){}))),!0}catch(t){return!1}}();return function(){var i,n=Ld(t);if(e){var o=Ld(this).constructor;i=Pd(n,arguments,o)}else i=n.apply(this,arguments);return Fd(this,i)}}var ef=function(t){Ad(i,t);var e=tf(i);function i(t,n,o){return Nn(this,i),e.call(this,t,n,o)}return Fn(i,[{key:"draw",value:function(t,e,i,n,o,r){return this._drawShape(t,"star",4,e,i,n,o,r)}},{key:"distanceToBorder",value:function(t,e){return this._distanceToBorder(t,e)}}]),i}(Bu);function nf(t){var e=function(){if("undefined"==typeof Reflect||!Pd)return!1;if(Pd.sham)return!1;if("function"==typeof Proxy)return!0;try{return Boolean.prototype.valueOf.call(Pd(Boolean,[],(function(){}))),!0}catch(t){return!1}}();return function(){var i,n=Ld(t);if(e){var o=Ld(this).constructor;i=Pd(n,arguments,o)}else i=n.apply(this,arguments);return Fd(this,i)}}var of=function(t){Ad(i,t);var e=nf(i);function i(t,n,o){var r;return Nn(this,i),(r=e.call(this,t,n,o))._setMargins(o),r}return Fn(i,[{key:"resize",value:function(t,e,i){this.needsRefresh(e,i)&&(this.textSize=this.labelModule.getTextSize(t,e,i),this.width=this.textSize.width+this.margin.right+this.margin.left,this.height=this.textSize.height+this.margin.top+this.margin.bottom,this.radius=.5*this.width)}},{key:"draw",value:function(t,e,i,n,o,r){this.resize(t,n,o),this.left=e-this.width/2,this.top=i-this.height/2,this.enableShadow(t,r),this.labelModule.draw(t,this.left+this.textSize.width/2+this.margin.left,this.top+this.textSize.height/2+this.margin.top,n,o),this.disableShadow(t,r),this.updateBoundingBox(e,i,t,n,o)}},{key:"distanceToBorder",value:function(t,e){return this._distanceToBorder(t,e)}}]),i}(xu);function rf(t){var e=function(){if("undefined"==typeof Reflect||!Pd)return!1;if(Pd.sham)return!1;if("function"==typeof Proxy)return!0;try{return Boolean.prototype.valueOf.call(Pd(Boolean,[],(function(){}))),!0}catch(t){return!1}}();return function(){var i,n=Ld(t);if(e){var o=Ld(this).constructor;i=Pd(n,arguments,o)}else i=n.apply(this,arguments);return Fd(this,i)}}var sf=function(t){Ad(i,t);var e=rf(i);function i(t,n,o){return Nn(this,i),e.call(this,t,n,o)}return Fn(i,[{key:"draw",value:function(t,e,i,n,o,r){return this._drawShape(t,"triangle",3,e,i,n,o,r)}},{key:"distanceToBorder",value:function(t,e){return this._distanceToBorder(t,e)}}]),i}(Bu);function af(t){var e=function(){if("undefined"==typeof Reflect||!Pd)return!1;if(Pd.sham)return!1;if("function"==typeof Proxy)return!0;try{return Boolean.prototype.valueOf.call(Pd(Boolean,[],(function(){}))),!0}catch(t){return!1}}();return function(){var i,n=Ld(t);if(e){var o=Ld(this).constructor;i=Pd(n,arguments,o)}else i=n.apply(this,arguments);return Fd(this,i)}}var hf=function(t){Ad(i,t);var e=af(i);function i(t,n,o){return Nn(this,i),e.call(this,t,n,o)}return Fn(i,[{key:"draw",value:function(t,e,i,n,o,r){return this._drawShape(t,"triangleDown",3,e,i,n,o,r)}},{key:"distanceToBorder",value:function(t,e){return this._distanceToBorder(t,e)}}]),i}(Bu);function lf(t,e){var i=zo(t);if(On){var n=On(t);e&&(n=mr(n).call(n,(function(e){return Mn(t,e).enumerable}))),i.push.apply(i,n)}return i}function df(t){for(var e=1;e<arguments.length;e++){var i,n=null!=arguments[e]?arguments[e]:{};if(e%2)Wo(i=lf(Object(n),!0)).call(i,(function(e){jn(t,e,n[e])}));else if(Dn)In(t,Dn(n));else{var o;Wo(o=lf(Object(n))).call(o,(function(e){zn(t,e,Mn(n,e))}))}}return t}var cf=function(){function t(e,i,n,o,r,s){Nn(this,t),this.options=Kh(r),this.globalOptions=r,this.defaultOptions=s,this.body=i,this.edges=[],this.id=void 0,this.imagelist=n,this.grouplist=o,this.x=void 0,this.y=void 0,this.baseSize=this.options.size,this.baseFontSize=this.options.font.size,this.predefinedPosition=!1,this.selected=!1,this.hover=!1,this.labelModule=new _u(this.body,this.options,!1),this.setOptions(e)}return Fn(t,[{key:"attachEdge",value:function(t){var e;-1===Hr(e=this.edges).call(e,t)&&this.edges.push(t)}},{key:"detachEdge",value:function(t){var e,i,n=Hr(e=this.edges).call(e,t);-1!=n&&er(i=this.edges).call(i,n,1)}},{key:"setOptions",value:function(e){var i=this.options.shape;if(e){if(void 0!==e.color&&(this._localColor=e.color),void 0!==e.id&&(this.id=e.id),void 0===this.id)throw new Error("Node must have an id");t.checkMass(e,this.id),void 0!==e.x&&(null===e.x?(this.x=void 0,this.predefinedPosition=!1):(this.x=Br(e.x),this.predefinedPosition=!0)),void 0!==e.y&&(null===e.y?(this.y=void 0,this.predefinedPosition=!1):(this.y=Br(e.y),this.predefinedPosition=!0)),void 0!==e.size&&(this.baseSize=e.size),void 0!==e.value&&(e.value=hu(e.value)),t.parseOptions(this.options,e,!0,this.globalOptions,this.grouplist);var n=[e,this.options,this.defaultOptions];return this.chooser=fu("node",n),this._load_images(),this.updateLabelModule(e),void 0!==e.opacity&&t.checkOpacity(e.opacity)&&(this.options.opacity=e.opacity),this.updateShape(i),void 0!==e.hidden||void 0!==e.physics}}},{key:"_load_images",value:function(){if(("circularImage"===this.options.shape||"image"===this.options.shape)&&void 0===this.options.image)throw new Error("Option image must be defined for node type '"+this.options.shape+"'");if(void 0!==this.options.image){if(void 0===this.imagelist)throw new Error("Internal Error: No images provided");if("string"==typeof this.options.image)this.imageObj=this.imagelist.load(this.options.image,this.options.brokenImage,this.id);else{if(void 0===this.options.image.unselected)throw new Error("No unselected image provided");this.imageObj=this.imagelist.load(this.options.image.unselected,this.options.brokenImage,this.id),void 0!==this.options.image.selected?this.imageObjAlt=this.imagelist.load(this.options.image.selected,this.options.brokenImage,this.id):this.imageObjAlt=void 0}}}},{key:"getFormattingValues",value:function(){var t={color:this.options.color.background,opacity:this.options.opacity,borderWidth:this.options.borderWidth,borderColor:this.options.color.border,size:this.options.size,borderDashes:this.options.shapeProperties.borderDashes,borderRadius:this.options.shapeProperties.borderRadius,shadow:this.options.shadow.enabled,shadowColor:this.options.shadow.color,shadowSize:this.options.shadow.size,shadowX:this.options.shadow.x,shadowY:this.options.shadow.y};if(this.selected||this.hover?!0===this.chooser?this.selected?(null!=this.options.borderWidthSelected?t.borderWidth=this.options.borderWidthSelected:t.borderWidth*=2,t.color=this.options.color.highlight.background,t.borderColor=this.options.color.highlight.border,t.shadow=this.options.shadow.enabled):this.hover&&(t.color=this.options.color.hover.background,t.borderColor=this.options.color.hover.border,t.shadow=this.options.shadow.enabled):"function"==typeof this.chooser&&(this.chooser(t,this.options.id,this.selected,this.hover),!1===t.shadow&&(t.shadowColor===this.options.shadow.color&&t.shadowSize===this.options.shadow.size&&t.shadowX===this.options.shadow.x&&t.shadowY===this.options.shadow.y||(t.shadow=!0))):t.shadow=this.options.shadow.enabled,void 0!==this.options.opacity){var e=this.options.opacity;t.borderColor=Fh(t.borderColor,e),t.color=Fh(t.color,e),t.shadowColor=Fh(t.shadowColor,e)}return t}},{key:"updateLabelModule",value:function(e){void 0!==this.options.label&&null!==this.options.label||(this.options.label=""),t.updateGroupOptions(this.options,df(df({},e),{},{color:e&&e.color||this._localColor||void 0}),this.grouplist);var i=this.grouplist.get(this.options.group,!1),n=[e,this.options,i,this.globalOptions,this.defaultOptions];this.labelModule.update(this.options,n),void 0!==this.labelModule.baseSize&&(this.baseFontSize=this.labelModule.baseSize)}},{key:"updateShape",value:function(t){if(t===this.options.shape&&this.shape)this.shape.setOptions(this.options,this.imageObj,this.imageObjAlt);else switch(this.options.shape){case"box":this.shape=new Ou(this.options,this.body,this.labelModule);break;case"circle":this.shape=new Mu(this.options,this.body,this.labelModule);break;case"circularImage":this.shape=new Du(this.options,this.body,this.labelModule,this.imageObj,this.imageObjAlt);break;case"custom":this.shape=new Fu(this.options,this.body,this.labelModule,this.options.ctxRenderer);break;case"database":this.shape=new Ru(this.options,this.body,this.labelModule);break;case"diamond":this.shape=new Hu(this.options,this.body,this.labelModule);break;case"dot":this.shape=new qu(this.options,this.body,this.labelModule);break;case"ellipse":this.shape=new Uu(this.options,this.body,this.labelModule);break;case"icon":this.shape=new Xu(this.options,this.body,this.labelModule);break;case"image":this.shape=new Ku(this.options,this.body,this.labelModule,this.imageObj,this.imageObjAlt);break;case"square":this.shape=new Zu(this.options,this.body,this.labelModule);break;case"hexagon":this.shape=new Ju(this.options,this.body,this.labelModule);break;case"star":this.shape=new ef(this.options,this.body,this.labelModule);break;case"text":this.shape=new of(this.options,this.body,this.labelModule);break;case"triangle":this.shape=new sf(this.options,this.body,this.labelModule);break;case"triangleDown":this.shape=new hf(this.options,this.body,this.labelModule);break;default:this.shape=new Uu(this.options,this.body,this.labelModule)}this.needsRefresh()}},{key:"select",value:function(){this.selected=!0,this.needsRefresh()}},{key:"unselect",value:function(){this.selected=!1,this.needsRefresh()}},{key:"needsRefresh",value:function(){this.shape.refreshNeeded=!0}},{key:"getTitle",value:function(){return this.options.title}},{key:"distanceToBorder",value:function(t,e){return this.shape.distanceToBorder(t,e)}},{key:"isFixed",value:function(){return this.options.fixed.x&&this.options.fixed.y}},{key:"isSelected",value:function(){return this.selected}},{key:"getValue",value:function(){return this.options.value}},{key:"getLabelSize",value:function(){return this.labelModule.size()}},{key:"setValueRange",value:function(t,e,i){if(void 0!==this.options.value){var n=this.options.scaling.customScalingFunction(t,e,i,this.options.value),o=this.options.scaling.max-this.options.scaling.min;if(!0===this.options.scaling.label.enabled){var r=this.options.scaling.label.max-this.options.scaling.label.min;this.options.font.size=this.options.scaling.label.min+n*r}this.options.size=this.options.scaling.min+n*o}else this.options.size=this.baseSize,this.options.font.size=this.baseFontSize;this.updateLabelModule()}},{key:"draw",value:function(t){var e=this.getFormattingValues();return this.shape.draw(t,this.x,this.y,this.selected,this.hover,e)||{}}},{key:"updateBoundingBox",value:function(t){this.shape.updateBoundingBox(this.x,this.y,t)}},{key:"resize",value:function(t){var e=this.getFormattingValues();this.shape.resize(t,this.selected,this.hover,e)}},{key:"getItemsOnPoint",value:function(t){var e=[];return this.labelModule.visible()&&pu(this.labelModule.getSize(),t)&&e.push({nodeId:this.id,labelId:0}),pu(this.shape.boundingBox,t)&&e.push({nodeId:this.id}),e}},{key:"isOverlappingWith",value:function(t){return this.shape.left<t.right&&this.shape.left+this.shape.width>t.left&&this.shape.top<t.bottom&&this.shape.top+this.shape.height>t.top}},{key:"isBoundingBoxOverlappingWith",value:function(t){return this.shape.boundingBox.left<t.right&&this.shape.boundingBox.right>t.left&&this.shape.boundingBox.top<t.bottom&&this.shape.boundingBox.bottom>t.top}}],[{key:"checkOpacity",value:function(t){return 0<=t&&t<=1}},{key:"checkCoordinateOrigin",value:function(t){return void 0===t||"center"===t||"top-left"===t}},{key:"updateGroupOptions",value:function(e,i,n){var o;if(void 0!==n){var r=e.group;if(void 0!==i&&void 0!==i.group&&r!==i.group)throw new Error("updateGroupOptions: group values in options don't match.");if("number"==typeof r||"string"==typeof r&&""!=r){var s=n.get(r);void 0!==s.opacity&&void 0===i.opacity&&(t.checkOpacity(s.opacity)||(console.error("Invalid option for node opacity. Value must be between 0 and 1, found: "+s.opacity),s.opacity=void 0));var a=mr(o=uu(i)).call(o,(function(t){return null!=i[t]}));a.push("font"),Oh(a,e,s),e.color=Rh(e.color)}}}},{key:"parseOptions",value:function(e,i){var n=arguments.length>2&&void 0!==arguments[2]&&arguments[2],o=arguments.length>3&&void 0!==arguments[3]?arguments[3]:{},r=arguments.length>4?arguments[4]:void 0,s=["color","fixed","shadow"];if(Oh(s,e,i,n),t.checkMass(i),void 0!==e.opacity&&(t.checkOpacity(e.opacity)||(console.error("Invalid option for node opacity. Value must be between 0 and 1, found: "+e.opacity),e.opacity=void 0)),void 0!==i.opacity&&(t.checkOpacity(i.opacity)||(console.error("Invalid option for node opacity. Value must be between 0 and 1, found: "+i.opacity),i.opacity=void 0)),i.shapeProperties&&!t.checkCoordinateOrigin(i.shapeProperties.coordinateOrigin)&&console.error("Invalid option for node coordinateOrigin, found: "+i.shapeProperties.coordinateOrigin),$h(e,i,"shadow",o),void 0!==i.color&&null!==i.color){var a=Rh(i.color);_h(e.color,a)}else!0===n&&null===i.color&&(e.color=Kh(o.color));void 0!==i.fixed&&null!==i.fixed&&("boolean"==typeof i.fixed?(e.fixed.x=i.fixed,e.fixed.y=i.fixed):(void 0!==i.fixed.x&&"boolean"==typeof i.fixed.x&&(e.fixed.x=i.fixed.x),void 0!==i.fixed.y&&"boolean"==typeof i.fixed.y&&(e.fixed.y=i.fixed.y))),!0===n&&null===i.font&&(e.font=Kh(o.font)),t.updateGroupOptions(e,i,r),void 0!==i.scaling&&$h(e.scaling,i.scaling,"label",o.scaling)}},{key:"checkMass",value:function(t,e){if(void 0!==t.mass&&t.mass<=0){var i="";void 0!==e&&(i=" in node id: "+e),console.error("%cNegative or zero mass disallowed"+i+", setting mass to 1.",cl),t.mass=1}}}]),t}();function uf(t,e){var i=void 0!==ko&&Bi(t)||t["@@iterator"];if(!i){if(So(t)||(i=function(t,e){var i;if(!t)return;if("string"==typeof t)return ff(t,e);var n=Oo(i=Object.prototype.toString.call(t)).call(i,8,-1);"Object"===n&&t.constructor&&(n=t.constructor.name);if("Map"===n||"Set"===n)return Ei(t);if("Arguments"===n||/^(?:Ui|I)nt(?:8|16|32)(?:Clamped)?Array$/.test(n))return ff(t,e)}(t))||e&&t&&"number"==typeof t.length){i&&(t=i);var n=0,o=function(){};return{s:o,n:function(){return n>=t.length?{done:!0}:{done:!1,value:t[n++]}},e:function(t){throw t},f:o}}throw new TypeError("Invalid attempt to iterate non-iterable instance.\nIn order to be iterable, non-array objects must have a [Symbol.iterator]() method.")}var r,s=!0,a=!1;return{s:function(){i=i.call(t)},n:function(){var t=i.next();return s=t.done,t},e:function(t){a=!0,r=t},f:function(){try{s||null==i.return||i.return()}finally{if(a)throw r}}}}function ff(t,e){(null==e||e>t.length)&&(e=t.length);for(var i=0,n=new Array(e);i<e;i++)n[i]=t[i];return n}var pf=function(){function t(e,i,n,o){var r,s=this;if(Nn(this,t),this.body=e,this.images=i,this.groups=n,this.layoutEngine=o,this.body.functions.createNode=Vt(r=this.create).call(r,this),this.nodesListeners={add:function(t,e){s.add(e.items)},update:function(t,e){s.update(e.items,e.data,e.oldData)},remove:function(t,e){s.remove(e.items)}},this.defaultOptions={borderWidth:1,borderWidthSelected:void 0,brokenImage:void 0,color:{border:"#2B7CE9",background:"#97C2FC",highlight:{border:"#2B7CE9",background:"#D2E5FF"},hover:{border:"#2B7CE9",background:"#D2E5FF"}},opacity:void 0,fixed:{x:!1,y:!1},font:{color:"#343434",size:14,face:"arial",background:"none",strokeWidth:0,strokeColor:"#ffffff",align:"center",vadjust:0,multi:!1,bold:{mod:"bold"},boldital:{mod:"bold italic"},ital:{mod:"italic"},mono:{mod:"",size:15,face:"monospace",vadjust:2}},group:void 0,hidden:!1,icon:{face:"FontAwesome",code:void 0,size:50,color:"#2B7CE9"},image:void 0,imagePadding:{top:0,right:0,bottom:0,left:0},label:void 0,labelHighlightBold:!0,level:void 0,margin:{top:5,right:5,bottom:5,left:5},mass:1,physics:!0,scaling:{min:10,max:30,label:{enabled:!1,min:14,max:30,maxVisible:30,drawThreshold:5},customScalingFunction:function(t,e,i,n){if(e===t)return.5;var o=1/(e-t);return Math.max(0,(n-t)*o)}},shadow:{enabled:!1,color:"rgba(0,0,0,0.5)",size:10,x:5,y:5},shape:"ellipse",shapeProperties:{borderDashes:!1,borderRadius:6,interpolation:!0,useImageSize:!1,useBorderWithImage:!1,coordinateOrigin:"center"},size:25,title:void 0,value:void 0,x:void 0,y:void 0},this.defaultOptions.mass<=0)throw"Internal error: mass in defaultOptions of NodesHandler may not be zero or negative";this.options=Kh(this.defaultOptions),this.bindEventListeners()}return Fn(t,[{key:"bindEventListeners",value:function(){var t,e,i=this;this.body.emitter.on("refreshNodes",Vt(t=this.refresh).call(t,this)),this.body.emitter.on("refresh",Vt(e=this.refresh).call(e,this)),this.body.emitter.on("destroy",(function(){Dh(i.nodesListeners,(function(t,e){i.body.data.nodes&&i.body.data.nodes.off(e,t)})),delete i.body.functions.createNode,delete i.nodesListeners.add,delete i.nodesListeners.update,delete i.nodesListeners.remove,delete i.nodesListeners}))}},{key:"setOptions",value:function(t){if(void 0!==t){if(cf.parseOptions(this.options,t),void 0!==t.opacity&&(yd(t.opacity)||!wd(t.opacity)||t.opacity<0||t.opacity>1?console.error("Invalid option for node opacity. Value must be between 0 and 1, found: "+t.opacity):this.options.opacity=t.opacity),void 0!==t.shape)for(var e in this.body.nodes)Object.prototype.hasOwnProperty.call(this.body.nodes,e)&&this.body.nodes[e].updateShape();if(void 0!==t.font||void 0!==t.widthConstraint||void 0!==t.heightConstraint)for(var i=0,n=zo(this.body.nodes);i<n.length;i++){var o=n[i];this.body.nodes[o].updateLabelModule(),this.body.nodes[o].needsRefresh()}if(void 0!==t.size)for(var r in this.body.nodes)Object.prototype.hasOwnProperty.call(this.body.nodes,r)&&this.body.nodes[r].needsRefresh();void 0===t.hidden&&void 0===t.physics||this.body.emitter.emit("_dataChanged")}}},{key:"setData",value:function(t){var e=arguments.length>1&&void 0!==arguments[1]&&arguments[1],i=this.body.data.nodes;if(nu("id",t))this.body.data.nodes=t;else if(So(t))this.body.data.nodes=new tu,this.body.data.nodes.add(t);else{if(t)throw new TypeError("Array or DataSet expected");this.body.data.nodes=new tu}if(i&&Dh(this.nodesListeners,(function(t,e){i.off(e,t)})),this.body.nodes={},this.body.data.nodes){var n=this;Dh(this.nodesListeners,(function(t,e){n.body.data.nodes.on(e,t)}));var o=this.body.data.nodes.getIds();this.add(o,!0)}!1===e&&this.body.emitter.emit("_dataChanged")}},{key:"add",value:function(t){for(var e,i=arguments.length>1&&void 0!==arguments[1]&&arguments[1],n=[],o=0;o<t.length;o++){e=t[o];var r=this.body.data.nodes.get(e),s=this.create(r);n.push(s),this.body.nodes[e]=s}this.layoutEngine.positionInitially(n),!1===i&&this.body.emitter.emit("_dataChanged")}},{key:"update",value:function(t,e,i){for(var n=this.body.nodes,o=!1,r=0;r<t.length;r++){var s=t[r],a=n[s],h=e[r];void 0!==a?a.setOptions(h)&&(o=!0):(o=!0,a=this.create(h),n[s]=a)}o||void 0===i||(o=Od(e).call(e,(function(t,e){var n=i[e];return n&&n.level!==t.level}))),!0===o?this.body.emitter.emit("_dataChanged"):this.body.emitter.emit("_dataUpdated")}},{key:"remove",value:function(t){for(var e=this.body.nodes,i=0;i<t.length;i++){delete e[t[i]]}this.body.emitter.emit("_dataChanged")}},{key:"create",value:function(t){var e=arguments.length>1&&void 0!==arguments[1]?arguments[1]:cf;return new e(t,this.body,this.images,this.groups,this.options,this.defaultOptions)}},{key:"refresh",value:function(){var t=this,e=arguments.length>0&&void 0!==arguments[0]&&arguments[0];Dh(this.body.nodes,(function(i,n){var o=t.body.data.nodes.get(n);void 0!==o&&(!0===e&&i.setOptions({x:null,y:null}),i.setOptions({fixed:!1}),i.setOptions(o))}))}},{key:"getPositions",value:function(t){var e={};if(void 0!==t){if(!0===So(t)){for(var i=0;i<t.length;i++)if(void 0!==this.body.nodes[t[i]]){var n=this.body.nodes[t[i]];e[t[i]]={x:Math.round(n.x),y:Math.round(n.y)}}}else if(void 0!==this.body.nodes[t]){var o=this.body.nodes[t];e[t]={x:Math.round(o.x),y:Math.round(o.y)}}}else for(var r=0;r<this.body.nodeIndices.length;r++){var s=this.body.nodes[this.body.nodeIndices[r]];e[this.body.nodeIndices[r]]={x:Math.round(s.x),y:Math.round(s.y)}}return e}},{key:"getPosition",value:function(t){if(null==t)throw new TypeError("No id was specified for getPosition method.");if(null==this.body.nodes[t])throw new ReferenceError("NodeId provided for getPosition does not exist. Provided: ".concat(t));return{x:Math.round(this.body.nodes[t].x),y:Math.round(this.body.nodes[t].y)}}},{key:"storePositions",value:function(){var t,e=[],i=this.body.data.nodes.getDataSet(),n=uf(i.get());try{for(n.s();!(t=n.n()).done;){var o=t.value,r=o.id,s=this.body.nodes[r],a=Math.round(s.x),h=Math.round(s.y);o.x===a&&o.y===h||e.push({id:r,x:a,y:h})}}catch(t){n.e(t)}finally{n.f()}i.update(e)}},{key:"getBoundingBox",value:function(t){if(void 0!==this.body.nodes[t])return this.body.nodes[t].shape.boundingBox}},{key:"getConnectedNodes",value:function(t,e){var i=[];if(void 0!==this.body.nodes[t])for(var n=this.body.nodes[t],o={},r=0;r<n.edges.length;r++){var s=n.edges[r];"to"!==e&&s.toId==n.id?void 0===o[s.fromId]&&(i.push(s.fromId),o[s.fromId]=!0):"from"!==e&&s.fromId==n.id&&void 0===o[s.toId]&&(i.push(s.toId),o[s.toId]=!0)}return i}},{key:"getConnectedEdges",value:function(t){var e=[];if(void 0!==this.body.nodes[t])for(var i=this.body.nodes[t],n=0;n<i.edges.length;n++)e.push(i.edges[n].id);else console.error("NodeId provided for getConnectedEdges does not exist. Provided: ",t);return e}},{key:"moveNode",value:function(t,e,i){var n=this;void 0!==this.body.nodes[t]?(this.body.nodes[t].x=Number(e),this.body.nodes[t].y=Number(i),rs((function(){n.body.emitter.emit("startSimulation")}),0)):console.error("Node id supplied to moveNode does not exist. Provided: ",t)}}]),t}();gt({target:"Reflect",stat:!0},{get:function t(e,i){var n,o,r=arguments.length<3?e:arguments[2];return dt(e)===r?e[i]:(n=tt.f(e,i))?j(n,"value")?n.value:void 0===n.get?void 0:n.get.call(r):w(o=Ee(e))?t(o,i,r):void 0}});var vf=k.Reflect.get,gf=Tn,yf=n((function(t){t.exports=function(t,e){for(;!Object.prototype.hasOwnProperty.call(t,e)&&null!==(t=Rd(t)););return t},t.exports.default=t.exports,t.exports.__esModule=!0}));i(yf);var mf=i(n((function(t){function e(i,n,o){return"undefined"!=typeof Reflect&&vf?(t.exports=e=vf,t.exports.default=t.exports,t.exports.__esModule=!0):(t.exports=e=function(t,e,i){var n=yf(t,e);if(n){var o=gf(n,e);return o.get?o.get.call(i):o.value}},t.exports.default=t.exports,t.exports.__esModule=!0),e(i,n,o||i)}t.exports=e,t.exports.default=t.exports,t.exports.__esModule=!0}))),bf=Math.hypot,wf=Math.abs,kf=Math.sqrt,_f=!!bf&&bf(1/0,NaN)!==1/0;gt({target:"Math",stat:!0,forced:_f},{hypot:function(t,e){for(var i,n,o=0,r=0,s=arguments.length,a=0;r<s;)a<(i=wf(arguments[r++]))?(o=o*(n=a/i)*n+1,a=i):o+=i>0?(n=i/a)*n:i;return a===1/0?1/0:a*kf(o)}});var xf=k.Math.hypot;function Ef(t){var e=function(){if("undefined"==typeof Reflect||!Pd)return!1;if(Pd.sham)return!1;if("function"==typeof Proxy)return!0;try{return Boolean.prototype.valueOf.call(Pd(Boolean,[],(function(){}))),!0}catch(t){return!1}}();return function(){var i,n=Ld(t);if(e){var o=Ld(this).constructor;i=Pd(n,arguments,o)}else i=n.apply(this,arguments);return Fd(this,i)}}var Of=function(){function t(){Nn(this,t)}return Fn(t,null,[{key:"transform",value:function(t,e){So(t)||(t=[t]);for(var i=e.point.x,n=e.point.y,o=e.angle,r=e.length,s=0;s<t.length;++s){var a=t[s],h=a.x*Math.cos(o)-a.y*Math.sin(o),l=a.x*Math.sin(o)+a.y*Math.cos(o);a.x=i+r*h,a.y=n+r*l}}},{key:"drawPath",value:function(t,e){t.beginPath(),t.moveTo(e[0].x,e[0].y);for(var i=1;i<e.length;++i)t.lineTo(e[i].x,e[i].y);t.closePath()}}]),t}(),Cf=function(t){Ad(i,t);var e=Ef(i);function i(){return Nn(this,i),e.apply(this,arguments)}return Fn(i,null,[{key:"draw",value:function(t,e){if(e.image){t.save(),t.translate(e.point.x,e.point.y),t.rotate(Math.PI/2+e.angle);var i=null!=e.imageWidth?e.imageWidth:e.image.width,n=null!=e.imageHeight?e.imageHeight:e.image.height;e.image.drawImageAtPosition(t,1,-i/2,0,i,n),t.restore()}return!1}}]),i}(Of),Sf=function(t){Ad(i,t);var e=Ef(i);function i(){return Nn(this,i),e.apply(this,arguments)}return Fn(i,null,[{key:"draw",value:function(t,e){var i=[{x:0,y:0},{x:-1,y:.3},{x:-.9,y:0},{x:-1,y:-.3}];return Of.transform(i,e),Of.drawPath(t,i),!0}}]),i}(Of),Tf=function(){function t(){Nn(this,t)}return Fn(t,null,[{key:"draw",value:function(t,e){var i=[{x:-1,y:0},{x:0,y:.3},{x:-.4,y:0},{x:0,y:-.3}];return Of.transform(i,e),Of.drawPath(t,i),!0}}]),t}(),Mf=function(){function t(){Nn(this,t)}return Fn(t,null,[{key:"draw",value:function(t,e){var i={x:-.4,y:0};Of.transform(i,e),t.strokeStyle=t.fillStyle,t.fillStyle="rgba(0, 0, 0, 0)";var n=Math.PI,o=e.angle-n/2,r=e.angle+n/2;return t.beginPath(),t.arc(i.x,i.y,.4*e.length,o,r,!1),t.stroke(),!0}}]),t}(),Pf=function(){function t(){Nn(this,t)}return Fn(t,null,[{key:"draw",value:function(t,e){var i={x:-.3,y:0};Of.transform(i,e),t.strokeStyle=t.fillStyle,t.fillStyle="rgba(0, 0, 0, 0)";var n=Math.PI,o=e.angle+n/2,r=e.angle+3*n/2;return t.beginPath(),t.arc(i.x,i.y,.4*e.length,o,r,!1),t.stroke(),!0}}]),t}(),Df=function(){function t(){Nn(this,t)}return Fn(t,null,[{key:"draw",value:function(t,e){var i=[{x:.02,y:0},{x:-1,y:.3},{x:-1,y:-.3}];return Of.transform(i,e),Of.drawPath(t,i),!0}}]),t}(),If=function(){function t(){Nn(this,t)}return Fn(t,null,[{key:"draw",value:function(t,e){var i=[{x:0,y:.3},{x:0,y:-.3},{x:-1,y:0}];return Of.transform(i,e),Of.drawPath(t,i),!0}}]),t}(),Bf=function(){function t(){Nn(this,t)}return Fn(t,null,[{key:"draw",value:function(t,e){var i={x:-.4,y:0};return Of.transform(i,e),Ut(t,i.x,i.y,.4*e.length),!0}}]),t}(),zf=function(){function t(){Nn(this,t)}return Fn(t,null,[{key:"draw",value:function(t,e){var i=[{x:0,y:.5},{x:0,y:-.5},{x:-.15,y:-.5},{x:-.15,y:.5}];return Of.transform(i,e),Of.drawPath(t,i),!0}}]),t}(),Nf=function(){function t(){Nn(this,t)}return Fn(t,null,[{key:"draw",value:function(t,e){var i=[{x:0,y:.3},{x:0,y:-.3},{x:-.6,y:-.3},{x:-.6,y:.3}];return Of.transform(i,e),Of.drawPath(t,i),!0}}]),t}(),Af=function(){function t(){Nn(this,t)}return Fn(t,null,[{key:"draw",value:function(t,e){var i=[{x:0,y:0},{x:-.5,y:-.3},{x:-1,y:0},{x:-.5,y:.3}];return Of.transform(i,e),Of.drawPath(t,i),!0}}]),t}(),Ff=function(){function t(){Nn(this,t)}return Fn(t,null,[{key:"draw",value:function(t,e){var i=[{x:-1,y:.3},{x:-.5,y:0},{x:-1,y:-.3},{x:0,y:0}];return Of.transform(i,e),Of.drawPath(t,i),!0}}]),t}(),jf=function(){function t(){Nn(this,t)}return Fn(t,null,[{key:"draw",value:function(t,e){var i;switch(e.type&&(i=e.type.toLowerCase()),i){case"image":return Cf.draw(t,e);case"circle":return Bf.draw(t,e);case"box":return Nf.draw(t,e);case"crow":return Tf.draw(t,e);case"curve":return Mf.draw(t,e);case"diamond":return Af.draw(t,e);case"inv_curve":return Pf.draw(t,e);case"triangle":return Df.draw(t,e);case"inv_triangle":return If.draw(t,e);case"bar":return zf.draw(t,e);case"vee":return Ff.draw(t,e);case"arrow":default:return Sf.draw(t,e)}}}]),t}();function Rf(t,e){var i=zo(t);if(On){var n=On(t);e&&(n=mr(n).call(n,(function(e){return Mn(t,e).enumerable}))),i.push.apply(i,n)}return i}function Lf(t){for(var e=1;e<arguments.length;e++){var i,n=null!=arguments[e]?arguments[e]:{};if(e%2)Wo(i=Rf(Object(n),!0)).call(i,(function(e){jn(t,e,n[e])}));else if(Dn)In(t,Dn(n));else{var o;Wo(o=Rf(Object(n))).call(o,(function(e){zn(t,e,Mn(n,e))}))}}return t}var Hf=function(){function t(e,i,n){Nn(this,t),this._body=i,this._labelModule=n,this.color={},this.colorDirty=!0,this.hoverWidth=1.5,this.selectionWidth=2,this.setOptions(e),this.fromPoint=this.from,this.toPoint=this.to}return Fn(t,[{key:"connect",value:function(){this.from=this._body.nodes[this.options.from],this.to=this._body.nodes[this.options.to]}},{key:"cleanup",value:function(){return!1}},{key:"setOptions",value:function(t){this.options=t,this.from=this._body.nodes[this.options.from],this.to=this._body.nodes[this.options.to],this.id=this.options.id}},{key:"drawLine",value:function(t,e,i,n){var o=arguments.length>4&&void 0!==arguments[4]?arguments[4]:this.getViaNode();t.strokeStyle=this.getColor(t,e),t.lineWidth=e.width,!1!==e.dashes?this._drawDashedLine(t,e,o):this._drawLine(t,e,o)}},{key:"_drawLine",value:function(t,e,i,n,o){if(this.from!=this.to)this._line(t,e,i,n,o);else{var r=this._getCircleData(t),s=uo(r,3),a=s[0],h=s[1],l=s[2];this._circle(t,e,a,h,l)}}},{key:"_drawDashedLine",value:function(t,e,i,n,o){t.lineCap="round";var r=So(e.dashes)?e.dashes:[5,5];if(void 0!==t.setLineDash){if(t.save(),t.setLineDash(r),t.lineDashOffset=0,this.from!=this.to)this._line(t,e,i);else{var s=this._getCircleData(t),a=uo(s,3),h=a[0],l=a[1],d=a[2];this._circle(t,e,h,l,d)}t.setLineDash([0]),t.lineDashOffset=0,t.restore()}else{if(this.from!=this.to)Kt(t,this.from.x,this.from.y,this.to.x,this.to.y,r);else{var c=this._getCircleData(t),u=uo(c,3),f=u[0],p=u[1],v=u[2];this._circle(t,e,f,p,v)}this.enableShadow(t,e),t.stroke(),this.disableShadow(t,e)}}},{key:"findBorderPosition",value:function(t,e,i){return this.from!=this.to?this._findBorderPosition(t,e,i):this._findBorderPositionCircle(t,e,i)}},{key:"findBorderPositions",value:function(t){if(this.from!=this.to)return{from:this._findBorderPosition(this.from,t),to:this._findBorderPosition(this.to,t)};var e,i=Oo(e=this._getCircleData(t)).call(e,0,2),n=uo(i,2),o=n[0],r=n[1];return{from:this._findBorderPositionCircle(this.from,t,{x:o,y:r,low:.25,high:.6,direction:-1}),to:this._findBorderPositionCircle(this.from,t,{x:o,y:r,low:.6,high:.8,direction:1})}}},{key:"_getCircleData",value:function(t){var e=this.options.selfReference.size;void 0!==t&&void 0===this.from.shape.width&&this.from.shape.resize(t);var i=gu(t,this.options.selfReference.angle,e,this.from);return[i.x,i.y,e]}},{key:"_pointOnCircle",value:function(t,e,i,n){var o=2*n*Math.PI;return{x:t+i*Math.cos(o),y:e-i*Math.sin(o)}}},{key:"_findBorderPositionCircle",value:function(t,e,i){var n,o=i.x,r=i.y,s=i.low,a=i.high,h=i.direction,l=this.options.selfReference.size,d=.5*(s+a),c=0;!0===this.options.arrowStrikethrough&&(-1===h?c=this.options.endPointOffset.from:1===h&&(c=this.options.endPointOffset.to));var u=0;do{d=.5*(s+a),n=this._pointOnCircle(o,r,l,d);var f=Math.atan2(t.y-n.y,t.x-n.x),p=t.distanceToBorder(e,f)+c-Math.sqrt(Math.pow(n.x-t.x,2)+Math.pow(n.y-t.y,2));if(Math.abs(p)<.05)break;p>0?h>0?s=d:a=d:h>0?a=d:s=d,++u}while(s<=a&&u<10);return Lf(Lf({},n),{},{t:d})}},{key:"getLineWidth",value:function(t,e){return!0===t?Math.max(this.selectionWidth,.3/this._body.view.scale):!0===e?Math.max(this.hoverWidth,.3/this._body.view.scale):Math.max(this.options.width,.3/this._body.view.scale)}},{key:"getColor",value:function(t,e){if(!1!==e.inheritsColor){if("both"===e.inheritsColor&&this.from.id!==this.to.id){var i=t.createLinearGradient(this.from.x,this.from.y,this.to.x,this.to.y),n=this.from.options.color.highlight.border,o=this.to.options.color.highlight.border;return!1===this.from.selected&&!1===this.to.selected?(n=Fh(this.from.options.color.border,e.opacity),o=Fh(this.to.options.color.border,e.opacity)):!0===this.from.selected&&!1===this.to.selected?o=this.to.options.color.border:!1===this.from.selected&&!0===this.to.selected&&(n=this.from.options.color.border),i.addColorStop(0,n),i.addColorStop(1,o),i}return"to"===e.inheritsColor?Fh(this.to.options.color.border,e.opacity):Fh(this.from.options.color.border,e.opacity)}return Fh(e.color,e.opacity)}},{key:"_circle",value:function(t,e,i,n,o){this.enableShadow(t,e);var r=0,s=2*Math.PI;if(!this.options.selfReference.renderBehindTheNode){var a=this.options.selfReference.angle,h=this.options.selfReference.angle+Math.PI,l=this._findBorderPositionCircle(this.from,t,{x:i,y:n,low:a,high:h,direction:-1}),d=this._findBorderPositionCircle(this.from,t,{x:i,y:n,low:a,high:h,direction:1});r=Math.atan2(l.y-n,l.x-i),s=Math.atan2(d.y-n,d.x-i)}t.beginPath(),t.arc(i,n,o,r,s,!1),t.stroke(),this.disableShadow(t,e)}},{key:"getDistanceToEdge",value:function(t,e,i,n,o,r){if(this.from!=this.to)return this._getDistanceToEdge(t,e,i,n,o,r);var s=this._getCircleData(void 0),a=uo(s,3),h=a[0],l=a[1],d=a[2],c=h-o,u=l-r;return Math.abs(Math.sqrt(c*c+u*u)-d)}},{key:"_getDistanceToLine",value:function(t,e,i,n,o,r){var s=i-t,a=n-e,h=((o-t)*s+(r-e)*a)/(s*s+a*a);h>1?h=1:h<0&&(h=0);var l=t+h*s-o,d=e+h*a-r;return Math.sqrt(l*l+d*d)}},{key:"getArrowData",value:function(t,e,i,n,o,r){var s,a,h,l,d,c,u,f=r.width;"from"===e?(h=this.from,l=this.to,d=r.fromArrowScale<0,c=Math.abs(r.fromArrowScale),u=r.fromArrowType):"to"===e?(h=this.to,l=this.from,d=r.toArrowScale<0,c=Math.abs(r.toArrowScale),u=r.toArrowType):(h=this.to,l=this.from,d=r.middleArrowScale<0,c=Math.abs(r.middleArrowScale),u=r.middleArrowType);var p=15*c+3*f;if(h!=l){var v=p/xf(h.x-l.x,h.y-l.y);if("middle"!==e)if(!0===this.options.smooth.enabled){var g=this._findBorderPosition(h,t,{via:i}),y=this.getPoint(g.t+v*("from"===e?1:-1),i);s=Math.atan2(g.y-y.y,g.x-y.x),a=g}else s=Math.atan2(h.y-l.y,h.x-l.x),a=this._findBorderPosition(h,t);else{var m=(d?-v:v)/2,b=this.getPoint(.5+m,i),w=this.getPoint(.5-m,i);s=Math.atan2(b.y-w.y,b.x-w.x),a=this.getPoint(.5,i)}}else{var k=this._getCircleData(t),_=uo(k,3),x=_[0],E=_[1],O=_[2];if("from"===e){var C=this.options.selfReference.angle,S=this.options.selfReference.angle+Math.PI,T=this._findBorderPositionCircle(this.from,t,{x:x,y:E,low:C,high:S,direction:-1});s=-2*T.t*Math.PI+1.5*Math.PI+.1*Math.PI,a=T}else if("to"===e){var M=this.options.selfReference.angle,P=this.options.selfReference.angle+Math.PI,D=this._findBorderPositionCircle(this.from,t,{x:x,y:E,low:M,high:P,direction:1});s=-2*D.t*Math.PI+1.5*Math.PI-1.1*Math.PI,a=D}else{var I=this.options.selfReference.angle/(2*Math.PI);a=this._pointOnCircle(x,E,O,I),s=-2*I*Math.PI+1.5*Math.PI+.1*Math.PI}}return{point:a,core:{x:a.x-.9*p*Math.cos(s),y:a.y-.9*p*Math.sin(s)},angle:s,length:p,type:u}}},{key:"drawArrowHead",value:function(t,e,i,n,o){t.strokeStyle=this.getColor(t,e),t.fillStyle=t.strokeStyle,t.lineWidth=e.width,jf.draw(t,o)&&(this.enableShadow(t,e),hs(t).call(t),this.disableShadow(t,e))}},{key:"enableShadow",value:function(t,e){!0===e.shadow&&(t.shadowColor=e.shadowColor,t.shadowBlur=e.shadowSize,t.shadowOffsetX=e.shadowX,t.shadowOffsetY=e.shadowY)}},{key:"disableShadow",value:function(t,e){!0===e.shadow&&(t.shadowColor="rgba(0,0,0,0)",t.shadowBlur=0,t.shadowOffsetX=0,t.shadowOffsetY=0)}},{key:"drawBackground",value:function(t,e){if(!1!==e.background){var i={strokeStyle:t.strokeStyle,lineWidth:t.lineWidth,dashes:t.dashes};t.strokeStyle=e.backgroundColor,t.lineWidth=e.backgroundSize,this.setStrokeDashed(t,e.backgroundDashes),t.stroke(),t.strokeStyle=i.strokeStyle,t.lineWidth=i.lineWidth,t.dashes=i.dashes,this.setStrokeDashed(t,e.dashes)}}},{key:"setStrokeDashed",value:function(t,e){if(!1!==e)if(void 0!==t.setLineDash){var i=So(e)?e:[5,5];t.setLineDash(i)}else console.warn("setLineDash is not supported in this browser. The dashed stroke cannot be used.");else void 0!==t.setLineDash?t.setLineDash([]):console.warn("setLineDash is not supported in this browser. The dashed stroke cannot be used.")}}]),t}();function Wf(t,e){var i=zo(t);if(On){var n=On(t);e&&(n=mr(n).call(n,(function(e){return Mn(t,e).enumerable}))),i.push.apply(i,n)}return i}function qf(t){for(var e=1;e<arguments.length;e++){var i,n=null!=arguments[e]?arguments[e]:{};if(e%2)Wo(i=Wf(Object(n),!0)).call(i,(function(e){jn(t,e,n[e])}));else if(Dn)In(t,Dn(n));else{var o;Wo(o=Wf(Object(n))).call(o,(function(e){zn(t,e,Mn(n,e))}))}}return t}function Vf(t){var e=function(){if("undefined"==typeof Reflect||!Pd)return!1;if(Pd.sham)return!1;if("function"==typeof Proxy)return!0;try{return Boolean.prototype.valueOf.call(Pd(Boolean,[],(function(){}))),!0}catch(t){return!1}}();return function(){var i,n=Ld(t);if(e){var o=Ld(this).constructor;i=Pd(n,arguments,o)}else i=n.apply(this,arguments);return Fd(this,i)}}var Uf=function(t){Ad(i,t);var e=Vf(i);function i(t,n,o){return Nn(this,i),e.call(this,t,n,o)}return Fn(i,[{key:"_findBorderPositionBezier",value:function(t,e){var i,n,o=arguments.length>2&&void 0!==arguments[2]?arguments[2]:this._getViaCoordinates(),r=10,s=.2,a=!1,h=1,l=0,d=this.to,c=this.options.endPointOffset?this.options.endPointOffset.to:0;t.id===this.from.id&&(d=this.from,a=!0,c=this.options.endPointOffset?this.options.endPointOffset.from:0),!1===this.options.arrowStrikethrough&&(c=0);var u=0;do{n=.5*(l+h),i=this.getPoint(n,o);var f=Math.atan2(d.y-i.y,d.x-i.x),p=d.distanceToBorder(e,f)+c,v=Math.sqrt(Math.pow(i.x-d.x,2)+Math.pow(i.y-d.y,2)),g=p-v;if(Math.abs(g)<s)break;g<0?!1===a?l=n:h=n:!1===a?h=n:l=n,++u}while(l<=h&&u<r);return qf(qf({},i),{},{t:n})}},{key:"_getDistanceToBezierEdge",value:function(t,e,i,n,o,r,s){var a,h,l,d,c,u=1e9,f=t,p=e;for(h=1;h<10;h++)l=.1*h,d=Math.pow(1-l,2)*t+2*l*(1-l)*s.x+Math.pow(l,2)*i,c=Math.pow(1-l,2)*e+2*l*(1-l)*s.y+Math.pow(l,2)*n,h>0&&(u=(a=this._getDistanceToLine(f,p,d,c,o,r))<u?a:u),f=d,p=c;return u}},{key:"_bezierCurve",value:function(t,e,i,n){t.beginPath(),t.moveTo(this.fromPoint.x,this.fromPoint.y),null!=i&&null!=i.x?null!=n&&null!=n.x?t.bezierCurveTo(i.x,i.y,n.x,n.y,this.toPoint.x,this.toPoint.y):t.quadraticCurveTo(i.x,i.y,this.toPoint.x,this.toPoint.y):t.lineTo(this.toPoint.x,this.toPoint.y),this.drawBackground(t,e),this.enableShadow(t,e),t.stroke(),this.disableShadow(t,e)}},{key:"getViaNode",value:function(){return this._getViaCoordinates()}}]),i}(Hf);function Yf(t){var e=function(){if("undefined"==typeof Reflect||!Pd)return!1;if(Pd.sham)return!1;if("function"==typeof Proxy)return!0;try{return Boolean.prototype.valueOf.call(Pd(Boolean,[],(function(){}))),!0}catch(t){return!1}}();return function(){var i,n=Ld(t);if(e){var o=Ld(this).constructor;i=Pd(n,arguments,o)}else i=n.apply(this,arguments);return Fd(this,i)}}var Xf=function(t){Ad(i,t);var e=Yf(i);function i(t,n,o){var r;return Nn(this,i),(r=e.call(this,t,n,o)).via=r.via,r._boundFunction=function(){r.positionBezierNode()},r._body.emitter.on("_repositionBezierNodes",r._boundFunction),r}return Fn(i,[{key:"setOptions",value:function(t){mf(Ld(i.prototype),"setOptions",this).call(this,t);var e=!1;this.options.physics!==t.physics&&(e=!0),this.options=t,this.id=this.options.id,this.from=this._body.nodes[this.options.from],this.to=this._body.nodes[this.options.to],this.setupSupportNode(),this.connect(),!0===e&&(this.via.setOptions({physics:this.options.physics}),this.positionBezierNode())}},{key:"connect",value:function(){this.from=this._body.nodes[this.options.from],this.to=this._body.nodes[this.options.to],void 0===this.from||void 0===this.to||!1===this.options.physics||this.from.id===this.to.id?this.via.setOptions({physics:!1}):this.via.setOptions({physics:!0})}},{key:"cleanup",value:function(){return this._body.emitter.off("_repositionBezierNodes",this._boundFunction),void 0!==this.via&&(delete this._body.nodes[this.via.id],this.via=void 0,!0)}},{key:"setupSupportNode",value:function(){if(void 0===this.via){var t="edgeId:"+this.id,e=this._body.functions.createNode({id:t,shape:"circle",physics:!0,hidden:!0});this._body.nodes[t]=e,this.via=e,this.via.parentEdgeId=this.id,this.positionBezierNode()}}},{key:"positionBezierNode",value:function(){void 0!==this.via&&void 0!==this.from&&void 0!==this.to?(this.via.x=.5*(this.from.x+this.to.x),this.via.y=.5*(this.from.y+this.to.y)):void 0!==this.via&&(this.via.x=0,this.via.y=0)}},{key:"_line",value:function(t,e,i){this._bezierCurve(t,e,i)}},{key:"_getViaCoordinates",value:function(){return this.via}},{key:"getViaNode",value:function(){return this.via}},{key:"getPoint",value:function(t){var e=arguments.length>1&&void 0!==arguments[1]?arguments[1]:this.via;if(this.from===this.to){var i=this._getCircleData(),n=uo(i,3),o=n[0],r=n[1],s=n[2],a=2*Math.PI*(1-t);return{x:o+s*Math.sin(a),y:r+s-s*(1-Math.cos(a))}}return{x:Math.pow(1-t,2)*this.fromPoint.x+2*t*(1-t)*e.x+Math.pow(t,2)*this.toPoint.x,y:Math.pow(1-t,2)*this.fromPoint.y+2*t*(1-t)*e.y+Math.pow(t,2)*this.toPoint.y}}},{key:"_findBorderPosition",value:function(t,e){return this._findBorderPositionBezier(t,e,this.via)}},{key:"_getDistanceToEdge",value:function(t,e,i,n,o,r){return this._getDistanceToBezierEdge(t,e,i,n,o,r,this.via)}}]),i}(Uf);function Gf(t){var e=function(){if("undefined"==typeof Reflect||!Pd)return!1;if(Pd.sham)return!1;if("function"==typeof Proxy)return!0;try{return Boolean.prototype.valueOf.call(Pd(Boolean,[],(function(){}))),!0}catch(t){return!1}}();return function(){var i,n=Ld(t);if(e){var o=Ld(this).constructor;i=Pd(n,arguments,o)}else i=n.apply(this,arguments);return Fd(this,i)}}var Kf=function(t){Ad(i,t);var e=Gf(i);function i(t,n,o){return Nn(this,i),e.call(this,t,n,o)}return Fn(i,[{key:"_line",value:function(t,e,i){this._bezierCurve(t,e,i)}},{key:"getViaNode",value:function(){return this._getViaCoordinates()}},{key:"_getViaCoordinates",value:function(){var t,e,i=this.options.smooth.roundness,n=this.options.smooth.type,o=Math.abs(this.from.x-this.to.x),r=Math.abs(this.from.y-this.to.y);if("discrete"===n||"diagonalCross"===n){var s,a;s=a=o<=r?i*r:i*o,this.from.x>this.to.x&&(s=-s),this.from.y>=this.to.y&&(a=-a);var h=this.from.x+s,l=this.from.y+a;return"discrete"===n&&(o<=r?h=o<i*r?this.from.x:h:l=r<i*o?this.from.y:l),{x:h,y:l}}if("straightCross"===n){var d=(1-i)*o,c=(1-i)*r;return o<=r?(d=0,this.from.y<this.to.y&&(c=-c)):(this.from.x<this.to.x&&(d=-d),c=0),{x:this.to.x+d,y:this.to.y+c}}if("horizontal"===n){var u=(1-i)*o;return this.from.x<this.to.x&&(u=-u),{x:this.to.x+u,y:this.from.y}}if("vertical"===n){var f=(1-i)*r;return this.from.y<this.to.y&&(f=-f),{x:this.from.x,y:this.to.y+f}}if("curvedCW"===n){o=this.to.x-this.from.x,r=this.from.y-this.to.y;var p=Math.sqrt(o*o+r*r),v=Math.PI,g=(Math.atan2(r,o)+(.5*i+.5)*v)%(2*v);return{x:this.from.x+(.5*i+.5)*p*Math.sin(g),y:this.from.y+(.5*i+.5)*p*Math.cos(g)}}if("curvedCCW"===n){o=this.to.x-this.from.x,r=this.from.y-this.to.y;var y=Math.sqrt(o*o+r*r),m=Math.PI,b=(Math.atan2(r,o)+(.5*-i+.5)*m)%(2*m);return{x:this.from.x+(.5*i+.5)*y*Math.sin(b),y:this.from.y+(.5*i+.5)*y*Math.cos(b)}}t=e=o<=r?i*r:i*o,this.from.x>this.to.x&&(t=-t),this.from.y>=this.to.y&&(e=-e);var w=this.from.x+t,k=this.from.y+e;return o<=r?w=this.from.x<=this.to.x?this.to.x<w?this.to.x:w:this.to.x>w?this.to.x:w:k=this.from.y>=this.to.y?this.to.y>k?this.to.y:k:this.to.y<k?this.to.y:k,{x:w,y:k}}},{key:"_findBorderPosition",value:function(t,e){var i=arguments.length>2&&void 0!==arguments[2]?arguments[2]:{};return this._findBorderPositionBezier(t,e,i.via)}},{key:"_getDistanceToEdge",value:function(t,e,i,n,o,r){var s=arguments.length>6&&void 0!==arguments[6]?arguments[6]:this._getViaCoordinates();return this._getDistanceToBezierEdge(t,e,i,n,o,r,s)}},{key:"getPoint",value:function(t){var e=arguments.length>1&&void 0!==arguments[1]?arguments[1]:this._getViaCoordinates(),i=t,n=Math.pow(1-i,2)*this.fromPoint.x+2*i*(1-i)*e.x+Math.pow(i,2)*this.toPoint.x,o=Math.pow(1-i,2)*this.fromPoint.y+2*i*(1-i)*e.y+Math.pow(i,2)*this.toPoint.y;return{x:n,y:o}}}]),i}(Uf);function $f(t){var e=function(){if("undefined"==typeof Reflect||!Pd)return!1;if(Pd.sham)return!1;if("function"==typeof Proxy)return!0;try{return Boolean.prototype.valueOf.call(Pd(Boolean,[],(function(){}))),!0}catch(t){return!1}}();return function(){var i,n=Ld(t);if(e){var o=Ld(this).constructor;i=Pd(n,arguments,o)}else i=n.apply(this,arguments);return Fd(this,i)}}function Zf(t){var e=function(){if("undefined"==typeof Reflect||!Pd)return!1;if(Pd.sham)return!1;if("function"==typeof Proxy)return!0;try{return Boolean.prototype.valueOf.call(Pd(Boolean,[],(function(){}))),!0}catch(t){return!1}}();return function(){var i,n=Ld(t);if(e){var o=Ld(this).constructor;i=Pd(n,arguments,o)}else i=n.apply(this,arguments);return Fd(this,i)}}var Qf=function(t){Ad(i,t);var e=Zf(i);function i(t,n,o){return Nn(this,i),e.call(this,t,n,o)}return Fn(i,[{key:"_line",value:function(t,e,i){var n=i[0],o=i[1];this._bezierCurve(t,e,n,o)}},{key:"_getViaCoordinates",value:function(){var t,e,i,n,o=this.from.x-this.to.x,r=this.from.y-this.to.y,s=this.options.smooth.roundness;return(Math.abs(o)>Math.abs(r)||!0===this.options.smooth.forceDirection||"horizontal"===this.options.smooth.forceDirection)&&"vertical"!==this.options.smooth.forceDirection?(e=this.from.y,n=this.to.y,t=this.from.x-s*o,i=this.to.x+s*o):(e=this.from.y-s*r,n=this.to.y+s*r,t=this.from.x,i=this.to.x),[{x:t,y:e},{x:i,y:n}]}},{key:"getViaNode",value:function(){return this._getViaCoordinates()}},{key:"_findBorderPosition",value:function(t,e){return this._findBorderPositionBezier(t,e)}},{key:"_getDistanceToEdge",value:function(t,e,i,n,o,r){var s=arguments.length>6&&void 0!==arguments[6]?arguments[6]:this._getViaCoordinates(),a=uo(s,2),h=a[0],l=a[1];return this._getDistanceToBezierEdge2(t,e,i,n,o,r,h,l)}},{key:"getPoint",value:function(t){var e=arguments.length>1&&void 0!==arguments[1]?arguments[1]:this._getViaCoordinates(),i=uo(e,2),n=i[0],o=i[1],r=t,s=[Math.pow(1-r,3),3*r*Math.pow(1-r,2),3*Math.pow(r,2)*(1-r),Math.pow(r,3)],a=s[0]*this.fromPoint.x+s[1]*n.x+s[2]*o.x+s[3]*this.toPoint.x,h=s[0]*this.fromPoint.y+s[1]*n.y+s[2]*o.y+s[3]*this.toPoint.y;return{x:a,y:h}}}]),i}(function(t){Ad(i,t);var e=$f(i);function i(t,n,o){return Nn(this,i),e.call(this,t,n,o)}return Fn(i,[{key:"_getDistanceToBezierEdge2",value:function(t,e,i,n,o,r,s,a){for(var h=1e9,l=t,d=e,c=[0,0,0,0],u=1;u<10;u++){var f=.1*u;c[0]=Math.pow(1-f,3),c[1]=3*f*Math.pow(1-f,2),c[2]=3*Math.pow(f,2)*(1-f),c[3]=Math.pow(f,3);var p=c[0]*t+c[1]*s.x+c[2]*a.x+c[3]*i,v=c[0]*e+c[1]*s.y+c[2]*a.y+c[3]*n;if(u>0){var g=this._getDistanceToLine(l,d,p,v,o,r);h=g<h?g:h}l=p,d=v}return h}}]),i}(Uf));function Jf(t){var e=function(){if("undefined"==typeof Reflect||!Pd)return!1;if(Pd.sham)return!1;if("function"==typeof Proxy)return!0;try{return Boolean.prototype.valueOf.call(Pd(Boolean,[],(function(){}))),!0}catch(t){return!1}}();return function(){var i,n=Ld(t);if(e){var o=Ld(this).constructor;i=Pd(n,arguments,o)}else i=n.apply(this,arguments);return Fd(this,i)}}var tp=function(t){Ad(i,t);var e=Jf(i);function i(t,n,o){return Nn(this,i),e.call(this,t,n,o)}return Fn(i,[{key:"_line",value:function(t,e){t.beginPath(),t.moveTo(this.fromPoint.x,this.fromPoint.y),t.lineTo(this.toPoint.x,this.toPoint.y),this.enableShadow(t,e),t.stroke(),this.disableShadow(t,e)}},{key:"getViaNode",value:function(){}},{key:"getPoint",value:function(t){return{x:(1-t)*this.fromPoint.x+t*this.toPoint.x,y:(1-t)*this.fromPoint.y+t*this.toPoint.y}}},{key:"_findBorderPosition",value:function(t,e){var i=this.to,n=this.from;t.id===this.from.id&&(i=this.from,n=this.to);var o=Math.atan2(i.y-n.y,i.x-n.x),r=i.x-n.x,s=i.y-n.y,a=Math.sqrt(r*r+s*s),h=(a-t.distanceToBorder(e,o))/a;return{x:(1-h)*n.x+h*i.x,y:(1-h)*n.y+h*i.y,t:0}}},{key:"_getDistanceToEdge",value:function(t,e,i,n,o,r){return this._getDistanceToLine(t,e,i,n,o,r)}}]),i}(Hf),ep=function(){function t(e,i,n,o,r){if(Nn(this,t),void 0===i)throw new Error("No body provided");this.options=Kh(o),this.globalOptions=o,this.defaultOptions=r,this.body=i,this.imagelist=n,this.id=void 0,this.fromId=void 0,this.toId=void 0,this.selected=!1,this.hover=!1,this.labelDirty=!0,this.baseWidth=this.options.width,this.baseFontSize=this.options.font.size,this.from=void 0,this.to=void 0,this.edgeType=void 0,this.connected=!1,this.labelModule=new _u(this.body,this.options,!0),this.setOptions(e)}return Fn(t,[{key:"setOptions",value:function(e){if(e){var i=void 0!==e.physics&&this.options.physics!==e.physics||void 0!==e.hidden&&(this.options.hidden||!1)!==(e.hidden||!1)||void 0!==e.from&&this.options.from!==e.from||void 0!==e.to&&this.options.to!==e.to;t.parseOptions(this.options,e,!0,this.globalOptions),void 0!==e.id&&(this.id=e.id),void 0!==e.from&&(this.fromId=e.from),void 0!==e.to&&(this.toId=e.to),void 0!==e.title&&(this.title=e.title),void 0!==e.value&&(e.value=hu(e.value));var n=[e,this.options,this.defaultOptions];return this.chooser=fu("edge",n),this.updateLabelModule(e),i=this.updateEdgeType()||i,this._setInteractionWidths(),this.connect(),i}}},{key:"getFormattingValues",value:function(){var t=!0===this.options.arrows.to||!0===this.options.arrows.to.enabled,e=!0===this.options.arrows.from||!0===this.options.arrows.from.enabled,i=!0===this.options.arrows.middle||!0===this.options.arrows.middle.enabled,n=this.options.color.inherit,o={toArrow:t,toArrowScale:this.options.arrows.to.scaleFactor,toArrowType:this.options.arrows.to.type,toArrowSrc:this.options.arrows.to.src,toArrowImageWidth:this.options.arrows.to.imageWidth,toArrowImageHeight:this.options.arrows.to.imageHeight,middleArrow:i,middleArrowScale:this.options.arrows.middle.scaleFactor,middleArrowType:this.options.arrows.middle.type,middleArrowSrc:this.options.arrows.middle.src,middleArrowImageWidth:this.options.arrows.middle.imageWidth,middleArrowImageHeight:this.options.arrows.middle.imageHeight,fromArrow:e,fromArrowScale:this.options.arrows.from.scaleFactor,fromArrowType:this.options.arrows.from.type,fromArrowSrc:this.options.arrows.from.src,fromArrowImageWidth:this.options.arrows.from.imageWidth,fromArrowImageHeight:this.options.arrows.from.imageHeight,arrowStrikethrough:this.options.arrowStrikethrough,color:n?void 0:this.options.color.color,inheritsColor:n,opacity:this.options.color.opacity,hidden:this.options.hidden,length:this.options.length,shadow:this.options.shadow.enabled,shadowColor:this.options.shadow.color,shadowSize:this.options.shadow.size,shadowX:this.options.shadow.x,shadowY:this.options.shadow.y,dashes:this.options.dashes,width:this.options.width,background:this.options.background.enabled,backgroundColor:this.options.background.color,backgroundSize:this.options.background.size,backgroundDashes:this.options.background.dashes};if(this.selected||this.hover)if(!0===this.chooser){if(this.selected){var r=this.options.selectionWidth;"function"==typeof r?o.width=r(o.width):"number"==typeof r&&(o.width+=r),o.width=Math.max(o.width,.3/this.body.view.scale),o.color=this.options.color.highlight,o.shadow=this.options.shadow.enabled}else if(this.hover){var s=this.options.hoverWidth;"function"==typeof s?o.width=s(o.width):"number"==typeof s&&(o.width+=s),o.width=Math.max(o.width,.3/this.body.view.scale),o.color=this.options.color.hover,o.shadow=this.options.shadow.enabled}}else"function"==typeof this.chooser&&(this.chooser(o,this.options.id,this.selected,this.hover),void 0!==o.color&&(o.inheritsColor=!1),!1===o.shadow&&(o.shadowColor===this.options.shadow.color&&o.shadowSize===this.options.shadow.size&&o.shadowX===this.options.shadow.x&&o.shadowY===this.options.shadow.y||(o.shadow=!0)));else o.shadow=this.options.shadow.enabled,o.width=Math.max(o.width,.3/this.body.view.scale);return o}},{key:"updateLabelModule",value:function(t){var e=[t,this.options,this.globalOptions,this.defaultOptions];this.labelModule.update(this.options,e),void 0!==this.labelModule.baseSize&&(this.baseFontSize=this.labelModule.baseSize)}},{key:"updateEdgeType",value:function(){var t=this.options.smooth,e=!1,i=!0;return void 0!==this.edgeType&&((this.edgeType instanceof Xf&&!0===t.enabled&&"dynamic"===t.type||this.edgeType instanceof Qf&&!0===t.enabled&&"cubicBezier"===t.type||this.edgeType instanceof Kf&&!0===t.enabled&&"dynamic"!==t.type&&"cubicBezier"!==t.type||this.edgeType instanceof tp&&!1===t.type.enabled)&&(i=!1),!0===i&&(e=this.cleanup())),!0===i?!0===t.enabled?"dynamic"===t.type?(e=!0,this.edgeType=new Xf(this.options,this.body,this.labelModule)):"cubicBezier"===t.type?this.edgeType=new Qf(this.options,this.body,this.labelModule):this.edgeType=new Kf(this.options,this.body,this.labelModule):this.edgeType=new tp(this.options,this.body,this.labelModule):this.edgeType.setOptions(this.options),e}},{key:"connect",value:function(){this.disconnect(),this.from=this.body.nodes[this.fromId]||void 0,this.to=this.body.nodes[this.toId]||void 0,this.connected=void 0!==this.from&&void 0!==this.to,!0===this.connected?(this.from.attachEdge(this),this.to.attachEdge(this)):(this.from&&this.from.detachEdge(this),this.to&&this.to.detachEdge(this)),this.edgeType.connect()}},{key:"disconnect",value:function(){this.from&&(this.from.detachEdge(this),this.from=void 0),this.to&&(this.to.detachEdge(this),this.to=void 0),this.connected=!1}},{key:"getTitle",value:function(){return this.title}},{key:"isSelected",value:function(){return this.selected}},{key:"getValue",value:function(){return this.options.value}},{key:"setValueRange",value:function(t,e,i){if(void 0!==this.options.value){var n=this.options.scaling.customScalingFunction(t,e,i,this.options.value),o=this.options.scaling.max-this.options.scaling.min;if(!0===this.options.scaling.label.enabled){var r=this.options.scaling.label.max-this.options.scaling.label.min;this.options.font.size=this.options.scaling.label.min+n*r}this.options.width=this.options.scaling.min+n*o}else this.options.width=this.baseWidth,this.options.font.size=this.baseFontSize;this._setInteractionWidths(),this.updateLabelModule()}},{key:"_setInteractionWidths",value:function(){"function"==typeof this.options.hoverWidth?this.edgeType.hoverWidth=this.options.hoverWidth(this.options.width):this.edgeType.hoverWidth=this.options.hoverWidth+this.options.width,"function"==typeof this.options.selectionWidth?this.edgeType.selectionWidth=this.options.selectionWidth(this.options.width):this.edgeType.selectionWidth=this.options.selectionWidth+this.options.width}},{key:"draw",value:function(t){var e=this.getFormattingValues();if(!e.hidden){var i=this.edgeType.getViaNode();this.edgeType.drawLine(t,e,this.selected,this.hover,i),this.drawLabel(t,i)}}},{key:"drawArrows",value:function(t){var e=this.getFormattingValues();if(!e.hidden){var i=this.edgeType.getViaNode(),n={};this.edgeType.fromPoint=this.edgeType.from,this.edgeType.toPoint=this.edgeType.to,e.fromArrow&&(n.from=this.edgeType.getArrowData(t,"from",i,this.selected,this.hover,e),!1===e.arrowStrikethrough&&(this.edgeType.fromPoint=n.from.core),e.fromArrowSrc&&(n.from.image=this.imagelist.load(e.fromArrowSrc)),e.fromArrowImageWidth&&(n.from.imageWidth=e.fromArrowImageWidth),e.fromArrowImageHeight&&(n.from.imageHeight=e.fromArrowImageHeight)),e.toArrow&&(n.to=this.edgeType.getArrowData(t,"to",i,this.selected,this.hover,e),!1===e.arrowStrikethrough&&(this.edgeType.toPoint=n.to.core),e.toArrowSrc&&(n.to.image=this.imagelist.load(e.toArrowSrc)),e.toArrowImageWidth&&(n.to.imageWidth=e.toArrowImageWidth),e.toArrowImageHeight&&(n.to.imageHeight=e.toArrowImageHeight)),e.middleArrow&&(n.middle=this.edgeType.getArrowData(t,"middle",i,this.selected,this.hover,e),e.middleArrowSrc&&(n.middle.image=this.imagelist.load(e.middleArrowSrc)),e.middleArrowImageWidth&&(n.middle.imageWidth=e.middleArrowImageWidth),e.middleArrowImageHeight&&(n.middle.imageHeight=e.middleArrowImageHeight)),e.fromArrow&&this.edgeType.drawArrowHead(t,e,this.selected,this.hover,n.from),e.middleArrow&&this.edgeType.drawArrowHead(t,e,this.selected,this.hover,n.middle),e.toArrow&&this.edgeType.drawArrowHead(t,e,this.selected,this.hover,n.to)}}},{key:"drawLabel",value:function(t,e){if(void 0!==this.options.label){var i,n=this.from,o=this.to;if(this.labelModule.differentState(this.selected,this.hover)&&this.labelModule.getTextSize(t,this.selected,this.hover),n.id!=o.id){this.labelModule.pointToSelf=!1,i=this.edgeType.getPoint(.5,e),t.save();var r=this._getRotation(t);0!=r.angle&&(t.translate(r.x,r.y),t.rotate(r.angle)),this.labelModule.draw(t,i.x,i.y,this.selected,this.hover),t.restore()}else{this.labelModule.pointToSelf=!0;var s=gu(t,this.options.selfReference.angle,this.options.selfReference.size,n);i=this._pointOnCircle(s.x,s.y,this.options.selfReference.size,this.options.selfReference.angle),this.labelModule.draw(t,i.x,i.y,this.selected,this.hover)}}}},{key:"getItemsOnPoint",value:function(t){var e=[];if(this.labelModule.visible()){var i=this._getRotation();pu(this.labelModule.getSize(),t,i)&&e.push({edgeId:this.id,labelId:0})}var n={left:t.x,top:t.y};return this.isOverlappingWith(n)&&e.push({edgeId:this.id}),e}},{key:"isOverlappingWith",value:function(t){if(this.connected){var e=this.from.x,i=this.from.y,n=this.to.x,o=this.to.y,r=t.left,s=t.top;return this.edgeType.getDistanceToEdge(e,i,n,o,r,s)<10}return!1}},{key:"_getRotation",value:function(t){var e=this.edgeType.getViaNode(),i=this.edgeType.getPoint(.5,e);void 0!==t&&this.labelModule.calculateLabelSize(t,this.selected,this.hover,i.x,i.y);var n={x:i.x,y:this.labelModule.size.yLine,angle:0};if(!this.labelModule.visible())return n;if("horizontal"===this.options.font.align)return n;var o=this.from.y-this.to.y,r=this.from.x-this.to.x,s=Math.atan2(o,r);return(s<-1&&r<0||s>0&&r<0)&&(s+=Math.PI),n.angle=s,n}},{key:"_pointOnCircle",value:function(t,e,i,n){return{x:t+i*Math.cos(n),y:e-i*Math.sin(n)}}},{key:"select",value:function(){this.selected=!0}},{key:"unselect",value:function(){this.selected=!1}},{key:"cleanup",value:function(){return this.edgeType.cleanup()}},{key:"remove",value:function(){this.cleanup(),this.disconnect(),delete this.body.edges[this.id]}},{key:"endPointsValid",value:function(){return void 0!==this.body.nodes[this.fromId]&&void 0!==this.body.nodes[this.toId]}}],[{key:"parseOptions",value:function(t,e){var i=arguments.length>2&&void 0!==arguments[2]&&arguments[2],n=arguments.length>3&&void 0!==arguments[3]?arguments[3]:{},o=arguments.length>4&&void 0!==arguments[4]&&arguments[4],r=["endPointOffset","arrowStrikethrough","id","from","hidden","hoverWidth","labelHighlightBold","length","line","opacity","physics","scaling","selectionWidth","selfReferenceSize","selfReference","to","title","value","width","font","chosen","widthConstraint"];if(Eh(r,t,e,i),void 0!==e.endPointOffset&&void 0!==e.endPointOffset.from&&(wd(e.endPointOffset.from)?t.endPointOffset.from=e.endPointOffset.from:(t.endPointOffset.from=void 0!==n.endPointOffset.from?n.endPointOffset.from:0,console.error("endPointOffset.from is not a valid number"))),void 0!==e.endPointOffset&&void 0!==e.endPointOffset.to&&(wd(e.endPointOffset.to)?t.endPointOffset.to=e.endPointOffset.to:(t.endPointOffset.to=void 0!==n.endPointOffset.to?n.endPointOffset.to:0,console.error("endPointOffset.to is not a valid number"))),vu(e.label)?t.label=e.label:vu(t.label)||(t.label=void 0),$h(t,e,"smooth",n),$h(t,e,"shadow",n),$h(t,e,"background",n),void 0!==e.dashes&&null!==e.dashes?t.dashes=e.dashes:!0===i&&null===e.dashes&&(t.dashes=Gr(n.dashes)),void 0!==e.scaling&&null!==e.scaling?(void 0!==e.scaling.min&&(t.scaling.min=e.scaling.min),void 0!==e.scaling.max&&(t.scaling.max=e.scaling.max),$h(t.scaling,e.scaling,"label",n.scaling)):!0===i&&null===e.scaling&&(t.scaling=Gr(n.scaling)),void 0!==e.arrows&&null!==e.arrows)if("string"==typeof e.arrows){var s=e.arrows.toLowerCase();t.arrows.to.enabled=-1!=Hr(s).call(s,"to"),t.arrows.middle.enabled=-1!=Hr(s).call(s,"middle"),t.arrows.from.enabled=-1!=Hr(s).call(s,"from")}else{if("object"!==go(e.arrows))throw new Error("The arrow newOptions can only be an object or a string. Refer to the documentation. You used:"+es(e.arrows));$h(t.arrows,e.arrows,"to",n.arrows),$h(t.arrows,e.arrows,"middle",n.arrows),$h(t.arrows,e.arrows,"from",n.arrows)}else!0===i&&null===e.arrows&&(t.arrows=Gr(n.arrows));if(void 0!==e.color&&null!==e.color){var a=bh(e.color)?{color:e.color,highlight:e.color,hover:e.color,inherit:!1,opacity:1}:e.color,h=t.color;if(o)Ch(h,n.color,!1,i);else for(var l in h)Object.prototype.hasOwnProperty.call(h,l)&&delete h[l];if(bh(h))h.color=h,h.highlight=h,h.hover=h,h.inherit=!1,void 0===a.opacity&&(h.opacity=1);else{var d=!1;void 0!==a.color&&(h.color=a.color,d=!0),void 0!==a.highlight&&(h.highlight=a.highlight,d=!0),void 0!==a.hover&&(h.hover=a.hover,d=!0),void 0!==a.inherit&&(h.inherit=a.inherit),void 0!==a.opacity&&(h.opacity=Math.min(1,Math.max(0,a.opacity))),!0===d?h.inherit=!1:void 0===h.inherit&&(h.inherit="from")}}else!0===i&&null===e.color&&(t.color=Kh(n.color));!0===i&&null===e.font&&(t.font=Kh(n.font)),Object.prototype.hasOwnProperty.call(e,"selfReferenceSize")&&(console.warn("The selfReferenceSize property has been deprecated. Please use selfReference property instead. The selfReference can be set like thise selfReference:{size:30, angle:Math.PI / 4}"),t.selfReference.size=e.selfReferenceSize)}}]),t}(),ip=function(){function t(e,i,n){var o,r=this;Nn(this,t),this.body=e,this.images=i,this.groups=n,this.body.functions.createEdge=Vt(o=this.create).call(o,this),this.edgesListeners={add:function(t,e){r.add(e.items)},update:function(t,e){r.update(e.items)},remove:function(t,e){r.remove(e.items)}},this.options={},this.defaultOptions={arrows:{to:{enabled:!1,scaleFactor:1,type:"arrow"},middle:{enabled:!1,scaleFactor:1,type:"arrow"},from:{enabled:!1,scaleFactor:1,type:"arrow"}},endPointOffset:{from:0,to:0},arrowStrikethrough:!0,color:{color:"#848484",highlight:"#848484",hover:"#848484",inherit:"from",opacity:1},dashes:!1,font:{color:"#343434",size:14,face:"arial",background:"none",strokeWidth:2,strokeColor:"#ffffff",align:"horizontal",multi:!1,vadjust:0,bold:{mod:"bold"},boldital:{mod:"bold italic"},ital:{mod:"italic"},mono:{mod:"",size:15,face:"courier new",vadjust:2}},hidden:!1,hoverWidth:1.5,label:void 0,labelHighlightBold:!0,length:void 0,physics:!0,scaling:{min:1,max:15,label:{enabled:!0,min:14,max:30,maxVisible:30,drawThreshold:5},customScalingFunction:function(t,e,i,n){if(e===t)return.5;var o=1/(e-t);return Math.max(0,(n-t)*o)}},selectionWidth:1.5,selfReference:{size:20,angle:Math.PI/4,renderBehindTheNode:!0},shadow:{enabled:!1,color:"rgba(0,0,0,0.5)",size:10,x:5,y:5},background:{enabled:!1,color:"rgba(111,111,111,1)",size:10,dashes:!1},smooth:{enabled:!0,type:"dynamic",forceDirection:"none",roundness:.5},title:void 0,width:1,value:void 0},Ch(this.options,this.defaultOptions),this.bindEventListeners()}return Fn(t,[{key:"bindEventListeners",value:function(){var t,e,i=this;this.body.emitter.on("_forceDisableDynamicCurves",(function(t){var e=!(arguments.length>1&&void 0!==arguments[1])||arguments[1];"dynamic"===t&&(t="continuous");var n=!1;for(var o in i.body.edges)if(Object.prototype.hasOwnProperty.call(i.body.edges,o)){var r=i.body.edges[o],s=i.body.data.edges.get(o);if(null!=s){var a=s.smooth;void 0!==a&&!0===a.enabled&&"dynamic"===a.type&&(void 0===t?r.setOptions({smooth:!1}):r.setOptions({smooth:{type:t}}),n=!0)}}!0===e&&!0===n&&i.body.emitter.emit("_dataChanged")})),this.body.emitter.on("_dataUpdated",(function(){i.reconnectEdges()})),this.body.emitter.on("refreshEdges",Vt(t=this.refresh).call(t,this)),this.body.emitter.on("refresh",Vt(e=this.refresh).call(e,this)),this.body.emitter.on("destroy",(function(){Dh(i.edgesListeners,(function(t,e){i.body.data.edges&&i.body.data.edges.off(e,t)})),delete i.body.functions.createEdge,delete i.edgesListeners.add,delete i.edgesListeners.update,delete i.edgesListeners.remove,delete i.edgesListeners}))}},{key:"setOptions",value:function(t){if(void 0!==t){ep.parseOptions(this.options,t,!0,this.defaultOptions,!0);var e=!1;if(void 0!==t.smooth)for(var i in this.body.edges)Object.prototype.hasOwnProperty.call(this.body.edges,i)&&(e=this.body.edges[i].updateEdgeType()||e);if(void 0!==t.font)for(var n in this.body.edges)Object.prototype.hasOwnProperty.call(this.body.edges,n)&&this.body.edges[n].updateLabelModule();void 0===t.hidden&&void 0===t.physics&&!0!==e||this.body.emitter.emit("_dataChanged")}}},{key:"setData",value:function(t){var e=this,i=arguments.length>1&&void 0!==arguments[1]&&arguments[1],n=this.body.data.edges;if(nu("id",t))this.body.data.edges=t;else if(So(t))this.body.data.edges=new tu,this.body.data.edges.add(t);else{if(t)throw new TypeError("Array or DataSet expected");this.body.data.edges=new tu}if(n&&Dh(this.edgesListeners,(function(t,e){n.off(e,t)})),this.body.edges={},this.body.data.edges){Dh(this.edgesListeners,(function(t,i){e.body.data.edges.on(i,t)}));var o=this.body.data.edges.getIds();this.add(o,!0)}this.body.emitter.emit("_adjustEdgesForHierarchicalLayout"),!1===i&&this.body.emitter.emit("_dataChanged")}},{key:"add",value:function(t){for(var e=arguments.length>1&&void 0!==arguments[1]&&arguments[1],i=this.body.edges,n=this.body.data.edges,o=0;o<t.length;o++){var r=t[o],s=i[r];s&&s.disconnect();var a=n.get(r,{showInternalIds:!0});i[r]=this.create(a)}this.body.emitter.emit("_adjustEdgesForHierarchicalLayout"),!1===e&&this.body.emitter.emit("_dataChanged")}},{key:"update",value:function(t){for(var e=this.body.edges,i=this.body.data.edges,n=!1,o=0;o<t.length;o++){var r=t[o],s=i.get(r),a=e[r];void 0!==a?(a.disconnect(),n=a.setOptions(s)||n,a.connect()):(this.body.edges[r]=this.create(s),n=!0)}!0===n?(this.body.emitter.emit("_adjustEdgesForHierarchicalLayout"),this.body.emitter.emit("_dataChanged")):this.body.emitter.emit("_dataUpdated")}},{key:"remove",value:function(t){var e=!(arguments.length>1&&void 0!==arguments[1])||arguments[1];if(0!==t.length){var i=this.body.edges;Dh(t,(function(t){var e=i[t];void 0!==e&&e.remove()})),e&&this.body.emitter.emit("_dataChanged")}}},{key:"refresh",value:function(){var t=this;Dh(this.body.edges,(function(e,i){var n=t.body.data.edges.get(i);void 0!==n&&e.setOptions(n)}))}},{key:"create",value:function(t){return new ep(t,this.body,this.images,this.options,this.defaultOptions)}},{key:"reconnectEdges",value:function(){var t,e=this.body.nodes,i=this.body.edges;for(t in e)Object.prototype.hasOwnProperty.call(e,t)&&(e[t].edges=[]);for(t in i)if(Object.prototype.hasOwnProperty.call(i,t)){var n=i[t];n.from=null,n.to=null,n.connect()}}},{key:"getConnectedNodes",value:function(t){var e=[];if(void 0!==this.body.edges[t]){var i=this.body.edges[t];void 0!==i.fromId&&e.push(i.fromId),void 0!==i.toId&&e.push(i.toId)}return e}},{key:"_updateState",value:function(){this._addMissingEdges(),this._removeInvalidEdges()}},{key:"_removeInvalidEdges",value:function(){var t=this,e=[];Dh(this.body.edges,(function(i,n){var o=t.body.nodes[i.toId],r=t.body.nodes[i.fromId];void 0!==o&&!0===o.isCluster||void 0!==r&&!0===r.isCluster||void 0!==o&&void 0!==r||e.push(n)})),this.remove(e,!1)}},{key:"_addMissingEdges",value:function(){var t=this.body.data.edges;if(null!=t){var e=this.body.edges,i=[];Wo(t).call(t,(function(t,n){void 0===e[n]&&i.push(n)})),this.add(i,!0)}}}]),t}(),np=function(){function t(e,i,n){Nn(this,t),this.body=e,this.physicsBody=i,this.barnesHutTree,this.setOptions(n),this._rng=ah("BARNES HUT SOLVER")}return Fn(t,[{key:"setOptions",value:function(t){this.options=t,this.thetaInversed=1/this.options.theta,this.overlapAvoidanceFactor=1-Math.max(0,Math.min(1,this.options.avoidOverlap))}},{key:"solve",value:function(){if(0!==this.options.gravitationalConstant&&this.physicsBody.physicsNodeIndices.length>0){var t,e=this.body.nodes,i=this.physicsBody.physicsNodeIndices,n=i.length,o=this._formBarnesHutTree(e,i);this.barnesHutTree=o;for(var r=0;r<n;r++)(t=e[i[r]]).options.mass>0&&this._getForceContributions(o.root,t)}}},{key:"_getForceContributions",value:function(t,e){this._getForceContribution(t.children.NW,e),this._getForceContribution(t.children.NE,e),this._getForceContribution(t.children.SW,e),this._getForceContribution(t.children.SE,e)}},{key:"_getForceContribution",value:function(t,e){if(t.childrenCount>0){var i=t.centerOfMass.x-e.x,n=t.centerOfMass.y-e.y,o=Math.sqrt(i*i+n*n);o*t.calcSize>this.thetaInversed?this._calculateForces(o,i,n,e,t):4===t.childrenCount?this._getForceContributions(t,e):t.children.data.id!=e.id&&this._calculateForces(o,i,n,e,t)}}},{key:"_calculateForces",value:function(t,e,i,n,o){0===t&&(e=t=.1),this.overlapAvoidanceFactor<1&&n.shape.radius&&(t=Math.max(.1+this.overlapAvoidanceFactor*n.shape.radius,t-n.shape.radius));var r=this.options.gravitationalConstant*o.mass*n.options.mass/Math.pow(t,3),s=e*r,a=i*r;this.physicsBody.forces[n.id].x+=s,this.physicsBody.forces[n.id].y+=a}},{key:"_formBarnesHutTree",value:function(t,e){for(var i,n=e.length,o=t[e[0]].x,r=t[e[0]].y,s=t[e[0]].x,a=t[e[0]].y,h=1;h<n;h++){var l=t[e[h]],d=l.x,c=l.y;l.options.mass>0&&(d<o&&(o=d),d>s&&(s=d),c<r&&(r=c),c>a&&(a=c))}var u=Math.abs(s-o)-Math.abs(a-r);u>0?(r-=.5*u,a+=.5*u):(o+=.5*u,s-=.5*u);var f=Math.max(1e-5,Math.abs(s-o)),p=.5*f,v=.5*(o+s),g=.5*(r+a),y={root:{centerOfMass:{x:0,y:0},mass:0,range:{minX:v-p,maxX:v+p,minY:g-p,maxY:g+p},size:f,calcSize:1/f,children:{data:null},maxWidth:0,level:0,childrenCount:4}};this._splitBranch(y.root);for(var m=0;m<n;m++)(i=t[e[m]]).options.mass>0&&this._placeInTree(y.root,i);return y}},{key:"_updateBranchMass",value:function(t,e){var i=t.centerOfMass,n=t.mass+e.options.mass,o=1/n;i.x=i.x*t.mass+e.x*e.options.mass,i.x*=o,i.y=i.y*t.mass+e.y*e.options.mass,i.y*=o,t.mass=n;var r=Math.max(Math.max(e.height,e.radius),e.width);t.maxWidth=t.maxWidth<r?r:t.maxWidth}},{key:"_placeInTree",value:function(t,e,i){1==i&&void 0!==i||this._updateBranchMass(t,e);var n,o=t.children.NW.range;n=o.maxX>e.x?o.maxY>e.y?"NW":"SW":o.maxY>e.y?"NE":"SE",this._placeInRegion(t,e,n)}},{key:"_placeInRegion",value:function(t,e,i){var n=t.children[i];switch(n.childrenCount){case 0:n.children.data=e,n.childrenCount=1,this._updateBranchMass(n,e);break;case 1:n.children.data.x===e.x&&n.children.data.y===e.y?(e.x+=this._rng(),e.y+=this._rng()):(this._splitBranch(n),this._placeInTree(n,e));break;case 4:this._placeInTree(n,e)}}},{key:"_splitBranch",value:function(t){var e=null;1===t.childrenCount&&(e=t.children.data,t.mass=0,t.centerOfMass.x=0,t.centerOfMass.y=0),t.childrenCount=4,t.children.data=null,this._insertRegion(t,"NW"),this._insertRegion(t,"NE"),this._insertRegion(t,"SW"),this._insertRegion(t,"SE"),null!=e&&this._placeInTree(t,e)}},{key:"_insertRegion",value:function(t,e){var i,n,o,r,s=.5*t.size;switch(e){case"NW":i=t.range.minX,n=t.range.minX+s,o=t.range.minY,r=t.range.minY+s;break;case"NE":i=t.range.minX+s,n=t.range.maxX,o=t.range.minY,r=t.range.minY+s;break;case"SW":i=t.range.minX,n=t.range.minX+s,o=t.range.minY+s,r=t.range.maxY;break;case"SE":i=t.range.minX+s,n=t.range.maxX,o=t.range.minY+s,r=t.range.maxY}t.children[e]={centerOfMass:{x:0,y:0},mass:0,range:{minX:i,maxX:n,minY:o,maxY:r},size:.5*t.size,calcSize:2*t.calcSize,children:{data:null},maxWidth:0,level:t.level+1,childrenCount:0}}},{key:"_debug",value:function(t,e){void 0!==this.barnesHutTree&&(t.lineWidth=1,this._drawBranch(this.barnesHutTree.root,t,e))}},{key:"_drawBranch",value:function(t,e,i){void 0===i&&(i="#FF0000"),4===t.childrenCount&&(this._drawBranch(t.children.NW,e),this._drawBranch(t.children.NE,e),this._drawBranch(t.children.SE,e),this._drawBranch(t.children.SW,e)),e.strokeStyle=i,e.beginPath(),e.moveTo(t.range.minX,t.range.minY),e.lineTo(t.range.maxX,t.range.minY),e.stroke(),e.beginPath(),e.moveTo(t.range.maxX,t.range.minY),e.lineTo(t.range.maxX,t.range.maxY),e.stroke(),e.beginPath(),e.moveTo(t.range.maxX,t.range.maxY),e.lineTo(t.range.minX,t.range.maxY),e.stroke(),e.beginPath(),e.moveTo(t.range.minX,t.range.maxY),e.lineTo(t.range.minX,t.range.minY),e.stroke()}}]),t}(),op=function(){function t(e,i,n){Nn(this,t),this._rng=ah("REPULSION SOLVER"),this.body=e,this.physicsBody=i,this.setOptions(n)}return Fn(t,[{key:"setOptions",value:function(t){this.options=t}},{key:"solve",value:function(){for(var t,e,i,n,o,r,s,a,h=this.body.nodes,l=this.physicsBody.physicsNodeIndices,d=this.physicsBody.forces,c=this.options.nodeDistance,u=-2/3/c,f=0;f<l.length-1;f++){s=h[l[f]];for(var p=f+1;p<l.length;p++)t=(a=h[l[p]]).x-s.x,e=a.y-s.y,0===(i=Math.sqrt(t*t+e*e))&&(t=i=.1*this._rng()),i<2*c&&(r=i<.5*c?1:u*i+1.3333333333333333,n=t*(r/=i),o=e*r,d[s.id].x-=n,d[s.id].y-=o,d[a.id].x+=n,d[a.id].y+=o)}}}]),t}(),rp=function(){function t(e,i,n){Nn(this,t),this.body=e,this.physicsBody=i,this.setOptions(n)}return Fn(t,[{key:"setOptions",value:function(t){this.options=t,this.overlapAvoidanceFactor=Math.max(0,Math.min(1,this.options.avoidOverlap||0))}},{key:"solve",value:function(){for(var t=this.body.nodes,e=this.physicsBody.physicsNodeIndices,i=this.physicsBody.forces,n=this.options.nodeDistance,o=0;o<e.length-1;o++)for(var r=t[e[o]],s=o+1;s<e.length;s++){var a=t[e[s]];if(r.level===a.level){var h=n+this.overlapAvoidanceFactor*((r.shape.radius||0)/2+(a.shape.radius||0)/2),l=a.x-r.x,d=a.y-r.y,c=Math.sqrt(l*l+d*d),u=void 0;u=c<h?-Math.pow(.05*c,2)+Math.pow(.05*h,2):0,0!==c&&(u/=c);var f=l*u,p=d*u;i[r.id].x-=f,i[r.id].y-=p,i[a.id].x+=f,i[a.id].y+=p}}}}]),t}(),sp=function(){function t(e,i,n){Nn(this,t),this.body=e,this.physicsBody=i,this.setOptions(n)}return Fn(t,[{key:"setOptions",value:function(t){this.options=t}},{key:"solve",value:function(){for(var t,e,i,n,o,r=this.physicsBody.physicsEdgeIndices,s=this.body.edges,a=0;a<r.length;a++)!0===(e=s[r[a]]).connected&&e.toId!==e.fromId&&void 0!==this.body.nodes[e.toId]&&void 0!==this.body.nodes[e.fromId]&&(void 0!==e.edgeType.via?(t=void 0===e.options.length?this.options.springLength:e.options.length,i=e.to,n=e.edgeType.via,o=e.from,this._calculateSpringForce(i,n,.5*t),this._calculateSpringForce(n,o,.5*t)):(t=void 0===e.options.length?1.5*this.options.springLength:e.options.length,this._calculateSpringForce(e.from,e.to,t)))}},{key:"_calculateSpringForce",value:function(t,e,i){var n=t.x-e.x,o=t.y-e.y,r=Math.max(Math.sqrt(n*n+o*o),.01),s=this.options.springConstant*(i-r)/r,a=n*s,h=o*s;void 0!==this.physicsBody.forces[t.id]&&(this.physicsBody.forces[t.id].x+=a,this.physicsBody.forces[t.id].y+=h),void 0!==this.physicsBody.forces[e.id]&&(this.physicsBody.forces[e.id].x-=a,this.physicsBody.forces[e.id].y-=h)}}]),t}(),ap=function(){function t(e,i,n){Nn(this,t),this.body=e,this.physicsBody=i,this.setOptions(n)}return Fn(t,[{key:"setOptions",value:function(t){this.options=t}},{key:"solve",value:function(){for(var t,e,i,n,o,r,s,a,h,l,d=this.body.edges,c=.5,u=this.physicsBody.physicsEdgeIndices,f=this.physicsBody.physicsNodeIndices,p=this.physicsBody.forces,v=0;v<f.length;v++){var g=f[v];p[g].springFx=0,p[g].springFy=0}for(var y=0;y<u.length;y++)!0===(e=d[u[y]]).connected&&(t=void 0===e.options.length?this.options.springLength:e.options.length,i=e.from.x-e.to.x,n=e.from.y-e.to.y,a=0===(a=Math.sqrt(i*i+n*n))?.01:a,o=i*(s=this.options.springConstant*(t-a)/a),r=n*s,e.to.level!=e.from.level?(void 0!==p[e.toId]&&(p[e.toId].springFx-=o,p[e.toId].springFy-=r),void 0!==p[e.fromId]&&(p[e.fromId].springFx+=o,p[e.fromId].springFy+=r)):(void 0!==p[e.toId]&&(p[e.toId].x-=c*o,p[e.toId].y-=c*r),void 0!==p[e.fromId]&&(p[e.fromId].x+=c*o,p[e.fromId].y+=c*r)));s=1;for(var m=0;m<f.length;m++){var b=f[m];h=Math.min(s,Math.max(-s,p[b].springFx)),l=Math.min(s,Math.max(-s,p[b].springFy)),p[b].x+=h,p[b].y+=l}for(var w=0,k=0,_=0;_<f.length;_++){var x=f[_];w+=p[x].x,k+=p[x].y}for(var E=w/f.length,O=k/f.length,C=0;C<f.length;C++){var S=f[C];p[S].x-=E,p[S].y-=O}}}]),t}(),hp=function(){function t(e,i,n){Nn(this,t),this.body=e,this.physicsBody=i,this.setOptions(n)}return Fn(t,[{key:"setOptions",value:function(t){this.options=t}},{key:"solve",value:function(){for(var t,e,i,n,o=this.body.nodes,r=this.physicsBody.physicsNodeIndices,s=this.physicsBody.forces,a=0;a<r.length;a++){t=-(n=o[r[a]]).x,e=-n.y,i=Math.sqrt(t*t+e*e),this._calculateForces(i,t,e,s,n)}}},{key:"_calculateForces",value:function(t,e,i,n,o){var r=0===t?0:this.options.centralGravity/t;n[o.id].x=e*r,n[o.id].y=i*r}}]),t}();function lp(t){var e=function(){if("undefined"==typeof Reflect||!Pd)return!1;if(Pd.sham)return!1;if("function"==typeof Proxy)return!0;try{return Boolean.prototype.valueOf.call(Pd(Boolean,[],(function(){}))),!0}catch(t){return!1}}();return function(){var i,n=Ld(t);if(e){var o=Ld(this).constructor;i=Pd(n,arguments,o)}else i=n.apply(this,arguments);return Fd(this,i)}}var dp=function(t){Ad(i,t);var e=lp(i);function i(t,n,o){var r;return Nn(this,i),(r=e.call(this,t,n,o))._rng=ah("FORCE ATLAS 2 BASED REPULSION SOLVER"),r}return Fn(i,[{key:"_calculateForces",value:function(t,e,i,n,o){0===t&&(e=t=.1*this._rng()),this.overlapAvoidanceFactor<1&&n.shape.radius&&(t=Math.max(.1+this.overlapAvoidanceFactor*n.shape.radius,t-n.shape.radius));var r=n.edges.length+1,s=this.options.gravitationalConstant*o.mass*n.options.mass*r/Math.pow(t,2),a=e*s,h=i*s;this.physicsBody.forces[n.id].x+=a,this.physicsBody.forces[n.id].y+=h}}]),i}(np);function cp(t){var e=function(){if("undefined"==typeof Reflect||!Pd)return!1;if(Pd.sham)return!1;if("function"==typeof Proxy)return!0;try{return Boolean.prototype.valueOf.call(Pd(Boolean,[],(function(){}))),!0}catch(t){return!1}}();return function(){var i,n=Ld(t);if(e){var o=Ld(this).constructor;i=Pd(n,arguments,o)}else i=n.apply(this,arguments);return Fd(this,i)}}var up=function(t){Ad(i,t);var e=cp(i);function i(t,n,o){return Nn(this,i),e.call(this,t,n,o)}return Fn(i,[{key:"_calculateForces",value:function(t,e,i,n,o){if(t>0){var r=o.edges.length+1,s=this.options.centralGravity*r*o.options.mass;n[o.id].x=e*s,n[o.id].y=i*s}}}]),i}(hp),fp=function(){function t(e){Nn(this,t),this.body=e,this.physicsBody={physicsNodeIndices:[],physicsEdgeIndices:[],forces:{},velocities:{}},this.physicsEnabled=!0,this.simulationInterval=1e3/60,this.requiresTimeout=!0,this.previousStates={},this.referenceState={},this.freezeCache={},this.renderTimer=void 0,this.adaptiveTimestep=!1,this.adaptiveTimestepEnabled=!1,this.adaptiveCounter=0,this.adaptiveInterval=3,this.stabilized=!1,this.startedStabilization=!1,this.stabilizationIterations=0,this.ready=!1,this.options={},this.defaultOptions={enabled:!0,barnesHut:{theta:.5,gravitationalConstant:-2e3,centralGravity:.3,springLength:95,springConstant:.04,damping:.09,avoidOverlap:0},forceAtlas2Based:{theta:.5,gravitationalConstant:-50,centralGravity:.01,springConstant:.08,springLength:100,damping:.4,avoidOverlap:0},repulsion:{centralGravity:.2,springLength:200,springConstant:.05,nodeDistance:100,damping:.09,avoidOverlap:0},hierarchicalRepulsion:{centralGravity:0,springLength:100,springConstant:.01,nodeDistance:120,damping:.09},maxVelocity:50,minVelocity:.75,solver:"barnesHut",stabilization:{enabled:!0,iterations:1e3,updateInterval:50,onlyDynamicEdges:!1,fit:!0},timestep:.5,adaptiveTimestep:!0,wind:{x:0,y:0}},At(this.options,this.defaultOptions),this.timestep=.5,this.layoutFailed=!1,this.bindEventListeners()}return Fn(t,[{key:"bindEventListeners",value:function(){var t=this;this.body.emitter.on("initPhysics",(function(){t.initPhysics()})),this.body.emitter.on("_layoutFailed",(function(){t.layoutFailed=!0})),this.body.emitter.on("resetPhysics",(function(){t.stopSimulation(),t.ready=!1})),this.body.emitter.on("disablePhysics",(function(){t.physicsEnabled=!1,t.stopSimulation()})),this.body.emitter.on("restorePhysics",(function(){t.setOptions(t.options),!0===t.ready&&t.startSimulation()})),this.body.emitter.on("startSimulation",(function(){!0===t.ready&&t.startSimulation()})),this.body.emitter.on("stopSimulation",(function(){t.stopSimulation()})),this.body.emitter.on("destroy",(function(){t.stopSimulation(!1),t.body.emitter.off()})),this.body.emitter.on("_dataChanged",(function(){t.updatePhysicsData()}))}},{key:"setOptions",value:function(t){if(void 0!==t)if(!1===t)this.options.enabled=!1,this.physicsEnabled=!1,this.stopSimulation();else if(!0===t)this.options.enabled=!0,this.physicsEnabled=!0,this.startSimulation();else{this.physicsEnabled=!0,Oh(["stabilization"],this.options,t),$h(this.options,t,"stabilization"),void 0===t.enabled&&(this.options.enabled=!0),!1===this.options.enabled&&(this.physicsEnabled=!1,this.stopSimulation());var e=this.options.wind;e&&(("number"!=typeof e.x||yd(e.x))&&(e.x=0),("number"!=typeof e.y||yd(e.y))&&(e.y=0)),this.timestep=this.options.timestep}this.init()}},{key:"init",value:function(){var t;"forceAtlas2Based"===this.options.solver?(t=this.options.forceAtlas2Based,this.nodesSolver=new dp(this.body,this.physicsBody,t),this.edgesSolver=new sp(this.body,this.physicsBody,t),this.gravitySolver=new up(this.body,this.physicsBody,t)):"repulsion"===this.options.solver?(t=this.options.repulsion,this.nodesSolver=new op(this.body,this.physicsBody,t),this.edgesSolver=new sp(this.body,this.physicsBody,t),this.gravitySolver=new hp(this.body,this.physicsBody,t)):"hierarchicalRepulsion"===this.options.solver?(t=this.options.hierarchicalRepulsion,this.nodesSolver=new rp(this.body,this.physicsBody,t),this.edgesSolver=new ap(this.body,this.physicsBody,t),this.gravitySolver=new hp(this.body,this.physicsBody,t)):(t=this.options.barnesHut,this.nodesSolver=new np(this.body,this.physicsBody,t),this.edgesSolver=new sp(this.body,this.physicsBody,t),this.gravitySolver=new hp(this.body,this.physicsBody,t)),this.modelOptions=t}},{key:"initPhysics",value:function(){!0===this.physicsEnabled&&!0===this.options.enabled?!0===this.options.stabilization.enabled?this.stabilize():(this.stabilized=!1,this.ready=!0,this.body.emitter.emit("fit",{},this.layoutFailed),this.startSimulation()):(this.ready=!0,this.body.emitter.emit("fit"))}},{key:"startSimulation",value:function(){var t;!0===this.physicsEnabled&&!0===this.options.enabled?(this.stabilized=!1,this.adaptiveTimestep=!1,this.body.emitter.emit("_resizeNodes"),void 0===this.viewFunction&&(this.viewFunction=Vt(t=this.simulationStep).call(t,this),this.body.emitter.on("initRedraw",this.viewFunction),this.body.emitter.emit("_startRendering"))):this.body.emitter.emit("_redraw")}},{key:"stopSimulation",value:function(){var t=!(arguments.length>0&&void 0!==arguments[0])||arguments[0];this.stabilized=!0,!0===t&&this._emitStabilized(),void 0!==this.viewFunction&&(this.body.emitter.off("initRedraw",this.viewFunction),this.viewFunction=void 0,!0===t&&this.body.emitter.emit("_stopRendering"))}},{key:"simulationStep",value:function(){var t=No();this.physicsTick(),(No()-t<.4*this.simulationInterval||!0===this.runDoubleSpeed)&&!1===this.stabilized&&(this.physicsTick(),this.runDoubleSpeed=!0),!0===this.stabilized&&this.stopSimulation()}},{key:"_emitStabilized",value:function(){var t=this,e=arguments.length>0&&void 0!==arguments[0]?arguments[0]:this.stabilizationIterations;(this.stabilizationIterations>1||!0===this.startedStabilization)&&rs((function(){t.body.emitter.emit("stabilized",{iterations:e}),t.startedStabilization=!1,t.stabilizationIterations=0}),0)}},{key:"physicsStep",value:function(){this.gravitySolver.solve(),this.nodesSolver.solve(),this.edgesSolver.solve(),this.moveNodes()}},{key:"adjustTimeStep",value:function(){!0===this._evaluateStepQuality()?this.timestep=1.2*this.timestep:this.timestep/1.2<this.options.timestep?this.timestep=this.options.timestep:(this.adaptiveCounter=-1,this.timestep=Math.max(this.options.timestep,this.timestep/1.2))}},{key:"physicsTick",value:function(){if(this._startStabilizing(),!0!==this.stabilized){if(!0===this.adaptiveTimestep&&!0===this.adaptiveTimestepEnabled)this.adaptiveCounter%this.adaptiveInterval==0?(this.timestep=2*this.timestep,this.physicsStep(),this.revert(),this.timestep=.5*this.timestep,this.physicsStep(),this.physicsStep(),this.adjustTimeStep()):this.physicsStep(),this.adaptiveCounter+=1;else this.timestep=this.options.timestep,this.physicsStep();!0===this.stabilized&&this.revert(),this.stabilizationIterations++}}},{key:"updatePhysicsData",value:function(){this.physicsBody.forces={},this.physicsBody.physicsNodeIndices=[],this.physicsBody.physicsEdgeIndices=[];var t=this.body.nodes,e=this.body.edges;for(var i in t)Object.prototype.hasOwnProperty.call(t,i)&&!0===t[i].options.physics&&this.physicsBody.physicsNodeIndices.push(t[i].id);for(var n in e)Object.prototype.hasOwnProperty.call(e,n)&&!0===e[n].options.physics&&this.physicsBody.physicsEdgeIndices.push(e[n].id);for(var o=0;o<this.physicsBody.physicsNodeIndices.length;o++){var r=this.physicsBody.physicsNodeIndices[o];this.physicsBody.forces[r]={x:0,y:0},void 0===this.physicsBody.velocities[r]&&(this.physicsBody.velocities[r]={x:0,y:0})}for(var s in this.physicsBody.velocities)void 0===t[s]&&delete this.physicsBody.velocities[s]}},{key:"revert",value:function(){var t=zo(this.previousStates),e=this.body.nodes,i=this.physicsBody.velocities;this.referenceState={};for(var n=0;n<t.length;n++){var o=t[n];void 0!==e[o]?!0===e[o].options.physics&&(this.referenceState[o]={positions:{x:e[o].x,y:e[o].y}},i[o].x=this.previousStates[o].vx,i[o].y=this.previousStates[o].vy,e[o].x=this.previousStates[o].x,e[o].y=this.previousStates[o].y):delete this.previousStates[o]}}},{key:"_evaluateStepQuality",value:function(){var t,e,i=this.body.nodes,n=this.referenceState;for(var o in this.referenceState)if(Object.prototype.hasOwnProperty.call(this.referenceState,o)&&void 0!==i[o]&&(t=i[o].x-n[o].positions.x,e=i[o].y-n[o].positions.y,Math.sqrt(Math.pow(t,2)+Math.pow(e,2))>.3))return!1;return!0}},{key:"moveNodes",value:function(){for(var t=this.physicsBody.physicsNodeIndices,e=0,i=0,n=0;n<t.length;n++){var o=t[n],r=this._performStep(o);e=Math.max(e,r),i+=r}this.adaptiveTimestepEnabled=i/t.length<5,this.stabilized=e<this.options.minVelocity}},{key:"calculateComponentVelocity",value:function(t,e,i){t+=(e-this.modelOptions.damping*t)/i*this.timestep;var n=this.options.maxVelocity||1e9;return Math.abs(t)>n&&(t=t>0?n:-n),t}},{key:"_performStep",value:function(t){var e=this.body.nodes[t],i=this.physicsBody.forces[t];this.options.wind&&(i.x+=this.options.wind.x,i.y+=this.options.wind.y);var n=this.physicsBody.velocities[t];return this.previousStates[t]={x:e.x,y:e.y,vx:n.x,vy:n.y},!1===e.options.fixed.x?(n.x=this.calculateComponentVelocity(n.x,i.x,e.options.mass),e.x+=n.x*this.timestep):(i.x=0,n.x=0),!1===e.options.fixed.y?(n.y=this.calculateComponentVelocity(n.y,i.y,e.options.mass),e.y+=n.y*this.timestep):(i.y=0,n.y=0),Math.sqrt(Math.pow(n.x,2)+Math.pow(n.y,2))}},{key:"_freezeNodes",value:function(){var t=this.body.nodes;for(var e in t)if(Object.prototype.hasOwnProperty.call(t,e)&&t[e].x&&t[e].y){var i=t[e].options.fixed;this.freezeCache[e]={x:i.x,y:i.y},i.x=!0,i.y=!0}}},{key:"_restoreFrozenNodes",value:function(){var t=this.body.nodes;for(var e in t)Object.prototype.hasOwnProperty.call(t,e)&&void 0!==this.freezeCache[e]&&(t[e].options.fixed.x=this.freezeCache[e].x,t[e].options.fixed.y=this.freezeCache[e].y);this.freezeCache={}}},{key:"stabilize",value:function(){var t=this,e=arguments.length>0&&void 0!==arguments[0]?arguments[0]:this.options.stabilization.iterations;"number"!=typeof e&&(e=this.options.stabilization.iterations,console.error("The stabilize method needs a numeric amount of iterations. Switching to default: ",e)),0!==this.physicsBody.physicsNodeIndices.length?(this.adaptiveTimestep=this.options.adaptiveTimestep,this.body.emitter.emit("_resizeNodes"),this.stopSimulation(),this.stabilized=!1,this.body.emitter.emit("_blockRedraw"),this.targetIterations=e,!0===this.options.stabilization.onlyDynamicEdges&&this._freezeNodes(),this.stabilizationIterations=0,rs((function(){return t._stabilizationBatch()}),0)):this.ready=!0}},{key:"_startStabilizing",value:function(){return!0!==this.startedStabilization&&(this.body.emitter.emit("startStabilizing"),this.startedStabilization=!0,!0)}},{key:"_stabilizationBatch",value:function(){var t=this,e=function(){return!1===t.stabilized&&t.stabilizationIterations<t.targetIterations},i=function(){t.body.emitter.emit("stabilizationProgress",{iterations:t.stabilizationIterations,total:t.targetIterations})};this._startStabilizing()&&i();for(var n,o=0;e()&&o<this.options.stabilization.updateInterval;)this.physicsTick(),o++;(i(),e())?rs(Vt(n=this._stabilizationBatch).call(n,this),0):this._finalizeStabilization()}},{key:"_finalizeStabilization",value:function(){this.body.emitter.emit("_allowRedraw"),!0===this.options.stabilization.fit&&this.body.emitter.emit("fit"),!0===this.options.stabilization.onlyDynamicEdges&&this._restoreFrozenNodes(),this.body.emitter.emit("stabilizationIterationsDone"),this.body.emitter.emit("_requestRedraw"),!0===this.stabilized?this._emitStabilized():this.startSimulation(),this.ready=!0}},{key:"_drawForces",value:function(t){for(var e=0;e<this.physicsBody.physicsNodeIndices.length;e++){var i=this.physicsBody.physicsNodeIndices[e],n=this.body.nodes[i],o=this.physicsBody.forces[i],r=Math.sqrt(Math.pow(o.x,2)+Math.pow(o.x,2)),s=Math.min(Math.max(5,r),15),a=3*s,h=Vh((180-180*Math.min(1,Math.max(0,.03*r)))/360,1,1),l={x:n.x+20*o.x,y:n.y+20*o.y};t.lineWidth=s,t.strokeStyle=h,t.beginPath(),t.moveTo(n.x,n.y),t.lineTo(l.x,l.y),t.stroke();var d=Math.atan2(o.y,o.x);t.fillStyle=h,jf.draw(t,{type:"arrow",point:l,angle:d,length:a}),hs(t).call(t)}}}]),t}(),pp=function(){function t(){Nn(this,t)}return Fn(t,null,[{key:"getRange",value:function(t){var e,i=arguments.length>1&&void 0!==arguments[1]?arguments[1]:[],n=1e9,o=-1e9,r=1e9,s=-1e9;if(i.length>0)for(var a=0;a<i.length;a++)r>(e=t[i[a]]).shape.boundingBox.left&&(r=e.shape.boundingBox.left),s<e.shape.boundingBox.right&&(s=e.shape.boundingBox.right),n>e.shape.boundingBox.top&&(n=e.shape.boundingBox.top),o<e.shape.boundingBox.bottom&&(o=e.shape.boundingBox.bottom);return 1e9===r&&-1e9===s&&1e9===n&&-1e9===o&&(n=0,o=0,r=0,s=0),{minX:r,maxX:s,minY:n,maxY:o}}},{key:"getRangeCore",value:function(t){var e,i=arguments.length>1&&void 0!==arguments[1]?arguments[1]:[],n=1e9,o=-1e9,r=1e9,s=-1e9;if(i.length>0)for(var a=0;a<i.length;a++)r>(e=t[i[a]]).x&&(r=e.x),s<e.x&&(s=e.x),n>e.y&&(n=e.y),o<e.y&&(o=e.y);return 1e9===r&&-1e9===s&&1e9===n&&-1e9===o&&(n=0,o=0,r=0,s=0),{minX:r,maxX:s,minY:n,maxY:o}}},{key:"findCenter",value:function(t){return{x:.5*(t.maxX+t.minX),y:.5*(t.maxY+t.minY)}}},{key:"cloneOptions",value:function(t,e){var i={};return void 0===e||"node"===e?(Ch(i,t.options,!0),i.x=t.x,i.y=t.y,i.amountOfConnections=t.edges.length):Ch(i,t.options,!0),i}}]),t}();function vp(t){var e=function(){if("undefined"==typeof Reflect||!Pd)return!1;if(Pd.sham)return!1;if("function"==typeof Proxy)return!0;try{return Boolean.prototype.valueOf.call(Pd(Boolean,[],(function(){}))),!0}catch(t){return!1}}();return function(){var i,n=Ld(t);if(e){var o=Ld(this).constructor;i=Pd(n,arguments,o)}else i=n.apply(this,arguments);return Fd(this,i)}}var gp=function(t){Ad(i,t);var e=vp(i);function i(t,n,o,r,s,a){var h;return Nn(this,i),(h=e.call(this,t,n,o,r,s,a)).isCluster=!0,h.containedNodes={},h.containedEdges={},h}return Fn(i,[{key:"_openChildCluster",value:function(t){var e=this,i=this.body.nodes[t];if(void 0===this.containedNodes[t])throw new Error("node with id: "+t+" not in current cluster");if(!i.isCluster)throw new Error("node with id: "+t+" is not a cluster");delete this.containedNodes[t],Dh(i.edges,(function(t){delete e.containedEdges[t.id]})),Dh(i.containedNodes,(function(t,i){e.containedNodes[i]=t})),i.containedNodes={},Dh(i.containedEdges,(function(t,i){e.containedEdges[i]=t})),i.containedEdges={},Dh(i.edges,(function(t){Dh(e.edges,(function(i){var n,o,r=Hr(n=i.clusteringEdgeReplacingIds).call(n,t.id);-1!==r&&(Dh(t.clusteringEdgeReplacingIds,(function(t){i.clusteringEdgeReplacingIds.push(t),e.body.edges[t].edgeReplacedById=i.id})),er(o=i.clusteringEdgeReplacingIds).call(o,r,1))}))})),i.edges=[]}}]),i}(cf),yp=function(){function t(e){var i=this;Nn(this,t),this.body=e,this.clusteredNodes={},this.clusteredEdges={},this.options={},this.defaultOptions={},At(this.options,this.defaultOptions),this.body.emitter.on("_resetData",(function(){i.clusteredNodes={},i.clusteredEdges={}}))}return Fn(t,[{key:"clusterByHubsize",value:function(t,e){void 0===t?t=this._getHubSize():"object"===go(t)&&(e=this._checkOptions(t),t=this._getHubSize());for(var i=[],n=0;n<this.body.nodeIndices.length;n++){var o=this.body.nodes[this.body.nodeIndices[n]];o.edges.length>=t&&i.push(o.id)}for(var r=0;r<i.length;r++)this.clusterByConnection(i[r],e,!0);this.body.emitter.emit("_dataChanged")}},{key:"cluster",value:function(){var t=this,e=arguments.length>0&&void 0!==arguments[0]?arguments[0]:{},i=!(arguments.length>1&&void 0!==arguments[1])||arguments[1];if(void 0===e.joinCondition)throw new Error("Cannot call clusterByNodeData without a joinCondition function in the options.");e=this._checkOptions(e);var n={},o={};Dh(this.body.nodes,(function(i,r){i.options&&!0===e.joinCondition(i.options)&&(n[r]=i,Dh(i.edges,(function(e){void 0===t.clusteredEdges[e.id]&&(o[e.id]=e)})))})),this._cluster(n,o,e,i)}},{key:"clusterByEdgeCount",value:function(t,e){var i=this,n=!(arguments.length>2&&void 0!==arguments[2])||arguments[2];e=this._checkOptions(e);for(var o,r,s,a=[],h={},l=function(n){var l={},d={},c=i.body.nodeIndices[n],u=i.body.nodes[c];if(void 0===h[c]){s=0,r=[];for(var f=0;f<u.edges.length;f++)o=u.edges[f],void 0===i.clusteredEdges[o.id]&&(o.toId!==o.fromId&&s++,r.push(o));if(s===t){for(var p=function(t){if(void 0===e.joinCondition||null===e.joinCondition)return!0;var i=pp.cloneOptions(t);return e.joinCondition(i)},v=!0,g=0;g<r.length;g++){o=r[g];var y=i._getConnectedId(o,c);if(!p(u)){v=!1;break}d[o.id]=o,l[c]=u,l[y]=i.body.nodes[y],h[c]=!0}if(zo(l).length>0&&zo(d).length>0&&!0===v){var m=function(){for(var t=0;t<a.length;++t)for(var e in l)if(void 0!==a[t].nodes[e])return a[t]}();if(void 0!==m){for(var b in l)void 0===m.nodes[b]&&(m.nodes[b]=l[b]);for(var w in d)void 0===m.edges[w]&&(m.edges[w]=d[w])}else a.push({nodes:l,edges:d})}}}},d=0;d<this.body.nodeIndices.length;d++)l(d);for(var c=0;c<a.length;c++)this._cluster(a[c].nodes,a[c].edges,e,!1);!0===n&&this.body.emitter.emit("_dataChanged")}},{key:"clusterOutliers",value:function(t){var e=!(arguments.length>1&&void 0!==arguments[1])||arguments[1];this.clusterByEdgeCount(1,t,e)}},{key:"clusterBridges",value:function(t){var e=!(arguments.length>1&&void 0!==arguments[1])||arguments[1];this.clusterByEdgeCount(2,t,e)}},{key:"clusterByConnection",value:function(t,e){var i,n=!(arguments.length>2&&void 0!==arguments[2])||arguments[2];if(void 0===t)throw new Error("No nodeId supplied to clusterByConnection!");if(void 0===this.body.nodes[t])throw new Error("The nodeId given to clusterByConnection does not exist!");var o=this.body.nodes[t];void 0===(e=this._checkOptions(e,o)).clusterNodeProperties.x&&(e.clusterNodeProperties.x=o.x),void 0===e.clusterNodeProperties.y&&(e.clusterNodeProperties.y=o.y),void 0===e.clusterNodeProperties.fixed&&(e.clusterNodeProperties.fixed={},e.clusterNodeProperties.fixed.x=o.options.fixed.x,e.clusterNodeProperties.fixed.y=o.options.fixed.y);var r={},s={},a=o.id,h=pp.cloneOptions(o);r[a]=o;for(var l=0;l<o.edges.length;l++){var d=o.edges[l];if(void 0===this.clusteredEdges[d.id]){var c=this._getConnectedId(d,a);if(void 0===this.clusteredNodes[c])if(c!==a)if(void 0===e.joinCondition)s[d.id]=d,r[c]=this.body.nodes[c];else{var u=pp.cloneOptions(this.body.nodes[c]);!0===e.joinCondition(h,u)&&(s[d.id]=d,r[c]=this.body.nodes[c])}else s[d.id]=d}}var f=Io(i=zo(r)).call(i,(function(t){return r[t].id}));for(var p in r)if(Object.prototype.hasOwnProperty.call(r,p))for(var v=r[p],g=0;g<v.edges.length;g++){var y=v.edges[g];Hr(f).call(f,this._getConnectedId(y,v.id))>-1&&(s[y.id]=y)}this._cluster(r,s,e,n)}},{key:"_createClusterEdges",value:function(t,e,i,n){for(var o,r,s,a,h,l,d=zo(t),c=[],u=0;u<d.length;u++){s=t[r=d[u]];for(var f=0;f<s.edges.length;f++)o=s.edges[f],void 0===this.clusteredEdges[o.id]&&(o.toId==o.fromId?e[o.id]=o:o.toId==r?(a=i.id,l=h=o.fromId):(a=o.toId,h=i.id,l=a),void 0===t[l]&&c.push({edge:o,fromId:h,toId:a}))}for(var p=[],v=function(t){for(var e=0;e<p.length;e++){var i=p[e],n=t.fromId===i.fromId&&t.toId===i.toId,o=t.fromId===i.toId&&t.toId===i.fromId;if(n||o)return i}return null},g=0;g<c.length;g++){var y=c[g],m=y.edge,b=v(y);null===b?(b=this._createClusteredEdge(y.fromId,y.toId,m,n),p.push(b)):b.clusteringEdgeReplacingIds.push(m.id),this.body.edges[m.id].edgeReplacedById=b.id,this._backupEdgeOptions(m),m.setOptions({physics:!1})}}},{key:"_checkOptions",value:function(){var t=arguments.length>0&&void 0!==arguments[0]?arguments[0]:{};return void 0===t.clusterEdgeProperties&&(t.clusterEdgeProperties={}),void 0===t.clusterNodeProperties&&(t.clusterNodeProperties={}),t}},{key:"_cluster",value:function(t,e,i){var n=!(arguments.length>3&&void 0!==arguments[3])||arguments[3],o=[];for(var r in t)Object.prototype.hasOwnProperty.call(t,r)&&void 0!==this.clusteredNodes[r]&&o.push(r);for(var s=0;s<o.length;++s)delete t[o[s]];if(0!=zo(t).length&&(1!=zo(t).length||1==i.clusterNodeProperties.allowSingleNodeCluster)){var a=Ch({},i.clusterNodeProperties);if(void 0!==i.processProperties){var h=[];for(var l in t)if(Object.prototype.hasOwnProperty.call(t,l)){var d=pp.cloneOptions(t[l]);h.push(d)}var c=[];for(var u in e)if(Object.prototype.hasOwnProperty.call(e,u)&&"clusterEdge:"!==u.substr(0,12)){var f=pp.cloneOptions(e[u],"edge");c.push(f)}if(!(a=i.processProperties(a,h,c)))throw new Error("The processProperties function does not return properties!")}void 0===a.id&&(a.id="cluster:"+Wc());var p=a.id;void 0===a.label&&(a.label="cluster");var v=void 0;void 0===a.x&&(v=this._getClusterPosition(t),a.x=v.x),void 0===a.y&&(void 0===v&&(v=this._getClusterPosition(t)),a.y=v.y),a.id=p;var g=this.body.functions.createNode(a,gp);g.containedNodes=t,g.containedEdges=e,g.clusterEdgeProperties=i.clusterEdgeProperties,this.body.nodes[a.id]=g,this._clusterEdges(t,e,a,i.clusterEdgeProperties),a.id=void 0,!0===n&&this.body.emitter.emit("_dataChanged")}}},{key:"_backupEdgeOptions",value:function(t){void 0===this.clusteredEdges[t.id]&&(this.clusteredEdges[t.id]={physics:t.options.physics})}},{key:"_restoreEdge",value:function(t){var e=this.clusteredEdges[t.id];void 0!==e&&(t.setOptions({physics:e.physics}),delete this.clusteredEdges[t.id])}},{key:"isCluster",value:function(t){return void 0!==this.body.nodes[t]?!0===this.body.nodes[t].isCluster:(console.error("Node does not exist."),!1)}},{key:"_getClusterPosition",value:function(t){for(var e,i=zo(t),n=t[i[0]].x,o=t[i[0]].x,r=t[i[0]].y,s=t[i[0]].y,a=1;a<i.length;a++)n=(e=t[i[a]]).x<n?e.x:n,o=e.x>o?e.x:o,r=e.y<r?e.y:r,s=e.y>s?e.y:s;return{x:.5*(n+o),y:.5*(r+s)}}},{key:"openCluster",value:function(t,e){var i=!(arguments.length>2&&void 0!==arguments[2])||arguments[2];if(void 0===t)throw new Error("No clusterNodeId supplied to openCluster.");var n=this.body.nodes[t];if(void 0===n)throw new Error("The clusterNodeId supplied to openCluster does not exist.");if(!0!==n.isCluster||void 0===n.containedNodes||void 0===n.containedEdges)throw new Error("The node:"+t+" is not a valid cluster.");var o=this.findNode(t),r=Hr(o).call(o,t)-1;if(r>=0){var s=o[r],a=this.body.nodes[s];return a._openChildCluster(t),delete this.body.nodes[t],void(!0===i&&this.body.emitter.emit("_dataChanged"))}var h=n.containedNodes,l=n.containedEdges;if(void 0!==e&&void 0!==e.releaseFunction&&"function"==typeof e.releaseFunction){var d={},c={x:n.x,y:n.y};for(var u in h)if(Object.prototype.hasOwnProperty.call(h,u)){var f=this.body.nodes[u];d[u]={x:f.x,y:f.y}}var p=e.releaseFunction(c,d);for(var v in h)if(Object.prototype.hasOwnProperty.call(h,v)){var g=this.body.nodes[v];void 0!==p[v]&&(g.x=void 0===p[v].x?n.x:p[v].x,g.y=void 0===p[v].y?n.y:p[v].y)}}else Dh(h,(function(t){!1===t.options.fixed.x&&(t.x=n.x),!1===t.options.fixed.y&&(t.y=n.y)}));for(var y in h)if(Object.prototype.hasOwnProperty.call(h,y)){var m=this.body.nodes[y];m.vx=n.vx,m.vy=n.vy,m.setOptions({physics:!0}),delete this.clusteredNodes[y]}for(var b=[],w=0;w<n.edges.length;w++)b.push(n.edges[w]);for(var k=0;k<b.length;k++){for(var _=b[k],x=this._getConnectedId(_,t),E=this.clusteredNodes[x],O=0;O<_.clusteringEdgeReplacingIds.length;O++){var C=_.clusteringEdgeReplacingIds[O],S=this.body.edges[C];if(void 0!==S)if(void 0!==E){var T=this.body.nodes[E.clusterId];T.containedEdges[S.id]=S,delete l[S.id];var M=S.fromId,P=S.toId;S.toId==x?P=E.clusterId:M=E.clusterId,this._createClusteredEdge(M,P,S,T.clusterEdgeProperties,{hidden:!1,physics:!0})}else this._restoreEdge(S)}_.remove()}for(var D in l)Object.prototype.hasOwnProperty.call(l,D)&&this._restoreEdge(l[D]);delete this.body.nodes[t],!0===i&&this.body.emitter.emit("_dataChanged")}},{key:"getNodesInCluster",value:function(t){var e=[];if(!0===this.isCluster(t)){var i=this.body.nodes[t].containedNodes;for(var n in i)Object.prototype.hasOwnProperty.call(i,n)&&e.push(this.body.nodes[n].id)}return e}},{key:"findNode",value:function(t){for(var e,i=[],n=0;void 0!==this.clusteredNodes[t]&&n<100;){if(void 0===(e=this.body.nodes[t]))return[];i.push(e.id),t=this.clusteredNodes[t].clusterId,n++}return void 0===(e=this.body.nodes[t])?[]:(i.push(e.id),Xo(i).call(i),i)}},{key:"updateClusteredNode",value:function(t,e){if(void 0===t)throw new Error("No clusteredNodeId supplied to updateClusteredNode.");if(void 0===e)throw new Error("No newOptions supplied to updateClusteredNode.");if(void 0===this.body.nodes[t])throw new Error("The clusteredNodeId supplied to updateClusteredNode does not exist.");this.body.nodes[t].setOptions(e),this.body.emitter.emit("_dataChanged")}},{key:"updateEdge",value:function(t,e){if(void 0===t)throw new Error("No startEdgeId supplied to updateEdge.");if(void 0===e)throw new Error("No newOptions supplied to updateEdge.");if(void 0===this.body.edges[t])throw new Error("The startEdgeId supplied to updateEdge does not exist.");for(var i=this.getClusteredEdges(t),n=0;n<i.length;n++){this.body.edges[i[n]].setOptions(e)}this.body.emitter.emit("_dataChanged")}},{key:"getClusteredEdges",value:function(t){for(var e=[],i=0;void 0!==t&&void 0!==this.body.edges[t]&&i<100;)e.push(this.body.edges[t].id),t=this.body.edges[t].edgeReplacedById,i++;return Xo(e).call(e),e}},{key:"getBaseEdge",value:function(t){return this.getBaseEdges(t)[0]}},{key:"getBaseEdges",value:function(t){for(var e=[t],i=[],n=[],o=0;e.length>0&&o<100;){var r=e.pop();if(void 0!==r){var s=this.body.edges[r];if(void 0!==s){o++;var a=s.clusteringEdgeReplacingIds;if(void 0===a)n.push(r);else for(var h=0;h<a.length;++h){var l=a[h];-1===Hr(e).call(e,a)&&-1===Hr(i).call(i,a)&&e.push(l)}i.push(r)}}}return n}},{key:"_getConnectedId",value:function(t,e){return t.toId!=e?t.toId:(t.fromId,t.fromId)}},{key:"_getHubSize",value:function(){for(var t=0,e=0,i=0,n=0,o=0;o<this.body.nodeIndices.length;o++){var r=this.body.nodes[this.body.nodeIndices[o]];r.edges.length>n&&(n=r.edges.length),t+=r.edges.length,e+=Math.pow(r.edges.length,2),i+=1}t/=i;var s=(e/=i)-Math.pow(t,2),a=Math.sqrt(s),h=Math.floor(t+2*a);return h>n&&(h=n),h}},{key:"_createClusteredEdge",value:function(t,e,i,n,o){var r=pp.cloneOptions(i,"edge");Ch(r,n),r.from=t,r.to=e,r.id="clusterEdge:"+Wc(),void 0!==o&&Ch(r,o);var s=this.body.functions.createEdge(r);return s.clusteringEdgeReplacingIds=[i.id],s.connect(),this.body.edges[s.id]=s,s}},{key:"_clusterEdges",value:function(t,e,i,n){if(e instanceof ep){var o=e,r={};r[o.id]=o,e=r}if(t instanceof cf){var s=t,a={};a[s.id]=s,t=a}if(null==i)throw new Error("_clusterEdges: parameter clusterNode required");for(var h in void 0===n&&(n=i.clusterEdgeProperties),this._createClusterEdges(t,e,i,n),e)if(Object.prototype.hasOwnProperty.call(e,h)&&void 0!==this.body.edges[h]){var l=this.body.edges[h];this._backupEdgeOptions(l),l.setOptions({physics:!1})}for(var d in t)Object.prototype.hasOwnProperty.call(t,d)&&(this.clusteredNodes[d]={clusterId:i.id,node:this.body.nodes[d]},this.body.nodes[d].setOptions({physics:!1}))}},{key:"_getClusterNodeForNode",value:function(t){if(void 0!==t){var e=this.clusteredNodes[t];if(void 0!==e){var i=e.clusterId;if(void 0!==i)return this.body.nodes[i]}}}},{key:"_filter",value:function(t,e){var i=[];return Dh(t,(function(t){e(t)&&i.push(t)})),i}},{key:"_updateState",value:function(){var t,e=this,i=[],n={},o=function(t){Dh(e.body.nodes,(function(e){!0===e.isCluster&&t(e)}))};for(t in this.clusteredNodes){if(Object.prototype.hasOwnProperty.call(this.clusteredNodes,t))void 0===this.body.nodes[t]&&i.push(t)}o((function(t){for(var e=0;e<i.length;e++)delete t.containedNodes[i[e]]}));for(var r=0;r<i.length;r++)delete this.clusteredNodes[i[r]];Dh(this.clusteredEdges,(function(t){var i=e.body.edges[t];void 0!==i&&i.endPointsValid()||(n[t]=t)})),o((function(t){Dh(t.containedEdges,(function(t,e){t.endPointsValid()||n[e]||(n[e]=e)}))})),Dh(this.body.edges,(function(t,i){var o=!0,r=t.clusteringEdgeReplacingIds;if(void 0!==r){var s=0;Dh(r,(function(t){var i=e.body.edges[t];void 0!==i&&i.endPointsValid()&&(s+=1)})),o=s>0}t.endPointsValid()&&o||(n[i]=i)})),o((function(t){Dh(n,(function(i){delete t.containedEdges[i],Dh(t.edges,(function(o,r){o.id!==i?o.clusteringEdgeReplacingIds=e._filter(o.clusteringEdgeReplacingIds,(function(t){return!n[t]})):t.edges[r]=null})),t.edges=e._filter(t.edges,(function(t){return null!==t}))}))})),Dh(n,(function(t){delete e.clusteredEdges[t]})),Dh(n,(function(t){delete e.body.edges[t]})),Dh(zo(this.body.edges),(function(t){var i=e.body.edges[t],n=e._isClusteredNode(i.fromId)||e._isClusteredNode(i.toId);if(n!==e._isClusteredEdge(i.id))if(n){var o=e._getClusterNodeForNode(i.fromId);void 0!==o&&e._clusterEdges(e.body.nodes[i.fromId],i,o);var r=e._getClusterNodeForNode(i.toId);void 0!==r&&e._clusterEdges(e.body.nodes[i.toId],i,r)}else delete e._clusterEdges[t],e._restoreEdge(i)}));for(var s=!1,a=!0,h=function(){var t=[];o((function(e){var i=zo(e.containedNodes).length,n=!0===e.options.allowSingleNodeCluster;(n&&i<1||!n&&i<2)&&t.push(e.id)}));for(var i=0;i<t.length;++i)e.openCluster(t[i],{},!1);a=t.length>0,s=s||a};a;)h();s&&this._updateState()}},{key:"_isClusteredNode",value:function(t){return void 0!==this.clusteredNodes[t]}},{key:"_isClusteredEdge",value:function(t){return void 0!==this.clusteredEdges[t]}}]),t}();function mp(t,e){var i=void 0!==ko&&Bi(t)||t["@@iterator"];if(!i){if(So(t)||(i=function(t,e){var i;if(!t)return;if("string"==typeof t)return bp(t,e);var n=Oo(i=Object.prototype.toString.call(t)).call(i,8,-1);"Object"===n&&t.constructor&&(n=t.constructor.name);if("Map"===n||"Set"===n)return Ei(t);if("Arguments"===n||/^(?:Ui|I)nt(?:8|16|32)(?:Clamped)?Array$/.test(n))return bp(t,e)}(t))||e&&t&&"number"==typeof t.length){i&&(t=i);var n=0,o=function(){};return{s:o,n:function(){return n>=t.length?{done:!0}:{done:!1,value:t[n++]}},e:function(t){throw t},f:o}}throw new TypeError("Invalid attempt to iterate non-iterable instance.\nIn order to be iterable, non-array objects must have a [Symbol.iterator]() method.")}var r,s=!0,a=!1;return{s:function(){i=i.call(t)},n:function(){var t=i.next();return s=t.done,t},e:function(t){a=!0,r=t},f:function(){try{s||null==i.return||i.return()}finally{if(a)throw r}}}}function bp(t,e){(null==e||e>t.length)&&(e=t.length);for(var i=0,n=new Array(e);i<e;i++)n[i]=t[i];return n}var wp=function(){function t(e,i){var n;Nn(this,t),void 0!==window&&(n=window.requestAnimationFrame||window.mozRequestAnimationFrame||window.webkitRequestAnimationFrame||window.msRequestAnimationFrame),window.requestAnimationFrame=void 0===n?function(t){t()}:n,this.body=e,this.canvas=i,this.redrawRequested=!1,this.renderTimer=void 0,this.requiresTimeout=!0,this.renderingActive=!1,this.renderRequests=0,this.allowRedraw=!0,this.dragging=!1,this.zooming=!1,this.options={},this.defaultOptions={hideEdgesOnDrag:!1,hideEdgesOnZoom:!1,hideNodesOnDrag:!1},At(this.options,this.defaultOptions),this._determineBrowserMethod(),this.bindEventListeners()}return Fn(t,[{key:"bindEventListeners",value:function(){var t,e=this;this.body.emitter.on("dragStart",(function(){e.dragging=!0})),this.body.emitter.on("dragEnd",(function(){e.dragging=!1})),this.body.emitter.on("zoom",(function(){e.zooming=!0,window.clearTimeout(e.zoomTimeoutId),e.zoomTimeoutId=rs((function(){var t;e.zooming=!1,Vt(t=e._requestRedraw).call(t,e)()}),250)})),this.body.emitter.on("_resizeNodes",(function(){e._resizeNodes()})),this.body.emitter.on("_redraw",(function(){!1===e.renderingActive&&e._redraw()})),this.body.emitter.on("_blockRedraw",(function(){e.allowRedraw=!1})),this.body.emitter.on("_allowRedraw",(function(){e.allowRedraw=!0,e.redrawRequested=!1})),this.body.emitter.on("_requestRedraw",Vt(t=this._requestRedraw).call(t,this)),this.body.emitter.on("_startRendering",(function(){e.renderRequests+=1,e.renderingActive=!0,e._startRendering()})),this.body.emitter.on("_stopRendering",(function(){e.renderRequests-=1,e.renderingActive=e.renderRequests>0,e.renderTimer=void 0})),this.body.emitter.on("destroy",(function(){e.renderRequests=0,e.allowRedraw=!1,e.renderingActive=!1,!0===e.requiresTimeout?clearTimeout(e.renderTimer):window.cancelAnimationFrame(e.renderTimer),e.body.emitter.off()}))}},{key:"setOptions",value:function(t){if(void 0!==t){Eh(["hideEdgesOnDrag","hideEdgesOnZoom","hideNodesOnDrag"],this.options,t)}}},{key:"_requestNextFrame",value:function(t,e){if("undefined"!=typeof window){var i,n=window;return!0===this.requiresTimeout?i=rs(t,e):n.requestAnimationFrame&&(i=n.requestAnimationFrame(t)),i}}},{key:"_startRendering",value:function(){var t;!0===this.renderingActive&&(void 0===this.renderTimer&&(this.renderTimer=this._requestNextFrame(Vt(t=this._renderStep).call(t,this),this.simulationInterval)))}},{key:"_renderStep",value:function(){!0===this.renderingActive&&(this.renderTimer=void 0,!0===this.requiresTimeout&&this._startRendering(),this._redraw(),!1===this.requiresTimeout&&this._startRendering())}},{key:"redraw",value:function(){this.body.emitter.emit("setSize"),this._redraw()}},{key:"_requestRedraw",value:function(){var t=this;!0!==this.redrawRequested&&!1===this.renderingActive&&!0===this.allowRedraw&&(this.redrawRequested=!0,this._requestNextFrame((function(){t._redraw(!1)}),0))}},{key:"_redraw",value:function(){var t=arguments.length>0&&void 0!==arguments[0]&&arguments[0];if(!0===this.allowRedraw){this.body.emitter.emit("initRedraw"),this.redrawRequested=!1;var e={drawExternalLabels:null};0!==this.canvas.frame.canvas.width&&0!==this.canvas.frame.canvas.height||this.canvas.setSize(),this.canvas.setTransform();var i=this.canvas.getContext(),n=this.canvas.frame.canvas.clientWidth,o=this.canvas.frame.canvas.clientHeight;if(i.clearRect(0,0,n,o),0===this.canvas.frame.clientWidth)return;if(i.save(),i.translate(this.body.view.translation.x,this.body.view.translation.y),i.scale(this.body.view.scale,this.body.view.scale),i.beginPath(),this.body.emitter.emit("beforeDrawing",i),i.closePath(),!1===t&&(!1===this.dragging||!0===this.dragging&&!1===this.options.hideEdgesOnDrag)&&(!1===this.zooming||!0===this.zooming&&!1===this.options.hideEdgesOnZoom)&&this._drawEdges(i),!1===this.dragging||!0===this.dragging&&!1===this.options.hideNodesOnDrag){var r=this._drawNodes(i,t),s=r.drawExternalLabels;e.drawExternalLabels=s}!1===t&&(!1===this.dragging||!0===this.dragging&&!1===this.options.hideEdgesOnDrag)&&(!1===this.zooming||!0===this.zooming&&!1===this.options.hideEdgesOnZoom)&&this._drawArrows(i),null!=e.drawExternalLabels&&e.drawExternalLabels(),!1===t&&this._drawSelectionBox(i),i.beginPath(),this.body.emitter.emit("afterDrawing",i),i.closePath(),i.restore(),!0===t&&i.clearRect(0,0,n,o)}}},{key:"_resizeNodes",value:function(){this.canvas.setTransform();var t=this.canvas.getContext();t.save(),t.translate(this.body.view.translation.x,this.body.view.translation.y),t.scale(this.body.view.scale,this.body.view.scale);var e,i=this.body.nodes;for(var n in i)Object.prototype.hasOwnProperty.call(i,n)&&((e=i[n]).resize(t),e.updateBoundingBox(t,e.selected));t.restore()}},{key:"_drawNodes",value:function(t){for(var e,i,n=arguments.length>1&&void 0!==arguments[1]&&arguments[1],o=this.body.nodes,r=this.body.nodeIndices,s=[],a=[],h=20,l=this.canvas.DOMtoCanvas({x:-h,y:-h}),d=this.canvas.DOMtoCanvas({x:this.canvas.frame.canvas.clientWidth+h,y:this.canvas.frame.canvas.clientHeight+h}),c={top:l.y,left:l.x,bottom:d.y,right:d.x},u=[],f=0;f<r.length;f++)if((e=o[r[f]]).hover)a.push(r[f]);else if(e.isSelected())s.push(r[f]);else if(!0===n){var p=e.draw(t);null!=p.drawExternalLabel&&u.push(p.drawExternalLabel)}else if(!0===e.isBoundingBoxOverlappingWith(c)){var v=e.draw(t);null!=v.drawExternalLabel&&u.push(v.drawExternalLabel)}else e.updateBoundingBox(t,e.selected);var g=s.length,y=a.length;for(i=0;i<g;i++){var m=(e=o[s[i]]).draw(t);null!=m.drawExternalLabel&&u.push(m.drawExternalLabel)}for(i=0;i<y;i++){var b=(e=o[a[i]]).draw(t);null!=b.drawExternalLabel&&u.push(b.drawExternalLabel)}return{drawExternalLabels:function(){var t,e=mp(u);try{for(e.s();!(t=e.n()).done;){(0,t.value)()}}catch(t){e.e(t)}finally{e.f()}}}}},{key:"_drawEdges",value:function(t){for(var e=this.body.edges,i=this.body.edgeIndices,n=0;n<i.length;n++){var o=e[i[n]];!0===o.connected&&o.draw(t)}}},{key:"_drawArrows",value:function(t){for(var e=this.body.edges,i=this.body.edgeIndices,n=0;n<i.length;n++){var o=e[i[n]];!0===o.connected&&o.drawArrows(t)}}},{key:"_determineBrowserMethod",value:function(){if("undefined"!=typeof window){var t=navigator.userAgent.toLowerCase();this.requiresTimeout=!1,(-1!=Hr(t).call(t,"msie 9.0")||-1!=Hr(t).call(t,"safari")&&Hr(t).call(t,"chrome")<=-1)&&(this.requiresTimeout=!0)}else this.requiresTimeout=!0}},{key:"_drawSelectionBox",value:function(t){if(this.body.selectionBox.show){t.beginPath();var e=this.body.selectionBox.position.end.x-this.body.selectionBox.position.start.x,i=this.body.selectionBox.position.end.y-this.body.selectionBox.position.start.y;t.rect(this.body.selectionBox.position.start.x,this.body.selectionBox.position.start.y,e,i),t.fillStyle="rgba(151, 194, 252, 0.2)",t.fillRect(this.body.selectionBox.position.start.x,this.body.selectionBox.position.start.y,e,i),t.strokeStyle="rgba(151, 194, 252, 1)",t.stroke()}else t.closePath()}}]),t}(),kp=k.setInterval;function _p(t,e){e.inputHandler=function(t){t.isFirst&&e(t)},t.on("hammer.input",e.inputHandler)}function xp(t,e){return e.inputHandler=function(t){t.isFinal&&e(t)},t.on("hammer.input",e.inputHandler)}var Ep=function(){function t(e){Nn(this,t),this.body=e,this.pixelRatio=1,this.cameraState={},this.initialized=!1,this.canvasViewCenter={},this._cleanupCallbacks=[],this.options={},this.defaultOptions={autoResize:!0,height:"100%",width:"100%"},At(this.options,this.defaultOptions),this.bindEventListeners()}return Fn(t,[{key:"bindEventListeners",value:function(){var t,e=this;this.body.emitter.once("resize",(function(t){0!==t.width&&(e.body.view.translation.x=.5*t.width),0!==t.height&&(e.body.view.translation.y=.5*t.height)})),this.body.emitter.on("setSize",Vt(t=this.setSize).call(t,this)),this.body.emitter.on("destroy",(function(){e.hammerFrame.destroy(),e.hammer.destroy(),e._cleanUp()}))}},{key:"setOptions",value:function(t){var e=this;if(void 0!==t){Eh(["width","height","autoResize"],this.options,t)}if(this._cleanUp(),!0===this.options.autoResize){var i;if(window.ResizeObserver){var n=new ResizeObserver((function(){!0===e.setSize()&&e.body.emitter.emit("_requestRedraw")})),o=this.frame;n.observe(o),this._cleanupCallbacks.push((function(){n.unobserve(o)}))}else{var r=kp((function(){!0===e.setSize()&&e.body.emitter.emit("_requestRedraw")}),1e3);this._cleanupCallbacks.push((function(){clearInterval(r)}))}var s=Vt(i=this._onResize).call(i,this);Bh(window,"resize",s),this._cleanupCallbacks.push((function(){zh(window,"resize",s)}))}}},{key:"_cleanUp",value:function(){var t,e,i;Wo(t=Xo(e=er(i=this._cleanupCallbacks).call(i,0)).call(e)).call(t,(function(t){try{t()}catch(t){console.error(t)}}))}},{key:"_onResize",value:function(){this.setSize(),this.body.emitter.emit("_redraw")}},{key:"_getCameraState",value:function(){var t=arguments.length>0&&void 0!==arguments[0]?arguments[0]:this.pixelRatio;!0===this.initialized&&(this.cameraState.previousWidth=this.frame.canvas.width/t,this.cameraState.previousHeight=this.frame.canvas.height/t,this.cameraState.scale=this.body.view.scale,this.cameraState.position=this.DOMtoCanvas({x:.5*this.frame.canvas.width/t,y:.5*this.frame.canvas.height/t}))}},{key:"_setCameraState",value:function(){if(void 0!==this.cameraState.scale&&0!==this.frame.canvas.clientWidth&&0!==this.frame.canvas.clientHeight&&0!==this.pixelRatio&&this.cameraState.previousWidth>0&&this.cameraState.previousHeight>0){var t=this.frame.canvas.width/this.pixelRatio/this.cameraState.previousWidth,e=this.frame.canvas.height/this.pixelRatio/this.cameraState.previousHeight,i=this.cameraState.scale;1!=t&&1!=e?i=.5*this.cameraState.scale*(t+e):1!=t?i=this.cameraState.scale*t:1!=e&&(i=this.cameraState.scale*e),this.body.view.scale=i;var n=this.DOMtoCanvas({x:.5*this.frame.canvas.clientWidth,y:.5*this.frame.canvas.clientHeight}),o={x:n.x-this.cameraState.position.x,y:n.y-this.cameraState.position.y};this.body.view.translation.x+=o.x*this.body.view.scale,this.body.view.translation.y+=o.y*this.body.view.scale}}},{key:"_prepareValue",value:function(t){if("number"==typeof t)return t+"px";if("string"==typeof t){if(-1!==Hr(t).call(t,"%")||-1!==Hr(t).call(t,"px"))return t;if(-1===Hr(t).call(t,"%"))return t+"px"}throw new Error("Could not use the value supplied for width or height:"+t)}},{key:"_create",value:function(){for(;this.body.container.hasChildNodes();)this.body.container.removeChild(this.body.container.firstChild);if(this.frame=document.createElement("div"),this.frame.className="vis-network",this.frame.style.position="relative",this.frame.style.overflow="hidden",this.frame.tabIndex=0,this.frame.canvas=document.createElement("canvas"),this.frame.canvas.style.position="relative",this.frame.appendChild(this.frame.canvas),this.frame.canvas.getContext)this._setPixelRatio(),this.setTransform();else{var t=document.createElement("DIV");t.style.color="red",t.style.fontWeight="bold",t.style.padding="10px",t.innerText="Error: your browser does not support HTML canvas",this.frame.canvas.appendChild(t)}this.body.container.appendChild(this.frame),this.body.view.scale=1,this.body.view.translation={x:.5*this.frame.canvas.clientWidth,y:.5*this.frame.canvas.clientHeight},this._bindHammer()}},{key:"_bindHammer",value:function(){var t=this;void 0!==this.hammer&&this.hammer.destroy(),this.drag={},this.pinch={},this.hammer=new ll(this.frame.canvas),this.hammer.get("pinch").set({enable:!0}),this.hammer.get("pan").set({threshold:5,direction:ll.DIRECTION_ALL}),_p(this.hammer,(function(e){t.body.eventListeners.onTouch(e)})),this.hammer.on("tap",(function(e){t.body.eventListeners.onTap(e)})),this.hammer.on("doubletap",(function(e){t.body.eventListeners.onDoubleTap(e)})),this.hammer.on("press",(function(e){t.body.eventListeners.onHold(e)})),this.hammer.on("panstart",(function(e){t.body.eventListeners.onDragStart(e)})),this.hammer.on("panmove",(function(e){t.body.eventListeners.onDrag(e)})),this.hammer.on("panend",(function(e){t.body.eventListeners.onDragEnd(e)})),this.hammer.on("pinch",(function(e){t.body.eventListeners.onPinch(e)})),this.frame.canvas.addEventListener("wheel",(function(e){t.body.eventListeners.onMouseWheel(e)})),this.frame.canvas.addEventListener("mousemove",(function(e){t.body.eventListeners.onMouseMove(e)})),this.frame.canvas.addEventListener("contextmenu",(function(e){t.body.eventListeners.onContext(e)})),this.hammerFrame=new ll(this.frame),xp(this.hammerFrame,(function(e){t.body.eventListeners.onRelease(e)}))}},{key:"setSize",value:function(){var t=arguments.length>0&&void 0!==arguments[0]?arguments[0]:this.options.width,e=arguments.length>1&&void 0!==arguments[1]?arguments[1]:this.options.height;t=this._prepareValue(t),e=this._prepareValue(e);var i=!1,n=this.frame.canvas.width,o=this.frame.canvas.height,r=this.pixelRatio;if(this._setPixelRatio(),t!=this.options.width||e!=this.options.height||this.frame.style.width!=t||this.frame.style.height!=e)this._getCameraState(r),this.frame.style.width=t,this.frame.style.height=e,this.frame.canvas.style.width="100%",this.frame.canvas.style.height="100%",this.frame.canvas.width=Math.round(this.frame.canvas.clientWidth*this.pixelRatio),this.frame.canvas.height=Math.round(this.frame.canvas.clientHeight*this.pixelRatio),this.options.width=t,this.options.height=e,this.canvasViewCenter={x:.5*this.frame.clientWidth,y:.5*this.frame.clientHeight},i=!0;else{var s=Math.round(this.frame.canvas.clientWidth*this.pixelRatio),a=Math.round(this.frame.canvas.clientHeight*this.pixelRatio);this.frame.canvas.width===s&&this.frame.canvas.height===a||this._getCameraState(r),this.frame.canvas.width!==s&&(this.frame.canvas.width=s,i=!0),this.frame.canvas.height!==a&&(this.frame.canvas.height=a,i=!0)}return!0===i&&(this.body.emitter.emit("resize",{width:Math.round(this.frame.canvas.width/this.pixelRatio),height:Math.round(this.frame.canvas.height/this.pixelRatio),oldWidth:Math.round(n/this.pixelRatio),oldHeight:Math.round(o/this.pixelRatio)}),this._setCameraState()),this.initialized=!0,i}},{key:"getContext",value:function(){return this.frame.canvas.getContext("2d")}},{key:"_determinePixelRatio",value:function(){var t=this.getContext();if(void 0===t)throw new Error("Could not get canvax context");var e=1;return"undefined"!=typeof window&&(e=window.devicePixelRatio||1),e/(t.webkitBackingStorePixelRatio||t.mozBackingStorePixelRatio||t.msBackingStorePixelRatio||t.oBackingStorePixelRatio||t.backingStorePixelRatio||1)}},{key:"_setPixelRatio",value:function(){this.pixelRatio=this._determinePixelRatio()}},{key:"setTransform",value:function(){var t=this.getContext();if(void 0===t)throw new Error("Could not get canvax context");t.setTransform(this.pixelRatio,0,0,this.pixelRatio,0,0)}},{key:"_XconvertDOMtoCanvas",value:function(t){return(t-this.body.view.translation.x)/this.body.view.scale}},{key:"_XconvertCanvasToDOM",value:function(t){return t*this.body.view.scale+this.body.view.translation.x}},{key:"_YconvertDOMtoCanvas",value:function(t){return(t-this.body.view.translation.y)/this.body.view.scale}},{key:"_YconvertCanvasToDOM",value:function(t){return t*this.body.view.scale+this.body.view.translation.y}},{key:"canvasToDOM",value:function(t){return{x:this._XconvertCanvasToDOM(t.x),y:this._YconvertCanvasToDOM(t.y)}}},{key:"DOMtoCanvas",value:function(t){return{x:this._XconvertDOMtoCanvas(t.x),y:this._YconvertDOMtoCanvas(t.y)}}}]),t}();function Op(t,e){var i=At({nodes:e,minZoomLevel:Number.MIN_VALUE,maxZoomLevel:1},null!=t?t:{});if(!So(i.nodes))throw new TypeError("Nodes has to be an array of ids.");if(0===i.nodes.length&&(i.nodes=e),!("number"==typeof i.minZoomLevel&&i.minZoomLevel>0))throw new TypeError("Min zoom level has to be a number higher than zero.");if(!("number"==typeof i.maxZoomLevel&&i.minZoomLevel<=i.maxZoomLevel))throw new TypeError("Max zoom level has to be a number higher than min zoom level.");return i}var Cp=function(){function t(e,i){var n,o,r=this;Nn(this,t),this.body=e,this.canvas=i,this.animationSpeed=1/this.renderRefreshRate,this.animationEasingFunction="easeInOutQuint",this.easingTime=0,this.sourceScale=0,this.targetScale=0,this.sourceTranslation=0,this.targetTranslation=0,this.lockedOnNodeId=void 0,this.lockedOnNodeOffset=void 0,this.touchTime=0,this.viewFunction=void 0,this.body.emitter.on("fit",Vt(n=this.fit).call(n,this)),this.body.emitter.on("animationFinished",(function(){r.body.emitter.emit("_stopRendering")})),this.body.emitter.on("unlockNode",Vt(o=this.releaseNode).call(o,this))}return Fn(t,[{key:"setOptions",value:function(){var t=arguments.length>0&&void 0!==arguments[0]?arguments[0]:{};this.options=t}},{key:"fit",value:function(t){var e=arguments.length>1&&void 0!==arguments[1]&&arguments[1];t=Op(t,this.body.nodeIndices);var i,n,o=this.canvas.frame.canvas.clientWidth,r=this.canvas.frame.canvas.clientHeight;if(0===o||0===r)n=1,i=pp.getRange(this.body.nodes,t.nodes);else if(!0===e){var s=0;for(var a in this.body.nodes)if(Object.prototype.hasOwnProperty.call(this.body.nodes,a)){var h=this.body.nodes[a];!0===h.predefinedPosition&&(s+=1)}if(s>.5*this.body.nodeIndices.length)return void this.fit(t,!1);i=pp.getRange(this.body.nodes,t.nodes);var l=this.body.nodeIndices.length;n=12.662/(l+7.4147)+.0964822;var d=Math.min(o/600,r/600);n*=d}else{this.body.emitter.emit("_resizeNodes"),i=pp.getRange(this.body.nodes,t.nodes);var c=1.1*Math.abs(i.maxX-i.minX),u=1.1*Math.abs(i.maxY-i.minY),f=o/c,p=r/u;n=f<=p?f:p}n>t.maxZoomLevel?n=t.maxZoomLevel:n<t.minZoomLevel&&(n=t.minZoomLevel);var v=pp.findCenter(i),g={position:v,scale:n,animation:t.animation};this.moveTo(g)}},{key:"focus",value:function(t){var e=arguments.length>1&&void 0!==arguments[1]?arguments[1]:{};if(void 0!==this.body.nodes[t]){var i={x:this.body.nodes[t].x,y:this.body.nodes[t].y};e.position=i,e.lockedOnNode=t,this.moveTo(e)}else console.error("Node: "+t+" cannot be found.")}},{key:"moveTo",value:function(t){if(void 0!==t){if(null!=t.offset){if(null!=t.offset.x){if(t.offset.x=+t.offset.x,!wd(t.offset.x))throw new TypeError('The option "offset.x" has to be a finite number.')}else t.offset.x=0;if(null!=t.offset.y){if(t.offset.y=+t.offset.y,!wd(t.offset.y))throw new TypeError('The option "offset.y" has to be a finite number.')}else t.offset.x=0}else t.offset={x:0,y:0};if(null!=t.position){if(null!=t.position.x){if(t.position.x=+t.position.x,!wd(t.position.x))throw new TypeError('The option "position.x" has to be a finite number.')}else t.position.x=0;if(null!=t.position.y){if(t.position.y=+t.position.y,!wd(t.position.y))throw new TypeError('The option "position.y" has to be a finite number.')}else t.position.x=0}else t.position=this.getViewPosition();if(null!=t.scale){if(t.scale=+t.scale,!(t.scale>0))throw new TypeError('The option "scale" has to be a number greater than zero.')}else t.scale=this.body.view.scale;void 0===t.animation&&(t.animation={duration:0}),!1===t.animation&&(t.animation={duration:0}),!0===t.animation&&(t.animation={}),void 0===t.animation.duration&&(t.animation.duration=1e3),void 0===t.animation.easingFunction&&(t.animation.easingFunction="easeInOutQuad"),this.animateView(t)}else t={}}},{key:"animateView",value:function(t){if(void 0!==t){this.animationEasingFunction=t.animation.easingFunction,this.releaseNode(),!0===t.locked&&(this.lockedOnNodeId=t.lockedOnNode,this.lockedOnNodeOffset=t.offset),0!=this.easingTime&&this._transitionRedraw(!0),this.sourceScale=this.body.view.scale,this.sourceTranslation=this.body.view.translation,this.targetScale=t.scale,this.body.view.scale=this.targetScale;var e,i,n=this.canvas.DOMtoCanvas({x:.5*this.canvas.frame.canvas.clientWidth,y:.5*this.canvas.frame.canvas.clientHeight}),o=n.x-t.position.x,r=n.y-t.position.y;if(this.targetTranslation={x:this.sourceTranslation.x+o*this.targetScale+t.offset.x,y:this.sourceTranslation.y+r*this.targetScale+t.offset.y},0===t.animation.duration)if(null!=this.lockedOnNodeId)this.viewFunction=Vt(e=this._lockedRedraw).call(e,this),this.body.emitter.on("initRedraw",this.viewFunction);else this.body.view.scale=this.targetScale,this.body.view.translation=this.targetTranslation,this.body.emitter.emit("_requestRedraw");else this.animationSpeed=1/(60*t.animation.duration*.001)||1/60,this.animationEasingFunction=t.animation.easingFunction,this.viewFunction=Vt(i=this._transitionRedraw).call(i,this),this.body.emitter.on("initRedraw",this.viewFunction),this.body.emitter.emit("_startRendering")}}},{key:"_lockedRedraw",value:function(){var t=this.body.nodes[this.lockedOnNodeId].x,e=this.body.nodes[this.lockedOnNodeId].y,i=this.canvas.DOMtoCanvas({x:.5*this.canvas.frame.canvas.clientWidth,y:.5*this.canvas.frame.canvas.clientHeight}),n=i.x-t,o=i.y-e,r=this.body.view.translation,s={x:r.x+n*this.body.view.scale+this.lockedOnNodeOffset.x,y:r.y+o*this.body.view.scale+this.lockedOnNodeOffset.y};this.body.view.translation=s}},{key:"releaseNode",value:function(){void 0!==this.lockedOnNodeId&&void 0!==this.viewFunction&&(this.body.emitter.off("initRedraw",this.viewFunction),this.lockedOnNodeId=void 0,this.lockedOnNodeOffset=void 0)}},{key:"_transitionRedraw",value:function(){var t=arguments.length>0&&void 0!==arguments[0]&&arguments[0];this.easingTime+=this.animationSpeed,this.easingTime=!0===t?1:this.easingTime;var e=Zh[this.animationEasingFunction](this.easingTime);if(this.body.view.scale=this.sourceScale+(this.targetScale-this.sourceScale)*e,this.body.view.translation={x:this.sourceTranslation.x+(this.targetTranslation.x-this.sourceTranslation.x)*e,y:this.sourceTranslation.y+(this.targetTranslation.y-this.sourceTranslation.y)*e},this.easingTime>=1){var i;if(this.body.emitter.off("initRedraw",this.viewFunction),this.easingTime=0,null!=this.lockedOnNodeId)this.viewFunction=Vt(i=this._lockedRedraw).call(i,this),this.body.emitter.on("initRedraw",this.viewFunction);this.body.emitter.emit("animationFinished")}}},{key:"getScale",value:function(){return this.body.view.scale}},{key:"getViewPosition",value:function(){return this.canvas.DOMtoCanvas({x:.5*this.canvas.frame.canvas.clientWidth,y:.5*this.canvas.frame.canvas.clientHeight})}}]),t}();function Sp(t){var e,i=t&&t.preventDefault||!1,n=t&&t.container||window,o={},r={keydown:{},keyup:{}},s={};for(e=97;e<=122;e++)s[String.fromCharCode(e)]={code:e-97+65,shift:!1};for(e=65;e<=90;e++)s[String.fromCharCode(e)]={code:e,shift:!0};for(e=0;e<=9;e++)s[""+e]={code:48+e,shift:!1};for(e=1;e<=12;e++)s["F"+e]={code:111+e,shift:!1};for(e=0;e<=9;e++)s["num"+e]={code:96+e,shift:!1};s["num*"]={code:106,shift:!1},s["num+"]={code:107,shift:!1},s["num-"]={code:109,shift:!1},s["num/"]={code:111,shift:!1},s["num."]={code:110,shift:!1},s.left={code:37,shift:!1},s.up={code:38,shift:!1},s.right={code:39,shift:!1},s.down={code:40,shift:!1},s.space={code:32,shift:!1},s.enter={code:13,shift:!1},s.shift={code:16,shift:void 0},s.esc={code:27,shift:!1},s.backspace={code:8,shift:!1},s.tab={code:9,shift:!1},s.ctrl={code:17,shift:!1},s.alt={code:18,shift:!1},s.delete={code:46,shift:!1},s.pageup={code:33,shift:!1},s.pagedown={code:34,shift:!1},s["="]={code:187,shift:!1},s["-"]={code:189,shift:!1},s["]"]={code:221,shift:!1},s["["]={code:219,shift:!1};var a=function(t){l(t,"keydown")},h=function(t){l(t,"keyup")},l=function(t,e){if(void 0!==r[e][t.keyCode]){for(var n=r[e][t.keyCode],o=0;o<n.length;o++)(void 0===n[o].shift||1==n[o].shift&&1==t.shiftKey||0==n[o].shift&&0==t.shiftKey)&&n[o].fn(t);1==i&&t.preventDefault()}};return o.bind=function(t,e,i){if(void 0===i&&(i="keydown"),void 0===s[t])throw new Error("unsupported key: "+t);void 0===r[i][s[t].code]&&(r[i][s[t].code]=[]),r[i][s[t].code].push({fn:e,shift:s[t].shift})},o.bindAll=function(t,e){for(var i in void 0===e&&(e="keydown"),s)s.hasOwnProperty(i)&&o.bind(i,t,e)},o.getKey=function(t){for(var e in s)if(s.hasOwnProperty(e)){if(1==t.shiftKey&&1==s[e].shift&&t.keyCode==s[e].code)return e;if(0==t.shiftKey&&0==s[e].shift&&t.keyCode==s[e].code)return e;if(t.keyCode==s[e].code&&"shift"==e)return e}return"unknown key, currently not supported"},o.unbind=function(t,e,i){if(void 0===i&&(i="keydown"),void 0===s[t])throw new Error("unsupported key: "+t);if(void 0!==e){var n=[],o=r[i][s[t].code];if(void 0!==o)for(var a=0;a<o.length;a++)o[a].fn==e&&o[a].shift==s[t].shift||n.push(r[i][s[t].code][a]);r[i][s[t].code]=n}else r[i][s[t].code]=[]},o.reset=function(){r={keydown:{},keyup:{}}},o.destroy=function(){r={keydown:{},keyup:{}},n.removeEventListener("keydown",a,!0),n.removeEventListener("keyup",h,!0)},n.addEventListener("keydown",a,!0),n.addEventListener("keyup",h,!0),o}var Tp=Object.freeze({__proto__:null,default:Sp}),Mp=function(){function t(e,i){var n=this;Nn(this,t),this.body=e,this.canvas=i,this.iconsCreated=!1,this.navigationHammers=[],this.boundFunctions={},this.touchTime=0,this.activated=!1,this.body.emitter.on("activate",(function(){n.activated=!0,n.configureKeyboardBindings()})),this.body.emitter.on("deactivate",(function(){n.activated=!1,n.configureKeyboardBindings()})),this.body.emitter.on("destroy",(function(){void 0!==n.keycharm&&n.keycharm.destroy()})),this.options={}}return Fn(t,[{key:"setOptions",value:function(t){void 0!==t&&(this.options=t,this.create())}},{key:"create",value:function(){!0===this.options.navigationButtons?!1===this.iconsCreated&&this.loadNavigationElements():!0===this.iconsCreated&&this.cleanNavigation(),this.configureKeyboardBindings()}},{key:"cleanNavigation",value:function(){if(0!=this.navigationHammers.length){for(var t=0;t<this.navigationHammers.length;t++)this.navigationHammers[t].destroy();this.navigationHammers=[]}this.navigationDOM&&this.navigationDOM.wrapper&&this.navigationDOM.wrapper.parentNode&&this.navigationDOM.wrapper.parentNode.removeChild(this.navigationDOM.wrapper),this.iconsCreated=!1}},{key:"loadNavigationElements",value:function(){var t=this;this.cleanNavigation(),this.navigationDOM={};var e=["up","down","left","right","zoomIn","zoomOut","zoomExtends"],i=["_moveUp","_moveDown","_moveLeft","_moveRight","_zoomIn","_zoomOut","_fit"];this.navigationDOM.wrapper=document.createElement("div"),this.navigationDOM.wrapper.className="vis-navigation",this.canvas.frame.appendChild(this.navigationDOM.wrapper);for(var n=0;n<e.length;n++){this.navigationDOM[e[n]]=document.createElement("div"),this.navigationDOM[e[n]].className="vis-button vis-"+e[n],this.navigationDOM.wrapper.appendChild(this.navigationDOM[e[n]]);var o,r,s=new ll(this.navigationDOM[e[n]]);if("_fit"===i[n])_p(s,Vt(o=this._fit).call(o,this));else _p(s,Vt(r=this.bindToRedraw).call(r,this,i[n]));this.navigationHammers.push(s)}var a=new ll(this.canvas.frame);xp(a,(function(){t._stopMovement()})),this.navigationHammers.push(a),this.iconsCreated=!0}},{key:"bindToRedraw",value:function(t){var e;void 0===this.boundFunctions[t]&&(this.boundFunctions[t]=Vt(e=this[t]).call(e,this),this.body.emitter.on("initRedraw",this.boundFunctions[t]),this.body.emitter.emit("_startRendering"))}},{key:"unbindFromRedraw",value:function(t){void 0!==this.boundFunctions[t]&&(this.body.emitter.off("initRedraw",this.boundFunctions[t]),this.body.emitter.emit("_stopRendering"),delete this.boundFunctions[t])}},{key:"_fit",value:function(){(new Date).valueOf()-this.touchTime>700&&(this.body.emitter.emit("fit",{duration:700}),this.touchTime=(new Date).valueOf())}},{key:"_stopMovement",value:function(){for(var t in this.boundFunctions)Object.prototype.hasOwnProperty.call(this.boundFunctions,t)&&(this.body.emitter.off("initRedraw",this.boundFunctions[t]),this.body.emitter.emit("_stopRendering"));this.boundFunctions={}}},{key:"_moveUp",value:function(){this.body.view.translation.y+=this.options.keyboard.speed.y}},{key:"_moveDown",value:function(){this.body.view.translation.y-=this.options.keyboard.speed.y}},{key:"_moveLeft",value:function(){this.body.view.translation.x+=this.options.keyboard.speed.x}},{key:"_moveRight",value:function(){this.body.view.translation.x-=this.options.keyboard.speed.x}},{key:"_zoomIn",value:function(){var t=this.body.view.scale,e=this.body.view.scale*(1+this.options.keyboard.speed.zoom),i=this.body.view.translation,n=e/t,o=(1-n)*this.canvas.canvasViewCenter.x+i.x*n,r=(1-n)*this.canvas.canvasViewCenter.y+i.y*n;this.body.view.scale=e,this.body.view.translation={x:o,y:r},this.body.emitter.emit("zoom",{direction:"+",scale:this.body.view.scale,pointer:null})}},{key:"_zoomOut",value:function(){var t=this.body.view.scale,e=this.body.view.scale/(1+this.options.keyboard.speed.zoom),i=this.body.view.translation,n=e/t,o=(1-n)*this.canvas.canvasViewCenter.x+i.x*n,r=(1-n)*this.canvas.canvasViewCenter.y+i.y*n;this.body.view.scale=e,this.body.view.translation={x:o,y:r},this.body.emitter.emit("zoom",{direction:"-",scale:this.body.view.scale,pointer:null})}},{key:"configureKeyboardBindings",value:function(){var t,e,i,n,o,r,s,a,h,l,d,c,u,f,p,v,g,y,m,b,w,k,_,x,E=this;(void 0!==this.keycharm&&this.keycharm.destroy(),!0===this.options.keyboard.enabled)&&(!0===this.options.keyboard.bindToWindow?this.keycharm=Sp({container:window,preventDefault:!0}):this.keycharm=Sp({container:this.canvas.frame,preventDefault:!0}),this.keycharm.reset(),!0===this.activated&&(Vt(t=this.keycharm).call(t,"up",(function(){E.bindToRedraw("_moveUp")}),"keydown"),Vt(e=this.keycharm).call(e,"down",(function(){E.bindToRedraw("_moveDown")}),"keydown"),Vt(i=this.keycharm).call(i,"left",(function(){E.bindToRedraw("_moveLeft")}),"keydown"),Vt(n=this.keycharm).call(n,"right",(function(){E.bindToRedraw("_moveRight")}),"keydown"),Vt(o=this.keycharm).call(o,"=",(function(){E.bindToRedraw("_zoomIn")}),"keydown"),Vt(r=this.keycharm).call(r,"num+",(function(){E.bindToRedraw("_zoomIn")}),"keydown"),Vt(s=this.keycharm).call(s,"num-",(function(){E.bindToRedraw("_zoomOut")}),"keydown"),Vt(a=this.keycharm).call(a,"-",(function(){E.bindToRedraw("_zoomOut")}),"keydown"),Vt(h=this.keycharm).call(h,"[",(function(){E.bindToRedraw("_zoomOut")}),"keydown"),Vt(l=this.keycharm).call(l,"]",(function(){E.bindToRedraw("_zoomIn")}),"keydown"),Vt(d=this.keycharm).call(d,"pageup",(function(){E.bindToRedraw("_zoomIn")}),"keydown"),Vt(c=this.keycharm).call(c,"pagedown",(function(){E.bindToRedraw("_zoomOut")}),"keydown"),Vt(u=this.keycharm).call(u,"up",(function(){E.unbindFromRedraw("_moveUp")}),"keyup"),Vt(f=this.keycharm).call(f,"down",(function(){E.unbindFromRedraw("_moveDown")}),"keyup"),Vt(p=this.keycharm).call(p,"left",(function(){E.unbindFromRedraw("_moveLeft")}),"keyup"),Vt(v=this.keycharm).call(v,"right",(function(){E.unbindFromRedraw("_moveRight")}),"keyup"),Vt(g=this.keycharm).call(g,"=",(function(){E.unbindFromRedraw("_zoomIn")}),"keyup"),Vt(y=this.keycharm).call(y,"num+",(function(){E.unbindFromRedraw("_zoomIn")}),"keyup"),Vt(m=this.keycharm).call(m,"num-",(function(){E.unbindFromRedraw("_zoomOut")}),"keyup"),Vt(b=this.keycharm).call(b,"-",(function(){E.unbindFromRedraw("_zoomOut")}),"keyup"),Vt(w=this.keycharm).call(w,"[",(function(){E.unbindFromRedraw("_zoomOut")}),"keyup"),Vt(k=this.keycharm).call(k,"]",(function(){E.unbindFromRedraw("_zoomIn")}),"keyup"),Vt(_=this.keycharm).call(_,"pageup",(function(){E.unbindFromRedraw("_zoomIn")}),"keyup"),Vt(x=this.keycharm).call(x,"pagedown",(function(){E.unbindFromRedraw("_zoomOut")}),"keyup")))}}]),t}();function Pp(t,e){var i=void 0!==ko&&Bi(t)||t["@@iterator"];if(!i){if(So(t)||(i=function(t,e){var i;if(!t)return;if("string"==typeof t)return Dp(t,e);var n=Oo(i=Object.prototype.toString.call(t)).call(i,8,-1);"Object"===n&&t.constructor&&(n=t.constructor.name);if("Map"===n||"Set"===n)return Ei(t);if("Arguments"===n||/^(?:Ui|I)nt(?:8|16|32)(?:Clamped)?Array$/.test(n))return Dp(t,e)}(t))||e&&t&&"number"==typeof t.length){i&&(t=i);var n=0,o=function(){};return{s:o,n:function(){return n>=t.length?{done:!0}:{done:!1,value:t[n++]}},e:function(t){throw t},f:o}}throw new TypeError("Invalid attempt to iterate non-iterable instance.\nIn order to be iterable, non-array objects must have a [Symbol.iterator]() method.")}var r,s=!0,a=!1;return{s:function(){i=i.call(t)},n:function(){var t=i.next();return s=t.done,t},e:function(t){a=!0,r=t},f:function(){try{s||null==i.return||i.return()}finally{if(a)throw r}}}}function Dp(t,e){(null==e||e>t.length)&&(e=t.length);for(var i=0,n=new Array(e);i<e;i++)n[i]=t[i];return n}var Ip=function(){function t(e,i,n){var o,r,s,a,h,l,d,c,u,f,p,v,g;Nn(this,t),this.body=e,this.canvas=i,this.selectionHandler=n,this.navigationHandler=new Mp(e,i),this.body.eventListeners.onTap=Vt(o=this.onTap).call(o,this),this.body.eventListeners.onTouch=Vt(r=this.onTouch).call(r,this),this.body.eventListeners.onDoubleTap=Vt(s=this.onDoubleTap).call(s,this),this.body.eventListeners.onHold=Vt(a=this.onHold).call(a,this),this.body.eventListeners.onDragStart=Vt(h=this.onDragStart).call(h,this),this.body.eventListeners.onDrag=Vt(l=this.onDrag).call(l,this),this.body.eventListeners.onDragEnd=Vt(d=this.onDragEnd).call(d,this),this.body.eventListeners.onMouseWheel=Vt(c=this.onMouseWheel).call(c,this),this.body.eventListeners.onPinch=Vt(u=this.onPinch).call(u,this),this.body.eventListeners.onMouseMove=Vt(f=this.onMouseMove).call(f,this),this.body.eventListeners.onRelease=Vt(p=this.onRelease).call(p,this),this.body.eventListeners.onContext=Vt(v=this.onContext).call(v,this),this.touchTime=0,this.drag={},this.pinch={},this.popup=void 0,this.popupObj=void 0,this.popupTimer=void 0,this.body.functions.getPointer=Vt(g=this.getPointer).call(g,this),this.options={},this.defaultOptions={dragNodes:!0,dragView:!0,hover:!1,keyboard:{enabled:!1,speed:{x:10,y:10,zoom:.02},bindToWindow:!0,autoFocus:!0},navigationButtons:!1,tooltipDelay:300,zoomView:!0,zoomSpeed:1},At(this.options,this.defaultOptions),this.bindEventListeners()}return Fn(t,[{key:"bindEventListeners",value:function(){var t=this;this.body.emitter.on("destroy",(function(){clearTimeout(t.popupTimer),delete t.body.functions.getPointer}))}},{key:"setOptions",value:function(t){if(void 0!==t){Oh(["hideEdgesOnDrag","hideEdgesOnZoom","hideNodesOnDrag","keyboard","multiselect","selectable","selectConnectedEdges"],this.options,t),$h(this.options,t,"keyboard"),t.tooltip&&(At(this.options.tooltip,t.tooltip),t.tooltip.color&&(this.options.tooltip.color=Rh(t.tooltip.color)))}this.navigationHandler.setOptions(this.options)}},{key:"getPointer",value:function(t){return{x:t.x-Mh(this.canvas.frame.canvas),y:t.y-Ph(this.canvas.frame.canvas)}}},{key:"onTouch",value:function(t){(new Date).valueOf()-this.touchTime>50&&(this.drag.pointer=this.getPointer(t.center),this.drag.pinched=!1,this.pinch.scale=this.body.view.scale,this.touchTime=(new Date).valueOf())}},{key:"onTap",value:function(t){var e=this.getPointer(t.center),i=this.selectionHandler.options.multiselect&&(t.changedPointers[0].ctrlKey||t.changedPointers[0].metaKey);this.checkSelectionChanges(e,i),this.selectionHandler.commitAndEmit(e,t),this.selectionHandler.generateClickEvent("click",t,e)}},{key:"onDoubleTap",value:function(t){var e=this.getPointer(t.center);this.selectionHandler.generateClickEvent("doubleClick",t,e)}},{key:"onHold",value:function(t){var e=this.getPointer(t.center),i=this.selectionHandler.options.multiselect;this.checkSelectionChanges(e,i),this.selectionHandler.commitAndEmit(e,t),this.selectionHandler.generateClickEvent("click",t,e),this.selectionHandler.generateClickEvent("hold",t,e)}},{key:"onRelease",value:function(t){if((new Date).valueOf()-this.touchTime>10){var e=this.getPointer(t.center);this.selectionHandler.generateClickEvent("release",t,e),this.touchTime=(new Date).valueOf()}}},{key:"onContext",value:function(t){var e=this.getPointer({x:t.clientX,y:t.clientY});this.selectionHandler.generateClickEvent("oncontext",t,e)}},{key:"checkSelectionChanges",value:function(t){var e=arguments.length>1&&void 0!==arguments[1]&&arguments[1];!0===e?this.selectionHandler.selectAdditionalOnPoint(t):this.selectionHandler.selectOnPoint(t)}},{key:"_determineDifference",value:function(t,e){var i=function(t,e){for(var i=[],n=0;n<t.length;n++){var o=t[n];-1===Hr(e).call(e,o)&&i.push(o)}return i};return{nodes:i(t.nodes,e.nodes),edges:i(t.edges,e.edges)}}},{key:"onDragStart",value:function(t){if(!this.drag.dragging){void 0===this.drag.pointer&&this.onTouch(t);var e=this.selectionHandler.getNodeAt(this.drag.pointer);if(this.drag.dragging=!0,this.drag.selection=[],this.drag.translation=At({},this.body.view.translation),this.drag.nodeId=void 0,t.srcEvent.shiftKey){this.body.selectionBox.show=!0;var i=this.getPointer(t.center);this.body.selectionBox.position.start={x:this.canvas._XconvertDOMtoCanvas(i.x),y:this.canvas._YconvertDOMtoCanvas(i.y)},this.body.selectionBox.position.end={x:this.canvas._XconvertDOMtoCanvas(i.x),y:this.canvas._YconvertDOMtoCanvas(i.y)}}if(void 0!==e&&!0===this.options.dragNodes){this.drag.nodeId=e.id,!1===e.isSelected()&&(this.selectionHandler.unselectAll(),this.selectionHandler.selectObject(e)),this.selectionHandler.generateClickEvent("dragStart",t,this.drag.pointer);var n,o=Pp(this.selectionHandler.getSelectedNodes());try{for(o.s();!(n=o.n()).done;){var r=n.value,s={id:r.id,node:r,x:r.x,y:r.y,xFixed:r.options.fixed.x,yFixed:r.options.fixed.y};r.options.fixed.x=!0,r.options.fixed.y=!0,this.drag.selection.push(s)}}catch(t){o.e(t)}finally{o.f()}}else this.selectionHandler.generateClickEvent("dragStart",t,this.drag.pointer,void 0,!0)}}},{key:"onDrag",value:function(t){var e=this;if(!0!==this.drag.pinched){this.body.emitter.emit("unlockNode");var i=this.getPointer(t.center),n=this.drag.selection;if(n&&n.length&&!0===this.options.dragNodes){this.selectionHandler.generateClickEvent("dragging",t,i);var o=i.x-this.drag.pointer.x,r=i.y-this.drag.pointer.y;Wo(n).call(n,(function(t){var i=t.node;!1===t.xFixed&&(i.x=e.canvas._XconvertDOMtoCanvas(e.canvas._XconvertCanvasToDOM(t.x)+o)),!1===t.yFixed&&(i.y=e.canvas._YconvertDOMtoCanvas(e.canvas._YconvertCanvasToDOM(t.y)+r))})),this.body.emitter.emit("startSimulation")}else{if(t.srcEvent.shiftKey){if(this.selectionHandler.generateClickEvent("dragging",t,i,void 0,!0),void 0===this.drag.pointer)return void this.onDragStart(t);this.body.selectionBox.position.end={x:this.canvas._XconvertDOMtoCanvas(i.x),y:this.canvas._YconvertDOMtoCanvas(i.y)},this.body.emitter.emit("_requestRedraw")}if(!0===this.options.dragView&&!t.srcEvent.shiftKey){if(this.selectionHandler.generateClickEvent("dragging",t,i,void 0,!0),void 0===this.drag.pointer)return void this.onDragStart(t);var s=i.x-this.drag.pointer.x,a=i.y-this.drag.pointer.y;this.body.view.translation={x:this.drag.translation.x+s,y:this.drag.translation.y+a},this.body.emitter.emit("_requestRedraw")}}}}},{key:"onDragEnd",value:function(t){var e=this;if(this.drag.dragging=!1,this.body.selectionBox.show){var i;this.body.selectionBox.show=!1;var n=this.body.selectionBox.position,o={minX:Math.min(n.start.x,n.end.x),minY:Math.min(n.start.y,n.end.y),maxX:Math.max(n.start.x,n.end.x),maxY:Math.max(n.start.y,n.end.y)},r=mr(i=this.body.nodeIndices).call(i,(function(t){var i=e.body.nodes[t];return i.x>=o.minX&&i.x<=o.maxX&&i.y>=o.minY&&i.y<=o.maxY}));Wo(r).call(r,(function(t){return e.selectionHandler.selectObject(e.body.nodes[t])}));var s=this.getPointer(t.center);this.selectionHandler.commitAndEmit(s,t),this.selectionHandler.generateClickEvent("dragEnd",t,this.getPointer(t.center),void 0,!0),this.body.emitter.emit("_requestRedraw")}else{var a=this.drag.selection;a&&a.length?(Wo(a).call(a,(function(t){t.node.options.fixed.x=t.xFixed,t.node.options.fixed.y=t.yFixed})),this.selectionHandler.generateClickEvent("dragEnd",t,this.getPointer(t.center)),this.body.emitter.emit("startSimulation")):(this.selectionHandler.generateClickEvent("dragEnd",t,this.getPointer(t.center),void 0,!0),this.body.emitter.emit("_requestRedraw"))}}},{key:"onPinch",value:function(t){var e=this.getPointer(t.center);this.drag.pinched=!0,void 0===this.pinch.scale&&(this.pinch.scale=1);var i=this.pinch.scale*t.scale;this.zoom(i,e)}},{key:"zoom",value:function(t,e){if(!0===this.options.zoomView){var i=this.body.view.scale;t<1e-5&&(t=1e-5),t>10&&(t=10);var n=void 0;void 0!==this.drag&&!0===this.drag.dragging&&(n=this.canvas.DOMtoCanvas(this.drag.pointer));var o=this.body.view.translation,r=t/i,s=(1-r)*e.x+o.x*r,a=(1-r)*e.y+o.y*r;if(this.body.view.scale=t,this.body.view.translation={x:s,y:a},null!=n){var h=this.canvas.canvasToDOM(n);this.drag.pointer.x=h.x,this.drag.pointer.y=h.y}this.body.emitter.emit("_requestRedraw"),i<t?this.body.emitter.emit("zoom",{direction:"+",scale:this.body.view.scale,pointer:e}):this.body.emitter.emit("zoom",{direction:"-",scale:this.body.view.scale,pointer:e})}}},{key:"onMouseWheel",value:function(t){if(!0===this.options.zoomView){if(0!==t.deltaY){var e=this.body.view.scale;e*=1+(t.deltaY<0?1:-1)*(.1*this.options.zoomSpeed);var i=this.getPointer({x:t.clientX,y:t.clientY});this.zoom(e,i)}t.preventDefault()}}},{key:"onMouseMove",value:function(t){var e=this,i=this.getPointer({x:t.clientX,y:t.clientY}),n=!1;void 0!==this.popup&&(!1===this.popup.hidden&&this._checkHidePopup(i),!1===this.popup.hidden&&(n=!0,this.popup.setPosition(i.x+3,i.y-5),this.popup.show())),this.options.keyboard.autoFocus&&!1===this.options.keyboard.bindToWindow&&!0===this.options.keyboard.enabled&&this.canvas.frame.focus(),!1===n&&(void 0!==this.popupTimer&&(clearInterval(this.popupTimer),this.popupTimer=void 0),this.drag.dragging||(this.popupTimer=rs((function(){return e._checkShowPopup(i)}),this.options.tooltipDelay))),!0===this.options.hover&&this.selectionHandler.hoverObject(t,i)}},{key:"_checkShowPopup",value:function(t){var e=this.canvas._XconvertDOMtoCanvas(t.x),i=this.canvas._YconvertDOMtoCanvas(t.y),n={left:e,top:i,right:e,bottom:i},o=void 0===this.popupObj?void 0:this.popupObj.id,r=!1,s="node";if(void 0===this.popupObj){for(var a,h=this.body.nodeIndices,l=this.body.nodes,d=[],c=0;c<h.length;c++)!0===(a=l[h[c]]).isOverlappingWith(n)&&(r=!0,void 0!==a.getTitle()&&d.push(h[c]));d.length>0&&(this.popupObj=l[d[d.length-1]],r=!0)}if(void 0===this.popupObj&&!1===r){for(var u,f=this.body.edgeIndices,p=this.body.edges,v=[],g=0;g<f.length;g++)!0===(u=p[f[g]]).isOverlappingWith(n)&&!0===u.connected&&void 0!==u.getTitle()&&v.push(f[g]);v.length>0&&(this.popupObj=p[v[v.length-1]],s="edge")}void 0!==this.popupObj?this.popupObj.id!==o&&(void 0===this.popup&&(this.popup=new dl(this.canvas.frame)),this.popup.popupTargetType=s,this.popup.popupTargetId=this.popupObj.id,this.popup.setPosition(t.x+3,t.y-5),this.popup.setText(this.popupObj.getTitle()),this.popup.show(),this.body.emitter.emit("showPopup",this.popupObj.id)):void 0!==this.popup&&(this.popup.hide(),this.body.emitter.emit("hidePopup"))}},{key:"_checkHidePopup",value:function(t){var e=this.selectionHandler._pointerToPositionObject(t),i=!1;if("node"===this.popup.popupTargetType){if(void 0!==this.body.nodes[this.popup.popupTargetId]&&!0===(i=this.body.nodes[this.popup.popupTargetId].isOverlappingWith(e))){var n=this.selectionHandler.getNodeAt(t);i=void 0!==n&&n.id===this.popup.popupTargetId}}else void 0===this.selectionHandler.getNodeAt(t)&&void 0!==this.body.edges[this.popup.popupTargetId]&&(i=this.body.edges[this.popup.popupTargetId].isOverlappingWith(e));!1===i&&(this.popupObj=void 0,this.popup.hide(),this.body.emitter.emit("hidePopup"))}}]),t}(),Bp=Jl.getWeakData,zp=we.set,Np=we.getterFor,Ap=Gi.find,Fp=Gi.findIndex,jp=0,Rp=function(t){return t.frozen||(t.frozen=new Lp)},Lp=function(){this.entries=[]},Hp=function(t,e){return Ap(t.entries,(function(t){return t[0]===e}))};Lp.prototype={get:function(t){var e=Hp(this,t);if(e)return e[1]},has:function(t){return!!Hp(this,t)},set:function(t,e){var i=Hp(this,t);i?i[1]=e:this.entries.push([t,e])},delete:function(t){var e=Fp(this.entries,(function(e){return e[0]===t}));return~e&&this.entries.splice(e,1),!!~e}};var Wp={getConstructor:function(t,e,i,n){var o=t((function(t,r){id(t,o,e),zp(t,{type:e,id:jp++,frozen:void 0}),null!=r&&ed(r,t[n],{that:t,AS_ENTRIES:i})})),r=Np(e),s=function(t,e,i){var n=r(t),o=Bp(dt(e),!0);return!0===o?Rp(n).set(e,i):o[n.id]=i,t};return hd(o.prototype,{delete:function(t){var e=r(this);if(!w(t))return!1;var i=Bp(t);return!0===i?Rp(e).delete(t):i&&j(i,e.id)&&delete i[e.id]},has:function(t){var e=r(this);if(!w(t))return!1;var i=Bp(t);return!0===i?Rp(e).has(t):i&&j(i,e.id)}}),hd(o.prototype,i?{get:function(t){var e=r(this);if(w(t)){var i=Bp(t);return!0===i?Rp(e).get(t):i?i[e.id]:void 0}},set:function(t,e){return s(this,t,e)}}:{add:function(t){return s(this,t,!0)}}),o}};n((function(t){var e,i=we.enforce,n=!a.ActiveXObject&&"ActiveXObject"in a,o=Object.isExtensible,r=function(t){return function(){return t(this,arguments.length?arguments[0]:void 0)}},s=t.exports=ad("WeakMap",r,Wp);if(ae&&n){e=Wp.getConstructor(r,"WeakMap",!0),Jl.enable();var h=s.prototype,l=h.delete,d=h.has,c=h.get,u=h.set;hd(h,{delete:function(t){if(w(t)&&!o(t)){var n=i(this);return n.frozen||(n.frozen=new e),l.call(this,t)||n.frozen.delete(t)}return l.call(this,t)},has:function(t){if(w(t)&&!o(t)){var n=i(this);return n.frozen||(n.frozen=new e),d.call(this,t)||n.frozen.has(t)}return d.call(this,t)},get:function(t){if(w(t)&&!o(t)){var n=i(this);return n.frozen||(n.frozen=new e),d.call(this,t)?c.call(this,t):n.frozen.get(t)}return c.call(this,t)},set:function(t,n){if(w(t)&&!o(t)){var r=i(this);r.frozen||(r.frozen=new e),d.call(this,t)?u.call(this,t,n):r.frozen.set(t,n)}else u.call(this,t,n);return this}})}}));var qp,Vp,Up,Yp,Xp,Gp=k.WeakMap;function Kp(t,e,i,n){if("a"===i&&!n)throw new TypeError("Private accessor was defined without a getter");if("function"==typeof e?t!==e||!n:!e.has(t))throw new TypeError("Cannot read private member from an object whose class did not declare it");return"m"===i?n:"a"===i?n.call(t):n?n.value:e.get(t)}function $p(t,e,i,n,o){if("m"===n)throw new TypeError("Private method is not writable");if("a"===n&&!o)throw new TypeError("Private accessor was defined without a setter");if("function"==typeof e?t!==e||!o:!e.has(t))throw new TypeError("Cannot write private member to an object whose class did not declare it");return"a"===n?o.call(t,i):o?o.value=i:e.set(t,i),i}function Zp(t,e){var i=void 0!==ko&&Bi(t)||t["@@iterator"];if(!i){if(So(t)||(i=function(t,e){var i;if(!t)return;if("string"==typeof t)return Qp(t,e);var n=Oo(i=Object.prototype.toString.call(t)).call(i,8,-1);"Object"===n&&t.constructor&&(n=t.constructor.name);if("Map"===n||"Set"===n)return Ei(t);if("Arguments"===n||/^(?:Ui|I)nt(?:8|16|32)(?:Clamped)?Array$/.test(n))return Qp(t,e)}(t))||e&&t&&"number"==typeof t.length){i&&(t=i);var n=0,o=function(){};return{s:o,n:function(){return n>=t.length?{done:!0}:{done:!1,value:t[n++]}},e:function(t){throw t},f:o}}throw new TypeError("Invalid attempt to iterate non-iterable instance.\nIn order to be iterable, non-array objects must have a [Symbol.iterator]() method.")}var r,s=!0,a=!1;return{s:function(){i=i.call(t)},n:function(){var t=i.next();return s=t.done,t},e:function(t){a=!0,r=t},f:function(){try{s||null==i.return||i.return()}finally{if(a)throw r}}}}function Qp(t,e){(null==e||e>t.length)&&(e=t.length);for(var i=0,n=new Array(e);i<e;i++)n[i]=t[i];return n}function Jp(t,e){var i,n=new ec,o=Zp(e);try{for(o.s();!(i=o.n()).done;){var r=i.value;t.has(r)||n.add(r)}}catch(t){o.e(t)}finally{o.f()}return n}var tv=function(){function t(){Nn(this,t),qp.set(this,new ec),Vp.set(this,new ec)}return Fn(t,[{key:"size",get:function(){return Kp(this,Vp,"f").size}},{key:"add",value:function(){for(var t=arguments.length,e=new Array(t),i=0;i<t;i++)e[i]=arguments[i];for(var n=0,o=e;n<o.length;n++){var r=o[n];Kp(this,Vp,"f").add(r)}}},{key:"delete",value:function(){for(var t=arguments.length,e=new Array(t),i=0;i<t;i++)e[i]=arguments[i];for(var n=0,o=e;n<o.length;n++){var r=o[n];Kp(this,Vp,"f").delete(r)}}},{key:"clear",value:function(){Kp(this,Vp,"f").clear()}},{key:"getSelection",value:function(){return wo(Kp(this,Vp,"f"))}},{key:"getChanges",value:function(){return{added:wo(Jp(Kp(this,qp,"f"),Kp(this,Vp,"f"))),deleted:wo(Jp(Kp(this,Vp,"f"),Kp(this,qp,"f"))),previous:wo(new ec(Kp(this,qp,"f"))),current:wo(new ec(Kp(this,Vp,"f")))}}},{key:"commit",value:function(){var t=this.getChanges();$p(this,qp,Kp(this,Vp,"f"),"f"),$p(this,Vp,new ec(Kp(this,qp,"f")),"f");var e,i=Zp(t.added);try{for(i.s();!(e=i.n()).done;){e.value.select()}}catch(t){i.e(t)}finally{i.f()}var n,o=Zp(t.deleted);try{for(o.s();!(n=o.n()).done;){n.value.unselect()}}catch(t){o.e(t)}finally{o.f()}return t}}]),t}();qp=new Gp,Vp=new Gp;var ev=function(){function t(){var e=arguments.length>0&&void 0!==arguments[0]?arguments[0]:function(){};Nn(this,t),Up.set(this,new tv),Yp.set(this,new tv),Xp.set(this,void 0),$p(this,Xp,e,"f")}return Fn(t,[{key:"sizeNodes",get:function(){return Kp(this,Up,"f").size}},{key:"sizeEdges",get:function(){return Kp(this,Yp,"f").size}},{key:"getNodes",value:function(){return Kp(this,Up,"f").getSelection()}},{key:"getEdges",value:function(){return Kp(this,Yp,"f").getSelection()}},{key:"addNodes",value:function(){var t;(t=Kp(this,Up,"f")).add.apply(t,arguments)}},{key:"addEdges",value:function(){var t;(t=Kp(this,Yp,"f")).add.apply(t,arguments)}},{key:"deleteNodes",value:function(t){Kp(this,Up,"f").delete(t)}},{key:"deleteEdges",value:function(t){Kp(this,Yp,"f").delete(t)}},{key:"clear",value:function(){Kp(this,Up,"f").clear(),Kp(this,Yp,"f").clear()}},{key:"commit",value:function(){for(var t,e,i={nodes:Kp(this,Up,"f").commit(),edges:Kp(this,Yp,"f").commit()},n=arguments.length,o=new Array(n),r=0;r<n;r++)o[r]=arguments[r];return(t=Kp(this,Xp,"f")).call.apply(t,Eo(e=[this,i]).call(e,o)),i}}]),t}();function iv(t,e){var i=void 0!==ko&&Bi(t)||t["@@iterator"];if(!i){if(So(t)||(i=function(t,e){var i;if(!t)return;if("string"==typeof t)return nv(t,e);var n=Oo(i=Object.prototype.toString.call(t)).call(i,8,-1);"Object"===n&&t.constructor&&(n=t.constructor.name);if("Map"===n||"Set"===n)return Ei(t);if("Arguments"===n||/^(?:Ui|I)nt(?:8|16|32)(?:Clamped)?Array$/.test(n))return nv(t,e)}(t))||e&&t&&"number"==typeof t.length){i&&(t=i);var n=0,o=function(){};return{s:o,n:function(){return n>=t.length?{done:!0}:{done:!1,value:t[n++]}},e:function(t){throw t},f:o}}throw new TypeError("Invalid attempt to iterate non-iterable instance.\nIn order to be iterable, non-array objects must have a [Symbol.iterator]() method.")}var r,s=!0,a=!1;return{s:function(){i=i.call(t)},n:function(){var t=i.next();return s=t.done,t},e:function(t){a=!0,r=t},f:function(){try{s||null==i.return||i.return()}finally{if(a)throw r}}}}function nv(t,e){(null==e||e>t.length)&&(e=t.length);for(var i=0,n=new Array(e);i<e;i++)n[i]=t[i];return n}Up=new Gp,Yp=new Gp,Xp=new Gp;var ov=function(){function t(e,i){var n=this;Nn(this,t),this.body=e,this.canvas=i,this._selectionAccumulator=new ev,this.hoverObj={nodes:{},edges:{}},this.options={},this.defaultOptions={multiselect:!1,selectable:!0,selectConnectedEdges:!0,hoverConnectedEdges:!0},At(this.options,this.defaultOptions),this.body.emitter.on("_dataChanged",(function(){n.updateSelection()}))}return Fn(t,[{key:"setOptions",value:function(t){if(void 0!==t){Eh(["multiselect","hoverConnectedEdges","selectable","selectConnectedEdges"],this.options,t)}}},{key:"selectOnPoint",value:function(t){var e=!1;if(!0===this.options.selectable){var i=this.getNodeAt(t)||this.getEdgeAt(t);this.unselectAll(),void 0!==i&&(e=this.selectObject(i)),this.body.emitter.emit("_requestRedraw")}return e}},{key:"selectAdditionalOnPoint",value:function(t){var e=!1;if(!0===this.options.selectable){var i=this.getNodeAt(t)||this.getEdgeAt(t);void 0!==i&&(e=!0,!0===i.isSelected()?this.deselectObject(i):this.selectObject(i),this.body.emitter.emit("_requestRedraw"))}return e}},{key:"_initBaseEvent",value:function(t,e){var i={};return i.pointer={DOM:{x:e.x,y:e.y},canvas:this.canvas.DOMtoCanvas(e)},i.event=t,i}},{key:"generateClickEvent",value:function(t,e,i,n){var o=arguments.length>4&&void 0!==arguments[4]&&arguments[4],r=this._initBaseEvent(e,i);if(!0===o)r.nodes=[],r.edges=[];else{var s=this.getSelection();r.nodes=s.nodes,r.edges=s.edges}void 0!==n&&(r.previousSelection=n),"click"==t&&(r.items=this.getClickedItems(i)),void 0!==e.controlEdge&&(r.controlEdge=e.controlEdge),this.body.emitter.emit(t,r)}},{key:"selectObject",value:function(t){var e=arguments.length>1&&void 0!==arguments[1]?arguments[1]:this.options.selectConnectedEdges;if(void 0!==t){if(t instanceof cf){var i;if(!0===e)(i=this._selectionAccumulator).addEdges.apply(i,wo(t.edges));this._selectionAccumulator.addNodes(t)}else this._selectionAccumulator.addEdges(t);return!0}return!1}},{key:"deselectObject",value:function(t){!0===t.isSelected()&&(t.selected=!1,this._removeFromSelection(t))}},{key:"_getAllNodesOverlappingWith",value:function(t){for(var e=[],i=this.body.nodes,n=0;n<this.body.nodeIndices.length;n++){var o=this.body.nodeIndices[n];i[o].isOverlappingWith(t)&&e.push(o)}return e}},{key:"_pointerToPositionObject",value:function(t){var e=this.canvas.DOMtoCanvas(t);return{left:e.x-1,top:e.y+1,right:e.x+1,bottom:e.y-1}}},{key:"getNodeAt",value:function(t){var e=!(arguments.length>1&&void 0!==arguments[1])||arguments[1],i=this._pointerToPositionObject(t),n=this._getAllNodesOverlappingWith(i);return n.length>0?!0===e?this.body.nodes[n[n.length-1]]:n[n.length-1]:void 0}},{key:"_getEdgesOverlappingWith",value:function(t,e){for(var i=this.body.edges,n=0;n<this.body.edgeIndices.length;n++){var o=this.body.edgeIndices[n];i[o].isOverlappingWith(t)&&e.push(o)}}},{key:"_getAllEdgesOverlappingWith",value:function(t){var e=[];return this._getEdgesOverlappingWith(t,e),e}},{key:"getEdgeAt",value:function(t){for(var e=!(arguments.length>1&&void 0!==arguments[1])||arguments[1],i=this.canvas.DOMtoCanvas(t),n=10,o=null,r=this.body.edges,s=0;s<this.body.edgeIndices.length;s++){var a=this.body.edgeIndices[s],h=r[a];if(h.connected){var l=h.from.x,d=h.from.y,c=h.to.x,u=h.to.y,f=h.edgeType.getDistanceToEdge(l,d,c,u,i.x,i.y);f<n&&(o=a,n=f)}}return null!==o?!0===e?this.body.edges[o]:o:void 0}},{key:"_addToHover",value:function(t){t instanceof cf?this.hoverObj.nodes[t.id]=t:this.hoverObj.edges[t.id]=t}},{key:"_removeFromSelection",value:function(t){var e;t instanceof cf?(this._selectionAccumulator.deleteNodes(t),(e=this._selectionAccumulator).deleteEdges.apply(e,wo(t.edges))):this._selectionAccumulator.deleteEdges(t)}},{key:"unselectAll",value:function(){this._selectionAccumulator.clear()}},{key:"getSelectedNodeCount",value:function(){return this._selectionAccumulator.sizeNodes}},{key:"getSelectedEdgeCount",value:function(){return this._selectionAccumulator.sizeEdges}},{key:"_hoverConnectedEdges",value:function(t){for(var e=0;e<t.edges.length;e++){var i=t.edges[e];i.hover=!0,this._addToHover(i)}}},{key:"emitBlurEvent",value:function(t,e,i){var n=this._initBaseEvent(t,e);!0===i.hover&&(i.hover=!1,i instanceof cf?(n.node=i.id,this.body.emitter.emit("blurNode",n)):(n.edge=i.id,this.body.emitter.emit("blurEdge",n)))}},{key:"emitHoverEvent",value:function(t,e,i){var n=this._initBaseEvent(t,e),o=!1;return!1===i.hover&&(i.hover=!0,this._addToHover(i),o=!0,i instanceof cf?(n.node=i.id,this.body.emitter.emit("hoverNode",n)):(n.edge=i.id,this.body.emitter.emit("hoverEdge",n))),o}},{key:"hoverObject",value:function(t,e){var i=this.getNodeAt(e);void 0===i&&(i=this.getEdgeAt(e));var n=!1;for(var o in this.hoverObj.nodes)Object.prototype.hasOwnProperty.call(this.hoverObj.nodes,o)&&(void 0===i||i instanceof cf&&i.id!=o||i instanceof ep)&&(this.emitBlurEvent(t,e,this.hoverObj.nodes[o]),delete this.hoverObj.nodes[o],n=!0);for(var r in this.hoverObj.edges)Object.prototype.hasOwnProperty.call(this.hoverObj.edges,r)&&(!0===n?(this.hoverObj.edges[r].hover=!1,delete this.hoverObj.edges[r]):(void 0===i||i instanceof ep&&i.id!=r||i instanceof cf&&!i.hover)&&(this.emitBlurEvent(t,e,this.hoverObj.edges[r]),delete this.hoverObj.edges[r],n=!0));if(void 0!==i){var s=zo(this.hoverObj.edges).length,a=zo(this.hoverObj.nodes).length;(n||i instanceof ep&&0===s&&0===a||i instanceof cf&&0===s&&0===a)&&(n=this.emitHoverEvent(t,e,i)),i instanceof cf&&!0===this.options.hoverConnectedEdges&&this._hoverConnectedEdges(i)}!0===n&&this.body.emitter.emit("_requestRedraw")}},{key:"commitWithoutEmitting",value:function(){this._selectionAccumulator.commit()}},{key:"commitAndEmit",value:function(t,e){var i=!1,n=this._selectionAccumulator.commit(),o={nodes:n.nodes.previous,edges:n.edges.previous};n.edges.deleted.length>0&&(this.generateClickEvent("deselectEdge",e,t,o),i=!0),n.nodes.deleted.length>0&&(this.generateClickEvent("deselectNode",e,t,o),i=!0),n.nodes.added.length>0&&(this.generateClickEvent("selectNode",e,t),i=!0),n.edges.added.length>0&&(this.generateClickEvent("selectEdge",e,t),i=!0),!0===i&&this.generateClickEvent("select",e,t)}},{key:"getSelection",value:function(){return{nodes:this.getSelectedNodeIds(),edges:this.getSelectedEdgeIds()}}},{key:"getSelectedNodes",value:function(){return this._selectionAccumulator.getNodes()}},{key:"getSelectedEdges",value:function(){return this._selectionAccumulator.getEdges()}},{key:"getSelectedNodeIds",value:function(){var t;return Io(t=this._selectionAccumulator.getNodes()).call(t,(function(t){return t.id}))}},{key:"getSelectedEdgeIds",value:function(){var t;return Io(t=this._selectionAccumulator.getEdges()).call(t,(function(t){return t.id}))}},{key:"setSelection",value:function(t){var e=arguments.length>1&&void 0!==arguments[1]?arguments[1]:{};if(!t||!t.nodes&&!t.edges)throw new TypeError("Selection must be an object with nodes and/or edges properties");if((e.unselectAll||void 0===e.unselectAll)&&this.unselectAll(),t.nodes){var i,n=iv(t.nodes);try{for(n.s();!(i=n.n()).done;){var o=i.value,r=this.body.nodes[o];if(!r)throw new RangeError('Node with id "'+o+'" not found');this.selectObject(r,e.highlightEdges)}}catch(t){n.e(t)}finally{n.f()}}if(t.edges){var s,a=iv(t.edges);try{for(a.s();!(s=a.n()).done;){var h=s.value,l=this.body.edges[h];if(!l)throw new RangeError('Edge with id "'+h+'" not found');this.selectObject(l)}}catch(t){a.e(t)}finally{a.f()}}this.body.emitter.emit("_requestRedraw"),this._selectionAccumulator.commit()}},{key:"selectNodes",value:function(t){var e=!(arguments.length>1&&void 0!==arguments[1])||arguments[1];if(!t||void 0===t.length)throw"Selection must be an array with ids";this.setSelection({nodes:t},{highlightEdges:e})}},{key:"selectEdges",value:function(t){if(!t||void 0===t.length)throw"Selection must be an array with ids";this.setSelection({edges:t})}},{key:"updateSelection",value:function(){for(var t in this._selectionAccumulator.getNodes())Object.prototype.hasOwnProperty.call(this.body.nodes,t.id)||this._selectionAccumulator.deleteNodes(t);for(var e in this._selectionAccumulator.getEdges())Object.prototype.hasOwnProperty.call(this.body.edges,e.id)||this._selectionAccumulator.deleteEdges(e)}},{key:"getClickedItems",value:function(t){for(var e=this.canvas.DOMtoCanvas(t),i=[],n=this.body.nodeIndices,o=this.body.nodes,r=n.length-1;r>=0;r--){var s=o[n[r]].getItemsOnPoint(e);i.push.apply(i,s)}for(var a=this.body.edgeIndices,h=this.body.edges,l=a.length-1;l>=0;l--){var d=h[a[l]].getItemsOnPoint(e);i.push.apply(i,d)}return i}}]),t}(),rv=n((function(t,e){!function(t){function e(t,e){if(!(t instanceof e))throw new TypeError("Cannot call a class as a function")}t.__esModule=!0,t.sort=v;var i=32,n=7,o=256,r=[1,10,100,1e3,1e4,1e5,1e6,1e7,1e8,1e9];function s(t){return t<1e5?t<100?t<10?0:1:t<1e4?t<1e3?2:3:4:t<1e7?t<1e6?5:6:t<1e9?t<1e8?7:8:9}function a(t,e){if(t===e)return 0;if(~~t===t&&~~e===e){if(0===t||0===e)return t<e?-1:1;if(t<0||e<0){if(e>=0)return-1;if(t>=0)return 1;t=-t,e=-e}var i=s(t),n=s(e),o=0;return i<n?(t*=r[n-i-1],e/=10,o=-1):i>n&&(e*=r[i-n-1],t/=10,o=1),t===e?o:t<e?-1:1}var a=String(t),h=String(e);return a===h?0:a<h?-1:1}function h(t){for(var e=0;t>=i;)e|=1&t,t>>=1;return t+e}function l(t,e,i,n){var o=e+1;if(o===i)return 1;if(n(t[o++],t[e])<0){for(;o<i&&n(t[o],t[o-1])<0;)o++;d(t,e,o)}else for(;o<i&&n(t[o],t[o-1])>=0;)o++;return o-e}function d(t,e,i){for(i--;e<i;){var n=t[e];t[e++]=t[i],t[i--]=n}}function c(t,e,i,n,o){for(n===e&&n++;n<i;n++){for(var r=t[n],s=e,a=n;s<a;){var h=s+a>>>1;o(r,t[h])<0?a=h:s=h+1}var l=n-s;switch(l){case 3:t[s+3]=t[s+2];case 2:t[s+2]=t[s+1];case 1:t[s+1]=t[s];break;default:for(;l>0;)t[s+l]=t[s+l-1],l--}t[s]=r}}function u(t,e,i,n,o,r){var s=0,a=0,h=1;if(r(t,e[i+o])>0){for(a=n-o;h<a&&r(t,e[i+o+h])>0;)s=h,(h=1+(h<<1))<=0&&(h=a);h>a&&(h=a),s+=o,h+=o}else{for(a=o+1;h<a&&r(t,e[i+o-h])<=0;)s=h,(h=1+(h<<1))<=0&&(h=a);h>a&&(h=a);var l=s;s=o-h,h=o-l}for(s++;s<h;){var d=s+(h-s>>>1);r(t,e[i+d])>0?s=d+1:h=d}return h}function f(t,e,i,n,o,r){var s=0,a=0,h=1;if(r(t,e[i+o])<0){for(a=o+1;h<a&&r(t,e[i+o-h])<0;)s=h,(h=1+(h<<1))<=0&&(h=a);h>a&&(h=a);var l=s;s=o-h,h=o-l}else{for(a=n-o;h<a&&r(t,e[i+o+h])>=0;)s=h,(h=1+(h<<1))<=0&&(h=a);h>a&&(h=a),s+=o,h+=o}for(s++;s<h;){var d=s+(h-s>>>1);r(t,e[i+d])<0?h=d:s=d+1}return h}var p=function(){function t(i,r){e(this,t),this.array=null,this.compare=null,this.minGallop=n,this.length=0,this.tmpStorageLength=o,this.stackLength=0,this.runStart=null,this.runLength=null,this.stackSize=0,this.array=i,this.compare=r,this.length=i.length,this.length<2*o&&(this.tmpStorageLength=this.length>>>1),this.tmp=new Array(this.tmpStorageLength),this.stackLength=this.length<120?5:this.length<1542?10:this.length<119151?19:40,this.runStart=new Array(this.stackLength),this.runLength=new Array(this.stackLength)}return t.prototype.pushRun=function(t,e){this.runStart[this.stackSize]=t,this.runLength[this.stackSize]=e,this.stackSize+=1},t.prototype.mergeRuns=function(){for(;this.stackSize>1;){var t=this.stackSize-2;if(t>=1&&this.runLength[t-1]<=this.runLength[t]+this.runLength[t+1]||t>=2&&this.runLength[t-2]<=this.runLength[t]+this.runLength[t-1])this.runLength[t-1]<this.runLength[t+1]&&t--;else if(this.runLength[t]>this.runLength[t+1])break;this.mergeAt(t)}},t.prototype.forceMergeRuns=function(){for(;this.stackSize>1;){var t=this.stackSize-2;t>0&&this.runLength[t-1]<this.runLength[t+1]&&t--,this.mergeAt(t)}},t.prototype.mergeAt=function(t){var e=this.compare,i=this.array,n=this.runStart[t],o=this.runLength[t],r=this.runStart[t+1],s=this.runLength[t+1];this.runLength[t]=o+s,t===this.stackSize-3&&(this.runStart[t+1]=this.runStart[t+2],this.runLength[t+1]=this.runLength[t+2]),this.stackSize--;var a=f(i[r],i,n,o,0,e);n+=a,0!=(o-=a)&&0!==(s=u(i[n+o-1],i,r,s,s-1,e))&&(o<=s?this.mergeLow(n,o,r,s):this.mergeHigh(n,o,r,s))},t.prototype.mergeLow=function(t,e,i,o){var r=this.compare,s=this.array,a=this.tmp,h=0;for(h=0;h<e;h++)a[h]=s[t+h];var l=0,d=i,c=t;if(s[c++]=s[d++],0!=--o)if(1!==e){for(var p=this.minGallop;;){var v=0,g=0,y=!1;do{if(r(s[d],a[l])<0){if(s[c++]=s[d++],g++,v=0,0==--o){y=!0;break}}else if(s[c++]=a[l++],v++,g=0,1==--e){y=!0;break}}while((v|g)<p);if(y)break;do{if(0!==(v=f(s[d],a,l,e,0,r))){for(h=0;h<v;h++)s[c+h]=a[l+h];if(c+=v,l+=v,(e-=v)<=1){y=!0;break}}if(s[c++]=s[d++],0==--o){y=!0;break}if(0!==(g=u(a[l],s,d,o,0,r))){for(h=0;h<g;h++)s[c+h]=s[d+h];if(c+=g,d+=g,0==(o-=g)){y=!0;break}}if(s[c++]=a[l++],1==--e){y=!0;break}p--}while(v>=n||g>=n);if(y)break;p<0&&(p=0),p+=2}if(this.minGallop=p,p<1&&(this.minGallop=1),1===e){for(h=0;h<o;h++)s[c+h]=s[d+h];s[c+o]=a[l]}else{if(0===e)throw new Error("mergeLow preconditions were not respected");for(h=0;h<e;h++)s[c+h]=a[l+h]}}else{for(h=0;h<o;h++)s[c+h]=s[d+h];s[c+o]=a[l]}else for(h=0;h<e;h++)s[c+h]=a[l+h]},t.prototype.mergeHigh=function(t,e,i,o){var r=this.compare,s=this.array,a=this.tmp,h=0;for(h=0;h<o;h++)a[h]=s[i+h];var l=t+e-1,d=o-1,c=i+o-1,p=0,v=0;if(s[c--]=s[l--],0!=--e)if(1!==o){for(var g=this.minGallop;;){var y=0,m=0,b=!1;do{if(r(a[d],s[l])<0){if(s[c--]=s[l--],y++,m=0,0==--e){b=!0;break}}else if(s[c--]=a[d--],m++,y=0,1==--o){b=!0;break}}while((y|m)<g);if(b)break;do{if(0!=(y=e-f(a[d],s,t,e,e-1,r))){for(e-=y,v=1+(c-=y),p=1+(l-=y),h=y-1;h>=0;h--)s[v+h]=s[p+h];if(0===e){b=!0;break}}if(s[c--]=a[d--],1==--o){b=!0;break}if(0!=(m=o-u(s[l],a,0,o,o-1,r))){for(o-=m,v=1+(c-=m),p=1+(d-=m),h=0;h<m;h++)s[v+h]=a[p+h];if(o<=1){b=!0;break}}if(s[c--]=s[l--],0==--e){b=!0;break}g--}while(y>=n||m>=n);if(b)break;g<0&&(g=0),g+=2}if(this.minGallop=g,g<1&&(this.minGallop=1),1===o){for(v=1+(c-=e),p=1+(l-=e),h=e-1;h>=0;h--)s[v+h]=s[p+h];s[c]=a[d]}else{if(0===o)throw new Error("mergeHigh preconditions were not respected");for(p=c-(o-1),h=0;h<o;h++)s[p+h]=a[h]}}else{for(v=1+(c-=e),p=1+(l-=e),h=e-1;h>=0;h--)s[v+h]=s[p+h];s[c]=a[d]}else for(p=c-(o-1),h=0;h<o;h++)s[p+h]=a[h]},t}();function v(t,e,n,o){if(!Array.isArray(t))throw new TypeError("Can only sort arrays");e?"function"!=typeof e&&(o=n,n=e,e=a):e=a,n||(n=0),o||(o=t.length);var r=o-n;if(!(r<2)){var s=0;if(r<i)c(t,n,o,n+(s=l(t,n,o,e)),e);else{var d=new p(t,e),u=h(r);do{if((s=l(t,n,o,e))<u){var f=r;f>u&&(f=u),c(t,n,n+f,n+s,e),s=f}d.pushRun(n,s),d.mergeRuns(),r-=s,n+=s}while(0!==r);d.forceMergeRuns()}}}}(e)}));i(rv);var sv=rv,av=sv.sort;function hv(t){var e=function(){if("undefined"==typeof Reflect||!Pd)return!1;if(Pd.sham)return!1;if("function"==typeof Proxy)return!0;try{return Boolean.prototype.valueOf.call(Pd(Boolean,[],(function(){}))),!0}catch(t){return!1}}();return function(){var i,n=Ld(t);if(e){var o=Ld(this).constructor;i=Pd(n,arguments,o)}else i=n.apply(this,arguments);return Fd(this,i)}}var lv=function(){function t(){Nn(this,t)}return Fn(t,[{key:"abstract",value:function(){throw new Error("Can't instantiate abstract class!")}},{key:"fake_use",value:function(){}},{key:"curveType",value:function(){return this.abstract()}},{key:"getPosition",value:function(t){return this.fake_use(t),this.abstract()}},{key:"setPosition",value:function(t,e){var i=arguments.length>2&&void 0!==arguments[2]?arguments[2]:void 0;this.fake_use(t,e,i),this.abstract()}},{key:"getTreeSize",value:function(t){return this.fake_use(t),this.abstract()}},{key:"sort",value:function(t){this.fake_use(t),this.abstract()}},{key:"fix",value:function(t,e){this.fake_use(t,e),this.abstract()}},{key:"shift",value:function(t,e){this.fake_use(t,e),this.abstract()}}]),t}(),dv=function(t){Ad(i,t);var e=hv(i);function i(t){var n;return Nn(this,i),(n=e.call(this)).layout=t,n}return Fn(i,[{key:"curveType",value:function(){return"horizontal"}},{key:"getPosition",value:function(t){return t.x}},{key:"setPosition",value:function(t,e){var i=arguments.length>2&&void 0!==arguments[2]?arguments[2]:void 0;void 0!==i&&this.layout.hierarchical.addToOrdering(t,i),t.x=e}},{key:"getTreeSize",value:function(t){var e=this.layout.hierarchical.getTreeSize(this.layout.body.nodes,t);return{min:e.min_x,max:e.max_x}}},{key:"sort",value:function(t){av(t,(function(t,e){return t.x-e.x}))}},{key:"fix",value:function(t,e){t.y=this.layout.options.hierarchical.levelSeparation*e,t.options.fixed.y=!0}},{key:"shift",value:function(t,e){this.layout.body.nodes[t].x+=e}}]),i}(lv),cv=function(t){Ad(i,t);var e=hv(i);function i(t){var n;return Nn(this,i),(n=e.call(this)).layout=t,n}return Fn(i,[{key:"curveType",value:function(){return"vertical"}},{key:"getPosition",value:function(t){return t.y}},{key:"setPosition",value:function(t,e){var i=arguments.length>2&&void 0!==arguments[2]?arguments[2]:void 0;void 0!==i&&this.layout.hierarchical.addToOrdering(t,i),t.y=e}},{key:"getTreeSize",value:function(t){var e=this.layout.hierarchical.getTreeSize(this.layout.body.nodes,t);return{min:e.min_y,max:e.max_y}}},{key:"sort",value:function(t){av(t,(function(t,e){return t.y-e.y}))}},{key:"fix",value:function(t,e){t.x=this.layout.options.hierarchical.levelSeparation*e,t.options.fixed.x=!0}},{key:"shift",value:function(t,e){this.layout.body.nodes[t].y+=e}}]),i}(lv),uv=Gi.every,fv=Ao("every");gt({target:"Array",proto:!0,forced:!fv},{every:function(t){return uv(this,t,arguments.length>1?arguments[1]:void 0)}});var pv=Ht("Array").every,vv=Array.prototype,gv=function(t){var e=t.every;return t===vv||t instanceof Array&&e===vv.every?pv:e};function yv(t,e){var i=void 0!==ko&&Bi(t)||t["@@iterator"];if(!i){if(So(t)||(i=function(t,e){var i;if(!t)return;if("string"==typeof t)return mv(t,e);var n=Oo(i=Object.prototype.toString.call(t)).call(i,8,-1);"Object"===n&&t.constructor&&(n=t.constructor.name);if("Map"===n||"Set"===n)return Ei(t);if("Arguments"===n||/^(?:Ui|I)nt(?:8|16|32)(?:Clamped)?Array$/.test(n))return mv(t,e)}(t))||e&&t&&"number"==typeof t.length){i&&(t=i);var n=0,o=function(){};return{s:o,n:function(){return n>=t.length?{done:!0}:{done:!1,value:t[n++]}},e:function(t){throw t},f:o}}throw new TypeError("Invalid attempt to iterate non-iterable instance.\nIn order to be iterable, non-array objects must have a [Symbol.iterator]() method.")}var r,s=!0,a=!1;return{s:function(){i=i.call(t)},n:function(){var t=i.next();return s=t.done,t},e:function(t){a=!0,r=t},f:function(){try{s||null==i.return||i.return()}finally{if(a)throw r}}}}function mv(t,e){(null==e||e>t.length)&&(e=t.length);for(var i=0,n=new Array(e);i<e;i++)n[i]=t[i];return n}function bv(t,e){var i=new ec;return Wo(t).call(t,(function(t){var e;Wo(e=t.edges).call(e,(function(t){t.connected&&i.add(t)}))})),Wo(i).call(i,(function(t){var i=t.from.id,n=t.to.id;null==e[i]&&(e[i]=0),(null==e[n]||e[i]>=e[n])&&(e[n]=e[i]+1)})),e}function wv(t,e,i,n){var o,r,s=Gr(null),a=Kd(o=wo(Dc(n).call(n))).call(o,(function(t,e){return t+1+e.edges.length}),0),h=i+"Id",l="to"===i?1:-1,d=yv(n);try{var c=function(){var o=uo(r.value,2),d=o[0],c=o[1];if(!n.has(d)||!t(c))return"continue";s[d]=0;for(var u=[c],f=0,p=void 0,v=function(){var t,o;if(!n.has(d))return"continue";var r=s[p.id]+l;if(Wo(t=mr(o=p.edges).call(o,(function(t){return t.connected&&t.to!==t.from&&t[i]!==p&&n.has(t.toId)&&n.has(t.fromId)}))).call(t,(function(t){var n=t[h],o=s[n];(null==o||e(r,o))&&(s[n]=r,u.push(t[i]))})),f>a)return{v:{v:bv(n,s)}};++f};p=u.pop();){var g=v();if("continue"!==g&&"object"===go(g))return g.v}};for(d.s();!(r=d.n()).done;){var u=c();if("continue"!==u&&"object"===go(u))return u.v}}catch(t){d.e(t)}finally{d.f()}return s}var kv=function(){function t(){Nn(this,t),this.childrenReference={},this.parentReference={},this.trees={},this.distributionOrdering={},this.levels={},this.distributionIndex={},this.isTree=!1,this.treeIndex=-1}return Fn(t,[{key:"addRelation",value:function(t,e){void 0===this.childrenReference[t]&&(this.childrenReference[t]=[]),this.childrenReference[t].push(e),void 0===this.parentReference[e]&&(this.parentReference[e]=[]),this.parentReference[e].push(t)}},{key:"checkIfTree",value:function(){for(var t in this.parentReference)if(this.parentReference[t].length>1)return void(this.isTree=!1);this.isTree=!0}},{key:"numTrees",value:function(){return this.treeIndex+1}},{key:"setTreeIndex",value:function(t,e){void 0!==e&&void 0===this.trees[t.id]&&(this.trees[t.id]=e,this.treeIndex=Math.max(e,this.treeIndex))}},{key:"ensureLevel",value:function(t){void 0===this.levels[t]&&(this.levels[t]=0)}},{key:"getMaxLevel",value:function(t){var e=this,i={};return function t(n){if(void 0!==i[n])return i[n];var o=e.levels[n];if(e.childrenReference[n]){var r=e.childrenReference[n];if(r.length>0)for(var s=0;s<r.length;s++)o=Math.max(o,t(r[s]))}return i[n]=o,o}(t)}},{key:"levelDownstream",value:function(t,e){void 0===this.levels[e.id]&&(void 0===this.levels[t.id]&&(this.levels[t.id]=0),this.levels[e.id]=this.levels[t.id]+1)}},{key:"setMinLevelToZero",value:function(t){var e=1e9;for(var i in t)Object.prototype.hasOwnProperty.call(t,i)&&void 0!==this.levels[i]&&(e=Math.min(this.levels[i],e));for(var n in t)Object.prototype.hasOwnProperty.call(t,n)&&void 0!==this.levels[n]&&(this.levels[n]-=e)}},{key:"getTreeSize",value:function(t,e){var i=1e9,n=-1e9,o=1e9,r=-1e9;for(var s in this.trees)if(Object.prototype.hasOwnProperty.call(this.trees,s)&&this.trees[s]===e){var a=t[s];i=Math.min(a.x,i),n=Math.max(a.x,n),o=Math.min(a.y,o),r=Math.max(a.y,r)}return{min_x:i,max_x:n,min_y:o,max_y:r}}},{key:"hasSameParent",value:function(t,e){var i=this.parentReference[t.id],n=this.parentReference[e.id];if(void 0===i||void 0===n)return!1;for(var o=0;o<i.length;o++)for(var r=0;r<n.length;r++)if(i[o]==n[r])return!0;return!1}},{key:"inSameSubNetwork",value:function(t,e){return this.trees[t.id]===this.trees[e.id]}},{key:"getLevels",value:function(){return zo(this.distributionOrdering)}},{key:"addToOrdering",value:function(t,e){void 0===this.distributionOrdering[e]&&(this.distributionOrdering[e]=[]);var i=!1,n=this.distributionOrdering[e];for(var o in n)if(n[o]===t){i=!0;break}i||(this.distributionOrdering[e].push(t),this.distributionIndex[t.id]=this.distributionOrdering[e].length-1)}}]),t}(),_v=function(){function t(e){Nn(this,t),this.body=e,this._resetRNG(Math.random()+":"+No()),this.setPhysics=!1,this.options={},this.optionsBackup={physics:{}},this.defaultOptions={randomSeed:void 0,improvedLayout:!0,clusterThreshold:150,hierarchical:{enabled:!1,levelSeparation:150,nodeSpacing:100,treeSpacing:200,blockShifting:!0,edgeMinimization:!0,parentCentralization:!0,direction:"UD",sortMethod:"hubsize"}},At(this.options,this.defaultOptions),this.bindEventListeners()}return Fn(t,[{key:"bindEventListeners",value:function(){var t=this;this.body.emitter.on("_dataChanged",(function(){t.setupHierarchicalLayout()})),this.body.emitter.on("_dataLoaded",(function(){t.layoutNetwork()})),this.body.emitter.on("_resetHierarchicalLayout",(function(){t.setupHierarchicalLayout()})),this.body.emitter.on("_adjustEdgesForHierarchicalLayout",(function(){if(!0===t.options.hierarchical.enabled){var e=t.direction.curveType();t.body.emitter.emit("_forceDisableDynamicCurves",e,!1)}}))}},{key:"setOptions",value:function(t,e){if(void 0!==t){var i=this.options.hierarchical,n=i.enabled;if(Eh(["randomSeed","improvedLayout","clusterThreshold"],this.options,t),$h(this.options,t,"hierarchical"),void 0!==t.randomSeed&&this._resetRNG(t.randomSeed),!0===i.enabled)return!0===n&&this.body.emitter.emit("refresh",!0),"RL"===i.direction||"DU"===i.direction?i.levelSeparation>0&&(i.levelSeparation*=-1):i.levelSeparation<0&&(i.levelSeparation*=-1),this.setDirectionStrategy(),this.body.emitter.emit("_resetHierarchicalLayout"),this.adaptAllOptionsForHierarchicalLayout(e);if(!0===n)return this.body.emitter.emit("refresh"),Ch(e,this.optionsBackup)}return e}},{key:"_resetRNG",value:function(t){this.initialRandomSeed=t,this._rng=ah(this.initialRandomSeed)}},{key:"adaptAllOptionsForHierarchicalLayout",value:function(t){if(!0===this.options.hierarchical.enabled){var e=this.optionsBackup.physics;void 0===t.physics||!0===t.physics?(t.physics={enabled:void 0===e.enabled||e.enabled,solver:"hierarchicalRepulsion"},e.enabled=void 0===e.enabled||e.enabled,e.solver=e.solver||"barnesHut"):"object"===go(t.physics)?(e.enabled=void 0===t.physics.enabled||t.physics.enabled,e.solver=t.physics.solver||"barnesHut",t.physics.solver="hierarchicalRepulsion"):!1!==t.physics&&(e.solver="barnesHut",t.physics={solver:"hierarchicalRepulsion"});var i=this.direction.curveType();if(void 0===t.edges)this.optionsBackup.edges={smooth:{enabled:!0,type:"dynamic"}},t.edges={smooth:!1};else if(void 0===t.edges.smooth)this.optionsBackup.edges={smooth:{enabled:!0,type:"dynamic"}},t.edges.smooth=!1;else if("boolean"==typeof t.edges.smooth)this.optionsBackup.edges={smooth:t.edges.smooth},t.edges.smooth={enabled:t.edges.smooth,type:i};else{var n=t.edges.smooth;void 0!==n.type&&"dynamic"!==n.type&&(i=n.type),this.optionsBackup.edges={smooth:{enabled:void 0===n.enabled||n.enabled,type:void 0===n.type?"dynamic":n.type,roundness:void 0===n.roundness?.5:n.roundness,forceDirection:void 0!==n.forceDirection&&n.forceDirection}},t.edges.smooth={enabled:void 0===n.enabled||n.enabled,type:i,roundness:void 0===n.roundness?.5:n.roundness,forceDirection:void 0!==n.forceDirection&&n.forceDirection}}this.body.emitter.emit("_forceDisableDynamicCurves",i)}return t}},{key:"positionInitially",value:function(t){if(!0!==this.options.hierarchical.enabled){this._resetRNG(this.initialRandomSeed);for(var e=t.length+50,i=0;i<t.length;i++){var n=t[i],o=2*Math.PI*this._rng();void 0===n.x&&(n.x=e*Math.cos(o)),void 0===n.y&&(n.y=e*Math.sin(o))}}}},{key:"layoutNetwork",value:function(){if(!0!==this.options.hierarchical.enabled&&!0===this.options.improvedLayout){for(var t=this.body.nodeIndices,e=0,i=0;i<t.length;i++){!0===this.body.nodes[t[i]].predefinedPosition&&(e+=1)}if(e<.5*t.length){var n=0,o=this.options.clusterThreshold,r={clusterNodeProperties:{shape:"ellipse",label:"",group:"",font:{multi:!1}},clusterEdgeProperties:{label:"",font:{multi:!1},smooth:{enabled:!1}}};if(t.length>o){for(var s=t.length;t.length>o&&n<=10;){n+=1;var a=t.length;if(n%3==0?this.body.modules.clustering.clusterBridges(r):this.body.modules.clustering.clusterOutliers(r),a==t.length&&n%3!=0)return this._declusterAll(),this.body.emitter.emit("_layoutFailed"),void console.info("This network could not be positioned by this version of the improved layout algorithm. Please disable improvedLayout for better performance.")}this.body.modules.kamadaKawai.setOptions({springLength:Math.max(150,2*s)})}n>10&&console.info("The clustering didn't succeed within the amount of interations allowed, progressing with partial result."),this.body.modules.kamadaKawai.solve(t,this.body.edgeIndices,!0),this._shiftToCenter();for(var h=0;h<t.length;h++){var l=this.body.nodes[t[h]];!1===l.predefinedPosition&&(l.x+=70*(.5-this._rng()),l.y+=70*(.5-this._rng()))}this._declusterAll(),this.body.emitter.emit("_repositionBezierNodes")}}}},{key:"_shiftToCenter",value:function(){for(var t=pp.getRangeCore(this.body.nodes,this.body.nodeIndices),e=pp.findCenter(t),i=0;i<this.body.nodeIndices.length;i++){var n=this.body.nodes[this.body.nodeIndices[i]];n.x-=e.x,n.y-=e.y}}},{key:"_declusterAll",value:function(){for(var t=!0;!0===t;){t=!1;for(var e=0;e<this.body.nodeIndices.length;e++)!0===this.body.nodes[this.body.nodeIndices[e]].isCluster&&(t=!0,this.body.modules.clustering.openCluster(this.body.nodeIndices[e],{},!1));!0===t&&this.body.emitter.emit("_dataChanged")}}},{key:"getSeed",value:function(){return this.initialRandomSeed}},{key:"setupHierarchicalLayout",value:function(){if(!0===this.options.hierarchical.enabled&&this.body.nodeIndices.length>0){var t,e,i=!1,n=!1;for(e in this.lastNodeOnLevel={},this.hierarchical=new kv,this.body.nodes)Object.prototype.hasOwnProperty.call(this.body.nodes,e)&&(void 0!==(t=this.body.nodes[e]).options.level?(i=!0,this.hierarchical.levels[e]=t.options.level):n=!0);if(!0===n&&!0===i)throw new Error("To use the hierarchical layout, nodes require either no predefined levels or levels have to be defined for all nodes.");if(!0===n){var o=this.options.hierarchical.sortMethod;"hubsize"===o?this._determineLevelsByHubsize():"directed"===o?this._determineLevelsDirected():"custom"===o&&this._determineLevelsCustomCallback()}for(var r in this.body.nodes)Object.prototype.hasOwnProperty.call(this.body.nodes,r)&&this.hierarchical.ensureLevel(r);var s=this._getDistribution();this._generateMap(),this._placeNodesByHierarchy(s),this._condenseHierarchy(),this._shiftToCenter()}}},{key:"_condenseHierarchy",value:function(){var t=this,e=!1,i={},n=function(e,i){var n=t.hierarchical.trees;for(var o in n)Object.prototype.hasOwnProperty.call(n,o)&&n[o]===e&&t.direction.shift(o,i)},o=function(){for(var e=[],i=0;i<t.hierarchical.numTrees();i++)e.push(t.direction.getTreeSize(i));return e},r=function e(i,n){if(!n[i.id]&&(n[i.id]=!0,t.hierarchical.childrenReference[i.id])){var o=t.hierarchical.childrenReference[i.id];if(o.length>0)for(var r=0;r<o.length;r++)e(t.body.nodes[o[r]],n)}},s=function(e){var i=arguments.length>1&&void 0!==arguments[1]?arguments[1]:1e9,n=1e9,o=1e9,r=1e9,s=-1e9;for(var a in e)if(Object.prototype.hasOwnProperty.call(e,a)){var h=t.body.nodes[a],l=t.hierarchical.levels[h.id],d=t.direction.getPosition(h),c=t._getSpaceAroundNode(h,e),u=uo(c,2),f=u[0],p=u[1];n=Math.min(f,n),o=Math.min(p,o),l<=i&&(r=Math.min(d,r),s=Math.max(d,s))}return[r,s,n,o]},a=function(e,i){var n=t.hierarchical.getMaxLevel(e.id),o=t.hierarchical.getMaxLevel(i.id);return Math.min(n,o)},h=function(e,i,n){for(var o=t.hierarchical,r=0;r<i.length;r++){var s=i[r],a=o.distributionOrdering[s];if(a.length>1)for(var h=0;h<a.length-1;h++){var l=a[h],d=a[h+1];o.hasSameParent(l,d)&&o.inSameSubNetwork(l,d)&&e(l,d,n)}}},l=function(i,n){var o=arguments.length>2&&void 0!==arguments[2]&&arguments[2],h=t.direction.getPosition(i),l=t.direction.getPosition(n),d=Math.abs(l-h),c=t.options.hierarchical.nodeSpacing;if(d>c){var u={},f={};r(i,u),r(n,f);var p=a(i,n),v=s(u,p),g=s(f,p),y=v[1],m=g[0],b=g[2],w=Math.abs(y-m);if(w>c){var k=y-m+c;k<-b+c&&(k=-b+c),k<0&&(t._shiftBlock(n.id,k),e=!0,!0===o&&t._centerParent(n))}}},d=function(n,o){for(var a=o.id,h=o.edges,l=t.hierarchical.levels[o.id],d=t.options.hierarchical.levelSeparation*t.options.hierarchical.levelSeparation,c={},u=[],f=0;f<h.length;f++){var p=h[f];if(p.toId!=p.fromId){var v=p.toId==a?p.from:p.to;c[h[f].id]=v,t.hierarchical.levels[v.id]<l&&u.push(p)}}var g=function(e,i){for(var n=0,o=0;o<i.length;o++)if(void 0!==c[i[o].id]){var r=t.direction.getPosition(c[i[o].id])-e;n+=r/Math.sqrt(r*r+d)}return n},y=function(e,i){for(var n=0,o=0;o<i.length;o++)if(void 0!==c[i[o].id]){var r=t.direction.getPosition(c[i[o].id])-e;n-=d*Math.pow(r*r+d,-1.5)}return n},m=function(e,i){for(var n=t.direction.getPosition(o),r={},s=0;s<e;s++){var a=g(n,i),h=y(n,i);if(void 0!==r[n-=Math.max(-40,Math.min(40,Math.round(a/h)))])break;r[n]=s}return n},b=m(n,u);!function(n){var a=t.direction.getPosition(o);if(void 0===i[o.id]){var h={};r(o,h),i[o.id]=h}var l=s(i[o.id]),d=l[2],c=l[3],u=n-a,f=0;u>0?f=Math.min(u,c-t.options.hierarchical.nodeSpacing):u<0&&(f=-Math.min(-u,d-t.options.hierarchical.nodeSpacing)),0!=f&&(t._shiftBlock(o.id,f),e=!0)}(b),function(i){var n=t.direction.getPosition(o),r=t._getSpaceAroundNode(o),s=uo(r,2),a=s[0],h=s[1],l=i-n,d=n;l>0?d=Math.min(n+(h-t.options.hierarchical.nodeSpacing),i):l<0&&(d=Math.max(n-(a-t.options.hierarchical.nodeSpacing),i)),d!==n&&(t.direction.setPosition(o,d),e=!0)}(b=m(n,h))};!0===this.options.hierarchical.blockShifting&&(function(i){var n=t.hierarchical.getLevels();n=Xo(n).call(n);for(var o=0;o<i&&(e=!1,h(l,n,!0),!0===e);o++);}(5),function(){for(var e in t.body.nodes)Object.prototype.hasOwnProperty.call(t.body.nodes,e)&&t._centerParent(t.body.nodes[e])}()),!0===this.options.hierarchical.edgeMinimization&&function(i){var n=t.hierarchical.getLevels();n=Xo(n).call(n);for(var o=0;o<i;o++){e=!1;for(var r=0;r<n.length;r++)for(var s=n[r],a=t.hierarchical.distributionOrdering[s],h=0;h<a.length;h++)d(1e3,a[h]);if(!0!==e)break}}(20),!0===this.options.hierarchical.parentCentralization&&function(){var e=t.hierarchical.getLevels();e=Xo(e).call(e);for(var i=0;i<e.length;i++)for(var n=e[i],o=t.hierarchical.distributionOrdering[n],r=0;r<o.length;r++)t._centerParent(o[r])}(),function(){for(var e=o(),i=0,r=0;r<e.length-1;r++){i+=e[r].max-e[r+1].min+t.options.hierarchical.treeSpacing,n(r+1,i)}}()}},{key:"_getSpaceAroundNode",value:function(t,e){var i=!0;void 0===e&&(i=!1);var n=this.hierarchical.levels[t.id];if(void 0!==n){var o=this.hierarchical.distributionIndex[t.id],r=this.direction.getPosition(t),s=this.hierarchical.distributionOrdering[n],a=1e9,h=1e9;if(0!==o){var l=s[o-1];if(!0===i&&void 0===e[l.id]||!1===i)a=r-this.direction.getPosition(l)}if(o!=s.length-1){var d=s[o+1];if(!0===i&&void 0===e[d.id]||!1===i){var c=this.direction.getPosition(d);h=Math.min(h,c-r)}}return[a,h]}return[0,0]}},{key:"_centerParent"
```
