## Supplementary material for "Evaluating Regional Diversity in Scientific Communication: A Comparative Analysis of COVID-19 Preprints and Peer-Reviewed Publications": visNetwork.js

```
// Production steps of ECMA-262, Edition 6, 22.1.2.1
// Référence : https://people.mozilla.org/~jorendorff/es6-draft.html#sec-array.from
if (!Array.from) {
  Array.from = (function () {
    var toStr = Object.prototype.toString;
    var isCallable = function (fn) { 
      return typeof fn === 'function' || toStr.call(fn) === '[object Function]';
    };
    var toInteger = function (value) { 
      var number = Number(value); 
      if (isNaN(number)) { return 0; }
      if (number === 0 || !isFinite(number)) { return number; }
      return (number > 0 ? 1 : -1) * Math.floor(Math.abs(number)); 
    };
    var maxSafeInteger = Math.pow(2, 53) - 1;
    var toLength = function (value) { 
      var len = toInteger(value);
      return Math.min(Math.max(len, 0), maxSafeInteger);
    }; 
  
    // La propriété length de la méthode vaut 1.
    return function from(arrayLike/*, mapFn, thisArg */) { 
      // 1. Soit C, la valeur this
      var C = this;
      
      // 2. Soit items le ToObject(arrayLike).
      var items = Object(arrayLike); 
      
      // 3. ReturnIfAbrupt(items).
      if (arrayLike == null) { 
        throw new TypeError("Array.from doit utiliser un objet semblable à un tableau - null ou undefined ne peuvent pas être utilisés");
      }
    
      // 4. Si mapfn est undefined, le mapping sera false.
      var mapFn = arguments.length > 1 ? arguments[1] : void undefined;
      var T;
      if (typeof mapFn !== 'undefined') {  
        // 5. sinon      
        // 5. a. si IsCallable(mapfn) est false, on lève une TypeError.
        if (!isCallable(mapFn)) { 
          throw new TypeError('Array.from: lorsqu il est utilisé le deuxième argument doit être une fonction'); 
        }
     
        // 5. b. si thisArg a été fourni, T sera thisArg ; sinon T sera undefined.
        if (arguments.length > 2) { 
          T = arguments[2];
        }
      }
    
      // 10. Soit lenValue pour Get(items, "length").
      // 11. Soit len pour ToLength(lenValue).
      var len = toLength(items.length);  
     
      // 13. Si IsConstructor(C) vaut true, alors
      // 13. a. Soit A le résultat de l'appel à la méthode interne [[Construct]] avec une liste en argument qui contient l'élément len.
      // 14. a. Sinon, soit A le résultat de ArrayCreate(len).
      var A = isCallable(C) ? Object(new C(len)) : new Array(len);
   
      // 16. Soit k égal à 0.
      var k = 0;  // 17. On répète tant que k < len… 
      var kValue;
      while (k < len) {
        kValue = items[k]; 
        if (mapFn) {
          A[k] = typeof T === 'undefined' ? mapFn(kValue, k) : mapFn.call(T, kValue, k); 
        } else {
          A[k] = kValue;
        }
        k += 1;
      }
      // 18. Soit putStatus égal à Put(A, "length", len, true).
      A.length = len;  // 20. On renvoie A.
      return A;
    };
  }());
};


// https://tc39.github.io/ecma262/#sec-array.prototype.includes
if (!Array.prototype.includes) {
  Object.defineProperty(Array.prototype, 'includes', {
    value: function(searchElement, fromIndex) {

      if (this == null) {
        throw new TypeError('"this" est nul ou non défini');
      }

      // 1. Soit o égal à ? Object(cette valeur).
      var o = Object(this);

      // 2. Soit len égal à ? Length(? Get(o, "length")).
      var len = o.length >>> 0;

      // 3. Si len = 0, renvoyer "false".
      if (len === 0) {
        return false;
      }

      // 4. Soit n = ? ToInteger(fromIndex).
      // Pour la cohérence du code, on gardera le nom anglais "fromIndex" pour la variable auparavant appelée "indiceDépart"
      //    (Si fromIndex n'est pas défini, cette étape produit la valeur 0.)
      var n = fromIndex | 0;

      // 5. Si n ≥ 0,
      //  a. Alors k = n.
      // 6. Sinon, si n < 0,
      //  a. Alors k = len + n.
      //  b. Si k < 0, alors k = 0.
      var k = Math.max(n >= 0 ? n : len - Math.abs(n), 0);

      function sameValueZero(x, y) {
        return x === y || (typeof x === 'number' && typeof y === 'number' && isNaN(x) && isNaN(y));
      }

      // 7. Répéter tant que k < len
      while (k < len) {
        // a. Soit elementK le résultat de ? Get(O, ! ToString(k)).
        // b. Si SameValueZero(searchElement, elementK) est vrai, renvoyer "true".
        if (sameValueZero(o[k], searchElement)) {
          return true;
        }
        // c. Augmenter la valeur de k de 1. 
        k++;
      }

      // 8. Renvoyer "false"
      return false;
    }
  });
}

// Add shim for Function.prototype.bind() from:
// https://developer.mozilla.org/en-US/docs/JavaScript/Reference/Global_Objects/Function/bind#Compatibility
// for fix some RStudio viewer bug (Desktop / windows)
if (!Function.prototype.bind) {
  Function.prototype.bind = function (oThis) {
    if (typeof this !== "function") {
      // closest thing possible to the ECMAScript 5 internal IsCallable function
      throw new TypeError("Function.prototype.bind - what is trying to be bound is not callable");
    }
    
    var aArgs = Array.prototype.slice.call(arguments, 1),
    fToBind = this,
    fNOP = function () {},
    fBound = function () {
      return fToBind.apply(this instanceof fNOP && oThis
                           ? this
                           : oThis,
                           aArgs.concat(Array.prototype.slice.call(arguments)));
    };
    
    fNOP.prototype = this.prototype;
    fBound.prototype = new fNOP();
    
    return fBound;
  };
}

//--------------------------------------------
// functions to reset edges after hard to read
//--------------------------------------------

// for edges
function edgeAsHardToRead(edge, hideColor1, hideColor2, network, type){
  //console.info("edgeAsHardToRead")
  
  if(type === "edge"){
    //console.info("edge")
    //console.info(edge.id)
    
    // saving color information (if we have)
    if (edge.hiddenColor === undefined && edge.color !== hideColor1 && edge.color !== hideColor2) {
      edge.hiddenColor = edge.color;
    }
    // set "hard to read" color
    edge.color = hideColor1;
    
    // reset and save label
    if (edge.hiddenLabel === undefined) {
      edge.hiddenLabel = edge.label;
      edge.label = undefined;
    }
    edge.isHardToRead = true;
  } else {
    //console.info("cluster")
    //console.info(edge.id)
    //console.info(edge)
    // saving color information (if we have)
    if (edge.hiddenColor === undefined && edge.color !== hideColor1 && edge.color !== hideColor2) {
      //network.clustering.updateEdge(edge.id, {hiddenColor : edge.color});
      edge.hiddenColor = edge.color;
    }
    // set "hard to read" color
    edge.color = hideColor1;
    //network.clustering.updateEdge(edge.id, {color : hideColor1});
    //edge.color = hideColor1;
    // reset and save label
    if (edge.hiddenLabel === undefined) {
      edge.hiddenLabel = edge.label;
      edge.label = undefined;
    }
    edge.isHardToRead = true;
  }

}

function resetOneEdge(edge, hideColor1, hideColor2, type){
  
  /*console.info("resetOneEdge")
  console.info(type)
  console.info(edge.id) 
  console.info(edge)
  console.info("edge.hiddenColor")
  console.info(edge.hiddenColor)*/
  
  var treat_egde = false;
  if(type === "cluster"){
    if(edge.isHardToRead !== undefined){ // we have to reset this node
      if(edge.isHardToRead){
        treat_egde = true;
      } else if(edge.isHardToRead === false && (edge.color.color === hideColor1 || edge.color.color === hideColor2)){
        treat_egde = true;
      }
    } else if(edge.color.color === hideColor1 || edge.color.color === hideColor2){
      treat_egde = true;
    }
    
    if(treat_egde){
      // get back color
      if (edge.hiddenColor !== undefined) {
        edge.color = edge.hiddenColor;
        edge.hiddenColor = undefined;
      }else{
        delete edge.color;
      }
        
      // finally, get back label
      if (edge.hiddenLabel !== undefined) {
        edge.label = edge.hiddenLabel;
        edge.hiddenLabel = undefined;
      }
      edge.isHardToRead = false;
    }
  } else {
    // get back color
    if (edge.hiddenColor !== undefined) {
      edge.color = edge.hiddenColor;
      edge.hiddenColor = undefined;
    }else{
      edge.color = null;
    }
    
    // finally, get back label
    if (edge.hiddenLabel !== undefined) {
      edge.label = edge.hiddenLabel;
      edge.hiddenLabel = undefined;
    }
    edge.isHardToRead = false;
  }
}

function resetAllEdges(edges, hideColor1, hideColor2, network){
  
  var edgesToReset = edges.get({
    fields: ['id', 'color', 'hiddenColor', 'label', 'hiddenLabel'],
    filter: function (item) {
      return item.isHardToRead === true;
    },
    returnType :'Array'
  });

  var is_cluster_edges = false;
  var edges_in_clusters;
  if(network !== undefined){
    edges_in_clusters = network.body.modules.clustering.clusteredEdges;
    if(Object.keys(edges_in_clusters).length > 0){
      is_cluster_edges = true;
      edges_in_clusters = Object.keys(edges_in_clusters);
    } else {
      edges_in_clusters = [];
    }
  }
  
  var treat_edges_in_clusters = [];
  // all edges get their own color and their label back
  for (var i = 0; i < edgesToReset.length; i++) {
    resetOneEdge(edgesToReset[i], hideColor1, hideColor2,type = "edge");
    if(is_cluster_edges){
      if(indexOf.call(edges_in_clusters, edgesToReset[i].id, true) > -1){
        var tmp_cluster_id = network.clustering.getClusteredEdges(edgesToReset[i].id);
        if(tmp_cluster_id.length > 1){
          tmp_cluster_id = tmp_cluster_id[0];
          treat_edges_in_clusters.push(tmp_cluster_id);
          resetOneEdge(network.body.edges[tmp_cluster_id].options, hideColor1, hideColor2, type = "cluster");
        }
      }
    }
  }
  
  // some misunderstood bug on some cluster edges... so have a (bad) fix...
  var edges_in_clusters_ctrl = edges_in_clusters.filter(function(word,index){
    if(word.match(/^clusterEdge/i)){
      if(indexOf.call(treat_edges_in_clusters, word, true) === -1){
        return true;
      } else {
        return false;
      }
        
    }else{
        return false;
    }
  });
  
  if(is_cluster_edges){
    if(edges_in_clusters_ctrl.length > 0){
       for (var j = 0; j < edges_in_clusters_ctrl.length; j++) {
         if(network.body.edges[edges_in_clusters_ctrl[j]] !== undefined){
           resetOneEdge(network.body.edges[edges_in_clusters_ctrl[j]].options, hideColor1, hideColor2, type = "cluster");
         }
        }
    }
  }

  edges.update(edgesToReset);
}

//--------------------------------------------
// functions to reset nodes after hard to read
//--------------------------------------------

// for classic node
function simpleResetNode(node, type){
  if(type === "node"){
    // get back color
    if (node.hiddenColor !== undefined) {
      node.color = node.hiddenColor;
      node.hiddenColor = undefined;
    }else{
      if(node.group !== undefined){
        node.color = undefined;
      } else {
        node.color = null;
      }
    }
  } else {
    if (Object.keys(node.options.hiddenColor).length > 2){
      node.setOptions({color : node.options.hiddenColor, hiddenColor : undefined});
    }else{
      if(node.options.group !== undefined){
        node.setOptions({color : undefined});
      } else {
        node.setOptions({color : null});
      }
    }
  }
}

// for icon node
function simpleIconResetNode(node, type){
  if(type === "node"){  
    // icon color
    node.icon.color = node.hiddenColor;
    node.hiddenColor = undefined;
    // get back color
    if (node.hiddenColorForLabel !== undefined) {
      node.color = node.hiddenColorForLabel;
      node.hiddenColorForLabel = undefined;
    }else{
      if(node.group !== undefined){
        node.color = undefined;
      } else {
        node.color = null;
      }
    }
  } else {
    node.setOptions({icon : { color : node.options.hiddenColor}, hiddenColor : undefined});
    if (node.options.hiddenColorForLabel !== undefined) {
      node.setOptions({color : node.options.hiddenColorForLabel, hiddenColorForLabel : undefined});
    }else{
      if(node.options.group !== undefined){
        node.setOptions({color : undefined});
      } else {
        node.setOptions({color : null});
      }
    }
  }
}

// for image node
function simpleImageResetNode(node, imageType, type){
  if(type === "node"){  
    // get back color
    if (node.hiddenColor !== undefined) {
      node.color = node.hiddenColor;
      node.hiddenColor = undefined;
    }else{
      if(node.group !== undefined){
        node.color = undefined;
      } else {
        node.color = null;
      }
    }
    // and set shape as image/circularImage
    node.shape = imageType;
  } else {
    if (Object.keys(node.options.hiddenColor).length > 2) {
      node.setOptions({color : node.options.hiddenColor, hiddenColor : undefined});
    }else{
      if(node.options.group !== undefined){
        node.setOptions({color : undefined});
      } else {
        node.setOptions({color : null});
      }
    }
    node.setOptions({shape : imageType});
  }
}

// Global function to reset one cluster
function resetOneCluster(node, options, network){
  if(node !== undefined){
    if(node.options.isHardToRead !== undefined){ // we have to reset this node
      if(node.options.isHardToRead){
        var final_shape;
        var shape_group = false;
        var is_group = false;
  	  // have a group information & a shape defined in group ?
        if(node.options.group !== undefined && options.groups !== undefined){
          if(options.groups[node.options.group] !== undefined){
            is_group = true;
            if(options.groups[node.options.group].shape !== undefined){
              shape_group = true;
            }
          }
        }
        // have a global shape in nodes options ?
        var shape_options = false;
        if(options.nodes !== undefined){
          if(options.nodes.shape !== undefined){
            shape_options = true;
          }
        }
        // set final shape (individual > group > global)
        if(node.options.hiddenImage !== undefined){
          final_shape = node.options.hiddenImage;
        } else if(node.options.shape !== undefined){
          final_shape = node.options.shape;
        } else if(shape_group){
          final_shape = options.groups[node.options.group].shape;
        } else if(shape_options){
          final_shape = options.nodes.shape;
        }
        
        node.setOptions({bodyHiddenColor : network.body.nodes[node.id].options.color});
        // and call good reset function
        if(final_shape === "icon"){
          simpleIconResetNode(node, "cluster");
        } else if(final_shape === "image"){
          simpleImageResetNode(node, "image", "cluster");
        } else if(final_shape === "circularImage"){
          simpleImageResetNode(node, "circularImage", "cluster");
        } else {
          simpleResetNode(node, "cluster");
        }
    	 // finally, get back label
        if (node.options.hiddenLabel !== undefined) {
          node.setOptions({label : node.options.hiddenLabel, hiddenLabel : undefined});
        }
        node.options.isHardToRead = false;
      }
    }
  }
}

// Global function to reset one node
function resetOneNode(node, options, network){
  if(node !== undefined){
    if(node.isHardToRead !== undefined){ // we have to reset this node
      if(node.isHardToRead){
        var final_shape;
        var shape_group = false;
        var is_group = false;
  	  // have a group information & a shape defined in group ?
        if(node.group !== undefined && options.groups !== undefined){
          if(options.groups[node.group] !== undefined){
            is_group = true;
            if(options.groups[node.group].shape !== undefined){
              shape_group = true;
            }
          }
        }
        // have a global shape in nodes options ?
        var shape_options = false;
        if(options.nodes !== undefined){
          if(options.nodes.shape !== undefined){
            shape_options = true;
          }
        }
        // set final shape (individual > group > global)
        if(node.hiddenImage !== undefined){
          final_shape = node.hiddenImage;
        } else if(node.shape !== undefined){
          final_shape = node.shape;
        } else if(shape_group){
          final_shape = options.groups[node.group].shape;
        } else if(shape_options){
          final_shape = options.nodes.shape;
        }
        
        // reset body information
        network.body.nodes[node.id].options.color = node.bodyHiddenColor;

        // and call good reset function
        if(final_shape === "icon"){
          simpleIconResetNode(node, "node");
        } else if(final_shape === "image"){
          simpleImageResetNode(node, "image", "node");
        } else if(final_shape === "circularImage"){
          simpleImageResetNode(node, "circularImage", "node");
        } else {
          simpleResetNode(node, "node");
        }
    	 // finally, get back label
    	  if (node.hiddenLabel !== undefined) {
          node.label = node.hiddenLabel;
          node.hiddenLabel = undefined;
        }
        node.isHardToRead = false;
      }
    }
  }
}

// Global function to reset all node
function resetAllNodes(nodes, update, options, network, all){
  
  if(all === false){
      var nodesToReset = nodes.get({
      filter: function (item) {
        return item.isHardToRead === true;
      },
      returnType :'Array'
    });
  } else {
      var nodesToReset = nodes.get({returnType :'Array'});
  }
  
  var have_cluster_nodes = false;
  var nodes_in_clusters;
  if(network !== undefined){
    nodes_in_clusters = network.body.modules.clustering.clusteredNodes;
    if(Object.keys(nodes_in_clusters).length > 0){
      have_cluster_nodes = true;
      nodes_in_clusters = Object.keys(nodes_in_clusters);
    } else {
      nodes_in_clusters = [];
    }
  }

  for (var i = 0; i < nodesToReset.length; i++) {
    resetOneNode(nodesToReset[i], options, network, type = "node");
	// reset coordinates
    nodesToReset[i].x = undefined;
    nodesToReset[i].y = undefined;
    if(have_cluster_nodes){
      if(indexOf.call(nodes_in_clusters, nodesToReset[i].id, true) > -1){
        var tmp_cluster_id = network.clustering.findNode(nodesToReset[i].id);
        // in case of multiple cluster...
        for(var j = 0; j < (tmp_cluster_id.length-1); j++) {
          resetOneCluster(network.body.nodes[tmp_cluster_id[j]], options, network);
        }
      }
    }
  }
  if(update){
    nodes.update(nodesToReset);
  }
}

//--------------------------------------------
// functions to set nodes as hard to read
//--------------------------------------------

// for classic node
function simpleNodeAsHardToRead(node, hideColor1, hideColor2, type){


  // classic nodes
  if(type === "node"){
    // saving color information (if we have)
    if (node.hiddenColor === undefined && node.color !== hideColor1 && node.color !== hideColor2) {
      node.hiddenColor = node.color;
    }
  
    // set "hard to read" color
    node.color = hideColor1;
    // reset and save label
    if (node.hiddenLabel === undefined) {
      node.hiddenLabel = node.label;
      node.label = undefined;
    }
  // cluster  
  } else {
    // saving color information (if we have)
    if (node.options.hiddenColor === undefined && node.options.color !== hideColor1 && node.options.color !== hideColor2) {
      node.setOptions({hiddenColor : node.options.color});
    }
    // set "hard to read" color
    node.setOptions({color : hideColor1});
    // reset and save label
    if (node.options.hiddenLabel === undefined) {
      node.setOptions({hiddenLabel : node.options.label});
      node.setOptions({label : undefined});
    }
  }
}

// for icon node
function iconsNodeAsHardToRead(node, hideColor1, hideColor2, icon_color, type){
  // classic nodes
  if(type === "node"){
    // individual information
    if(node.icon !== undefined && node.icon !== null && node.icon !== {}){
      node.iconDefined = true;
    } else { // information in group : have to as individual
      node.icon = {};
      node.iconDefined = false;
    }
    // set "hard to read" color
    node.icon.color = hideColor1;
    node.hiddenColor = icon_color;
    // for edges....saving color information (if we have)
    if (node.hiddenColorForLabel === undefined && node.color !== hideColor1 && node.color !== hideColor2) {
      node.hiddenColorForLabel = node.color;
    }
    // set "hard to read" color
    node.color = hideColor1;
    // reset and save label
    if (node.hiddenLabel === undefined) {
      node.hiddenLabel = node.label;
      node.label = undefined;
    }
  } else {
    // individual information
    if(node.options.icon !== undefined && node.options.icon !== null && node.options.icon !== {}){
      node.setOptions({iconDefined : true});
    } else { // information in group : have to as individual
      node.setOptions({iconDefined : false, icon:{}});
    }
    // set "hard to read" color
    node.setOptions({hiddenColor : icon_color, icon:{color : hideColor1}});
    // for edges....saving color information (if we have)
    if (node.options.hiddenColorForLabel === undefined && node.options.color !== hideColor1 && node.options.color !== hideColor2) {
      node.setOptions({hiddenColorForLabel : node.options.color});
    }
    // set "hard to read" color
    node.setOptions({color : hideColor1});
    // reset and save label
    if (node.options.hiddenLabel === undefined) {
      node.setOptions({hiddenLabel : node.options.label, label : undefined});
    }
  }
}

// for image node
function imageNodeAsHardToRead(node, imageType, hideColor1, hideColor2, type){
  // classic nodes
  if(type === "node"){
    // saving color information (if we have)
    if (node.hiddenColor === undefined && node.color !== hideColor1 && node.color !== hideColor2) {
      node.hiddenColor = node.color;
    }
    // set "hard to read" color
    node.color = hideColor1;
    // reset and save label
    if (node.hiddenLabel === undefined) {
      node.hiddenLabel = node.label;
      node.label = undefined;
    }
    // keep shape information, and set a new
    if(imageType === "image"){
      node.hiddenImage = imageType;
      node.shape = "square";
    }else if(imageType === "circularImage"){
      node.hiddenImage = imageType;
      node.shape = "dot";
    }
  } else {
    // saving color information (if we have)
    if (node.options.hiddenColor === undefined && node.options.color !== hideColor1 && node.options.color !== hideColor2) {
      node.setOptions({hiddenColor : node.options.color});
    }
    // set "hard to read" color
    node.setOptions({color : hideColor1});
    // reset and save label
    if (node.options.hiddenLabel === undefined) {
      node.setOptions({hiddenLabel : node.options.label, label : undefined});
    }
    if(imageType === "image"){
      node.setOptions({hiddenImage : "image", shape : "square"});
    } else if(imageType === "circularImage"){
      node.setOptions({hiddenImage : "circularImage", shape : "dot"});
    }
    node.hiddenImage = imageType;
  }
}

// Global function to set one node as hard to read
function nodeAsHardToRead(node, options, hideColor1, hideColor2, network, type){
  var final_shape;
  var shape_group = false;
  var is_group = false;


  if(node.isHardToRead === false || node.isHardToRead === undefined){

    // have a group information & a shape defined in group ?
    if(node.group !== undefined && options.groups !== undefined){
      if(options.groups[node.group] !== undefined){
        is_group = true;
        if(options.groups[node.group].shape !== undefined){
          shape_group = true;
        }
      }
    }
    // have a group information & a shape defined in group ?
    var shape_options = false;
    if(options.nodes !== undefined){
      if(options.nodes.shape !== undefined){
        shape_options = true;
      }
    }
    // set final shape (individual > group > global)
    if(node.shape !== undefined){
      final_shape = node.shape;
    } else if(shape_group){
      final_shape = options.groups[node.group].shape;
    } else if(shape_options){
      final_shape = options.nodes.shape;
    }
    
    // information save in body nodes
    if(type === "node"){
      node.bodyHiddenColor = clone(network.body.nodes[node.id].options.color);
    } else {
      node.setOptions({bodyHiddenColor : clone(network.body.nodes[node.id].options.color)});
    }
  
    // and call good function
    if(final_shape === "icon"){
      // find color for icon
      var icon_color = "#2B7CE9";
      var find_color = false;
      // in nodes ?
      if(node.icon !== undefined){
        if(node.icon.color !== undefined){
          icon_color = node.icon.color;
          find_color = true;
        }
      } 
      // or in group ?
      if(find_color === false && is_group && options.groups !== undefined && options.groups[node.group].icon !== undefined){
        if(options.groups[node.group].icon.color !== undefined){
          icon_color = options.groups[node.group].icon.color;
          find_color = true;
        }
      }
      // in global node ?
      if(find_color === false && options.nodes.icon !== undefined){
        if(options.nodes.icon.color !== undefined){
          icon_color = options.nodes.icon.color;
        }
      } 
      iconsNodeAsHardToRead(node, hideColor1, hideColor2, icon_color, type);
    } else if(final_shape === "image"){
      imageNodeAsHardToRead(node, "image", hideColor1, hideColor2, type);
    } else if(final_shape === "circularImage"){
      imageNodeAsHardToRead(node, "circularImage", hideColor1, hideColor2, type);
    } else {
      simpleNodeAsHardToRead(node, hideColor1, hideColor2, type);
    }
    
    // finally set isHardToRead
    if(type === "node"){
      node.isHardToRead = true;
    } else {
      node.setOptions({isHardToRead : true});
    }
  // special case of just to label  
  } else if(node.isHardToRead === true && node.label !== undefined){
    if(type === "node"){
      node.hiddenLabel = node.label;
      node.label = undefined;
    } else {
      node.setOptions({hiddenLabel : node.options.label, label : undefined})
    }
  }
}

//----------------------------------------------------------------
// Revrite HTMLWidgets.dataframeToD3() for passing custom
// properties directly in data.frame (color.background) for example
//----------------------------------------------------------------
function visNetworkdataframeToD3(df, type) {

  // variables we have specially to control
  /*var nodesctrl = ["color", "fixed", "font", "icon", "shadow", "scaling", "shapeProperties", "chosen", "heightConstraint", "image", "margin", "widthConstraint"];
  var edgesctrl = ["color", "font", "arrows", "shadow", "smooth", "scaling", "chosen", "widthConstraint"];*/
  
  var names = [];
  var colnames = [];
  var length;
  var toctrl;
  var ctrlname;
  
  for (var name in df) {
    if (df.hasOwnProperty(name))
      colnames.push(name);
      ctrlname = name.split(".");
      if(ctrlname.length === 1){
        names.push( new Array(name));
      } else {
        /*if(type === "nodes"){
         toctrl = indexOf.call(nodesctrl, ctrlname[0], true);
        } else if(type === "edges"){
         toctrl = indexOf.call(edgesctrl, ctrlname[0], true);
        }
        if(toctrl > -1){*/
          names.push(ctrlname);
        /*} else {
          names.push(new Array(name));
        }*/
      }
      if (typeof(df[name]) !== "object" || typeof(df[name].length) === "undefined") {
          throw new Error("All fields must be arrays");
      } else if (typeof(length) !== "undefined" && length !== df[name].length) {
          throw new Error("All fields must be arrays of the same length");
      }
      length = df[name].length;
  }

  var results = [];
  var item;
    for (var row = 0; row < length; row++) {
      item = {};
      for (var col = 0; col < names.length; col++) {
        if(df[colnames[col]][row] !== null){
          if(names[col].length === 1){
            if(names[col][0] === "dashes"){
              item[names[col]] = eval(df[colnames[col]][row]);
            } else {
              item[names[col]] = df[colnames[col]][row];
            }
          } else if(names[col].length === 2){
            if(item[names[col][0]] === undefined){
              item[names[col][0]] = {};
            }
            if(names[col][0] === "icon" && names[col][1] === "code"){
              item[names[col][0]][names[col][1]] = JSON.parse( '"'+'\\u' + df[colnames[col]][row] + '"');
            } else if(names[col][0] === "icon" && names[col][1] === "color"){
              item.color = df[colnames[col]][row];
              item[names[col][0]][names[col][1]] = df[colnames[col]][row];
            } else if(names[col][0] === "icon" && names[col][1] === "face"){
              if(df[colnames[col]][row] === "'Font Awesome 5 Free'"){
                item.icon.weight = "bold";
              }
              item[names[col][0]][names[col][1]] = df[colnames[col]][row];
            } else{
              item[names[col][0]][names[col][1]] = df[colnames[col]][row];
            }
          } else if(names[col].length === 3){
            if(item[names[col][0]] === undefined){
              item[names[col][0]] = {};
            }
            if(item[names[col][0]][names[col][1]] === undefined){
              item[names[col][0]][names[col][1]] = {};
            }
            item[names[col][0]][names[col][1]][names[col][2]] = df[colnames[col]][row];
          } else if(names[col].length === 4){
            if(item[names[col][0]] === undefined){
              item[names[col][0]] = {};
            }
            if(item[names[col][0]][names[col][1]] === undefined){
              item[names[col][0]][names[col][1]] = {};
            }
            if(item[names[col][0]][names[col][1]][names[col][2]] === undefined){
              item[names[col][0]][names[col][1]][names[col][2]] = {};
            }
            item[names[col][0]][names[col][1]][names[col][2]][names[col][3]] = df[colnames[col]][row];
          }
        }
      }
      results.push(item);
    }
  return results;
}

//----------------------------------------------------------------
// Some utils functions
//---------------------------------------------------------------- 

//unique element in array
function uniqueArray(arr, exclude_cluster, network) {
  var a = [];
  for (var i=0, l=arr.length; i<l; i++){
    if (a.indexOf(arr[i]) === -1 && arr[i] !== ''){
      if(exclude_cluster === false){
        a.push(arr[i]);
      } else if(network.isCluster(arr[i]) === false){
        a.push(arr[i]);
      }
    }
  }

  return a;
}

function uniqueShiny(arr) {
  return arr.filter(function (value, index, self) { 
    return self.indexOf(value) === index;
  });
};
// clone an object
function clone(obj) {
    if(obj === null || typeof(obj) != 'object')
        return obj;    
    var temp = new obj.constructor(); 
    for(var key in obj)
        temp[key] = clone(obj[key]);    
    return temp;
}
// update a list
function update(source, target) {
	Object.keys(target).forEach(function (k) {
		if (typeof target[k] === 'object' && k !== "container") {
			source[k] = source[k] || {};
			update(source[k], target[k]);
		} else {
			source[k] = target[k];
		}
	});
}
// for find element
function indexOf(needle, str) {
        indexOf = function(needle, str) {
            var i = -1, index = -1;
            if(str){
                  needle = ''+needle;
            }
            for(i = 0; i < this.length; i++) {
                var val = this[i];
                if(str){
                  val = ''+val;
                }
                if(val === needle) {
                    index = i;
                    break;
                }
            }
            return index;
        };
    return indexOf.call(this, needle, str);
};
// reset a html list
function resetList(list_name, id, shiny_input_name) {
  var list = document.getElementById(list_name + id);
  list.value = "";
  if (window.Shiny){
    Shiny.onInputChange(id + '_' + shiny_input_name, "");
  }
}
// id node list selection init
function setNodeIdList(selectList, params, nodes){
  if(params.style !== undefined){
    selectList.setAttribute('style', params.style);
  }
  selectList.style.display = 'inline';
      
  option = document.createElement("option");
  option.value = "";
  if(params.main === undefined){
    option.text = "Select by id";
  } else {
    option.text = params.main;
  }
  
  selectList.appendChild(option);
      
  // have to set for all nodes ?
  if(params.values === undefined){
    var info_node_list = nodes.get({
      fields: ['id', 'label'],
      returnType :'Array'
    });
    for (var i = 0; i < info_node_list.length; i++) {
      option = document.createElement("option");
      option.value = info_node_list[i].id;
      if(info_node_list[i].label && params.useLabels){
        option.text = info_node_list[i].label;
      }else{
        option.text = info_node_list[i].id;
      }
      selectList.appendChild(option);
    }
  } else {
    var tmp_node;
    for(var tmp_id = 0 ; tmp_id < params.values.length; tmp_id++){
      tmp_node = nodes.get({
        fields: ['id', 'label'],
        filter: function (item) {
          return (item.id === params.values[tmp_id]) ;
        },
        returnType :'Array'
      });
      if(tmp_node !== undefined){
        option = document.createElement("option");
        option.value = tmp_node[0].id;
        if(tmp_node[0].label && params.useLabels){
          option.text = tmp_node[0].label;
        }else{
          option.text = tmp_node[0].id;
        }
        selectList.appendChild(option);
      }
    }
  }
}

//----------------------------------------------------------------
// Collapsed function
//---------------------------------------------------------------- 

function networkOpenCluster(params){
  if (params.nodes.length === 1) {
    if (this.isCluster(params.nodes[0]) === true) {
      var elid = this.body.container.id.substring(5);
      var fit = document.getElementById(elid).collapseFit;
      var resetHighlight = document.getElementById(elid).collapseResetHighlight;
      
      if(document.getElementById(elid).collapseKeepCoord){
        this.openCluster(params.nodes[0], 
        {releaseFunction : function(clusterPosition, containedNodesPositions) {
              return containedNodesPositions;
            }
        });
      } else {
        this.openCluster(params.nodes[0]);
      }

      
      if(resetHighlight){
        document.getElementById("nodeSelect"+elid).value = "";
        document.getElementById("nodeSelect"+elid).onchange();
      }
      if(fit){
        this.fit();
      }
    }
  }
}

function collapsedNetwork(nodes, fit, resetHighlight, clusterParams, labelSuffix, treeParams, network, elid) {
  
  var set_position = true;
  var selectedNode;
  var j;
  
  if(nodes[0] !== undefined){
    
    for (var inodes = 0; inodes < nodes.length; inodes++) {
      
      selectedNode = nodes[inodes];
      if(selectedNode !== undefined){
        if(network.isCluster(selectedNode)){
          //network.openCluster(selectedNode)
          /*instance.network.openCluster(selectedNode, 
          {releaseFunction : function(clusterPosition, containedNodesPositions) {
            return tmp_position;
          }})*/
          //networkOpenCluster(selectedNode)
        } else {
          var firstLevelNodes = [];
          var otherLevelNodes = [];
          var connectedToNodes = [];
      
          item = network.body.data.nodes.get({
            filter: function (item) {
              return item.id == selectedNode;
            }
          });
            
          connectedToNodes = network.body.data.edges.get({
          fields: ['id','to'],
            filter: function (item) {
              return item.from == selectedNode;
            },
            returnType :'Array'
          });
          
          
          
          for (j = 0; j < connectedToNodes.length; j++) {
            firstLevelNodes = firstLevelNodes.concat(connectedToNodes[j].to);
          }
    
          var currentConnectedToNodes = firstLevelNodes;
          while(currentConnectedToNodes.length !== 0){
            connectedToNodes = network.body.data.edges.get({
              fields: ['id', 'to'],
                filter: function (item) {
                  return indexOf.call(currentConnectedToNodes, item.from, true) > -1;
                },
                returnType :'Array'
            });
                
            currentConnectedToNodes = [];
            var currentlength = otherLevelNodes.length;
            for (j = 0; j < connectedToNodes.length; j++) {
              otherLevelNodes = uniqueArray(otherLevelNodes.concat(connectedToNodes[j].to), false, network);
              currentConnectedToNodes = uniqueArray(currentConnectedToNodes.concat(connectedToNodes[j].to), false, network);
            }
            if (otherLevelNodes.length === currentlength) { break; }
          }
              
          var finalFirstLevelNodes = [];
          for (j = 0; j < firstLevelNodes.length; j++) {
            var findnode = network.clustering.findNode(firstLevelNodes[j])
            if(findnode.length === 1){
              finalFirstLevelNodes = finalFirstLevelNodes.concat(firstLevelNodes[j]);
            } else {
              finalFirstLevelNodes = finalFirstLevelNodes.concat(findnode[0]);
            }
          }
          
          var finalClusterNodes = [];
          for (j = 0; j < otherLevelNodes.length; j++) {
            var findnode = network.clustering.findNode(otherLevelNodes[j])
            if(findnode.length === 1){
              finalClusterNodes = finalClusterNodes.concat(otherLevelNodes[j]);
            } else {
              finalClusterNodes = finalClusterNodes.concat(findnode[0]);
            }
          }

          if(set_position){ 
            network.storePositions();
          }
    
          var clusterOptions = {
            joinCondition: function (nodesOptions) {
              return nodesOptions.id === selectedNode || indexOf.call(finalFirstLevelNodes, nodesOptions.id, true) > -1 || 
                  indexOf.call(finalClusterNodes, nodesOptions.id, true) > -1; 
              },
              processProperties: function(clusterOptions, childNodes) {
                var click_node = network.body.data.nodes.get({
                  filter: function (item) {
                    return item.id == selectedNode;
                  },
                  returnType :'Array'
                });
                
                var is_hard_to_read = false;
                if(click_node[0].isHardToRead !== undefined){
                  is_hard_to_read = click_node[0].isHardToRead;
                }
                
                for (var i in click_node[0]) {
                  if(i !== "id" && i !== "isHardToRead"){
                    if(i === "label" && is_hard_to_read){
                      clusterOptions[i]=  click_node[0]["hiddenLabel"];
                    } else if(i === "color" && is_hard_to_read) {
                      clusterOptions[i]=  click_node[0]["hiddenColor"];
                    } else {
                       clusterOptions[i]=  click_node[0][i];
                    }
                  }
                }
                        
                // gestion des tree
                if(treeParams !== undefined){
                  if(treeParams.updateShape){
                    clusterOptions.label = clusterOptions.labelClust
                    clusterOptions.color = clusterOptions.colorClust
                    clusterOptions.shape = treeParams.shapeY
                  }
                }
                        
                if(clusterOptions.label !== undefined){
                  clusterOptions.label = clusterOptions.label + " " + labelSuffix;
                } else {
                  clusterOptions.label =  labelSuffix;
                }
                        
                if(clusterOptions.borderWidth !== undefined){
                  clusterOptions.borderWidth = clusterOptions.borderWidth * 3;
                } else {
                  clusterOptions.borderWidth =  3;
                }
                        
                if(set_position){
                  if(click_node[0].x !== undefined){
                    clusterOptions.x = click_node[0].x;
                  }
                  if(click_node[0].y !== undefined){
                    clusterOptions.y = click_node[0].y;
                  }
                }
                      
                if(clusterParams !== undefined){
                  for (var j in clusterParams) {
                    clusterOptions[j]=  clusterParams[j];
                  }
                }
                    
              return clusterOptions;
            },
            clusterNodeProperties: {
              allowSingleNodeCluster: false
            }
          }
          network.cluster(clusterOptions);
        }
      }
      
    }
    if(resetHighlight){
      document.getElementById("nodeSelect"+elid).value = "";
      document.getElementById("nodeSelect"+elid).onchange();
    }
    if(fit){
      network.fit();
    }
  }
};

function uncollapsedNetwork(nodes, fit, resetHighlight, keepCoord, network, elid) {
  var selectedNode;
  var j;
  var arr_nodes = [];
  var cluster_node;
  
  var nodes_in_clusters = network.body.modules.clustering.clusteredNodes;
  if(Object.keys(nodes_in_clusters).length > 0){
    nodes_in_clusters = Object.keys(nodes_in_clusters);
  } else {
    nodes_in_clusters = []
  }
    
  if(nodes !== undefined && nodes !== null){
    arr_nodes = nodes
  } else {
    arr_nodes = nodes_in_clusters;
  }

  for (var inodes = 0; inodes < arr_nodes.length; inodes++) {
    selectedNode = '' + arr_nodes[inodes];
    if(selectedNode !== undefined){
        if(network.isCluster(selectedNode)){
          if(keepCoord){
            network.openCluster(selectedNode, 
              {releaseFunction : function(clusterPosition, containedNodesPositions) {
                    return containedNodesPositions;
                  }
              });
          } else {
            network.openCluster(selectedNode)
          }
        } else {
          if(indexOf.call(nodes_in_clusters, selectedNode, true) > -1){
            // not a cluster into a cluster...
            if(selectedNode.search(/^cluster/i) === -1){
              cluster_node = network.clustering.findNode(selectedNode)[0];
              if(network.isCluster(cluster_node)){
                if(keepCoord){
                  network.openCluster(cluster_node, 
                    {releaseFunction : function(clusterPosition, containedNodesPositions) {
                          return containedNodesPositions;
                        }
                    });
                } else {
                  network.openCluster(cluster_node)
                }
              }
            }
          }
        } 
      }
    }
  if(resetHighlight){
    document.getElementById("nodeSelect"+elid).value = "";
    document.getElementById("nodeSelect"+elid).onchange();
  }
  if(fit){
    network.fit();
  }
};

//----------------------------------------------------------------
// All available functions/methods with visNetworkProxy
//--------------------------------------------------------------- 
if (HTMLWidgets.shinyMode){
  
  // collapsed method
  Shiny.addCustomMessageHandler('visShinyCollapse', function(data){
      // get container id
      var el = document.getElementById("graph"+data.id);
      if(el){
        collapsedNetwork(data.nodes, data.fit, data.resetHighlight, data.clusterOptions, data.labelSuffix, undefined, el.chart, data.id)
      }
  });
  
  // uncollapsed method
  Shiny.addCustomMessageHandler('visShinyUncollapse', function(data){
      // get container id
      var el = document.getElementById("graph"+data.id);
      if(el){
        uncollapsedNetwork(data.nodes, data.fit, data.resetHighlight, data.keepCoord, el.chart, data.id)
      }
  });

  // event method
  Shiny.addCustomMessageHandler('visShinyEvents', function(data){
      // get container id
      var el = document.getElementById("graph"+data.id);
      if(el){
        var network = el.chart;
        
        if(data.type === "once"){
          for (var key in data.events) {
            eval('network.once("' + key + '",' + data.events[key] + ')');
          }
        } else if(data.type === "on"){
          for (var key in data.events) {
            eval('network.on("' + key + '",' + data.events[key] + ')');
          }
        } else if(data.type === "off"){
          for (var key in data.events) {
            eval('network.off("' + key + '",' + data.events[key] + ')');
          }
        }
      }
  });
  
  // moveNode method
  Shiny.addCustomMessageHandler('visShinyMoveNode', function(data){
      // get container id
      var el = document.getElementById("graph"+data.id);
      if(el){
        var network = el.chart;
        network.moveNode(data.nodeId, data.x, data.y);
      }
  });
  
  // unselectAll method
  Shiny.addCustomMessageHandler('visShinyUnselectAll', function(data){
      // get container id
      var el = document.getElementById("graph"+data.id);
      if(el){
        var network = el.chart;
        
        // reset selection
        document.getElementById("nodeSelect"+data.id).value = "";
        document.getElementById("nodeSelect"+data.id).onchange();
        
        if(document.getElementById(data.id).selectActive === true){
            document.getElementById("selectedBy"+data.id).value = "";
            document.getElementById("selectedBy"+data.id).onchange();
        }
        
        network.unselectAll();
      }
  });
  
  // updateOptions in the network
  Shiny.addCustomMessageHandler('visShinyOptions', function(data){
      // get container id
      var el = document.getElementById("graph"+data.id);
      
      if(el){
        var network = el.chart;
        var options = el.options;
        // configure
        if(data.options.configure !== undefined){
          if(data.options.configure.container !== undefined){
            var dom_conf = document.getElementById(data.options.configure.container);
            if(dom_conf !== null){
              data.options.configure.container = dom_conf;
            } else {
              data.options.configure.container = undefined;
            }
          }
        }
    
        //*************************
        // pre-treatment for icons (unicode)
        //*************************
        if(data.options.groups){
          for (var gr in data.options.groups){
            if(data.options.groups[gr].icon){
              if(data.options.groups[gr].icon.code){
                data.options.groups[gr].icon.code = JSON.parse( '"'+'\\u' + data.options.groups[gr].icon.code + '"');
              }
              if(data.options.groups[gr].icon.face){
                if(data.options.groups[gr].icon.face === "'Font Awesome 5 Free'"){
                  data.options.groups[gr].icon.weight = "bold"
                }
              }
              if(data.options.groups[gr].icon.color){
                data.options.groups[gr].color = data.options.groups[gr].icon.color;
              }
            }
          }
        }
        
        if(data.options.nodes){
          if(data.options.nodes.icon){
            if(data.options.nodes.icon.code){
              data.options.nodes.icon.code = JSON.parse( '"'+'\\u' + data.options.nodes.icon.code + '"');
            }
            if(data.options.nodes.icon.face){
              if(data.options.nodes.icon.face === "'Font Awesome 5 Free'"){
                  data.options.nodes.icon.weight = "bold"
              }
            }
            if(data.options.nodes.icon.color){
              data.options.nodes.color = data.options.nodes.icon.color;
            }
          }
        }

        
        update(options, data.options);
        network.setOptions(options);
      }
  });
  
  // setData the network
  Shiny.addCustomMessageHandler('visShinySetData', function(data){
      // get container id
      var el = document.getElementById("graph"+data.id);
      if(el){
        var network = el.chart;
        var newnodes = new vis.DataSet();
        var newedges = new vis.DataSet();
		
        newnodes.add(visNetworkdataframeToD3(data.nodes, "nodes"));
        newedges.add(visNetworkdataframeToD3(data.edges, "edges"));
        var newdata = {
          nodes: newnodes,
          edges: newedges
        };
        network.setData(newdata);
      }
  });
  
  // fit to a specific node
  Shiny.addCustomMessageHandler('visShinyFit', function(data){
    // get container id
    var el = document.getElementById("graph"+data.id);
    if(el){
        var network = el.chart;
        network.fit(data.options);
      }
  });
  
  // focus on a node in the network
  Shiny.addCustomMessageHandler('visShinyFocus', function(data){
      // get container id
      var el = document.getElementById("graph"+data.id);
      if(el){
        var network = el.chart;
        network.focus(data.focusId, data.options);
      }
  });
  
  // stabilize the network
  Shiny.addCustomMessageHandler('visShinyStabilize', function(data){
      // get container id
      var el = document.getElementById("graph"+data.id);
      if(el){
        var network = el.chart;
        network.stabilize(data.options);
      }
  });

  // startSimulation on network
  Shiny.addCustomMessageHandler('visShinyStartSimulation', function(data){
      // get container id
      var el = document.getElementById("graph"+data.id);
      if(el){
        var network = el.chart;
        network.startSimulation();
      }
  });
  
  // stopSimulation on network
  Shiny.addCustomMessageHandler('visShinyStopSimulation', function(data){
      // get container id
      var el = document.getElementById("graph"+data.id);
      if(el){
        var network = el.chart;
        network.stopSimulation();
      }
  });
  
  // get positions of the network
  Shiny.addCustomMessageHandler('visShinyGetPositions', function(data){
      // get container id
      var el = document.getElementById("graph"+data.id);
      if(el){
        var network = el.chart;
        var pos;
        
        if(data.nodes !== undefined){
          pos = network.getPositions(data.nodes);
        }else{
          pos = network.getPositions();
        }
		// return positions in shiny
        Shiny.onInputChange(data.input, pos);
      }
  });
  
  // get edges data
  Shiny.addCustomMessageHandler('visShinyGetEdges', function(data){
      // get container id
      var el = document.getElementById("graph"+data.id);
      if(el){
        // return data in shiny
        Shiny.onInputChange(data.input, el.edges.get({returnType:"Object"}));
      }
  });
  
  // get nodes data
  Shiny.addCustomMessageHandler('visShinyGetNodes', function(data){
      // get container id
      var el = document.getElementById("graph"+data.id);
      if(el){
        if(data.addCoordinates){
          el.chart.storePositions();
        }
        // return data in shiny
        Shiny.onInputChange(data.input, el.nodes.get({returnType:"Object"}));
      }
  });
  
  // get selected edges
  Shiny.addCustomMessageHandler('visShinyGetSelectedEdges', function(data){
      // get container id
      var el = document.getElementById("graph"+data.id);
      if(el){
        var network = el.chart;
        var pos = network.getSelectedEdges();
		    // return  in shiny
        Shiny.onInputChange(data.input, pos);
      }
  });
  
  // get selected nodes
  Shiny.addCustomMessageHandler('visShinyGetSelectedNodes', function(data){
      // get container id
      var el = document.getElementById("graph"+data.id);
      if(el){
        var network = el.chart;
        var pos = network.getSelectedNodes();
		    // return  in shiny
        Shiny.onInputChange(data.input, pos);
      }
  });
  
  // getConnectedEdges
  Shiny.addCustomMessageHandler('visShinyGetConnectedEdges', function(data){
      // get container id
      var el = document.getElementById("graph"+data.id);
      if(el){
        var network = el.chart;
        var pos = network.getConnectedEdges(data.nodeId);
        // return  in shiny
        Shiny.onInputChange(data.input, pos);
      }
  });
  
  // getConnectedNodes
  Shiny.addCustomMessageHandler('visShinyGetConnectedNodes', function(data){
      // get container id
      var el = document.getElementById("graph"+data.id);
      if(el){
        var network = el.chart;
        var pos = network.getConnectedNodes(data.nodeId);
        // return  in shiny
        Shiny.onInputChange(data.input, pos);
      }
  });
  
  // getBoundingBox
  Shiny.addCustomMessageHandler('visShinyGetBoundingBox', function(data){
      // get container id
      var el = document.getElementById("graph"+data.id);
      if(el){
        var network = el.chart;
        var pos = network.getBoundingBox(data.nodeId);
        // return  in shiny
        Shiny.onInputChange(data.input, pos);
      }
  });
  
  // get selection
  Shiny.addCustomMessageHandler('visShinyGetSelection', function(data){
      // get container id
      var el = document.getElementById("graph"+data.id);
      if(el){
        var network = el.chart;
        var pos;
        
        pos = network.getSelection();

		    // return  in shiny
        Shiny.onInputChange(data.input, pos);
      }
  });
  
  // get scale
  Shiny.addCustomMessageHandler('visShinyGetScale', function(data){
      // get container id
      var el = document.getElementById("graph"+data.id);
      if(el){
        var network = el.chart;
        var pos;
        
        pos = network.getScale();

		    // return  in shiny
        Shiny.onInputChange(data.input, pos);
      }
  });
  
  // store positions
  Shiny.addCustomMessageHandler('visShinyStorePositions', function(data){
      // get container id
      var el = document.getElementById("graph"+data.id);
      if(el){
        var network = el.chart;
        network.storePositions();
      }
  });
  
  // get view position
  Shiny.addCustomMessageHandler('visShinyGetViewPosition', function(data){
      // get container id
      var el = document.getElementById("graph"+data.id);
      if(el){
        var network = el.chart;
        var pos;
        
        pos = network.getViewPosition();

		    // return  in shiny
        Shiny.onInputChange(data.input, pos);
      }
  });
  
  // get view position
  Shiny.addCustomMessageHandler('visShinyGetOptionsFromConfigurator', function(data){
      // get container id
      var el = document.getElementById("graph"+data.id);
      if(el){
        var network = el.chart;
		    // return  in shiny
        Shiny.onInputChange(data.input, network.getOptionsFromConfigurator());
      }
  });
  
  // Redraw the network
  Shiny.addCustomMessageHandler('visShinyRedraw', function(data){
      // get container id
      var el = document.getElementById("graph"+data.id);
      if(el){
        el.chart.redraw();
      }
  });
  
  // select nodes
  Shiny.addCustomMessageHandler('visShinySelectNodes', function(data){
      // get container id
      var el = document.getElementById("graph"+data.id);
      if(el){
        var network = el.chart;
        if(data.selid !== null){
          network.selectNodes(data.selid, data.highlightEdges);
          if(data.clickEvent){
            el.myclick({nodes : data.selid});
          }
        }else{
          if(data.clickEvent){
            el.myclick({nodes : []});
          }
        }
      }
  });
  
  // select edges
  Shiny.addCustomMessageHandler('visShinySelectEdges', function(data){
      // get container id
      var el = document.getElementById("graph"+data.id);
      if(el){
        var network = el.chart;
        if(data.selid !== null){
          network.selectEdges(data.selid);
        }
      }
  });
  
  // set selection
  Shiny.addCustomMessageHandler('visShinySetSelection', function(data){
      // get container id
      var el = document.getElementById("graph"+data.id);
      if(el){
        var network = el.chart;
        if(data.selection.nodes !== null || data.selection.edges !== null){
          network.setSelection(data.selection, data.options);
        }
        if(data.clickEvent){
          if(data.selection.nodes !== null){
            el.myclick({nodes : data.selection.nodes});
          } else {
           el.myclick({nodes : []}); 
          }
        }
      }
  });
  
  function updateVisOptions(data){
        // get container id
        var graph = document.getElementById("graph"+data.id);
        var el = document.getElementById(data.id);
        var do_loop_by = false;
        var option2;
        var selectList2;
        var selectList;
        var reset = false;
        
        if(graph){
          // reset nodes before ?
          if(document.getElementById(el.id).highlight){
            // need reset nodes
            if(document.getElementById(el.id).highlightActive === true){
              reset = true;
            }
          }
          if(reset){
            document.getElementById("nodeSelect"+data.id).value = "";
            document.getElementById("nodeSelect"+data.id).onchange();
          }
          
          // collapse init
          if(data.options.collapse !== undefined){
            el.collapse = data.options.collapse.enabled;
            el.collapseFit = data.options.collapse.fit;
            el.collapseResetHighlight = data.options.collapse.resetHighlight;
            el.collapseKeepCoord = data.options.collapse.keepCoord;
            el.collapseLabelSuffix = data.options.collapse.labelSuffix;
            el.clusterOptions = data.options.collapse.clusterOptions;
          }
          
          // highlight init
          if(data.options.highlight !== undefined){
            el.highlight = data.options.highlight.enabled;
            el.degree = data.options.highlight.degree;
            el.hoverNearest = data.options.highlight.hoverNearest;
            el.highlightColor = data.options.highlight.hideColor;
            el.highlightAlgorithm = data.options.highlight.algorithm;
            el.highlightLabelOnly = data.options.labelOnly;
          }

          // byselection init
          if(data.options.byselection !== undefined){
            if(data.options.byselection.selected !== undefined){
              document.getElementById("selectedBy"+data.id).value = data.options.byselection.selected;
              document.getElementById("selectedBy"+data.id).onchange();
            }
            if(data.options.byselection.hideColor){
              el.byselectionColor = data.options.byselection.hideColor;
            }
            if(data.options.byselection.highlight !== undefined){
              el.byselectionHighlight = data.options.byselection.highlight;
            }
          }
          
          if(data.options.byselection !== undefined){
            selectList2 = document.getElementById("selectedBy"+data.id)
            selectList2.options.length = 0;
            if(data.options.byselection.enabled === true){
              option2 = document.createElement("option");
              option2.value = "";
              if(data.options.byselection.main === undefined){
                option2.text = "Select by " + data.options.byselection.variable;
              } else {
                option2.text = data.options.byselection.main;
              }
              
              selectList2.appendChild(option2);
      
              if(data.options.byselection.values !== undefined){
                for (var i = 0; i < data.options.byselection.values.length; i++) {
                  option2 = document.createElement("option");
                  option2.value = data.options.byselection.values[i];
                  option2.text = data.options.byselection.values[i];
                  selectList2.appendChild(option2);
                }
              }else{
                do_loop_by = true;
              }

              el.byselection_variable = data.options.byselection.variable;
              el.byselection_multiple = data.options.byselection.multiple;
              selectList2.style.display = 'inline';
              if(data.options.byselection.style !== undefined){
                selectList2.setAttribute('style', data.options.byselection.style);
              }
              el.byselection = true;
            } else {
              selectList2.style.display = 'none';
              el.byselection = false;
              // reset selection
              if(el.selectActive === true){
                document.getElementById("selectedBy"+data.id).value = "";
                document.getElementById("selectedBy"+data.id).onchange();
              }
            }
          }else{
            // reset selection
            if(el.selectActive === true){
              document.getElementById("selectedBy"+data.id).value = "";
              document.getElementById("selectedBy"+data.id).onchange();
            }
          }
          
          if(do_loop_by){
              var allNodes = graph.nodes.get({returnType:"Object"});
              var byselection_values = [];
              for (var nodeId in allNodes) {
                if(do_loop_by){
                  var current_sel_value = allNodes[nodeId][data.options.byselection.variable];
                  if(data.options.byselection.multiple){
                    current_sel_value = current_sel_value.split(",").map(Function.prototype.call, String.prototype.trim);
                  }else{
                    current_sel_value = [current_sel_value];
                  }
                  for(var ind_c in current_sel_value){
                    if(indexOf.call(byselection_values, current_sel_value[ind_c], false) === -1){
                      option2 = document.createElement("option");
                      option2.value = current_sel_value[ind_c];
                      option2.text = current_sel_value[ind_c];
                      selectList2.appendChild(option2);
                      byselection_values.push(current_sel_value[ind_c]);
                    }
                  }
                }
              } 
          }
          
          // node id selection init
          if(data.options.idselection !== undefined){
            selectList = document.getElementById("nodeSelect"+data.id)
            selectList.options.length = 0;
            if(data.options.idselection.enabled === true){
              setNodeIdList(selectList, data.options.idselection, graph.nodes)
              el.idselection = true;
            } else {
              selectList.style.display = 'none';
              el.idselection = false;
            }
            if(data.options.idselection.useLabels !== undefined){
              el.idselection_useLabels = data.options.idselection.useLabels
            }
          }
          
          if(data.options.idselection !== undefined){
            if(data.options.idselection.enabled === true && data.options.idselection.selected !== undefined){
              document.getElementById("nodeSelect"+data.id).value = data.options.idselection.selected;
              document.getElementById("nodeSelect"+data.id).onchange();
            }
          }
        }
  };
      
  Shiny.addCustomMessageHandler('visShinyCustomOptions', updateVisOptions);
  
  // udpate nodes data
  Shiny.addCustomMessageHandler('visShinyUpdateNodes', function(data){
      // get container id
      var el = document.getElementById("graph"+data.id);
      var main_el = document.getElementById(data.id);
      
      if(data.legend === false){
        if(el){
          // get & transform nodes object
          var tmpnodes = visNetworkdataframeToD3(data.nodes, "nodes");
          
          // reset some parameters / data before
          if (main_el.selectActive === true | main_el.highlightActive === true) {

            //reset nodes
            resetAllEdges(el.edges, el.highlightColor, el.byselectionColor, el.chart);
            resetAllNodes(el.nodes, true, el.options, el.chart, false);
            
            if (main_el.selectActive === true){
              main_el.selectActive = false;
              resetList('selectedBy', data.id, 'selectedBy');
            }
            if (main_el.highlightActive === true){
              main_el.highlightActive = false;
              resetList('nodeSelect', data.id, 'selected');
            }
          }
          // update nodes
          el.nodes.update(tmpnodes);
          
          // update options ?
          if(data.updateOptions){
            var dataOptions = {};
            dataOptions.options = {};
          
            var updateOpts = false;
            if(document.getElementById("nodeSelect"+data.id).style.display === 'inline'){
              updateOpts = true;
              dataOptions.id  = data.id;
              dataOptions.options.idselection = {enabled : true, useLabels : main_el.idselection_useLabels};
            }
      
            if(document.getElementById("selectedBy"+data.id).style.display === 'inline'){
              updateOpts = true;
              dataOptions.id  = data.id;
              dataOptions.options.byselection = {enabled : true, variable : main_el.byselection_variable, multiple : main_el.byselection_multiple};
            }
          
            if(updateOpts){
              updateVisOptions(dataOptions);
            }
          }
        }
      } else if(data.legend === true){
        var legend_network = document.getElementById("legend"+data.id);
        if(legend_network){
          // get & transform nodes object
          var tmpnodes = visNetworkdataframeToD3(data.nodes, "nodes");
          // update nodes
          legend_network.network.body.data.nodes.update(tmpnodes);
          // fit
          legend_network.network.fit();
        }
      }
  });

  // udpate edges data
  Shiny.addCustomMessageHandler('visShinyUpdateEdges', function(data){
      // get container id
      var el = document.getElementById("graph"+data.id);
      if(data.legend === false){
        if(el){
          // get edges object
          var tmpedges = visNetworkdataframeToD3(data.edges, "edges");
          // reset edges
          resetAllEdges(el.edges, el.highlightColor,  el.byselectionColor, el.chart)
          el.edges.update(tmpedges);
        }
      } else if(data.legend === true){
        var legend_network = document.getElementById("legend"+data.id);
        if(legend_network){
          // get & transform nodes object
          var tmpedges = visNetworkdataframeToD3(data.edges, "edges");
          // update edges
          legend_network.network.body.data.edges.update(tmpedges);
          // fit
          legend_network.network.fit();
        }
      }
  });
  
  // remove nodes
  Shiny.addCustomMessageHandler('visShinyRemoveNodes', function(data){
      // get container id
      var el = document.getElementById("graph"+data.id);
      var main_el = document.getElementById(data.id);
      if(data.legend === false){
        if(el){
          // reset some parameters / date before
          if (main_el.selectActive === true | main_el.highlightActive === true) {
            //reset nodes
            resetAllNodes(el.nodes, true, el.options, el.chart, false);
            
            if (main_el.selectActive === true){
              main_el.selectActive = false;
              resetList('selectedBy', data.id, 'selectedBy');
            }
            if (main_el.highlightActive === true){
              main_el.highlightActive = false;
              resetList('nodeSelect', data.id, 'selected');
            }
          }
          // remove nodes
          el.nodes.remove(data.rmid);
  
          // update options ?
          if(data.updateOptions){
            var dataOptions = {};
            dataOptions.options = {};
          
            var updateOpts = false;
            if(document.getElementById("nodeSelect"+data.id).style.display === 'inline'){
              updateOpts = true;
              dataOptions.id  = data.id;
              dataOptions.options.idselection = {enabled : true, useLabels : main_el.idselection_useLabels};
            }
      
            if(document.getElementById("selectedBy"+data.id).style.display === 'inline'){
              updateOpts = true;
              dataOptions.id  = data.id;
              dataOptions.options.byselection = {enabled : true, variable : main_el.byselection_variable, multiple : main_el.byselection_multiple};
            }
          
            if(updateOpts){
              updateVisOptions(dataOptions);
            }
          }
        }
      } else if(data.legend === true){
        var legend_network = document.getElementById("legend"+data.id);
        if(legend_network){
          // remove nodes
          legend_network.network.body.data.nodes.remove(data.rmid);
          // fit
          legend_network.network.fit();
        }
      }
  });
  
  // remove edges
  Shiny.addCustomMessageHandler('visShinyRemoveEdges', function(data){
      // get container id
      var el = document.getElementById("graph"+data.id);
      if(data.legend === false){
        if(el){
          // reset edges
          resetAllEdges(el.edges, el.highlightColor,  el.byselectionColor, el.chart)
          el.edges.remove(data.rmid);
        }
      } else if(data.legend === true){
        var legend_network = document.getElementById("legend"+data.id);
        if(legend_network){
          // remove edges
          legend_network.network.body.data.edges.remove(data.rmid);
          // fit
          legend_network.network.fit();
        }
      }
  });
  
  // remove edges
  Shiny.addCustomMessageHandler('visShinySetTitle', function(data){
    if(data.main !== null){
      var div_title = document.getElementById("title" + data.id);
      if(div_title !== null){
        if(data.main.hidden === true){
          div_title.style.display = 'none';
        } else {
          if(data.main.text !== undefined){
            if(data.main.text !== null){
              if(data.main.text.length > 0){
                div_title.innerHTML = data.main.text;
              } else {
                div_title.innerHTML = "";
              }
            }
          }
          if(data.main.style !== undefined){
            if(data.main.style !== null){
              if(data.main.style.length > 0){
                div_title.setAttribute('style',  data.main.style);
              }
            }
          }
          div_title.style.display = 'block';
        } 
      }
    }
    if(data.submain !== null){
      var div_subtitle = document.getElementById("subtitle" + data.id);
      if(div_subtitle !== null){
        if(data.submain.hidden === true){
          div_subtitle.style.display = 'none';
        } else {
          if(data.submain.text !== undefined){
            if(data.submain.text !== null){
              if(data.submain.text.length > 0){
                div_subtitle.innerHTML = data.submain.text;
              } else {
                div_subtitle.innerHTML = "";
              }
            }
          }
          if(data.submain.style !== undefined){
            if(data.submain.style !== null){
              if(data.submain.style.length > 0){
                div_subtitle.setAttribute('style',  data.submain.style);
              }
            }
          }
          div_subtitle.style.display = 'block';
        }
      }
    }
    if(data.footer !== null){
      var div_footer = document.getElementById("footer" + data.id);
      if(div_footer !== null){
        if(data.footer.hidden === true){
          div_footer.style.display = 'none';
        } else {
          if(data.footer.text !== undefined){
            if(data.footer.text !== null){
              if(data.footer.text.length > 0){
                div_footer.innerHTML = data.footer.text;
              } else {
                div_footer.innerHTML = "";
              }
            }
          }
          if(data.footer.style !== undefined){
            if(data.footer.style !== null){
              if(data.footer.style.length > 0){
                div_footer.setAttribute('style',  data.footer.style);
              }
            }
          }
          div_footer.style.display = 'block';
        } 
      }
    }
  });

  // updateTree
  Shiny.addCustomMessageHandler('visShinyUpdateTree', function(data){
      // get container id
      var el = document.getElementById(data.id);
      if(el){
      if(el.tree){
          if(data.tree.updateShape != undefined){
            el.tree.updateShape = data.tree.updateShape
          }
          if(data.tree.shapeVar != undefined){
            el.tree.shapeVar = data.tree.shapeVar
          }
          if(data.tree.shapeY != undefined){
            el.tree.shapeY = data.tree.shapeY
          }
        }
      }
  });
}

//----------------------------------------------------------------
// HTMLWidgets.widget Definition
//--------------------------------------------------------------- 
HTMLWidgets.widget({
  
  name: 'visNetwork',
  
  type: 'output',
  
  initialize: function(el, width, height) {
    return {
    };
  },
  
  renderValue: function(el, x, instance) {
    var data;
    var nodes;
    var edges;


    // clustergin by zoom variables
    var clusterIndex = 0;
    var clusters = [];
    var lastClusterZoomLevel = 0;
    var clusterFactor;
    var ctrlwait = 0;
    
    // legend control
    var addlegend = false;

    // main div el.id
    var el_id = document.getElementById(el.id);
    
    // test background
    el_id.style.background = x.background;
    
    // clear el.id (for shiny...)
    el_id.innerHTML = "";  
    
    // shared control with proxy function (is there a better way ?)
    el_id.highlightActive = false;
    el_id.selectActive = false;
    el_id.idselection = x.idselection.enabled;
    el_id.byselection = x.byselection.enabled;
    
    if(x.highlight !== undefined){
      el_id.highlight = x.highlight.enabled;
      el_id.highlightColor = x.highlight.hideColor;
      el_id.hoverNearest = x.highlight.hoverNearest;
      el_id.degree = x.highlight.degree;
      el_id.highlightAlgorithm = x.highlight.algorithm;
      el_id.highlightLabelOnly = x.highlight.labelOnly;
    } else {
      el_id.highlight = false;
      el_id.hoverNearest = false;
      el_id.highlightColor = 'rgba(200,200,200,0.5)';
      el_id.degree = 1;
      el_id.highlightAlgorithm = "all";
      el_id.highlightLabelOnly = true;
    }

    if(x.byselection.enabled){
      el_id.byselectionColor = x.byselection.hideColor;
      el_id.byselectionHighlight = x.byselection.highlight;
    } else {
      el_id.byselectionColor = 'rgba(200,200,200,0.5)';
      el_id.byselectionHighlight = false;
    }
    
    if(x.idselection.enabled){
      el_id.idselection_useLabels = true;
    } else {
      el_id.idselection_useLabels = false;
    }
    
    if(x.collapse !== undefined){
      if(x.collapse.enabled){
        el_id.collapse = true;
        el_id.collapseFit = x.collapse.fit;
        el_id.collapseResetHighlight = x.collapse.resetHighlight;
        el_id.collapseKeepCoord = x.collapse.keepCoord;
        el_id.collapseLabelSuffix = x.collapse.labelSuffix;
        el_id.clusterOptions = x.collapse.clusterOptions;
      }
    } else {
      el_id.collapse = false;
      el_id.collapseFit = false;
      el_id.collapseResetHighlight = false;
      el_id.collapseKeepCoord = true;
      el_id.collapseLabelSuffix = " (cluster)";
      el_id.clusterOptions = undefined;
    }
    
    if(x.tree !== undefined){
      el_id.tree = x.tree;
    }

    // configure
    if(x.options.configure !== undefined){
      if(x.options.configure.container !== undefined){
        var dom_conf = document.getElementById(x.options.configure.container);
        if(dom_conf !== null){
          x.options.configure.container = dom_conf;
        } else {
          x.options.configure.container = undefined;
        }
      }
    }
    
    var changeInput = function(id, data) {
            Shiny.onInputChange(el.id + '_' + id, data);
    };
          
    //*************************
    //title
    //*************************
    var div_title = document.createElement('div');
    div_title.id = "title"+el.id;
    div_title.setAttribute('style','font-family:Georgia, Times New Roman, Times, serif;font-weight:bold;font-size:20px;text-align:center;');
    div_title.style.display = 'none';
    el_id.appendChild(div_title);  
    if(x.main !== null){
      div_title.innerHTML = x.main.text;
      div_title.setAttribute('style',  x.main.style + ";background-color: inherit;");
      div_title.style.display = 'block';
    }
    
    //*************************
    //subtitle
    //*************************
    var div_subtitle = document.createElement('div');
    div_subtitle.id = "subtitle"+el.id;
    div_subtitle.setAttribute('style',  'font-family:Georgia, Times New Roman, Times, serif;font-size:12px;text-align:center;');
    div_subtitle.style.display = 'none';
    el_id.appendChild(div_subtitle); 
    if(x.submain !== null){
      div_subtitle.innerHTML = x.submain.text;
      div_subtitle.setAttribute('style',  x.submain.style + ";background-color: inherit;");
      div_title.style.display = 'block';
    }
 
    //*************************
    //init idselection
    //*************************
    function onIdChange(id, init) {
      if(id === ""){
        instance.network.selectNodes([]);
      }else{
        instance.network.selectNodes([id]);
      }
      if(el_id.highlight){
        neighbourhoodHighlight(instance.network.getSelection().nodes, "click", el_id.highlightAlgorithm, true);
      }else{
        if(init){
          selectNode = document.getElementById('nodeSelect'+el.id);
          if(x.idselection.values !== undefined){
            if(indexOf.call(x.idselection.values, id, true) > -1){
              selectNode.value = id;
            }else{
              selectNode.value = "";
            }
          }else{
            selectNode.value = id;
          }
        }
      }
      if (window.Shiny){
        changeInput('selected', document.getElementById("nodeSelect"+el.id).value);
      }
      if(el_id.byselection){
        resetList('selectedBy', el.id, 'selectedBy');
      }
    }
      
    // id nodes selection : add a list on top left
    // actually only with nodes + edges data (not dot and gephi)
    var idList = document.createElement("select");
    idList.setAttribute('class', 'dropdown');
    idList.style.display = 'none';
    idList.id = "nodeSelect"+el.id;
    el_id.appendChild(idList);
      
    idList.onchange =  function(){
      if(instance.network){
        onIdChange(document.getElementById("nodeSelect"+el.id).value, false);
      }
    };
      
    var hr = document.createElement("hr");
    hr.setAttribute('style', 'height:0px; visibility:hidden; margin-bottom:-1px;');
    el_id.appendChild(hr);  
      
    //*************************
    //selectedBy
    //*************************
    function onByChange(value) {
        if(instance.network){
          selectedHighlight(value);
        }
        if (window.Shiny){
          changeInput('selectedBy', value);
        }
        if(el_id.idselection){
          resetList('nodeSelect', el.id, 'selected');
        }
    }
    
    // selectedBy : add a list on top left
    // actually only with nodes + edges data (not dot and gephi)
    //Create and append select list
    var byList = document.createElement("select");
    byList.setAttribute('class', 'dropdown');
    byList.style.display = 'none';
    byList.id = "selectedBy"+el.id;
    el_id.appendChild(byList);

    byList.onchange =  function(){
      onByChange(document.getElementById("selectedBy"+el.id).value);
    };
      
    if(el_id.byselection){

      el_id.byselection_values = x.byselection.values;
      el_id.byselection_variable = x.byselection.variable;
      el_id.byselection_multiple = x.byselection.multiple;
      var option2;
      
      //Create and append select list
      var selectList2 = document.getElementById("selectedBy"+el.id);
      selectList2.setAttribute('style', x.byselection.style);
      selectList2.style.display = 'inline';
      
      option2 = document.createElement("option");
      option2.value = "";
      if(x.byselection.main === undefined){
        option2.text = "Select by " + x.byselection.variable;
      } else {
        option2.text = x.byselection.main;
      }

      selectList2.appendChild(option2);
      
      //Create and append the options
      for (var i2 = 0; i2 < x.byselection.values.length; i2++) {
        option2 = document.createElement("option");
        option2.value = x.byselection.values[i2];
        option2.text = x.byselection.values[i2];
        selectList2.appendChild(option2);
      }
      
      if (window.Shiny){
        changeInput('selectedBy', document.getElementById("selectedBy"+el.id).value);
      }
    }
    
    //*************************
    // pre-treatment for icons (unicode)
    //*************************
    if(x.options.groups){
      for (var gr in x.options.groups){
        if(x.options.groups[gr].icon){
          if(x.options.groups[gr].icon.code){
            x.options.groups[gr].icon.code = JSON.parse( '"'+'\\u' + x.options.groups[gr].icon.code + '"');
          }
          if(x.options.groups[gr].icon.face){
            if(x.options.groups[gr].icon.face === "'Font Awesome 5 Free'"){
                x.options.groups[gr].icon.weight = "bold"
            }
          }
          if(x.options.groups[gr].icon.color){
            x.options.groups[gr].color = x.options.groups[gr].icon.color;
          }
        }
      }
    }
    
    if(x.options.nodes.icon){
        if(x.options.nodes.icon.code){
          x.options.nodes.icon.code = JSON.parse( '"'+'\\u' + x.options.nodes.icon.code + '"');
        }
        if(x.options.nodes.icon.face){
          if(x.options.nodes.icon.face === "'Font Awesome 5 Free'"){
              x.options.nodes.icon.weight = "bold"
          }
        }
        if(x.options.nodes.icon.color){
          x.options.nodes.color = x.options.nodes.icon.color;
        }
    }
    
    //*************************
    //page structure
    //*************************
    
    // divide page
    var maindiv  = document.createElement('div');
    maindiv.id = "maindiv"+el.id;
    maindiv.setAttribute('style', 'height:95%;background-color: inherit;');
    el_id.appendChild(maindiv);
    
    var graph = document.createElement('div');
    graph.id = "graph"+el.id;
    
    if(x.legend !== undefined){
      if((x.groups && x.legend.useGroups) || (x.legend.nodes !== undefined) || (x.legend.edges !== undefined)){
        addlegend = true;
      }
    }
    
    //legend
    if(addlegend){
      var legendwidth = x.legend.width*100;
      var legend = document.createElement('div');
      
      var pos = x.legend.position;
      var pos2 = "right";
      if(pos == "right"){
        pos2 = "left";
      }
      
      legend.id = "legend"+el.id;
      legend.setAttribute('style', 'float:' + pos + '; width:'+legendwidth+'%;height:100%');
      
      //legend title
      if(x.legend.main !== undefined){
        var legend_title = document.createElement('div');
        legend_title.innerHTML = x.legend.main.text;
        legend_title.setAttribute('style',  x.legend.main.style);
        legend.appendChild(legend_title);  
        
        legend.id = "legend_main"+el.id;
        var legend_network = document.createElement('div');
        legend_network.id = "legend"+el.id;
        legend_network.setAttribute('style', 'height:100%');
        legend.appendChild(legend_network); 
      }
      
      document.getElementById("maindiv"+el.id).appendChild(legend);
      graph.setAttribute('style', 'float:' + pos2 + '; width:'+(100-legendwidth)+'%;height:100%;background-color: inherit;');
    }else{
      graph.setAttribute('style', 'float:right; width:100%;height:100%;background-color: inherit;');
    }
    
    document.getElementById("maindiv"+el.id).appendChild(graph);
    
    //*************************
    //legend definition
    //*************************
    if(addlegend){
      
      var legendnodes = new vis.DataSet();
      var legendedges = null;
      var datalegend;
      var tmpnodes;
      
      // set some options
      var optionslegend = {
        interaction:{
          dragNodes: false,
          dragView: false,
          selectable: false,
          zoomView: x.legend.zoom
        },
        physics:{
          stabilization: false
        }
      };
      
      function range(start, length, step, rep){
        var a=[], b=start;
        while(a.length < length){
          for (var i = 0; i < rep; i++){
            a.push(b);
            if(a.length === length){
              break;
            }
          }
          b+=step;
        }
        return a;
      };
      
      var mynetwork = document.getElementById('legend'+el.id);
      var lx = mynetwork.clientWidth / 2 + 50;
      var ly = mynetwork.clientHeight / 2 + 50;
      var edge_ly = ly;
      var ncol = x.legend.ncol;
      var step_x = x.legend.stepX;
      var step_y = x.legend.stepY;
      var tmp_ly;
      var tmp_lx = lx;
      var tmp_lx2;
      var all_tmp_y = [];
      if(tmp_lx === 0){
        tmp_lx = 1
      }
      
      // construct nodes data if needed
      if(x.legend.nodes !== undefined){
        if(x.legend.nodesToDataframe){ // data in data.frame
          tmpnodes = visNetworkdataframeToD3(x.legend.nodes, "nodes")
        } else { // data in list
          tmpnodes = x.legend.nodes;
        }
        // only one element   
        if(tmpnodes.length === undefined){
          tmpnodes = new Array(tmpnodes);
        }
      }
      
      // array of y position 
      if(x.groups && x.legend.useGroups && x.legend.nodes !== undefined){
        all_tmp_y = range(ly, x.groups.length + tmpnodes.length, step_y, ncol);
      } else if(x.groups && x.legend.useGroups && x.legend.nodes === undefined){
        all_tmp_y = range(ly, x.groups.length, step_y, ncol);
      } else if(x.legend.useGroups === false && x.legend.nodes !== undefined){
        all_tmp_y = range(ly, tmpnodes.length, step_y, ncol);
      }
      
      // want to view groups in legend
      if(x.groups && x.legend.useGroups){
        // create data
        for (var g1 = 0; g1 < x.groups.length; g1++){
          
          if(g1 === 0){
            tmp_lx = lx;
          } else {
            tmp_lx = lx + g1%ncol * step_x;
          }
          
          tmp_ly = all_tmp_y[g1];
          if(tmp_ly === 0){
            tmp_ly = 1
          }
          
          legendnodes.add({id: null, x : tmp_lx, y : tmp_ly, label: x.groups[g1], group: x.groups[g1], value: 1, mass:1});
          edge_ly = tmp_ly;
        }
        // control icon size
        if(x.options.groups){
          optionslegend.groups = clone(x.options.groups);
          for (var grp in optionslegend.groups) {
            if(optionslegend.groups[grp].shape === "icon"){
              optionslegend.groups[grp].icon.size = 50;
            }
          }
        }
      }
      // want to add custom nodes
      if(x.legend.nodes !== undefined){
        
        // control icon
        for (var nd in tmpnodes){
          if(tmpnodes[nd].icon  && !x.legend.nodesToDataframe){
            tmpnodes[nd].icon.code = JSON.parse( '"'+'\\u' + tmpnodes[nd].icon.code + '"');
          }
          if(tmpnodes[nd].icon  && tmpnodes[nd].icon.face){
            if(tmpnodes[nd].icon.face === "'Font Awesome 5 Free'"){
              tmpnodes[nd].icon.weight = "bold"
            }
          }
        }
        // group control for y
        var add_gr_y = 0;
        if(x.groups && x.legend.useGroups){
          add_gr_y = x.groups.length;
        }
        // set coordinates
        for (var g = 0; g < tmpnodes.length; g++){
          if((g+legendnodes.length) === 0){
            tmp_lx = lx;
          } else {
            tmp_lx = lx + (g+legendnodes.length)%ncol * step_x;
          }
          
          tmp_ly = all_tmp_y[add_gr_y + g];
          if(tmp_lx === 0){
            tmp_lx = 1
          }
          if(tmp_ly === 0){
            tmp_ly = 1
          }
          tmpnodes[g].x = tmp_lx;
          tmpnodes[g].y = tmp_ly;
          
          if(tmpnodes[g].value === undefined && tmpnodes[g].size === undefined){
            tmpnodes[g].value = 1;
          }
          /*if(tmpnodes[g].id !== undefined){
            tmpnodes[g].id = null;
          }*/
          tmpnodes[g].mass = 1;
          edge_ly = tmp_ly;
        }
        legendnodes.add(tmpnodes);
      }
      // want to add custom edges
      if(x.legend.edges !== undefined){
        if(x.legend.edgesToDataframe){ // data in data.frame
          legendedges = visNetworkdataframeToD3(x.legend.edges, "edges")
        } else {  // data in list
          legendedges = x.legend.edges;
        }
        // only one element 
        if(legendedges.length === undefined){
          legendedges = new Array(legendedges);
        }

        // set coordinates and options
        for (var edg = 0; edg < (legendedges.length); edg++){
          
          var tmp_int = Math.floor(Math.random() * 1001);
          legendedges[edg].from = edg + "tmp_leg_edges_" + tmp_int + "_1";
          legendedges[edg].to = edg + "tmp_leg_edges_" + tmp_int + "_2";
          legendedges[edg].physics = false;
          legendedges[edg].smooth = false;
          legendedges[edg].value = undefined;

          if(legendedges[edg].arrows === undefined){
            legendedges[edg].arrows = 'to';
          }
          
          if(legendedges[edg].width === undefined){
            legendedges[edg].width = 1;
          }

          tmp_ly = edge_ly + (edg+1)*step_y;
          if(tmp_ly === 0){
            tmp_ly = 1
          }
          
          if(ncol === 1){
            tmp_lx = lx - mynetwork.clientWidth/3;
            tmp_lx2 = lx + mynetwork.clientWidth/3;
          } else {
            tmp_lx = lx;
            tmp_lx2 = lx + (ncol-1) * step_x;
          }
          
          if(tmp_lx === 0){
            tmp_lx = 1
          }
          
          if(tmp_lx2 === 0){
            tmp_lx2 = 1
          }
          
          legendnodes.add({id: edg + "tmp_leg_edges_" + tmp_int + "_1", x : tmp_lx, y : tmp_ly, size : 0.0001, hidden : false, shape : "square", mass:1});
          legendnodes.add({id: edg + "tmp_leg_edges_" + tmp_int + "_2", x : tmp_lx2, y : tmp_ly, size : 0.0001, hidden : false, shape : "square", mass:1});
        }
      }
      
      // render legend network
      datalegend = {
        nodes: legendnodes, 
        edges: legendedges       
      };

      
      instance.legend = new vis.Network(document.getElementById("legend"+el.id), datalegend, optionslegend);
      //link network for update for re-use and update
      document.getElementById("legend"+el.id).network = instance.legend;
    }
    
    //*************************
    // Main Network rendering
    //*************************
    if(x.nodes){
      
      // network
      nodes = new vis.DataSet();
      edges = new vis.DataSet();
      
      var tmpnodes;
      if(x.nodesToDataframe){ // data in data.frame
        tmpnodes = visNetworkdataframeToD3(x.nodes, "nodes")
      } else { // data in list
        tmpnodes = x.nodes;
      }
      // only one element   
      if(tmpnodes.length === undefined){
        tmpnodes = new Array(tmpnodes);
      }
        
      // update coordinates if igraph
      if(x.igraphlayout !== undefined){
        // to improved
        var zoomLevel = -232.622349 / (tmpnodes.length + 91.165919)  +2.516861;
        var igclientWidth = document.getElementById("graph"+el.id).clientWidth;
        var scalex = 100;
        var scaley = 100;
        
        // current div visibled
        if(igclientWidth !== 0){
          var factor = igclientWidth / 1890;
          zoomLevel = zoomLevel/factor;
          var scalex = (igclientWidth / 2) * zoomLevel;
          var scaley = scalex;
          if(x.igraphlayout.type !== "square"){
            scaley = (document.getElementById("graph"+el.id).clientHeight / 2) * zoomLevel;
          }
        } else {
          // current div not visibled....
          igclientWidth = parseInt(el_id.style.width);
          if(igclientWidth !== 0){
            var factor = igclientWidth / 1890;
            zoomLevel = zoomLevel/factor;
            var scalex = (igclientWidth / 2) * zoomLevel;
            var scaley = scalex;
            if(x.igraphlayout.type !== "square"){
              scaley = (parseInt(el_id.style.height) / 2) * zoomLevel;
            }
          }
        }
        
        for (var nd in tmpnodes) {
          tmpnodes[nd].x = tmpnodes[nd].x * scalex;
          tmpnodes[nd].y = tmpnodes[nd].y * scaley;
        }
      }
      
      nodes.add(tmpnodes);
      
      var tmpedges;
      if(x.edgesToDataframe){ // data in data.frame
        tmpedges = visNetworkdataframeToD3(x.edges, "edges")
      } else { // data in list
        tmpedges = x.edges;
      }
      // only one element
      if(tmpedges !== null){
        if(tmpedges.length === undefined){
          tmpedges = new Array(tmpedges);
        }
        edges.add(tmpedges);  
      }
      
      // reset tmpnodes
      tmpnodes = null;
      
      data = {
        nodes: nodes,
        edges: edges
      };
      
      //save data for re-use and update
      document.getElementById("graph"+el.id).nodes = nodes;
      document.getElementById("graph"+el.id).edges = edges;

    }else if(x.dot){
      data = {
        dot: x.dot
      };
    }else if(x.gephi){
      data = {
        gephi: x.gephi
      };
    } 
    
    var options = x.options;

    //*************************
    //manipulation
    //*************************
    if(x.options.manipulation.enabled){

      var style = document.createElement('style');
      style.type = 'text/css';
      style.appendChild(document.createTextNode(x.opts_manipulation.datacss));
      document.getElementsByTagName("head")[0].appendChild(style);

      var div_addnode = document.createElement('div');
      div_addnode.id = 'addnode-popUp';
      div_addnode.classList.add('network-popUp');
      div_addnode.innerHTML = x.opts_manipulation.tab_add_node;
      el_id.appendChild(div_addnode);

      var div_editnode = document.createElement('div');
      div_editnode.id = 'editnode-popUp';
      div_editnode.classList.add('network-popUp');
      div_editnode.innerHTML = x.opts_manipulation.tab_edit_node;
      el_id.appendChild(div_editnode);
      
      var div_editedge = document.createElement('div');
      div_editedge.id = 'editedge-popUp';
      div_editedge.classList.add('network-popUp');
      div_editedge.innerHTML = x.opts_manipulation.tab_edit_edge;
      el_id.appendChild(div_editedge);
      
      if(x.options.manipulation.addNode === undefined){
        options.manipulation.addNode = function(data, callback) {
          document.getElementById('addnode-operation').innerHTML = "Add Node";
          for (var nodecol = 0; nodecol < x.opts_manipulation.addNodeCols.length; nodecol++){
            document.getElementById('addnode-' + x.opts_manipulation.addNodeCols[nodecol]).value = data[x.opts_manipulation.addNodeCols[nodecol]];
          }
          document.getElementById('addnode-saveButton').onclick = saveNode.bind(this, data, callback, "addNode");
          document.getElementById('addnode-cancelButton').onclick = clearPopUp.bind();
          document.getElementById('addnode-popUp').style.display = 'block';
        };
      } else if(typeof(x.options.manipulation.addNode) === typeof(true)){
        if(x.options.manipulation.addNode){
          options.manipulation.addNode = function(data, callback) {
            document.getElementById('addnode-operation').innerHTML = "Add Node";
            for (var nodecol = 0; nodecol < x.opts_manipulation.addNodeCols.length; nodecol++){
              document.getElementById('addnode-' + x.opts_manipulation.addNodeCols[nodecol]).value = data[x.opts_manipulation.addNodeCols[nodecol]];
            }
            document.getElementById('addnode-saveButton').onclick = saveNode.bind(this, data, callback, "addNode");
            document.getElementById('addnode-cancelButton').onclick = clearPopUp.bind();
            document.getElementById('addnode-popUp').style.display = 'block';
          };
        } else {
          options.manipulation.addNode = false;
        }
      } else {
        options.manipulation.addNode = x.options.manipulation.addNode;
      }

      if(x.options.manipulation.editNode === undefined){
        options.manipulation.editNode = function(data, callback) {
            var node_data = nodes.get(data.id);
            document.getElementById('editnode-operation').innerHTML = "Edit Node";
            for (var nodecol = 0; nodecol < x.opts_manipulation.editNodeCols.length; nodecol++){
              document.getElementById('editnode-' + x.opts_manipulation.editNodeCols[nodecol]).value = node_data[x.opts_manipulation.editNodeCols[nodecol]];
            }
            document.getElementById('editnode-saveButton').onclick = saveNode.bind(this, data, callback, "editNode");
            document.getElementById('editnode-cancelButton').onclick = cancelEdit.bind(this,callback);
            document.getElementById('editnode-popUp').style.display = 'block';
          };
      } else if(typeof(x.options.manipulation.editNode) === typeof(true)){
        if(x.options.manipulation.editNode){
          options.manipulation.editNode = function(data, callback) {
            var node_data = nodes.get(data.id);
            document.getElementById('editnode-operation').innerHTML = "Edit Node";
            for (var nodecol = 0; nodecol < x.opts_manipulation.editNodeCols.length; nodecol++){
              document.getElementById('editnode-' + x.opts_manipulation.editNodeCols[nodecol]).value = node_data[x.opts_manipulation.editNodeCols[nodecol]];
            }
            document.getElementById('editnode-saveButton').onclick = saveNode.bind(this, data, callback, "editNode");
            document.getElementById('editnode-cancelButton').onclick = cancelEdit.bind(this,callback);
            document.getElementById('editnode-popUp').style.display = 'block';
            };
        } else {
          options.manipulation.editNode = false;
        }
      } else {
        options.manipulation.editNode = x.options.manipulation.editNode;
      }
  
      if(x.options.manipulation.deleteNode === undefined){
        options.manipulation.deleteNode = function(data, callback) {
            var r = confirm("Do you want to delete " + data.nodes.length + " node(s) and " + data.edges.length + " edges ?");
            if (r === true) {
              deleteSubGraph(data, callback);
            } else { clearPopUp(); callback(null); }
        };
      } else if(typeof(x.options.manipulation.deleteNode) === typeof(true)){
        if(x.options.manipulation.deleteNode){
          options.manipulation.deleteNode = function(data, callback) {
              var r = confirm("Do you want to delete " + data.nodes.length + " node(s) and " + data.edges.length + " edges ?");
              if (r === true) {
                deleteSubGraph(data, callback);
              } else { clearPopUp(); callback(null); }
          };
        } else {
          options.manipulation.deleteNode = false;
        }
      } else {
        options.manipulation.deleteNode = x.options.manipulation.deleteNode;
      }

      if(x.options.manipulation.deleteEdge === undefined){
        options.manipulation.deleteEdge = function(data, callback) {
            var r = confirm("Do you want to delete " + data.edges.length + " edges ?");
            if (r === true) {
              deleteSubGraph(data, callback);
            } else { clearPopUp(); callback(null); }
        };
      } else if(typeof(x.options.manipulation.deleteEdge) === typeof(true)){
        if(x.options.manipulation.deleteEdge){
          options.manipulation.deleteEdge = function(data, callback) {
              var r = confirm("Do you want to delete " + data.edges.length + " edges ?");
              if (r === true) {
                deleteSubGraph(data, callback);
              } else { clearPopUp(); callback(null); }
          };
        } else {
          options.manipulation.deleteEdge = false;
        }
      } else {
        options.manipulation.deleteEdge = x.options.manipulation.deleteEdge;
      }

      if(x.options.manipulation.addEdge === undefined){
        options.manipulation.addEdge = function(data, callback) {
          if (data.from == data.to) {
            var r = confirm("Do you want to connect the node to itself?");
            if (r === true) {
              saveEdge(data, callback, "addEdge");
            }
          }
          else {
            saveEdge(data, callback, "addEdge");
          }
        };
      } else if(typeof(x.options.manipulation.addEdge) === typeof(true)){
        if(x.options.manipulation.addEdge){
          options.manipulation.addEdge = function(data, callback) {
            if (data.from == data.to) {
              var r = confirm("Do you want to connect the node to itself?");
              if (r === true) {
                saveEdge(data, callback, "addEdge");
              }
            }
            else {
              saveEdge(data, callback, "addEdge");
            }
          };
        } else {
          options.manipulation.addEdge = false;
        }
      } else {
        options.manipulation.addEdge = x.options.manipulation.addEdge;
      }

      if(x.options.manipulation.editEdge === undefined){
          if(x.opts_manipulation.tab_edit_edge){
            options.manipulation.editEdge = {editWithoutDrag : function(data, callback) {
              var edge_data = edges.get(data.id);
              document.getElementById('editedge-operation').innerHTML = "Edit Edge";
              for (var edgecol = 0; edgecol < x.opts_manipulation.editEdgeCols.length; edgecol++){
                document.getElementById('editedge-' + x.opts_manipulation.editEdgeCols[edgecol]).value = edge_data[x.opts_manipulation.editEdgeCols[edgecol]];
              }
              document.getElementById('editedge-saveButton').onclick = saveEdge.bind(this, data, callback, "editEdgeCols");
              document.getElementById('editedge-cancelButton').onclick = cancelEdit.bind(this,callback);
              document.getElementById('editedge-popUp').style.display = 'block';
            }
            }
          } else {
            options.manipulation.editEdge = function(data, callback) {
              if (data.from == data.to) {
                var r = confirm("Do you want to connect the node to itself?");
                if (r === true) {
                  saveEdge(data, callback, "editEdge");
                }
              }
              else {
                saveEdge(data, callback, "editEdge");
              }
            };
          }
      } else if(typeof(x.options.manipulation.editEdge) === typeof(true)){
        if(x.options.manipulation.editEdge){
          if(x.opts_manipulation.tab_edit_edge){
            options.manipulation.editEdge = {editWithoutDrag : function(data, callback) {
              var edge_data = edges.get(data.id);
              document.getElementById('editedge-operation').innerHTML = "Edit Edge";
              for (var edgecol = 0; edgecol < x.opts_manipulation.editEdgeCols.length; edgecol++){
                document.getElementById('editedge-' + x.opts_manipulation.editEdgeCols[edgecol]).value = edge_data[x.opts_manipulation.editEdgeCols[edgecol]];
              }
              document.getElementById('editedge-saveButton').onclick = saveEdge.bind(this, data, callback, "editEdgeCols");
              document.getElementById('editedge-cancelButton').onclick = cancelEdit.bind(this,callback);
              document.getElementById('editedge-popUp').style.display = 'block';
            }
            }
          } else {
            options.manipulation.editEdge = function(data, callback) {
              if (data.from == data.to) {
                var r = confirm("Do you want to connect the node to itself?");
                if (r === true) {
                  saveEdge(data, callback, "editEdge");
                }
              }
              else {
                saveEdge(data, callback, "editEdge");
              }
            };
          }
        } else {
          options.manipulation.editEdge = false;
        }
      } else {
        options.manipulation.editEdge = x.options.manipulation.editEdge;
      }
    }
    
    // create network
    instance.network = new vis.Network(document.getElementById("graph"+el.id), data, options);
    if (window.Shiny){
      Shiny.onInputChange(el.id + '_initialized', true);
    }
    
    //*************************
    //add values to idselection
    //*************************
    
    if(el_id.idselection){  
      var selectList = document.getElementById("nodeSelect"+el.id)
      setNodeIdList(selectList, x.idselection, nodes)
      
      if (window.Shiny){
        changeInput('selected', document.getElementById("nodeSelect"+el.id).value);
      }
    }
      
    //console.info(instance.network)  
    //save data for re-use and update
    document.getElementById("graph"+el.id).chart = instance.network;
    document.getElementById("graph"+el.id).options = options;
    
    /////////
    // popup
    /////////
    
    // Temporary variables to hold mouse x-y pos.s
    var tempX = 0
    var tempY = 0

    // Main function to retrieve mouse x-y pos.s
    function getMouseXY(e) {
      tempX = e.clientX
      tempY = e.clientY
      // catch possible negative values in NS
      if (tempX < 0){tempX = 0}
      if (tempY < 0){tempY = 0}
    }

    document.addEventListener('mousemove', getMouseXY);

   //this.body.emitter.emit("showPopup",{id:this.popupObj.id,x:t.x+3,y:t.y-5}))

    // popup for title
    var popupState = false;
    var popupTimeout = null;
    var vispopup = document.createElement("div");
   
    // disable vis.js tooltip 
    var style = document.createElement('style');
    style.type = 'text/css';
    style.innerHTML = 'div.vis-tooltip {display : none}';
    document.getElementsByTagName('head')[0].appendChild(style);

    var popupStyle = 'position: fixed;visibility:hidden;padding: 5px;font-family: verdana;font-size:14px;font-color:#000000;background-color: #f5f4ed;-moz-border-radius: 3px;-webkit-border-radius: 3px;border-radius: 3px;border: 1px solid #808074;box-shadow: 3px 3px 10px rgba(0, 0, 0, 0.2);max-width:400px;word-break: break-all'
    
    if(x.tooltipStyle !== undefined){
      popupStyle = x.tooltipStyle
    }
    var popupStay = 300;
    if(x.tooltipStay !== undefined){
      popupStay = x.tooltipStay
    }
    vispopup.setAttribute('style', popupStyle)
    
    document.getElementById("graph"+el.id).appendChild(vispopup);
    
    // add some event listeners to avoid it disappearing when the mouse if over it.
    vispopup.addEventListener('mouseover',function () {
      if (popupTimeout !== null) {
        clearTimeout(popupTimeout);
        popupTimeout = null;
      }
    });
  
    // set the timeout when the mouse leaves it.
    vispopup.addEventListener('mouseout',function () {
      if (popupTimeout === null) {
        myHidePopup(100);
      }
    });
    
    // use the popup event to show
    instance.network.on("showPopup", function(params) {
      popupState = true;  
      myShowPopup(params);
    })
  
    // use the hide event to hide it
    instance.network.on("hidePopup", function(params) {
      // avoid double firing of this event, bug in 4.2.0
      if (popupState === true) {
        popupState = false;
        myHidePopup(popupStay);
      }
    })
  
    // hiding the popup through css and a timeout
    function myHidePopup(delay) {
      popupTimeout = setTimeout(function() {vispopup.style.visibility = 'hidden';}, delay);
    }
  
    // showing the popup
    function myShowPopup(id) {
      // get the data from the vis.DataSet
      var nodeData = nodes.get([id]);
      var edgeData = edges.get([id]);
      
      // a node ?
      if(nodeData[0] !== null && nodeData[0] !== undefined){
        vispopup.innerHTML = nodeData[0].title;
        // show and place the tooltip.
        vispopup.style.visibility = 'visible';
        vispopup.style.top = tempY - 20 +  "px";
        vispopup.style.left = tempX + 5 + "px";
        
      } else if(edgeData[0] !== null && edgeData[0] !== undefined){
        // so it's perhaps a edge ?
        vispopup.innerHTML = edgeData[0].title;
        // show and place the tooltip.
        vispopup.style.visibility = 'visible';
        vispopup.style.top = tempY - 20 +  "px";
        vispopup.style.left = tempX + 5 + "px";
      } else {
        // or a cluster ?
        var node_cluster = instance.network.body.nodes[id]
        if(node_cluster !== undefined){
          vispopup.innerHTML = node_cluster.options.title;
          // show and place the tooltip.
          vispopup.style.visibility = 'visible';
          vispopup.style.top = tempY - 20 +  "px";
          vispopup.style.left = tempX + 5 + "px";
        }
      }
      
      // for sparkline. Eval script...
      var any_script= vispopup.getElementsByTagName('script')
      for (var n = 0; n < any_script.length; n++){
        if(any_script[n].getAttribute("type") === "text/javascript"){
           eval(any_script[n].innerHTML);
        }
      }
    }
  
    //*************************
    // Events
    //*************************
    if(x.events !== undefined){
      for (var key in x.events) {
          instance.network.on(key, x.events[key]);
      }
    }

    if(x.OnceEvents !== undefined){
      for (var key in x.OnceEvents) {
          instance.network.once(key, x.OnceEvents[key]);
      }
    }
    
    if(x.ResetEvents !== undefined){
      for (var key in x.ResetEvents) {
          instance.network.off(key);
      }
    }
    //*************************
    // Selected Highlight
    //*************************

    function selectedHighlight(value) {
      // get current nodes
      var allNodes = nodes.get({returnType:"Object"});
       
      // first resetEdges
      resetAllEdges(edges, el_id.byselectionColor, el_id.highlightColor, instance.network);
      var connectedNodes = [];  
      
      // get variable
      var sel = el_id.byselection_variable;
      // need to make an update?
      var update = !(el_id.selectActive === false && value === "");

      if (value !== "") {
        var updateArray = [];
        el_id.selectActive = true;
        
        // mark all nodes as hard to read.
        for (var nodeId in allNodes) {
          var value_in = false;
          // unique selection
          if(el_id.byselection_multiple === false){
            if(sel == "label"){
              value_in = ((allNodes[nodeId]["label"] + "") === value) || ((allNodes[nodeId]["hiddenLabel"] + "") === value);
            }else if(sel == "color"){
              value_in = ((allNodes[nodeId]["color"] + "") === value) || ((allNodes[nodeId]["hiddenColor"] + "") === value);
            }else {
              value_in = (allNodes[nodeId][sel] + "") === value;
            }
          }else{ // multiple selection
            if(sel == "label"){
              var current_value = allNodes[nodeId]["label"] + "";
              var value_split = current_value.split(",").map(Function.prototype.call, String.prototype.trim);
              var current_value2 = allNodes[nodeId]["hiddenLabel"] + "";
              var value_split2 = current_value.split(",").map(Function.prototype.call, String.prototype.trim);
              value_in = (value_split.indexOf(value) !== -1) || (value_split2.indexOf(value) !== -1);
            }else if(sel == "color"){
              var current_value = allNodes[nodeId]["color"] + "";
              var value_split = current_value.split(",").map(Function.prototype.call, String.prototype.trim);
              var current_value2 = allNodes[nodeId]["hiddenColor"] + "";
              var value_split2 = current_value.split(",").map(Function.prototype.call, String.prototype.trim);
              value_in = (value_split.indexOf(value) !== -1) || (value_split2.indexOf(value) !== -1);
            }else {
              var current_value = allNodes[nodeId][sel] + "";
              var value_split = current_value.split(",").map(Function.prototype.call, String.prototype.trim);
              value_in = value_split.indexOf(value) !== -1;
            }
          }
          if(value_in === false){ // not in selection, so as hard to read
            nodeAsHardToRead(allNodes[nodeId], options, el_id.byselectionColor, el_id.highlightColor, instance.network, "node");
          } else { // in selection, so reset if needed
            connectedNodes = connectedNodes.concat(allNodes[nodeId].id);
            resetOneNode(allNodes[nodeId], options, instance.network);
          }
          allNodes[nodeId].x = undefined;
          allNodes[nodeId].y = undefined;
          // update data
          if (allNodes.hasOwnProperty(nodeId) && update) {
            updateArray.push(allNodes[nodeId]);
          }
        }
        if(update){
          // set some edges as hard to read
          var edgesHardToRead = edges.get({
            fields: ['id', 'color', 'hiddenColor', 'hiddenLabel', 'label'],
            filter: function (item) {
              return (indexOf.call(connectedNodes, item.from, true) === -1) ;
            },
            returnType :'Array'
          });

          // all in degree nodes get their own color and their label back
          for (i = 0; i < edgesHardToRead.length; i++) {
            edgeAsHardToRead(edgesHardToRead[i], el_id.byselectionColor, el_id.highlightColor, instance.network, type = "edge")
          }
          edges.update(edgesHardToRead);
            
          nodes.update(updateArray);
          
          // select for highlight
          if(el_id.highlight && x.nodes && el_id.byselectionHighlight){
              neighbourhoodHighlight(connectedNodes, "click", el_id.highlightAlgorithm, false);
              instance.network.selectNodes(connectedNodes)
          }
        }
      }
      else if (el_id.selectActive === true) {
        //reset nodes
        resetAllNodes(nodes, update, options, instance.network, false)
        el_id.selectActive = false
      }
    } 
  
    //*************************
    //Highlight
    //*************************
    var is_hovered = false;
    var is_clicked = false;
    
    function neighbourhoodHighlight(params, action_type, algorithm, reset_selectedBy) {

      var nodes_in_clusters = instance.network.body.modules.clustering.clusteredNodes;
      var have_cluster_nodes = false;
      if(Object.keys(nodes_in_clusters).length > 0){
        have_cluster_nodes = true;
        nodes_in_clusters = Object.keys(nodes_in_clusters);
        edges_in_clusters = Object.keys(instance.network.body.modules.clustering.clusteredEdges);
      } else {
        nodes_in_clusters = [];
        edges_in_clusters = [];
      }
      
      var selectNode;
      // get nodes data
      var allNodes = nodes.get({returnType:"Object"});

      // cluster
      var array_cluster_id;
      
      // update 
      var update = !(el_id.highlightActive === false && params.length === 0) | (el_id.selectActive === true && params.length === 0);

      if(!(action_type == "hover" && is_clicked)){
        
        // first resetEdges
        resetAllEdges(edges, el_id.highlightColor, el_id.byselectionColor, instance.network);

        if (params.length > 0) {
          var is_cluster = instance.network.isCluster(params[0]);
          var selectedNode;
          
          if(is_cluster){
            selectedNode = instance.network.getNodesInCluster(params[0]);
          } else {
            selectedNode = params;
          }

          var updateArray = [];
          if(el_id.idselection){
            selectNode = document.getElementById('nodeSelect'+el.id);
            if(is_cluster === false){
              if(x.idselection.values !== undefined){
                if(indexOf.call(x.idselection.values, selectedNode[0], true) > -1){
                  selectNode.value = selectedNode[0];
                }else{
                  selectNode.value = "";
                }
              }else{
                selectNode.value = selectedNode[0];
              }
              if (window.Shiny){
                changeInput('selected', selectNode.value);
              }
            }
          }
          
          el_id.highlightActive = true;
          var i,j;
          var degrees = el_id.degree;
          
          // mark all nodes as hard to read.
          for (var nodeId in instance.network.body.nodes) {
            if(instance.network.isCluster(nodeId)){
              nodeAsHardToRead(instance.network.body.nodes[nodeId], options, el_id.highlightColor, el_id.byselectionColor, instance.network, "cluster");
            }else {
              var tmp_node = allNodes[nodeId];
              if(tmp_node !== undefined){
                nodeAsHardToRead(tmp_node, options, el_id.highlightColor, el_id.byselectionColor, instance.network, "node");
                tmp_node.x = undefined;
                tmp_node.y = undefined;
              }
            }
          }
 
          if(algorithm === "all"){
            var connectedNodes;
            if(degrees > 0){
              connectedNodes = [];
              for (j = 0; j < selectedNode.length; j++) {
                connectedNodes = connectedNodes.concat(instance.network.getConnectedNodes(selectedNode[j], true));
              }
              connectedNodes = uniqueArray(connectedNodes, true, instance.network);
            }else{
              connectedNodes = selectedNode;
            }
            
            var allConnectedNodes = [];
            // get the nodes to color
            if(degrees >= 2){
              for (i = 2; i <= degrees; i++) {
                var previous_connectedNodes = connectedNodes;
                var currentlength = connectedNodes.length;
                for (j = 0; j < currentlength; j++) {
                  connectedNodes = uniqueArray(connectedNodes.concat(instance.network.getConnectedNodes(connectedNodes[j])), true, instance.network);
                }
                if (connectedNodes.length === previous_connectedNodes.length) { break; }
              }
            }
            // nodes to just label
            for (j = 0; j < connectedNodes.length; j++) {
                allConnectedNodes = allConnectedNodes.concat(instance.network.getConnectedNodes(connectedNodes[j]));
            }
            allConnectedNodes = uniqueArray(allConnectedNodes, true, instance.network);

            if(el_id.highlightLabelOnly === true){
              // all higher degree nodes get a different color and their label back
              array_cluster_id = [];
              for (i = 0; i < allConnectedNodes.length; i++) {
                if (allNodes[allConnectedNodes[i]].hiddenLabel !== undefined) {
                  allNodes[allConnectedNodes[i]].label = allNodes[allConnectedNodes[i]].hiddenLabel;
                  allNodes[allConnectedNodes[i]].hiddenLabel = undefined;
                  if(have_cluster_nodes){
                    if(indexOf.call(nodes_in_clusters, allConnectedNodes[i], true) > -1){
  
                      array_cluster_id = array_cluster_id.concat(instance.network.clustering.findNode(allConnectedNodes[i])[0]);
                    }
                  }
                }
              }
  
              if(array_cluster_id.length > 0){
                array_cluster_id = uniqueArray(array_cluster_id, false, instance.network);
                for (i = 0; i < array_cluster_id.length; i++) {
                  instance.network.body.nodes[array_cluster_id[i]].setOptions({label : instance.network.body.nodes[array_cluster_id[i]].options.hiddenLabel, hiddenLabel:undefined})
                }
              }
            }

            // all in degree nodes get their own color and their label back + main nodes
            connectedNodes = connectedNodes.concat(selectedNode);
            
            if (window.Shiny){
              Shiny.onInputChange(el.id + '_highlight_color_id', uniqueShiny(connectedNodes));
            }
            if(el_id.highlightLabelOnly === true){
              if (window.Shiny){
                Shiny.onInputChange(el.id + '_highlight_label_id', allConnectedNodes.filter(function(x){ return !connectedNodes.includes(x)}));
              }
            }  
   
            array_cluster_id = [];
            for (i = 0; i < connectedNodes.length; i++) {
              resetOneNode(allNodes[connectedNodes[i]], options, instance.network);
              if(have_cluster_nodes){
                if(indexOf.call(nodes_in_clusters, connectedNodes[i], true) > -1){
                  array_cluster_id = array_cluster_id.concat(instance.network.clustering.findNode(connectedNodes[i])[0]);
                }
              }
            }
            
            if(array_cluster_id.length > 0){
              array_cluster_id = uniqueArray(array_cluster_id, false, instance.network);
              for (i = 0; i < array_cluster_id.length; i++) {
                resetOneCluster(instance.network.body.nodes[array_cluster_id[i]], options, instance.network);
              }
            }
            
            // set some edges as hard to read
            var edgesHardToRead = edges.get({
              fields: ['id', 'color', 'hiddenColor', 'hiddenLabel', 'label'],
              filter: function (item) {
                return ((indexOf.call(connectedNodes, item.from, true) === -1)) ;
              },
              returnType :'Array'
            });

            // all in degree nodes get their own color and their label back
            array_cluster_id = [];
            var tmp_cluster_id;
            for (i = 0; i < edgesHardToRead.length; i++) {
              edgeAsHardToRead(edgesHardToRead[i], el_id.highlightColor, el_id.byselectionColor, instance.network, type = "edge")
              if(have_cluster_nodes){
                if(indexOf.call(edges_in_clusters, edgesHardToRead[i].id, true) > -1){
                  tmp_cluster_id = instance.network.clustering.getClusteredEdges(edgesHardToRead[i].id);
                  if(tmp_cluster_id.length > 1){
                    array_cluster_id = array_cluster_id.concat(tmp_cluster_id[0]);
                  }
                }
              }
            }
            
            if(array_cluster_id.length > 0){
              array_cluster_id = uniqueArray(array_cluster_id, false, instance.network);
              for (i = 0; i < array_cluster_id.length; i++) {
                edgeAsHardToRead(instance.network.body.edges[array_cluster_id[i]].options, el_id.highlightColor, el_id.byselectionColor, instance.network, type = "cluster")
              }
            }
            edges.update(edgesHardToRead);
          } else if(algorithm === "hierarchical"){
            
            var degree_from = degrees.from;
            var degree_to = degrees.to;
            degrees = Math.max(degree_from, degree_to);
            
            var allConnectedNodes = [];
            var currentConnectedFromNodes = [];
            var currentConnectedToNodes = [];
            var connectedFromNodes = [];
            var connectedToNodes = [];
            
            if(degree_from > 0){
              connectedFromNodes = edges.get({
                fields: ['from'],
                filter: function (item) {
                  return ((indexOf.call(selectedNode, item.to, true) !== -1)) ;
                },
                returnType :'Array'
              });
            }

            if(degree_to > 0){
              connectedToNodes = edges.get({
                fields: ['to'],
                filter: function (item) {
                  return ((indexOf.call(selectedNode, item.from, true) !== -1)) ;
                },
                returnType :'Array'
              });
            }
            for (j = 0; j < connectedFromNodes.length; j++) {
                allConnectedNodes = allConnectedNodes.concat(connectedFromNodes[j].from);
                currentConnectedFromNodes = currentConnectedFromNodes.concat(connectedFromNodes[j].from);
            }
            
            for (j = 0; j < connectedToNodes.length; j++) {
                allConnectedNodes = allConnectedNodes.concat(connectedToNodes[j].to);
                currentConnectedToNodes = currentConnectedToNodes.concat(connectedToNodes[j].to);
            }
            
            var go_from;
            var go_to;
                
            if(degrees > 1){
              for (i = 2; i <= degrees; i++) {
                go_from = false;
                go_to = false;
                if(currentConnectedFromNodes.length > 0 && i <= degree_from){
                  connectedFromNodes = edges.get({
                    fields: ['from'],
                    filter: function (item) {
                      return indexOf.call(currentConnectedFromNodes, item.to, true) > -1;
                    },
                    returnType :'Array'
                  });
                  go_from = true;
                }

                if(currentConnectedToNodes.length > 0 && i <= degree_to){
                  connectedToNodes = edges.get({
                    fields: ['to'],
                    filter: function (item) {
                      return indexOf.call(currentConnectedToNodes, item.from, true) > -1;
                    },
                    returnType :'Array'
                  });
                  go_to = true;
                }

                if(go_from === true){
                  currentConnectedFromNodes = [];
                  for (j = 0; j < connectedFromNodes.length; j++) {
                    allConnectedNodes = allConnectedNodes.concat(connectedFromNodes[j].from);
                    currentConnectedFromNodes = currentConnectedFromNodes.concat(connectedFromNodes[j].from);
                  }
                }

                if(go_to === true){
                  currentConnectedToNodes = [];
                  for (j = 0; j < connectedToNodes.length; j++) {
                    allConnectedNodes = allConnectedNodes.concat(connectedToNodes[j].to);
                    currentConnectedToNodes = currentConnectedToNodes.concat(connectedToNodes[j].to);
                  } 
                }
                
                if (go_from === false &&  go_to === false) { break;}
              }
            }
            
            allConnectedNodes = uniqueArray(allConnectedNodes, true, instance.network).concat(selectedNode);

            var nodesWithLabel = [];
            if(el_id.highlightLabelOnly === true){
              if(degrees > 0){
                // nodes to just label
                for (j = 0; j < currentConnectedToNodes.length; j++) {
                    nodesWithLabel = nodesWithLabel.concat(instance.network.getConnectedNodes(currentConnectedToNodes[j]));
                }
                
                for (j = 0; j < currentConnectedFromNodes.length; j++) {
                    nodesWithLabel = nodesWithLabel.concat(instance.network.getConnectedNodes(currentConnectedFromNodes[j]));
                }
                nodesWithLabel = uniqueArray(nodesWithLabel, true, instance.network);
              } else{
                nodesWithLabel = currentConnectedToNodes;
                nodesWithLabel = nodesWithLabel.concat(currentConnectedFromNodes);
                nodesWithLabel = uniqueArray(nodesWithLabel, true, instance.network);
              }
            }
            // all higher degree nodes get a different color and their label back
            array_cluster_id = [];
            for (i = 0; i < nodesWithLabel.length; i++) {
              if (allNodes[nodesWithLabel[i]].hiddenLabel !== undefined) {
                allNodes[nodesWithLabel[i]].label = allNodes[nodesWithLabel[i]].hiddenLabel;
                allNodes[nodesWithLabel[i]].hiddenLabel = undefined;
                if(have_cluster_nodes){
                  if(indexOf.call(nodes_in_clusters, nodesWithLabel[i], true) > -1){
                    array_cluster_id = array_cluster_id.concat(instance.network.clustering.findNode(nodesWithLabel[i])[0]);
                  }
                }
              }
            }
            
            if(array_cluster_id.length > 0){
              array_cluster_id = uniqueArray(array_cluster_id, false, instance.network);
              for (i = 0; i < array_cluster_id.length; i++) {
                instance.network.body.nodes[array_cluster_id[i]].setOptions({label : instance.network.body.nodes[array_cluster_id[i]].options.hiddenLabel, hiddenLabel:undefined})
              }
            }

            // all in degree nodes get their own color and their label back
            array_cluster_id = [];
            for (i = 0; i < allConnectedNodes.length; i++) {
              resetOneNode(allNodes[allConnectedNodes[i]], options, instance.network);
              if(have_cluster_nodes){
                if(indexOf.call(nodes_in_clusters, allConnectedNodes[i], true) > -1){
                  array_cluster_id = array_cluster_id.concat(instance.network.clustering.findNode(allConnectedNodes[i])[0]);
                }
              }
            }
            
            if(array_cluster_id.length > 0){
              array_cluster_id = uniqueArray(array_cluster_id, false, instance.network);
              for (i = 0; i < array_cluster_id.length; i++) {
                 resetOneCluster(instance.network.body.nodes[array_cluster_id[i]], options, instance.network);
              }
            }
             
            if (window.Shiny){ 
              Shiny.onInputChange(el.id + '_highlight_color_id', uniqueShiny(allConnectedNodes));
            }
            if(el_id.highlightLabelOnly === true){
              if (window.Shiny){
                Shiny.onInputChange(el.id + '_highlight_label_id', nodesWithLabel.filter(function(x) {return !allConnectedNodes.includes(x)}));
              }
            }  
            
            // set some edges as hard to read
            var edgesHardToRead = edges.get({
              fields: ['id', 'color', 'hiddenColor', 'hiddenLabel', 'label'],
              filter: function (item) {
                return ((indexOf.call(allConnectedNodes, item.from, true) === -1)  || (indexOf.call(allConnectedNodes, item.to, true) === -1)) ;
              },
              returnType :'Array'
            });

            array_cluster_id = [];
            for (i = 0; i < edgesHardToRead.length; i++) {
              edgeAsHardToRead(edgesHardToRead[i], el_id.highlightColor, el_id.byselectionColor, instance.network, type = "edge")
              if(have_cluster_nodes){
                if(indexOf.call(edges_in_clusters, edgesHardToRead[i].id, true) > -1){
                  var tmp_cluster_id = instance.network.clustering.getClusteredEdges(edgesHardToRead[i].id);
                  if(tmp_cluster_id.length > 1){
                    array_cluster_id = array_cluster_id.concat(tmp_cluster_id[0]);
                  }
                }
              }
            }

            if(array_cluster_id.length > 0){
              array_cluster_id = uniqueArray(array_cluster_id, false, instance.network);
              for (i = 0; i < array_cluster_id.length; i++) {
                 edgeAsHardToRead(instance.network.body.edges[array_cluster_id[i]].options, el_id.highlightColor, el_id.byselectionColor, instance.network, type = "cluster");
              }
            }
            
            edges.update(edgesHardToRead);
            
          }

          if(update){
            if(!(action_type == "hover")){
               is_clicked = true;
            }
            // transform the object into an array
            var updateArray = [];
            for (nodeId in allNodes) {
              if (allNodes.hasOwnProperty(nodeId)) {
                updateArray.push(allNodes[nodeId]);
              }
            }
            nodes.update(updateArray);
          }else{
            is_clicked = false;
          }
        
        }
        else if (el_id.highlightActive === true | el_id.selectActive === true) {
          // reset nodeSelect list if actived
          if(el_id.idselection){
            resetList("nodeSelect", el.id, 'selected');
          }
          //reset nodes
          resetAllNodes(nodes, update, options, instance.network, false)
          el_id.highlightActive = false;
          is_clicked = false;
          
          if (window.Shiny){
            Shiny.onInputChange(el.id + '_highlight_label_id', null)
            Shiny.onInputChange(el.id + '_highlight_color_id', null)
          }
        }
      }
      // reset selectedBy list if actived
      if(el_id.byselection && reset_selectedBy){
        resetList("selectedBy", el.id, 'selectedBy');
      }
    }
    
    function onClickIDSelection(selectedItems) {
      var selectNode;
      if(el_id.idselection){
        if (selectedItems.nodes.length !== 0) {
          selectNode = document.getElementById('nodeSelect'+el.id);
          if(x.idselection.values !== undefined){
            if(indexOf.call(x.idselection.values, selectedItems.nodes[0], true) > -1){
              selectNode.value = selectedItems.nodes;
            }else{
              selectNode.value = "";
            }
          }else{
            selectNode.value = selectedItems.nodes;
          }
          if (window.Shiny){
            changeInput('selected', selectNode.value);
          }
        }else{
          resetList("nodeSelect", el.id, 'selected');
        } 
      }
      if(el_id.byselection){
        // reset selectedBy list if actived
        if (selectedItems.nodes.length === 0) {
          resetList("selectedBy", el.id, 'selectedBy');
          selectedHighlight("");
        }
      }
    }
    
    // shared click function (selectedNodes)
    document.getElementById("graph"+el.id).myclick = function(params){
        if(el_id.highlight && x.nodes){
          neighbourhoodHighlight(params.nodes, "click", el_id.highlightAlgorithm, true);
        }else if((el_id.idselection || el_id.byselection) && x.nodes){
          onClickIDSelection(params)
        }
    };
    
    // Set event in relation with highlightNearest      
    instance.network.on("click", function(params){
        if(el_id.highlight && x.nodes){
          neighbourhoodHighlight(params.nodes, "click", el_id.highlightAlgorithm, true);
        }else if((el_id.idselection || el_id.byselection) && x.nodes){
          onClickIDSelection(params)
        } 
    });
    
    instance.network.on("hoverNode", function(params){
      if(el_id.hoverNearest && x.nodes){
        neighbourhoodHighlight([params.node], "hover", el_id.highlightAlgorithm, true);
      } 
    });

    instance.network.on("blurNode", function(params){
      if(el_id.hoverNearest && x.nodes){
        neighbourhoodHighlight([], "hover", el_id.highlightAlgorithm, true);
      }      
    });
    
    //*************************
    //collapse
    //*************************
    instance.network.on("doubleClick", function(params){
      if(el_id.collapse){
        collapsedNetwork(params.nodes, el_id.collapseFit, el_id.collapseResetHighlight, 
          el_id.clusterOptions, el_id.collapseLabelSuffix,
          el_id.tree, instance.network, el.id) 
      }
    }); 
    
    if(el_id.collapse){
      instance.network.on("doubleClick", networkOpenCluster);
    }
    
    //*************************
    //footer
    //*************************
    var div_footer = document.createElement('div');
    div_footer.id = "footer"+el.id;
    div_footer.setAttribute('style',  'font-family:Georgia, Times New Roman, Times, serif;font-size:12px;text-align:center;background-color: inherit;');
    div_footer.style.display = 'none';

    document.getElementById("graph" + el.id).appendChild(div_footer);  
    if(x.footer !== null){
      div_footer.innerHTML = x.footer.text;
      div_footer.setAttribute('style',  x.footer.style + ';background-color: inherit;');
      div_footer.style.display = 'block'; 
    }
    
    //*************************
    // export
    //*************************
    if(x.export !== undefined){
      
      var downloaddiv = document.createElement('div');
      downloaddiv.setAttribute('style', 'float:right; width:100%;background-color: inherit;');
      
      var downloadbutton = document.createElement("button");
      downloadbutton.setAttribute('style', x.export.css);
      downloadbutton.style.position = "relative";
      downloadbutton.id = "download"+el.id;
      downloadbutton.appendChild(document.createTextNode(x.export.label)); 
      downloaddiv.appendChild(downloadbutton);
      
      var hr = document.createElement("hr");
      hr.setAttribute('style', 'height:5px; visibility:hidden; margin-bottom:-1px;');
      downloaddiv.appendChild(hr);  
      
      document.getElementById("maindiv"+el.id).appendChild(downloaddiv);
      
      document.getElementById("download"+el.id).onclick = function() {

      // height control for export
      var addHeightExport = document.getElementById("graph" + el.id).offsetHeight + idList.offsetHeight + byList.offsetHeight + downloaddiv.offsetHeight;
      if(div_title.style.display !== 'none'){
        addHeightExport = addHeightExport + div_title.offsetHeight;
      }
      if(div_subtitle.style.display !== 'none'){
        addHeightExport = addHeightExport + div_subtitle.offsetHeight;
      }
      if(div_footer.style.display !== 'none'){
        addHeightExport = addHeightExport + div_footer.offsetHeight;
      } else {
        addHeightExport = addHeightExport + 15;
      }

      downloadbutton.style.display = 'none';
      var export_background = x.export.background;
      if(x.background !== "transparent" && x.background !== "rgba(0, 0, 0, 0)"){
        export_background = x.background
      }
      
      if(x.export.type !== "pdf"){
        html2canvas(el_id, {
          background: export_background,
          height : addHeightExport,
          onrendered: function(canvas) {
            canvas.toBlobHD(function(blob) {
              saveAs(blob, x.export.name);
            }, "image/"+x.export.type);
          }
        });
      } else {
        html2canvas(el_id, {
          background: export_background,
          height : addHeightExport,
          onrendered: function(canvas) {
            var myImage = canvas.toDataURL("image/png", 1.0);
            //var imgWidth = (canvas.width * 25.4) / 24;
            //var imgHeight = (canvas.height * 25.4) / 24; 
            var table = new jsPDF('l', 'pt', [canvas.width, canvas.height]);
            table.addImage(myImage, 'JPEG', 0, 0, canvas.width, canvas.height);
            table.save(x.export.name);
          } 
        });
      }

      downloadbutton.style.display = 'block';
      };
    }

    //*************************
    // dataManipulation
    //*************************
    function clearPopUp() {
      if(x.opts_manipulation.tab_add_node){
        document.getElementById('addnode-saveButton').onclick = null;
        document.getElementById('addnode-cancelButton').onclick = null;
        document.getElementById('addnode-popUp').style.display = 'none';
      }

      if(x.opts_manipulation.tab_edit_node){
        document.getElementById('editnode-saveButton').onclick = null;
        document.getElementById('editnode-cancelButton').onclick = null;
        document.getElementById('editnode-popUp').style.display = 'none';
      }
      
      if(x.opts_manipulation.tab_edit_edge){
        document.getElementById('editedge-saveButton').onclick = null;
        document.getElementById('editedge-cancelButton').onclick = null;
        document.getElementById('editedge-popUp').style.display = 'none';
      }
    }

    function saveNode(data, callback, cmd) {
      var iname;
      var prediv;
      if(cmd === "addNode"){
        iname = "addNodeCols";
        prediv = 'addnode-';
      } else  {
        iname = "editNodeCols";
        prediv = 'editnode-';
      }
      var obj = {id : data.id}
      for (var nodecol = 0; nodecol < x.opts_manipulation[iname].length; nodecol++){
        var add_node_val = document.getElementById(prediv + x.opts_manipulation[iname][nodecol]).value;
        var add_node_type = document.getElementById(prediv + x.opts_manipulation[iname][nodecol]).type;
        if(add_node_type && add_node_type === "number"){
          add_node_val = parseFloat(add_node_val)
        }
        if(add_node_val !== "undefined"){
          obj[x.opts_manipulation[iname][nodecol]] = add_node_val
        }
      }

      var update_obj = clone(obj);
      update_obj.x = data.x;
      update_obj.y = data.y;
      nodes.update(update_obj);
      
      if (window.Shiny){
        obj.cmd = cmd;
        Shiny.onInputChange(el.id + '_graphChange', obj);
      }
      clearPopUp();
      callback(null);
    }

    function saveEdge(data, callback, cmd) {
      if(cmd === "editEdge"){
        callback(data); //must be first called for egde id !
        if (window.Shiny){
          var obj = {cmd: cmd, id: data.id, from: data.from, to: data.to};
          Shiny.onInputChange(el.id + '_graphChange', obj);
        }
      } else if(cmd === "addEdge"){
        callback(data); //must be first called for egde id !
        if (window.Shiny){
          var obj = {cmd: cmd, id: data.id, from: data.from, to: data.to};
          Shiny.onInputChange(el.id + '_graphChange', obj);
        }
      } else if(cmd === "editEdgeCols"){
        for (var edgecol = 0; edgecol < x.opts_manipulation.editEdgeCols.length; edgecol++){
          var add_edge_val = document.getElementById("editedge-" + x.opts_manipulation.editEdgeCols[edgecol]).value;
          var add_edge_type = document.getElementById("editedge-" + x.opts_manipulation.editEdgeCols[edgecol]).type;
          if(add_edge_type && add_edge_type === "number"){
            add_edge_val = parseFloat(add_edge_val)
          }
          if(add_edge_val !== "undefined"){
            data[x.opts_manipulation.editEdgeCols[edgecol]] = add_edge_val
          }
        }
        if (window.Shiny){
          var obj = {cmd: "editEdge", id : data.id}
          for (var edgecol = 0; edgecol < x.opts_manipulation.editEdgeCols.length; edgecol++){
            if(data[x.opts_manipulation.editEdgeCols[edgecol]] !== "undefined"){
              obj[x.opts_manipulation.editEdgeCols[edgecol]] = data[x.opts_manipulation.editEdgeCols[edgecol]];
            }
          }
          Shiny.onInputChange(el.id + '_graphChange', obj);
        }
        callback(data);
        clearPopUp();
      }
    }

    function deleteSubGraph(data, callback) {
      if (window.Shiny){
        var obj = {cmd: "deleteElements", nodes: data.nodes, edges: data.edges}
        Shiny.onInputChange(el.id + '_graphChange', obj);
      }
      callback(data);
    }

    function cancelEdit(callback) {
      clearPopUp();
      callback(null);
    }
    
    //*************************
    // CLUSTERING
    //*************************
    if(x.clusteringGroup || x.clusteringColor || x.clusteringHubsize || x.clusteringConnection){
      
      var clusterbutton = document.createElement("input");
      clusterbutton.id = "backbtn"+el.id;
      clusterbutton.setAttribute('type', 'button');  
      clusterbutton.setAttribute('value', 'Reinitialize clustering'); 
      clusterbutton.setAttribute('style', 'background-color:#FFFFFF;border: none');
      el_id.appendChild(clusterbutton);
      
      clusterbutton.onclick =  function(){
        // reset some parameters / data before
        if (el_id.selectActive === true | el_id.highlightActive === true) {
          //reset nodes
          neighbourhoodHighlight([], "click", el_id.highlightAlgorithm, true);
          if (el_id.selectActive === true){
            el_id.selectActive = false;
            resetList('selectedBy',el.id, 'selectedBy');
          }
          if (el_id.highlightActive === true){
            el_id.highlightActive = false;
            resetList('nodeSelect', el.id, 'selected');
          }
        }
        //instance.network.setData(data);
        if(x.clusteringColor){
          clusterByColor();
        }
        if(x.clusteringGroup){
          clusterByGroup(x.clusteringGroup.groups);
        }
        if(x.clusteringHubsize){
          clusterByHubsize();
        }
        if(x.clusteringConnection){
          clusterByConnection();
        }
        instance.network.fit();
      }
    }
    
    if(x.clusteringGroup || x.clusteringColor || x.clusteringOutliers || x.clusteringHubsize || x.clusteringConnection){
      // if we click on a node, we want to open it up!
      instance.network.on("doubleClick", function (params){
        if (params.nodes.length === 1) {
          if (instance.network.isCluster(params.nodes[0]) === true) {
            is_clicked = false;
            instance.network.openCluster(params.nodes[0], {releaseFunction : function(clusterPosition, containedNodesPositions) {
              return containedNodesPositions;
            }});
            // must be better...
            resetAllEdges(edges, el_id.highlightColor, el_id.byselectionColor, instance.network);
            resetAllNodes(nodes, true, options, instance.network, true);
          } else {
            if(x.clusteringGroup){
              var array_group = nodes.get({
                fields: ['group'],
                filter: function (item) {
                  return  item.id === params.nodes[0] ;
                },
                returnType :'Array'
              });
            
              clusterByGroup([array_group[0].group]);
            }
          }
        }
      });
    }
    //*************************
    //clustering Connection
    //*************************
    if(x.clusteringConnection){
      
      function clusterByConnection() {
        for (var i = 0; i < x.clusteringConnection.nodes.length; i++) {
          instance.network.clusterByConnection(x.clusteringConnection.nodes[i])
        }
      }
      clusterByConnection();
    }
    
    //*************************
    //clustering hubsize
    //*************************
    if(x.clusteringHubsize){
      
      function clusterByHubsize() {
        var clusterOptionsByData = {
          processProperties: function(clusterOptions, childNodes) {
            var cluster_level = 9999999
                  for (var i = 0; i < childNodes.length; i++) {
                      //totalMass += childNodes[i].mass;
                      if(childNodes[i].level){
                        cluster_level = Math.min(cluster_level, childNodes[i].level)
                      }
                      if(i === 0){
                        //clusterOptions.shape =  childNodes[i].shape;
                        clusterOptions.color =  childNodes[i].color.background;
                      }else{
                        //if(childNodes[i].shape !== clusterOptions.shape){
                          //clusterOptions.shape = 'database';
                        //}
                        if(childNodes[i].color.background !== clusterOptions.color){
                          clusterOptions.color = 'grey';
                        }
                      }
                  }
            clusterOptions.label = "[" + childNodes.length + "]";
            if(cluster_level !== 9999999){
              clusterOptions.level = cluster_level
            }
            return clusterOptions;
          },
          clusterNodeProperties: {borderWidth:3, shape:'box', font:{size:30}}
        }
        if(x.clusteringHubsize.size > 0){
          instance.network.clusterByHubsize(x.clusteringHubsize.size, clusterOptionsByData);
        }else{
          instance.network.clusterByHubsize(undefined, clusterOptionsByData);
        }
      }
      
      clusterByHubsize();
    }
    
    if(x.clusteringColor){
      
    //*************************
    //clustering color
    //*************************
    function clusterByColor() {
        var colors = x.clusteringColor.colors
        var clusterOptionsByData;
        for (var i = 0; i < colors.length; i++) {
          var color = colors[i];
          var sh = x.clusteringColor.shape[i];
          var force = x.clusteringColor.force[i];
          clusterOptionsByData = {
              joinCondition: function (childOptions) {
                  return childOptions.color.background == color; // the color is fully defined in the node.
              },
              processProperties: function (clusterOptions, childNodes, childEdges) {
                  var totalMass = 0;
                  var cluster_level = 9999999;
                  for (var i = 0; i < childNodes.length; i++) {
                      totalMass += childNodes[i].mass;
                      if(childNodes[i].level){
                        cluster_level = Math.min(cluster_level, childNodes[i].level)
                      }
                      if(force === false){
                        if(i === 0){
                          clusterOptions.shape =  childNodes[i].shape;
                        }else{
                          if(childNodes[i].shape !== clusterOptions.shape){
                            clusterOptions.shape = sh;
                          }
                        }
                      } else {
                        clusterOptions.shape = sh;
                      }

                  }
                  clusterOptions.value = totalMass;
                  if(cluster_level !== 9999999){
                    clusterOptions.level = cluster_level
                  }
                  return clusterOptions;
              },
              clusterNodeProperties: {id: 'cluster:' + color, borderWidth: 3, color:color, label: x.clusteringColor.label + color}
          }
          instance.network.cluster(clusterOptionsByData);
        }
      }
      
      clusterByColor();
    }

    //*************************
    //clustering groups
    //*************************
    if(x.clusteringGroup){
      
      function clusterByGroup(groups) {
        var clusterOptionsByData;
        for (var i = 0; i < groups.length; i++) {
          var group = groups[i];
          var j = x.clusteringGroup.groups.indexOf(group);
          if(j !== -1) {
            var col = x.clusteringGroup.color[j];
            var sh = x.clusteringGroup.shape[j];
            var force = x.clusteringGroup.force[j];
            var sc_size = x.clusteringGroup.scale_size[j];
            
            clusterOptionsByData = {
                joinCondition: function (childOptions) {
                    return childOptions.group == group; //
                },
                processProperties: function (clusterOptions, childNodes, childEdges) {
                    var totalMass = 0;
                    var cluster_level = 9999999;
                    for (var i = 0; i < childNodes.length; i++) {
                        totalMass += childNodes[i].mass;
                        if(childNodes[i].level){
                          cluster_level = Math.min(cluster_level, childNodes[i].level)
                        }
                        if(force === false){
                          if(i === 0){
                            clusterOptions.shape =  childNodes[i].shape;
                            clusterOptions.color =  childNodes[i].color.background;
                          }else{
                            if(childNodes[i].shape !== clusterOptions.shape){
                              clusterOptions.shape = sh;
                            }
                            if(childNodes[i].color.background !== clusterOptions.color){
                              clusterOptions.color = col;
                            }
                          }
                        } else {
                          clusterOptions.shape = sh;
                          clusterOptions.color = col;
                        }
                    }
                    if(sc_size){
                       clusterOptions.value = totalMass;
                    }
                    if(cluster_level !== 9999999){
                      clusterOptions.level = cluster_level
                    }
                    return clusterOptions;
                },
                clusterNodeProperties: {id: 'cluster:' + group, borderWidth: 3, label:x.clusteringGroup.label + group}
            }
            instance.network.cluster(clusterOptionsByData);
          }

        }
      }
      clusterByGroup(x.clusteringGroup.groups);
    }
  
    //*************************
    //clustering by zoom
    //*************************
    if(x.clusteringOutliers){
      
      clusterFactor = x.clusteringOutliers.clusterFactor;
      
      // set the first initial zoom level
      instance.network.on('initRedraw', function() {
        if (lastClusterZoomLevel === 0) {
          lastClusterZoomLevel = instance.network.getScale();
        }
      });

      // we use the zoom event for our clustering
      instance.network.on('zoom', function (params) {
        if(ctrlwait === 0){
        if (params.direction == '-') {
          if (params.scale < lastClusterZoomLevel*clusterFactor) {
            makeClusters(params.scale);
            lastClusterZoomLevel = params.scale;
          }
        }
        else {
          openClusters(params.scale);
        }
        }
      });
    }

    // make the clusters
    function makeClusters(scale) {
        ctrlwait = 1;
        var clusterOptionsByData = {
            processProperties: function (clusterOptions, childNodes) {
                clusterIndex = clusterIndex + 1;
                var childrenCount = 0;
                var cluster_level = 9999999;
                for (var i = 0; i < childNodes.length; i++) {
                    childrenCount += childNodes[i].childrenCount || 1;
                    if(childNodes[i].level){
                      cluster_level = Math.min(cluster_level, childNodes[i].level)
                    }
                }
                clusterOptions.childrenCount = childrenCount;
                clusterOptions.label = "# " + childrenCount + "";
                clusterOptions.font = {size: childrenCount*5+30}
                clusterOptions.id = 'cluster:' + clusterIndex;
                clusters.push({id:'cluster:' + clusterIndex, scale:scale});
                
                if(cluster_level !== 9999999){
                  clusterOptions.level = cluster_level
                }
                return clusterOptions;
            },
            clusterNodeProperties: {borderWidth: 3, shape: 'database', font: {size: 30}}
        }
        instance.network.clusterOutliers(clusterOptionsByData);
        if (x.clusteringOutliers.stabilize) {
            instance.network.stabilize();
        };
        ctrlwait = 0;
    }

    // open them back up!
    function openClusters(scale) {
        ctrlwait = 1;
        var newClusters = [];
        var declustered = false;
        for (var i = 0; i < clusters.length; i++) {
            if (clusters[i].scale < scale) {
                instance.network.openCluster(clusters[i].id);
                lastClusterZoomLevel = scale;
                declustered = true;
            }
            else {
                newClusters.push(clusters[i])
            }
        }
        clusters = newClusters;
        if (x.clusteringOutliers.stabilize) {
            instance.network.stabilize();
        };
        ctrlwait = 0;
    }
    
    //******************
    // init selection
    //******************
    if(el_id.idselection && x.nodes && x.idselection.selected !== undefined){ 
      onIdChange(''+ x.idselection.selected, true);
    }
      
    if(el_id.byselection && x.nodes && x.byselection.selected !== undefined){ 
      onByChange(x.byselection.selected);
      selectNode = document.getElementById('selectedBy'+el.id);
      selectNode.value = x.byselection.selected;
    }
    
    // try to fix icons loading css bug...
    function iconsRedraw() {
      setTimeout(function(){
        if(instance.network)
          instance.network.redraw();
        if(instance.legend)
          instance.legend.redraw();
      }, 250);
    };
    
    if(x.iconsRedraw !== undefined){
      if(x.iconsRedraw){
        iconsRedraw();
        instance.network.once("stabilized", function(){iconsRedraw();})
      }
    }
  }, 
  
  resize: function(el, width, height, instance) {
      if(instance.network)
        instance.network.fit();
      if(instance.legend)
        instance.legend.fit();
  }
  
});
```
