## Supplementary material for "Evaluating Regional Diversity in Scientific Communication: A Comparative Analysis of COVID-19 Preprints and Peer-Reviewed Publications": d3-bundle.min.js

```
// https://d3js.org/d3-selection/ v1.4.2 Copyright 2020 Mike Bostock
!function(t,n){"object"==typeof exports&&"undefined"!=typeof module?n(exports):"function"==typeof define&&define.amd?define(["exports"],n):n((t=t||self).d3=t.d3||{})}(this,function(t){"use strict";var n="http://www.w3.org/1999/xhtml",e={svg:"http://www.w3.org/2000/svg",xhtml:n,xlink:"http://www.w3.org/1999/xlink",xml:"http://www.w3.org/XML/1998/namespace",xmlns:"http://www.w3.org/2000/xmlns/"};function r(t){var n=t+="",r=n.indexOf(":");return r>=0&&"xmlns"!==(n=t.slice(0,r))&&(t=t.slice(r+1)),e.hasOwnProperty(n)?{space:e[n],local:t}:t}function i(t){var e=r(t);return(e.local?function(t){return function(){return this.ownerDocument.createElementNS(t.space,t.local)}}:function(t){return function(){var e=this.ownerDocument,r=this.namespaceURI;return r===n&&e.documentElement.namespaceURI===n?e.createElement(t):e.createElementNS(r,t)}})(e)}function o(){}function u(t){return null==t?o:function(){return this.querySelector(t)}}function c(){return[]}function s(t){return null==t?c:function(){return this.querySelectorAll(t)}}function a(t){return function(){return this.matches(t)}}function l(t){return new Array(t.length)}function f(t,n){this.ownerDocument=t.ownerDocument,this.namespaceURI=t.namespaceURI,this._next=null,this._parent=t,this.__data__=n}f.prototype={constructor:f,appendChild:function(t){return this._parent.insertBefore(t,this._next)},insertBefore:function(t,n){return this._parent.insertBefore(t,n)},querySelector:function(t){return this._parent.querySelector(t)},querySelectorAll:function(t){return this._parent.querySelectorAll(t)}};var h="$";function p(t,n,e,r,i,o){for(var u,c=0,s=n.length,a=o.length;c<a;++c)(u=n[c])?(u.__data__=o[c],r[c]=u):e[c]=new f(t,o[c]);for(;c<s;++c)(u=n[c])&&(i[c]=u)}function _(t,n,e,r,i,o,u){var c,s,a,l={},p=n.length,_=o.length,v=new Array(p);for(c=0;c<p;++c)(s=n[c])&&(v[c]=a=h+u.call(s,s.__data__,c,n),a in l?i[c]=s:l[a]=s);for(c=0;c<_;++c)(s=l[a=h+u.call(t,o[c],c,o)])?(r[c]=s,s.__data__=o[c],l[a]=null):e[c]=new f(t,o[c]);for(c=0;c<p;++c)(s=n[c])&&l[v[c]]===s&&(i[c]=s)}function v(t,n){return t<n?-1:t>n?1:t>=n?0:NaN}function d(t){return t.ownerDocument&&t.ownerDocument.defaultView||t.document&&t||t.defaultView}function y(t,n){return t.style.getPropertyValue(n)||d(t).getComputedStyle(t,null).getPropertyValue(n)}function m(t){return t.trim().split(/^|\s+/)}function g(t){return t.classList||new w(t)}function w(t){this._node=t,this._names=m(t.getAttribute("class")||"")}function A(t,n){for(var e=g(t),r=-1,i=n.length;++r<i;)e.add(n[r])}function x(t,n){for(var e=g(t),r=-1,i=n.length;++r<i;)e.remove(n[r])}function S(){this.textContent=""}function b(){this.innerHTML=""}function E(){this.nextSibling&&this.parentNode.appendChild(this)}function N(){this.previousSibling&&this.parentNode.insertBefore(this,this.parentNode.firstChild)}function P(){return null}function C(){var t=this.parentNode;t&&t.removeChild(this)}function L(){var t=this.cloneNode(!1),n=this.parentNode;return n?n.insertBefore(t,this.nextSibling):t}function T(){var t=this.cloneNode(!0),n=this.parentNode;return n?n.insertBefore(t,this.nextSibling):t}w.prototype={add:function(t){this._names.indexOf(t)<0&&(this._names.push(t),this._node.setAttribute("class",this._names.join(" ")))},remove:function(t){var n=this._names.indexOf(t);n>=0&&(this._names.splice(n,1),this._node.setAttribute("class",this._names.join(" ")))},contains:function(t){return this._names.indexOf(t)>=0}};var B={};(t.event=null,"undefined"!=typeof document)&&("onmouseenter"in document.documentElement||(B={mouseenter:"mouseover",mouseleave:"mouseout"}));function q(t,n,e){return t=D(t,n,e),function(n){var e=n.relatedTarget;e&&(e===this||8&e.compareDocumentPosition(this))||t.call(this,n)}}function D(n,e,r){return function(i){var o=t.event;t.event=i;try{n.call(this,this.__data__,e,r)}finally{t.event=o}}}function M(t){return function(){var n=this.__on;if(n){for(var e,r=0,i=-1,o=n.length;r<o;++r)e=n[r],t.type&&e.type!==t.type||e.name!==t.name?n[++i]=e:this.removeEventListener(e.type,e.listener,e.capture);++i?n.length=i:delete this.__on}}}function O(t,n,e){var r=B.hasOwnProperty(t.type)?q:D;return function(i,o,u){var c,s=this.__on,a=r(n,o,u);if(s)for(var l=0,f=s.length;l<f;++l)if((c=s[l]).type===t.type&&c.name===t.name)return this.removeEventListener(c.type,c.listener,c.capture),this.addEventListener(c.type,c.listener=a,c.capture=e),void(c.value=n);this.addEventListener(t.type,a,e),c={type:t.type,name:t.name,value:n,listener:a,capture:e},s?s.push(c):this.__on=[c]}}function V(t,n,e){var r=d(t),i=r.CustomEvent;"function"==typeof i?i=new i(n,e):(i=r.document.createEvent("Event"),e?(i.initEvent(n,e.bubbles,e.cancelable),i.detail=e.detail):i.initEvent(n,!1,!1)),t.dispatchEvent(i)}var j=[null];function R(t,n){this._groups=t,this._parents=n}function H(){return new R([[document.documentElement]],j)}function I(t){return"string"==typeof t?new R([[document.querySelector(t)]],[document.documentElement]):new R([[t]],j)}R.prototype=H.prototype={constructor:R,select:function(t){"function"!=typeof t&&(t=u(t));for(var n=this._groups,e=n.length,r=new Array(e),i=0;i<e;++i)for(var o,c,s=n[i],a=s.length,l=r[i]=new Array(a),f=0;f<a;++f)(o=s[f])&&(c=t.call(o,o.__data__,f,s))&&("__data__"in o&&(c.__data__=o.__data__),l[f]=c);return new R(r,this._parents)},selectAll:function(t){"function"!=typeof t&&(t=s(t));for(var n=this._groups,e=n.length,r=[],i=[],o=0;o<e;++o)for(var u,c=n[o],a=c.length,l=0;l<a;++l)(u=c[l])&&(r.push(t.call(u,u.__data__,l,c)),i.push(u));return new R(r,i)},filter:function(t){"function"!=typeof t&&(t=a(t));for(var n=this._groups,e=n.length,r=new Array(e),i=0;i<e;++i)for(var o,u=n[i],c=u.length,s=r[i]=[],l=0;l<c;++l)(o=u[l])&&t.call(o,o.__data__,l,u)&&s.push(o);return new R(r,this._parents)},data:function(t,n){if(!t)return d=new Array(this.size()),l=-1,this.each(function(t){d[++l]=t}),d;var e,r=n?_:p,i=this._parents,o=this._groups;"function"!=typeof t&&(e=t,t=function(){return e});for(var u=o.length,c=new Array(u),s=new Array(u),a=new Array(u),l=0;l<u;++l){var f=i[l],h=o[l],v=h.length,d=t.call(f,f&&f.__data__,l,i),y=d.length,m=s[l]=new Array(y),g=c[l]=new Array(y);r(f,h,m,g,a[l]=new Array(v),d,n);for(var w,A,x=0,S=0;x<y;++x)if(w=m[x]){for(x>=S&&(S=x+1);!(A=g[S])&&++S<y;);w._next=A||null}}return(c=new R(c,i))._enter=s,c._exit=a,c},enter:function(){return new R(this._enter||this._groups.map(l),this._parents)},exit:function(){return new R(this._exit||this._groups.map(l),this._parents)},join:function(t,n,e){var r=this.enter(),i=this,o=this.exit();return r="function"==typeof t?t(r):r.append(t+""),null!=n&&(i=n(i)),null==e?o.remove():e(o),r&&i?r.merge(i).order():i},merge:function(t){for(var n=this._groups,e=t._groups,r=n.length,i=e.length,o=Math.min(r,i),u=new Array(r),c=0;c<o;++c)for(var s,a=n[c],l=e[c],f=a.length,h=u[c]=new Array(f),p=0;p<f;++p)(s=a[p]||l[p])&&(h[p]=s);for(;c<r;++c)u[c]=n[c];return new R(u,this._parents)},order:function(){for(var t=this._groups,n=-1,e=t.length;++n<e;)for(var r,i=t[n],o=i.length-1,u=i[o];--o>=0;)(r=i[o])&&(u&&4^r.compareDocumentPosition(u)&&u.parentNode.insertBefore(r,u),u=r);return this},sort:function(t){function n(n,e){return n&&e?t(n.__data__,e.__data__):!n-!e}t||(t=v);for(var e=this._groups,r=e.length,i=new Array(r),o=0;o<r;++o){for(var u,c=e[o],s=c.length,a=i[o]=new Array(s),l=0;l<s;++l)(u=c[l])&&(a[l]=u);a.sort(n)}return new R(i,this._parents).order()},call:function(){var t=arguments[0];return arguments[0]=this,t.apply(null,arguments),this},nodes:function(){var t=new Array(this.size()),n=-1;return this.each(function(){t[++n]=this}),t},node:function(){for(var t=this._groups,n=0,e=t.length;n<e;++n)for(var r=t[n],i=0,o=r.length;i<o;++i){var u=r[i];if(u)return u}return null},size:function(){var t=0;return this.each(function(){++t}),t},empty:function(){return!this.node()},each:function(t){for(var n=this._groups,e=0,r=n.length;e<r;++e)for(var i,o=n[e],u=0,c=o.length;u<c;++u)(i=o[u])&&t.call(i,i.__data__,u,o);return this},attr:function(t,n){var e=r(t);if(arguments.length<2){var i=this.node();return e.local?i.getAttributeNS(e.space,e.local):i.getAttribute(e)}return this.each((null==n?e.local?function(t){return function(){this.removeAttributeNS(t.space,t.local)}}:function(t){return function(){this.removeAttribute(t)}}:"function"==typeof n?e.local?function(t,n){return function(){var e=n.apply(this,arguments);null==e?this.removeAttributeNS(t.space,t.local):this.setAttributeNS(t.space,t.local,e)}}:function(t,n){return function(){var e=n.apply(this,arguments);null==e?this.removeAttribute(t):this.setAttribute(t,e)}}:e.local?function(t,n){return function(){this.setAttributeNS(t.space,t.local,n)}}:function(t,n){return function(){this.setAttribute(t,n)}})(e,n))},style:function(t,n,e){return arguments.length>1?this.each((null==n?function(t){return function(){this.style.removeProperty(t)}}:"function"==typeof n?function(t,n,e){return function(){var r=n.apply(this,arguments);null==r?this.style.removeProperty(t):this.style.setProperty(t,r,e)}}:function(t,n,e){return function(){this.style.setProperty(t,n,e)}})(t,n,null==e?"":e)):y(this.node(),t)},property:function(t,n){return arguments.length>1?this.each((null==n?function(t){return function(){delete this[t]}}:"function"==typeof n?function(t,n){return function(){var e=n.apply(this,arguments);null==e?delete this[t]:this[t]=e}}:function(t,n){return function(){this[t]=n}})(t,n)):this.node()[t]},classed:function(t,n){var e=m(t+"");if(arguments.length<2){for(var r=g(this.node()),i=-1,o=e.length;++i<o;)if(!r.contains(e[i]))return!1;return!0}return this.each(("function"==typeof n?function(t,n){return function(){(n.apply(this,arguments)?A:x)(this,t)}}:n?function(t){return function(){A(this,t)}}:function(t){return function(){x(this,t)}})(e,n))},text:function(t){return arguments.length?this.each(null==t?S:("function"==typeof t?function(t){return function(){var n=t.apply(this,arguments);this.textContent=null==n?"":n}}:function(t){return function(){this.textContent=t}})(t)):this.node().textContent},html:function(t){return arguments.length?this.each(null==t?b:("function"==typeof t?function(t){return function(){var n=t.apply(this,arguments);this.innerHTML=null==n?"":n}}:function(t){return function(){this.innerHTML=t}})(t)):this.node().innerHTML},raise:function(){return this.each(E)},lower:function(){return this.each(N)},append:function(t){var n="function"==typeof t?t:i(t);return this.select(function(){return this.appendChild(n.apply(this,arguments))})},insert:function(t,n){var e="function"==typeof t?t:i(t),r=null==n?P:"function"==typeof n?n:u(n);return this.select(function(){return this.insertBefore(e.apply(this,arguments),r.apply(this,arguments)||null)})},remove:function(){return this.each(C)},clone:function(t){return this.select(t?T:L)},datum:function(t){return arguments.length?this.property("__data__",t):this.node().__data__},on:function(t,n,e){var r,i,o=function(t){return t.trim().split(/^|\s+/).map(function(t){var n="",e=t.indexOf(".");return e>=0&&(n=t.slice(e+1),t=t.slice(0,e)),{type:t,name:n}})}(t+""),u=o.length;if(!(arguments.length<2)){for(c=n?O:M,null==e&&(e=!1),r=0;r<u;++r)this.each(c(o[r],n,e));return this}var c=this.node().__on;if(c)for(var s,a=0,l=c.length;a<l;++a)for(r=0,s=c[a];r<u;++r)if((i=o[r]).type===s.type&&i.name===s.name)return s.value},dispatch:function(t,n){return this.each(("function"==typeof n?function(t,n){return function(){return V(this,t,n.apply(this,arguments))}}:function(t,n){return function(){return V(this,t,n)}})(t,n))}};var U=0;function z(){return new G}function G(){this._="@"+(++U).toString(36)}function X(){for(var n,e=t.event;n=e.sourceEvent;)e=n;return e}function k(t,n){var e=t.ownerSVGElement||t;if(e.createSVGPoint){var r=e.createSVGPoint();return r.x=n.clientX,r.y=n.clientY,[(r=r.matrixTransform(t.getScreenCTM().inverse())).x,r.y]}var i=t.getBoundingClientRect();return[n.clientX-i.left-t.clientLeft,n.clientY-i.top-t.clientTop]}G.prototype=z.prototype={constructor:G,get:function(t){for(var n=this._;!(n in t);)if(!(t=t.parentNode))return;return t[n]},set:function(t,n){return t[this._]=n},remove:function(t){return this._ in t&&delete t[this._]},toString:function(){return this._}},t.clientPoint=k,t.create=function(t){return I(i(t).call(document.documentElement))},t.creator=i,t.customEvent=function(n,e,r,i){var o=t.event;n.sourceEvent=t.event,t.event=n;try{return e.apply(r,i)}finally{t.event=o}},t.local=z,t.matcher=a,t.mouse=function(t){var n=X();return n.changedTouches&&(n=n.changedTouches[0]),k(t,n)},t.namespace=r,t.namespaces=e,t.select=I,t.selectAll=function(t){return"string"==typeof t?new R([document.querySelectorAll(t)],[document.documentElement]):new R([null==t?[]:t],j)},t.selection=H,t.selector=u,t.selectorAll=s,t.style=y,t.touch=function(t,n,e){arguments.length<3&&(e=n,n=X().changedTouches);for(var r,i=0,o=n?n.length:0;i<o;++i)if((r=n[i]).identifier===e)return k(t,r);return null},t.touches=function(t,n){null==n&&(n=X().touches);for(var e=0,r=n?n.length:0,i=new Array(r);e<r;++e)i[e]=k(t,n[e]);return i},t.window=d,Object.defineProperty(t,"__esModule",{value:!0})});

// https://d3js.org/d3-color/ v1.4.1 Copyright 2020 Mike Bostock
!function(t,e){"object"==typeof exports&&"undefined"!=typeof module?e(exports):"function"==typeof define&&define.amd?define(["exports"],e):e((t=t||self).d3=t.d3||{})}(this,function(t){"use strict";function e(t,e,n){t.prototype=e.prototype=n,n.constructor=t}function n(t,e){var n=Object.create(t.prototype);for(var i in e)n[i]=e[i];return n}function i(){}var r="\\s*([+-]?\\d+)\\s*",a="\\s*([+-]?\\d*\\.?\\d+(?:[eE][+-]?\\d+)?)\\s*",s="\\s*([+-]?\\d*\\.?\\d+(?:[eE][+-]?\\d+)?)%\\s*",o=/^#([0-9a-f]{3,8})$/,h=new RegExp("^rgb\\("+[r,r,r]+"\\)$"),l=new RegExp("^rgb\\("+[s,s,s]+"\\)$"),u=new RegExp("^rgba\\("+[r,r,r,a]+"\\)$"),c=new RegExp("^rgba\\("+[s,s,s,a]+"\\)$"),g=new RegExp("^hsl\\("+[a,s,s]+"\\)$"),f=new RegExp("^hsla\\("+[a,s,s,a]+"\\)$"),d={aliceblue:15792383,antiquewhite:16444375,aqua:65535,aquamarine:8388564,azure:15794175,beige:16119260,bisque:16770244,black:0,blanchedalmond:16772045,blue:255,blueviolet:9055202,brown:10824234,burlywood:14596231,cadetblue:6266528,chartreuse:8388352,chocolate:13789470,coral:16744272,cornflowerblue:6591981,cornsilk:16775388,crimson:14423100,cyan:65535,darkblue:139,darkcyan:35723,darkgoldenrod:12092939,darkgray:11119017,darkgreen:25600,darkgrey:11119017,darkkhaki:12433259,darkmagenta:9109643,darkolivegreen:5597999,darkorange:16747520,darkorchid:10040012,darkred:9109504,darksalmon:15308410,darkseagreen:9419919,darkslateblue:4734347,darkslategray:3100495,darkslategrey:3100495,darkturquoise:52945,darkviolet:9699539,deeppink:16716947,deepskyblue:49151,dimgray:6908265,dimgrey:6908265,dodgerblue:2003199,firebrick:11674146,floralwhite:16775920,forestgreen:2263842,fuchsia:16711935,gainsboro:14474460,ghostwhite:16316671,gold:16766720,goldenrod:14329120,gray:8421504,green:32768,greenyellow:11403055,grey:8421504,honeydew:15794160,hotpink:16738740,indianred:13458524,indigo:4915330,ivory:16777200,khaki:15787660,lavender:15132410,lavenderblush:16773365,lawngreen:8190976,lemonchiffon:16775885,lightblue:11393254,lightcoral:15761536,lightcyan:14745599,lightgoldenrodyellow:16448210,lightgray:13882323,lightgreen:9498256,lightgrey:13882323,lightpink:16758465,lightsalmon:16752762,lightseagreen:2142890,lightskyblue:8900346,lightslategray:7833753,lightslategrey:7833753,lightsteelblue:11584734,lightyellow:16777184,lime:65280,limegreen:3329330,linen:16445670,magenta:16711935,maroon:8388608,mediumaquamarine:6737322,mediumblue:205,mediumorchid:12211667,mediumpurple:9662683,mediumseagreen:3978097,mediumslateblue:8087790,mediumspringgreen:64154,mediumturquoise:4772300,mediumvioletred:13047173,midnightblue:1644912,mintcream:16121850,mistyrose:16770273,moccasin:16770229,navajowhite:16768685,navy:128,oldlace:16643558,olive:8421376,olivedrab:7048739,orange:16753920,orangered:16729344,orchid:14315734,palegoldenrod:15657130,palegreen:10025880,paleturquoise:11529966,palevioletred:14381203,papayawhip:16773077,peachpuff:16767673,peru:13468991,pink:16761035,plum:14524637,powderblue:11591910,purple:8388736,rebeccapurple:6697881,red:16711680,rosybrown:12357519,royalblue:4286945,saddlebrown:9127187,salmon:16416882,sandybrown:16032864,seagreen:3050327,seashell:16774638,sienna:10506797,silver:12632256,skyblue:8900331,slateblue:6970061,slategray:7372944,slategrey:7372944,snow:16775930,springgreen:65407,steelblue:4620980,tan:13808780,teal:32896,thistle:14204888,tomato:16737095,turquoise:4251856,violet:15631086,wheat:16113331,white:16777215,whitesmoke:16119285,yellow:16776960,yellowgreen:10145074};function p(){return this.rgb().formatHex()}function b(){return this.rgb().formatRgb()}function y(t){var e,n;return t=(t+"").trim().toLowerCase(),(e=o.exec(t))?(n=e[1].length,e=parseInt(e[1],16),6===n?w(e):3===n?new M(e>>8&15|e>>4&240,e>>4&15|240&e,(15&e)<<4|15&e,1):8===n?m(e>>24&255,e>>16&255,e>>8&255,(255&e)/255):4===n?m(e>>12&15|e>>8&240,e>>8&15|e>>4&240,e>>4&15|240&e,((15&e)<<4|15&e)/255):null):(e=h.exec(t))?new M(e[1],e[2],e[3],1):(e=l.exec(t))?new M(255*e[1]/100,255*e[2]/100,255*e[3]/100,1):(e=u.exec(t))?m(e[1],e[2],e[3],e[4]):(e=c.exec(t))?m(255*e[1]/100,255*e[2]/100,255*e[3]/100,e[4]):(e=g.exec(t))?R(e[1],e[2]/100,e[3]/100,1):(e=f.exec(t))?R(e[1],e[2]/100,e[3]/100,e[4]):d.hasOwnProperty(t)?w(d[t]):"transparent"===t?new M(NaN,NaN,NaN,0):null}function w(t){return new M(t>>16&255,t>>8&255,255&t,1)}function m(t,e,n,i){return i<=0&&(t=e=n=NaN),new M(t,e,n,i)}function N(t){return t instanceof i||(t=y(t)),t?new M((t=t.rgb()).r,t.g,t.b,t.opacity):new M}function k(t,e,n,i){return 1===arguments.length?N(t):new M(t,e,n,null==i?1:i)}function M(t,e,n,i){this.r=+t,this.g=+e,this.b=+n,this.opacity=+i}function v(){return"#"+q(this.r)+q(this.g)+q(this.b)}function x(){var t=this.opacity;return(1===(t=isNaN(t)?1:Math.max(0,Math.min(1,t)))?"rgb(":"rgba(")+Math.max(0,Math.min(255,Math.round(this.r)||0))+", "+Math.max(0,Math.min(255,Math.round(this.g)||0))+", "+Math.max(0,Math.min(255,Math.round(this.b)||0))+(1===t?")":", "+t+")")}function q(t){return((t=Math.max(0,Math.min(255,Math.round(t)||0)))<16?"0":"")+t.toString(16)}function R(t,e,n,i){return i<=0?t=e=n=NaN:n<=0||n>=1?t=e=NaN:e<=0&&(t=NaN),new H(t,e,n,i)}function E(t){if(t instanceof H)return new H(t.h,t.s,t.l,t.opacity);if(t instanceof i||(t=y(t)),!t)return new H;if(t instanceof H)return t;var e=(t=t.rgb()).r/255,n=t.g/255,r=t.b/255,a=Math.min(e,n,r),s=Math.max(e,n,r),o=NaN,h=s-a,l=(s+a)/2;return h?(o=e===s?(n-r)/h+6*(n<r):n===s?(r-e)/h+2:(e-n)/h+4,h/=l<.5?s+a:2-s-a,o*=60):h=l>0&&l<1?0:o,new H(o,h,l,t.opacity)}function $(t,e,n,i){return 1===arguments.length?E(t):new H(t,e,n,null==i?1:i)}function H(t,e,n,i){this.h=+t,this.s=+e,this.l=+n,this.opacity=+i}function j(t,e,n){return 255*(t<60?e+(n-e)*t/60:t<180?n:t<240?e+(n-e)*(240-t)/60:e)}e(i,y,{copy:function(t){return Object.assign(new this.constructor,this,t)},displayable:function(){return this.rgb().displayable()},hex:p,formatHex:p,formatHsl:function(){return E(this).formatHsl()},formatRgb:b,toString:b}),e(M,k,n(i,{brighter:function(t){return t=null==t?1/.7:Math.pow(1/.7,t),new M(this.r*t,this.g*t,this.b*t,this.opacity)},darker:function(t){return t=null==t?.7:Math.pow(.7,t),new M(this.r*t,this.g*t,this.b*t,this.opacity)},rgb:function(){return this},displayable:function(){return-.5<=this.r&&this.r<255.5&&-.5<=this.g&&this.g<255.5&&-.5<=this.b&&this.b<255.5&&0<=this.opacity&&this.opacity<=1},hex:v,formatHex:v,formatRgb:x,toString:x})),e(H,$,n(i,{brighter:function(t){return t=null==t?1/.7:Math.pow(1/.7,t),new H(this.h,this.s,this.l*t,this.opacity)},darker:function(t){return t=null==t?.7:Math.pow(.7,t),new H(this.h,this.s,this.l*t,this.opacity)},rgb:function(){var t=this.h%360+360*(this.h<0),e=isNaN(t)||isNaN(this.s)?0:this.s,n=this.l,i=n+(n<.5?n:1-n)*e,r=2*n-i;return new M(j(t>=240?t-240:t+120,r,i),j(t,r,i),j(t<120?t+240:t-120,r,i),this.opacity)},displayable:function(){return(0<=this.s&&this.s<=1||isNaN(this.s))&&0<=this.l&&this.l<=1&&0<=this.opacity&&this.opacity<=1},formatHsl:function(){var t=this.opacity;return(1===(t=isNaN(t)?1:Math.max(0,Math.min(1,t)))?"hsl(":"hsla(")+(this.h||0)+", "+100*(this.s||0)+"%, "+100*(this.l||0)+"%"+(1===t?")":", "+t+")")}}));var O=Math.PI/180,P=180/Math.PI,I=.96422,S=1,_=.82521,z=4/29,C=6/29,L=3*C*C,A=C*C*C;function B(t){if(t instanceof F)return new F(t.l,t.a,t.b,t.opacity);if(t instanceof V)return W(t);t instanceof M||(t=N(t));var e,n,i=Q(t.r),r=Q(t.g),a=Q(t.b),s=G((.2225045*i+.7168786*r+.0606169*a)/S);return i===r&&r===a?e=n=s:(e=G((.4360747*i+.3850649*r+.1430804*a)/I),n=G((.0139322*i+.0971045*r+.7141733*a)/_)),new F(116*s-16,500*(e-s),200*(s-n),t.opacity)}function D(t,e,n,i){return 1===arguments.length?B(t):new F(t,e,n,null==i?1:i)}function F(t,e,n,i){this.l=+t,this.a=+e,this.b=+n,this.opacity=+i}function G(t){return t>A?Math.pow(t,1/3):t/L+z}function J(t){return t>C?t*t*t:L*(t-z)}function K(t){return 255*(t<=.0031308?12.92*t:1.055*Math.pow(t,1/2.4)-.055)}function Q(t){return(t/=255)<=.04045?t/12.92:Math.pow((t+.055)/1.055,2.4)}function T(t){if(t instanceof V)return new V(t.h,t.c,t.l,t.opacity);if(t instanceof F||(t=B(t)),0===t.a&&0===t.b)return new V(NaN,0<t.l&&t.l<100?0:NaN,t.l,t.opacity);var e=Math.atan2(t.b,t.a)*P;return new V(e<0?e+360:e,Math.sqrt(t.a*t.a+t.b*t.b),t.l,t.opacity)}function U(t,e,n,i){return 1===arguments.length?T(t):new V(t,e,n,null==i?1:i)}function V(t,e,n,i){this.h=+t,this.c=+e,this.l=+n,this.opacity=+i}function W(t){if(isNaN(t.h))return new F(t.l,0,0,t.opacity);var e=t.h*O;return new F(t.l,Math.cos(e)*t.c,Math.sin(e)*t.c,t.opacity)}e(F,D,n(i,{brighter:function(t){return new F(this.l+18*(null==t?1:t),this.a,this.b,this.opacity)},darker:function(t){return new F(this.l-18*(null==t?1:t),this.a,this.b,this.opacity)},rgb:function(){var t=(this.l+16)/116,e=isNaN(this.a)?t:t+this.a/500,n=isNaN(this.b)?t:t-this.b/200;return new M(K(3.1338561*(e=I*J(e))-1.6168667*(t=S*J(t))-.4906146*(n=_*J(n))),K(-.9787684*e+1.9161415*t+.033454*n),K(.0719453*e-.2289914*t+1.4052427*n),this.opacity)}})),e(V,U,n(i,{brighter:function(t){return new V(this.h,this.c,this.l+18*(null==t?1:t),this.opacity)},darker:function(t){return new V(this.h,this.c,this.l-18*(null==t?1:t),this.opacity)},rgb:function(){return W(this).rgb()}}));var X=-.14861,Y=1.78277,Z=-.29227,tt=-.90649,et=1.97294,nt=et*tt,it=et*Y,rt=Y*Z-tt*X;function at(t,e,n,i){return 1===arguments.length?function(t){if(t instanceof st)return new st(t.h,t.s,t.l,t.opacity);t instanceof M||(t=N(t));var e=t.r/255,n=t.g/255,i=t.b/255,r=(rt*i+nt*e-it*n)/(rt+nt-it),a=i-r,s=(et*(n-r)-Z*a)/tt,o=Math.sqrt(s*s+a*a)/(et*r*(1-r)),h=o?Math.atan2(s,a)*P-120:NaN;return new st(h<0?h+360:h,o,r,t.opacity)}(t):new st(t,e,n,null==i?1:i)}function st(t,e,n,i){this.h=+t,this.s=+e,this.l=+n,this.opacity=+i}e(st,at,n(i,{brighter:function(t){return t=null==t?1/.7:Math.pow(1/.7,t),new st(this.h,this.s,this.l*t,this.opacity)},darker:function(t){return t=null==t?.7:Math.pow(.7,t),new st(this.h,this.s,this.l*t,this.opacity)},rgb:function(){var t=isNaN(this.h)?0:(this.h+120)*O,e=+this.l,n=isNaN(this.s)?0:this.s*e*(1-e),i=Math.cos(t),r=Math.sin(t);return new M(255*(e+n*(X*i+Y*r)),255*(e+n*(Z*i+tt*r)),255*(e+n*(et*i)),this.opacity)}})),t.color=y,t.cubehelix=at,t.gray=function(t,e){return new F(t,0,0,null==e?1:e)},t.hcl=U,t.hsl=$,t.lab=D,t.lch=function(t,e,n,i){return 1===arguments.length?T(t):new V(n,e,t,null==i?1:i)},t.rgb=k,Object.defineProperty(t,"__esModule",{value:!0})});

// https://d3js.org/d3-dispatch/ v1.0.6 Copyright 2019 Mike Bostock
!function(n,e){"object"==typeof exports&&"undefined"!=typeof module?e(exports):"function"==typeof define&&define.amd?define(["exports"],e):e((n=n||self).d3=n.d3||{})}(this,function(n){"use strict";var e={value:function(){}};function t(){for(var n,e=0,t=arguments.length,o={};e<t;++e){if(!(n=arguments[e]+"")||n in o||/[\s.]/.test(n))throw new Error("illegal type: "+n);o[n]=[]}return new r(o)}function r(n){this._=n}function o(n,e){return n.trim().split(/^|\s+/).map(function(n){var t="",r=n.indexOf(".");if(r>=0&&(t=n.slice(r+1),n=n.slice(0,r)),n&&!e.hasOwnProperty(n))throw new Error("unknown type: "+n);return{type:n,name:t}})}function i(n,e){for(var t,r=0,o=n.length;r<o;++r)if((t=n[r]).name===e)return t.value}function f(n,t,r){for(var o=0,i=n.length;o<i;++o)if(n[o].name===t){n[o]=e,n=n.slice(0,o).concat(n.slice(o+1));break}return null!=r&&n.push({name:t,value:r}),n}r.prototype=t.prototype={constructor:r,on:function(n,e){var t,r=this._,l=o(n+"",r),u=-1,a=l.length;if(!(arguments.length<2)){if(null!=e&&"function"!=typeof e)throw new Error("invalid callback: "+e);for(;++u<a;)if(t=(n=l[u]).type)r[t]=f(r[t],n.name,e);else if(null==e)for(t in r)r[t]=f(r[t],n.name,null);return this}for(;++u<a;)if((t=(n=l[u]).type)&&(t=i(r[t],n.name)))return t},copy:function(){var n={},e=this._;for(var t in e)n[t]=e[t].slice();return new r(n)},call:function(n,e){if((t=arguments.length-2)>0)for(var t,r,o=new Array(t),i=0;i<t;++i)o[i]=arguments[i+2];if(!this._.hasOwnProperty(n))throw new Error("unknown type: "+n);for(i=0,t=(r=this._[n]).length;i<t;++i)r[i].value.apply(e,o)},apply:function(n,e,t){if(!this._.hasOwnProperty(n))throw new Error("unknown type: "+n);for(var r=this._[n],o=0,i=r.length;o<i;++o)r[o].value.apply(e,t)}},n.dispatch=t,Object.defineProperty(n,"__esModule",{value:!0})});

// https://d3js.org/d3-ease/ v1.0.7 Copyright 2020 Mike Bostock
!function(n,e){"object"==typeof exports&&"undefined"!=typeof module?e(exports):"function"==typeof define&&define.amd?define(["exports"],e):e((n=n||self).d3=n.d3||{})}(this,function(n){"use strict";function e(n){return((n*=2)<=1?n*n:--n*(2-n)+1)/2}function t(n){return((n*=2)<=1?n*n*n:(n-=2)*n*n+2)/2}var u=function n(e){function t(n){return Math.pow(n,e)}return e=+e,t.exponent=n,t}(3),r=function n(e){function t(n){return 1-Math.pow(1-n,e)}return e=+e,t.exponent=n,t}(3),a=function n(e){function t(n){return((n*=2)<=1?Math.pow(n,e):2-Math.pow(2-n,e))/2}return e=+e,t.exponent=n,t}(3),o=Math.PI,i=o/2;function c(n){return(1-Math.cos(o*n))/2}function s(n){return 1.0009775171065494*(Math.pow(2,-10*n)-.0009765625)}function f(n){return((n*=2)<=1?s(1-n):2-s(n-1))/2}function h(n){return((n*=2)<=1?1-Math.sqrt(1-n*n):Math.sqrt(1-(n-=2)*n)+1)/2}var p=4/11,M=6/11,d=8/11,I=.75,l=9/11,O=10/11,x=.9375,v=21/22,m=63/64,y=1/p/p;function B(n){return(n=+n)<p?y*n*n:n<d?y*(n-=M)*n+I:n<O?y*(n-=l)*n+x:y*(n-=v)*n+m}var C=function n(e){function t(n){return(n=+n)*n*(e*(n-1)+n)}return e=+e,t.overshoot=n,t}(1.70158),E=function n(e){function t(n){return--n*n*((n+1)*e+n)+1}return e=+e,t.overshoot=n,t}(1.70158),P=function n(e){function t(n){return((n*=2)<1?n*n*((e+1)*n-e):(n-=2)*n*((e+1)*n+e)+2)/2}return e=+e,t.overshoot=n,t}(1.70158),b=2*Math.PI,w=function n(e,t){var u=Math.asin(1/(e=Math.max(1,e)))*(t/=b);function r(n){return e*s(- --n)*Math.sin((u-n)/t)}return r.amplitude=function(e){return n(e,t*b)},r.period=function(t){return n(e,t)},r}(1,.3),k=function n(e,t){var u=Math.asin(1/(e=Math.max(1,e)))*(t/=b);function r(n){return 1-e*s(n=+n)*Math.sin((n+u)/t)}return r.amplitude=function(e){return n(e,t*b)},r.period=function(t){return n(e,t)},r}(1,.3),q=function n(e,t){var u=Math.asin(1/(e=Math.max(1,e)))*(t/=b);function r(n){return((n=2*n-1)<0?e*s(-n)*Math.sin((u-n)/t):2-e*s(n)*Math.sin((u+n)/t))/2}return r.amplitude=function(e){return n(e,t*b)},r.period=function(t){return n(e,t)},r}(1,.3);n.easeBack=P,n.easeBackIn=C,n.easeBackInOut=P,n.easeBackOut=E,n.easeBounce=B,n.easeBounceIn=function(n){return 1-B(1-n)},n.easeBounceInOut=function(n){return((n*=2)<=1?1-B(1-n):B(n-1)+1)/2},n.easeBounceOut=B,n.easeCircle=h,n.easeCircleIn=function(n){return 1-Math.sqrt(1-n*n)},n.easeCircleInOut=h,n.easeCircleOut=function(n){return Math.sqrt(1- --n*n)},n.easeCubic=t,n.easeCubicIn=function(n){return n*n*n},n.easeCubicInOut=t,n.easeCubicOut=function(n){return--n*n*n+1},n.easeElastic=k,n.easeElasticIn=w,n.easeElasticInOut=q,n.easeElasticOut=k,n.easeExp=f,n.easeExpIn=function(n){return s(1-+n)},n.easeExpInOut=f,n.easeExpOut=function(n){return 1-s(n)},n.easeLinear=function(n){return+n},n.easePoly=a,n.easePolyIn=u,n.easePolyInOut=a,n.easePolyOut=r,n.easeQuad=e,n.easeQuadIn=function(n){return n*n},n.easeQuadInOut=e,n.easeQuadOut=function(n){return n*(2-n)},n.easeSin=c,n.easeSinIn=function(n){return 1==+n?1:1-Math.cos(n*i)},n.easeSinInOut=c,n.easeSinOut=function(n){return Math.sin(n*i)},Object.defineProperty(n,"__esModule",{value:!0})});

// https://d3js.org/d3-interpolate/ v1.4.0 Copyright 2019 Mike Bostock
!function(t,n){"object"==typeof exports&&"undefined"!=typeof module?n(exports,require("d3-color")):"function"==typeof define&&define.amd?define(["exports","d3-color"],n):n((t=t||self).d3=t.d3||{},t.d3)}(this,function(t,n){"use strict";function r(t,n,r,e,a){var o=t*t,u=o*t;return((1-3*t+3*o-u)*n+(4-6*o+3*u)*r+(1+3*t+3*o-3*u)*e+u*a)/6}function e(t){var n=t.length-1;return function(e){var a=e<=0?e=0:e>=1?(e=1,n-1):Math.floor(e*n),o=t[a],u=t[a+1],i=a>0?t[a-1]:2*o-u,l=a<n-1?t[a+2]:2*u-o;return r((e-a/n)*n,i,o,u,l)}}function a(t){var n=t.length;return function(e){var a=Math.floor(((e%=1)<0?++e:e)*n),o=t[(a+n-1)%n],u=t[a%n],i=t[(a+1)%n],l=t[(a+2)%n];return r((e-a/n)*n,o,u,i,l)}}function o(t){return function(){return t}}function u(t,n){return function(r){return t+r*n}}function i(t,n){var r=n-t;return r?u(t,r>180||r<-180?r-360*Math.round(r/360):r):o(isNaN(t)?n:t)}function l(t){return 1==(t=+t)?c:function(n,r){return r-n?function(t,n,r){return t=Math.pow(t,r),n=Math.pow(n,r)-t,r=1/r,function(e){return Math.pow(t+e*n,r)}}(n,r,t):o(isNaN(n)?r:n)}}function c(t,n){var r=n-t;return r?u(t,r):o(isNaN(t)?n:t)}var f=function t(r){var e=l(r);function a(t,r){var a=e((t=n.rgb(t)).r,(r=n.rgb(r)).r),o=e(t.g,r.g),u=e(t.b,r.b),i=c(t.opacity,r.opacity);return function(n){return t.r=a(n),t.g=o(n),t.b=u(n),t.opacity=i(n),t+""}}return a.gamma=t,a}(1);function s(t){return function(r){var e,a,o=r.length,u=new Array(o),i=new Array(o),l=new Array(o);for(e=0;e<o;++e)a=n.rgb(r[e]),u[e]=a.r||0,i[e]=a.g||0,l[e]=a.b||0;return u=t(u),i=t(i),l=t(l),a.opacity=1,function(t){return a.r=u(t),a.g=i(t),a.b=l(t),a+""}}}var p=s(e),h=s(a);function v(t,n){n||(n=[]);var r,e=t?Math.min(n.length,t.length):0,a=n.slice();return function(o){for(r=0;r<e;++r)a[r]=t[r]*(1-o)+n[r]*o;return a}}function g(t){return ArrayBuffer.isView(t)&&!(t instanceof DataView)}function d(t,n){var r,e=n?n.length:0,a=t?Math.min(e,t.length):0,o=new Array(a),u=new Array(e);for(r=0;r<a;++r)o[r]=X(t[r],n[r]);for(;r<e;++r)u[r]=n[r];return function(t){for(r=0;r<a;++r)u[r]=o[r](t);return u}}function y(t,n){var r=new Date;return t=+t,n=+n,function(e){return r.setTime(t*(1-e)+n*e),r}}function m(t,n){return t=+t,n=+n,function(r){return t*(1-r)+n*r}}function M(t,n){var r,e={},a={};for(r in null!==t&&"object"==typeof t||(t={}),null!==n&&"object"==typeof n||(n={}),n)r in t?e[r]=X(t[r],n[r]):a[r]=n[r];return function(t){for(r in e)a[r]=e[r](t);return a}}var b=/[-+]?(?:\d+\.?\d*|\.?\d+)(?:[eE][-+]?\d+)?/g,x=new RegExp(b.source,"g");function w(t,n){var r,e,a,o=b.lastIndex=x.lastIndex=0,u=-1,i=[],l=[];for(t+="",n+="";(r=b.exec(t))&&(e=x.exec(n));)(a=e.index)>o&&(a=n.slice(o,a),i[u]?i[u]+=a:i[++u]=a),(r=r[0])===(e=e[0])?i[u]?i[u]+=e:i[++u]=e:(i[++u]=null,l.push({i:u,x:m(r,e)})),o=x.lastIndex;return o<n.length&&(a=n.slice(o),i[u]?i[u]+=a:i[++u]=a),i.length<2?l[0]?function(t){return function(n){return t(n)+""}}(l[0].x):function(t){return function(){return t}}(n):(n=l.length,function(t){for(var r,e=0;e<n;++e)i[(r=l[e]).i]=r.x(t);return i.join("")})}function X(t,r){var e,a=typeof r;return null==r||"boolean"===a?o(r):("number"===a?m:"string"===a?(e=n.color(r))?(r=e,f):w:r instanceof n.color?f:r instanceof Date?y:g(r)?v:Array.isArray(r)?d:"function"!=typeof r.valueOf&&"function"!=typeof r.toString||isNaN(r)?M:m)(t,r)}var A,N,C,Y,j=180/Math.PI,q={translateX:0,translateY:0,rotate:0,skewX:0,scaleX:1,scaleY:1};function k(t,n,r,e,a,o){var u,i,l;return(u=Math.sqrt(t*t+n*n))&&(t/=u,n/=u),(l=t*r+n*e)&&(r-=t*l,e-=n*l),(i=Math.sqrt(r*r+e*e))&&(r/=i,e/=i,l/=i),t*e<n*r&&(t=-t,n=-n,l=-l,u=-u),{translateX:a,translateY:o,rotate:Math.atan2(n,t)*j,skewX:Math.atan(l)*j,scaleX:u,scaleY:i}}function D(t,n,r,e){function a(t){return t.length?t.pop()+" ":""}return function(o,u){var i=[],l=[];return o=t(o),u=t(u),function(t,e,a,o,u,i){if(t!==a||e!==o){var l=u.push("translate(",null,n,null,r);i.push({i:l-4,x:m(t,a)},{i:l-2,x:m(e,o)})}else(a||o)&&u.push("translate("+a+n+o+r)}(o.translateX,o.translateY,u.translateX,u.translateY,i,l),function(t,n,r,o){t!==n?(t-n>180?n+=360:n-t>180&&(t+=360),o.push({i:r.push(a(r)+"rotate(",null,e)-2,x:m(t,n)})):n&&r.push(a(r)+"rotate("+n+e)}(o.rotate,u.rotate,i,l),function(t,n,r,o){t!==n?o.push({i:r.push(a(r)+"skewX(",null,e)-2,x:m(t,n)}):n&&r.push(a(r)+"skewX("+n+e)}(o.skewX,u.skewX,i,l),function(t,n,r,e,o,u){if(t!==r||n!==e){var i=o.push(a(o)+"scale(",null,",",null,")");u.push({i:i-4,x:m(t,r)},{i:i-2,x:m(n,e)})}else 1===r&&1===e||o.push(a(o)+"scale("+r+","+e+")")}(o.scaleX,o.scaleY,u.scaleX,u.scaleY,i,l),o=u=null,function(t){for(var n,r=-1,e=l.length;++r<e;)i[(n=l[r]).i]=n.x(t);return i.join("")}}}var R=D(function(t){return"none"===t?q:(A||(A=document.createElement("DIV"),N=document.documentElement,C=document.defaultView),A.style.transform=t,t=C.getComputedStyle(N.appendChild(A),null).getPropertyValue("transform"),N.removeChild(A),k(+(t=t.slice(7,-1).split(","))[0],+t[1],+t[2],+t[3],+t[4],+t[5]))},"px, ","px)","deg)"),S=D(function(t){return null==t?q:(Y||(Y=document.createElementNS("http://www.w3.org/2000/svg","g")),Y.setAttribute("transform",t),(t=Y.transform.baseVal.consolidate())?k((t=t.matrix).a,t.b,t.c,t.d,t.e,t.f):q)},", ",")",")"),V=Math.SQRT2,B=2,E=4,H=1e-12;function I(t){return((t=Math.exp(t))+1/t)/2}function L(t){return function(r,e){var a=t((r=n.hsl(r)).h,(e=n.hsl(e)).h),o=c(r.s,e.s),u=c(r.l,e.l),i=c(r.opacity,e.opacity);return function(t){return r.h=a(t),r.s=o(t),r.l=u(t),r.opacity=i(t),r+""}}}var T=L(i),O=L(c);function P(t){return function(r,e){var a=t((r=n.hcl(r)).h,(e=n.hcl(e)).h),o=c(r.c,e.c),u=c(r.l,e.l),i=c(r.opacity,e.opacity);return function(t){return r.h=a(t),r.c=o(t),r.l=u(t),r.opacity=i(t),r+""}}}var _=P(i),z=P(c);function Q(t){return function r(e){function a(r,a){var o=t((r=n.cubehelix(r)).h,(a=n.cubehelix(a)).h),u=c(r.s,a.s),i=c(r.l,a.l),l=c(r.opacity,a.opacity);return function(t){return r.h=o(t),r.s=u(t),r.l=i(Math.pow(t,e)),r.opacity=l(t),r+""}}return e=+e,a.gamma=r,a}(1)}var Z=Q(i),F=Q(c);t.interpolate=X,t.interpolateArray=function(t,n){return(g(n)?v:d)(t,n)},t.interpolateBasis=e,t.interpolateBasisClosed=a,t.interpolateCubehelix=Z,t.interpolateCubehelixLong=F,t.interpolateDate=y,t.interpolateDiscrete=function(t){var n=t.length;return function(r){return t[Math.max(0,Math.min(n-1,Math.floor(r*n)))]}},t.interpolateHcl=_,t.interpolateHclLong=z,t.interpolateHsl=T,t.interpolateHslLong=O,t.interpolateHue=function(t,n){var r=i(+t,+n);return function(t){var n=r(t);return n-360*Math.floor(n/360)}},t.interpolateLab=function(t,r){var e=c((t=n.lab(t)).l,(r=n.lab(r)).l),a=c(t.a,r.a),o=c(t.b,r.b),u=c(t.opacity,r.opacity);return function(n){return t.l=e(n),t.a=a(n),t.b=o(n),t.opacity=u(n),t+""}},t.interpolateNumber=m,t.interpolateNumberArray=v,t.interpolateObject=M,t.interpolateRgb=f,t.interpolateRgbBasis=p,t.interpolateRgbBasisClosed=h,t.interpolateRound=function(t,n){return t=+t,n=+n,function(r){return Math.round(t*(1-r)+n*r)}},t.interpolateString=w,t.interpolateTransformCss=R,t.interpolateTransformSvg=S,t.interpolateZoom=function(t,n){var r,e,a=t[0],o=t[1],u=t[2],i=n[0],l=n[1],c=n[2],f=i-a,s=l-o,p=f*f+s*s;if(p<H)e=Math.log(c/u)/V,r=function(t){return[a+t*f,o+t*s,u*Math.exp(V*t*e)]};else{var h=Math.sqrt(p),v=(c*c-u*u+E*p)/(2*u*B*h),g=(c*c-u*u-E*p)/(2*c*B*h),d=Math.log(Math.sqrt(v*v+1)-v),y=Math.log(Math.sqrt(g*g+1)-g);e=(y-d)/V,r=function(t){var n,r=t*e,i=I(d),l=u/(B*h)*(i*(n=V*r+d,((n=Math.exp(2*n))-1)/(n+1))-function(t){return((t=Math.exp(t))-1/t)/2}(d));return[a+l*f,o+l*s,u*i/I(V*r+d)]}}return r.duration=1e3*e,r},t.piecewise=function(t,n){for(var r=0,e=n.length-1,a=n[0],o=new Array(e<0?0:e);r<e;)o[r]=t(a,a=n[++r]);return function(t){var n=Math.max(0,Math.min(e-1,Math.floor(t*=e)));return o[n](t-n)}},t.quantize=function(t,n){for(var r=new Array(n),e=0;e<n;++e)r[e]=t(e/(n-1));return r},Object.defineProperty(t,"__esModule",{value:!0})});

// https://d3js.org/d3-timer/ v1.0.10 Copyright 2019 Mike Bostock
!function(t,n){"object"==typeof exports&&"undefined"!=typeof module?n(exports):"function"==typeof define&&define.amd?define(["exports"],n):n((t=t||self).d3=t.d3||{})}(this,function(t){"use strict";var n,e,o=0,i=0,r=0,u=1e3,l=0,c=0,f=0,a="object"==typeof performance&&performance.now?performance:Date,s="object"==typeof window&&window.requestAnimationFrame?window.requestAnimationFrame.bind(window):function(t){setTimeout(t,17)};function _(){return c||(s(m),c=a.now()+f)}function m(){c=0}function p(){this._call=this._time=this._next=null}function w(t,n,e){var o=new p;return o.restart(t,n,e),o}function d(){_(),++o;for(var t,e=n;e;)(t=c-e._time)>=0&&e._call.call(null,t),e=e._next;--o}function h(){c=(l=a.now())+f,o=i=0;try{d()}finally{o=0,function(){var t,o,i=n,r=1/0;for(;i;)i._call?(r>i._time&&(r=i._time),t=i,i=i._next):(o=i._next,i._next=null,i=t?t._next=o:n=o);e=t,v(r)}(),c=0}}function y(){var t=a.now(),n=t-l;n>u&&(f-=n,l=t)}function v(t){o||(i&&(i=clearTimeout(i)),t-c>24?(t<1/0&&(i=setTimeout(h,t-a.now()-f)),r&&(r=clearInterval(r))):(r||(l=a.now(),r=setInterval(y,u)),o=1,s(h)))}p.prototype=w.prototype={constructor:p,restart:function(t,o,i){if("function"!=typeof t)throw new TypeError("callback is not a function");i=(null==i?_():+i)+(null==o?0:+o),this._next||e===this||(e?e._next=this:n=this,e=this),this._call=t,this._time=i,v()},stop:function(){this._call&&(this._call=null,this._time=1/0,v())}},t.interval=function(t,n,e){var o=new p,i=n;return null==n?(o.restart(t,n,e),o):(n=+n,e=null==e?_():+e,o.restart(function r(u){u+=i,o.restart(r,i+=n,e),t(u)},n,e),o)},t.now=_,t.timeout=function(t,n,e){var o=new p;return n=null==n?0:+n,o.restart(function(e){o.stop(),t(e+n)},n,e),o},t.timer=w,t.timerFlush=d,Object.defineProperty(t,"__esModule",{value:!0})});

// https://d3js.org/d3-transition/ v1.3.2 Copyright 2019 Mike Bostock
!function(t,n){"object"==typeof exports&&"undefined"!=typeof module?n(exports,require("d3-selection"),require("d3-dispatch"),require("d3-timer"),require("d3-interpolate"),require("d3-color"),require("d3-ease")):"function"==typeof define&&define.amd?define(["exports","d3-selection","d3-dispatch","d3-timer","d3-interpolate","d3-color","d3-ease"],n):n((t=t||self).d3=t.d3||{},t.d3,t.d3,t.d3,t.d3,t.d3,t.d3)}(this,function(t,n,e,r,i,o,u){"use strict";var a=e.dispatch("start","end","cancel","interrupt"),s=[],l=0,f=1,c=2,h=3,d=4,_=5,v=6;function p(t,n,e,i,o,u){var p=t.__transition;if(p){if(e in p)return}else t.__transition={};!function(t,n,e){var i,o=t.__transition;function u(l){var _,p,y,m;if(e.state!==f)return s();for(_ in o)if((m=o[_]).name===e.name){if(m.state===h)return r.timeout(u);m.state===d?(m.state=v,m.timer.stop(),m.on.call("interrupt",t,t.__data__,m.index,m.group),delete o[_]):+_<n&&(m.state=v,m.timer.stop(),m.on.call("cancel",t,t.__data__,m.index,m.group),delete o[_])}if(r.timeout(function(){e.state===h&&(e.state=d,e.timer.restart(a,e.delay,e.time),a(l))}),e.state=c,e.on.call("start",t,t.__data__,e.index,e.group),e.state===c){for(e.state=h,i=new Array(y=e.tween.length),_=0,p=-1;_<y;++_)(m=e.tween[_].value.call(t,t.__data__,e.index,e.group))&&(i[++p]=m);i.length=p+1}}function a(n){for(var r=n<e.duration?e.ease.call(null,n/e.duration):(e.timer.restart(s),e.state=_,1),o=-1,u=i.length;++o<u;)i[o].call(t,r);e.state===_&&(e.on.call("end",t,t.__data__,e.index,e.group),s())}function s(){for(var r in e.state=v,e.timer.stop(),delete o[n],o)return;delete t.__transition}o[n]=e,e.timer=r.timer(function(t){e.state=f,e.timer.restart(u,e.delay,e.time),e.delay<=t&&u(t-e.delay)},0,e.time)}(t,e,{name:n,index:i,group:o,on:a,tween:s,time:u.time,delay:u.delay,duration:u.duration,ease:u.ease,timer:null,state:l})}function y(t,n){var e=w(t,n);if(e.state>l)throw new Error("too late; already scheduled");return e}function m(t,n){var e=w(t,n);if(e.state>h)throw new Error("too late; already running");return e}function w(t,n){var e=t.__transition;if(!e||!(e=e[n]))throw new Error("transition not found");return e}function g(t,n){var e,r,i,o=t.__transition,u=!0;if(o){for(i in n=null==n?null:n+"",o)(e=o[i]).name===n?(r=e.state>c&&e.state<_,e.state=v,e.timer.stop(),e.on.call(r?"interrupt":"cancel",t,t.__data__,e.index,e.group),delete o[i]):u=!1;u&&delete t.__transition}}function b(t,n,e){var r=t._id;return t.each(function(){var t=m(this,r);(t.value||(t.value={}))[n]=e.apply(this,arguments)}),function(t){return w(t,r).value[n]}}function x(t,n){var e;return("number"==typeof n?i.interpolateNumber:n instanceof o.color?i.interpolateRgb:(e=o.color(n))?(n=e,i.interpolateRgb):i.interpolateString)(t,n)}var A=n.selection.prototype.constructor;function E(t){return function(){this.style.removeProperty(t)}}var T=0;function N(t,n,e,r){this._groups=t,this._parents=n,this._name=e,this._id=r}function S(t){return n.selection().transition(t)}function q(){return++T}var C=n.selection.prototype;N.prototype=S.prototype={constructor:N,select:function(t){var e=this._name,r=this._id;"function"!=typeof t&&(t=n.selector(t));for(var i=this._groups,o=i.length,u=new Array(o),a=0;a<o;++a)for(var s,l,f=i[a],c=f.length,h=u[a]=new Array(c),d=0;d<c;++d)(s=f[d])&&(l=t.call(s,s.__data__,d,f))&&("__data__"in s&&(l.__data__=s.__data__),h[d]=l,p(h[d],e,r,d,h,w(s,r)));return new N(u,this._parents,e,r)},selectAll:function(t){var e=this._name,r=this._id;"function"!=typeof t&&(t=n.selectorAll(t));for(var i=this._groups,o=i.length,u=[],a=[],s=0;s<o;++s)for(var l,f=i[s],c=f.length,h=0;h<c;++h)if(l=f[h]){for(var d,_=t.call(l,l.__data__,h,f),v=w(l,r),y=0,m=_.length;y<m;++y)(d=_[y])&&p(d,e,r,y,_,v);u.push(_),a.push(l)}return new N(u,a,e,r)},filter:function(t){"function"!=typeof t&&(t=n.matcher(t));for(var e=this._groups,r=e.length,i=new Array(r),o=0;o<r;++o)for(var u,a=e[o],s=a.length,l=i[o]=[],f=0;f<s;++f)(u=a[f])&&t.call(u,u.__data__,f,a)&&l.push(u);return new N(i,this._parents,this._name,this._id)},merge:function(t){if(t._id!==this._id)throw new Error;for(var n=this._groups,e=t._groups,r=n.length,i=e.length,o=Math.min(r,i),u=new Array(r),a=0;a<o;++a)for(var s,l=n[a],f=e[a],c=l.length,h=u[a]=new Array(c),d=0;d<c;++d)(s=l[d]||f[d])&&(h[d]=s);for(;a<r;++a)u[a]=n[a];return new N(u,this._parents,this._name,this._id)},selection:function(){return new A(this._groups,this._parents)},transition:function(){for(var t=this._name,n=this._id,e=q(),r=this._groups,i=r.length,o=0;o<i;++o)for(var u,a=r[o],s=a.length,l=0;l<s;++l)if(u=a[l]){var f=w(u,n);p(u,t,e,l,a,{time:f.time+f.delay+f.duration,delay:0,duration:f.duration,ease:f.ease})}return new N(r,this._parents,t,e)},call:C.call,nodes:C.nodes,node:C.node,size:C.size,empty:C.empty,each:C.each,on:function(t,n){var e=this._id;return arguments.length<2?w(this.node(),e).on.on(t):this.each(function(t,n,e){var r,i,o=function(t){return(t+"").trim().split(/^|\s+/).every(function(t){var n=t.indexOf(".");return n>=0&&(t=t.slice(0,n)),!t||"start"===t})}(n)?y:m;return function(){var u=o(this,t),a=u.on;a!==r&&(i=(r=a).copy()).on(n,e),u.on=i}}(e,t,n))},attr:function(t,e){var r=n.namespace(t),o="transform"===r?i.interpolateTransformSvg:x;return this.attrTween(t,"function"==typeof e?(r.local?function(t,n,e){var r,i,o;return function(){var u,a,s=e(this);if(null!=s)return(u=this.getAttributeNS(t.space,t.local))===(a=s+"")?null:u===r&&a===i?o:(i=a,o=n(r=u,s));this.removeAttributeNS(t.space,t.local)}}:function(t,n,e){var r,i,o;return function(){var u,a,s=e(this);if(null!=s)return(u=this.getAttribute(t))===(a=s+"")?null:u===r&&a===i?o:(i=a,o=n(r=u,s));this.removeAttribute(t)}})(r,o,b(this,"attr."+t,e)):null==e?(r.local?function(t){return function(){this.removeAttributeNS(t.space,t.local)}}:function(t){return function(){this.removeAttribute(t)}})(r):(r.local?function(t,n,e){var r,i,o=e+"";return function(){var u=this.getAttributeNS(t.space,t.local);return u===o?null:u===r?i:i=n(r=u,e)}}:function(t,n,e){var r,i,o=e+"";return function(){var u=this.getAttribute(t);return u===o?null:u===r?i:i=n(r=u,e)}})(r,o,e))},attrTween:function(t,e){var r="attr."+t;if(arguments.length<2)return(r=this.tween(r))&&r._value;if(null==e)return this.tween(r,null);if("function"!=typeof e)throw new Error;var i=n.namespace(t);return this.tween(r,(i.local?function(t,n){var e,r;function i(){var i=n.apply(this,arguments);return i!==r&&(e=(r=i)&&function(t,n){return function(e){this.setAttributeNS(t.space,t.local,n.call(this,e))}}(t,i)),e}return i._value=n,i}:function(t,n){var e,r;function i(){var i=n.apply(this,arguments);return i!==r&&(e=(r=i)&&function(t,n){return function(e){this.setAttribute(t,n.call(this,e))}}(t,i)),e}return i._value=n,i})(i,e))},style:function(t,e,r){var o="transform"==(t+="")?i.interpolateTransformCss:x;return null==e?this.styleTween(t,function(t,e){var r,i,o;return function(){var u=n.style(this,t),a=(this.style.removeProperty(t),n.style(this,t));return u===a?null:u===r&&a===i?o:o=e(r=u,i=a)}}(t,o)).on("end.style."+t,E(t)):"function"==typeof e?this.styleTween(t,function(t,e,r){var i,o,u;return function(){var a=n.style(this,t),s=r(this),l=s+"";return null==s&&(this.style.removeProperty(t),l=s=n.style(this,t)),a===l?null:a===i&&l===o?u:(o=l,u=e(i=a,s))}}(t,o,b(this,"style."+t,e))).each(function(t,n){var e,r,i,o,u="style."+n,a="end."+u;return function(){var s=m(this,t),l=s.on,f=null==s.value[u]?o||(o=E(n)):void 0;l===e&&i===f||(r=(e=l).copy()).on(a,i=f),s.on=r}}(this._id,t)):this.styleTween(t,function(t,e,r){var i,o,u=r+"";return function(){var a=n.style(this,t);return a===u?null:a===i?o:o=e(i=a,r)}}(t,o,e),r).on("end.style."+t,null)},styleTween:function(t,n,e){var r="style."+(t+="");if(arguments.length<2)return(r=this.tween(r))&&r._value;if(null==n)return this.tween(r,null);if("function"!=typeof n)throw new Error;return this.tween(r,function(t,n,e){var r,i;function o(){var o=n.apply(this,arguments);return o!==i&&(r=(i=o)&&function(t,n,e){return function(r){this.style.setProperty(t,n.call(this,r),e)}}(t,o,e)),r}return o._value=n,o}(t,n,null==e?"":e))},text:function(t){return this.tween("text","function"==typeof t?function(t){return function(){var n=t(this);this.textContent=null==n?"":n}}(b(this,"text",t)):function(t){return function(){this.textContent=t}}(null==t?"":t+""))},textTween:function(t){var n="text";if(arguments.length<1)return(n=this.tween(n))&&n._value;if(null==t)return this.tween(n,null);if("function"!=typeof t)throw new Error;return this.tween(n,function(t){var n,e;function r(){var r=t.apply(this,arguments);return r!==e&&(n=(e=r)&&function(t){return function(n){this.textContent=t.call(this,n)}}(r)),n}return r._value=t,r}(t))},remove:function(){return this.on("end.remove",function(t){return function(){var n=this.parentNode;for(var e in this.__transition)if(+e!==t)return;n&&n.removeChild(this)}}(this._id))},tween:function(t,n){var e=this._id;if(t+="",arguments.length<2){for(var r,i=w(this.node(),e).tween,o=0,u=i.length;o<u;++o)if((r=i[o]).name===t)return r.value;return null}return this.each((null==n?function(t,n){var e,r;return function(){var i=m(this,t),o=i.tween;if(o!==e)for(var u=0,a=(r=e=o).length;u<a;++u)if(r[u].name===n){(r=r.slice()).splice(u,1);break}i.tween=r}}:function(t,n,e){var r,i;if("function"!=typeof e)throw new Error;return function(){var o=m(this,t),u=o.tween;if(u!==r){i=(r=u).slice();for(var a={name:n,value:e},s=0,l=i.length;s<l;++s)if(i[s].name===n){i[s]=a;break}s===l&&i.push(a)}o.tween=i}})(e,t,n))},delay:function(t){var n=this._id;return arguments.length?this.each(("function"==typeof t?function(t,n){return function(){y(this,t).delay=+n.apply(this,arguments)}}:function(t,n){return n=+n,function(){y(this,t).delay=n}})(n,t)):w(this.node(),n).delay},duration:function(t){var n=this._id;return arguments.length?this.each(("function"==typeof t?function(t,n){return function(){m(this,t).duration=+n.apply(this,arguments)}}:function(t,n){return n=+n,function(){m(this,t).duration=n}})(n,t)):w(this.node(),n).duration},ease:function(t){var n=this._id;return arguments.length?this.each(function(t,n){if("function"!=typeof n)throw new Error;return function(){m(this,t).ease=n}}(n,t)):w(this.node(),n).ease},end:function(){var t,n,e=this,r=e._id,i=e.size();return new Promise(function(o,u){var a={value:u},s={value:function(){0==--i&&o()}};e.each(function(){var e=m(this,r),i=e.on;i!==t&&((n=(t=i).copy())._.cancel.push(a),n._.interrupt.push(a),n._.end.push(s)),e.on=n})})}};var P={time:null,delay:0,duration:250,ease:u.easeCubicInOut};function z(t,n){for(var e;!(e=t.__transition)||!(e=e[n]);)if(!(t=t.parentNode))return P.time=r.now(),P;return e}n.selection.prototype.interrupt=function(t){return this.each(function(){g(this,t)})},n.selection.prototype.transition=function(t){var n,e;t instanceof N?(n=t._id,t=t._name):(n=q(),(e=P).time=r.now(),t=null==t?null:t+"");for(var i=this._groups,o=i.length,u=0;u<o;++u)for(var a,s=i[u],l=s.length,f=0;f<l;++f)(a=s[f])&&p(a,t,n,f,s,e||z(a,n));return new N(i,this._parents,t,n)};var O=[null];t.active=function(t,n){var e,r,i=t.__transition;if(i)for(r in n=null==n?null:n+"",i)if((e=i[r]).state>f&&e.name===n)return new N([[t]],O,n,+r);return null},t.interrupt=g,t.transition=S,Object.defineProperty(t,"__esModule",{value:!0})});

// https://d3js.org/d3-drag/ v1.2.5 Copyright 2019 Mike Bostock
!function(t,e){"object"==typeof exports&&"undefined"!=typeof module?e(exports,require("d3-dispatch"),require("d3-selection")):"function"==typeof define&&define.amd?define(["exports","d3-dispatch","d3-selection"],e):e((t=t||self).d3=t.d3||{},t.d3,t.d3)}(this,function(t,e,n){"use strict";function o(){n.event.stopImmediatePropagation()}function i(){n.event.preventDefault(),n.event.stopImmediatePropagation()}function r(t){var e=t.document.documentElement,o=n.select(t).on("dragstart.drag",i,!0);"onselectstart"in e?o.on("selectstart.drag",i,!0):(e.__noselect=e.style.MozUserSelect,e.style.MozUserSelect="none")}function c(t,e){var o=t.document.documentElement,r=n.select(t).on("dragstart.drag",null);e&&(r.on("click.drag",i,!0),setTimeout(function(){r.on("click.drag",null)},0)),"onselectstart"in o?r.on("selectstart.drag",null):(o.style.MozUserSelect=o.__noselect,delete o.__noselect)}function u(t){return function(){return t}}function s(t,e,n,o,i,r,c,u,s,a){this.target=t,this.type=e,this.subject=n,this.identifier=o,this.active=i,this.x=r,this.y=c,this.dx=u,this.dy=s,this._=a}function a(){return!n.event.ctrlKey&&!n.event.button}function l(){return this.parentNode}function d(t){return null==t?{x:n.event.x,y:n.event.y}:t}function f(){return navigator.maxTouchPoints||"ontouchstart"in this}s.prototype.on=function(){var t=this._.on.apply(this._,arguments);return t===this._?this:t},t.drag=function(){var t,h,v,p,g=a,m=l,y=d,b=f,_={},w=e.dispatch("start","drag","end"),x=0,T=0;function j(t){t.on("mousedown.drag",k).filter(b).on("touchstart.drag",P).on("touchmove.drag",q).on("touchend.drag touchcancel.drag",z).style("touch-action","none").style("-webkit-tap-highlight-color","rgba(0,0,0,0)")}function k(){if(!p&&g.apply(this,arguments)){var e=D("mouse",m.apply(this,arguments),n.mouse,this,arguments);e&&(n.select(n.event.view).on("mousemove.drag",E,!0).on("mouseup.drag",M,!0),r(n.event.view),o(),v=!1,t=n.event.clientX,h=n.event.clientY,e("start"))}}function E(){if(i(),!v){var e=n.event.clientX-t,o=n.event.clientY-h;v=e*e+o*o>T}_.mouse("drag")}function M(){n.select(n.event.view).on("mousemove.drag mouseup.drag",null),c(n.event.view,v),i(),_.mouse("end")}function P(){if(g.apply(this,arguments)){var t,e,i=n.event.changedTouches,r=m.apply(this,arguments),c=i.length;for(t=0;t<c;++t)(e=D(i[t].identifier,r,n.touch,this,arguments))&&(o(),e("start"))}}function q(){var t,e,o=n.event.changedTouches,r=o.length;for(t=0;t<r;++t)(e=_[o[t].identifier])&&(i(),e("drag"))}function z(){var t,e,i=n.event.changedTouches,r=i.length;for(p&&clearTimeout(p),p=setTimeout(function(){p=null},500),t=0;t<r;++t)(e=_[i[t].identifier])&&(o(),e("end"))}function D(t,e,o,i,r){var c,u,a,l=o(e,t),d=w.copy();if(n.customEvent(new s(j,"beforestart",c,t,x,l[0],l[1],0,0,d),function(){return null!=(n.event.subject=c=y.apply(i,r))&&(u=c.x-l[0]||0,a=c.y-l[1]||0,!0)}))return function f(h){var v,p=l;switch(h){case"start":_[t]=f,v=x++;break;case"end":delete _[t],--x;case"drag":l=o(e,t),v=x}n.customEvent(new s(j,h,c,t,v,l[0]+u,l[1]+a,l[0]-p[0],l[1]-p[1],d),d.apply,d,[h,i,r])}}return j.filter=function(t){return arguments.length?(g="function"==typeof t?t:u(!!t),j):g},j.container=function(t){return arguments.length?(m="function"==typeof t?t:u(t),j):m},j.subject=function(t){return arguments.length?(y="function"==typeof t?t:u(t),j):y},j.touchable=function(t){return arguments.length?(b="function"==typeof t?t:u(!!t),j):b},j.on=function(){var t=w.on.apply(w,arguments);return t===w?j:t},j.clickDistance=function(t){return arguments.length?(T=(t=+t)*t,j):Math.sqrt(T)},j},t.dragDisable=r,t.dragEnable=c,Object.defineProperty(t,"__esModule",{value:!0})});

// https://d3js.org/d3-zoom/ v1.8.3 Copyright 2019 Mike Bostock
!function(t,n){"object"==typeof exports&&"undefined"!=typeof module?n(exports,require("d3-dispatch"),require("d3-drag"),require("d3-interpolate"),require("d3-selection"),require("d3-transition")):"function"==typeof define&&define.amd?define(["exports","d3-dispatch","d3-drag","d3-interpolate","d3-selection","d3-transition"],n):n((t=t||self).d3=t.d3||{},t.d3,t.d3,t.d3,t.d3,t.d3)}(this,function(t,n,e,o,i,u){"use strict";function r(t){return function(){return t}}function s(t,n,e){this.target=t,this.type=n,this.transform=e}function h(t,n,e){this.k=t,this.x=n,this.y=e}h.prototype={constructor:h,scale:function(t){return 1===t?this:new h(this.k*t,this.x,this.y)},translate:function(t,n){return 0===t&0===n?this:new h(this.k,this.x+this.k*t,this.y+this.k*n)},apply:function(t){return[t[0]*this.k+this.x,t[1]*this.k+this.y]},applyX:function(t){return t*this.k+this.x},applyY:function(t){return t*this.k+this.y},invert:function(t){return[(t[0]-this.x)/this.k,(t[1]-this.y)/this.k]},invertX:function(t){return(t-this.x)/this.k},invertY:function(t){return(t-this.y)/this.k},rescaleX:function(t){return t.copy().domain(t.range().map(this.invertX,this).map(t.invert,t))},rescaleY:function(t){return t.copy().domain(t.range().map(this.invertY,this).map(t.invert,t))},toString:function(){return"translate("+this.x+","+this.y+") scale("+this.k+")"}};var c=new h(1,0,0);function a(t){for(;!t.__zoom;)if(!(t=t.parentNode))return c;return t.__zoom}function f(){i.event.stopImmediatePropagation()}function l(){i.event.preventDefault(),i.event.stopImmediatePropagation()}function m(){return!i.event.ctrlKey&&!i.event.button}function p(){var t=this;return t instanceof SVGElement?(t=t.ownerSVGElement||t).hasAttribute("viewBox")?[[(t=t.viewBox.baseVal).x,t.y],[t.x+t.width,t.y+t.height]]:[[0,0],[t.width.baseVal.value,t.height.baseVal.value]]:[[0,0],[t.clientWidth,t.clientHeight]]}function v(){return this.__zoom||c}function d(){return-i.event.deltaY*(1===i.event.deltaMode?.05:i.event.deltaMode?1:.002)}function y(){return navigator.maxTouchPoints||"ontouchstart"in this}function z(t,n,e){var o=t.invertX(n[0][0])-e[0][0],i=t.invertX(n[1][0])-e[1][0],u=t.invertY(n[0][1])-e[0][1],r=t.invertY(n[1][1])-e[1][1];return t.translate(i>o?(o+i)/2:Math.min(0,o)||Math.max(0,i),r>u?(u+r)/2:Math.min(0,u)||Math.max(0,r))}a.prototype=h.prototype,t.zoom=function(){var t,a,_=m,g=p,x=z,k=d,w=y,M=[0,1/0],T=[[-1/0,-1/0],[1/0,1/0]],b=250,Y=o.interpolateZoom,X=n.dispatch("start","zoom","end"),q=500,E=150,V=0;function B(t){t.property("__zoom",v).on("wheel.zoom",K).on("mousedown.zoom",A).on("dblclick.zoom",H).filter(w).on("touchstart.zoom",N).on("touchmove.zoom",O).on("touchend.zoom touchcancel.zoom",W).style("touch-action","none").style("-webkit-tap-highlight-color","rgba(0,0,0,0)")}function D(t,n){return(n=Math.max(M[0],Math.min(M[1],n)))===t.k?t:new h(n,t.x,t.y)}function P(t,n,e){var o=n[0]-e[0]*t.k,i=n[1]-e[1]*t.k;return o===t.x&&i===t.y?t:new h(t.k,o,i)}function I(t){return[(+t[0][0]+ +t[1][0])/2,(+t[0][1]+ +t[1][1])/2]}function S(t,n,e){t.on("start.zoom",function(){j(this,arguments).start()}).on("interrupt.zoom end.zoom",function(){j(this,arguments).end()}).tween("zoom",function(){var t=this,o=arguments,i=j(t,o),u=g.apply(t,o),r=null==e?I(u):"function"==typeof e?e.apply(t,o):e,s=Math.max(u[1][0]-u[0][0],u[1][1]-u[0][1]),c=t.__zoom,a="function"==typeof n?n.apply(t,o):n,f=Y(c.invert(r).concat(s/c.k),a.invert(r).concat(s/a.k));return function(t){if(1===t)t=a;else{var n=f(t),e=s/n[2];t=new h(e,r[0]-n[0]*e,r[1]-n[1]*e)}i.zoom(null,t)}})}function j(t,n,e){return!e&&t.__zooming||new G(t,n)}function G(t,n){this.that=t,this.args=n,this.active=0,this.extent=g.apply(t,n),this.taps=0}function K(){if(_.apply(this,arguments)){var t=j(this,arguments),n=this.__zoom,e=Math.max(M[0],Math.min(M[1],n.k*Math.pow(2,k.apply(this,arguments)))),o=i.mouse(this);if(t.wheel)t.mouse[0][0]===o[0]&&t.mouse[0][1]===o[1]||(t.mouse[1]=n.invert(t.mouse[0]=o)),clearTimeout(t.wheel);else{if(n.k===e)return;t.mouse=[o,n.invert(o)],u.interrupt(this),t.start()}l(),t.wheel=setTimeout(function(){t.wheel=null,t.end()},E),t.zoom("mouse",x(P(D(n,e),t.mouse[0],t.mouse[1]),t.extent,T))}}function A(){if(!a&&_.apply(this,arguments)){var t=j(this,arguments,!0),n=i.select(i.event.view).on("mousemove.zoom",function(){if(l(),!t.moved){var n=i.event.clientX-r,e=i.event.clientY-s;t.moved=n*n+e*e>V}t.zoom("mouse",x(P(t.that.__zoom,t.mouse[0]=i.mouse(t.that),t.mouse[1]),t.extent,T))},!0).on("mouseup.zoom",function(){n.on("mousemove.zoom mouseup.zoom",null),e.dragEnable(i.event.view,t.moved),l(),t.end()},!0),o=i.mouse(this),r=i.event.clientX,s=i.event.clientY;e.dragDisable(i.event.view),f(),t.mouse=[o,this.__zoom.invert(o)],u.interrupt(this),t.start()}}function H(){if(_.apply(this,arguments)){var t=this.__zoom,n=i.mouse(this),e=t.invert(n),o=t.k*(i.event.shiftKey?.5:2),u=x(P(D(t,o),n,e),g.apply(this,arguments),T);l(),b>0?i.select(this).transition().duration(b).call(S,u,n):i.select(this).call(B.transform,u)}}function N(){if(_.apply(this,arguments)){var n,e,o,r,s=i.event.touches,h=s.length,c=j(this,arguments,i.event.changedTouches.length===h);for(f(),e=0;e<h;++e)o=s[e],r=[r=i.touch(this,s,o.identifier),this.__zoom.invert(r),o.identifier],c.touch0?c.touch1||c.touch0[2]===r[2]||(c.touch1=r,c.taps=0):(c.touch0=r,n=!0,c.taps=1+!!t);t&&(t=clearTimeout(t)),n&&(c.taps<2&&(t=setTimeout(function(){t=null},q)),u.interrupt(this),c.start())}}function O(){if(this.__zooming){var n,e,o,u,r=j(this,arguments),s=i.event.changedTouches,h=s.length;for(l(),t&&(t=clearTimeout(t)),r.taps=0,n=0;n<h;++n)e=s[n],o=i.touch(this,s,e.identifier),r.touch0&&r.touch0[2]===e.identifier?r.touch0[0]=o:r.touch1&&r.touch1[2]===e.identifier&&(r.touch1[0]=o);if(e=r.that.__zoom,r.touch1){var c=r.touch0[0],a=r.touch0[1],f=r.touch1[0],m=r.touch1[1],p=(p=f[0]-c[0])*p+(p=f[1]-c[1])*p,v=(v=m[0]-a[0])*v+(v=m[1]-a[1])*v;e=D(e,Math.sqrt(p/v)),o=[(c[0]+f[0])/2,(c[1]+f[1])/2],u=[(a[0]+m[0])/2,(a[1]+m[1])/2]}else{if(!r.touch0)return;o=r.touch0[0],u=r.touch0[1]}r.zoom("touch",x(P(e,o,u),r.extent,T))}}function W(){if(this.__zooming){var t,n,e=j(this,arguments),o=i.event.changedTouches,u=o.length;for(f(),a&&clearTimeout(a),a=setTimeout(function(){a=null},q),t=0;t<u;++t)n=o[t],e.touch0&&e.touch0[2]===n.identifier?delete e.touch0:e.touch1&&e.touch1[2]===n.identifier&&delete e.touch1;if(e.touch1&&!e.touch0&&(e.touch0=e.touch1,delete e.touch1),e.touch0)e.touch0[1]=this.__zoom.invert(e.touch0[0]);else if(e.end(),2===e.taps){var r=i.select(this).on("dblclick.zoom");r&&r.apply(this,arguments)}}}return B.transform=function(t,n,e){var o=t.selection?t.selection():t;o.property("__zoom",v),t!==o?S(t,n,e):o.interrupt().each(function(){j(this,arguments).start().zoom(null,"function"==typeof n?n.apply(this,arguments):n).end()})},B.scaleBy=function(t,n,e){B.scaleTo(t,function(){var t=this.__zoom.k,e="function"==typeof n?n.apply(this,arguments):n;return t*e},e)},B.scaleTo=function(t,n,e){B.transform(t,function(){var t=g.apply(this,arguments),o=this.__zoom,i=null==e?I(t):"function"==typeof e?e.apply(this,arguments):e,u=o.invert(i),r="function"==typeof n?n.apply(this,arguments):n;return x(P(D(o,r),i,u),t,T)},e)},B.translateBy=function(t,n,e){B.transform(t,function(){return x(this.__zoom.translate("function"==typeof n?n.apply(this,arguments):n,"function"==typeof e?e.apply(this,arguments):e),g.apply(this,arguments),T)})},B.translateTo=function(t,n,e,o){B.transform(t,function(){var t=g.apply(this,arguments),i=this.__zoom,u=null==o?I(t):"function"==typeof o?o.apply(this,arguments):o;return x(c.translate(u[0],u[1]).scale(i.k).translate("function"==typeof n?-n.apply(this,arguments):-n,"function"==typeof e?-e.apply(this,arguments):-e),t,T)},o)},G.prototype={start:function(){return 1==++this.active&&(this.that.__zooming=this,this.emit("start")),this},zoom:function(t,n){return this.mouse&&"mouse"!==t&&(this.mouse[1]=n.invert(this.mouse[0])),this.touch0&&"touch"!==t&&(this.touch0[1]=n.invert(this.touch0[0])),this.touch1&&"touch"!==t&&(this.touch1[1]=n.invert(this.touch1[0])),this.that.__zoom=n,this.emit("zoom"),this},end:function(){return 0==--this.active&&(delete this.that.__zooming,this.emit("end")),this},emit:function(t){i.customEvent(new s(B,t,this.that.__zoom),X.apply,X,[t,this.that,this.args])}},B.wheelDelta=function(t){return arguments.length?(k="function"==typeof t?t:r(+t),B):k},B.filter=function(t){return arguments.length?(_="function"==typeof t?t:r(!!t),B):_},B.touchable=function(t){return arguments.length?(w="function"==typeof t?t:r(!!t),B):w},B.extent=function(t){return arguments.length?(g="function"==typeof t?t:r([[+t[0][0],+t[0][1]],[+t[1][0],+t[1][1]]]),B):g},B.scaleExtent=function(t){return arguments.length?(M[0]=+t[0],M[1]=+t[1],B):[M[0],M[1]]},B.translateExtent=function(t){return arguments.length?(T[0][0]=+t[0][0],T[1][0]=+t[1][0],T[0][1]=+t[0][1],T[1][1]=+t[1][1],B):[[T[0][0],T[0][1]],[T[1][0],T[1][1]]]},B.constrain=function(t){return arguments.length?(x=t,B):x},B.duration=function(t){return arguments.length?(b=+t,B):b},B.interpolate=function(t){return arguments.length?(Y=t,B):Y},B.on=function(){var t=X.on.apply(X,arguments);return t===X?B:t},B.clickDistance=function(t){return arguments.length?(V=(t=+t)*t,B):Math.sqrt(V)},B},t.zoomIdentity=c,t.zoomTransform=a,Object.defineProperty(t,"__esModule",{value:!0})});
```
