## Supplementary material for "Evaluating Regional Diversity in Scientific Communication: A Comparative Analysis of COVID-19 Preprints and Peer-Reviewed Publications": d3-lasso.min.js

```
!function(t,n){"object"==typeof exports&&"undefined"!=typeof module?n(exports,require("d3-selection"),require("d3-drag")):"function"==typeof define&&define.amd?define(["exports","d3-selection","d3-drag"],n):n(t.d3=t.d3||{},t.d3,t.d3)}(this,function(t,n,r){"use strict";function e(t,n){return n={exports:{}},t(n,n.exports),n.exports}function o(){function t(t){function u(){p=[],h="",_.attr("d",null),m.attr("d",null),r.nodes().forEach(function(t){t.__lasso.possible=!1,t.__lasso.selected=!1,t.__lasso.hoverSelect=!1,t.__lasso.loopSelect=!1;var n=t.getBoundingClientRect();t.__lasso.lassoPoint=[Math.round(n.left+n.width/2),Math.round(n.top+n.height/2)]}),s&&r.on("mouseover.lasso",function(){this.__lasso.hoverSelect=!0}),i.start()}function l(){var t,n;"touchmove"===d3.event.sourceEvent.type?(t=d3.event.sourceEvent.touches[0].clientX,n=d3.event.sourceEvent.touches[0].clientY):(t=d3.event.sourceEvent.clientX,n=d3.event.sourceEvent.clientY);var s=d3.mouse(this)[0],u=d3.mouse(this)[1];""===h?(h=h+"M "+s+" "+u,v=[t,n],d=[s,u],b.attr("cx",s).attr("cy",u).attr("r",7).attr("display",null)):h=h+" L "+s+" "+u,p.push([t,n]);var l=Math.sqrt(Math.pow(t-v[0],2)+Math.pow(n-v[1],2)),f="M "+s+" "+u+" L "+d[0]+" "+d[1];_.attr("d",h),m.attr("d",f),a=l<=e,a&&o?m.attr("display",null):m.attr("display","none"),r.nodes().forEach(function(t){t.__lasso.loopSelect=!(!a||!o)&&c(p,t.__lasso.lassoPoint)<1,t.__lasso.possible=t.__lasso.hoverSelect||t.__lasso.loopSelect}),i.draw()}function f(){r.on("mouseover.lasso",null),r.nodes().forEach(function(t){t.__lasso.selected=t.__lasso.possible,t.__lasso.possible=!1}),_.attr("d",null),m.attr("d",null),b.attr("display","none"),i.end()}var h,v,d,p,g=t.append("g").attr("class","lasso"),_=g.append("path").attr("class","drawn"),m=g.append("path").attr("class","loop_close"),b=g.append("circle").attr("class","origin"),M=d3.drag().on("start",u).on("drag",l).on("end",f);n.call(M)}var n,r=[],e=75,o=!0,a=!1,s=!0,i={start:function(){},draw:function(){},end:function(){}};return t.items=function(n){if(!arguments.length)return r;r=n;var e=r.nodes();return e.forEach(function(t){t.__lasso={possible:!1,selected:!1}}),t},t.possibleItems=function(){return r.filter(function(){return this.__lasso.possible})},t.selectedItems=function(){return r.filter(function(){return this.__lasso.selected})},t.notPossibleItems=function(){return r.filter(function(){return!this.__lasso.possible})},t.notSelectedItems=function(){return r.filter(function(){return!this.__lasso.selected})},t.closePathDistance=function(n){return arguments.length?(e=n,t):e},t.closePathSelect=function(n){return arguments.length?(o=n===!0,t):o},t.isPathClosed=function(n){return arguments.length?(a=n===!0,t):a},t.hoverSelect=function(n){return arguments.length?(s=n===!0,t):s},t.on=function(n,r){if(!arguments.length)return i;if(1===arguments.length)return i[n];var e=["start","draw","end"];return e.indexOf(n)>-1&&(i[n]=r),t},t.targetArea=function(r){return arguments.length?(n=r,t):n},t}var a=e(function(t){function n(t,n,e){var o=t*n,a=r*t,s=a-t,i=a-s,u=t-i,l=r*n,f=l-n,c=l-f,h=n-c,v=o-i*c,d=v-u*c,p=d-i*h,g=u*h-p;return e?(e[0]=g,e[1]=o,e):[g,o]}t.exports=n;var r=+(Math.pow(2,27)+1)}),s=e(function(t){function n(t,n){var r=t+n,e=r-t,o=r-e,a=n-e,s=t-o,i=s+a;return i?[i,r]:[r]}function r(t,r){var e=0|t.length,o=0|r.length;if(1===e&&1===o)return n(t[0],r[0]);var a,s,i=e+o,u=new Array(i),l=0,f=0,c=0,h=Math.abs,v=t[f],d=h(v),p=r[c],g=h(p);d<g?(s=v,f+=1,f<e&&(v=t[f],d=h(v))):(s=p,c+=1,c<o&&(p=r[c],g=h(p))),f<e&&d<g||c>=o?(a=v,f+=1,f<e&&(v=t[f],d=h(v))):(a=p,c+=1,c<o&&(p=r[c],g=h(p)));for(var _,m,b,M,y,w=a+s,x=w-a,j=s-x,E=j,A=w;f<e&&c<o;)d<g?(a=v,f+=1,f<e&&(v=t[f],d=h(v))):(a=p,c+=1,c<o&&(p=r[c],g=h(p))),s=E,w=a+s,x=w-a,j=s-x,j&&(u[l++]=j),_=A+w,m=_-A,b=_-m,M=w-m,y=A-b,E=y+M,A=_;for(;f<e;)a=v,s=E,w=a+s,x=w-a,j=s-x,j&&(u[l++]=j),_=A+w,m=_-A,b=_-m,M=w-m,y=A-b,E=y+M,A=_,f+=1,f<e&&(v=t[f]);for(;c<o;)a=p,s=E,w=a+s,x=w-a,j=s-x,j&&(u[l++]=j),_=A+w,m=_-A,b=_-m,M=w-m,y=A-b,E=y+M,A=_,c+=1,c<o&&(p=r[c]);return E&&(u[l++]=E),A&&(u[l++]=A),l||(u[l++]=0),u.length=l,u}t.exports=r}),i=e(function(t){function n(t,n,r){var e=t+n,o=e-t,a=e-o,s=n-o,i=t-a;return r?(r[0]=i+s,r[1]=e,r):[i+s,e]}t.exports=n}),u=e(function(t){function n(t,n){var o=t.length;if(1===o){var a=r(t[0],n);return a[0]?a:[a[1]]}var s=new Array(2*o),i=[.1,.1],u=[.1,.1],l=0;r(t[0],n,i),i[0]&&(s[l++]=i[0]);for(var f=1;f<o;++f){r(t[f],n,u);var c=i[1];e(c,u[0],i),i[0]&&(s[l++]=i[0]);var h=u[1],v=i[1],d=h+v,p=d-h,g=v-p;i[1]=d,g&&(s[l++]=g)}return i[1]&&(s[l++]=i[1]),0===l&&(s[l++]=0),s.length=l,s}var r=a,e=i;t.exports=n}),l=e(function(t){function n(t,n){var r=t+n,e=r-t,o=r-e,a=n-e,s=t-o,i=s+a;return i?[i,r]:[r]}function r(t,r){var e=0|t.length,o=0|r.length;if(1===e&&1===o)return n(t[0],-r[0]);var a,s,i=e+o,u=new Array(i),l=0,f=0,c=0,h=Math.abs,v=t[f],d=h(v),p=-r[c],g=h(p);d<g?(s=v,f+=1,f<e&&(v=t[f],d=h(v))):(s=p,c+=1,c<o&&(p=-r[c],g=h(p))),f<e&&d<g||c>=o?(a=v,f+=1,f<e&&(v=t[f],d=h(v))):(a=p,c+=1,c<o&&(p=-r[c],g=h(p)));for(var _,m,b,M,y,w=a+s,x=w-a,j=s-x,E=j,A=w;f<e&&c<o;)d<g?(a=v,f+=1,f<e&&(v=t[f],d=h(v))):(a=p,c+=1,c<o&&(p=-r[c],g=h(p))),s=E,w=a+s,x=w-a,j=s-x,j&&(u[l++]=j),_=A+w,m=_-A,b=_-m,M=w-m,y=A-b,E=y+M,A=_;for(;f<e;)a=v,s=E,w=a+s,x=w-a,j=s-x,j&&(u[l++]=j),_=A+w,m=_-A,b=_-m,M=w-m,y=A-b,E=y+M,A=_,f+=1,f<e&&(v=t[f]);for(;c<o;)a=p,s=E,w=a+s,x=w-a,j=s-x,j&&(u[l++]=j),_=A+w,m=_-A,b=_-m,M=w-m,y=A-b,E=y+M,A=_,c+=1,c<o&&(p=-r[c]);return E&&(u[l++]=E),A&&(u[l++]=A),l||(u[l++]=0),u.length=l,u}t.exports=r}),f=e(function(t){function n(t,n){for(var r=new Array(t.length-1),e=1;e<t.length;++e)for(var o=r[e-1]=new Array(t.length-1),a=0,s=0;a<t.length;++a)a!==n&&(o[s++]=t[e][a]);return r}function r(t){for(var n=new Array(t),r=0;r<t;++r){n[r]=new Array(t);for(var e=0;e<t;++e)n[r][e]=["m",e,"[",t-r-1,"]"].join("")}return n}function e(t){return 1&t?"-":""}function o(t){if(1===t.length)return t[0];if(2===t.length)return["sum(",t[0],",",t[1],")"].join("");var n=t.length>>1;return["sum(",o(t.slice(0,n)),",",o(t.slice(n)),")"].join("")}function i(t){if(2===t.length)return[["sum(prod(",t[0][0],",",t[1][1],"),prod(-",t[0][1],",",t[1][0],"))"].join("")];for(var r=[],a=0;a<t.length;++a)r.push(["scale(",o(i(n(t,a))),",",e(a),t[0][a],")"].join(""));return r}function f(t){for(var e=[],a=[],s=r(t),u=[],l=0;l<t;++l)0===(1&l)?e.push.apply(e,i(n(s,l))):a.push.apply(a,i(n(s,l))),u.push("m"+l);var f=o(e),c=o(a),h="orientation"+t+"Exact",_=["function ",h,"(",u.join(),"){var p=",f,",n=",c,",d=sub(p,n);return d[d.length-1];};return ",h].join(""),m=new Function("sum","prod","scale","sub",_);return m(d,v,p,g)}function c(t){var n=x[t.length];return n||(n=x[t.length]=f(t.length)),n.apply(void 0,t)}function h(){for(;x.length<=_;)x.push(f(x.length));for(var n=[],r=["slow"],e=0;e<=_;++e)n.push("a"+e),r.push("o"+e);for(var o=["function getOrientation(",n.join(),"){switch(arguments.length){case 0:case 1:return 0;"],e=2;e<=_;++e)o.push("case ",e,":return o",e,"(",n.slice(0,e).join(),");");o.push("}var s=new Array(arguments.length);for(var i=0;i<arguments.length;++i){s[i]=arguments[i]};return slow(s);}return getOrientation"),r.push(o.join(""));var a=Function.apply(void 0,r);t.exports=a.apply(void 0,[c].concat(x));for(var e=0;e<=_;++e)t.exports[e]=x[e]}var v=a,d=s,p=u,g=l,_=5,m=1.1102230246251565e-16,b=(3+16*m)*m,M=(7+56*m)*m,y=f(3),w=f(4),x=[function(){return 0},function(){return 0},function(t,n){return n[0]-t[0]},function(t,n,r){var e,o=(t[1]-r[1])*(n[0]-r[0]),a=(t[0]-r[0])*(n[1]-r[1]),s=o-a;if(o>0){if(a<=0)return s;e=o+a}else{if(!(o<0))return s;if(a>=0)return s;e=-(o+a)}var i=b*e;return s>=i||s<=-i?s:y(t,n,r)},function(t,n,r,e){var o=t[0]-e[0],a=n[0]-e[0],s=r[0]-e[0],i=t[1]-e[1],u=n[1]-e[1],l=r[1]-e[1],f=t[2]-e[2],c=n[2]-e[2],h=r[2]-e[2],v=a*l,d=s*u,p=s*i,g=o*l,_=o*u,m=a*i,b=f*(v-d)+c*(p-g)+h*(_-m),y=(Math.abs(v)+Math.abs(d))*Math.abs(f)+(Math.abs(p)+Math.abs(g))*Math.abs(c)+(Math.abs(_)+Math.abs(m))*Math.abs(h),x=M*y;return b>x||-b>x?b:w(t,n,r,e)}];h()}),c=e(function(t){function n(t,n){for(var e=n[0],o=n[1],a=t.length,s=1,i=a,u=0,l=a-1;u<i;l=u++){var f=t[u],c=t[l],h=f[1],v=c[1];if(v<h){if(v<o&&o<h){var d=r(f,c,n);if(0===d)return 0;s^=0<d|0}else if(o===h){var p=t[(u+1)%a],g=p[1];if(h<g){var d=r(f,c,n);if(0===d)return 0;s^=0<d|0}}}else if(h<v){if(h<o&&o<v){var d=r(f,c,n);if(0===d)return 0;s^=d<0|0}else if(o===h){var p=t[(u+1)%a],g=p[1];if(g<h){var d=r(f,c,n);if(0===d)return 0;s^=d<0|0}}}else if(o===h){var _=Math.min(f[0],c[0]),m=Math.max(f[0],c[0]);if(0===u){for(;l>0;){var b=(l+a-1)%a,M=t[b];if(M[1]!==o)break;var y=M[0];_=Math.min(_,y),m=Math.max(m,y),l=b}if(0===l)return _<=e&&e<=m?0:1;i=l+1}for(var w=t[(l+a-1)%a][1];u+1<i;){var M=t[u+1];if(M[1]!==o)break;var y=M[0];_=Math.min(_,y),m=Math.max(m,y),u+=1}if(_<=e&&e<=m)return 0;var x=t[(u+1)%a][1];e<_&&w<o!=x<o&&(s^=1)}}return 2*s-1}t.exports=n;var r=f});t.lasso=o,Object.defineProperty(t,"__esModule",{value:!0})});
```
