## Supplementary material for "Evaluating Regional Diversity in Scientific Communication: A Comparative Analysis of COVID-19 Preprints and Peer-Reviewed Publications": flatbush.min.js

```
!function(t,s){"object"==typeof exports&&"undefined"!=typeof module?module.exports=s():"function"==typeof define&&define.amd?define(s):(t="undefined"!=typeof globalThis?globalThis:t||self).Flatbush=s()}(this,function(){"use strict";class a{constructor(){this.ids=[],this.values=[],this.length=0}clear(){this.length=0}push(t,s){let i=this.length++;for(;0<i;){var e=i-1>>1,h=this.values[e];if(h<=s)break;this.ids[i]=this.ids[e],this.values[i]=h,i=e}this.ids[i]=t,this.values[i]=s}pop(){if(0!==this.length){var t=this.ids[0];if(this.length--,0<this.length){var s=this.ids[0]=this.ids[this.length],h=this.values[0]=this.values[this.length],i=this.length>>1;let e=0;for(;e<i;){let t=1+(e<<1);var r=t+1;let s=this.ids[t],i=this.values[t];var n=this.values[r];if(r<this.length&&n<i&&(t=r,s=this.ids[r],i=n),i>=h)break;this.ids[e]=s,this.values[e]=i,e=t}this.ids[e]=s,this.values[e]=h}return t}}peek(){if(0!==this.length)return this.ids[0]}peekValue(){if(0!==this.length)return this.values[0]}shrink(){this.ids.length=this.values.length=this.length}}const l=[Int8Array,Uint8Array,Uint8ClampedArray,Int16Array,Uint16Array,Int32Array,Uint32Array,Float32Array,Float64Array];function f(t,s,i){return t<s?s-t:t<=i?0:t-i}function x(t,s){let i=0,e=s.length-1;for(;i<e;){var h=i+e>>1;s[h]>t?e=h:i=1+h}return s[i]}function m(t,s,i,e,h){var r=t[e],t=(t[e]=t[h],t[h]=r,4*e),r=4*h,n=s[t],o=s[1+t],a=s[2+t],l=s[3+t],t=(s[t]=s[r],s[1+t]=s[1+r],s[2+t]=s[2+r],s[3+t]=s[3+r],s[r]=n,s[1+r]=o,s[2+r]=a,s[3+r]=l,i[e]);i[e]=i[h],i[h]=t}return class h{static from(t){if(!(t instanceof ArrayBuffer))throw new Error("Data must be an instance of ArrayBuffer.");var[s,i]=new Uint8Array(t,0,2);if(251!==s)throw new Error("Data does not appear to be in a Flatbush format.");if(i>>4!=3)throw new Error(`Got v${i>>4} data when expected v3.`);var[s]=new Uint16Array(t,2,1),[e]=new Uint32Array(t,4,1);return new h(e,s,l[15&i],t)}constructor(t,s=16,i=Float64Array,e){if(void 0===t)throw new Error("Missing required argument: numItems.");if(isNaN(t)||t<=0)throw new Error(`Unpexpected numItems value: ${t}.`);this.numItems=+t,this.nodeSize=Math.min(Math.max(+s,2),65535);let h=t,r=h;for(this._levelBounds=[4*h];h=Math.ceil(h/this.nodeSize),r+=h,this._levelBounds.push(4*r),1!==h;);this.ArrayType=i||Float64Array,this.IndexArrayType=r<16384?Uint16Array:Uint32Array;var n=l.indexOf(this.ArrayType),o=4*r*this.ArrayType.BYTES_PER_ELEMENT;if(n<0)throw new Error(`Unexpected typed array class: ${i}.`);e&&e instanceof ArrayBuffer?(this.data=e,this._boxes=new this.ArrayType(this.data,8,4*r),this._indices=new this.IndexArrayType(this.data,8+o,r),this._pos=4*r,this.minX=this._boxes[this._pos-4],this.minY=this._boxes[this._pos-3],this.maxX=this._boxes[this._pos-2],this.maxY=this._boxes[this._pos-1]):(this.data=new ArrayBuffer(8+o+r*this.IndexArrayType.BYTES_PER_ELEMENT),this._boxes=new this.ArrayType(this.data,8,4*r),this._indices=new this.IndexArrayType(this.data,8+o,r),this._pos=0,this.minX=1/0,this.minY=1/0,this.maxX=-1/0,this.maxY=-1/0,new Uint8Array(this.data,0,2).set([251,48+n]),new Uint16Array(this.data,2,1)[0]=s,new Uint32Array(this.data,4,1)[0]=t),this._queue=new a}add(t,s,i,e){var h=this._pos>>2;return this._indices[h]=h,this._boxes[this._pos++]=t,this._boxes[this._pos++]=s,this._boxes[this._pos++]=i,this._boxes[this._pos++]=e,t<this.minX&&(this.minX=t),s<this.minY&&(this.minY=s),i>this.maxX&&(this.maxX=i),e>this.maxY&&(this.maxY=e),h}finish(){if(this._pos>>2!==this.numItems)throw new Error(`Added ${this._pos>>2} items when expected ${this.numItems}.`);if(this.numItems<=this.nodeSize)this._boxes[this._pos++]=this.minX,this._boxes[this._pos++]=this.minY,this._boxes[this._pos++]=this.maxX,this._boxes[this._pos++]=this.maxY;else{var s,i,e,h,r,n,o=this.maxX-this.minX||1,a=this.maxY-this.minY||1;const p=new Uint32Array(this.numItems);for(let t=0;t<this.numItems;t++){var l=4*t,d=this._boxes[l++],_=this._boxes[l++],u=this._boxes[l++],l=this._boxes[+l],d=Math.floor(65535*((d+u)/2-this.minX)/o),u=Math.floor(65535*((_+l)/2-this.minY)/a);p[t]=(h=e=n=r=i=s=l=_=void 0,e=(_=d^u)|(l=65535^_)>>1,r=(s=65535^(d|u))>>1^l&(i=d&(65535^u))>>1^s,n=_&s>>1^i>>1^i,l=h=_>>1^_,e=(_=e)&_>>2^h&h>>2,r^=_&(s=r)>>2^h&n>>2,n^=h&s>>2^(_^h)&(i=n)>>2,l=h=_&h>>2^h&(_^h)>>2,e=(_=e)&_>>4^h&h>>4,r^=_&(s=r)>>4^h&n>>4,n^=h&s>>4^(_^h)&(i=n)>>4,l=h=_&h>>4^h&(_^h)>>4,r^=e&(s=r)>>8^h&n>>8,h=(l=(n^=h&s>>8^((_=e)^(l=h))&(i=n)>>8)^n>>1)|65535^((e=d^u)|(_=r^r>>1)),e=1431655765&((e=858993459&((e=252645135&((e=16711935&(e|e<<8))|e<<4))|e<<2))|e<<1),((h=1431655765&((h=858993459&((h=252645135&((h=16711935&(h|h<<8))|h<<4))|h<<2))|h<<1))<<1|e)>>>0)}!function i(e,h,r,n,o,a){if(!(Math.floor(n/a)>=Math.floor(o/a))){const l=e[n+o>>1];let t=n-1,s=o+1;for(;;){for(;t++,e[t]<l;);for(;s--,e[s]>l;);if(t>=s)break;m(e,h,r,t,s)}i(e,h,r,n,s,a),i(e,h,r,s+1,o,a)}}(p,this._boxes,this._indices,0,this.numItems-1,this.nodeSize);for(let t=0,r=0;t<this._levelBounds.length-1;t++)for(var f=this._levelBounds[t];r<f;){var x=r;let s=1/0,i=1/0,e=-1/0,h=-1/0;for(let t=0;t<this.nodeSize&&r<f;t++)s=Math.min(s,this._boxes[r++]),i=Math.min(i,this._boxes[r++]),e=Math.max(e,this._boxes[r++]),h=Math.max(h,this._boxes[r++]);this._indices[this._pos>>2]=x,this._boxes[this._pos++]=s,this._boxes[this._pos++]=i,this._boxes[this._pos++]=e,this._boxes[this._pos++]=h}}}search(s,i,e,h,r){if(this._pos!==this._boxes.length)throw new Error("Data not yet indexed - call index.finish().");let n=this._boxes.length-4;const o=[],a=[];for(;void 0!==n;){var l,d=Math.min(n+4*this.nodeSize,x(n,this._levelBounds));for(let t=n;t<d;t+=4)e<this._boxes[t]||h<this._boxes[t+1]||s>this._boxes[t+2]||i>this._boxes[t+3]||(l=0|this._indices[t>>2],n<4*this.numItems?void 0!==r&&!r(l)||a.push(l):o.push(l));n=o.pop()}return a}neighbors(s,i,t=1/0,e=1/0,h){if(this._pos!==this._boxes.length)throw new Error("Data not yet indexed - call index.finish().");let r=this._boxes.length-4;const n=this._queue,o=[];for(var a=e*e;void 0!==r;){var l=Math.min(r+4*this.nodeSize,x(r,this._levelBounds));for(let t=r;t<l;t+=4){var d=0|this._indices[t>>2],_=f(s,this._boxes[t],this._boxes[t+2]),u=f(i,this._boxes[t+1],this._boxes[t+3]),_=_*_+u*u;r<4*this.numItems?void 0!==h&&!h(d)||n.push(1+(d<<1),_):n.push(d<<1,_)}for(;n.length&&1&n.peek();){if(a<n.peekValue())return n.clear(),o;if(o.push(n.pop()>>1),o.length===t)return n.clear(),o}r=n.pop()>>1}return n.clear(),o}}});
```
