## Supplementary material for "Evaluating Regional Diversity in Scientific Communication: A Comparative Analysis of COVID-19 Preprints and Peer-Reviewed Publications": ggiraphjs.min.js

```
!function(t,e){"object"==typeof exports&&"object"==typeof module?module.exports=e(require("d3"),require("Flatbush")):"function"==typeof define&&define.amd?define(["d3","Flatbush"],e):"object"==typeof exports?exports.ggiraphjs=e(require("d3"),require("Flatbush")):t.ggiraphjs=e(t.d3,t.Flatbush)}(window,(function(t,e){return function(t){var e={};function n(r){if(e[r])return e[r].exports;var i=e[r]={i:r,l:!1,exports:{}};return t[r].call(i.exports,i,i.exports,n),i.l=!0,i.exports}return n.m=t,n.c=e,n.d=function(t,e,r){n.o(t,e)||Object.defineProperty(t,e,{enumerable:!0,get:r})},n.r=function(t){"undefined"!=typeof Symbol&&Symbol.toStringTag&&Object.defineProperty(t,Symbol.toStringTag,{value:"Module"}),Object.defineProperty(t,"__esModule",{value:!0})},n.t=function(t,e){if(1&e&&(t=n(t)),8&e)return t;if(4&e&&"object"==typeof t&&t&&t.__esModule)return t;var r=Object.create(null);if(n.r(r),Object.defineProperty(r,"default",{enumerable:!0,value:t}),2&e&&"string"!=typeof t)for(var i in t)n.d(r,i,function(e){return t[e]}.bind(null,i));return r},n.n=function(t){var e=t&&t.__esModule?function(){return t.default}:function(){return t};return n.d(e,"a",e),e},n.o=function(t,e){return Object.prototype.hasOwnProperty.call(t,e)},n.p="",n(n.s=245)}([function(e,n){e.exports=t},function(t,e,n){var r=n(63),i=Function.prototype,o=i.bind,s=i.call,a=r&&o.bind(s,s);t.exports=r?function(t){return t&&a(t)}:function(t){return t&&function(){return s.apply(t,arguments)}}},function(t,e){t.exports=function(t){try{return!!t()}catch(t){return!0}}},function(t,e,n){var r=n(4),i=n(44).f,o=n(23),s=n(18),a=n(87),u=n(120),c=n(93);t.exports=function(t,e){var n,f,l,h,d,p=t.target,y=t.global,v=t.stat;if(n=y?r:v?r[p]||a(p,{}):(r[p]||{}).prototype)for(f in e){if(h=e[f],l=t.dontCallGetSet?(d=i(n,f))&&d.value:n[f],!c(y?f:p+(v?".":"#")+f,t.forced)&&void 0!==l){if(typeof h==typeof l)continue;u(h,l)}(t.sham||l&&l.sham)&&o(h,"sham",!0),s(n,f,h,t)}}},function(t,e,n){(function(e){var n=function(t){return t&&t.Math==Math&&t};t.exports=n("object"==typeof globalThis&&globalThis)||n("object"==typeof window&&window)||n("object"==typeof self&&self)||n("object"==typeof e&&e)||function(){return this}()||Function("return this")()}).call(this,n(166))},function(t,e,n){"use strict";var r,i,o,s=n(153),a=n(8),u=n(4),c=n(7),f=n(13),l=n(10),h=n(32),d=n(35),p=n(23),y=n(18),v=n(11).f,g=n(28),m=n(69),x=n(59),b=n(6),w=n(52),S=n(22),A=S.enforce,E=S.get,O=u.Int8Array,k=O&&O.prototype,T=u.Uint8ClampedArray,I=T&&T.prototype,M=O&&m(O),_=k&&m(k),P=Object.prototype,j=u.TypeError,N=b("toStringTag"),R=w("TYPED_ARRAY_TAG"),V=s&&!!x&&"Opera"!==h(u.opera),C=!1,D={Int8Array:1,Uint8Array:1,Uint8ClampedArray:1,Int16Array:2,Uint16Array:2,Int32Array:4,Uint32Array:4,Float32Array:4,Float64Array:8},z={BigInt64Array:8,BigUint64Array:8},F=function(t){var e=m(t);if(f(e)){var n=E(e);return n&&l(n,"TypedArrayConstructor")?n.TypedArrayConstructor:F(e)}},B=function(t){if(!f(t))return!1;var e=h(t);return l(D,e)||l(z,e)};for(r in D)(o=(i=u[r])&&i.prototype)?A(o).TypedArrayConstructor=i:V=!1;for(r in z)(o=(i=u[r])&&i.prototype)&&(A(o).TypedArrayConstructor=i);if((!V||!c(M)||M===Function.prototype)&&(M=function(){throw j("Incorrect invocation")},V))for(r in D)u[r]&&x(u[r],M);if((!V||!_||_===P)&&(_=M.prototype,V))for(r in D)u[r]&&x(u[r].prototype,_);if(V&&m(I)!==_&&x(I,_),a&&!l(_,N))for(r in C=!0,v(_,N,{get:function(){return f(this)?this[R]:void 0}}),D)u[r]&&p(u[r],R,r);t.exports={NATIVE_ARRAY_BUFFER_VIEWS:V,TYPED_ARRAY_TAG:C&&R,aTypedArray:function(t){if(B(t))return t;throw j("Target is not a typed array")},aTypedArrayConstructor:function(t){if(c(t)&&(!x||g(M,t)))return t;throw j(d(t)+" is not a typed array constructor")},exportTypedArrayMethod:function(t,e,n,r){if(a){if(n)for(var i in D){var o=u[i];if(o&&l(o.prototype,t))try{delete o.prototype[t]}catch(n){try{o.prototype[t]=e}catch(t){}}}_[t]&&!n||y(_,t,n?e:V&&k[t]||e,r)}},exportTypedArrayStaticMethod:function(t,e,n){var r,i;if(a){if(x){if(n)for(r in D)if((i=u[r])&&l(i,t))try{delete i[t]}catch(t){}if(M[t]&&!n)return;try{return y(M,t,n?e:V&&M[t]||e)}catch(t){}}for(r in D)!(i=u[r])||i[t]&&!n||y(i,t,e)}},getTypedArrayConstructor:F,isView:function(t){if(!f(t))return!1;var e=h(t);return"DataView"===e||l(D,e)||l(z,e)},isTypedArray:B,TypedArray:M,TypedArrayPrototype:_}},function(t,e,n){var r=n(4),i=n(38),o=n(10),s=n(52),a=n(40),u=n(112),c=i("wks"),f=r.Symbol,l=f&&f.for,h=u?f:f&&f.withoutSetter||s;t.exports=function(t){if(!o(c,t)||!a&&"string"!=typeof c[t]){var e="Symbol."+t;a&&o(f,t)?c[t]=f[t]:c[t]=u&&l?l(e):h(e)}return c[t]}},function(t,e){t.exports=function(t){return"function"==typeof t}},function(t,e,n){var r=n(2);t.exports=!r((function(){return 7!=Object.defineProperty({},1,{get:function(){return 7}})[1]}))},function(t,e,n){var r=n(63),i=Function.prototype.call;t.exports=r?i.bind(i):function(){return i.apply(i,arguments)}},function(t,e,n){var r=n(1),i=n(16),o=r({}.hasOwnProperty);t.exports=Object.hasOwn||function(t,e){return o(i(t),e)}},function(t,e,n){var r=n(8),i=n(115),o=n(114),s=n(12),a=n(53),u=TypeError,c=Object.defineProperty,f=Object.getOwnPropertyDescriptor;e.f=r?o?function(t,e,n){if(s(t),e=a(e),s(n),"function"==typeof t&&"prototype"===e&&"value"in n&&"writable"in n&&!n.writable){var r=f(t,e);r&&r.writable&&(t[e]=n.value,n={configurable:"configurable"in n?n.configurable:r.configurable,enumerable:"enumerable"in n?n.enumerable:r.enumerable,writable:!1})}return c(t,e,n)}:c:function(t,e,n){if(s(t),e=a(e),s(n),i)try{return c(t,e,n)}catch(t){}if("get"in n||"set"in n)throw u("Accessors not supported");return"value"in n&&(t[e]=n.value),t}},function(t,e,n){var r=n(13),i=String,o=TypeError;t.exports=function(t){if(r(t))return t;throw o(i(t)+" is not an object")}},function(t,e,n){var r=n(7),i="object"==typeof document&&document.all,o=void 0===i&&void 0!==i;t.exports=o?function(t){return"object"==typeof t?null!==t:r(t)||t===i}:function(t){return"object"==typeof t?null!==t:r(t)}},function(t,e,n){var r=n(21);t.exports=function(t){return r(t.length)}},function(t,e,n){var r=n(32),i=String;t.exports=function(t){if("Symbol"===r(t))throw TypeError("Cannot convert a Symbol value to a string");return i(t)}},function(t,e,n){var r=n(25),i=Object;t.exports=function(t){return i(r(t))}},function(t,e,n){var r=n(45),i=n(1),o=n(62),s=n(16),a=n(14),u=n(101),c=i([].push),f=function(t){var e=1==t,n=2==t,i=3==t,f=4==t,l=6==t,h=7==t,d=5==t||l;return function(p,y,v,g){for(var m,x,b=s(p),w=o(b),S=r(y,v),A=a(w),E=0,O=g||u,k=e?O(p,A):n||h?O(p,0):void 0;A>E;E++)if((d||E in w)&&(x=S(m=w[E],E,b),t))if(e)k[E]=x;else if(x)switch(t){case 3:return!0;case 5:return m;case 6:return E;case 2:c(k,m)}else switch(t){case 4:return!1;case 7:c(k,m)}return l?-1:i||f?f:k}};t.exports={forEach:f(0),map:f(1),filter:f(2),some:f(3),every:f(4),find:f(5),findIndex:f(6),filterReject:f(7)}},function(t,e,n){var r=n(7),i=n(11),o=n(171),s=n(87);t.exports=function(t,e,n,a){a||(a={});var u=a.enumerable,c=void 0!==a.name?a.name:e;if(r(n)&&o(n,c,a),a.global)u?t[e]=n:s(e,n);else{try{a.unsafe?t[e]&&(u=!0):delete t[e]}catch(t){}u?t[e]=n:i.f(t,e,{value:n,enumerable:!1,configurable:!a.nonConfigurable,writable:!a.nonWritable})}return t}},function(t,e,n){var r=n(62),i=n(25);t.exports=function(t){return r(i(t))}},function(t,e,n){var r=n(168);t.exports=function(t){var e=+t;return e!=e||0===e?0:r(e)}},function(t,e,n){var r=n(20),i=Math.min;t.exports=function(t){return t>0?i(r(t),9007199254740991):0}},function(t,e,n){var r,i,o,s=n(170),a=n(4),u=n(1),c=n(13),f=n(23),l=n(10),h=n(86),d=n(67),p=n(56),y=a.TypeError,v=a.WeakMap;if(s||h.state){var g=h.state||(h.state=new v),m=u(g.get),x=u(g.has),b=u(g.set);r=function(t,e){if(x(g,t))throw y("Object already initialized");return e.facade=t,b(g,t,e),e},i=function(t){return m(g,t)||{}},o=function(t){return x(g,t)}}else{var w=d("state");p[w]=!0,r=function(t,e){if(l(t,w))throw y("Object already initialized");return e.facade=t,f(t,w,e),e},i=function(t){return l(t,w)?t[w]:{}},o=function(t){return l(t,w)}}t.exports={set:r,get:i,has:o,enforce:function(t){return o(t)?i(t):r(t,{})},getterFor:function(t){return function(e){var n;if(!c(e)||(n=i(e)).type!==t)throw y("Incompatible receiver, "+t+" required");return n}}}},function(t,e,n){var r=n(8),i=n(11),o=n(43);t.exports=r?function(t,e,n){return i.f(t,e,o(1,n))}:function(t,e,n){return t[e]=n,t}},function(t,e,n){var r=n(3),i=n(8),o=n(11).f;r({target:"Object",stat:!0,forced:Object.defineProperty!==o,sham:!i},{defineProperty:o})},function(t,e,n){var r=n(26),i=TypeError;t.exports=function(t){if(r(t))throw i("Can't call method on "+t);return t}},function(t,e){t.exports=function(t){return null==t}},function(t,e,n){var r=n(4),i=n(7),o=function(t){return i(t)?t:void 0};t.exports=function(t,e){return arguments.length<2?o(r[t]):r[t]&&r[t][e]}},function(t,e,n){var r=n(1);t.exports=r({}.isPrototypeOf)},function(t,e,n){var r=n(97),i=n(18),o=n(181);r||i(Object.prototype,"toString",o,{unsafe:!0})},function(t,e,n){var r,i=n(12),o=n(113),s=n(90),a=n(56),u=n(169),c=n(88),f=n(67),l=f("IE_PROTO"),h=function(){},d=function(t){return"<script>"+t+"<\/script>"},p=function(t){t.write(d("")),t.close();var e=t.parentWindow.Object;return t=null,e},y=function(){try{r=new ActiveXObject("htmlfile")}catch(t){}var t,e;y="undefined"!=typeof document?document.domain&&r?p(r):((e=c("iframe")).style.display="none",u.appendChild(e),e.src=String("javascript:"),(t=e.contentWindow.document).open(),t.write(d("document.F=Object")),t.close(),t.F):p(r);for(var n=s.length;n--;)delete y.prototype[s[n]];return y()};a[l]=!0,t.exports=Object.create||function(t,e){var n;return null!==t?(h.prototype=i(t),n=new h,h.prototype=null,n[l]=t):n=y(),void 0===e?n:o.f(n,e)}},function(t,e,n){var r=n(20),i=Math.max,o=Math.min;t.exports=function(t,e){var n=r(t);return n<0?i(n+e,0):o(n,e)}},function(t,e,n){var r=n(97),i=n(7),o=n(34),s=n(6)("toStringTag"),a=Object,u="Arguments"==o(function(){return arguments}());t.exports=r?o:function(t){var e,n,r;return void 0===t?"Undefined":null===t?"Null":"string"==typeof(n=function(t,e){try{return t[e]}catch(t){}}(e=a(t),s))?n:u?o(e):"Object"==(r=o(e))&&i(e.callee)?"Arguments":r}},function(t,e,n){"use strict";var r=n(3),i=n(133);r({target:"Array",proto:!0,forced:[].forEach!=i},{forEach:i})},function(t,e,n){var r=n(1),i=r({}.toString),o=r("".slice);t.exports=function(t){return o(i(t),8,-1)}},function(t,e){var n=String;t.exports=function(t){try{return n(t)}catch(t){return"Object"}}},function(t,e,n){var r=n(117),i=n(90).concat("length","prototype");e.f=Object.getOwnPropertyNames||function(t){return r(t,i)}},function(t,e,n){var r=n(4),i=n(131),o=n(132),s=n(133),a=n(23),u=function(t){if(t&&t.forEach!==s)try{a(t,"forEach",s)}catch(e){t.forEach=s}};for(var c in i)i[c]&&u(r[c]&&r[c].prototype);u(o)},function(t,e,n){var r=n(39),i=n(86);(t.exports=function(t,e){return i[t]||(i[t]=void 0!==e?e:{})})("versions",[]).push({version:"3.25.1",mode:r?"pure":"global",copyright:"© 2014-2022 Denis Pushkarev (zloirock.ru)",license:"https://github.com/zloirock/core-js/blob/v3.25.1/LICENSE",source:"https://github.com/zloirock/core-js"})},function(t,e){t.exports=!1},function(t,e,n){var r=n(65),i=n(2);t.exports=!!Object.getOwnPropertySymbols&&!i((function(){var t=Symbol();return!String(t)||!(Object(t)instanceof Symbol)||!Symbol.sham&&r&&r<41}))},function(t,e,n){var r=n(42),i=n(26);t.exports=function(t,e){var n=t[e];return i(n)?void 0:r(n)}},function(t,e,n){var r=n(7),i=n(35),o=TypeError;t.exports=function(t){if(r(t))return t;throw o(i(t)+" is not a function")}},function(t,e){t.exports=function(t,e){return{enumerable:!(1&t),configurable:!(2&t),writable:!(4&t),value:e}}},function(t,e,n){var r=n(8),i=n(9),o=n(118),s=n(43),a=n(19),u=n(53),c=n(10),f=n(115),l=Object.getOwnPropertyDescriptor;e.f=r?l:function(t,e){if(t=a(t),e=u(e),f)try{return l(t,e)}catch(t){}if(c(t,e))return s(!i(o.f,t,e),t[e])}},function(t,e,n){var r=n(1),i=n(42),o=n(63),s=r(r.bind);t.exports=function(t,e){return i(t),void 0===e?t:o?s(t,e):function(){return t.apply(e,arguments)}}},function(t,e,n){var r=n(34);t.exports=Array.isArray||function(t){return"Array"==r(t)}},function(t,e,n){"use strict";var r=n(2);t.exports=function(t,e){var n=[][t];return!!n&&r((function(){n.call(null,e||function(){return 1},1)}))}},function(t,e,n){"use strict";var r=n(3),i=n(76);r({target:"RegExp",proto:!0,forced:/./.exec!==i},{exec:i})},function(t,e,n){var r=n(63),i=Function.prototype,o=i.apply,s=i.call;t.exports="object"==typeof Reflect&&Reflect.apply||(r?s.bind(o):function(){return s.apply(o,arguments)})},function(t,e,n){"use strict";var r=n(3),i=n(1),o=n(55).indexOf,s=n(47),a=i([].indexOf),u=!!a&&1/a([1],1,-0)<0,c=s("indexOf");r({target:"Array",proto:!0,forced:u||!c},{indexOf:function(t){var e=arguments.length>1?arguments[1]:void 0;return u?a(this,t,e)||0:o(this,t,e)}})},function(t,e,n){"use strict";var r=n(19),i=n(64),o=n(57),s=n(22),a=n(11).f,u=n(91),c=n(94),f=n(39),l=n(8),h=s.set,d=s.getterFor("Array Iterator");t.exports=u(Array,"Array",(function(t,e){h(this,{type:"Array Iterator",target:r(t),index:0,kind:e})}),(function(){var t=d(this),e=t.target,n=t.kind,r=t.index++;return!e||r>=e.length?(t.target=void 0,c(void 0,!0)):c("keys"==n?r:"values"==n?e[r]:[r,e[r]],!1)}),"values");var p=o.Arguments=o.Array;if(i("keys"),i("values"),i("entries"),!f&&l&&"values"!==p.name)try{a(p,"name",{value:"values"})}catch(t){}},function(t,e,n){var r=n(1),i=0,o=Math.random(),s=r(1..toString);t.exports=function(t){return"Symbol("+(void 0===t?"":t)+")_"+s(++i+o,36)}},function(t,e,n){var r=n(116),i=n(54);t.exports=function(t){var e=r(t,"string");return i(e)?e:e+""}},function(t,e,n){var r=n(27),i=n(7),o=n(28),s=n(112),a=Object;t.exports=s?function(t){return"symbol"==typeof t}:function(t){var e=r("Symbol");return i(e)&&o(e.prototype,a(t))}},function(t,e,n){var r=n(19),i=n(31),o=n(14),s=function(t){return function(e,n,s){var a,u=r(e),c=o(u),f=i(s,c);if(t&&n!=n){for(;c>f;)if((a=u[f++])!=a)return!0}else for(;c>f;f++)if((t||f in u)&&u[f]===n)return t||f||0;return!t&&-1}};t.exports={includes:s(!0),indexOf:s(!1)}},function(t,e){t.exports={}},function(t,e){t.exports={}},function(t,e,n){var r=n(11).f,i=n(10),o=n(6)("toStringTag");t.exports=function(t,e,n){t&&!n&&(t=t.prototype),t&&!i(t,o)&&r(t,o,{configurable:!0,value:e})}},function(t,e,n){var r=n(1),i=n(12),o=n(175);t.exports=Object.setPrototypeOf||("__proto__"in{}?function(){var t,e=!1,n={};try{(t=r(Object.getOwnPropertyDescriptor(Object.prototype,"__proto__").set))(n,[]),e=n instanceof Array}catch(t){}return function(n,r){return i(n),o(r),e?t(n,r):n.__proto__=r,n}}():void 0)},function(t,e,n){n(3)({target:"Array",stat:!0},{isArray:n(46)})},function(t,e,n){"use strict";var r=n(3),i=n(17).map;r({target:"Array",proto:!0,forced:!n(81)("map")},{map:function(t){return i(this,t,arguments.length>1?arguments[1]:void 0)}})},function(t,e,n){var r=n(1),i=n(2),o=n(34),s=Object,a=r("".split);t.exports=i((function(){return!s("z").propertyIsEnumerable(0)}))?function(t){return"String"==o(t)?a(t,""):s(t)}:s},function(t,e,n){var r=n(2);t.exports=!r((function(){var t=function(){}.bind();return"function"!=typeof t||t.hasOwnProperty("prototype")}))},function(t,e,n){var r=n(6),i=n(30),o=n(11).f,s=r("unscopables"),a=Array.prototype;null==a[s]&&o(a,s,{configurable:!0,value:i(null)}),t.exports=function(t){a[s][t]=!0}},function(t,e,n){var r,i,o=n(4),s=n(66),a=o.process,u=o.Deno,c=a&&a.versions||u&&u.version,f=c&&c.v8;f&&(i=(r=f.split("."))[0]>0&&r[0]<4?1:+(r[0]+r[1])),!i&&s&&(!(r=s.match(/Edge\/(\d+)/))||r[1]>=74)&&(r=s.match(/Chrome\/(\d+)/))&&(i=+r[1]),t.exports=i},function(t,e,n){var r=n(27);t.exports=r("navigator","userAgent")||""},function(t,e,n){var r=n(38),i=n(52),o=r("keys");t.exports=function(t){return o[t]||(o[t]=i(t))}},function(t,e,n){var r=n(8),i=n(10),o=Function.prototype,s=r&&Object.getOwnPropertyDescriptor,a=i(o,"name"),u=a&&"something"===function(){}.name,c=a&&(!r||r&&s(o,"name").configurable);t.exports={EXISTS:a,PROPER:u,CONFIGURABLE:c}},function(t,e,n){var r=n(10),i=n(7),o=n(16),s=n(67),a=n(174),u=s("IE_PROTO"),c=Object,f=c.prototype;t.exports=a?c.getPrototypeOf:function(t){var e=o(t);if(r(e,u))return e[u];var n=e.constructor;return i(n)&&e instanceof n?n.prototype:e instanceof c?f:null}},function(t,e,n){var r=n(31),i=n(14),o=n(71),s=Array,a=Math.max;t.exports=function(t,e,n){for(var u=i(t),c=r(e,u),f=r(void 0===n?u:n,u),l=s(a(f-c,0)),h=0;c<f;c++,h++)o(l,h,t[c]);return l.length=h,l}},function(t,e,n){"use strict";var r=n(53),i=n(11),o=n(43);t.exports=function(t,e,n){var s=r(e);s in t?i.f(t,s,o(0,n)):t[s]=n}},function(t,e,n){var r=n(32),i=n(41),o=n(26),s=n(57),a=n(6)("iterator");t.exports=function(t){if(!o(t))return i(t,a)||i(t,"@@iterator")||s[r(t)]}},function(t,e,n){var r=n(28),i=TypeError;t.exports=function(t,e){if(r(e,t))return t;throw i("Incorrect invocation")}},function(t,e,n){"use strict";var r=n(129).charAt,i=n(15),o=n(22),s=n(91),a=n(94),u=o.set,c=o.getterFor("String Iterator");s(String,"String",(function(t){u(this,{type:"String Iterator",string:i(t),index:0})}),(function(){var t,e=c(this),n=e.string,i=e.index;return i>=n.length?a(void 0,!0):(t=r(n,i),e.index+=t.length,a(t,!1))}))},function(t,e,n){var r=n(1),i=n(2),o=n(7),s=n(32),a=n(27),u=n(119),c=function(){},f=[],l=a("Reflect","construct"),h=/^\s*(?:class|function)\b/,d=r(h.exec),p=!h.exec(c),y=function(t){if(!o(t))return!1;try{return l(c,f,t),!0}catch(t){return!1}},v=function(t){if(!o(t))return!1;switch(s(t)){case"AsyncFunction":case"GeneratorFunction":case"AsyncGeneratorFunction":return!1}try{return p||!!d(h,u(t))}catch(t){return!0}};v.sham=!0,t.exports=!l||i((function(){var t;return y(y.call)||!y(Object)||!y((function(){t=!0}))||t}))?v:y},function(t,e,n){"use strict";var r,i,o=n(9),s=n(1),a=n(15),u=n(134),c=n(102),f=n(38),l=n(30),h=n(22).get,d=n(135),p=n(136),y=f("native-string-replace",String.prototype.replace),v=RegExp.prototype.exec,g=v,m=s("".charAt),x=s("".indexOf),b=s("".replace),w=s("".slice),S=(i=/b*/g,o(v,r=/a/,"a"),o(v,i,"a"),0!==r.lastIndex||0!==i.lastIndex),A=c.BROKEN_CARET,E=void 0!==/()??/.exec("")[1];(S||E||A||d||p)&&(g=function(t){var e,n,r,i,s,c,f,d=this,p=h(d),O=a(t),k=p.raw;if(k)return k.lastIndex=d.lastIndex,e=o(g,k,O),d.lastIndex=k.lastIndex,e;var T=p.groups,I=A&&d.sticky,M=o(u,d),_=d.source,P=0,j=O;if(I&&(M=b(M,"y",""),-1===x(M,"g")&&(M+="g"),j=w(O,d.lastIndex),d.lastIndex>0&&(!d.multiline||d.multiline&&"\n"!==m(O,d.lastIndex-1))&&(_="(?: "+_+")",j=" "+j,P++),n=new RegExp("^(?:"+_+")",M)),E&&(n=new RegExp("^"+_+"$(?!\\s)",M)),S&&(r=d.lastIndex),i=o(v,I?n:d,j),I?i?(i.input=w(i.input,P),i[0]=w(i[0],P),i.index=d.lastIndex,d.lastIndex+=i[0].length):d.lastIndex=0:S&&i&&(d.lastIndex=d.global?i.index+i[0].length:r),E&&i&&i.length>1&&o(y,i[0],n,(function(){for(s=1;s<arguments.length-2;s++)void 0===arguments[s]&&(i[s]=void 0)})),i&&T)for(i.groups=c=l(null),s=0;s<T.length;s++)c[(f=T[s])[0]]=i[f[1]];return i}),t.exports=g},function(t,e,n){"use strict";var r=n(49),i=n(9),o=n(1),s=n(103),a=n(2),u=n(12),c=n(7),f=n(26),l=n(20),h=n(21),d=n(15),p=n(25),y=n(104),v=n(41),g=n(183),m=n(105),x=n(6)("replace"),b=Math.max,w=Math.min,S=o([].concat),A=o([].push),E=o("".indexOf),O=o("".slice),k="$0"==="a".replace(/./,"$0"),T=!!/./[x]&&""===/./[x]("a","$0");s("replace",(function(t,e,n){var o=T?"$":"$0";return[function(t,n){var r=p(this),o=f(t)?void 0:v(t,x);return o?i(o,t,r,n):i(e,d(r),t,n)},function(t,i){var s=u(this),a=d(t);if("string"==typeof i&&-1===E(i,o)&&-1===E(i,"$<")){var f=n(e,s,a,i);if(f.done)return f.value}var p=c(i);p||(i=d(i));var v=s.global;if(v){var x=s.unicode;s.lastIndex=0}for(var k=[];;){var T=m(s,a);if(null===T)break;if(A(k,T),!v)break;""===d(T[0])&&(s.lastIndex=y(a,h(s.lastIndex),x))}for(var I,M="",_=0,P=0;P<k.length;P++){for(var j=d((T=k[P])[0]),N=b(w(l(T.index),a.length),0),R=[],V=1;V<T.length;V++)A(R,void 0===(I=T[V])?I:String(I));var C=T.groups;if(p){var D=S([j],R,N,a);void 0!==C&&A(D,C);var z=d(r(i,void 0,D))}else z=g(j,a,N,R,C,i);N>=_&&(M+=O(a,_,N)+z,_=N+j.length)}return M+O(a,_)}]}),!!a((function(){var t=/./;return t.exec=function(){var t=[];return t.groups={a:"7"},t},"7"!=="".replace(t,"$<a>")}))||!k||T)},function(t,e,n){"use strict";var r=n(3),i=n(1),o=n(62),s=n(19),a=n(47),u=i([].join),c=o!=Object,f=a("join",",");r({target:"Array",proto:!0,forced:c||!f},{join:function(t){return u(s(this),void 0===t?",":t)}})},function(t,e,n){"use strict";var r=n(3),i=n(55).includes,o=n(2),s=n(64);r({target:"Array",proto:!0,forced:o((function(){return!Array(1).includes()}))},{includes:function(t){return i(this,t,arguments.length>1?arguments[1]:void 0)}}),s("includes")},function(t,e,n){"use strict";var r=n(3),i=n(1),o=n(137),s=n(25),a=n(15),u=n(138),c=i("".indexOf);r({target:"String",proto:!0,forced:!u("includes")},{includes:function(t){return!!~c(a(s(this)),a(o(t)),arguments.length>1?arguments[1]:void 0)}})},function(t,e,n){var r=n(2),i=n(6),o=n(65),s=i("species");t.exports=function(t){return o>=51||!r((function(){var e=[];return(e.constructor={})[s]=function(){return{foo:1}},1!==e[t](Boolean).foo}))}},function(t,e,n){var r=n(1);t.exports=r([].slice)},function(t,e,n){"use strict";var r=n(3),i=n(17).filter;r({target:"Array",proto:!0,forced:!n(81)("filter")},{filter:function(t){return i(this,t,arguments.length>1?arguments[1]:void 0)}})},function(t,e,n){"use strict";var r=n(3),i=n(17).find,o=n(64),s=!0;"find"in[]&&Array(1).find((function(){s=!1})),r({target:"Array",proto:!0,forced:s},{find:function(t){return i(this,t,arguments.length>1?arguments[1]:void 0)}}),o("find")},function(t,e,n){var r=n(5),i=n(107),o=r.aTypedArrayConstructor,s=r.getTypedArrayConstructor;t.exports=function(t){return o(i(t,s(t)))}},function(t,e,n){var r=n(4),i=n(87),o=r["__core-js_shared__"]||i("__core-js_shared__",{});t.exports=o},function(t,e,n){var r=n(4),i=Object.defineProperty;t.exports=function(t,e){try{i(r,t,{value:e,configurable:!0,writable:!0})}catch(n){r[t]=e}return e}},function(t,e,n){var r=n(4),i=n(13),o=r.document,s=i(o)&&i(o.createElement);t.exports=function(t){return s?o.createElement(t):{}}},function(t,e,n){var r=n(117),i=n(90);t.exports=Object.keys||function(t){return r(t,i)}},function(t,e){t.exports=["constructor","hasOwnProperty","isPrototypeOf","propertyIsEnumerable","toLocaleString","toString","valueOf"]},function(t,e,n){"use strict";var r=n(3),i=n(9),o=n(39),s=n(68),a=n(7),u=n(173),c=n(69),f=n(59),l=n(58),h=n(23),d=n(18),p=n(6),y=n(57),v=n(121),g=s.PROPER,m=s.CONFIGURABLE,x=v.IteratorPrototype,b=v.BUGGY_SAFARI_ITERATORS,w=p("iterator"),S=function(){return this};t.exports=function(t,e,n,s,p,v,A){u(n,e,s);var E,O,k,T=function(t){if(t===p&&j)return j;if(!b&&t in _)return _[t];switch(t){case"keys":case"values":case"entries":return function(){return new n(this,t)}}return function(){return new n(this)}},I=e+" Iterator",M=!1,_=t.prototype,P=_[w]||_["@@iterator"]||p&&_[p],j=!b&&P||T(p),N="Array"==e&&_.entries||P;if(N&&(E=c(N.call(new t)))!==Object.prototype&&E.next&&(o||c(E)===x||(f?f(E,x):a(E[w])||d(E,w,S)),l(E,I,!0,!0),o&&(y[I]=S)),g&&"values"==p&&P&&"values"!==P.name&&(!o&&m?h(_,"name","values"):(M=!0,j=function(){return i(P,this)})),p)if(O={values:T("values"),keys:v?j:T("keys"),entries:T("entries")},A)for(k in O)(b||M||!(k in _))&&d(_,k,O[k]);else r({target:e,proto:!0,forced:b||M},O);return o&&!A||_[w]===j||d(_,w,j,{name:p}),y[e]=j,O}},function(t,e){e.f=Object.getOwnPropertySymbols},function(t,e,n){var r=n(2),i=n(7),o=/#|\.prototype\./,s=function(t,e){var n=u[a(t)];return n==f||n!=c&&(i(e)?r(e):!!e)},a=s.normalize=function(t){return String(t).replace(o,".").toLowerCase()},u=s.data={},c=s.NATIVE="N",f=s.POLYFILL="P";t.exports=s},function(t,e){t.exports=function(t,e){return{value:t,done:e}}},function(t,e,n){var r=n(6),i=n(57),o=r("iterator"),s=Array.prototype;t.exports=function(t){return void 0!==t&&(i.Array===t||s[o]===t)}},function(t,e,n){var r=n(9),i=n(42),o=n(12),s=n(35),a=n(72),u=TypeError;t.exports=function(t,e){var n=arguments.length<2?a(t):e;if(i(n))return o(r(n,t));throw u(s(t)+" is not iterable")}},function(t,e,n){var r={};r[n(6)("toStringTag")]="z",t.exports="[object z]"===String(r)},function(t,e,n){var r=n(6)("iterator"),i=!1;try{var o=0,s={next:function(){return{done:!!o++}},return:function(){i=!0}};s[r]=function(){return this},Array.from(s,(function(){throw 2}))}catch(t){}t.exports=function(t,e){if(!e&&!i)return!1;var n=!1;try{var o={};o[r]=function(){return{next:function(){return{done:n=!0}}}},t(o)}catch(t){}return n}},function(t,e,n){var r=n(7),i=n(13),o=n(59);t.exports=function(t,e,n){var s,a;return o&&r(s=e.constructor)&&s!==n&&i(a=s.prototype)&&a!==n.prototype&&o(t,a),t}},function(t,e,n){"use strict";var r=n(27),i=n(11),o=n(6),s=n(8),a=o("species");t.exports=function(t){var e=r(t),n=i.f;s&&e&&!e[a]&&n(e,a,{configurable:!0,get:function(){return this}})}},function(t,e,n){var r=n(182);t.exports=function(t,e){return new(r(t))(0===e?0:e)}},function(t,e,n){var r=n(2),i=n(4).RegExp,o=r((function(){var t=i("a","y");return t.lastIndex=2,null!=t.exec("abcd")})),s=o||r((function(){return!i("a","y").sticky})),a=o||r((function(){var t=i("^r","gy");return t.lastIndex=2,null!=t.exec("str")}));t.exports={BROKEN_CARET:a,MISSED_STICKY:s,UNSUPPORTED_Y:o}},function(t,e,n){"use strict";n(48);var r=n(1),i=n(18),o=n(76),s=n(2),a=n(6),u=n(23),c=a("species"),f=RegExp.prototype;t.exports=function(t,e,n,l){var h=a(t),d=!s((function(){var e={};return e[h]=function(){return 7},7!=""[t](e)})),p=d&&!s((function(){var e=!1,n=/a/;return"split"===t&&((n={}).constructor={},n.constructor[c]=function(){return n},n.flags="",n[h]=/./[h]),n.exec=function(){return e=!0,null},n[h](""),!e}));if(!d||!p||n){var y=r(/./[h]),v=e(h,""[t],(function(t,e,n,i,s){var a=r(t),u=e.exec;return u===o||u===f.exec?d&&!s?{done:!0,value:y(e,n,i)}:{done:!0,value:a(n,e,i)}:{done:!1}}));i(String.prototype,t,v[0]),i(f,h,v[1])}l&&u(f[h],"sham",!0)}},function(t,e,n){"use strict";var r=n(129).charAt;t.exports=function(t,e,n){return e+(n?r(t,e).length:1)}},function(t,e,n){var r=n(9),i=n(12),o=n(7),s=n(34),a=n(76),u=TypeError;t.exports=function(t,e){var n=t.exec;if(o(n)){var c=r(n,t,e);return null!==c&&i(c),c}if("RegExp"===s(t))return r(a,t,e);throw u("RegExp#exec called on incompatible receiver")}},function(t,e,n){var r=n(13),i=n(34),o=n(6)("match");t.exports=function(t){var e;return r(t)&&(void 0!==(e=t[o])?!!e:"RegExp"==i(t))}},function(t,e,n){var r=n(12),i=n(141),o=n(26),s=n(6)("species");t.exports=function(t,e){var n,a=r(t).constructor;return void 0===a||o(n=r(a)[s])?e:i(n)}},function(t,e,n){"use strict";var r=n(35),i=TypeError;t.exports=function(t,e){if(!delete t[e])throw i("Cannot delete property "+r(e)+" of "+r(t))}},function(t,e,n){"use strict";var r=n(3),i=n(186),o=n(16),s=n(14),a=n(20),u=n(101);r({target:"Array",proto:!0},{flat:function(){var t=arguments.length?arguments[0]:void 0,e=o(this),n=s(e),r=u(e,0);return r.length=i(r,e,e,n,0,void 0===t?1:a(t)),r}})},function(t,e,n){n(64)("flat")},function(t,e,n){var r=n(3),i=n(238);r({target:"Array",stat:!0,forced:!n(98)((function(t){Array.from(t)}))},{from:i})},function(t,e,n){var r=n(40);t.exports=r&&!Symbol.sham&&"symbol"==typeof Symbol.iterator},function(t,e,n){var r=n(8),i=n(114),o=n(11),s=n(12),a=n(19),u=n(89);e.f=r&&!i?Object.defineProperties:function(t,e){s(t);for(var n,r=a(e),i=u(e),c=i.length,f=0;c>f;)o.f(t,n=i[f++],r[n]);return t}},function(t,e,n){var r=n(8),i=n(2);t.exports=r&&i((function(){return 42!=Object.defineProperty((function(){}),"prototype",{value:42,writable:!1}).prototype}))},function(t,e,n){var r=n(8),i=n(2),o=n(88);t.exports=!r&&!i((function(){return 7!=Object.defineProperty(o("div"),"a",{get:function(){return 7}}).a}))},function(t,e,n){var r=n(9),i=n(13),o=n(54),s=n(41),a=n(167),u=n(6),c=TypeError,f=u("toPrimitive");t.exports=function(t,e){if(!i(t)||o(t))return t;var n,u=s(t,f);if(u){if(void 0===e&&(e="default"),n=r(u,t,e),!i(n)||o(n))return n;throw c("Can't convert object to primitive value")}return void 0===e&&(e="number"),a(t,e)}},function(t,e,n){var r=n(1),i=n(10),o=n(19),s=n(55).indexOf,a=n(56),u=r([].push);t.exports=function(t,e){var n,r=o(t),c=0,f=[];for(n in r)!i(a,n)&&i(r,n)&&u(f,n);for(;e.length>c;)i(r,n=e[c++])&&(~s(f,n)||u(f,n));return f}},function(t,e,n){"use strict";var r={}.propertyIsEnumerable,i=Object.getOwnPropertyDescriptor,o=i&&!r.call({1:2},1);e.f=o?function(t){var e=i(this,t);return!!e&&e.enumerable}:r},function(t,e,n){var r=n(1),i=n(7),o=n(86),s=r(Function.toString);i(o.inspectSource)||(o.inspectSource=function(t){return s(t)}),t.exports=o.inspectSource},function(t,e,n){var r=n(10),i=n(172),o=n(44),s=n(11);t.exports=function(t,e,n){for(var a=i(e),u=s.f,c=o.f,f=0;f<a.length;f++){var l=a[f];r(t,l)||n&&r(n,l)||u(t,l,c(e,l))}}},function(t,e,n){"use strict";var r,i,o,s=n(2),a=n(7),u=n(13),c=n(30),f=n(69),l=n(18),h=n(6),d=n(39),p=h("iterator"),y=!1;[].keys&&("next"in(o=[].keys())?(i=f(f(o)))!==Object.prototype&&(r=i):y=!0),!u(r)||s((function(){var t={};return r[p].call(t)!==t}))?r={}:d&&(r=c(r)),a(r[p])||l(r,p,(function(){return this})),t.exports={IteratorPrototype:r,BUGGY_SAFARI_ITERATORS:y}},function(t,e,n){"use strict";var r=n(3),i=n(4),o=n(1),s=n(93),a=n(18),u=n(123),c=n(125),f=n(73),l=n(7),h=n(26),d=n(13),p=n(2),y=n(98),v=n(58),g=n(99);t.exports=function(t,e,n){var m=-1!==t.indexOf("Map"),x=-1!==t.indexOf("Weak"),b=m?"set":"add",w=i[t],S=w&&w.prototype,A=w,E={},O=function(t){var e=o(S[t]);a(S,t,"add"==t?function(t){return e(this,0===t?0:t),this}:"delete"==t?function(t){return!(x&&!d(t))&&e(this,0===t?0:t)}:"get"==t?function(t){return x&&!d(t)?void 0:e(this,0===t?0:t)}:"has"==t?function(t){return!(x&&!d(t))&&e(this,0===t?0:t)}:function(t,n){return e(this,0===t?0:t,n),this})};if(s(t,!l(w)||!(x||S.forEach&&!p((function(){(new w).entries().next()})))))A=n.getConstructor(e,t,m,b),u.enable();else if(s(t,!0)){var k=new A,T=k[b](x?{}:-0,1)!=k,I=p((function(){k.has(1)})),M=y((function(t){new w(t)})),_=!x&&p((function(){for(var t=new w,e=5;e--;)t[b](e,e);return!t.has(-0)}));M||((A=e((function(t,e){f(t,S);var n=g(new w,t,A);return h(e)||c(e,n[b],{that:n,AS_ENTRIES:m}),n}))).prototype=S,S.constructor=A),(I||_)&&(O("delete"),O("has"),m&&O("get")),(_||T)&&O(b),x&&S.clear&&delete S.clear}return E[t]=A,r({global:!0,constructor:!0,forced:A!=w},E),v(A,t),x||n.setStrong(A,t,m),A}},function(t,e,n){var r=n(3),i=n(1),o=n(56),s=n(13),a=n(10),u=n(11).f,c=n(36),f=n(124),l=n(178),h=n(52),d=n(180),p=!1,y=h("meta"),v=0,g=function(t){u(t,y,{value:{objectID:"O"+v++,weakData:{}}})},m=t.exports={enable:function(){m.enable=function(){},p=!0;var t=c.f,e=i([].splice),n={};n[y]=1,t(n).length&&(c.f=function(n){for(var r=t(n),i=0,o=r.length;i<o;i++)if(r[i]===y){e(r,i,1);break}return r},r({target:"Object",stat:!0,forced:!0},{getOwnPropertyNames:f.f}))},fastKey:function(t,e){if(!s(t))return"symbol"==typeof t?t:("string"==typeof t?"S":"P")+t;if(!a(t,y)){if(!l(t))return"F";if(!e)return"E";g(t)}return t[y].objectID},getWeakData:function(t,e){if(!a(t,y)){if(!l(t))return!0;if(!e)return!1;g(t)}return t[y].weakData},onFreeze:function(t){return d&&p&&l(t)&&!a(t,y)&&g(t),t}};o[y]=!0},function(t,e,n){var r=n(34),i=n(19),o=n(36).f,s=n(70),a="object"==typeof window&&window&&Object.getOwnPropertyNames?Object.getOwnPropertyNames(window):[];t.exports.f=function(t){return a&&"Window"==r(t)?function(t){try{return o(t)}catch(t){return s(a)}}(t):o(i(t))}},function(t,e,n){var r=n(45),i=n(9),o=n(12),s=n(35),a=n(95),u=n(14),c=n(28),f=n(96),l=n(72),h=n(126),d=TypeError,p=function(t,e){this.stopped=t,this.result=e},y=p.prototype;t.exports=function(t,e,n){var v,g,m,x,b,w,S,A=n&&n.that,E=!(!n||!n.AS_ENTRIES),O=!(!n||!n.IS_RECORD),k=!(!n||!n.IS_ITERATOR),T=!(!n||!n.INTERRUPTED),I=r(e,A),M=function(t){return v&&h(v,"normal",t),new p(!0,t)},_=function(t){return E?(o(t),T?I(t[0],t[1],M):I(t[0],t[1])):T?I(t,M):I(t)};if(O)v=t.iterator;else if(k)v=t;else{if(!(g=l(t)))throw d(s(t)+" is not iterable");if(a(g)){for(m=0,x=u(t);x>m;m++)if((b=_(t[m]))&&c(y,b))return b;return new p(!1)}v=f(t,g)}for(w=O?t.next:v.next;!(S=i(w,v)).done;){try{b=_(S.value)}catch(t){h(v,"throw",t)}if("object"==typeof b&&b&&c(y,b))return b}return new p(!1)}},function(t,e,n){var r=n(9),i=n(12),o=n(41);t.exports=function(t,e,n){var s,a;i(t);try{if(!(s=o(t,"return"))){if("throw"===e)throw n;return n}s=r(s,t)}catch(t){a=!0,s=t}if("throw"===e)throw n;if(a)throw s;return i(s),n}},function(t,e,n){"use strict";var r=n(11).f,i=n(30),o=n(128),s=n(45),a=n(73),u=n(26),c=n(125),f=n(91),l=n(94),h=n(100),d=n(8),p=n(123).fastKey,y=n(22),v=y.set,g=y.getterFor;t.exports={getConstructor:function(t,e,n,f){var l=t((function(t,r){a(t,h),v(t,{type:e,index:i(null),first:void 0,last:void 0,size:0}),d||(t.size=0),u(r)||c(r,t[f],{that:t,AS_ENTRIES:n})})),h=l.prototype,y=g(e),m=function(t,e,n){var r,i,o=y(t),s=x(t,e);return s?s.value=n:(o.last=s={index:i=p(e,!0),key:e,value:n,previous:r=o.last,next:void 0,removed:!1},o.first||(o.first=s),r&&(r.next=s),d?o.size++:t.size++,"F"!==i&&(o.index[i]=s)),t},x=function(t,e){var n,r=y(t),i=p(e);if("F"!==i)return r.index[i];for(n=r.first;n;n=n.next)if(n.key==e)return n};return o(h,{clear:function(){for(var t=y(this),e=t.index,n=t.first;n;)n.removed=!0,n.previous&&(n.previous=n.previous.next=void 0),delete e[n.index],n=n.next;t.first=t.last=void 0,d?t.size=0:this.size=0},delete:function(t){var e=y(this),n=x(this,t);if(n){var r=n.next,i=n.previous;delete e.index[n.index],n.removed=!0,i&&(i.next=r),r&&(r.previous=i),e.first==n&&(e.first=r),e.last==n&&(e.last=i),d?e.size--:this.size--}return!!n},forEach:function(t){for(var e,n=y(this),r=s(t,arguments.length>1?arguments[1]:void 0);e=e?e.next:n.first;)for(r(e.value,e.key,this);e&&e.removed;)e=e.previous},has:function(t){return!!x(this,t)}}),o(h,n?{get:function(t){var e=x(this,t);return e&&e.value},set:function(t,e){return m(this,0===t?0:t,e)}}:{add:function(t){return m(this,t=0===t?0:t,t)}}),d&&r(h,"size",{get:function(){return y(this).size}}),l},setStrong:function(t,e,n){var r=e+" Iterator",i=g(e),o=g(r);f(t,e,(function(t,e){v(this,{type:r,target:t,state:i(t),kind:e,last:void 0})}),(function(){for(var t=o(this),e=t.kind,n=t.last;n&&n.removed;)n=n.previous;return t.target&&(t.last=n=n?n.next:t.state.first)?l("keys"==e?n.key:"values"==e?n.value:[n.key,n.value],!1):(t.target=void 0,l(void 0,!0))}),n?"entries":"values",!n,!0),h(e)}}},function(t,e,n){var r=n(18);t.exports=function(t,e,n){for(var i in e)r(t,i,e[i],n);return t}},function(t,e,n){var r=n(1),i=n(20),o=n(15),s=n(25),a=r("".charAt),u=r("".charCodeAt),c=r("".slice),f=function(t){return function(e,n){var r,f,l=o(s(e)),h=i(n),d=l.length;return h<0||h>=d?t?"":void 0:(r=u(l,h))<55296||r>56319||h+1===d||(f=u(l,h+1))<56320||f>57343?t?a(l,h):r:t?c(l,h,h+2):f-56320+(r-55296<<10)+65536}};t.exports={codeAt:f(!1),charAt:f(!0)}},function(t,e,n){var r=n(4),i=n(131),o=n(132),s=n(51),a=n(23),u=n(6),c=u("iterator"),f=u("toStringTag"),l=s.values,h=function(t,e){if(t){if(t[c]!==l)try{a(t,c,l)}catch(e){t[c]=l}if(t[f]||a(t,f,e),i[e])for(var n in s)if(t[n]!==s[n])try{a(t,n,s[n])}catch(e){t[n]=s[n]}}};for(var d in i)h(r[d]&&r[d].prototype,d);h(o,"DOMTokenList")},function(t,e){t.exports={CSSRuleList:0,CSSStyleDeclaration:0,CSSValueList:0,ClientRectList:0,DOMRectList:0,DOMStringList:0,DOMTokenList:1,DataTransferItemList:0,FileList:0,HTMLAllCollection:0,HTMLCollection:0,HTMLFormElement:0,HTMLSelectElement:0,MediaList:0,MimeTypeArray:0,NamedNodeMap:0,NodeList:1,PaintRequestList:0,Plugin:0,PluginArray:0,SVGLengthList:0,SVGNumberList:0,SVGPathSegList:0,SVGPointList:0,SVGStringList:0,SVGTransformList:0,SourceBufferList:0,StyleSheetList:0,TextTrackCueList:0,TextTrackList:0,TouchList:0}},function(t,e,n){var r=n(88)("span").classList,i=r&&r.constructor&&r.constructor.prototype;t.exports=i===Object.prototype?void 0:i},function(t,e,n){"use strict";var r=n(17).forEach,i=n(47)("forEach");t.exports=i?[].forEach:function(t){return r(this,t,arguments.length>1?arguments[1]:void 0)}},function(t,e,n){"use strict";var r=n(12);t.exports=function(){var t=r(this),e="";return t.hasIndices&&(e+="d"),t.global&&(e+="g"),t.ignoreCase&&(e+="i"),t.multiline&&(e+="m"),t.dotAll&&(e+="s"),t.unicode&&(e+="u"),t.unicodeSets&&(e+="v"),t.sticky&&(e+="y"),e}},function(t,e,n){var r=n(2),i=n(4).RegExp;t.exports=r((function(){var t=i(".","s");return!(t.dotAll&&t.exec("\n")&&"s"===t.flags)}))},function(t,e,n){var r=n(2),i=n(4).RegExp;t.exports=r((function(){var t=i("(?<a>b)","g");return"b"!==t.exec("b").groups.a||"bc"!=="b".replace(t,"$<a>c")}))},function(t,e,n){var r=n(106),i=TypeError;t.exports=function(t){if(r(t))throw i("The method doesn't accept regular expressions");return t}},function(t,e,n){var r=n(6)("match");t.exports=function(t){var e=/./;try{"/./"[t](e)}catch(n){try{return e[r]=!1,"/./"[t](e)}catch(t){}}return!1}},function(t,e,n){"use strict";var r=n(49),i=n(19),o=n(20),s=n(14),a=n(47),u=Math.min,c=[].lastIndexOf,f=!!c&&1/[1].lastIndexOf(1,-0)<0,l=a("lastIndexOf"),h=f||!l;t.exports=h?function(t){if(f)return r(c,this,arguments)||0;var e=i(this),n=s(e),a=n-1;for(arguments.length>1&&(a=u(a,o(arguments[1]))),a<0&&(a=n+a);a>=0;a--)if(a in e&&e[a]===t)return a||0;return-1}:c},function(t,e,n){"use strict";var r=n(49),i=n(9),o=n(1),s=n(103),a=n(12),u=n(26),c=n(106),f=n(25),l=n(107),h=n(104),d=n(21),p=n(15),y=n(41),v=n(70),g=n(105),m=n(76),x=n(102),b=n(2),w=x.UNSUPPORTED_Y,S=Math.min,A=[].push,E=o(/./.exec),O=o(A),k=o("".slice);s("split",(function(t,e,n){var o;return o="c"=="abbc".split(/(b)*/)[1]||4!="test".split(/(?:)/,-1).length||2!="ab".split(/(?:ab)*/).length||4!=".".split(/(.?)(.?)/).length||".".split(/()()/).length>1||"".split(/.?/).length?function(t,n){var o=p(f(this)),s=void 0===n?4294967295:n>>>0;if(0===s)return[];if(void 0===t)return[o];if(!c(t))return i(e,o,t,s);for(var a,u,l,h=[],d=(t.ignoreCase?"i":"")+(t.multiline?"m":"")+(t.unicode?"u":"")+(t.sticky?"y":""),y=0,g=new RegExp(t.source,d+"g");(a=i(m,g,o))&&!((u=g.lastIndex)>y&&(O(h,k(o,y,a.index)),a.length>1&&a.index<o.length&&r(A,h,v(a,1)),l=a[0].length,y=u,h.length>=s));)g.lastIndex===a.index&&g.lastIndex++;return y===o.length?!l&&E(g,"")||O(h,""):O(h,k(o,y)),h.length>s?v(h,0,s):h}:"0".split(void 0,0).length?function(t,n){return void 0===t&&0===n?[]:i(e,this,t,n)}:e,[function(e,n){var r=f(this),s=u(e)?void 0:y(e,t);return s?i(s,e,r,n):i(o,p(r),e,n)},function(t,r){var i=a(this),s=p(t),u=n(o,i,s,r,o!==e);if(u.done)return u.value;var c=l(i,RegExp),f=i.unicode,y=(i.ignoreCase?"i":"")+(i.multiline?"m":"")+(i.unicode?"u":"")+(w?"g":"y"),v=new c(w?"^(?:"+i.source+")":i,y),m=void 0===r?4294967295:r>>>0;if(0===m)return[];if(0===s.length)return null===g(v,s)?[s]:[];for(var x=0,b=0,A=[];b<s.length;){v.lastIndex=w?0:b;var E,T=g(v,w?k(s,b):s);if(null===T||(E=S(d(v.lastIndex+(w?b:0)),s.length))===x)b=h(s,b,f);else{if(O(A,k(s,x,b)),A.length===m)return A;for(var I=1;I<=T.length-1;I++)if(O(A,T[I]),A.length===m)return A;b=x=E}}return O(A,k(s,x)),A}]}),!!b((function(){var t=/(?:)/,e=t.exec;t.exec=function(){return e.apply(this,arguments)};var n="ab".split(t);return 2!==n.length||"a"!==n[0]||"b"!==n[1]})),w)},function(t,e,n){var r=n(75),i=n(35),o=TypeError;t.exports=function(t){if(r(t))return t;throw o(i(t)+" is not a constructor")}},function(t,e,n){"use strict";var r=n(9),i=n(103),o=n(12),s=n(26),a=n(21),u=n(15),c=n(25),f=n(41),l=n(104),h=n(105);i("match",(function(t,e,n){return[function(e){var n=c(this),i=s(e)?void 0:f(e,t);return i?r(i,e,n):new RegExp(e)[t](u(n))},function(t){var r=o(this),i=u(t),s=n(e,r,i);if(s.done)return s.value;if(!r.global)return h(r,i);var c=r.unicode;r.lastIndex=0;for(var f,d=[],p=0;null!==(f=h(r,i));){var y=u(f[0]);d[p]=y,""===y&&(r.lastIndex=l(i,a(r.lastIndex),c)),p++}return 0===p?null:d}]}))},function(t,e,n){"use strict";var r=n(3),i=n(46),o=n(75),s=n(13),a=n(31),u=n(14),c=n(19),f=n(71),l=n(6),h=n(81),d=n(82),p=h("slice"),y=l("species"),v=Array,g=Math.max;r({target:"Array",proto:!0,forced:!p},{slice:function(t,e){var n,r,l,h=c(this),p=u(h),m=a(t,p),x=a(void 0===e?p:e,p);if(i(h)&&(n=h.constructor,(o(n)&&(n===v||i(n.prototype))||s(n)&&null===(n=n[y]))&&(n=void 0),n===v||void 0===n))return d(h,m,x);for(r=new(void 0===n?v:n)(g(x-m,0)),l=0;m<x;m++,l++)m in h&&f(r,l,h[m]);return r.length=l,r}})},function(t,e,n){"use strict";var r=n(3),i=n(16),o=n(31),s=n(20),a=n(14),u=n(185),c=n(145),f=n(101),l=n(71),h=n(108),d=n(81)("splice"),p=Math.max,y=Math.min;r({target:"Array",proto:!0,forced:!d},{splice:function(t,e){var n,r,d,v,g,m,x=i(this),b=a(x),w=o(t,b),S=arguments.length;for(0===S?n=r=0:1===S?(n=0,r=b-w):(n=S-2,r=y(p(s(e),0),b-w)),c(b+n-r),d=f(x,r),v=0;v<r;v++)(g=w+v)in x&&l(d,v,x[g]);if(d.length=r,n<r){for(v=w;v<b-r;v++)m=v+n,(g=v+r)in x?x[m]=x[g]:h(x,m);for(v=b;v>b-r+n;v--)h(x,v-1)}else if(n>r)for(v=b-r;v>w;v--)m=v+n-1,(g=v+r-1)in x?x[m]=x[g]:h(x,m);for(v=0;v<n;v++)x[v+w]=arguments[v+2];return u(x,b-r+n),d}})},function(t,e){var n=TypeError;t.exports=function(t){if(t>9007199254740991)throw n("Maximum allowed index exceeded");return t}},function(t,e,n){var r=n(8),i=n(4),o=n(1),s=n(93),a=n(99),u=n(23),c=n(36).f,f=n(28),l=n(106),h=n(15),d=n(147),p=n(102),y=n(187),v=n(18),g=n(2),m=n(10),x=n(22).enforce,b=n(100),w=n(6),S=n(135),A=n(136),E=w("match"),O=i.RegExp,k=O.prototype,T=i.SyntaxError,I=o(k.exec),M=o("".charAt),_=o("".replace),P=o("".indexOf),j=o("".slice),N=/^\?<[^\s\d!#%&*+<=>@^][^\s!#%&*+<=>@^]*>/,R=/a/g,V=/a/g,C=new O(R)!==R,D=p.MISSED_STICKY,z=p.UNSUPPORTED_Y,F=r&&(!C||D||S||A||g((function(){return V[E]=!1,O(R)!=R||O(V)==V||"/a/i"!=O(R,"i")})));if(s("RegExp",F)){for(var B=function(t,e){var n,r,i,o,s,c,p=f(k,this),y=l(t),v=void 0===e,g=[],b=t;if(!p&&y&&v&&t.constructor===B)return t;if((y||f(k,t))&&(t=t.source,v&&(e=d(b))),t=void 0===t?"":h(t),e=void 0===e?"":h(e),b=t,S&&"dotAll"in R&&(r=!!e&&P(e,"s")>-1)&&(e=_(e,/s/g,"")),n=e,D&&"sticky"in R&&(i=!!e&&P(e,"y")>-1)&&z&&(e=_(e,/y/g,"")),A&&(t=(o=function(t){for(var e,n=t.length,r=0,i="",o=[],s={},a=!1,u=!1,c=0,f="";r<=n;r++){if("\\"===(e=M(t,r)))e+=M(t,++r);else if("]"===e)a=!1;else if(!a)switch(!0){case"["===e:a=!0;break;case"("===e:I(N,j(t,r+1))&&(r+=2,u=!0),i+=e,c++;continue;case">"===e&&u:if(""===f||m(s,f))throw new T("Invalid capture group name");s[f]=!0,o[o.length]=[f,c],u=!1,f="";continue}u?f+=e:i+=e}return[i,o]}(t))[0],g=o[1]),s=a(O(t,e),p?this:k,B),(r||i||g.length)&&(c=x(s),r&&(c.dotAll=!0,c.raw=B(function(t){for(var e,n=t.length,r=0,i="",o=!1;r<=n;r++)"\\"!==(e=M(t,r))?o||"."!==e?("["===e?o=!0:"]"===e&&(o=!1),i+=e):i+="[\\s\\S]":i+=e+M(t,++r);return i}(t),n)),i&&(c.sticky=!0),g.length&&(c.groups=g)),t!==b)try{u(s,"source",""===b?"(?:)":b)}catch(t){}return s},L=c(O),G=0;L.length>G;)y(B,O,L[G++]);k.constructor=B,B.prototype=k,v(i,"RegExp",B,{constructor:!0})}b("RegExp")},function(t,e,n){var r=n(9),i=n(10),o=n(28),s=n(134),a=RegExp.prototype;t.exports=function(t){var e=t.flags;return void 0!==e||"flags"in a||i(t,"flags")||!o(a,t)?e:r(s,t)}},function(t,e,n){"use strict";var r=n(68).PROPER,i=n(18),o=n(12),s=n(15),a=n(2),u=n(147),c=RegExp.prototype.toString,f=a((function(){return"/a/b"!=c.call({source:"a",flags:"b"})})),l=r&&"toString"!=c.name;(f||l)&&i(RegExp.prototype,"toString",(function(){var t=o(this);return"/"+s(t.source)+"/"+s(u(t))}),{unsafe:!0})},function(t,e){t.exports="\t\n\v\f\r                　\u2028\u2029\ufeff"},function(t,e,n){var r=n(6);e.f=r},function(t,e,n){var r=n(40);t.exports=r&&!!Symbol.for&&!!Symbol.keyFor},function(t,e,n){"use strict";var r=n(4),i=n(1),o=n(8),s=n(153),a=n(68),u=n(23),c=n(128),f=n(2),l=n(73),h=n(20),d=n(21),p=n(154),y=n(204),v=n(69),g=n(59),m=n(36).f,x=n(11).f,b=n(155),w=n(70),S=n(58),A=n(22),E=a.PROPER,O=a.CONFIGURABLE,k=A.get,T=A.set,I=r.ArrayBuffer,M=I,_=M&&M.prototype,P=r.DataView,j=P&&P.prototype,N=Object.prototype,R=r.Array,V=r.RangeError,C=i(b),D=i([].reverse),z=y.pack,F=y.unpack,B=function(t){return[255&t]},L=function(t){return[255&t,t>>8&255]},G=function(t){return[255&t,t>>8&255,t>>16&255,t>>24&255]},U=function(t){return t[3]<<24|t[2]<<16|t[1]<<8|t[0]},H=function(t){return z(t,23,4)},W=function(t){return z(t,52,8)},Y=function(t,e){x(t.prototype,e,{get:function(){return k(this)[e]}})},$=function(t,e,n,r){var i=p(n),o=k(t);if(i+e>o.byteLength)throw V("Wrong index");var s=k(o.buffer).bytes,a=i+o.byteOffset,u=w(s,a,a+e);return r?u:D(u)},q=function(t,e,n,r,i,o){var s=p(n),a=k(t);if(s+e>a.byteLength)throw V("Wrong index");for(var u=k(a.buffer).bytes,c=s+a.byteOffset,f=r(+i),l=0;l<e;l++)u[c+l]=f[o?l:e-l-1]};if(s){var K=E&&"ArrayBuffer"!==I.name;if(f((function(){I(1)}))&&f((function(){new I(-1)}))&&!f((function(){return new I,new I(1.5),new I(NaN),1!=I.length||K&&!O})))K&&O&&u(I,"name","ArrayBuffer");else{(M=function(t){return l(this,_),new I(p(t))}).prototype=_;for(var X,J=m(I),Z=0;J.length>Z;)(X=J[Z++])in M||u(M,X,I[X]);_.constructor=M}g&&v(j)!==N&&g(j,N);var Q=new P(new M(2)),tt=i(j.setInt8);Q.setInt8(0,2147483648),Q.setInt8(1,2147483649),!Q.getInt8(0)&&Q.getInt8(1)||c(j,{setInt8:function(t,e){tt(this,t,e<<24>>24)},setUint8:function(t,e){tt(this,t,e<<24>>24)}},{unsafe:!0})}else _=(M=function(t){l(this,_);var e=p(t);T(this,{bytes:C(R(e),0),byteLength:e}),o||(this.byteLength=e)}).prototype,j=(P=function(t,e,n){l(this,j),l(t,_);var r=k(t).byteLength,i=h(e);if(i<0||i>r)throw V("Wrong offset");if(i+(n=void 0===n?r-i:d(n))>r)throw V("Wrong length");T(this,{buffer:t,byteLength:n,byteOffset:i}),o||(this.buffer=t,this.byteLength=n,this.byteOffset=i)}).prototype,o&&(Y(M,"byteLength"),Y(P,"buffer"),Y(P,"byteLength"),Y(P,"byteOffset")),c(j,{getInt8:function(t){return $(this,1,t)[0]<<24>>24},getUint8:function(t){return $(this,1,t)[0]},getInt16:function(t){var e=$(this,2,t,arguments.length>1?arguments[1]:void 0);return(e[1]<<8|e[0])<<16>>16},getUint16:function(t){var e=$(this,2,t,arguments.length>1?arguments[1]:void 0);return e[1]<<8|e[0]},getInt32:function(t){return U($(this,4,t,arguments.length>1?arguments[1]:void 0))},getUint32:function(t){return U($(this,4,t,arguments.length>1?arguments[1]:void 0))>>>0},getFloat32:function(t){return F($(this,4,t,arguments.length>1?arguments[1]:void 0),23)},getFloat64:function(t){return F($(this,8,t,arguments.length>1?arguments[1]:void 0),52)},setInt8:function(t,e){q(this,1,t,B,e)},setUint8:function(t,e){q(this,1,t,B,e)},setInt16:function(t,e){q(this,2,t,L,e,arguments.length>2?arguments[2]:void 0)},setUint16:function(t,e){q(this,2,t,L,e,arguments.length>2?arguments[2]:void 0)},setInt32:function(t,e){q(this,4,t,G,e,arguments.length>2?arguments[2]:void 0)},setUint32:function(t,e){q(this,4,t,G,e,arguments.length>2?arguments[2]:void 0)},setFloat32:function(t,e){q(this,4,t,H,e,arguments.length>2?arguments[2]:void 0)},setFloat64:function(t,e){q(this,8,t,W,e,arguments.length>2?arguments[2]:void 0)}});S(M,"ArrayBuffer"),S(P,"DataView"),t.exports={ArrayBuffer:M,DataView:P}},function(t,e){t.exports="undefined"!=typeof ArrayBuffer&&"undefined"!=typeof DataView},function(t,e,n){var r=n(20),i=n(21),o=RangeError;t.exports=function(t){if(void 0===t)return 0;var e=r(t),n=i(e);if(e!==n)throw o("Wrong length or index");return n}},function(t,e,n){"use strict";var r=n(16),i=n(31),o=n(14);t.exports=function(t){for(var e=r(this),n=o(e),s=arguments.length,a=i(s>1?arguments[1]:void 0,n),u=s>2?arguments[2]:void 0,c=void 0===u?n:i(u,n);c>a;)e[a++]=t;return e}},function(t,e,n){var r=n(209),i=RangeError;t.exports=function(t,e){var n=r(t);if(n%e)throw i("Wrong offset");return n}},function(t,e,n){var r=n(116),i=TypeError;t.exports=function(t){var e=r(t,"number");if("number"==typeof e)throw i("Can't convert number to bigint");return BigInt(e)}},function(t,e,n){var r=n(42),i=n(16),o=n(62),s=n(14),a=TypeError,u=function(t){return function(e,n,u,c){r(n);var f=i(e),l=o(f),h=s(f),d=t?h-1:0,p=t?-1:1;if(u<2)for(;;){if(d in l){c=l[d],d+=p;break}if(d+=p,t?d<0:h<=d)throw a("Reduce of empty array with no initial value")}for(;t?d>=0:h>d;d+=p)d in l&&(c=n(c,l[d],d,f));return c}};t.exports={left:u(!1),right:u(!0)}},function(t,e,n){var r=n(70),i=Math.floor,o=function(t,e){var n=t.length,u=i(n/2);return n<8?s(t,e):a(t,o(r(t,0,u),e),o(r(t,u),e),e)},s=function(t,e){for(var n,r,i=t.length,o=1;o<i;){for(r=o,n=t[o];r&&e(t[r-1],n)>0;)t[r]=t[--r];r!==o++&&(t[r]=n)}return t},a=function(t,e,n,r){for(var i=e.length,o=n.length,s=0,a=0;s<i||a<o;)t[s+a]=s<i&&a<o?r(e[s],n[a])<=0?e[s++]:n[a++]:s<i?e[s++]:n[a++];return t};t.exports=o},function(t,e,n){var r=n(66).match(/firefox\/(\d+)/i);t.exports=!!r&&+r[1]},function(t,e,n){var r=n(66);t.exports=/MSIE|Trident/.test(r)},function(t,e,n){var r=n(66).match(/AppleWebKit\/(\d+)\./);t.exports=!!r&&+r[1]},function(t,e,n){"use strict";var r=n(3),i=n(17).every;r({target:"Array",proto:!0,forced:!n(47)("every")},{every:function(t){return i(this,t,arguments.length>1?arguments[1]:void 0)}})},function(t,e,n){"use strict";var r=n(3),i=n(1),o=n(42),s=n(16),a=n(14),u=n(108),c=n(15),f=n(2),l=n(159),h=n(47),d=n(160),p=n(161),y=n(65),v=n(162),g=[],m=i(g.sort),x=i(g.push),b=f((function(){g.sort(void 0)})),w=f((function(){g.sort(null)})),S=h("sort"),A=!f((function(){if(y)return y<70;if(!(d&&d>3)){if(p)return!0;if(v)return v<603;var t,e,n,r,i="";for(t=65;t<76;t++){switch(e=String.fromCharCode(t),t){case 66:case 69:case 70:case 72:n=3;break;case 68:case 71:n=4;break;default:n=2}for(r=0;r<47;r++)g.push({k:e+r,v:n})}for(g.sort((function(t,e){return e.v-t.v})),r=0;r<g.length;r++)e=g[r].k.charAt(0),i.charAt(i.length-1)!==e&&(i+=e);return"DGBEFHACIJK"!==i}}));r({target:"Array",proto:!0,forced:b||!w||!S||!A},{sort:function(t){void 0!==t&&o(t);var e=s(this);if(A)return void 0===t?m(e):m(e,t);var n,r,i=[],f=a(e);for(r=0;r<f;r++)r in e&&x(i,e[r]);for(l(i,function(t){return function(e,n){return void 0===n?-1:void 0===e?1:void 0!==t?+t(e,n)||0:c(e)>c(n)?1:-1}}(t)),n=a(i),r=0;r<n;)e[r]=i[r++];for(;r<f;)u(e,r++);return e}})},function(t,n){t.exports=e},function(t,e){var n;n=function(){return this}();try{n=n||new Function("return this")()}catch(t){"object"==typeof window&&(n=window)}t.exports=n},function(t,e,n){var r=n(9),i=n(7),o=n(13),s=TypeError;t.exports=function(t,e){var n,a;if("string"===e&&i(n=t.toString)&&!o(a=r(n,t)))return a;if(i(n=t.valueOf)&&!o(a=r(n,t)))return a;if("string"!==e&&i(n=t.toString)&&!o(a=r(n,t)))return a;throw s("Can't convert object to primitive value")}},function(t,e){var n=Math.ceil,r=Math.floor;t.exports=Math.trunc||function(t){var e=+t;return(e>0?r:n)(e)}},function(t,e,n){var r=n(27);t.exports=r("document","documentElement")},function(t,e,n){var r=n(4),i=n(7),o=r.WeakMap;t.exports=i(o)&&/native code/.test(String(o))},function(t,e,n){var r=n(2),i=n(7),o=n(10),s=n(8),a=n(68).CONFIGURABLE,u=n(119),c=n(22),f=c.enforce,l=c.get,h=Object.defineProperty,d=s&&!r((function(){return 8!==h((function(){}),"length",{value:8}).length})),p=String(String).split("String"),y=t.exports=function(t,e,n){"Symbol("===String(e).slice(0,7)&&(e="["+String(e).replace(/^Symbol\(([^)]*)\)/,"$1")+"]"),n&&n.getter&&(e="get "+e),n&&n.setter&&(e="set "+e),(!o(t,"name")||a&&t.name!==e)&&(s?h(t,"name",{value:e,configurable:!0}):t.name=e),d&&n&&o(n,"arity")&&t.length!==n.arity&&h(t,"length",{value:n.arity});try{n&&o(n,"constructor")&&n.constructor?s&&h(t,"prototype",{writable:!1}):t.prototype&&(t.prototype=void 0)}catch(t){}var r=f(t);return o(r,"source")||(r.source=p.join("string"==typeof e?e:"")),t};Function.prototype.toString=y((function(){return i(this)&&l(this).source||u(this)}),"toString")},function(t,e,n){var r=n(27),i=n(1),o=n(36),s=n(92),a=n(12),u=i([].concat);t.exports=r("Reflect","ownKeys")||function(t){var e=o.f(a(t)),n=s.f;return n?u(e,n(t)):e}},function(t,e,n){"use strict";var r=n(121).IteratorPrototype,i=n(30),o=n(43),s=n(58),a=n(57),u=function(){return this};t.exports=function(t,e,n,c){var f=e+" Iterator";return t.prototype=i(r,{next:o(+!c,n)}),s(t,f,!1,!0),a[f]=u,t}},function(t,e,n){var r=n(2);t.exports=!r((function(){function t(){}return t.prototype.constructor=null,Object.getPrototypeOf(new t)!==t.prototype}))},function(t,e,n){var r=n(7),i=String,o=TypeError;t.exports=function(t){if("object"==typeof t||r(t))return t;throw o("Can't set "+i(t)+" as a prototype")}},function(t,e,n){n(177)},function(t,e,n){"use strict";n(122)("Map",(function(t){return function(){return t(this,arguments.length?arguments[0]:void 0)}}),n(127))},function(t,e,n){var r=n(2),i=n(13),o=n(34),s=n(179),a=Object.isExtensible,u=r((function(){a(1)}));t.exports=u||s?function(t){return!!i(t)&&((!s||"ArrayBuffer"!=o(t))&&(!a||a(t)))}:a},function(t,e,n){var r=n(2);t.exports=r((function(){if("function"==typeof ArrayBuffer){var t=new ArrayBuffer(8);Object.isExtensible(t)&&Object.defineProperty(t,"a",{value:8})}}))},function(t,e,n){var r=n(2);t.exports=!r((function(){return Object.isExtensible(Object.preventExtensions({}))}))},function(t,e,n){"use strict";var r=n(97),i=n(32);t.exports=r?{}.toString:function(){return"[object "+i(this)+"]"}},function(t,e,n){var r=n(46),i=n(75),o=n(13),s=n(6)("species"),a=Array;t.exports=function(t){var e;return r(t)&&(e=t.constructor,(i(e)&&(e===a||r(e.prototype))||o(e)&&null===(e=e[s]))&&(e=void 0)),void 0===e?a:e}},function(t,e,n){var r=n(1),i=n(16),o=Math.floor,s=r("".charAt),a=r("".replace),u=r("".slice),c=/\$([$&'`]|\d{1,2}|<[^>]*>)/g,f=/\$([$&'`]|\d{1,2})/g;t.exports=function(t,e,n,r,l,h){var d=n+t.length,p=r.length,y=f;return void 0!==l&&(l=i(l),y=c),a(h,y,(function(i,a){var c;switch(s(a,0)){case"$":return"$";case"&":return t;case"`":return u(e,0,n);case"'":return u(e,d);case"<":c=l[u(a,1,-1)];break;default:var f=+a;if(0===f)return i;if(f>p){var h=o(f/10);return 0===h?i:h<=p?void 0===r[h-1]?s(a,1):r[h-1]+s(a,1):i}c=r[f-1]}return void 0===c?"":c}))}},function(t,e,n){var r=n(3),i=n(139);r({target:"Array",proto:!0,forced:i!==[].lastIndexOf},{lastIndexOf:i})},function(t,e,n){"use strict";var r=n(8),i=n(46),o=TypeError,s=Object.getOwnPropertyDescriptor,a=r&&!function(){if(void 0!==this)return!0;try{Object.defineProperty([],"length",{writable:!1}).length=1}catch(t){return t instanceof TypeError}}();t.exports=a?function(t,e){if(i(t)&&!s(t,"length").writable)throw o("Cannot set read only .length");return t.length=e}:function(t,e){return t.length=e}},function(t,e,n){"use strict";var r=n(46),i=n(14),o=n(145),s=n(45),a=function(t,e,n,u,c,f,l,h){for(var d,p,y=c,v=0,g=!!l&&s(l,h);v<u;)v in n&&(d=g?g(n[v],v,e):n[v],f>0&&r(d)?(p=i(d),y=a(t,e,d,p,y,f-1)-1):(o(y+1),t[y]=d),y++),v++;return y};t.exports=a},function(t,e,n){var r=n(11).f;t.exports=function(t,e,n){n in t||r(t,n,{configurable:!0,get:function(){return e[n]},set:function(t){e[n]=t}})}},function(t,e,n){var r=n(3),i=n(189);r({global:!0,forced:parseFloat!=i},{parseFloat:i})},function(t,e,n){var r=n(4),i=n(2),o=n(1),s=n(15),a=n(190).trim,u=n(149),c=o("".charAt),f=r.parseFloat,l=r.Symbol,h=l&&l.iterator,d=1/f(u+"-0")!=-1/0||h&&!i((function(){f(Object(h))}));t.exports=d?function(t){var e=a(s(t)),n=f(e);return 0===n&&"-"==c(e,0)?-0:n}:f},function(t,e,n){var r=n(1),i=n(25),o=n(15),s=n(149),a=r("".replace),u="["+s+"]",c=RegExp("^"+u+u+"*"),f=RegExp(u+u+"*$"),l=function(t){return function(e){var n=o(i(e));return 1&t&&(n=a(n,c,"")),2&t&&(n=a(n,f,"")),n}};t.exports={start:l(1),end:l(2),trim:l(3)}},function(t,e,n){var r=n(3),i=n(2),o=n(19),s=n(44).f,a=n(8),u=i((function(){s(1)}));r({target:"Object",stat:!0,forced:!a||u,sham:!a},{getOwnPropertyDescriptor:function(t,e){return s(o(t),e)}})},function(t,e,n){n(193),n(197),n(198),n(199),n(200)},function(t,e,n){"use strict";var r=n(3),i=n(4),o=n(9),s=n(1),a=n(39),u=n(8),c=n(40),f=n(2),l=n(10),h=n(28),d=n(12),p=n(19),y=n(53),v=n(15),g=n(43),m=n(30),x=n(89),b=n(36),w=n(124),S=n(92),A=n(44),E=n(11),O=n(113),k=n(118),T=n(18),I=n(38),M=n(67),_=n(56),P=n(52),j=n(6),N=n(150),R=n(194),V=n(196),C=n(58),D=n(22),z=n(17).forEach,F=M("hidden"),B=D.set,L=D.getterFor("Symbol"),G=Object.prototype,U=i.Symbol,H=U&&U.prototype,W=i.TypeError,Y=i.QObject,$=A.f,q=E.f,K=w.f,X=k.f,J=s([].push),Z=I("symbols"),Q=I("op-symbols"),tt=I("wks"),et=!Y||!Y.prototype||!Y.prototype.findChild,nt=u&&f((function(){return 7!=m(q({},"a",{get:function(){return q(this,"a",{value:7}).a}})).a}))?function(t,e,n){var r=$(G,e);r&&delete G[e],q(t,e,n),r&&t!==G&&q(G,e,r)}:q,rt=function(t,e){var n=Z[t]=m(H);return B(n,{type:"Symbol",tag:t,description:e}),u||(n.description=e),n},it=function(t,e,n){t===G&&it(Q,e,n),d(t);var r=y(e);return d(n),l(Z,r)?(n.enumerable?(l(t,F)&&t[F][r]&&(t[F][r]=!1),n=m(n,{enumerable:g(0,!1)})):(l(t,F)||q(t,F,g(1,{})),t[F][r]=!0),nt(t,r,n)):q(t,r,n)},ot=function(t,e){d(t);var n=p(e),r=x(n).concat(ct(n));return z(r,(function(e){u&&!o(st,n,e)||it(t,e,n[e])})),t},st=function(t){var e=y(t),n=o(X,this,e);return!(this===G&&l(Z,e)&&!l(Q,e))&&(!(n||!l(this,e)||!l(Z,e)||l(this,F)&&this[F][e])||n)},at=function(t,e){var n=p(t),r=y(e);if(n!==G||!l(Z,r)||l(Q,r)){var i=$(n,r);return!i||!l(Z,r)||l(n,F)&&n[F][r]||(i.enumerable=!0),i}},ut=function(t){var e=K(p(t)),n=[];return z(e,(function(t){l(Z,t)||l(_,t)||J(n,t)})),n},ct=function(t){var e=t===G,n=K(e?Q:p(t)),r=[];return z(n,(function(t){!l(Z,t)||e&&!l(G,t)||J(r,Z[t])})),r};c||(T(H=(U=function(){if(h(H,this))throw W("Symbol is not a constructor");var t=arguments.length&&void 0!==arguments[0]?v(arguments[0]):void 0,e=P(t),n=function(t){this===G&&o(n,Q,t),l(this,F)&&l(this[F],e)&&(this[F][e]=!1),nt(this,e,g(1,t))};return u&&et&&nt(G,e,{configurable:!0,set:n}),rt(e,t)}).prototype,"toString",(function(){return L(this).tag})),T(U,"withoutSetter",(function(t){return rt(P(t),t)})),k.f=st,E.f=it,O.f=ot,A.f=at,b.f=w.f=ut,S.f=ct,N.f=function(t){return rt(j(t),t)},u&&(q(H,"description",{configurable:!0,get:function(){return L(this).description}}),a||T(G,"propertyIsEnumerable",st,{unsafe:!0}))),r({global:!0,constructor:!0,wrap:!0,forced:!c,sham:!c},{Symbol:U}),z(x(tt),(function(t){R(t)})),r({target:"Symbol",stat:!0,forced:!c},{useSetter:function(){et=!0},useSimple:function(){et=!1}}),r({target:"Object",stat:!0,forced:!c,sham:!u},{create:function(t,e){return void 0===e?m(t):ot(m(t),e)},defineProperty:it,defineProperties:ot,getOwnPropertyDescriptor:at}),r({target:"Object",stat:!0,forced:!c},{getOwnPropertyNames:ut}),V(),C(U,"Symbol"),_[F]=!0},function(t,e,n){var r=n(195),i=n(10),o=n(150),s=n(11).f;t.exports=function(t){var e=r.Symbol||(r.Symbol={});i(e,t)||s(e,t,{value:o.f(t)})}},function(t,e,n){var r=n(4);t.exports=r},function(t,e,n){var r=n(9),i=n(27),o=n(6),s=n(18);t.exports=function(){var t=i("Symbol"),e=t&&t.prototype,n=e&&e.valueOf,a=o("toPrimitive");e&&!e[a]&&s(e,a,(function(t){return r(n,this)}),{arity:1})}},function(t,e,n){var r=n(3),i=n(27),o=n(10),s=n(15),a=n(38),u=n(151),c=a("string-to-symbol-registry"),f=a("symbol-to-string-registry");r({target:"Symbol",stat:!0,forced:!u},{for:function(t){var e=s(t);if(o(c,e))return c[e];var n=i("Symbol")(e);return c[e]=n,f[n]=e,n}})},function(t,e,n){var r=n(3),i=n(10),o=n(54),s=n(35),a=n(38),u=n(151),c=a("symbol-to-string-registry");r({target:"Symbol",stat:!0,forced:!u},{keyFor:function(t){if(!o(t))throw TypeError(s(t)+" is not a symbol");if(i(c,t))return c[t]}})},function(t,e,n){var r=n(3),i=n(27),o=n(49),s=n(9),a=n(1),u=n(2),c=n(46),f=n(7),l=n(13),h=n(54),d=n(82),p=n(40),y=i("JSON","stringify"),v=a(/./.exec),g=a("".charAt),m=a("".charCodeAt),x=a("".replace),b=a(1..toString),w=/[\uD800-\uDFFF]/g,S=/^[\uD800-\uDBFF]$/,A=/^[\uDC00-\uDFFF]$/,E=!p||u((function(){var t=i("Symbol")();return"[null]"!=y([t])||"{}"!=y({a:t})||"{}"!=y(Object(t))})),O=u((function(){return'"\\udf06\\ud834"'!==y("\udf06\ud834")||'"\\udead"'!==y("\udead")})),k=function(t,e){var n=d(arguments),r=e;if((l(e)||void 0!==t)&&!h(t))return c(e)||(e=function(t,e){if(f(r)&&(e=s(r,this,t,e)),!h(e))return e}),n[1]=e,o(y,null,n)},T=function(t,e,n){var r=g(n,e-1),i=g(n,e+1);return v(S,t)&&!v(A,i)||v(A,t)&&!v(S,r)?"\\u"+b(m(t,0),16):t};y&&r({target:"JSON",stat:!0,arity:3,forced:E||O},{stringify:function(t,e,n){var r=d(arguments),i=o(E?k:y,null,r);return O&&"string"==typeof i?x(i,w,T):i}})},function(t,e,n){var r=n(3),i=n(40),o=n(2),s=n(92),a=n(16);r({target:"Object",stat:!0,forced:!i||o((function(){s.f(1)}))},{getOwnPropertySymbols:function(t){var e=s.f;return e?e(a(t)):[]}})},function(t,e,n){"use strict";var r=n(3),i=n(8),o=n(4),s=n(1),a=n(10),u=n(7),c=n(28),f=n(15),l=n(11).f,h=n(120),d=o.Symbol,p=d&&d.prototype;if(i&&u(d)&&(!("description"in p)||void 0!==d().description)){var y={},v=function(){var t=arguments.length<1||void 0===arguments[0]?void 0:f(arguments[0]),e=c(p,this)?new d(t):void 0===t?d():d(t);return""===t&&(y[e]=!0),e};h(v,d),v.prototype=p,p.constructor=v;var g="Symbol(test)"==String(d("test")),m=s(p.valueOf),x=s(p.toString),b=/^Symbol\((.*)\)[^)]+$/,w=s("".replace),S=s("".slice);l(p,"description",{configurable:!0,get:function(){var t=m(this);if(a(y,t))return"";var e=x(t),n=g?S(e,7,-1):w(e,b,"$1");return""===n?void 0:n}}),r({global:!0,constructor:!0,forced:!0},{Symbol:v})}},function(t,e,n){n(3)({target:"Object",stat:!0,sham:!n(8)},{create:n(30)})},function(t,e,n){"use strict";var r=n(3),i=n(1),o=n(2),s=n(152),a=n(12),u=n(31),c=n(21),f=n(107),l=s.ArrayBuffer,h=s.DataView,d=h.prototype,p=i(l.prototype.slice),y=i(d.getUint8),v=i(d.setUint8);r({target:"ArrayBuffer",proto:!0,unsafe:!0,forced:o((function(){return!new l(2).slice(1,void 0).byteLength}))},{slice:function(t,e){if(p&&void 0===e)return p(a(this),t);for(var n=a(this).byteLength,r=u(t,n),i=u(void 0===e?n:e,n),o=new(f(this,l))(c(i-r)),s=new h(this),d=new h(o),g=0;r<i;)v(d,g++,y(s,r++));return o}})},function(t,e){var n=Array,r=Math.abs,i=Math.pow,o=Math.floor,s=Math.log,a=Math.LN2;t.exports={pack:function(t,e,u){var c,f,l,h=n(u),d=8*u-e-1,p=(1<<d)-1,y=p>>1,v=23===e?i(2,-24)-i(2,-77):0,g=t<0||0===t&&1/t<0?1:0,m=0;for((t=r(t))!=t||t===1/0?(f=t!=t?1:0,c=p):(c=o(s(t)/a),t*(l=i(2,-c))<1&&(c--,l*=2),(t+=c+y>=1?v/l:v*i(2,1-y))*l>=2&&(c++,l/=2),c+y>=p?(f=0,c=p):c+y>=1?(f=(t*l-1)*i(2,e),c+=y):(f=t*i(2,y-1)*i(2,e),c=0));e>=8;)h[m++]=255&f,f/=256,e-=8;for(c=c<<e|f,d+=e;d>0;)h[m++]=255&c,c/=256,d-=8;return h[--m]|=128*g,h},unpack:function(t,e){var n,r=t.length,o=8*r-e-1,s=(1<<o)-1,a=s>>1,u=o-7,c=r-1,f=t[c--],l=127&f;for(f>>=7;u>0;)l=256*l+t[c--],u-=8;for(n=l&(1<<-u)-1,l>>=-u,u+=e;u>0;)n=256*n+t[c--],u-=8;if(0===l)l=1-a;else{if(l===s)return n?NaN:f?-1/0:1/0;n+=i(2,e),l-=a}return(f?-1:1)*n*i(2,l-e)}}},function(t,e,n){n(206)("Float64",(function(t){return function(e,n,r){return t(this,e,n,r)}}))},function(t,e,n){"use strict";var r=n(3),i=n(4),o=n(9),s=n(8),a=n(207),u=n(5),c=n(152),f=n(73),l=n(43),h=n(23),d=n(208),p=n(21),y=n(154),v=n(156),g=n(53),m=n(10),x=n(32),b=n(13),w=n(54),S=n(30),A=n(28),E=n(59),O=n(36).f,k=n(210),T=n(17).forEach,I=n(100),M=n(11),_=n(44),P=n(22),j=n(99),N=P.get,R=P.set,V=P.enforce,C=M.f,D=_.f,z=Math.round,F=i.RangeError,B=c.ArrayBuffer,L=B.prototype,G=c.DataView,U=u.NATIVE_ARRAY_BUFFER_VIEWS,H=u.TYPED_ARRAY_TAG,W=u.TypedArray,Y=u.TypedArrayPrototype,$=u.aTypedArrayConstructor,q=u.isTypedArray,K=function(t,e){$(t);for(var n=0,r=e.length,i=new t(r);r>n;)i[n]=e[n++];return i},X=function(t,e){C(t,e,{get:function(){return N(this)[e]}})},J=function(t){var e;return A(L,t)||"ArrayBuffer"==(e=x(t))||"SharedArrayBuffer"==e},Z=function(t,e){return q(t)&&!w(e)&&e in t&&d(+e)&&e>=0},Q=function(t,e){return e=g(e),Z(t,e)?l(2,t[e]):D(t,e)},tt=function(t,e,n){return e=g(e),!(Z(t,e)&&b(n)&&m(n,"value"))||m(n,"get")||m(n,"set")||n.configurable||m(n,"writable")&&!n.writable||m(n,"enumerable")&&!n.enumerable?C(t,e,n):(t[e]=n.value,t)};s?(U||(_.f=Q,M.f=tt,X(Y,"buffer"),X(Y,"byteOffset"),X(Y,"byteLength"),X(Y,"length")),r({target:"Object",stat:!0,forced:!U},{getOwnPropertyDescriptor:Q,defineProperty:tt}),t.exports=function(t,e,n){var s=t.match(/\d+$/)[0]/8,u=t+(n?"Clamped":"")+"Array",c="get"+t,l="set"+t,d=i[u],g=d,m=g&&g.prototype,x={},w=function(t,e){C(t,e,{get:function(){return function(t,e){var n=N(t);return n.view[c](e*s+n.byteOffset,!0)}(this,e)},set:function(t){return function(t,e,r){var i=N(t);n&&(r=(r=z(r))<0?0:r>255?255:255&r),i.view[l](e*s+i.byteOffset,r,!0)}(this,e,t)},enumerable:!0})};U?a&&(g=e((function(t,e,n,r){return f(t,m),j(b(e)?J(e)?void 0!==r?new d(e,v(n,s),r):void 0!==n?new d(e,v(n,s)):new d(e):q(e)?K(g,e):o(k,g,e):new d(y(e)),t,g)})),E&&E(g,W),T(O(d),(function(t){t in g||h(g,t,d[t])})),g.prototype=m):(g=e((function(t,e,n,r){f(t,m);var i,a,u,c=0,l=0;if(b(e)){if(!J(e))return q(e)?K(g,e):o(k,g,e);i=e,l=v(n,s);var h=e.byteLength;if(void 0===r){if(h%s)throw F("Wrong length");if((a=h-l)<0)throw F("Wrong length")}else if((a=p(r)*s)+l>h)throw F("Wrong length");u=a/s}else u=y(e),i=new B(a=u*s);for(R(t,{buffer:i,byteOffset:l,byteLength:a,length:u,view:new G(i)});c<u;)w(t,c++)})),E&&E(g,W),m=g.prototype=S(Y)),m.constructor!==g&&h(m,"constructor",g),V(m).TypedArrayConstructor=g,H&&h(m,H,u);var A=g!=d;x[u]=g,r({global:!0,constructor:!0,forced:A,sham:!U},x),"BYTES_PER_ELEMENT"in g||h(g,"BYTES_PER_ELEMENT",s),"BYTES_PER_ELEMENT"in m||h(m,"BYTES_PER_ELEMENT",s),I(u)}):t.exports=function(){}},function(t,e,n){var r=n(4),i=n(2),o=n(98),s=n(5).NATIVE_ARRAY_BUFFER_VIEWS,a=r.ArrayBuffer,u=r.Int8Array;t.exports=!s||!i((function(){u(1)}))||!i((function(){new u(-1)}))||!o((function(t){new u,new u(null),new u(1.5),new u(t)}),!0)||i((function(){return 1!==new u(new a(2),1,void 0).length}))},function(t,e,n){var r=n(13),i=Math.floor;t.exports=Number.isInteger||function(t){return!r(t)&&isFinite(t)&&i(t)===t}},function(t,e,n){var r=n(20),i=RangeError;t.exports=function(t){var e=r(t);if(e<0)throw i("The argument can't be less than 0");return e}},function(t,e,n){var r=n(45),i=n(9),o=n(141),s=n(16),a=n(14),u=n(96),c=n(72),f=n(95),l=n(211),h=n(5).aTypedArrayConstructor,d=n(157);t.exports=function(t){var e,n,p,y,v,g,m,x,b=o(this),w=s(t),S=arguments.length,A=S>1?arguments[1]:void 0,E=void 0!==A,O=c(w);if(O&&!f(O))for(x=(m=u(w,O)).next,w=[];!(g=i(x,m)).done;)w.push(g.value);for(E&&S>2&&(A=r(A,arguments[2])),n=a(w),p=new(h(b))(n),y=l(p),e=0;n>e;e++)v=E?A(w[e],e):w[e],p[e]=y?d(v):+v;return p}},function(t,e,n){var r=n(32),i=n(1)("".slice);t.exports=function(t){return"Big"===i(r(t),0,3)}},function(t,e,n){"use strict";var r=n(1),i=n(5),o=r(n(213)),s=i.aTypedArray;(0,i.exportTypedArrayMethod)("copyWithin",(function(t,e){return o(s(this),t,e,arguments.length>2?arguments[2]:void 0)}))},function(t,e,n){"use strict";var r=n(16),i=n(31),o=n(14),s=n(108),a=Math.min;t.exports=[].copyWithin||function(t,e){var n=r(this),u=o(n),c=i(t,u),f=i(e,u),l=arguments.length>2?arguments[2]:void 0,h=a((void 0===l?u:i(l,u))-f,u-c),d=1;for(f<c&&c<f+h&&(d=-1,f+=h-1,c+=h-1);h-- >0;)f in n?n[c]=n[f]:s(n,c),c+=d,f+=d;return n}},function(t,e,n){"use strict";var r=n(5),i=n(17).every,o=r.aTypedArray;(0,r.exportTypedArrayMethod)("every",(function(t){return i(o(this),t,arguments.length>1?arguments[1]:void 0)}))},function(t,e,n){"use strict";var r=n(5),i=n(155),o=n(157),s=n(32),a=n(9),u=n(1),c=n(2),f=r.aTypedArray,l=r.exportTypedArrayMethod,h=u("".slice);l("fill",(function(t){var e=arguments.length;f(this);var n="Big"===h(s(this),0,3)?o(t):+t;return a(i,this,n,e>1?arguments[1]:void 0,e>2?arguments[2]:void 0)}),c((function(){var t=0;return new Int8Array(2).fill({valueOf:function(){return t++}}),1!==t})))},function(t,e,n){"use strict";var r=n(5),i=n(17).filter,o=n(217),s=r.aTypedArray;(0,r.exportTypedArrayMethod)("filter",(function(t){var e=i(s(this),t,arguments.length>1?arguments[1]:void 0);return o(this,e)}))},function(t,e,n){var r=n(218),i=n(85);t.exports=function(t,e){return r(i(t),e)}},function(t,e,n){var r=n(14);t.exports=function(t,e){for(var n=0,i=r(e),o=new t(i);i>n;)o[n]=e[n++];return o}},function(t,e,n){"use strict";var r=n(5),i=n(17).find,o=r.aTypedArray;(0,r.exportTypedArrayMethod)("find",(function(t){return i(o(this),t,arguments.length>1?arguments[1]:void 0)}))},function(t,e,n){"use strict";var r=n(5),i=n(17).findIndex,o=r.aTypedArray;(0,r.exportTypedArrayMethod)("findIndex",(function(t){return i(o(this),t,arguments.length>1?arguments[1]:void 0)}))},function(t,e,n){"use strict";var r=n(5),i=n(17).forEach,o=r.aTypedArray;(0,r.exportTypedArrayMethod)("forEach",(function(t){i(o(this),t,arguments.length>1?arguments[1]:void 0)}))},function(t,e,n){"use strict";var r=n(5),i=n(55).includes,o=r.aTypedArray;(0,r.exportTypedArrayMethod)("includes",(function(t){return i(o(this),t,arguments.length>1?arguments[1]:void 0)}))},function(t,e,n){"use strict";var r=n(5),i=n(55).indexOf,o=r.aTypedArray;(0,r.exportTypedArrayMethod)("indexOf",(function(t){return i(o(this),t,arguments.length>1?arguments[1]:void 0)}))},function(t,e,n){"use strict";var r=n(4),i=n(2),o=n(1),s=n(5),a=n(51),u=n(6)("iterator"),c=r.Uint8Array,f=o(a.values),l=o(a.keys),h=o(a.entries),d=s.aTypedArray,p=s.exportTypedArrayMethod,y=c&&c.prototype,v=!i((function(){y[u].call([1])})),g=!!y&&y.values&&y[u]===y.values&&"values"===y.values.name,m=function(){return f(d(this))};p("entries",(function(){return h(d(this))}),v),p("keys",(function(){return l(d(this))}),v),p("values",m,v||!g,{name:"values"}),p(u,m,v||!g,{name:"values"})},function(t,e,n){"use strict";var r=n(5),i=n(1),o=r.aTypedArray,s=r.exportTypedArrayMethod,a=i([].join);s("join",(function(t){return a(o(this),t)}))},function(t,e,n){"use strict";var r=n(5),i=n(49),o=n(139),s=r.aTypedArray;(0,r.exportTypedArrayMethod)("lastIndexOf",(function(t){var e=arguments.length;return i(o,s(this),e>1?[t,arguments[1]]:[t])}))},function(t,e,n){"use strict";var r=n(5),i=n(17).map,o=n(85),s=r.aTypedArray;(0,r.exportTypedArrayMethod)("map",(function(t){return i(s(this),t,arguments.length>1?arguments[1]:void 0,(function(t,e){return new(o(t))(e)}))}))},function(t,e,n){"use strict";var r=n(5),i=n(158).left,o=r.aTypedArray;(0,r.exportTypedArrayMethod)("reduce",(function(t){var e=arguments.length;return i(o(this),t,e,e>1?arguments[1]:void 0)}))},function(t,e,n){"use strict";var r=n(5),i=n(158).right,o=r.aTypedArray;(0,r.exportTypedArrayMethod)("reduceRight",(function(t){var e=arguments.length;return i(o(this),t,e,e>1?arguments[1]:void 0)}))},function(t,e,n){"use strict";var r=n(5),i=r.aTypedArray,o=r.exportTypedArrayMethod,s=Math.floor;o("reverse",(function(){for(var t,e=i(this).length,n=s(e/2),r=0;r<n;)t=this[r],this[r++]=this[--e],this[e]=t;return this}))},function(t,e,n){"use strict";var r=n(4),i=n(9),o=n(5),s=n(14),a=n(156),u=n(16),c=n(2),f=r.RangeError,l=r.Int8Array,h=l&&l.prototype,d=h&&h.set,p=o.aTypedArray,y=o.exportTypedArrayMethod,v=!c((function(){var t=new Uint8ClampedArray(2);return i(d,t,{length:1,0:3},1),3!==t[1]})),g=v&&o.NATIVE_ARRAY_BUFFER_VIEWS&&c((function(){var t=new l(2);return t.set(1),t.set("2",1),0!==t[0]||2!==t[1]}));y("set",(function(t){p(this);var e=a(arguments.length>1?arguments[1]:void 0,1),n=u(t);if(v)return i(d,this,n,e);var r=this.length,o=s(n),c=0;if(o+e>r)throw f("Wrong length");for(;c<o;)this[e+c]=n[c++]}),!v||g)},function(t,e,n){"use strict";var r=n(5),i=n(85),o=n(2),s=n(82),a=r.aTypedArray;(0,r.exportTypedArrayMethod)("slice",(function(t,e){for(var n=s(a(this),t,e),r=i(this),o=0,u=n.length,c=new r(u);u>o;)c[o]=n[o++];return c}),o((function(){new Int8Array(1).slice()})))},function(t,e,n){"use strict";var r=n(5),i=n(17).some,o=r.aTypedArray;(0,r.exportTypedArrayMethod)("some",(function(t){return i(o(this),t,arguments.length>1?arguments[1]:void 0)}))},function(t,e,n){"use strict";var r=n(4),i=n(1),o=n(2),s=n(42),a=n(159),u=n(5),c=n(160),f=n(161),l=n(65),h=n(162),d=u.aTypedArray,p=u.exportTypedArrayMethod,y=r.Uint16Array,v=y&&i(y.prototype.sort),g=!(!v||o((function(){v(new y(2),null)}))&&o((function(){v(new y(2),{})}))),m=!!v&&!o((function(){if(l)return l<74;if(c)return c<67;if(f)return!0;if(h)return h<602;var t,e,n=new y(516),r=Array(516);for(t=0;t<516;t++)e=t%4,n[t]=515-t,r[t]=t-2*e+3;for(v(n,(function(t,e){return(t/4|0)-(e/4|0)})),t=0;t<516;t++)if(n[t]!==r[t])return!0}));p("sort",(function(t){return void 0!==t&&s(t),m?v(this,t):a(d(this),function(t){return function(e,n){return void 0!==t?+t(e,n)||0:n!=n?-1:e!=e?1:0===e&&0===n?1/e>0&&1/n<0?1:-1:e>n}}(t))}),!m||g)},function(t,e,n){"use strict";var r=n(5),i=n(21),o=n(31),s=n(85),a=r.aTypedArray;(0,r.exportTypedArrayMethod)("subarray",(function(t,e){var n=a(this),r=n.length,u=o(t,r);return new(s(n))(n.buffer,n.byteOffset+u*n.BYTES_PER_ELEMENT,i((void 0===e?r:o(e,r))-u))}))},function(t,e,n){"use strict";var r=n(4),i=n(49),o=n(5),s=n(2),a=n(82),u=r.Int8Array,c=o.aTypedArray,f=o.exportTypedArrayMethod,l=[].toLocaleString,h=!!u&&s((function(){l.call(new u(1))}));f("toLocaleString",(function(){return i(l,h?a(c(this)):c(this),a(arguments))}),s((function(){return[1,2].toLocaleString()!=new u([1,2]).toLocaleString()}))||!s((function(){u.prototype.toLocaleString.call([1,2])})))},function(t,e,n){"use strict";var r=n(5).exportTypedArrayMethod,i=n(2),o=n(4),s=n(1),a=o.Uint8Array,u=a&&a.prototype||{},c=[].toString,f=s([].join);i((function(){c.call({})}))&&(c=function(){return f(this)});var l=u.toString!=c;r("toString",c,l)},function(t,e,n){"use strict";var r=n(45),i=n(9),o=n(16),s=n(239),a=n(95),u=n(75),c=n(14),f=n(71),l=n(96),h=n(72),d=Array;t.exports=function(t){var e=o(t),n=u(this),p=arguments.length,y=p>1?arguments[1]:void 0,v=void 0!==y;v&&(y=r(y,p>2?arguments[2]:void 0));var g,m,x,b,w,S,A=h(e),E=0;if(!A||this===d&&a(A))for(g=c(e),m=n?new this(g):d(g);g>E;E++)S=v?y(e[E],E):e[E],f(m,E,S);else for(w=(b=l(e,A)).next,m=n?new this:[];!(x=i(w,b)).done;E++)S=v?s(b,y,[x.value,E],!0):x.value,f(m,E,S);return m.length=E,m}},function(t,e,n){var r=n(12),i=n(126);t.exports=function(t,e,n,o){try{return o?e(r(n)[0],n[1]):e(n)}catch(e){i(t,"throw",e)}}},function(t,e,n){n(241)},function(t,e,n){"use strict";n(122)("Set",(function(t){return function(){return t(this,arguments.length?arguments[0]:void 0)}}),n(127))},function(t,e,n){var r=n(1),i=n(18),o=Date.prototype,s=r(o.toString),a=r(o.getTime);"Invalid Date"!=String(new Date(NaN))&&i(o,"toString",(function(){var t=a(this);return t==t?s(this):"Invalid Date"}))},function(t,e,n){var r=n(3),i=n(16),o=n(89);r({target:"Object",stat:!0,forced:n(2)((function(){o(1)}))},{keys:function(t){return o(i(t))}})},function(t,e,n){"use strict";var r,i=n(3),o=n(1),s=n(44).f,a=n(21),u=n(15),c=n(137),f=n(25),l=n(138),h=n(39),d=o("".startsWith),p=o("".slice),y=Math.min,v=l("startsWith");i({target:"String",proto:!0,forced:!!(h||v||(r=s(String.prototype,"startsWith"),!r||r.writable))&&!v},{startsWith:function(t){var e=u(f(this));c(t);var n=a(y(arguments.length>1?arguments[1]:void 0,e.length)),r=u(t);return d?d(e,r,n):p(e,n,n+r.length)===r}})},function(t,e,n){"use strict";n.r(e),n.d(e,"factory",(function(){return nt}));n(51),n(176),n(29),n(74),n(130),n(33),n(37),n(48),n(77),n(78),n(79),n(80),n(24);var r=n(0);n(50),n(184),n(140),n(142),n(143),n(144);function i(){var t,e,n,r=(e=navigator.userAgent,n=e.match(/(opera|chrome|safari|firefox|msie|trident(?=\/))\/?\s*(\d+)/i)||[],/trident/i.test(n[1])?"IE "+((t=/\brv[ :]+(\d+)/g.exec(e)||[])[1]||""):"Chrome"===n[1]&&null!==(t=e.match(/\b(OPR|Edge)\/(\d+)/))?t.slice(1).join(" ").replace("OPR","Opera"):(n=n[2]?[n[1],n[2]]:[navigator.appName,navigator.appVersion,"-?"],null!==(t=e.match(/version\/(\d+)/i))&&n.splice(1,1,t[1]),n.join(" ")));return"IE 11"==r||"MSIE"===r.substring(0,4)}n(60),n(83),n(109),n(110),n(61);function o(t,e){for(var n=0;n<e.length;n++){var r=e[n];r.enumerable=r.enumerable||!1,r.configurable=!0,"value"in r&&(r.writable=!0),Object.defineProperty(t,r.key,r)}}var s=function(){function t(e,n,r){!function(t,e){if(!(t instanceof e))throw new TypeError("Cannot call a class as a function")}(this,t),this.containerid=e,this.svgid=n,this.clsName=r.clsName,this.position=r.position,this.delay_over=r.delay_over,this.delay_out=r.delay_out,this.tooltips=r.tooltips?r.tooltips:{},this.hidden=Array.isArray(r.hidden)?r.hidden:[r.hidden],this.on=!1}var e,n,i;return e=t,(n=[{key:"init",value:function(t){if(!t.length)return!1;var e=r.select("#"+this.containerid+" .girafe_container_std").append("div").attr("id",this.svgid+"_toolbar").classed(this.clsName,!0).classed(this.clsName+"-"+this.position,!0).attr("data-forsvg",this.svgid),n=this,i=t.map((function(t){return t.getButtons().map((function(t){return t.block}))})).flat().filter((function(t,e,n){return n.indexOf(t)===e})).filter((function(t){return!n.hidden.includes(t)}));if(!i.length)return e.remove(),!1;var o={};return i.forEach((function(t){o[t]=e.append("xhtml:div").classed(n.clsName+"-block",!0).classed(n.clsName+"-block-"+t,!0).attr("data-forsvg",n.svgid)})),t.forEach((function(t){t.getButtons().forEach((function(t){if(!n.hidden.includes(t.key)&&!n.hidden.includes(t.block)){var e=t.states,r=t.key,i=t;t.states&&(i=e[r=t.current_state]),n.tooltips[r]&&(i.tooltip=n.tooltips[r]),o[t.block].append("xhtml:a").classed(n.clsName+"-icon",!0).classed(n.clsName+"-icon-"+t.key,!0).classed(i.class,!0).classed(i.unclass,!1).attr("title",i.tooltip).attr("data-forsvg",n.svgid).html(i.icon).datum(e).on("click",t.onclick)}}))})),!!e.selectAll("."+this.clsName+"-icon").size()||(e.remove(),!1)}},{key:"destroy",value:function(){var t=r.select("#"+this.svgid+"_toolbar");try{t.selectAll("."+this.clsName+"-icon").on("click",null)}catch(t){console.error(t)}t.remove()}},{key:"clear",value:function(t){this.on&&!this.isValidTarget(t.relatedTarget)&&(this.on=!1,r.select("#"+this.containerid).select("."+this.clsName).transition().duration(this.delay_out).style("opacity",0))}},{key:"isValidTarget",value:function(t){return t&&t.getAttribute("data-forsvg")===this.svgid}},{key:"applyOn",value:function(){this.on||(this.on=!0,r.select("#"+this.svgid+"_toolbar").transition().duration(this.delay_over).style("opacity",.8))}}])&&o(e.prototype,n),i&&o(e,i),Object.defineProperty(e,"prototype",{writable:!1}),t}();function a(t,e){for(var n=0;n<e.length;n++){var r=e[n];r.enumerable=r.enumerable||!1,r.configurable=!0,"value"in r&&(r.writable=!0),Object.defineProperty(t,r.key,r)}}var u=function(){function t(e,n){!function(t,e){if(!(t instanceof e))throw new TypeError("Cannot call a class as a function")}(this,t),this.svgid=e,this.min=n.min,this.max=n.max,this.duration=n.duration,this.zoomer=null,this.on=!1}var e,n,i;return e=t,(n=[{key:"init",value:function(){return!(this.max<=this.min||(this.zoomer=r.zoom().scaleExtent([this.min,this.max]).duration(this.duration),this.on=!1,0))}},{key:"destroy",value:function(){this.zoomOff(),this.zoomer=null}},{key:"getButtons",value:function(){var t=this;return[{key:"zoom_onoff",block:"zoom",current_state:"zoom_on",states:{zoom_on:{class:"neutral",unclass:"drop",tooltip:"activate pan/zoom",icon:"<svg xmlns='http://www.w3.org/2000/svg' role='img' viewBox='0 0 512 512'><g><ellipse ry='150' rx='150' cy='213' cx='203.5' stroke-width='50' fill='transparent'/><line y2='455.5' x2='416' y1='331.5' x1='301' stroke-width='50'/></g></svg>"},zoom_off:{class:"drop",unclass:"neutral",tooltip:"deactivate pan/zoom",icon:"<svg xmlns='http://www.w3.org/2000/svg' role='img' viewBox='0 0 512 512'><g><ellipse ry='150' rx='150' cy='213' cx='203.5' stroke-width='50' fill='transparent'/><line y2='455.5' x2='416' y1='331.5' x1='301' stroke-width='50'/><line y2='455' x2='0' y1='0' x1='416' stroke-width='30'/></g></svg>"}},onclick:function(e){var n,i=r.select(this);t.on?(t.zoomOff(),n=e.zoom_on):(t.zoomOn(),n=e.zoom_off),i.attr("title",n.tooltip).classed(n.class,!0).classed(n.unclass,!1).html(n.icon)}},{key:"zoom_rect",block:"zoom",class:"neutral",tooltip:"zoom with rectangle",icon:"<svg xmlns='http://www.w3.org/2000/svg' role='img' viewBox='0 0 512 512'><g><ellipse ry='150' rx='150' cy='213' cx='203.5' stroke-width='50' fill='transparent'/><line y2='455.5' x2='416' y1='331.5' x1='301' stroke-width='50'/><rect x='126' y='153' width='152' height='116' stroke-width='25' fill='transparent'/></g></svg>",onclick:function(){t.zoomRect()}},{key:"zoom_reset",block:"zoom",class:"neutral",tooltip:"reset pan/zoom",icon:"<svg xmlns='http://www.w3.org/2000/svg' role='img' viewBox='0 0 512 512'><g><polygon points='274,209.7 337.9,145.9 288,96 416,96 416,224 366.1,174.1 302.3,238 '/><polygon points='274,302.3 337.9,366.1 288,416 416,416 416,288 366.1,337.9 302.3,274'/><polygon points='238,302.3 174.1,366.1 224,416 96,416 96,288 145.9,337.9 209.7,274'/><polygon points='238,209.7 174.1,145.9 224,96 96,96 96,224 145.9,174.1 209.7,238'/></g></svg>",onclick:function(){t.zoomIdentity()}}]}},{key:"zoomOn",value:function(){if(!this.on){this.on=!0;var t=r.select("#"+this.svgid+"_rootg");r.select("#"+this.svgid).call(this.zoomer.on("start",(function(){t.style("cursor","move")})).on("zoom",(function(){t.attr("transform",r.event.transform)})).on("end",(function(){t.style("cursor","auto")})))}}},{key:"zoomIdentity",value:function(){var t=r.zoomTransform(r.select("#"+this.svgid+"_rootg").node());if(1!==t.k||0!==t.x||0!==t.y){var e=this.on;e||this.zoomOn();var n=this;r.select("#"+this.svgid).transition().duration(this.duration).on("end",(function(){e||n.zoomOff()})).call(this.zoomer.transform,r.zoomIdentity)}}},{key:"zoomOff",value:function(){this.on&&(this.on=!1,r.select("#"+this.svgid).on(".zoom",null))}},{key:"zoomRect",value:function(){var t=this.on;t||this.zoomOn();var e=r.select("#"+this.svgid),n=r.select("#"+this.svgid+"_rootg"),i=this,o=function(){var t,e=new DOMRect,n={start:function(){},draw:function(){},end:function(){}};function i(i){var o,s,a=i.append("g").attr("class","dragrect").append("rect"),u=r.drag().on("start",(function(){o=null,s=null,n.start()})).on("drag",(function(){var t=r.mouse(this)[0],i=r.mouse(this)[1];o?(s.x=t,s.y=i):(o=new DOMPoint(t,i),s=new DOMPoint(t,i));var u=Math.min(o.x,s.x),c=Math.max(o.x,s.x),f=Math.min(o.y,s.y),l=Math.max(o.y,s.y);e.x=u,e.y=f,e.width=c-u,e.height=l-f,a.attr("x",e.x),a.attr("y",e.y),a.attr("width",e.width),a.attr("height",e.height),n.draw()})).on("end",(function(){a.attr("display","none"),n.end()}));t.call(u)}return i.on=function(t,e){if(!arguments.length)return n;if(1===arguments.length)return n[t];var r=["start","draw","end"];return r.indexOf(t)>-1&&(n[t]=e),i},i.targetArea=function(e){return arguments.length?(t=e,i):t},i.rect=function(){return e},i}();try{n.call(o.targetArea(n).on("start",(function(){n.style("cursor","zoom-in")})).on("end",(function(){try{n.style("cursor","auto"),n.on(".dragstart",null).on(".drag",null).on(".dragend",null),n.selectAll("g.dragrect").remove();var s=o.rect();if(s.width>=10&&s.height>=10){var a=s.x+s.width/2,u=s.y+s.height/2,c=n.node().getBBox(),f=c.x+c.width/2,l=c.y+c.height/2,h=c.width/c.height;s.height>s.width?(s.width=s.height*h,s.x=a-s.width/2):(s.height=s.width/h,s.y=u-s.height/2);var d=Math.max(Math.min(c.width/s.width,i.max),i.min);s.width=c.width/d,s.height=c.height/d,s.x=a-s.width/2,s.y=u-s.height/2;var p=(f-a-(c.width-s.width)/2)*d,y=(l-u-(c.height-s.height)/2)*d;e.transition().duration(i.duration).on("end",(function(){t||i.zoomOff()})).call(i.zoomer.transform,r.zoomIdentity.translate(p,y).scale(d))}}catch(t){console.error(t)}})))}catch(t){console.error(t)}}}])&&a(e.prototype,n),i&&a(e,i),Object.defineProperty(e,"prototype",{writable:!1}),t}();n(84),n(146),n(148),n(188),n(191),n(192),n(201),n(202),n(203),n(205),n(212),n(214),n(215),n(216),n(219),n(220),n(221),n(222),n(223),n(224),n(225),n(226),n(227),n(228),n(229),n(230),n(231),n(232),n(233),n(234),n(235),n(236),n(237);function c(t,e){for(var n=0;n<e.length;n++){var r=e[n];r.enumerable=r.enumerable||!1,r.configurable=!0,"value"in r&&(r.writable=!0),Object.defineProperty(t,r.key,r)}}function f(t,e,n){return e&&c(t.prototype,e),n&&c(t,n),Object.defineProperty(t,"prototype",{writable:!1}),t}function l(t,e){if(!(t instanceof e))throw new TypeError("Cannot call a class as a function")}function h(t){var e=t.ownerDocument,n=e.documentElement,r=e.defaultView;return new window.DOMRect(r.pageXOffset||n.scrollLeft||0,r.pageYOffset||n.scrollTop||0,Math.max(n.clientWidth||0,r.innerWidth||0),Math.max(n.clientHeight||0,r.innerHeight||0))}function d(t){for(var e=new window.DOMMatrix,n=e,r=t,i=t.ownerDocument.defaultView||window,o=h(t);null!=r&&r!==r.ownerDocument.documentElement;){var s=i.getComputedStyle(r,void 0).transform||"none";n=("none"===s?e:new window.DOMMatrix(s)).multiply(n),r=r.parentNode}var a=t.offsetWidth,u=t.offsetHeight,c=new window.DOMPoint(0,0).matrixTransform(n),f=new window.DOMPoint(a,0).matrixTransform(n),l=new window.DOMPoint(a,u).matrixTransform(n),d=new window.DOMPoint(0,u).matrixTransform(n),p=Math.min(c.x,f.x,l.x,d.x),y=Math.min(c.y,f.y,l.y,d.y),v=t.getBoundingClientRect();return n=e.translate(o.x+v.left-p,o.y+v.top-y,0).multiply(n)}function p(t,e){return Math.sqrt(Math.pow(e.x-t.x,2)+Math.pow(e.y-t.y,2))}function y(t,e,n){if(e.x===n.x&&e.y===n.y)return p(t,e);var r=function(t,e){return Math.pow(e.x-t.x,2)+Math.pow(e.y-t.y,2)}(e,n),i=((t.x-e.x)*(n.x-e.x)+(t.y-e.y)*(n.y-e.y))/r;if(i<=0)return p(t,e);if(i>=1)return p(t,n);var o=((e.y-t.y)*(n.x-e.x)-(e.x-t.x)*(n.y-e.y))/r;return Math.abs(o)*Math.sqrt(r)}function v(t,e){var n,r,i,o,s,a,u,c,f,l,h,d,p,y,v=t.x,g=t.y,m=t.x+t.width,x=t.y+t.height;if(e.find((function(t){return t.x>=v&&t.x<=m&&t.y>=g&&t.y<=x})))return!0;if(1===e.length&&e[0].r){var b=e[0],w=Math.abs(b.x-(t.x+t.width/2)),S=Math.abs(b.y-(t.y+t.height/2));return!(w>t.width/2+b.r)&&(!(S>t.height/2+b.r)&&(w<=t.width/2||(S<=t.height/2||(w-t.width/2^S-t.height/2+2^2)<=(2^b.r))))}if(e.length>1)for(n=0;n<e.length;n++)if((r=n+1)==e.length&&(r=0),i=v,o=g,s=m,a=x,u=e[n].x,c=e[n].y,f=e[r].x,l=e[r].y,h=void 0,d=void 0,p=void 0,y=void 0,h=!1,p=(f-u)*(o-c)-(l-c)*(i-u),y=(s-i)*(o-c)-(a-o)*(i-u),0==(d=(l-c)*(s-i)-(f-u)*(a-o))?0==p&&(i==s?o<c&&Math.max(o,a)<Math.min(c,l)||c<o&&Math.max(c,l)<Math.min(o,a)||(h=!0):i<u&&Math.max(i,s)<Math.min(u,f)||u<i&&Math.max(u,f)<Math.min(i,s)||(h=!0)):(y/=d,(p/=d)>0&&p<1&&y>0&&y<1&&(h=!0)),h)return!0;return!1}function g(t){var e,n,r,i,o,s,a=new RegExp("([\\d\\.]+) ([\\d\\.]+)","g");if(t instanceof SVGRect||t instanceof DOMRect)return[new DOMPoint(t.x,t.y),new DOMPoint(t.x+t.width,t.y),new DOMPoint(t.x+t.width,t.y+t.height),new DOMPoint(t.x,t.y+t.height)];if(t instanceof SVGRectElement||t instanceof SVGImageElement)return[new DOMPoint(t.x.baseVal.value,t.y.baseVal.value),new DOMPoint(t.x.baseVal.value+t.width.baseVal.value,t.y.baseVal.value),new DOMPoint(t.x.baseVal.value+t.width.baseVal.value,t.y.baseVal.value+t.height.baseVal.value),new DOMPoint(t.x.baseVal.value,t.y.baseVal.value+t.height.baseVal.value)];if(t instanceof SVGCircleElement)return(n=new DOMPoint(t.cx.baseVal.value,t.cy.baseVal.value)).r=t.r.baseVal.value,[n];if(t instanceof SVGLineElement)return(e=[new DOMPoint(t.x1.baseVal.value,t.y1.baseVal.value),new DOMPoint(t.x2.baseVal.value,t.y2.baseVal.value)]).shape="polyline",e;if(t instanceof SVGPolylineElement||t instanceof SVGPolygonElement){for(e=[],r=t.points,i=0;i<r.length;i++)(n=r.getItem(i))instanceof DOMPoint||(n=new DOMPoint(n.x,n.y)),e.push(n);return e.shape=t instanceof SVGPolylineElement?"polyline":"polygon",e}if(t instanceof SVGPathElement){for(e=[],o=t.getAttribute("d");null!==(s=a.exec(o));)e.push(new DOMPoint(parseFloat(s[1]),parseFloat(s[2])));return e}throw"Error in extractPoints: unimplemented object type: "+Object.prototype.toString.call(t)}function m(t){var e=t.map((function(t){return t.x})),n=t.map((function(t){return t.y}));return{minx:Math.min.apply(null,e),maxx:Math.max.apply(null,e),miny:Math.min.apply(null,n),maxy:Math.max.apply(null,n)}}function x(t,e){var n=e.createSVGRect();return n.x=t.minx,n.y=t.miny,n.width=t.maxx-t.minx,n.height=t.maxy-t.miny,n}function b(t,e){return x(m(t),e)}function w(t,e,n){var r,i=arguments.length>3&&void 0!==arguments[3]?arguments[3]:"#ff000033",o=arguments.length>4&&void 0!==arguments[4]?arguments[4]:"#ff0000",s="http://www.w3.org/2000/svg",a=document.getElementById(n);if(Array.isArray(t)&&1===t.length&&(t=t[0]),a?r=a.localName:(r=t.shape||"polygon",t instanceof SVGPoint||t instanceof DOMPoint?r="circle":(t instanceof SVGRect||t instanceof DOMRect)&&(r="rect"),(a=document.createElementNS(s,r)).setAttribute("id",n),"polyline"!==r&&a.setAttribute("fill",i),a.setAttribute("stroke",o),a.setAttribute("style","pointer-events: none;"),e.appendChild(a)),"circle"===r)a.setAttribute("cx",t.x),a.setAttribute("cy",t.y),a.setAttribute("r",t.r||"3pt");else if("rect"===r)a.setAttribute("x",t.x),a.setAttribute("y",t.y),a.setAttribute("width",t.width),a.setAttribute("height",t.height);else if("polygon"===r||"polyline"===r){var u="";t.forEach((function(t){u+=t.x+" "+t.y+" "})),a.setAttribute("points",u)}return a}window.DOMPoint=window.DOMPoint||f((function t(){var e=arguments.length>0&&void 0!==arguments[0]?arguments[0]:0,n=arguments.length>1&&void 0!==arguments[1]?arguments[1]:0,r=arguments.length>2&&void 0!==arguments[2]?arguments[2]:0,i=arguments.length>3&&void 0!==arguments[3]?arguments[3]:1;l(this,t),this.x=e,this.y=n,this.z=r,this.w=i})),window.DOMPoint.prototype.matrixTransform=window.DOMPoint.prototype.matrixTransform||function(t){return(t.is2D||t instanceof SVGMatrix)&&0===this.z&&1===this.w?new DOMPoint(this.x*t.a+this.y*t.c+t.e,this.x*t.b+this.y*t.d+t.f,0,1):new DOMPoint(this.x*t.m11+this.y*t.m21+this.z*t.m31+this.w*t.m41,this.x*t.m12+this.y*t.m22+this.z*t.m32+this.w*t.m42,this.x*t.m13+this.y*t.m23+this.z*t.m33+this.w*t.m43,this.x*t.m14+this.y*t.m24+this.z*t.m34+this.w*t.m44)};for(var S=function(){function t(){var e=arguments.length>0&&void 0!==arguments[0]?arguments[0]:0,n=arguments.length>1&&void 0!==arguments[1]?arguments[1]:0,r=arguments.length>2&&void 0!==arguments[2]?arguments[2]:0,i=arguments.length>3&&void 0!==arguments[3]?arguments[3]:0;l(this,t),this.x=e,this.y=n,this.width=r,this.height=i}return f(t,[{key:"top",get:function(){return this.y}},{key:"left",get:function(){return this.x}},{key:"right",get:function(){return this.x+this.width}},{key:"bottom",get:function(){return this.y+this.height}}]),t}(),A=0,E=["top","right","bottom","left"];A<E.length;A++){var O=E[A],k=Object.getOwnPropertyDescriptor(S.prototype,O);k.enumerable=!0,Object.defineProperty(S.prototype,O,k)}window.DOMRect=window.DOMRect||S;for(var T=Symbol(),I=Symbol(),M=function(t){var e=t.split(/\(/,1)[0];if("matrix"===e)return function(t){var e=t.replace(/matrix\(/,"");if(6!==(e=e.split(/,/,7)).length)throw new Error("Failed to parse ".concat(t));return[(e=e.map(parseFloat))[0],e[1],0,0,e[2],e[3],0,0,0,0,1,0,e[4],e[5],0,1]}(t);if("matrix3d"===e)return function(t){var e=t.replace(/matrix3d\(/,"");if(16!==(e=e.split(/,/,17)).length)throw new Error("Failed to parse ".concat(t));return e.map(parseFloat)}(t);throw new Error("".concat(e," parsing not implemented"))},_=function(t,e,n){if("number"!=typeof n)throw new TypeError("Expected number");t[T][e]=n},P=function(t,e,n){if("number"!=typeof n)throw new TypeError("Expected number");10===e||15===e?1!==n&&(t[I]=!1):0!==n&&(t[I]=!1),t[T][e]=n},j=function(t){var e=Object.create(R.prototype);return e.constructor=R,e[I]=!0,e[T]=t,e},N=function(t,e){for(var n=new Float64Array(16),r=0;r<4;r++)for(var i=0;i<4;i++){for(var o=0,s=0;s<4;s++)o+=t[4*r+s]*e[4*s+i];n[4*r+i]=o}return n},R=function(){function t(e){if(l(this,t),this[I]=!0,this[T]=new Float64Array([1,0,0,0,0,1,0,0,0,0,1,0,0,0,0,1]),"string"==typeof e){if(""===e)return;var n=e.split(/\)\s+/,20).map(M);if(0===n.length)return;e=n[0];for(var r=1;r<n.length;r++)e=N(n[r],e)}var i=0;if(e&&6===e.length)_(this,0,e[i++]),_(this,1,e[i++]),_(this,4,e[i++]),_(this,5,e[i++]),_(this,12,e[i++]),_(this,13,e[i++]);else if(e&&16===e.length)_(this,0,e[i++]),_(this,1,e[i++]),P(this,2,e[i++]),P(this,3,e[i++]),_(this,4,e[i++]),_(this,5,e[i++]),P(this,6,e[i++]),P(this,7,e[i++]),P(this,8,e[i++]),P(this,9,e[i++]),P(this,10,e[i++]),P(this,11,e[i++]),_(this,12,e[i++]),_(this,13,e[i++]),P(this,14,e[i++]),P(this,15,e[i]);else if(void 0!==e)throw new TypeError("Expected string or array.")}return f(t,[{key:"m11",get:function(){return this[T][0]},set:function(t){_(this,0,t)}},{key:"m12",get:function(){return this[T][1]},set:function(t){_(this,1,t)}},{key:"m13",get:function(){return this[T][2]},set:function(t){P(this,2,t)}},{key:"m14",get:function(){return this[T][3]},set:function(t){P(this,3,t)}},{key:"m21",get:function(){return this[T][4]},set:function(t){_(this,4,t)}},{key:"m22",get:function(){return this[T][5]},set:function(t){_(this,5,t)}},{key:"m23",get:function(){return this[T][6]},set:function(t){P(this,6,t)}},{key:"m24",get:function(){return this[T][7]},set:function(t){P(this,7,t)}},{key:"m31",get:function(){return this[T][8]},set:function(t){P(this,8,t)}},{key:"m32",get:function(){return this[T][9]},set:function(t){P(this,9,t)}},{key:"m33",get:function(){return this[T][10]},set:function(t){P(this,10,t)}},{key:"m34",get:function(){return this[T][11]},set:function(t){P(this,11,t)}},{key:"m41",get:function(){return this[T][12]},set:function(t){_(this,12,t)}},{key:"m42",get:function(){return this[T][13]},set:function(t){_(this,13,t)}},{key:"m43",get:function(){return this[T][14]},set:function(t){P(this,14,t)}},{key:"m44",get:function(){return this[T][15]},set:function(t){P(this,15,t)}},{key:"a",get:function(){return this[T][0]},set:function(t){_(this,0,t)}},{key:"b",get:function(){return this[T][1]},set:function(t){_(this,1,t)}},{key:"c",get:function(){return this[T][4]},set:function(t){_(this,4,t)}},{key:"d",get:function(){return this[T][5]},set:function(t){_(this,5,t)}},{key:"e",get:function(){return this[T][12]},set:function(t){_(this,12,t)}},{key:"f",get:function(){return this[T][13]},set:function(t){_(this,13,t)}},{key:"is2D",get:function(){return this[I]}},{key:"isIdentity",get:function(){var t=this[T];return 1===t[0]&&0===t[1]&&0===t[2]&&0===t[3]&&0===t[4]&&1===t[5]&&0===t[6]&&0===t[7]&&0===t[8]&&0===t[9]&&1===t[10]&&0===t[11]&&0===t[12]&&0===t[13]&&0===t[14]&&1===t[15]}},{key:"multiply",value:function(t){return j(this[T]).multiplySelf(t)}},{key:"multiplySelf",value:function(t){return this[T]=N(t[T],this[T]),t.is2D||(this[I]=!1),this}},{key:"translate",value:function(t,e,n){return j(this[T]).translateSelf(t,e,n)}},{key:"translateSelf",value:function(){var t=arguments.length>0&&void 0!==arguments[0]?arguments[0]:0,e=arguments.length>1&&void 0!==arguments[1]?arguments[1]:0,n=arguments.length>2&&void 0!==arguments[2]?arguments[2]:0;return this[T]=N([1,0,0,0,0,1,0,0,0,0,1,0,t,e,n,1],this[T]),0!==n&&(this[I]=!1),this}},{key:"scale",value:function(t,e,n,r,i,o){return j(this[T]).scaleSelf(t,e,n,r,i,o)}},{key:"scale3d",value:function(t,e,n,r){return j(this[T]).scale3dSelf(t,e,n,r)}},{key:"scale3dSelf",value:function(t,e,n,r){return this.scaleSelf(t,t,t,e,n,r)}},{key:"scaleSelf",value:function(t,e,n,r,i,o){return"number"!=typeof r&&(r=0),"number"!=typeof i&&(i=0),"number"!=typeof o&&(o=0),this.translateSelf(r,i,o),"number"!=typeof t&&(t=1),"number"!=typeof e&&(e=t),"number"!=typeof n&&(n=1),this[T]=N([t,0,0,0,0,e,0,0,0,0,n,0,0,0,0,1],this[T]),this.translateSelf(-r,-i,-o),1===n&&0===o||(this[I]=!1),this}},{key:"inverse",value:function(){return j(this[T]).invertSelf()}},{key:"invertSelf",value:function(){if(!this[I])throw new Error("3D matrix inversion is not implemented.");var e=this[T][0]*this[T][5]-this[T][1]*this[T][4];if(0!==e){var n=new t;return n.a=this[T][5]/e,n.b=-this[T][1]/e,n.c=-this[T][4]/e,n.d=this[T][0]/e,n.e=(this[T][4]*this[T][13]-this[T][5]*this[T][12])/e,n.f=(this[T][1]*this[T][12]-this[T][0]*this[T][13])/e,n}this[I]=!1,this[T]=[NaN,NaN,NaN,NaN,NaN,NaN,NaN,NaN,NaN,NaN,NaN,NaN,NaN,NaN,NaN,NaN]}}]),t}(),V=0,C=["a","b","c","d","e","f","m11","m12","m13","m14","m21","m22","m23","m24","m31","m32","m33","m34","m41","m42","m43","m44","is2D","isIdentity"];V<C.length;V++){var D=C[V],z=Object.getOwnPropertyDescriptor(R.prototype,D);z.enumerable=!0,Object.defineProperty(R.prototype,D,z)}function F(t,e){for(var n=0;n<e.length;n++){var r=e[n];r.enumerable=r.enumerable||!1,r.configurable=!0,"value"in r&&(r.writable=!0),Object.defineProperty(t,r.key,r)}}window.DOMMatrix=window.DOMMatrix||R;var B=function(){function t(e,n){!function(t,e){if(!(t instanceof e))throw new TypeError("Cannot call a class as a function")}(this,t),this.svgid=e,this.clsName=n.classPrefix+"_"+this.svgid,this.placement=n.placement,this.opacity=n.opacity,this.offx=n.offx,this.offy=n.offy,this.use_cursor_pos=n.use_cursor_pos,this.use_fill=n.use_fill,this.use_stroke=n.use_stroke,this.delay_over=n.delay_over,this.delay_out=n.delay_out,this.lastTargetId=null}var e,n,i;return e=t,(n=[{key:"init",value:function(){var t=document.getElementById(this.svgid+"_rootg");return!!t.querySelector("*[title]")&&(r.select("."+this.clsName).remove(),("doc"==this.placement?r.select("body"):r.select(t.ownerSVGElement.parentNode)).append("xhtml:div").classed(this.clsName,!0).style("position","absolute").style("opacity",0),this.decodingTextarea=document.createElementNS("http://www.w3.org/1999/xhtml","textarea"),!0)}},{key:"destroy",value:function(){this.lastTargetId=null,r.select("."+this.clsName).remove()}},{key:"clear",value:function(){this.lastTargetId&&(this.lastTargetId=null,r.select("div."+this.clsName).transition().duration(this.delay_out).style("opacity",0))}},{key:"isValidTarget",value:function(t){return t instanceof SVGGraphicsElement&&!(t instanceof SVGSVGElement)&&t.hasAttribute("title")}},{key:"applyOn",value:function(t,e){try{if(this.isValidTarget(t)){var n,i=new DOMPoint(e.pageX,e.pageY),o=r.select("div."+this.clsName);if(t.id===this.lastTargetId)n=this.calculatePosition(o,t,i),o.style("left",n.x+"px").style("top",n.y+"px");else{if(this.lastTargetId=t.id,this.use_fill){var s=t.getAttribute("tooltip_fill");s||(s=t.getAttribute("fill")),s&&o.style("background-color",s)}this.use_stroke&&o.style("border-color",t.getAttribute("stroke")),o.html(this.decodeContent(t.getAttribute("title"))),n=this.calculatePosition(o,t,i),o.style("left",n.x+"px").style("top",n.y+"px").transition().duration(this.delay_over).style("opacity",this.opacity)}return!0}}catch(t){console.error(t)}return!1}},{key:"decodeContent",value:function(t){return this.decodingTextarea.innerHTML=t,this.decodingTextarea.value}},{key:"calculatePosition",value:function(t,e,n){var r,i,o=e.ownerSVGElement,s=t.node(),a=o.parentNode;if(this.use_cursor_pos){var u=Math.ceil(s.offsetWidth),c=Math.ceil(s.offsetHeight),f=h(o),l=new DOMPoint(f.left,f.top),p=new DOMPoint(f.right,f.bottom);r=n,"doc"!=this.placement&&(i=d(a).inverse(),r=r.matrixTransform(i),l=l.matrixTransform(i),p=p.matrixTransform(i));var y=e.getBoundingClientRect(),v=[],g=p.x-(r.x+this.offx),m=r.x-this.offx-l.x;g>=u&&v.push(Math.max(l.x,r.x+this.offx)),m>=u&&v.push(Math.min(p.x-u,r.x-this.offx-u)),v.push(Math.max(l.x,Math.min(p.x,r.x+u/2)-u));var x=[],b=p.y-(r.y+this.offy),w=r.y-this.offy-l.y;b>=c&&x.push(Math.max(l.y,r.y+this.offy)),w>=c&&x.push(Math.min(p.y-c,r.y-this.offy-c)),x.push(Math.max(l.y,Math.min(p.y,r.y+c/2)-c));var S=[];v.forEach((function(t){x.forEach((function(e){S.push(new DOMRect(t,e,u,c))}))}));var A=S.find((function(t){return n=y,!((e=t).x<=n.x&&e.x+e.width>=n.x+n.width&&e.y<=n.y&&e.y+e.height>=n.y+n.height);var e,n}));A||(A=S[0]),r.x=A.x,r.y=A.y}else r=new DOMPoint(this.offx,this.offy),"doc"==this.placement&&(i=d(a),r=r.matrixTransform(i));return r}}])&&F(e.prototype,n),i&&F(e,i),Object.defineProperty(e,"prototype",{writable:!1}),t}();n(163);function L(t,e){for(var n=0;n<e.length;n++){var r=e[n];r.enumerable=r.enumerable||!1,r.configurable=!0,"value"in r&&(r.writable=!0),Object.defineProperty(t,r.key,r)}}var G=function(){function t(e,n){!function(t,e){if(!(t instanceof e))throw new TypeError("Cannot call a class as a function")}(this,t),this.svgid=e,this.clsName=n.classPrefix+"_"+e,this.invClsName=n.invClassPrefix?n.invClassPrefix+"_"+e:null,this.attrName=n.attrName,this.shinyInputId=n.shinyInputId,this.shinyMessageId=n.shinyMessageId,this.nodeIds=[],this.dataHovered=[],this.lastTargetId=null}var e,n,i;return e=t,(n=[{key:"init",value:function(){var t=document.getElementById(this.svgid+"_rootg").querySelectorAll("*["+this.attrName+"]");if(!t.length)return!1;this.nodeIds=Array(t.length);var e=0;t.forEach((function(t){this.nodeIds[e++]=t.id}),this);var n=this;return this.shinyMessageId&&Shiny.addCustomMessageHandler(this.shinyMessageId,(function(t){"string"==typeof t?n.setHovered([t]):Array.isArray(t)&&n.setHovered(t)})),!0}},{key:"destroy",value:function(){if(this.lastTargetId=null,this.nodeIds=[],this.shinyMessageId)try{Shiny.addCustomMessageHandler(this.shinyMessageId,(function(t){}))}catch(t){console.error(t)}this.dataHovered=[]}},{key:"clear",value:function(){this.lastTargetId&&(this.lastTargetId=null,this.setHovered([]))}},{key:"isValidTarget",value:function(t){return t instanceof SVGGraphicsElement&&!(t instanceof SVGSVGElement)&&t.hasAttribute(this.attrName)}},{key:"applyOn",value:function(t,e){try{if(this.isValidTarget(t))return t.id!==this.lastTargetId&&(this.lastTargetId=t.id,this.setHovered([t.getAttribute(this.attrName)])),!0}catch(t){console.error(t)}return!1}},{key:"setHovered",value:function(t){this.dataHovered.length===t.length&&this.dataHovered.every((function(e){return t.includes(e)}))||(this.dataHovered=t,this.refreshHovered(),this.shinyInputId&&Shiny.onInputChange(this.shinyInputId,this.dataHovered))}},{key:"refreshHovered",value:function(){var t,e,n;this.nodeIds.forEach((function(i){(t=document.getElementById(i))&&(e=this.dataHovered.includes(t.getAttribute(this.attrName)),(n=r.select(t)).classed(this.clsName,e),this.invClsName&&(this.dataHovered.length?n.classed(this.invClsName,!e):n.classed(this.invClsName,!1)))}),this)}}])&&L(e.prototype,n),i&&L(e,i),Object.defineProperty(e,"prototype",{writable:!1}),t}();n(111);function U(t,e){for(var n=0;n<e.length;n++){var r=e[n];r.enumerable=r.enumerable||!1,r.configurable=!0,"value"in r&&(r.writable=!0),Object.defineProperty(t,r.key,r)}}var H=function(){function t(e,n){!function(t,e){if(!(t instanceof e))throw new TypeError("Cannot call a class as a function")}(this,t),this.svgid=e,this.clsName=n.classPrefix+"_"+e,this.invClsName=n.invClassPrefix?n.invClassPrefix+"_"+e:null,this.attrName=n.attrName,this.shinyInputId=n.shinyInputId,this.shinyMessageId=n.shinyMessageId,this.type=n.type,this.initialSelection=n.selected,this.nodeIds=[],this.dataSelected=[]}var e,n,i;return e=t,(n=[{key:"init",value:function(){if("single"!==this.type&&"multiple"!==this.type)return!1;var t=document.getElementById(this.svgid+"_rootg").querySelectorAll("*["+this.attrName+"]");if(!t.length)return!1;var e=this;this.nodeIds=Array(t.length);var n=0;return t.forEach((function(t){this.nodeIds[n++]=t.id}),this),this.shinyMessageId&&Shiny.addCustomMessageHandler(this.shinyMessageId,(function(t){"string"==typeof t?e.setSelected([t]):Array.isArray(t)&&e.setSelected(t)})),"string"==typeof this.initialSelection?this.setSelected([this.initialSelection]):Array.isArray(this.initialSelection)&&"multiple"==this.type&&this.setSelected(this.initialSelection),this.initialSelection=null,!0}},{key:"destroy",value:function(){if(this.nodeIds=[],this.dataSelected=[],this.shinyMessageId)try{Shiny.addCustomMessageHandler(this.shinyMessageId,(function(t){}))}catch(t){console.error(t)}}},{key:"getButtons",value:function(){var t=this;return[{key:"lasso_select",block:"selection",class:"neutral",tooltip:"lasso selection",icon:"<svg xmlns='http://www.w3.org/2000/svg' role='img' viewBox='0 0 230 230'><g><ellipse ry='65.5' rx='86.5' cy='94' cx='115.5' stroke-width='20' fill='transparent'/><ellipse ry='11.500001' rx='10.5' cy='153' cx='91.5' stroke-width='20' fill='transparent'/><line y2='210.5' x2='105' y1='164.5' x1='96' stroke-width='20'/></g></svg>",onclick:function(){t.lasso(!0)}},{key:"lasso_deselect",block:"selection",class:"drop",tooltip:"lasso deselection",icon:"<svg xmlns='http://www.w3.org/2000/svg' role='img' viewBox='0 0 230 230'><g><ellipse ry='65.5' rx='86.5' cy='94' cx='115.5' stroke-width='20' fill='transparent'/><ellipse ry='11.500001' rx='10.5' cy='153' cx='91.5' stroke-width='20' fill='transparent'/><line y2='210.5' x2='105' y1='164.5' x1='96' stroke-width='20'/></g></svg>",onclick:function(){t.lasso(!1)}}]}},{key:"clear",value:function(){this.setSelected([])}},{key:"isValidTarget",value:function(t){return t instanceof SVGGraphicsElement&&!(t instanceof SVGSVGElement)&&t.hasAttribute(this.attrName)}},{key:"applyOn",value:function(t,e){try{if(this.isValidTarget(t)){var n=t.getAttribute(this.attrName),r=Array.from(this.dataSelected),i=r.indexOf(n);return"multiple"==this.type?i<0?r.push(n):r.splice(i,1):r=i<0?[n]:[],this.setSelected(r),!0}}catch(t){console.error(t)}return!1}},{key:"setSelected",value:function(t){this.dataSelected.length===t.length&&this.dataSelected.every((function(e){return t.includes(e)}))||(this.dataSelected=t,this.refreshSelected(),this.shinyInputId&&Shiny.onInputChange(this.shinyInputId,this.dataSelected))}},{key:"refreshSelected",value:function(){var t,e,n;this.nodeIds.forEach((function(i){(t=document.getElementById(i))&&(e=this.dataSelected.includes(t.getAttribute(this.attrName)),(n=r.select(t)).classed(this.clsName,e),this.invClsName&&(this.dataSelected.length?n.classed(this.invClsName,!e):n.classed(this.invClsName,!1)))}),this)}},{key:"lasso",value:function(t){var e=r.select("#"+this.svgid+"_rootg"),n=this,i=r.lasso();try{i=i.closePathSelect(!0).closePathDistance(100).items(e.selectAll("*["+this.attrName+"]")).targetArea(e).on("start",(function(){e.style("cursor","crosshair")})).on("draw",(function(){})).on("end",(function(){try{e.style("cursor","auto"),e.on(".dragstart",null).on(".drag",null).on(".dragend",null),e.selectAll("g.lasso").remove();var r=Array.from(n.dataSelected);i.selectedItems().each((function(e,i){var o=this.getAttribute(n.attrName),s=r.indexOf(o);s<0&&t?r.push(o):s>=0&&!t&&r.splice(s,1)})),n.setSelected(r)}catch(t){console.error(t)}})),e.call(i)}catch(t){console.error(t)}}}])&&U(e.prototype,n),i&&U(e,i),Object.defineProperty(e,"prototype",{writable:!1}),t}();function W(t,e){for(var n=0;n<e.length;n++){var r=e[n];r.enumerable=r.enumerable||!1,r.configurable=!0,"value"in r&&(r.writable=!0),Object.defineProperty(t,r.key,r)}}var Y=["mouseover","mouseout","mousemove","mousedown","wheel","click","nearest"],$=function(){function t(e,n,r){!function(t,e){if(!(t instanceof e))throw new TypeError("Cannot call a class as a function")}(this,t),this.svgid=e,this.handlers=n,this.nearestHandler=r}var e,n,r;return e=t,(n=[{key:"init",value:function(){var t=this;if(this.event_types=Y.filter((function(t){return"nearest"!==t})).filter((function(e){return t.handlers.get(e)&&t.handlers.get(e).length})),!this.event_types.length)return!1;this.handlers.get("nearest")&&this.handlers.get("nearest").length||(this.nearestHandler=null);var e,n=document.getElementById(this.svgid);return this.event_types.forEach((function(t){e="mousemove"!==t,n.addEventListener(t,this,e)}),this),!0}},{key:"destroy",value:function(){try{var t=document.getElementById(this.svgid);this.event_types.forEach((function(e){t.removeEventListener(e,this)}),this)}catch(t){console.error(t)}}},{key:"handleEvent",value:function(t){try{var e=t.target,n=!1,r=null;"mouseout"===t.type?this.handlers.get(t.type).forEach((function(e){return e.clear(t)})):"mouseover"!==t.type||t.buttons?"mousemove"!==t.type||t.buttons?"mousedown"===t.type||"wheel"===t.type?this.handlers.get(t.type).forEach((function(e){return e.clear(t)})):"click"===t.type&&this.svgid!==e.id&&this.handlers.get(t.type).map((function(n){return n.applyOn(e,t)})):(this.svgid!==e.id&&(n=this.handlers.get(t.type).map((function(n){return n.applyOn(e,t)})).find((function(t){return!!t}))),this.nearestHandler&&!n&&(t.fromNearest=!0,(r=this.nearestHandler.applyOn(e,t))?this.handlers.get("nearest").forEach((function(e){return e.applyOn(r,t)})):this.handlers.get("nearest").forEach((function(e){return e.clear(t)})))):this.handlers.get(t.type).forEach((function(n){return n.applyOn(e,t)}))}catch(t){console.error(t)}}}])&&W(e.prototype,n),r&&W(e,r),Object.defineProperty(e,"prototype",{writable:!1}),t}(),q=(n(240),n(164),n(165)),K=n.n(q);function X(t,e){for(var n=0;n<e.length;n++){var r=e[n];r.enumerable=r.enumerable||!1,r.configurable=!0,"value"in r&&(r.writable=!0),Object.defineProperty(t,r.key,r)}}var J=function(){function t(e,n,r){!function(t,e){if(!(t instanceof e))throw new TypeError("Cannot call a class as a function")}(this,t),this.svgid=e,this.attrNames=n,this.maximum_distance=r,this.spatialIndex=null,this.nodeIds=null,this.debug=!1}var e,n,i;return e=t,(n=[{key:"init",value:function(){if(0===this.maximum_distance)return!1;this.maximum_distance||(this.maximum_distance=1/0);var t=document.getElementById(this.svgid+"_rootg"),e=this.attrNames.map((function(t){return"*[nearest]["+t+"]"})).join(", "),n=Array.from(t.querySelectorAll(e)).filter((function(t){return t instanceof SVGCircleElement||t instanceof SVGImageElement||t instanceof SVGLineElement||t instanceof SVGPathElement||t instanceof SVGPolygonElement||t instanceof SVGPolylineElement||t instanceof SVGRectElement||t instanceof SVGTextElement}));if(!n.length)return!1;this.spatialIndex=new K.a(n.length);var r,i,o,s,a,u=t.ownerSVGElement,c=t.getCTM().inverse(),f=new Set,l=0;return this.nodeIds=Array(n.length),n.forEach((function(e){this.nodeIds[l++]=e.id,r=e.getCTM(),i=e.getBBox(),o=g(i).map((function(t){return t.matrixTransform(r).matrixTransform(c)})),s=m(o),this.spatialIndex.add(s.minx,s.miny,s.maxx,s.maxy),this.debug&&w(x(s,u),u,e.id+"_extent"),e instanceof SVGRectElement||e instanceof SVGImageElement||e instanceof SVGTextElement?(e.gpoints=o,e.gpoints.shape="polygon"):(e.gpoints=g(e).map((function(t){return t.matrixTransform(r).matrixTransform(c)})),e instanceof SVGCircleElement?(e.gpoints[0].r=e.r.baseVal.value,e.gpoints.shape="circle"):(e instanceof SVGLineElement||e instanceof SVGPolylineElement)&&(e.gpoints.shape="polyline")),this.debug&&w(e.gpoints,u,e.id+"_points"),e.grect=x(s,u),(a=e.parentElement.getAttribute("clip-path"))?f.add(a.replace(/url\(#|\)/g,"")):f.add(t.id)}),this),this.spatialIndex.finish(),this.nodeIds=n.map((function(t){return t.id})),i=t.getBBox(),r=t.getCTM(),t.gpoints=g(i).map((function(t){return t.matrixTransform(r).matrixTransform(c)})),this.debug&&w(t.gpoints,u,t.id+"_rootpoints","none","#00ff00"),0==f.size&&f.add(t.id),f.has(t.id)?this.clips=null:(this.clips=Array.from(f.values()).map((function(t){return document.getElementById(t)})).map((function(t){return Array.from(t.childNodes).filter((function(t){return t instanceof SVGGraphicsElement})).map((function(t){if(r=t.getCTM(),0===(i=t.getBBox()).width||0===i.height){if(!(t instanceof SVGRectElement))return null;i.x=t.x.baseVal.value,i.y=t.y.baseVal.value,i.width=t.width.baseVal.value,i.height=t.height.baseVal.value}return x(m(o=g(i).map((function(t){return t.matrixTransform(r).matrixTransform(c)}))),u)})).flat().filter((function(t){return!!t}))})).flat(),this.clips.length||(this.clips=null)),!0}},{key:"destroy",value:function(){this.spatialIndex=null,this.nodeIds=null,this.clips=null}},{key:"isValidTarget",value:function(t){return t instanceof SVGGraphicsElement}},{key:"applyOn",value:function(t,e){var n=this;try{if(!this.isValidTarget(t))return null;var i=new DOMPoint(e.clientX,e.clientY),o=document.getElementById(this.svgid),s=document.getElementById(this.svgid+"_rootg");if(o===t&&(t=s),i=i.matrixTransform(t.getScreenCTM().inverse()),this.debug&&w([i],o,o.id+"_mouse"),this.clips&&!this.clips.find((function(t){return v(t,[i])})))return null;var a,u,c=this.spatialIndex.neighbors(i.x,i.y,10,this.maximum_distance).filter((function(t,e,n){return n.indexOf(t)===e})).map((function(t){return n.nodeIds[t]})).sort().map((function(t){return document.getElementById(t)}));if(c.length){var f=r.zoomTransform(s);f=new DOMMatrix([f.k,0,0,f.k,f.x,f.y]).inverse(),u=s.gpoints.map((function(t){return t.matrixTransform(f)})),a=b(u,o),this.debug&&w(a,o,s.id+"_view","none","#00aa00"),c=c.filter((function(t){return e=a,n=t.grect,e.x<n.x+n.width&&e.x+e.width>n.x&&e.y<n.y+n.height&&e.y+e.height>n.y;var e,n}))}if(c.length&&o.checkIntersection){var l=s.getCTM();u=u.map((function(t){return t.matrixTransform(l)})),a=b(u,o),c=c.filter((function(t){return o.checkIntersection(t,a)}))}else c.length&&(c=c.filter((function(t){return v(a,t.gpoints)})));var h=null;if(1===c.length)h=c[0];else if(c.length){var d=1/0,g=1/0;c.forEach((function(t){var e,n;t instanceof SVGCircleElement?(e=i,n=t.gpoints[0],d=p(e,n)-n.r):d=function(t,e){var n,r,i=1/0;if(e.length>1)for(var o=0;o<e.length;o++)(r=o+1)==e.length&&(r=0),(n=y(t,e[o],e[r]))<=i&&(i=n);return i}(i,t.gpoints),d<=g&&d<=this.maximum_distance&&(g=d,h=t)}),this)}return h}catch(t){console.error(t)}return null}}])&&X(e.prototype,n),i&&X(e,i),Object.defineProperty(e,"prototype",{writable:!1}),t}();n(242),n(243),n(244);function Z(t,e){for(var n=0;n<e.length;n++){var r=e[n];r.enumerable=r.enumerable||!1,r.configurable=!0,"value"in r&&(r.writable=!0),Object.defineProperty(t,r.key,r)}}var Q=function(){function t(e,n){!function(t,e){if(!(t instanceof e))throw new TypeError("Cannot call a class as a function")}(this,t),this.svgid=e,this.pngname=n.pngname+".png"}var e,n,r;return e=t,(n=[{key:"init",value:function(){return"function"==typeof saveSvgAsPng&&"undefined"!=typeof Promise&&-1!==Promise.toString().indexOf("[native code]")}},{key:"destroy",value:function(){this.fonts=null}},{key:"getButtons",value:function(){var t=this;return[{key:"saveaspng",block:"misc",class:"neutral",tooltip:"download png",icon:"<svg xmlns='http://www.w3.org/2000/svg' role='img' viewBox='0 0 512 512'><g><polygon points='95 275 95 415 415 415 415 275 375 275 375 380 135 380 135 275'/><polygon points='220 30 220 250 150 175 150 245 250 345 350 245 350 175 280 250 280 30'/></g></svg>",onclick:function(){t.save_as_png()}}]}},{key:"save_as_png",value:function(){this.fonts||(this.fonts=function(){var t=["woff","ttf","otf","woff2","eot"],e={woff2:"font/woff2",woff:"font/woff",otf:"application/x-font-opentype",ttf:"application/x-font-ttf",eot:"application/vnd.ms-fontobject"},n=/(^.+src:\s*)(.+?)(;.*)$/,r=/url\(["']?(.+?)["']?\)(\s*format\(.+?\))?/g,i=new RegExp("url\\([\"\\']?(.+?\\.("+Object.keys(e).join("|")+")(\\?.*?)?)[\"\\']?\\)");function o(o){var s=o.cssText.match(n);if(s){var a=s[1],u=s[2],c=s[3],f=u.match(r);if(Array.isArray(f))return f.map((function(t){return function(t,e){var n=t.match(i),r=n&&n[1]||"";if(r&&!r.match(/^data:/)&&"about:blank"!==r)return r.startsWith("../")?r=e+"/../"+r:r.startsWith("./")&&(r=e+"/."+r),{fmt:n[2],url:r,css:t}}(t,o.href)})).filter((function(t){return!!t})).map((function(e){return e.order=t.indexOf(e.fmt),e})).sort((function(t,e){return t.order-e.order})).slice(0,1).map((function(t){return{text:a+t.css+c,format:e[t.fmt],url:t.url}})).pop()}}return Array.from(document.styleSheets).map((function(t){try{return{cssRules:t.cssRules,href:t.href}}catch(e){console.warn("Stylesheet could not be loaded: "+t.href,e)}})).map((function(t){return t&&t.cssRules?Array.from(t.cssRules).filter((function(t){return t.cssText.match(/^@font-face/)})).map((function(e){return{href:t.href,cssText:e.cssText}})):[]})).flat().map((function(t){return o(t)})).filter((function(t){return!!t}))}()),saveSvgAsPng(document.getElementById(this.svgid),this.pngname,{encoderOptions:1,fonts:this.fonts})}}])&&Z(e.prototype,n),r&&Z(e,r),Object.defineProperty(e,"prototype",{writable:!1}),t}();function tt(t,e){for(var n=0;n<e.length;n++){var r=e[n];r.enumerable=r.enumerable||!1,r.configurable=!0,"value"in r&&(r.writable=!0),Object.defineProperty(t,r.key,r)}}var et=function(){function t(e,n){!function(t,e){if(!(t instanceof e))throw new TypeError("Cannot call a class as a function")}(this,t),this.containerid=e,this.svgid=null,this.handlers=new Map,this.shinyMode=n}var e,n,o;return e=t,(n=[{key:"clear",value:function(){this.handlers.forEach((function(t){return t.destroy()})),this.handlers.clear(),r.select("#"+this.containerid+" style").remove(),r.select("#"+this.containerid+" div.girafe_container_std").remove()}},{key:"setSvgId",value:function(t){this.svgid=t}},{key:"addStyle",value:function(t){var e=t.join("\n").replace(/SVGID_/g,this.svgid);r.select("#"+this.containerid).append("style").attr("type","text/css").text(e)}},{key:"addSvg",value:function(t,e){r.select("#"+this.containerid).append("div").attr("class","girafe_container_std").html(t),e&&function(t){var e=t.indexOf("{"),n=t.substring(e+1,t.lastIndexOf("}")),r=t.substring(0,e),i=r.substring(r.indexOf("(")+1,r.lastIndexOf(")")).split(",");function o(){return Function.apply(this,i)}return i.push(n),o.prototype=Function.prototype,new o}(e)()}},{key:"IEFixResize",value:function(t,e){i()&&(r.select("#"+this.svgid).classed("girafe_svg_ie",!0),r.select("#"+this.containerid+" div").classed("girafe_container_ie",!0).style("width",Math.round(100*t)+"%").style("padding-bottom",Math.round(t*e*100)+"%"))}},{key:"autoScale",value:function(t){r.select("#"+this.svgid).style("width",t).style("height","100%").style("margin-left","auto").style("margin-right","auto")}},{key:"fixSize",value:function(t,e){r.select("#"+this.svgid).attr("preserveAspectRatio","xMidYMin meet"),r.select("#"+this.containerid+" .girafe_container_std").style("width","100%"),r.select("#"+this.svgid).attr("width",t).attr("height",e),r.select("#"+this.svgid).style("width",t+"px").style("height",e+"px")}},{key:"setSizeLimits",value:function(t,e,n,i){r.select("#"+this.svgid).style("max-width",t).style("max-height",n).style("min-width",e).style("min-height",i)}},{key:"removeContainerLimits",value:function(){i()||r.select("#"+this.containerid).style("width",null).style("height",null)}},{key:"setupTooltip",value:function(t){try{t.classPrefix="tooltip";var e=new B(this.svgid,t);e.init()&&this.handlers.set("tooltip",e)}catch(t){console.error(t)}}},{key:"setupHover",value:function(t,e,n,r){t.classPrefix="hover_data",t.attrName="data-id",t.shinyInputSuffix="_hovered",t.shinyMessageSuffix="_hovered_set",t.invClassPrefix=e.css.length>0?"hover_inv":null,n.classPrefix="hover_key",n.attrName="key-id",n.shinyInputSuffix="_key_hovered",n.shinyMessageSuffix="_key_hovered_set",r.classPrefix="hover_theme",r.attrName="theme-id",r.shinyInputSuffix="_theme_hovered",r.shinyMessageSuffix="_theme_hovered_set";var i,o=[t,n,r];try{o.forEach((function(t){t.shinyInputId=this.shinyMode&&t.reactive?this.containerid+t.shinyInputSuffix:null,t.shinyMessageId=this.shinyMode&&t.reactive?this.containerid+t.shinyMessageSuffix:null,(i=new G(this.svgid,t)).init()&&this.handlers.set(t.classPrefix,i)}),this)}catch(t){console.error(t)}try{(i=new J(this.svgid,[t.attrName],t.nearest_distance)).init()&&this.handlers.set("nearest",i)}catch(t){console.error(t)}}},{key:"setupSelection",value:function(t,e,n,r){t.classPrefix="select_data",t.attrName="data-id",t.shinyInputSuffix="_selected",t.shinyMessageSuffix="_set",t.invClassPrefix=e.css.length>0?"select_inv":null,n.classPrefix="select_key",n.attrName="key-id",n.shinyInputSuffix="_key_selected",n.shinyMessageSuffix="_key_set",r.classPrefix="select_theme",r.attrName="theme-id",r.shinyInputSuffix="_theme_selected",r.shinyMessageSuffix="_theme_set";var i,o=[t,n,r];try{o.forEach((function(t){!this.shinyMode&&t.only_shiny||(t.shinyInputId=this.shinyMode?this.containerid+t.shinyInputSuffix:null,t.shinyMessageId=this.shinyMode?this.containerid+t.shinyMessageSuffix:null,(i=new H(this.svgid,t)).init()&&this.handlers.set(t.classPrefix,i))}),this)}catch(t){console.error(t)}}},{key:"setupMouse",value:function(){try{var t=new Map;Y.forEach((function(e){return t.set(e,[])})),this.handlers.forEach((function(e){e instanceof B?(t.get("mouseover").push(e),t.get("mouseout").push(e),t.get("mousemove").push(e),t.get("mousedown").push(e),t.get("wheel").push(e),t.get("nearest").push(e)):e instanceof G?(t.get("mouseover").push(e),t.get("mouseout").push(e),t.get("nearest").push(e)):e instanceof H?t.get("click").push(e):e instanceof s&&(t.get("mouseover").push(e),t.get("mouseout").push(e))}));var e=new $(this.svgid,t,this.handlers.get("nearest"));e.init()&&this.handlers.set("mouse",e)}catch(t){console.error(t)}}},{key:"setupZoom",value:function(t){try{var e=new u(this.svgid,t);e.init()&&this.handlers.set("zoom",e)}catch(t){console.error(t)}}},{key:"setupToolbar",value:function(t){var e;try{t.hidden.includes("saveaspng")||(e=new Q(this.svgid,{pngname:t.pngname})).init()&&this.handlers.set("png",e)}catch(t){console.error(t)}try{var n=[];this.handlers.forEach((function(t){(t instanceof Q||t instanceof u||t instanceof H&&"data-id"==t.attrName&&"multiple"==t.type)&&n.push(t)})),t.clsName="ggiraph-toolbar",(e=new s(this.containerid,this.svgid,t)).init(n)&&this.handlers.set("toolbar",e)}catch(t){console.error(t)}}}])&&tt(e.prototype,n),o&&tt(e,o),Object.defineProperty(e,"prototype",{writable:!1}),t}();function nt(t){return function(e,n,r){var i=new et(e.id,t);return{renderValue:function(n){i.clear(),i.setSvgId(n.uid),i.addStyle([n.settings.tooltip.css,n.settings.select_inv.css,n.settings.select.css,n.settings.select_key.css,n.settings.select_theme.css,n.settings.hover_inv.css,n.settings.hover.css,n.settings.hover_key.css,n.settings.hover_theme.css]),i.addSvg(n.html,n.js);var r=d3.select("#"+i.svgid).property("viewBox").baseVal;n.settings.sizing.rescale?t?(i.autoScale("100%"),i.IEFixResize(1,1/n.ratio),i.setSizeLimits(d3.select(e).style("width"),0,d3.select(e).style("height"),0)):(i.autoScale(Math.round(100*n.settings.sizing.width)+"%"),i.IEFixResize(n.settings.sizing.width,1/n.ratio),i.setSizeLimits("unset","unset","unset","unset"),i.removeContainerLimits()):(i.fixSize(r.width,r.height),d3.select(e).style("width",null).style("height",null)),i.setupHover(n.settings.hover,n.settings.hover_inv,n.settings.hover_key,n.settings.hover_theme),i.setupSelection(n.settings.select,n.settings.select_inv,n.settings.select_key,n.settings.select_theme),i.setupZoom(n.settings.zoom),i.setupToolbar(n.settings.toolbar),i.setupTooltip(n.settings.tooltip),i.setupMouse()},resize:function(t,e){}}}}}])}));
```
