## Supplementary material for "Evaluating Regional Diversity in Scientific Communication: A Comparative Analysis of COVID-19 Preprints and Peer-Reviewed Publications": save-svg-as-png.min.js

```
"use strict";!function(){function i(e){if(!r(e))throw new Error("an HTMLElement or SVGElement is required; got "+e)}function o(n){return new Promise(function(e,t){r(n)?e(n):t(new Error("an HTMLElement or SVGElement is required; got "+n))})}function a(e){for(var t=window.atob(e.split(",")[1]),e=e.split(",")[0].split(":")[1].split(";")[0],n=new ArrayBuffer(t.length),r=new Uint8Array(n),o=0;o<t.length;o++)r[o]=t.charCodeAt(o);return new Blob([n],{type:e})}function s(){if(!(navigator.msSaveOrOpenBlob||"download"in document.createElement("a")))return{popup:window.open()}}var c="undefined"!=typeof exports&&exports||"undefined"!=typeof define&&{}||this||window,C=("undefined"!=typeof define&&define("save-svg-as-png",[],function(){return c}),c.default=c,"http://www.w3.org/2000/xmlns/"),T="http://www.w3.org/2000/svg",u=/url\(["']?(.+?)["']?\)/,n={woff2:"font/woff2",woff:"font/woff",otf:"application/x-font-opentype",ttf:"application/x-font-ttf",eot:"application/vnd.ms-fontobject",sfnt:"application/font-sfnt",svg:"image/svg+xml"},r=function(e){return e instanceof HTMLElement||e instanceof SVGElement},l=function(e){return e&&0===e.lastIndexOf("http",0)&&-1===e.lastIndexOf(window.location.host)},d=function(t){var e=Object.keys(n).filter(function(e){return 0<t.indexOf("."+e)}).map(function(e){return n[e]});return e?e[0]:(console.error("Unknown font format for "+t+". Fonts may not be working correctly."),"application/octet-stream")},f=function(e){for(var t="",n=new Uint8Array(e),r=0;r<n.byteLength;r++)t+=String.fromCharCode(n[r]);return window.btoa(t)},U=function(e,t,n){t=e.viewBox&&e.viewBox.baseVal&&e.viewBox.baseVal[n]||null!==t.getAttribute(n)&&!t.getAttribute(n).match(/%$/)&&parseInt(t.getAttribute(n))||e.getBoundingClientRect()[n]||parseInt(t.style[n])||parseInt(window.getComputedStyle(e).getPropertyValue(n));return null==t||isNaN(parseFloat(t))?0:t},k=function(e,t){if(t)try{return e.querySelector(t)||e.parentNode&&e.parentNode.querySelector(t)}catch(e){console.warn('Invalid CSS selector "'+t+'"',e)}},L=function(e,t){var n=e.cssText.match(u),n=n&&n[1]||"";if(n&&!n.match(/^data:/)&&"about:blank"!==n)return t=n.startsWith("../")?t+"/../"+n:n.startsWith("./")?t+"/."+n:n,{text:e.cssText,format:d(t),url:t}},h={},O=function(e){return Promise.all(e.map(function(r){return new Promise(function(t,e){if(h[r.url])return t(h[r.url]);var n=new XMLHttpRequest;n.addEventListener("load",function(){var e=f(n.response),e=r.text.replace(u,'url("data:'+r.format+";base64,"+e+'")')+"\n";h[r.url]=e,t(e)}),n.addEventListener("error",function(e){console.warn("Failed to load font from: "+r.url,e),h[r.url]=null,t(null)}),n.addEventListener("abort",function(e){console.warn("Aborted loading font from: "+r.url,e),t(null)}),n.open("GET",r.url),n.responseType="arraybuffer",n.send()})})).then(function(e){return e.filter(function(e){return e}).join("")})},e=null,B=function(){return e=e||Array.from(document.styleSheets).map(function(t){try{return{rules:t.cssRules,href:t.href}}catch(e){return console.warn("Stylesheet could not be loaded: "+t.href,e),{}}})};c.prepareSvg=function(g,p,m){i(g);var e=p||{},t=e.left,v=void 0===t?0:t,t=e.top,b=void 0===t?0:t,y=e.width,x=e.height,t=e.scale,A=void 0===t?1:t,t=e.responsive,S=void 0!==t&&t,t=e.excludeCss,E=void 0!==t&&t;return Promise.all(Array.from(g.querySelectorAll("image")).map(function(o){var i=o.getAttributeNS("http://www.w3.org/1999/xlink","href")||o.getAttribute("href");return i?(l(i)&&(i+=(-1===i.indexOf("?")?"?":"&")+"t="+(new Date).valueOf()),new Promise(function(e,t){var n=document.createElement("canvas"),r=new Image;r.crossOrigin="anonymous",r.src=i,r.onerror=function(){return t(new Error("Could not load "+i))},r.onload=function(){n.width=r.width,n.height=r.height,n.getContext("2d").drawImage(r,0,0),o.setAttributeNS("http://www.w3.org/1999/xlink","href",n.toDataURL("image/png")),e(!0)}})):Promise.resolve(null)})).then(function(){var e,r,n,o,t,i,a,s,u,l,c=g.cloneNode(!0),d=(c.style.backgroundColor=(p||{}).backgroundColor||g.style.backgroundColor,t=c,e=y,w=x,"svg"===(d=g).tagName?{width:e||U(d,t,"width"),height:w||U(d,t,"height")}:d.getBBox?(w=(e=d.getBBox()).x,t=e.y,{width:w+e.width,height:t+e.height}):void 0),f=d.width,h=d.height;if("svg"!==g.tagName){if(!g.getBBox)return void console.error("Attempted to render non-SVG element",g);null!=c.getAttribute("transform")&&c.setAttribute("transform",c.getAttribute("transform").replace(/translate\(.*?\)/,""));var w=document.createElementNS("http://www.w3.org/2000/svg","svg");w.appendChild(c),c=w}return c.setAttribute("version","1.1"),c.setAttribute("viewBox",[v,b,f,h].join(" ")),c.getAttribute("xmlns")||c.setAttributeNS(C,"xmlns",T),c.getAttribute("xmlns:xlink")||c.setAttributeNS(C,"xmlns:xlink","http://www.w3.org/1999/xlink"),S?(c.removeAttribute("width"),c.removeAttribute("height"),c.setAttribute("preserveAspectRatio","xMinYMin meet")):(c.setAttribute("width",f*A),c.setAttribute("height",h*A)),Array.from(c.querySelectorAll("foreignObject > *")).forEach(function(e){e.setAttributeNS(C,"xmlns","svg"===e.tagName?T:"http://www.w3.org/1999/xhtml")}),E?((t=document.createElement("div")).appendChild(c),e=t.innerHTML,"function"!=typeof m?{src:e,width:f,height:h}:void m(e,f,h)):(r=g,n=(d=(d=p)||{}).selectorRemap,o=d.modifyStyle,w=d.modifyCss,t=d.fonts,i=d.excludeUnusedCss,a=w||function(e,t){return(n?n(e):e)+"{"+(o?o(t):t)+"}\n"},s=[],u=void 0===t,l=t||[],B().forEach(function(e){var t=e.rules,n=e.href;t&&Array.from(t).forEach(function(e){var t;void 0!==e.style&&(k(r,e.selectorText)?s.push(a(e.selectorText,e.style.cssText)):u&&e.cssText.match(/^@font-face/)?(t=L(e,n))&&l.push(t):i||s.push(e.cssText))})}),O(l).then(function(e){return s.join("\n")+e}).then(function(e){var t=document.createElement("style"),e=(t.setAttribute("type","text/css"),t.innerHTML="<![CDATA[\n"+e+"\n]]>",document.createElement("defs")),t=(e.appendChild(t),c.insertBefore(e,c.firstChild),document.createElement("div")),e=(t.appendChild(c),t.innerHTML.replace(/NS\d+:href/gi,'xmlns:xlink="http://www.w3.org/1999/xlink" xlink:href'));if("function"!=typeof m)return{src:e,width:f,height:h};m(e,f,h)}))})},c.svgAsDataUri=function(e,t,r){return i(e),c.prepareSvg(e,t).then(function(e){var t=e.src,n=e.width,e=e.height,t="data:image/svg+xml;base64,"+window.btoa(decodeURIComponent(encodeURIComponent('<?xml version="1.0" standalone="no"?><!DOCTYPE svg PUBLIC "-//W3C//DTD SVG 1.1//EN" "http://www.w3.org/Graphics/SVG/1.1/DTD/svg11.dtd" [<!ENTITY nbsp "&#160;">]>'+t).replace(/%([0-9A-F]{2})/g,function(e,t){t=String.fromCharCode("0x"+t);return"%"===t?"%25":t})));return"function"==typeof r&&r(t,n,e),t})},c.svgAsPngUri=function(e,t,a){i(e);function o(e){var t=e.src,n=e.width,e=e.height,r=document.createElement("canvas"),o=r.getContext("2d"),i=window.devicePixelRatio||1,n=(r.width=n*i,r.height=e*i,r.style.width=r.width+"px",r.style.height=r.height+"px",o.setTransform(i,0,0,i,0,0),l?l(r,t):o.drawImage(t,0,0),void 0);try{n=r.toDataURL(s,u)}catch(e){if("undefined"!=typeof SecurityError&&e instanceof SecurityError||"SecurityError"===e.name)return void console.error("Rendered SVG images cannot be downloaded in this browser.");throw e}return"function"==typeof a&&a(n,r.width,r.height),Promise.resolve(n)}var n=t||{},r=n.encoderType,s=void 0===r?"image/png":r,r=n.encoderOptions,u=void 0===r?.8:r,l=n.canvg;return l?c.prepareSvg(e,t).then(o):c.svgAsDataUri(e,t).then(function(r){return new Promise(function(e,t){var n=new Image;n.onload=function(){return e(o({src:n,width:n.width,height:n.height}))},n.onerror=function(){t("There was an error loading the data URI as an image on the following SVG\n"+window.atob(r.slice(26))+"Open the following link to see browser's diagnosis\n"+r)},n.src=r})})},c.download=function(e,t,n){if(navigator.msSaveOrOpenBlob)navigator.msSaveOrOpenBlob(a(t),e);else{var r=document.createElement("a");if("download"in r){r.download=e,r.style.display="none",document.body.appendChild(r);try{var o=a(t),i=URL.createObjectURL(o);r.href=i,r.onclick=function(){return requestAnimationFrame(function(){return URL.revokeObjectURL(i)})}}catch(e){console.error(e),console.warn("Error while getting object URL. Falling back to string URL."),r.href=t}r.click(),document.body.removeChild(r)}else n&&n.popup&&(n.popup.document.title=e,n.popup.location.replace(t))}},c.saveSvg=function(e,t,n){var r=s();return o(e).then(function(e){return c.svgAsDataUri(e,n||{})}).then(function(e){return c.download(t,e,r)})},c.saveSvgAsPng=function(e,t,n){var r=s();return o(e).then(function(e){return c.svgAsPngUri(e,n||{})}).then(function(e){return c.download(t,e,r)})}}();
```
