## Supplementary figures and images for "Evaluating Regional Diversity in Scientific Communication: A Comparative Analysis of COVID-19 Preprints and Peer-Reviewed Publications"

### acceptDeleteIcon.png

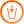

### addNodeIcon.png

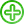

### backIcon.png

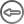

### connectIcon.png

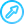

### cross2.png

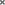

### cross.png

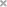

### deleteIcon.png

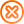

### downArrow.png

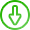

### editIcon.png

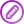

### leftArrow.png

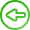

### minus.png

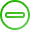

### plus.png

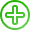

### rightArrow.png

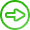

### upArrow.png

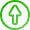

### zoomExtends.png

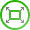
