## Supplementary Material for "Evaluating Regional Diversity in Scientific Communication: A Comparative Analysis of COVID-19 Preprints and Peer-Reviewed Publications"

**Supplementary Figure 1. Dataflow Chart**

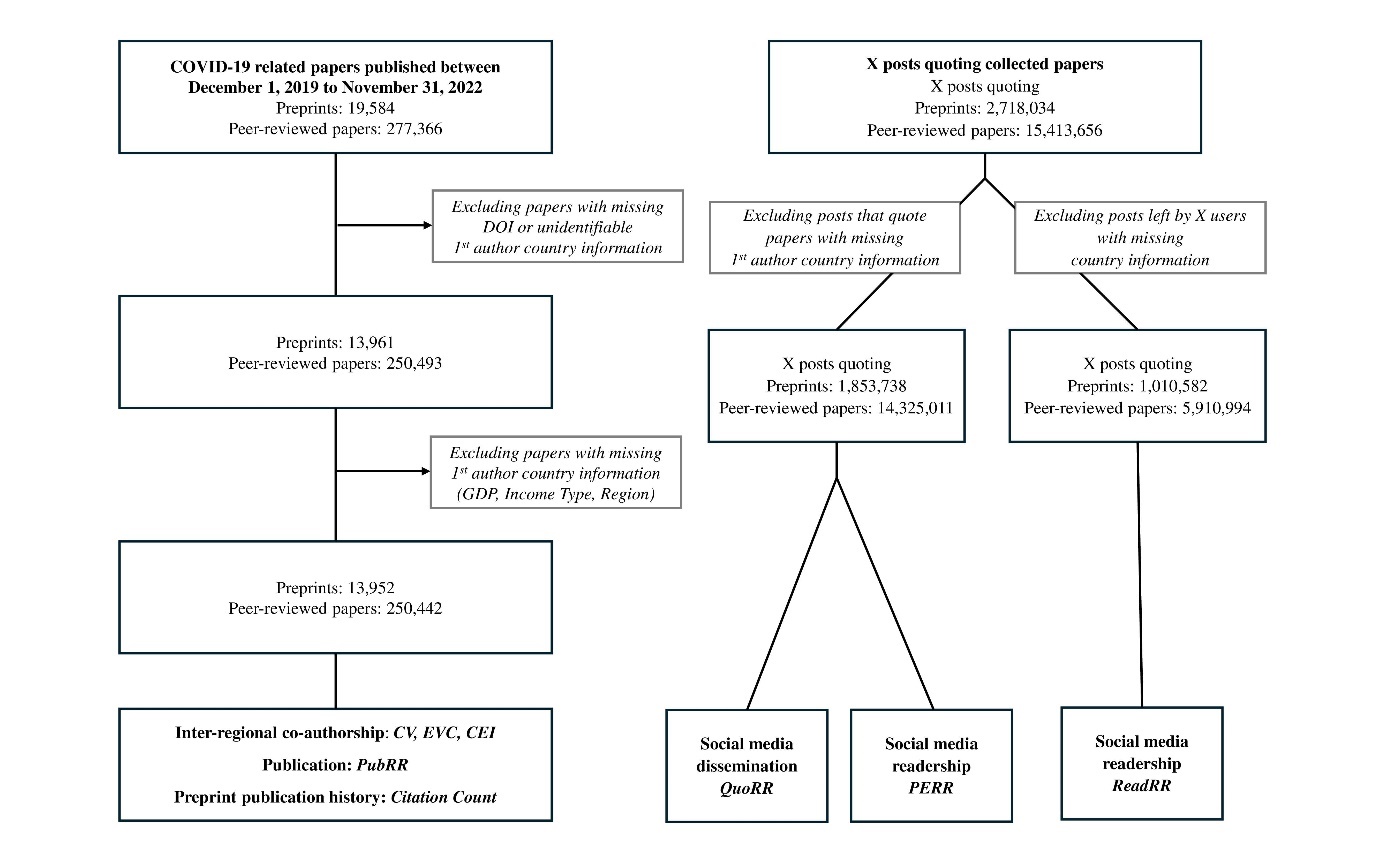

The monthly publication trends of peer-reviewed papers and preprints are categorized by region, ranging from December 2019 to October 2022, with November 2022 excluded due to a lack of publications. The y-axis reflects the proportion of publications within each paper type for a given month. A vertical line indicates when half of the publications were reached. Preprints show a notable surge at the onset of the COVID-19 pandemic across all regions, demonstrating a capacity for rapid response and dissemination of numerous studies. Preprints reached half of the publications faster than peer-reviewed papers in all regions. The publication time difference between preprints and their corresponding subsequent peer-reviewed papers is approximately 196 days when considering the latest version of preprints and around 215 days when considering the first version of preprints.
**Abbreviations**: COVID-19, Coronavirus Disease 2019; DOI, Digitial Object Identifier; GDP, Gross Domestic Product; CV, Coefficient of Variation; EVC, Eigenvector Centrality; CEI, Co-authorship Enhancement Index; PubRR, Publication Relative Ratio; QuoRR, Social Media Quotation Relative Ratio; ReadRR, Social Media Readership Relative Ratio; PERR, Public Engagement Relative Ratio.

**Supplementary Figure 2. Matrix of inter-regional co-authorship network
(A) Higher paper type (B) Difference ratio**

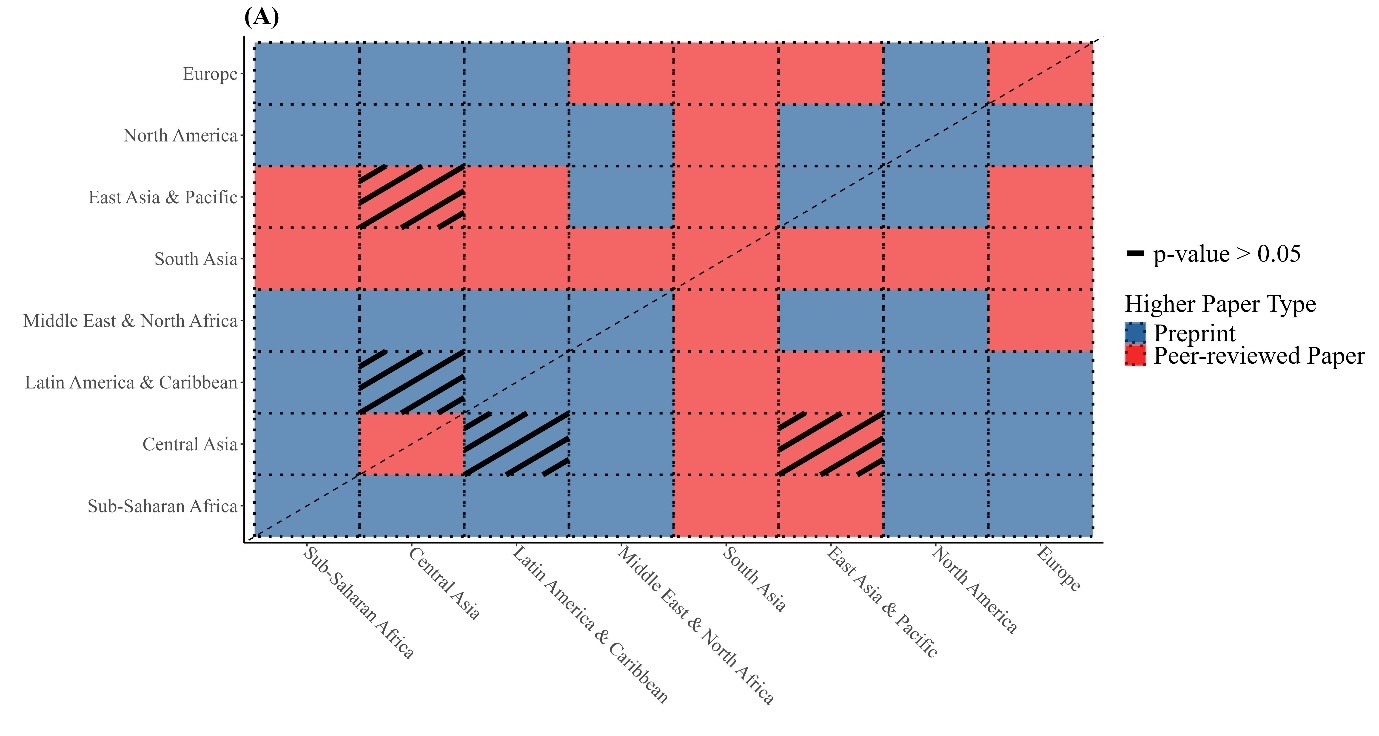

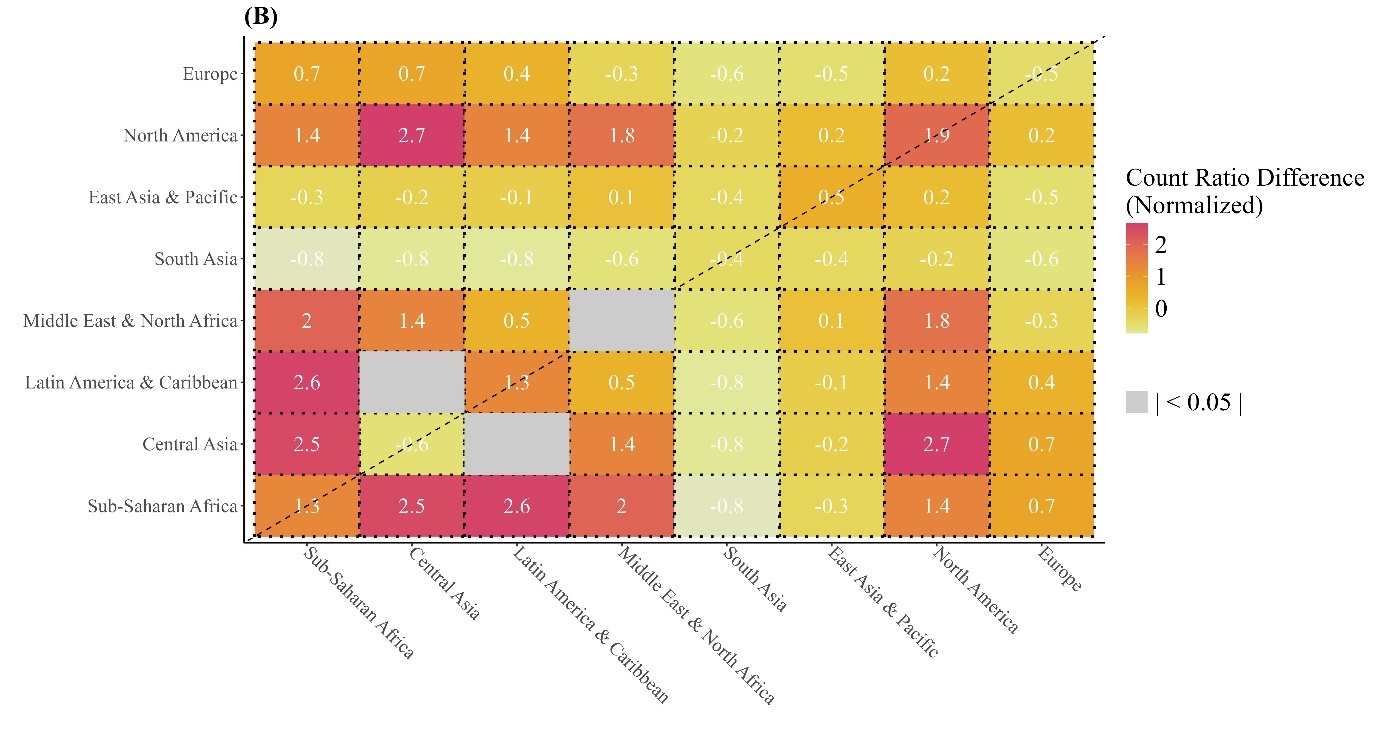

Supplementary Figure 2 is a set of adjacency matrices of inter-regional co-authorship networks among regions depicted in Figure 1. Figure (a) shows what type of paper has a higher inter-regional co-authorship count ratio between regions, with regions arranged according to the average GDP of their constituent countries. If the difference in ratio was shown to be non-significant in two-sample proportion test, black slash was marked in that specific cell. Figure (b) further goes on to show the % difference between two types of paper calculated by (Inter-regional Co-authorship Count Ratio in Preprints – Inter-regional Co-authorship Count Ratio in Peer-reviewed Papers) / (Inter-regional Co-authorship Count Ratio in Peer-reviewed Papers), rounded up to the first decimal point.

**Supplementary Figure 3. (A) Country-specific PubRR by descending order (top to bottom) of the country’s GDP (B) Bar Graph: Percentage of Countries with PubRR > 1 by Region**

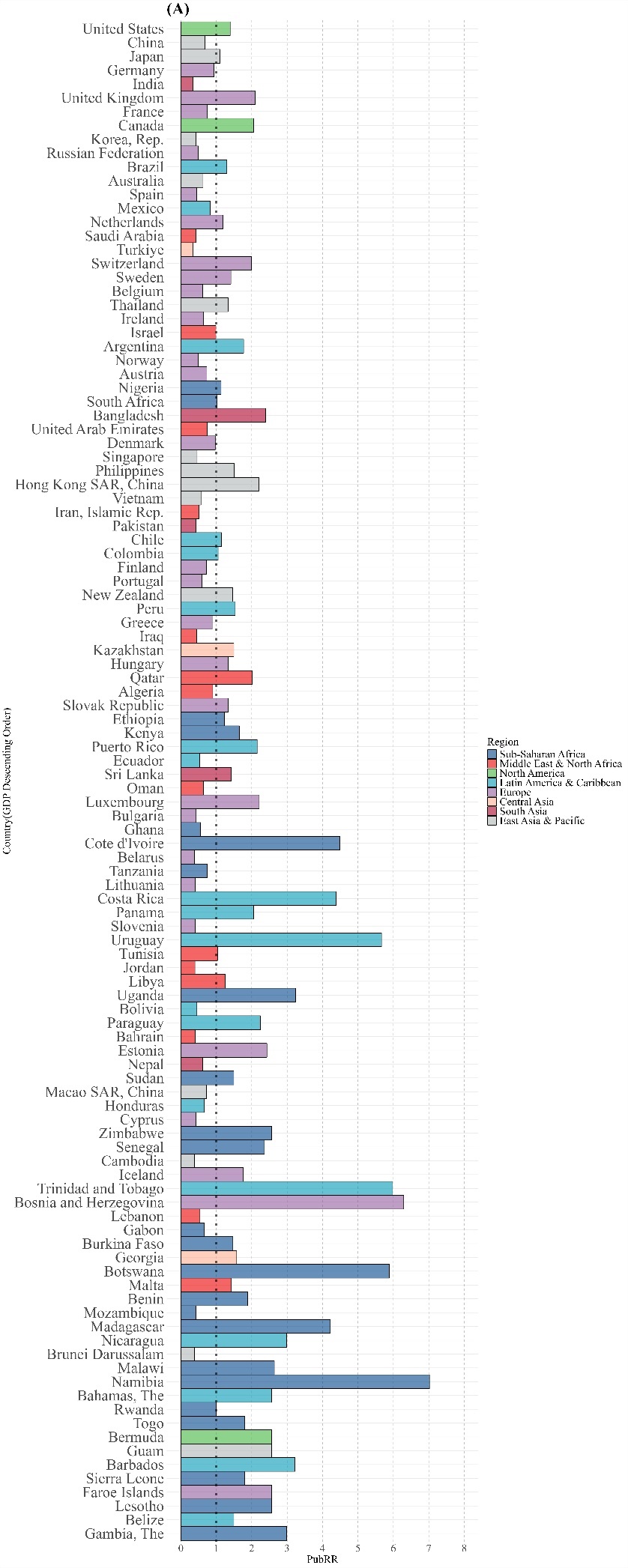

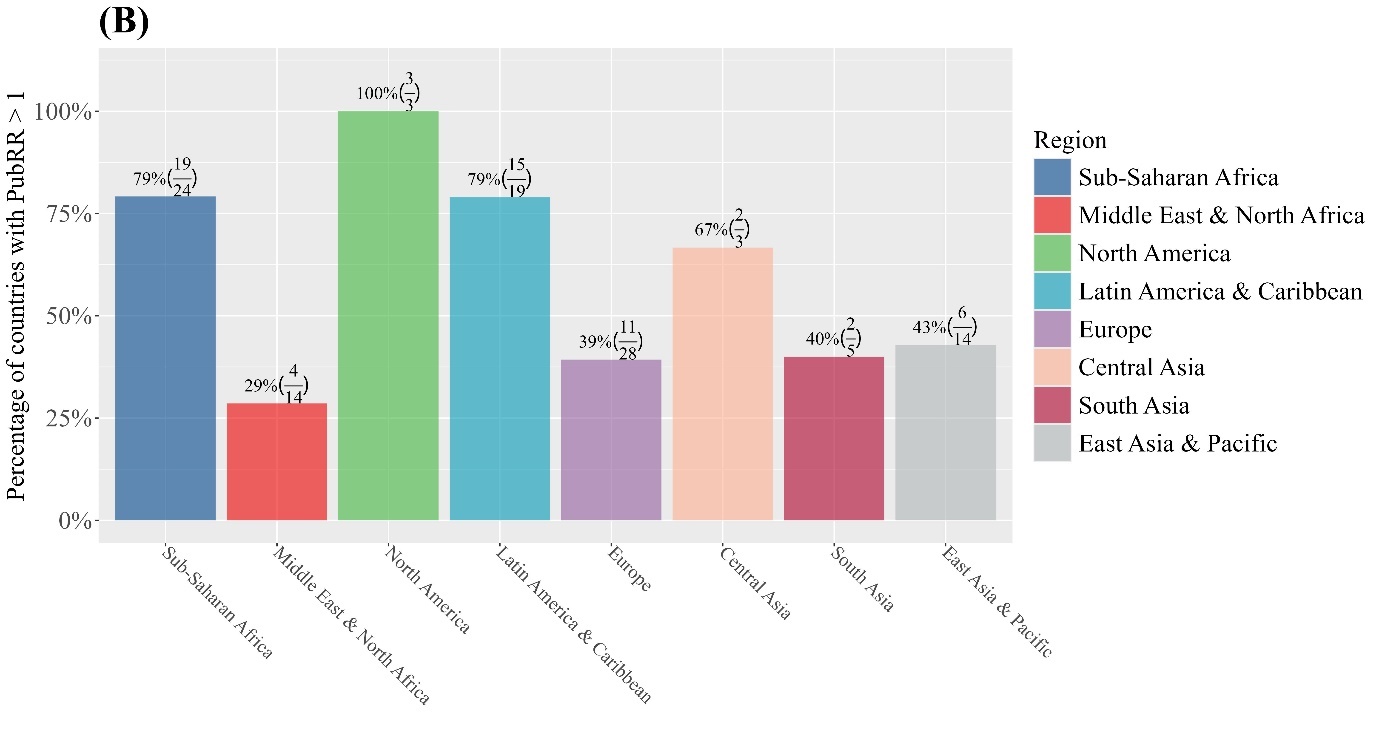

Supplementary Figure 3 (a) shows countries’ PubRR by descending order of the country’s GDP. Top 10% and bottom 10% outliers of PubRR values were excluded like in Figure 2. The countries were colored by the region they belong to. Figure (b) shows the percentage of the number of countries with PubRR value exceeding 1 in each region, along with the total number of countries and the number of countries with PubRR value exceeding 1 countries as well. **Abbreviations:** GDP, Gross Domestic Product; PubRR, Publication Relative Ratio.

**Supplementary Figure 4. Monthly Publication Rate from December 2019 to October 2022 by Region and Paper Type**

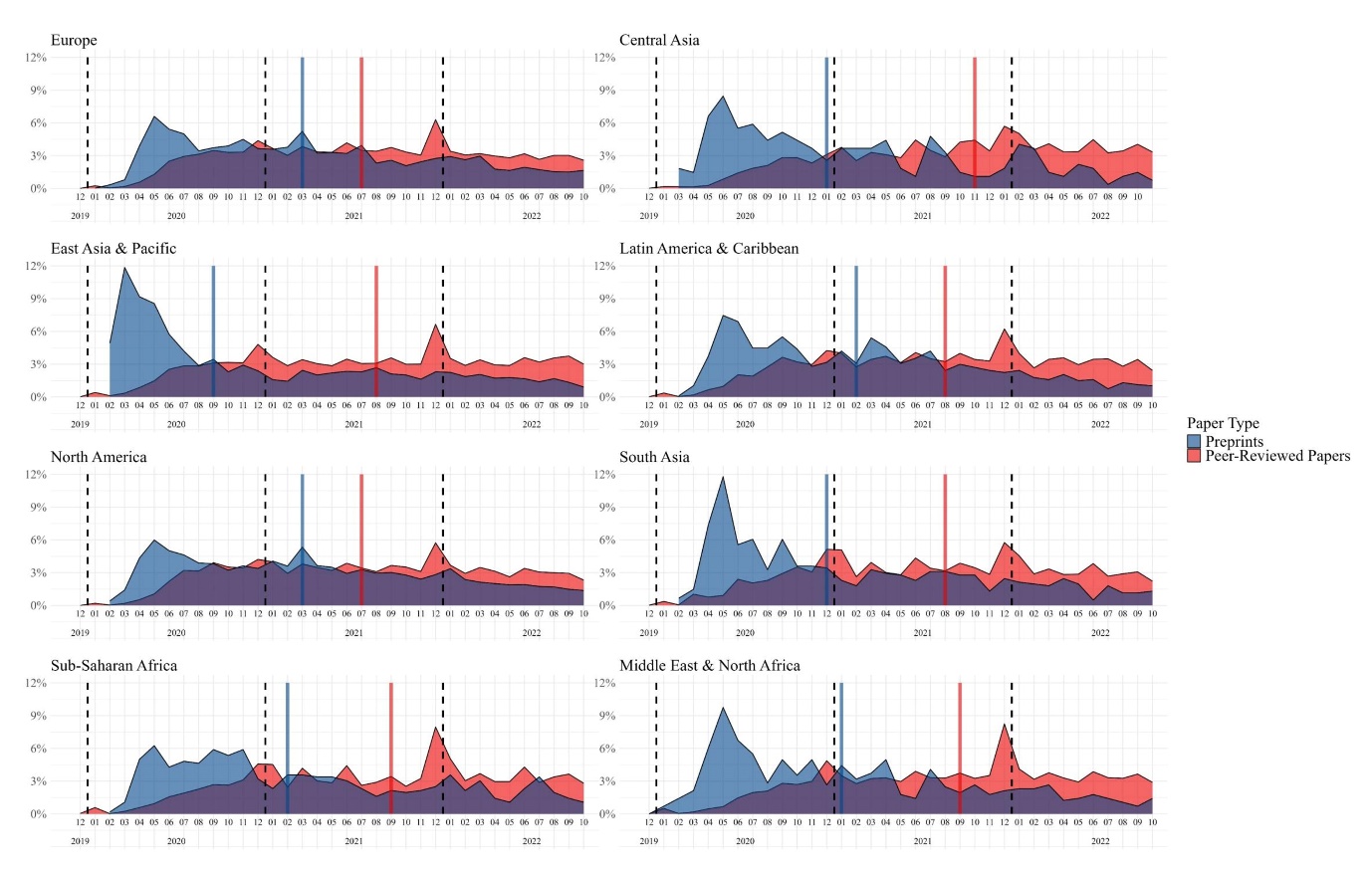

The monthly publication trends of peer-reviewed papers and preprints are categorized by region, ranging from December 2019 to October 2022, with November 2022 excluded due to a lack of publications. The y-axis reflects the proportion of publications within each paper type for a given month. A vertical line indicates when half of the publications were reached. Preprints show a notable surge at the onset of the COVID-19 pandemic across all regions, demonstrating a capacity for rapid response and dissemination of numerous studies. Preprints reached half of the publications faster than peer-reviewed papers in all regions. The publication time difference between preprints and their corresponding subsequent peer-reviewed papers is approximately 196 days when considering the latest version of preprints and around 215 days when considering the first version of preprints.

**Supplementary Figure 5. (A) Country-specific QuoRR by descending order (top to bottom) of the country GDP (B) Bar Graph: Percentage of countries with QuoRR > 1 in each region) (C) QQ-Plots of Social Media Quotation Count by Income Group**

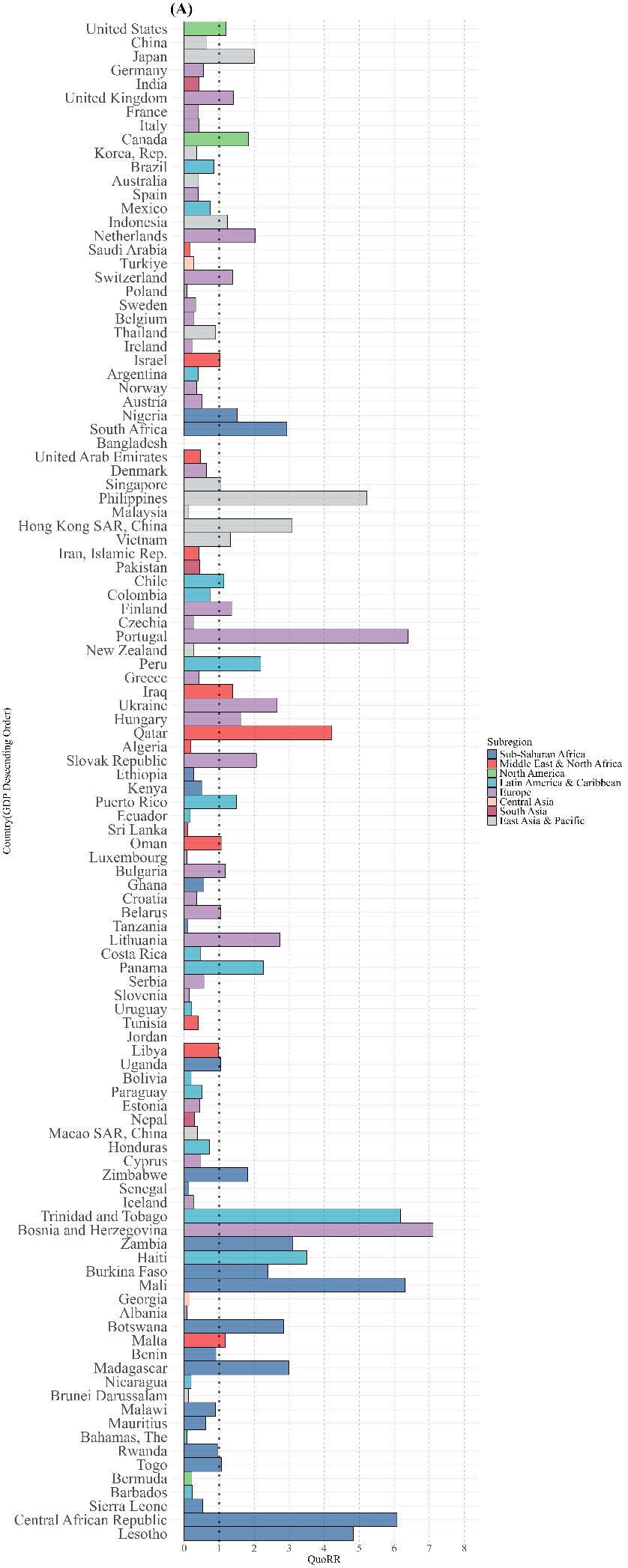

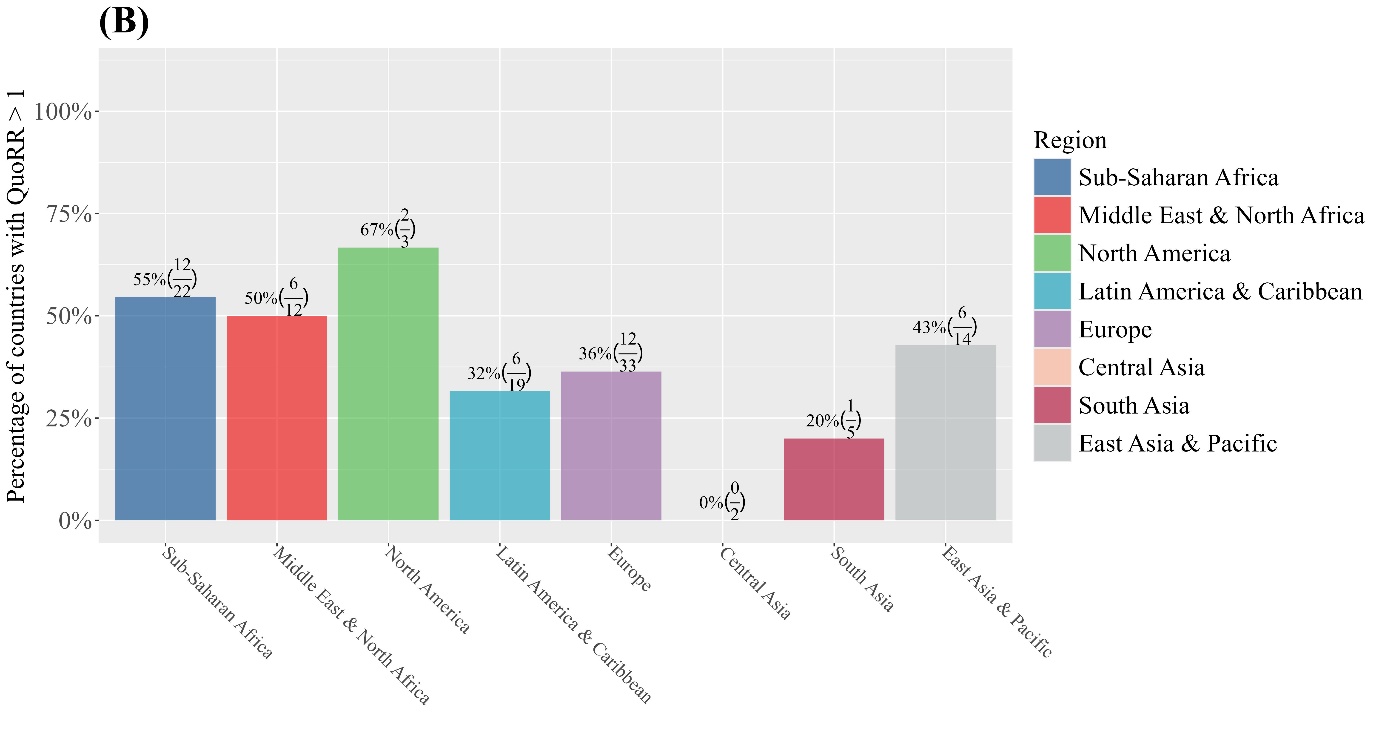

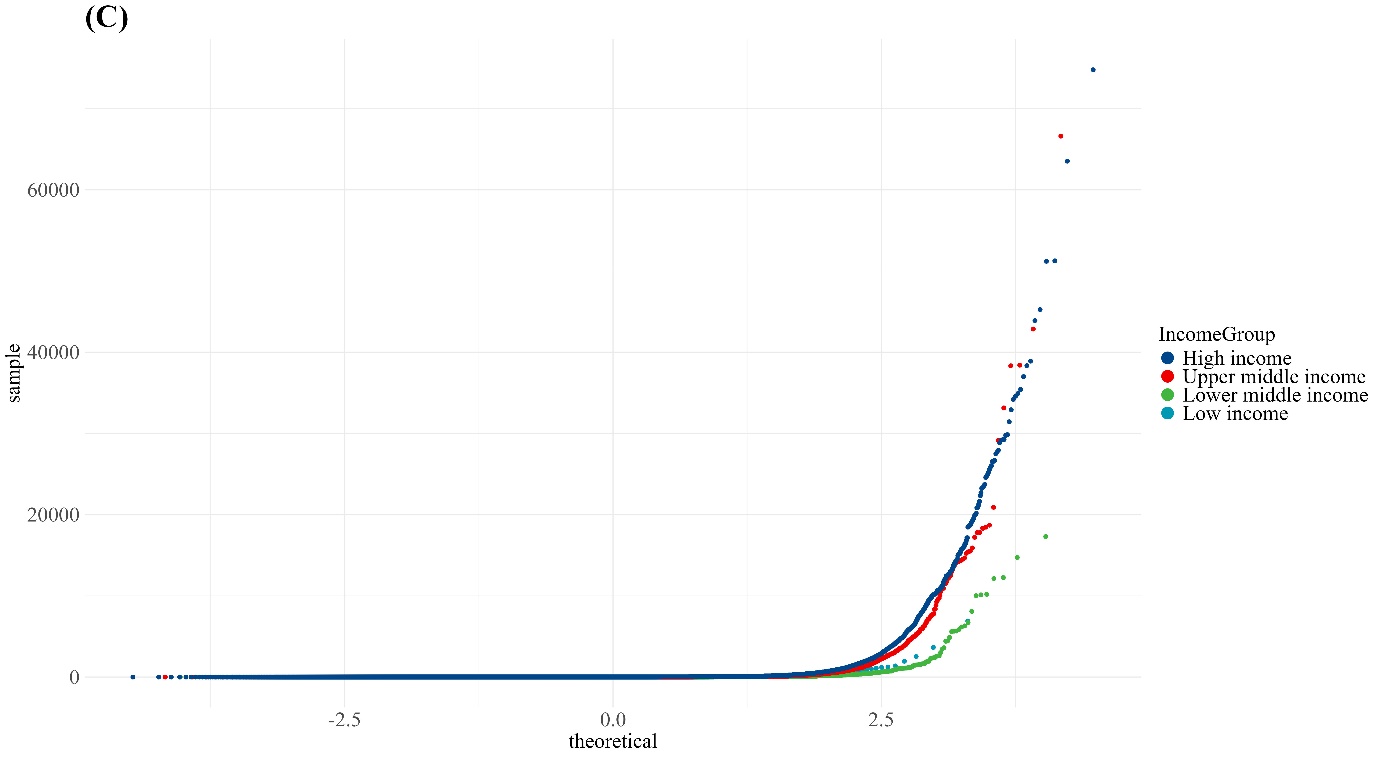

Supplementary Figure 5 (a) shows countries’ QuoRR by descending order of the country’s GDP. Top 10% and bottom 10% outliers of QuoRR values were excluded. Figure (b) shows a bar graph of percentage of the number of countries with QuoRR value exceeding 1 in each region. Under the percentage is written both the total number of countries and the number of countries with QuoRR value exceeding 1 countries in each region as well. Figure (c) shows Quantile-Quantile plots of social media quotation count to ensure non-normality when comparing via Wilcox test. Due to the abundance of data (n > 5,000), shapiro-wilk test was not valid.  **Abbreviations:** GDP, Gross Domestic Product; QuoRR, Social Media Quotation Relative Ratio.

**Supplementary Figure 6. (A) Country-specific ReadRR by descending order of the country’s GDP (B) Geographic heatmap of ReadRR (C) Bar Graph: Percentage of countries with ReadRR > 1 in each region**

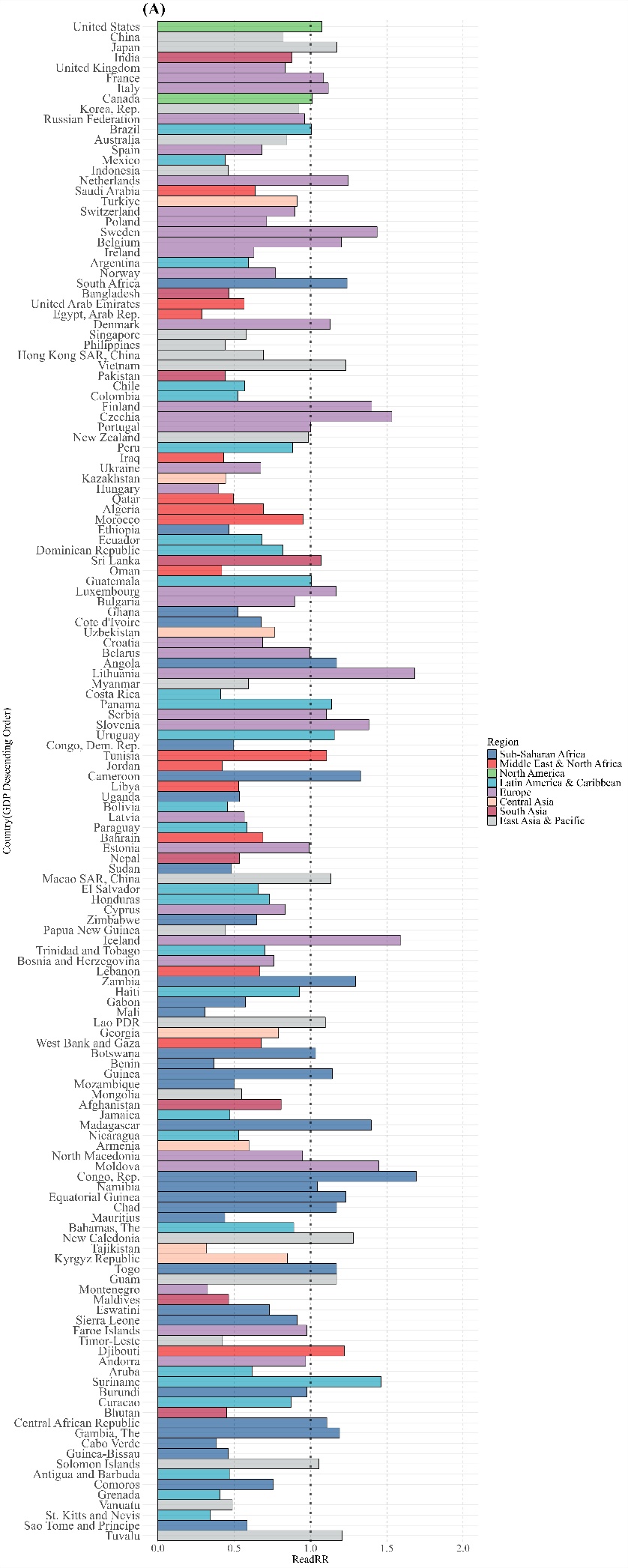

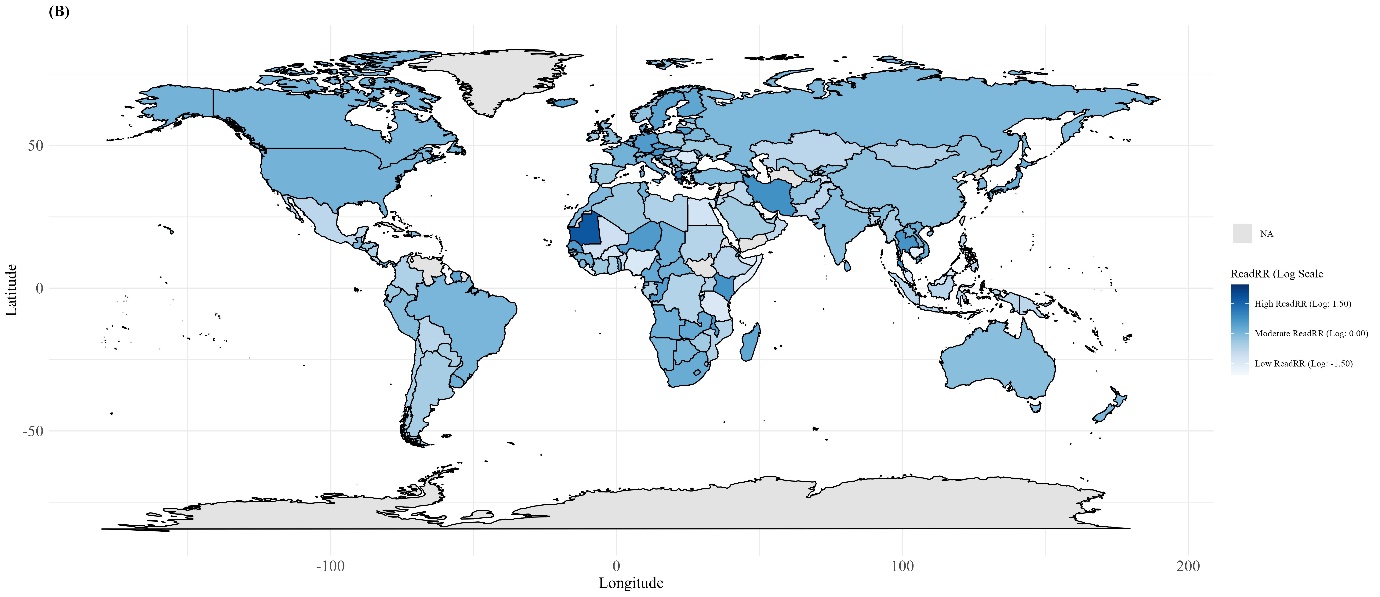

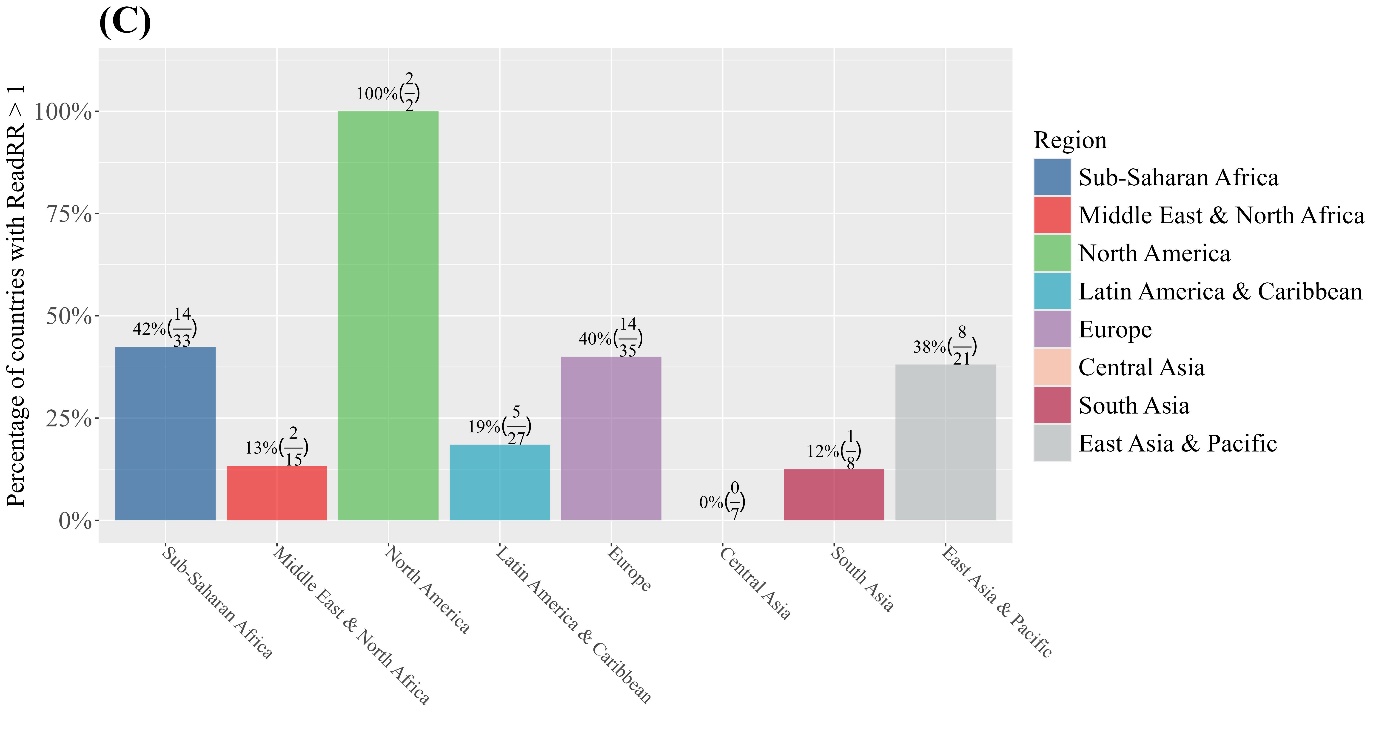

Countries’ ReadRR values are displayed in descending order of GDP, with top and bottom 10% outliers excluded. The geographical heatmap features countries with missing ReadRR values in gray, and the bar graph shows the percentage of countries with ReadRR values exceeding 1 in each region. The non-significant relationship between country-specific ReadRR and GDP is evident in the relatively uniform bar plot, which aligns with the analysis and pattern depicted in Figure 4.
**Abbreviations:** GDP, Gross Domestic Product; ReadRR, Social Media Readership Relative Ratio; NA, Not Available.

**Supplementary Figure 7. Ratio of Social Media Quotation Counts to Papers (A) Peer-reviewed Paper (B) Preprint**

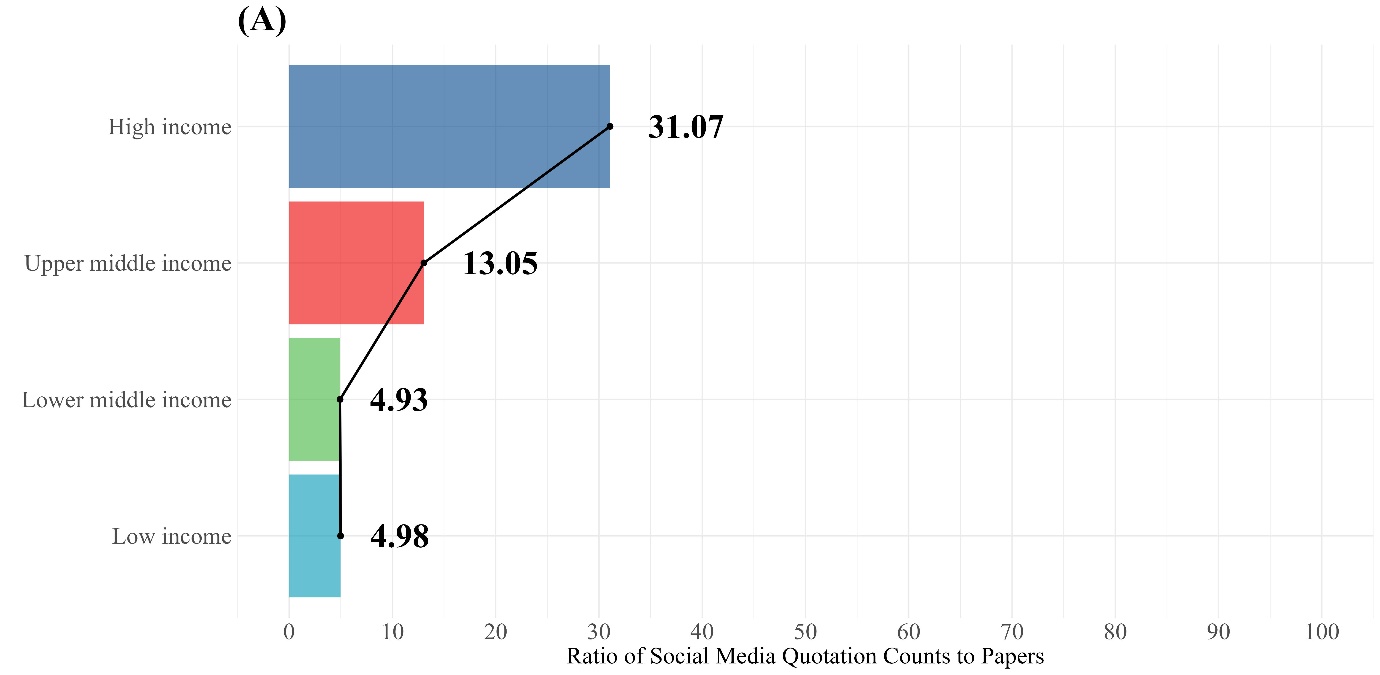

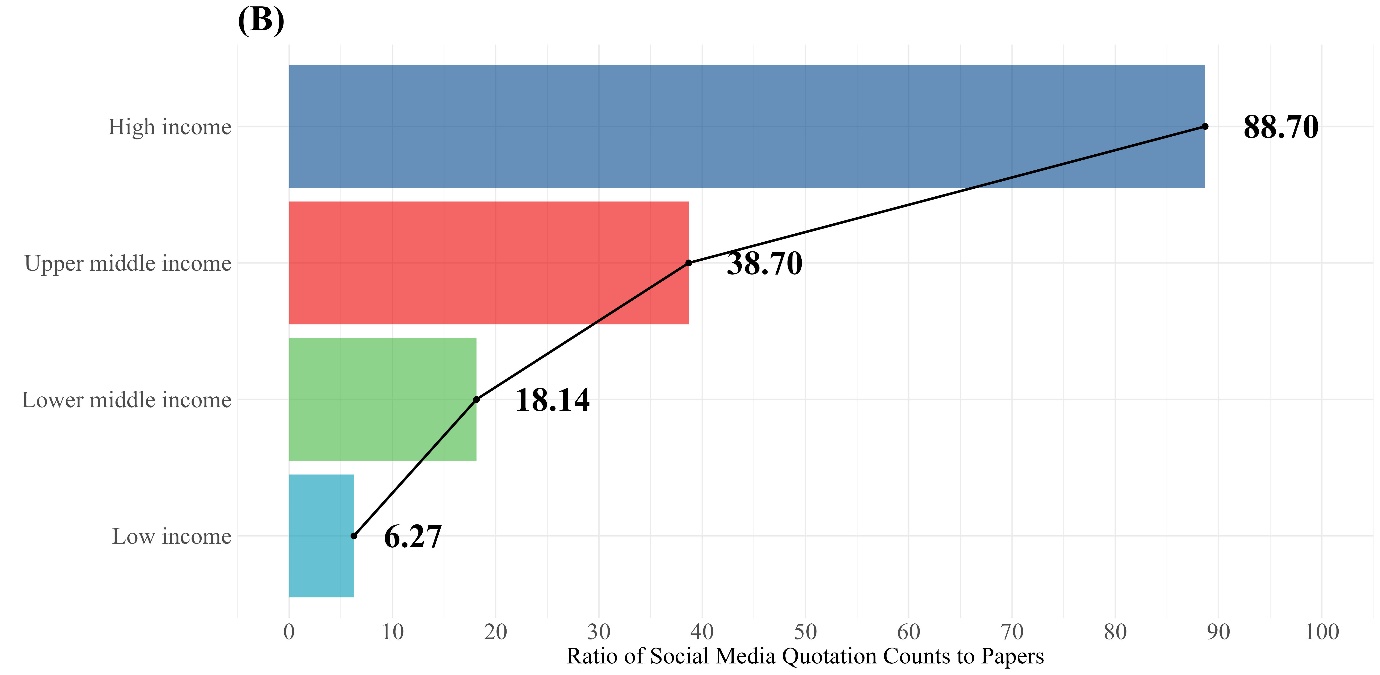

Both peer-reviewed papers and preprints exhibit a similar decline in the ratio of total social media quotation count to total paper counts as income levels decrease, moving from high-income to low-income groups. This trend is further supported by a two-sample proportion test, which shows no significant difference in the proportion of social media quotation count for low-income (p-value = 0.287) and high-income groups (p-value = 1.000) between the two types of papers.

**Supplementary Figure 8. Overall X Demographics by Paper type**

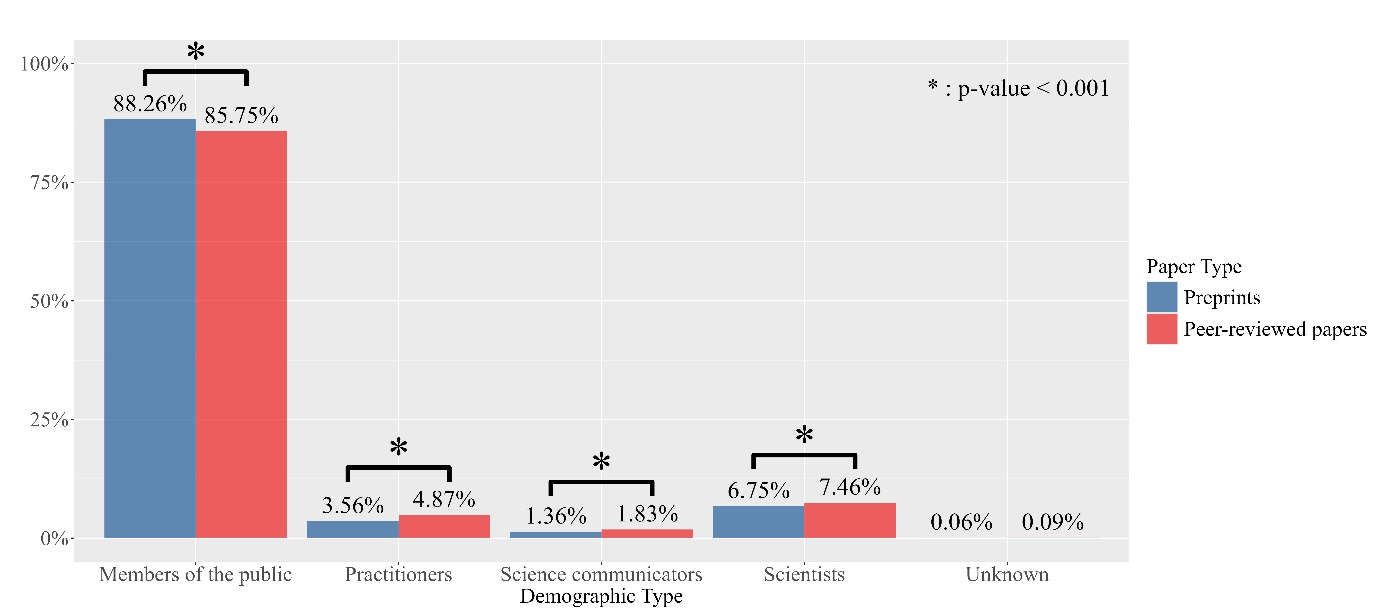

A bar graph of quoted post counts classified by a comprehensive demographic profile of X users for both peer-reviewed papers and preprints shows that the ratio of quoted posts from “members of the public” is approximately 2.5% higher for preprints, with a statistically significant p-value of < 0.001. Other demographic profiles all show statistically significant differences according to the chi-squared test.

**Supplementary Figure 9. (A) Scatter plot of PERR (B) Geographic heatmap of PERR (C) Country-specific PERR by descending order (top to bottom) of the country’s GDP (D) Bar Graph: Percentage of countries with PERR > 1 in each region**

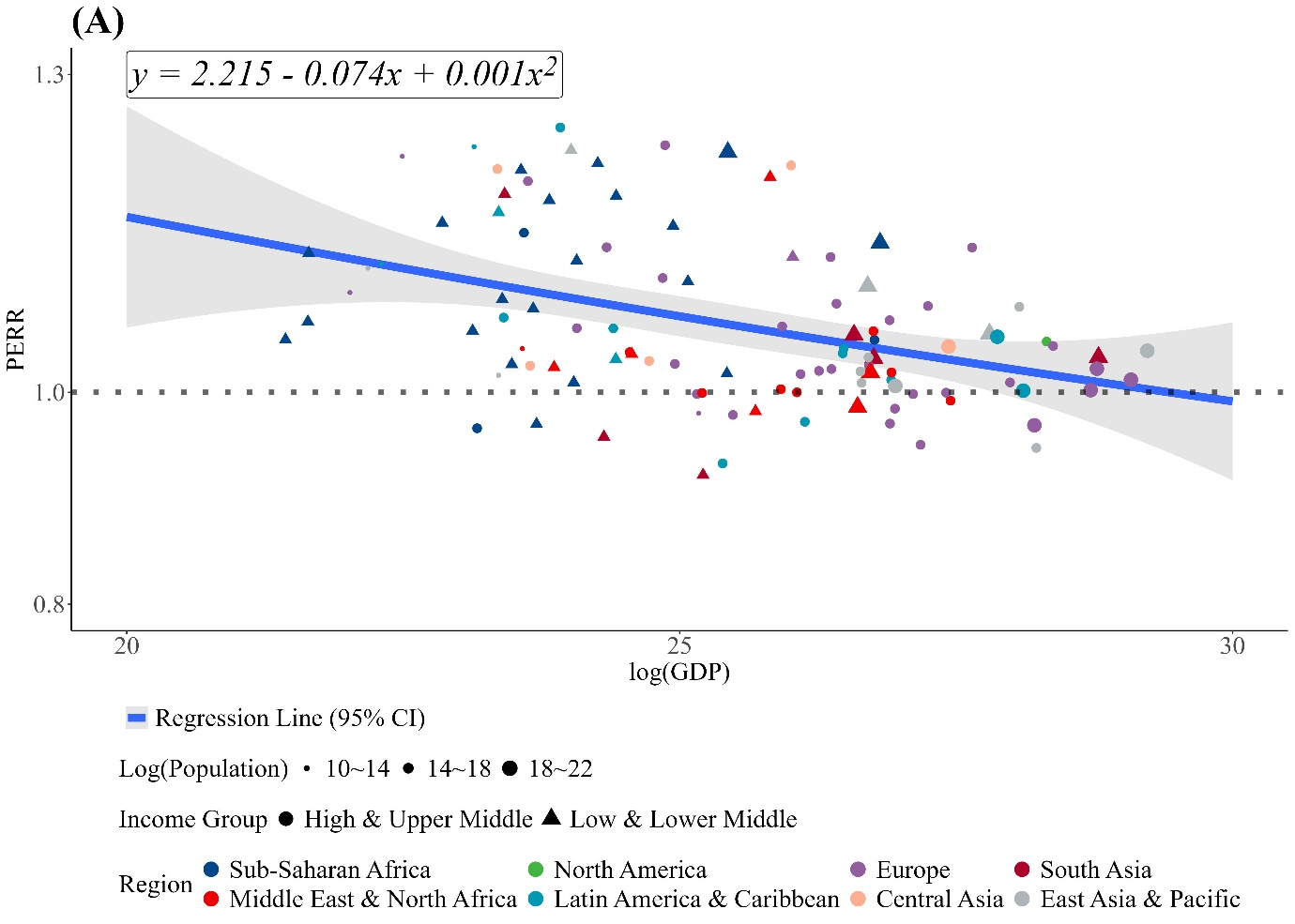

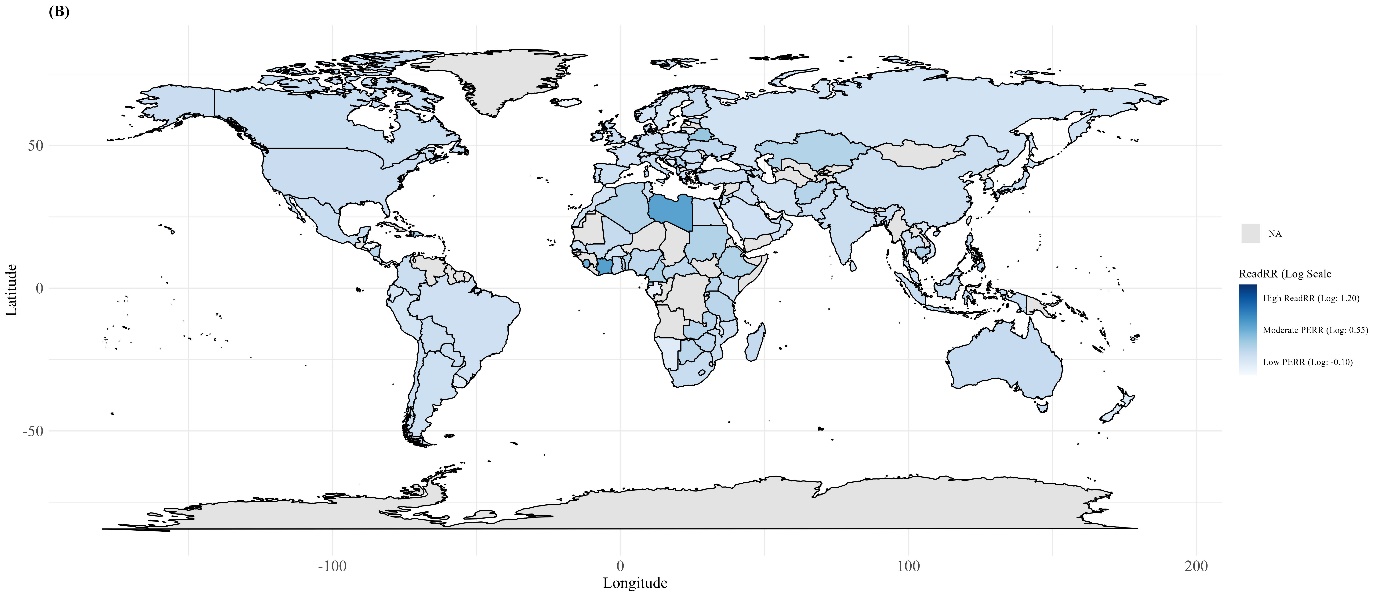

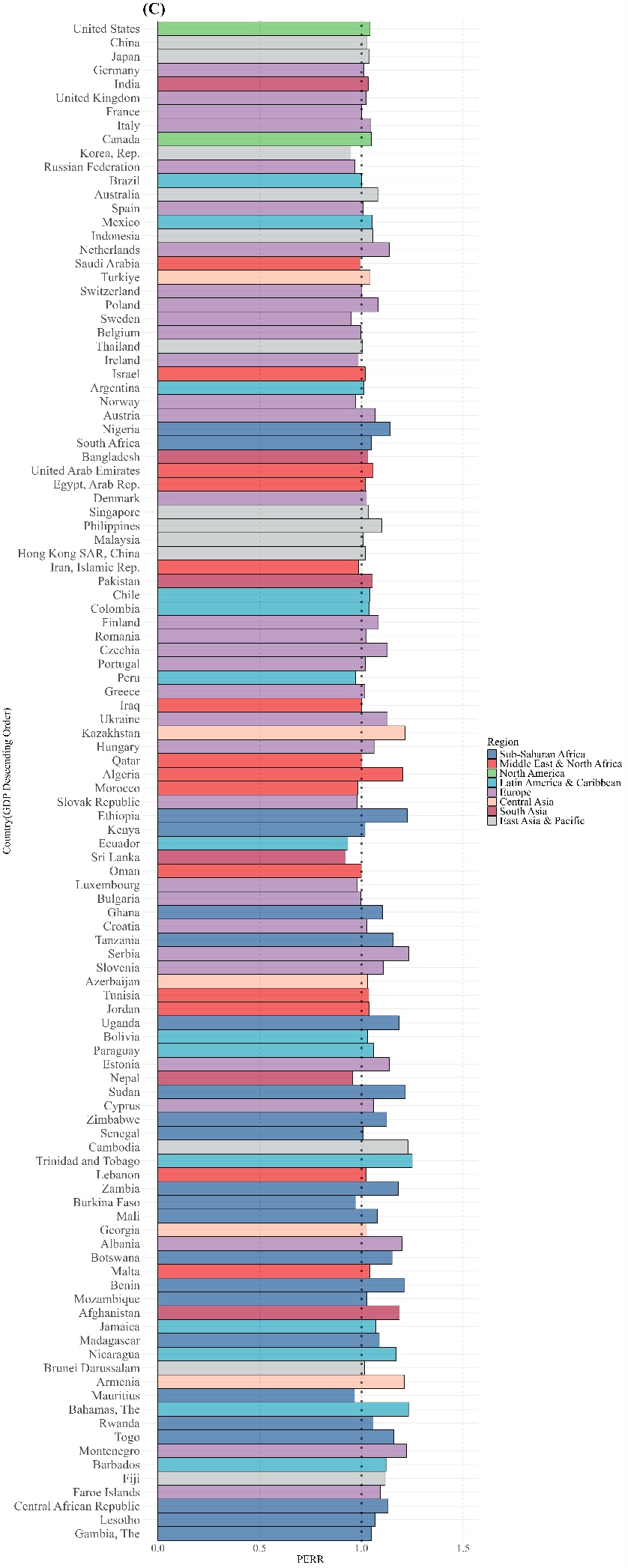

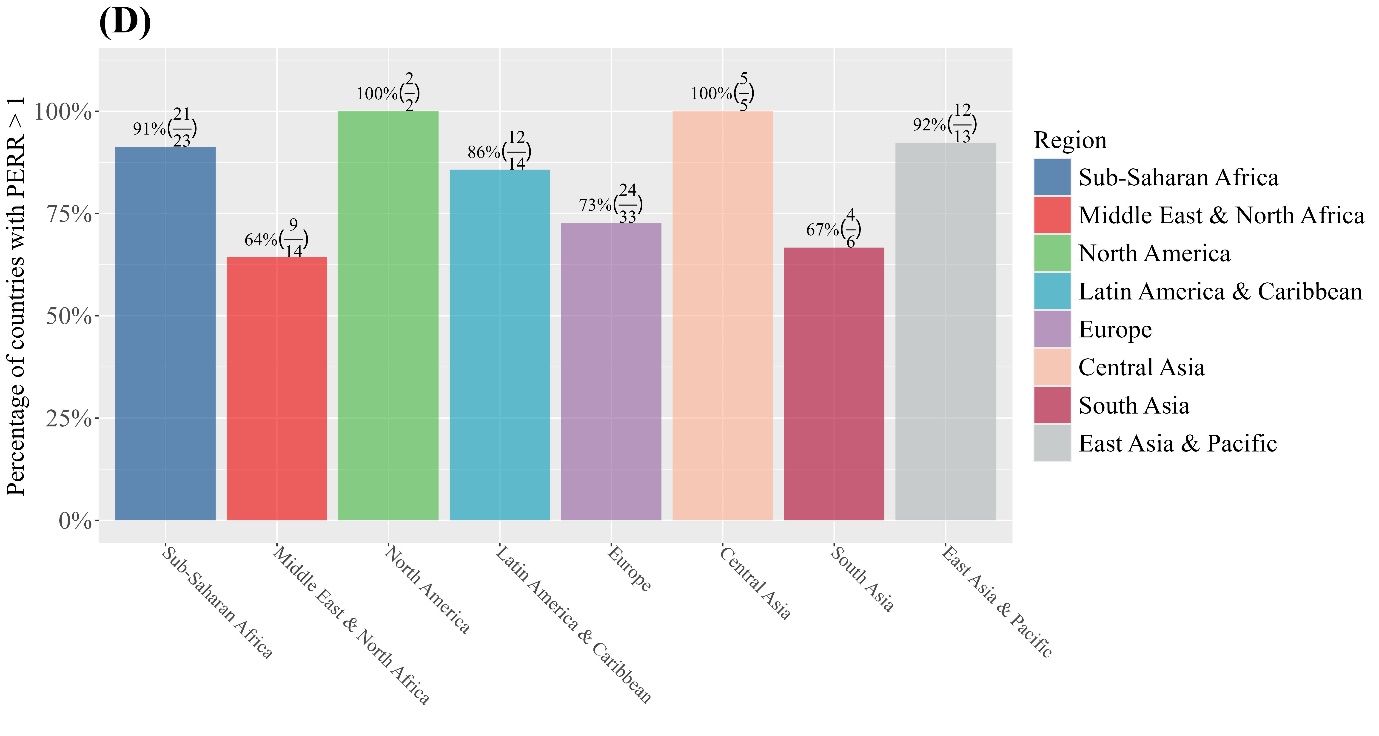

A significant negative correlation between the PERR and GDP was found, with a Pearson correlation coefficient of -0.20 (p-value = 0.021). A second-order polynomial trend line (y = 2.21 – 0.07x + 0.001x², R² = 0.16) illustrates this relationship in the scatter plot, indicating higher public engagement with preprints in countries with lower GDP compared to peer-reviewed papers. Countries’ ratio values are shown in descending order of GDP, excluding the top and bottom 10% outliers. The geographical heatmap highlights countries with high ratio values, such as Palestine, Cameroon, and Cambodia. **Abbreviations:** GDP, Gross Domestic Product; PERR, Public Engagement Relative Ratio; CI, Confidence Interval; Log, Common Logarithm; HMIC, High-Middle Income Countries; LMIC, Low-Middle Income Countries; NA, Not Available.

**Supplementary Figure 10. Median of Citation Count by descending order(top to bottom) of the country’s GDP (A) Peer-reviewed Paper (B) Preprint (C) QQ-Plots of Peer-reviewed papers' citation counts**

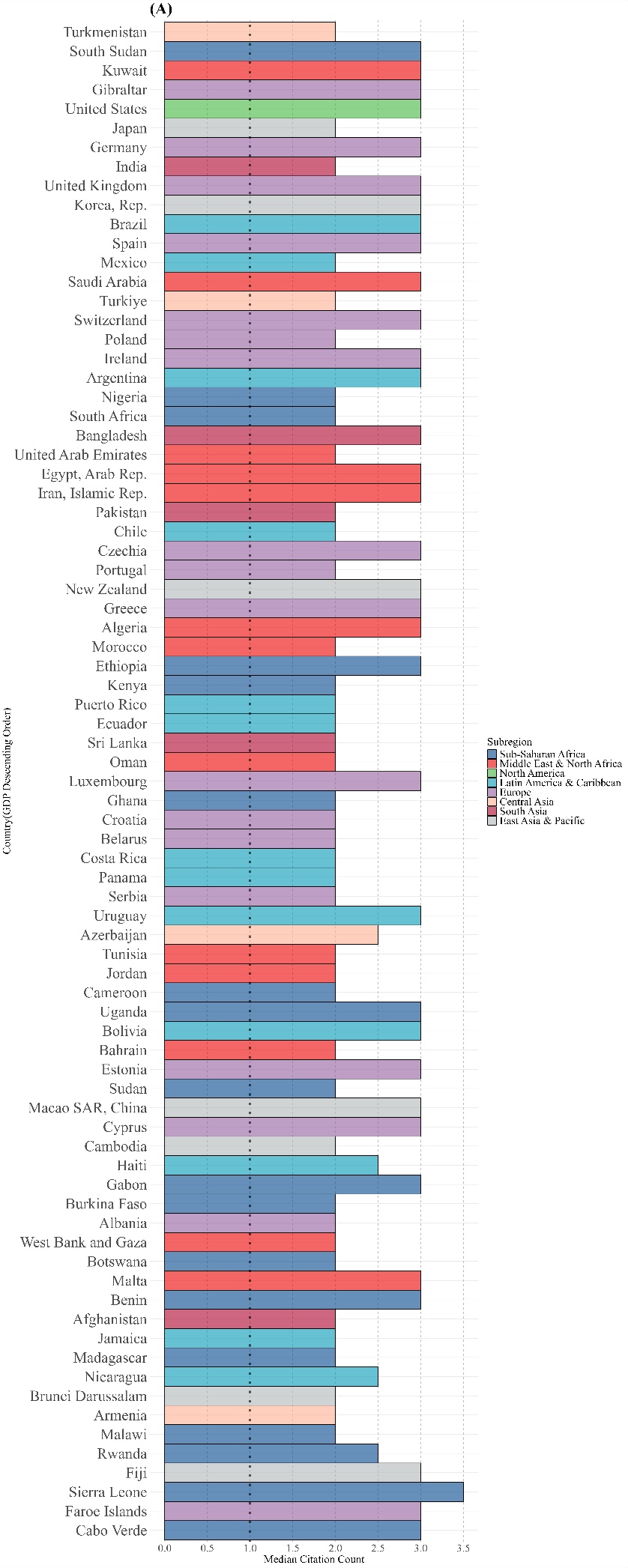

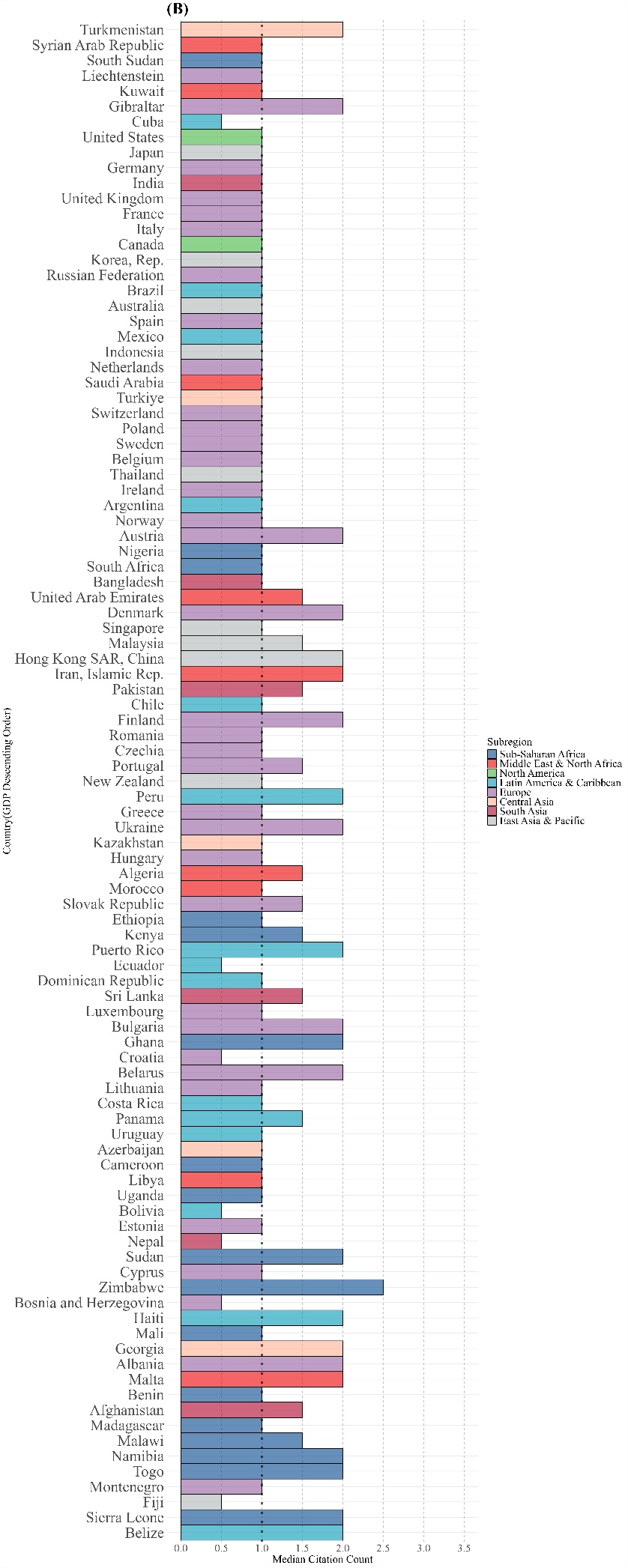

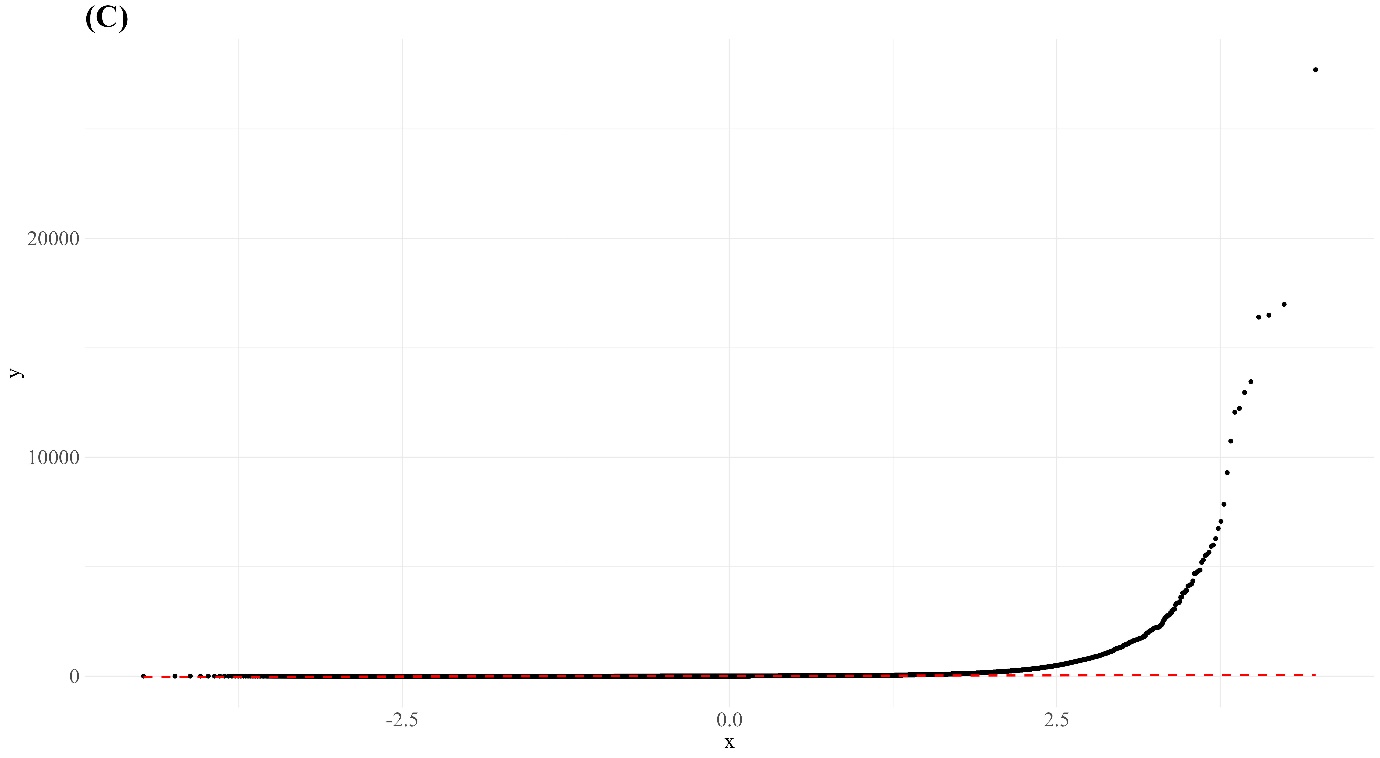

Countries’ median citation counts are shown in descending order of GDP for each type of paper, with the top and bottom 10% outliers excluded. A quantile-quantile plot of peer-reviewed papers’ citation counts was used, revealing non-normality. Consequently, a Wilcox test was conducted for comparisons of citation counts of peer-reviewed papers with and without publication history, as shown in Figure 5.
**Abbreviations:** GDP, Gross Domestic Product; QQ-plot, Quantile-Quantile Plot.

**Supplementary Table 1. Paper Counts Per Region and Income group**

|  | **Peer-reviewed papers** | **Preprints** |  |
| --- | --- | --- | --- |
| **Region** | | | ***p*-value** |
| **East Asia & Pacific** | 53,890 (21.5%) | 2092 (15.0%) | < 0.001 |
| **South Asia** | 19,964 (8.0%) | 611 (4.4%) | < 0.001 |
| **Central Asia** | 5,791 (2.3%) | 272 (1.9%) | 0.006 |
| **Europe** | 81,511 (32.5%) | 4183 (30.0%) | < 0.001 |
| **Latin America & Caribbean** | 14,196 (5.7%) | 1063 (7.6%) | < 0.001 |
| **North America** | 54,365 (21.7%) | 4607 (33.0%) | < 0.001 |
| **Middle East & North Africa** | 16,063 (6.4%) | 564 (4.0%) | < 0.001 |
| **Sub-Saharan Africa** | 4,662 (1.9%) | 560 (4.0%) | < 0.001 |
| **Total** | **250,442 (100.0%)** | **13952 (99.9%)** |  |
| **Income Group** | | | **p-value** |
| **High** | 163,353 (65.2%) | 10,027 (71.9%) | < 0.001 |
| **Upper Middle** | 50,898 (20.3%) | 2,513 (18.0%) | < 0.001 |
| **Lower Middle** | 34,881 (13.9%) | 1,275 (9.1%) | < 0.001 |
| **Low** | 1,310 (0.5%) | 137 (1.0%) | < 0.001 |
| **Total** | **250,442 (99.9%)** | **13,952 (100.0%)** |  |

Paper counts by region and income group for both preprints and peer-reviewed papers are categorized into 8 regions and 4 income groups. The p-value column reports the results of a two-sample proportion test for each row, all of which are significant. According to the first authors' countries, North America has a higher percentage of preprints (33.0%) compared to peer-reviewed papers (21.7%), while East Asia & Pacific and Europe have larger proportions of peer-reviewed papers. Preprints are more frequently represented in Sub-Saharan Africa and Latin America & the Caribbean by region, and in high-income and low-income groups by income level.

**Supplementary Table 2. Social Media Quotation Counts Per Region and Income group**

**(A) Social Media Quotation Counts Per Region and Income group (By Paper)**

|  | **Peer-reviewed papers** | **Preprints** |  |
| --- | --- | --- | --- |
| **Region** | | | ***p*-value** |
| **East Asia & Pacific** | 2,858,895 (20.0%) | 262,087 (14.1%) | < 0.001 |
| **South Asia** | 260,241 (1.8%) | 41,217 (2.2%) | < 0.001 |
| **Central Asia** | 169,808 (1.2%) | 12,163 (0.7%) | < 0.001 |
| **Europe** | 5,215,531 (36.4%) | 574,757 (31.0%) | < 0.001 |
| **Latin America & Caribbean** | 677,435 (4.7%) | 68,997 (3.7%) | < 0.001 |
| **North America** | 4,293,892 (30.0%) | 718,293 (38.8%) | < 0.001 |
| **Middle East & North Africa** | 732,486 (5.1%) | 112,730 (6.1%) | < 0.001 |
| **Sub-Saharan Africa** | 116,723 (0.8%) | 63,494 (3.4%) | < 0.001 |
| **Total** | **14,325,011 (99.9%)** | **1,853,738 (100.0%)** |  |
| **Income Group** | | | **p-value** |
| **High** | 11,633,706 (81.2%) | 1,527,919 (82.4%) | < 0.001 |
| **Upper Middle** | 2,224,448 (15.5%) | 234,551 (12.7.%) | < 0.001 |
| **Lower Middle** | 427,481 (3.0%) | 87,212 (4.7.%) | < 0.001 |
| **Low** | 39,376 (0.3%) | 4,056 (0.2%) | < 0.001 |
| **Total** | **14,325,011 (100.0%)** | **1,853,738 (100.0%)** |  |

Social media quotation counts for peer-reviewed papers and preprints are categorized by 8 regions and 4 income groups based on the country of the lead authors of the papers, which are also used for analysis of dissemination. The p-value column reports the results of a two-sample proportion test for each row, all of which are statistically significant. North America and Europe have the highest counts for both peer-reviewed papers and preprints. High-income groups dominate the quotation counts, with 81.2% for peer-reviewed papers and 82.4% for preprints. Other regions, such as East Asia & Pacific, South Asia, and Latin America & Caribbean, as well as upper-middle and lower-middle income groups, display varying proportions, all showing significant differences between peer-reviewed papers and preprints.

**(B) Social Media Quotation Counts Per Region and Income group (By User)**

|  | **Peer-reviewed papers** | **Preprints** |  |
| --- | --- | --- | --- |
| **Region** | | | ***p*-value** |
| **East Asia & Pacific** | 775,469 (13.1%) | 149,965 (14.8%) | < 0.001 |
| **South Asia** | 119,371 (2.0%) | 17,327 (1.7%) | < 0.001 |
| **Central Asia** | 44,526 (0.8%) | 6,870 (0.7%) | < 0.001 |
| **Europe** | 2,009,131 (34.0%) | 333,300 (33.0%) | < 0.001 |
| **Latin America & Caribbean** | 564,430 (9.6%) | 68,564 (6.8%) | < 0.001 |
| **North America** | 2,333,122 (39.5%) | 424,188 (42.0%) | < 0.001 |
| **Middle East & North Africa** | 26,316 (0.5%) | 4,032 (0.4%) | < 0.001 |
| **Sub-Saharan Africa** | 38,629 (0.7%) | 6,359 (0.6%) | 0.005 |
| **Total** | **5,910,994 (100.2%)** | **1,010,605 (100.0%)** |  |
| **Income Group** | | | **p-value** |
| **High** | 5,071,358 (85.8%) | 889,414 (88.0%) | < 0.001 |
| **Upper Middle** | 662,978 (11.2%) | 97,210 (9.6%) | < 0.001 |
| **Lower Middle** | 170,709 (2.9%) | 23,134 (2.3%) | < 0.001 |
| **Low** | 5,949 (0.1%) | 847 (0.1%) | < 0.001 |
| **Total** | **5,910,994 (100.0%)** | **1,010,605 (100.0%)** |  |

Social media quotation counts for peer-reviewed papers and preprints are categorized by region and income group based on the country information of the X users citing the papers. 8 regions and 4 income groups were used to classify the quotations and also used for analysis of readership. The p-value column displays the results of a two-sample proportion test for each row, all showing statistical significance. The data indicates that North America has the highest social media quotation counts for both peer-reviewed papers (39.5%) and preprints (42.0%). High-income groups are prominently represented in the social media quotation counts, with 85.8% for peer-reviewed papers and 88.0% for preprints. Other regions, including East Asia & Pacific, Europe, and Latin America & Caribbean, and income groups such as upper-middle and lower-middle, show varying proportions, all showing significant differences between peer-reviewed papers and preprints. The discrepancy in total quotation counts between (A) and (B) arises from the lack of X users’ country information due to X’s stringent privacy policies.

**Supplementary Table 3. Inter-regional Co-authorship Counts**

**(A) Peer-reviewed papers**

|  | East Asia & Pacific | South Asia | Central Asia | Europe | Latin America & Caribbean | North America | Middle East & North Africa | Sub-Saharan Africa |
| --- | --- | --- | --- | --- | --- | --- | --- | --- |
| East Asia & Pacific | 443,115 (6.26) |  |  |  |  |  |  |  |
| South Asia | 96,315 (1.36) | 218,603 (3.09) |  |  |  |  |  |  |
| Central Asia | 2,005 (0.03) | 2,868 (0.04) | 831 (0.01) |  |  |  |  |  |
| Europe | 402,249 (5.68) | 166,189 (2.35) | 4,073 (0.06) | 3,166,938 (44.75) |  |  |  |  |
| Latin America & Caribbean | 35,430 (0.50) | 35,073 (0.50) | 770 (0.01) | 151,710 (2.14) | 132,241 (1.87) |  |  |  |
| North America | 155,914 (2.20) | 85,986 (1.22) | 1,549 (0.02) | 551,951 (7.80) | 60,331 (0.85) | 531,010 (7.50) |  |  |
| Middle East & North Africa | 45,233 (0.64) | 48,048 (0.68) | 946 (0.01) | 234,140 (3.31) | 21,093 (0.30) | 63,193 (0.89) | 135,856 (1.92) |  |
| Sub-Saharan Africa | 30,692 (0.43) | 49,676 (0.70) | 851 (0.01) | 67,142 (0.95) | 13,400 (0.19) | 37,531 (0.53) | 23,155 (0.33) | 60,496 (0.85) |

**(B) Preprints**

|  | East Asia & Pacific | South Asia | Central Asia | Europe | Latin America & Caribbean | North America | Middle East & North Africa | Sub-Saharan Africa |
| --- | --- | --- | --- | --- | --- | --- | --- | --- |
| East Asia & Pacific | 23,490 (9.55) |  |  |  |  |  |  |  |
| South Asia | 2,104 (0.86) | 4,492 (1.83) |  |  |  |  |  |  |
| Central Asia | 58 (0.02) | 24 (0.01) | 12 (0.00) |  |  |  |  |  |
| Europe | 6,470 (2.63) | 2,073 (0.84) | 240 (0.10) | 58,586 (23.81) |  |  |  |  |
| Latin America & Caribbean | 1,078 (0.44) | 282 (0.11) | 28 (0.01) | 7,592 (3.09) | 10,685 (4.34) |  |  |  |
| North America | 6,742 (2.74) | 2,267 (0.92) | 197 (0.08) | 23,134 (9.40) | 5,067 (2.06) | 53,609 (21.79) |  |  |
| Middle East & North Africa | 1,794 (0.73) | 640 (0.26) | 79 (0.03) | 5,862 (2.38) | 1,070 (0.43) | 6,046 (2.46) | 4,923 (2.00) |  |
| Sub-Saharan Africa | 740 (0.30) | 377 (0.15) | 103 (0.04) | 3,992 (1.62) | 1,677 (0.68) | 3,149 (1.28) | 2,440 (0.99) | 4,889 (1.99) |

Supplementary Table 3 presents inter-regional co-authorship counts in a diagonal table format, where only the lower triangular half is populated to avoid redundancy. Table (A) shows counts for peer-reviewed papers, while Table (B) shows counts for preprints. Figure 1 is based on this edge weight distribution, divided into six quantiles with the following cutoff values: for peer-reviewed papers, the cutoffs are 2,725.2, 33,622.9, 55,040.8, 108,458.0, and 221,283.1; for preprints, the cutoffs are 181.4, 960.8, 2,186.1, 4,900.5, and 6,888.6.

**Supplementary Table 4. Statistics for countries exhibiting high PubRR values**

| Country name | Number of Peer-reviewed papers  Publications | Number of Preprints  Publications | PubRR |
| --- | --- | --- | --- |
| Mali | 26 | 23 | 15.87 |
| Cameroon | 270 | 168 | 11.16 |
| South Sudan | 7 | 13 | 33.32 |
| Ukraine | 42 | 23 | 9.83 |
| Turkmenistan | 2 | 4 | 35.88 |

Statistics for countries exhibiting high PubRR values, as shown in the geographical heatmap in Figure 2 (B), are shown. The table includes the number of peer-reviewed paper publications and preprint publications, along with the calculated PubRR values. A high PubRR value indicates that, relative to the distribution of peer-reviewed papers, there were more preprints published in that area, suggesting a publication preference for preprints.
**Abbreviations:** PubRR, Publication Relative Ratio.

**Supplementary Table 5. Average of Social Media Quotation Counts Per Region and Income Group**

|  | **Median value (Q1–Q3)** | | **Mean value (SD)** | |  |
| --- | --- | --- | --- | --- | --- |
|  | **Peer-reviewed** | **Preprint** | **Peer-reviewed** | **Preprint** |  |
| **Region** | | | | | **p-value** |
| East Asia & Pacific | 4 (2-13) | 7 (4-20) | 85.33 (890.67) | 125.28 (888.01) | < 0.001 |
| South Asia | 3 (1-8) | 5 (4-10) | 27.20 (299.58) | 67.46 (718.83) | < 0.001 |
| Central Asia | 3 (1-9) | 7 (4-13.25) | 51.49 (441.62) | 44.72 (271.00) | < 0.001 |
| Europe | 6 (2-19) | 10 (5-32) | 90.56 (891.02) | 137.40 (808.25) | < 0.001 |
| Latin America & Caribbean | 5 (2-15) | 7 (4-21) | 73.59 (770.91) | 64.91 (311.40) | < 0.001 |
| North America | 7 (2-21) | 9 (5-32) | 100.43 (810.35) | 156.02 (975.77) | < 0.001 |
| Middle East & North Africa | 3 (1-10) | 7 (4-26) | 74.75 (752.67) | 200.23 (920.25) | < 0.001 |
| Sub-Saharan Africa | 4 (2-13) | 6 (4-14) | 39.78 (300.75) | 113.38 (821.06) | < 0.001 |
| **Income Group** | | | | | **p-value** |
| High | 6 (2–22) | 9 (5–32) | 95.50 (855.03) | 152.41 (923.85) | < 0.001 |
| Upper-middle | 3 (1–12) | 7 (4–17) | 72.74 (808.9) | 93.34 (619.77) | < 0.001 |
| Lower-middle | 3 (1–10) | 6 (4–11) | 28.52 (256.96) | 68.40 (646.96) | < 0.001 |
| Low | 4 (2–14) | 6 (4–13) | 45.33 (289.77) | 29.61 (94.02) | < 0.001 |

Descriptive statistics for social media quotation counts of both peer-reviewed papers and preprints are presented, classified by 8 regions and 4 income groups based on the paper’s country. The p-value column shows the results of Wilcox rank-sum test comparing the two types, all indicating significance. Preprints have consistently higher median quotation counts compared to peer-reviewed papers across all regions and income groups. This suggests greater engagement with preprints on X, indicating broader dissemination compared to peer-reviewed papers regardless of the region or economic status of the lead authors' countries. **Abbreviations:** SD, Standard Deviation.

**Supplementary Table 6. Statistics for countries exhibiting high QuoRR values**

| Country name | Number of Peer-reviewed Paper Quotations | Number of Preprint Quotations | QuoRR |
| --- | --- | --- | --- |
| Sudan | 253 | 1,202 | 36.71 |
| South Sudan | 90 | 732 | 62.85 |
| Cameroon | 3,426 | 35,823 | 80.80 |
| Turkmenistan | 12 | 30 | 19.32 |
| Kazakhstan | 284 | 483 | 13.14 |

The number of peer-reviewed paper quotations, preprint quotations for countries with high QuoRR values, highlighted in the geographical heatmap in Figure 3 (C), are shown. QuoRR values are included as well. A high QuoRR means that, relative to the quoted posts for peer-reviewed papers, there were more social media quotations for preprints with lead authors from that area, indicating higher engagement with preprints on X.
**Abbreviations:** QuoRR, Social Media Quotation Relative Ratio.

**Supplementary Table 7. Statistics for countries exhibiting high PERR values**

| Country name | Members of the public rate in  Peer-reviewed paper quotations | Members of the public rate in  Preprint quotations | PERR |
| --- | --- | --- | --- |
| Algeria | 0.72 | 0.87 | 1.21 |
| Palestine | 0.20 | 0.87 | 4.36 |
| Cameroon | 0.71 | 0.93 | 1.26 |
| Kazakhstan | 0.73 | 0.89 | 1.21 |
| Armenia | 0.83 | 1.00 | 1.21 |
| Cambodia | 0.81 | 1.00 | 1.23 |

The members of the public rate in peer-reviewed paper quotations and preprint quotations, for countries with high PERR values, are shown. These countries are also shown in the geographical heatmap in Supplementary Figure 7 (B). A high PERR indicates that in that area, the proportion of public engagement with preprints is relatively higher compared to peer-reviewed papers, showing greater public interest in preprint quotations.
**Abbreviations:** PERR, Public Engagement Relative Ratio.

**Supplementary Table 8. Average of Citation Counts Per Region and Income group**

|  | **Median value (Q1–Q3)** | | **Mean value (SD)** | |  |
| --- | --- | --- | --- | --- | --- |
|  | **Peer-reviewed** | **Preprint** | **Peer-reviewed** | **Preprint** |  |
| **Region** | | | | | **p-value** |
| East Asia & Pacific | 3 (0-12) | 1 (0-7) | 26.37 (244.82) | 9.33 (28.63) | < 0.001 |
| South Asia | 2 (0-8) | 1 (0-4) | 10.16 (37.18) | 3.63 (8.42) | < 0.001 |
| Central Asia | 2 (0-8) | 1 (0-4) | 11.71 (57.50) | 4.18 (9.09) | < 0.001 |
| Europe | 3 (1-12) | 1 (0-4) | 17.95 (94.45) | 5.27 (16.99) | < 0.001 |
| Latin America & Caribbean | 2 (0-9) | 1 (0-3) | 12.68 (86.40) | 3.98 (18.77) | < 0.001 |
| North America | 3 (1-12) | 1 (0-5) | 18.79 (85.42) | 5.94 (16.64) | < 0.001 |
| Middle East & North Africa | 3 (1—11) | 2 (0-6) | 13.79 (47.96) | 5.99 (15.59) | < 0.001 |
| Sub-Saharan Africa | 2 (0-8) | 1 (0-3) | 11.09 (49.25) | 4.38 (15.20) | < 0.001 |
| **Income Group** | | | | | **p-value** |
| High | 3 (1–12) | 1 (0–5) | 18.40 (91.91) | 5.59 (16.58) | < 0.001 |
| Upper-middle | 3 (0–11) | 2 (0–6) | 24.91 (247.87) | 8.31 (27.98) | < 0.001 |
| Lower-middle | 2 (0–8) | 1 (0–4) | 9.85 (37.34) | 3.85 (11.70) | < 0.001 |
| Low | 3 (0–9) | 1 (0–3) | 10.04 (25.06) | 3.55 (12.61) | < 0.001 |

Descriptive statistics of papers’ citation counts by region and income group for both peer-reviewed papers and preprints are presented, classified into 8 regions and 4 income groups. The p-value column shows Wilcox rank-sum test results, all indicating statistical significance. Peer-reviewed papers consistently receive higher median citations than preprints across all regions and income groups. East Asia & Pacific and Europe have the highest median citations for peer-reviewed papers, while South Asia and Central Asia have lower values. High-income and upper-middle-income countries both show a median citation value of 3, while lower-middle and low-income groups have values of 2 and 3, respectively. Preprints generally have lower median citations, with most regions showing a median value of 1, except for the Middle East & North Africa, which has a value of 2. High-income countries have a median citation value of 1, upper-middle-income countries 2, and lower-middle and low-income groups a value of 1.
**Abbreviatons:** SD, Standard Deviation.

**Supplementary Table 9. Income group transition of lead author’s country from preprint to peer-reviewed paper (Column: Peer-reviewed paper, Row: Preprint)**

|  | Low | Lower-middle | Upper-middle | High | Total |
| --- | --- | --- | --- | --- | --- |
| Low | 22 | 1 | 2 | 26 | 51 |
| Lower-middle | 9 | 209 | 29 | 164 | 411 |
| Upper-middle | 2 | 21 | 770 | 224 | 1,017 |
| High | 24 | 119 | 278 | 4,317 | 4,738 |
| Total | 57 | 350 | 1,079 | 4,731 |  |

The transition of lead authors' country income groups from preprints to peer-reviewed papers is detailed. Preprints by researchers from lower-income countries often get published in peer-reviewed journals with lead authors from high-income countries. Notably, 26 out of 51 preprints (51%) with lead authors from low-income countries transitioned to having high-income lead authors in peer-reviewed publications. Similarly, 164 out of 411 preprints (40%) from lower-middle-income countries experienced the same transition. This pattern is consistent with previous studies suggesting that authors from low-income groups, such as African researchers, frequently remain as middle authors in traditional research papers.

**Supplementary File 1. Country and region information**

**Supplementary File 2. Inter-country co-authorship networks for peer-reviewed papers and preprints**

**Supplementary File 3. Interactive visualizations for scatter plots, geographic heatmaps, and bar plots**

**Supplementary Appendix 1. Search queries**

TITLE-ABS-KEY(sars-cov-2 OR ‘coronavirus 2’ OR ‘corona virus 2’ OR covid-19 OR {novel coronavirus} OR {novel corona virus} OR 2019-ncov OR covid OR covid19 OR ncovid-19 OR ‘coronavirus disease 2019’ OR ‘corona virus disease 2019’ OR corona-19 OR SARS-nCoV OR ncov-2019)
